# Bayesian Inference of State-Level COVID-19 Basic Reproduction Numbers across the United States

**DOI:** 10.1101/2021.09.27.21264188

**Authors:** Abhishek Mallela, Jacob Neumann, Ely F. Miller, Ye Chen, Richard G. Posner, Yen Ting Lin, William S. Hlavacek

## Abstract

Although many persons in the United States have acquired immunity to COVID-19, either through vaccination or infection with SARS-CoV-2, COVID-19 will pose an ongoing threat to non-immune persons so long as disease transmission continues. We can estimate when sustained disease transmission will end in a population by calculating the population-specific basic reproduction number ℛ_0_, the expected number of secondary cases generated by an infected person in the absence of any interventions. The value of ℛ_0_ relates to a herd immunity threshold (HIT), which is given by 1 − 1/ℛ_0_. When the immune fraction of a population exceeds this threshold, sustained disease transmission becomes exponentially unlikely (barring mutations allowing SARS-CoV-2 to escape immunity). Here, we report state-level ℛ_0_ estimates obtained using Bayesian inference. Maximum *a posteriori* estimates range from 7.1 for New Jersey to 2.3 for Wyoming, indicating that disease transmission varies considerably across states and that reaching herd immunity will be more difficult in some states than others. ℛ_0_ estimates were obtained from compartmental models via the next-generation matrix approach after each model was parameterized using regional daily confirmed case reports of COVID-19 from 21-January-2020 to 21-June-2020. Our ℛ_0_ estimates characterize infectiousness of ancestral strains, but they can be used to determine HITs for a distinct, currently dominant circulating strain, such as SARS-CoV-2 variant Delta (lineage B.1.617.2), if the relative infectiousness of the strain can be ascertained. On the basis of Delta-adjusted HITs, vaccination data, and seroprevalence survey data, we find that no state has achieved herd immunity as of 20-September-2021.

**Significance Statement:** COVID-19 will continue to threaten non-immune persons in the presence of ongoing disease transmission. We can estimate when sustained disease transmission will end by calculating the population-specific basic reproduction number ℛ_0_, which relates to a herd immunity threshold (HIT), given by 1 − 1/ℛ_0_. When the immune fraction of a population exceeds this threshold, sustained disease transmission becomes exponentially unlikely. Here, we report state-level ℛ_0_ estimates indicating that disease transmission varies considerably across states. Our ℛ_0_ estimates can also be used to determine HITs for the Delta variant of COVID-19. On the basis of Delta-adjusted HITs, vaccination data, and serological survey results, we find that no state has yet achieved herd immunity.

## Introduction

Vaccines to protect against coronavirus disease 2019 (COVID-19) became available in the United States (US) in December 2020 (1). As of September 20, 2021, 181,728,072 persons have been fully vaccinated, an additional 30,307,256 persons have been partially vaccinated, and an uncertain number of persons have acquired immunity through infection (2). The entire US population does not need to be vaccinated to end sustained COVID-19 transmission because of the phenomenon of herd immunity (3), which is reached when a critical fraction of the population becomes immune. This fraction is called the herd immunity threshold (HIT).

The HIT for a population relates to the basic reproduction number, ℛ_0_, as follows (3): HIT = 1 − 1/ℛ_0_. ℛ_0_ is defined as the expected number of secondary infections arising from a primary case in the absence of any immunity or intervention. As is well known, ℛ_0_ and HIT are population-specific (4-5), which means that the effort required to control the local COVID-19 epidemic may vary from community to community. However, knowledge of the HIT for a given region is insufficient to determine when disease transmission within the region will end. One also needs to know the fraction of the population that has immunity. Estimating the immune fraction is difficult, because we cannot simply count the number of persons who have been vaccinated or the number of persons detected to be infected. Immunity is acquired not only through vaccination but also through infection (6), and case detection is imperfect. Insight into the immune fraction can be obtained from seroprevalence surveys, which use blood tests to identify persons who have antibodies against the SARS-CoV-2 virus (acquired through vaccination or infection).

Various estimates of ℛ_0_ for transmission of COVID-19 have been provided in the literature (7). The estimates that have received the most attention are those given for China and Italy (8-12), which were among the first regions to be impacted by COVID-19. However, the relevance of these estimates for populations within the US (or elsewhere outside of China and Italy) is unclear. Several studies have estimated ℛ_0_ for the US at the national level (13-15), the state level (16-18), and the county level (19-20). The usefulness of a national estimate is unclear given the heterogeneity of the US, and none of the county-level estimates are comprehensive. Some state-level estimates are also incomplete (16, 18). Because responses to COVID-19 within the US have been and continue to be driven mainly by governors of US states (21), we undertook a study to generate comprehensive state-level ℛ_0_ estimates through Bayesian inference. With this approach, we were able to quantify uncertainty in each estimate through a parameter posterior distribution.

In earlier work, we developed a compartmental model for COVID-19 transmission dynamics that reproduces surveillance data and generates accurate forecasts for the 15 most populous metropolitan statistical areas (MSAs) in the US (22). Here, for each of the 50 states, we found a state-specific parameter posterior conditioned on this model from state-level COVID-19 surveillance data available from January 21 to June 21, 2020 (23). From these parameter posteriors, we then obtained region-specific ℛ_0_ and HIT posteriors and maximum *a posteriori* (MAP) estimates. The MAP estimates for HITs together with other data—vaccination tracking data (24), serological survey data (25-26), and quantitative estimates of the increased transmissibility of the recently introduced SARS-CoV-2 variant Delta (lineage B.1.617.2) (27-28)—provide insight into the progress of each state toward herd immunity.

## Materials and Methods

### Model

To obtain regional ℛ_0_ and HIT estimates, we used a compartmental model developed previously (22). We found region-specific parameterizations that allow the model to reproduce surveillance data (daily reports of new confirmed COVID-19 cases) available for each region of interest over a defined period (e.g., January 21 to June 21, 2020). The model is able to account for a variable number of social-distancing periods. We considered versions of the model accounting for one, two, and three social-distancing periods. The number of social-distancing periods deemed best (i.e., to provide the most parsimonious explanation of the data) for a given time period was determined using the model selection procedure described by Lin et al. (22). As in the study of Lin et al. (22), the model has 14 parameters with universal fixed values (applicable to all regions). The model also has 3(*n* + 1) + 3 parameters with region-specific adjustable values determined through Bayesian inference, where *n* + 1 denotes the number of social-distancing periods. In this study, for a given region, we censored case-reporting data whenever the cumulative reported case count was less than 10 cases. We also specified the onset time of the first social-distancing period as the earliest day on which the cumulative reported case count was 200 cases or more. A full description of model parameters is given in Lin et al. (22).

### Simulations

Each region-specific model consists of a coupled system of ordinary differential equations (ODEs), which are given by Lin et al. (22). The ODEs were numerically integrated using the SciPy (29) interface to LSODA (30) and the BioNetGen (31) interface to CVODE (32). Python code was converted to machine code using Numba (33). The initial conditions were determined as in Lin et al. (22).

### Calculation of epidemic parameters ℛ_0_ and λ

To find the basic reproduction number ℛ_0_, we considered a reduced form of the model of Lin et al. (22), which is given in Eqs. 1-8 of the Supplementary Information (SI). The reduced model omits consideration of interventions, including social distancing, quarantine, and self-isolation, which are all considered in the full model. From the reduced model, we derived an expression for ℛ_0_ by applying the next-generation matrix method (34). In this procedure, ℛ_0_ is determined as the spectral radius of the so-called next-generation matrix. Denoting this matrix as 𝒩, the (*i, j*) entry of 𝒩 is the expected number of new infections in the *i*^*th*^ compartment produced by persons initially in the *j*^*th*^ compartment. The expression for ℛ_0_ given in the Results section below was obtained using Mathematica (35). The matrix 𝒩 was obtained using Mathematica’s *LinearSolve* function and ℛ_0_ was computed as the dominant eigenvalue of 𝒩.

To characterize the initial rate of exponential growth for a local epidemic within a given region, we computed the epidemic growth rate λ as the dominant eigenvalue of the Jacobian of the reduced model linearized at the disease-free equilibrium (36). The derivation of λ is provided in the SI.

### Bayesian inference

To infer region-specific values of adjustable model parameters (and ℛ_0_ and HIT estimates), we followed the Bayesian inference approach of Lin et al. (22). In inferences, we used all region-relevant confirmed COVID-19 case-count data available in the GitHub repository maintained by *The New York Times* newspaper (23) for the period starting on 21-January-2020 and ending on 21-May-2020, 21-June-2020, or 21-July-2020 (inclusive dates). Markov Chain Monte Carlo (MCMC) sampling was performed using the Python code of Lin et al. (22) and a new release of PyBioNetFit (37), version 1.1.9, which includes an implementation of the adaptive MCMC method used in the study of Lin et al. (22). Inference job setup files for PyBioNetFit, including data files, are provided for each of the 50 states online (https://github.com/lanl/PyBNF/tree/master/examples/Mallela2021States). Results from both methods were found to be consistent (SI Fig S1). To ensure that MCMC sampling procedures converged, we visually inspected trace plots for log-likelihood (SI Fig S2) and parameters (SI Fig S3) and pairs plots (SI Fig S4). We also performed simulations using maximum likelihood estimates (MLEs) for parameter values to assess agreement of the simulations with the training data (SI Fig S5).

The maximum *a posteriori* (MAP) estimate of a parameter is the value of the parameter corresponding to the peak of its marginal posterior distribution, where probability density is highest. Because we assumed a proper uniform prior distribution for each of the adjustable parameters, as in the study of Lin et al. (22), the MAP estimates are MLEs.

## Results

### Bayesian uncertainty quantification

Following the Bayesian inference approach of Lin et al. (22), we quantified uncertainty in predicted trajectories of confirmed case counts for all 50 states, using data from January 21 to June 21, 2020. As illustrated in Fig 1 for the states of New Jersey, Wyoming, Florida, and Alaska, we find that each region-specific model parameterized on the basis of our MCMC sampling procedure reproduces the corresponding surveillance data over the period of interest. Results for the remaining states are shown in SI Fig S5. At the end of each MCMC sampling procedure, we obtained a marginal posterior distribution for *β* (the rate constant in the model for disease transmission) which provides a probabilistic characterization of region-specific SARS-CoV-2 transmissibility. If the marginal posterior is narrow, we have high confidence in the MAP estimate of *β*; if it is wide, we have less confidence in its value. Each state-specific marginal posterior yields a MAP estimate for *β*.

**Figure 1.**
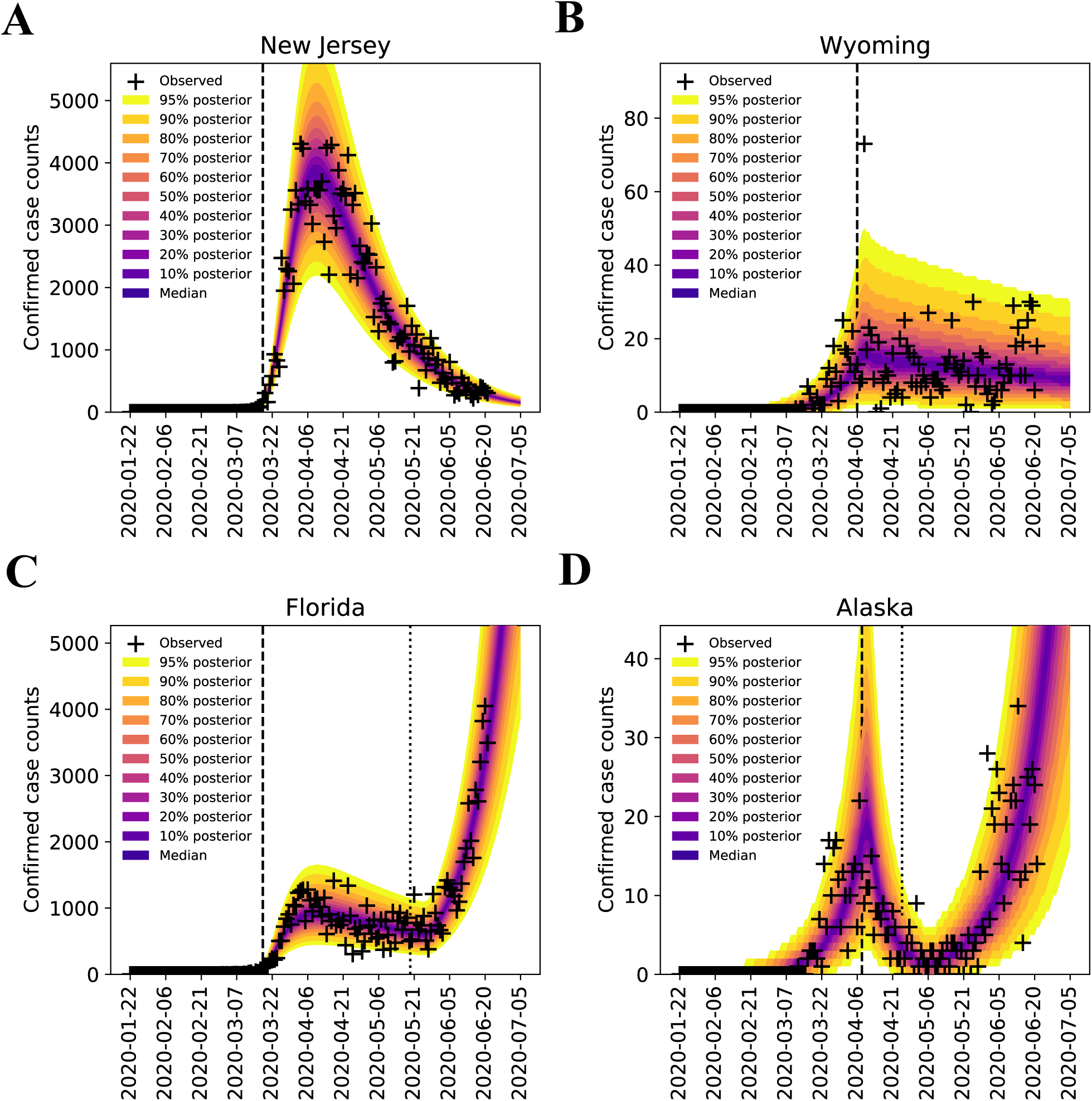
Bayesian predictive inferences for daily confirmed case counts of COVID-19 in (A) New Jersey (B) Wyoming (C) Florida (D) Alaska, from January 21 to June 21, 2020 (inclusive dates). The compartmental model (22) accounts for an initial social distancing period followed by *n* additional periods. We considered *n =* 0, 1 and 2 and selected the best *n* using the model selection procedure of Lin et al. (22). Plus signs indicate daily case reports. The shaded region indicates the prediction uncertainty and inferred noise in detection of new cases. The color-coded bands within the shaded region indicate the median and different credible intervals (e.g., dark purple band corresponds to the median, the band with lightest shade of yellow corresponds to the 95% credible interval, and gradations of color between these two extremes correspond to different credible intervals as indicated in the legend). In each panel, the vertical broken line indicates the onset time of the first social-distancing period. For states with *n* = 1 (Alaska and Florida), there is an additional broken line, which indicates the onset time of the second social-distancing period. The model was used to make forecasts of new case detection for 14 days after June 21, 2020. The last prediction date was July 5, 2020.

We can propagate the uncertainty in *β* into uncertainty in ℛ_0_ and HIT estimates, using the formula for ℛ_0_ given below and HIT = 1 − 1/ℛ_0_. In Fig 2, we show marginal posterior distributions for ℛ_0_ and HIT for the states of New Jersey, Wyoming, Florida, and Alaska. We provide MAP estimates of the model parameters for all states in SI Table S1. Model parameters were found to be identifiable in practice. (We have no proof of identifiability.) MAP estimates for ℛ_0_ and HIT for all 50 states are provided in SI Table S2. These tables also provide 95% credible intervals. These estimates characterize the infectiousness of SARS-CoV-2 ancestral strains in each region of interest.

**Figure 2.**
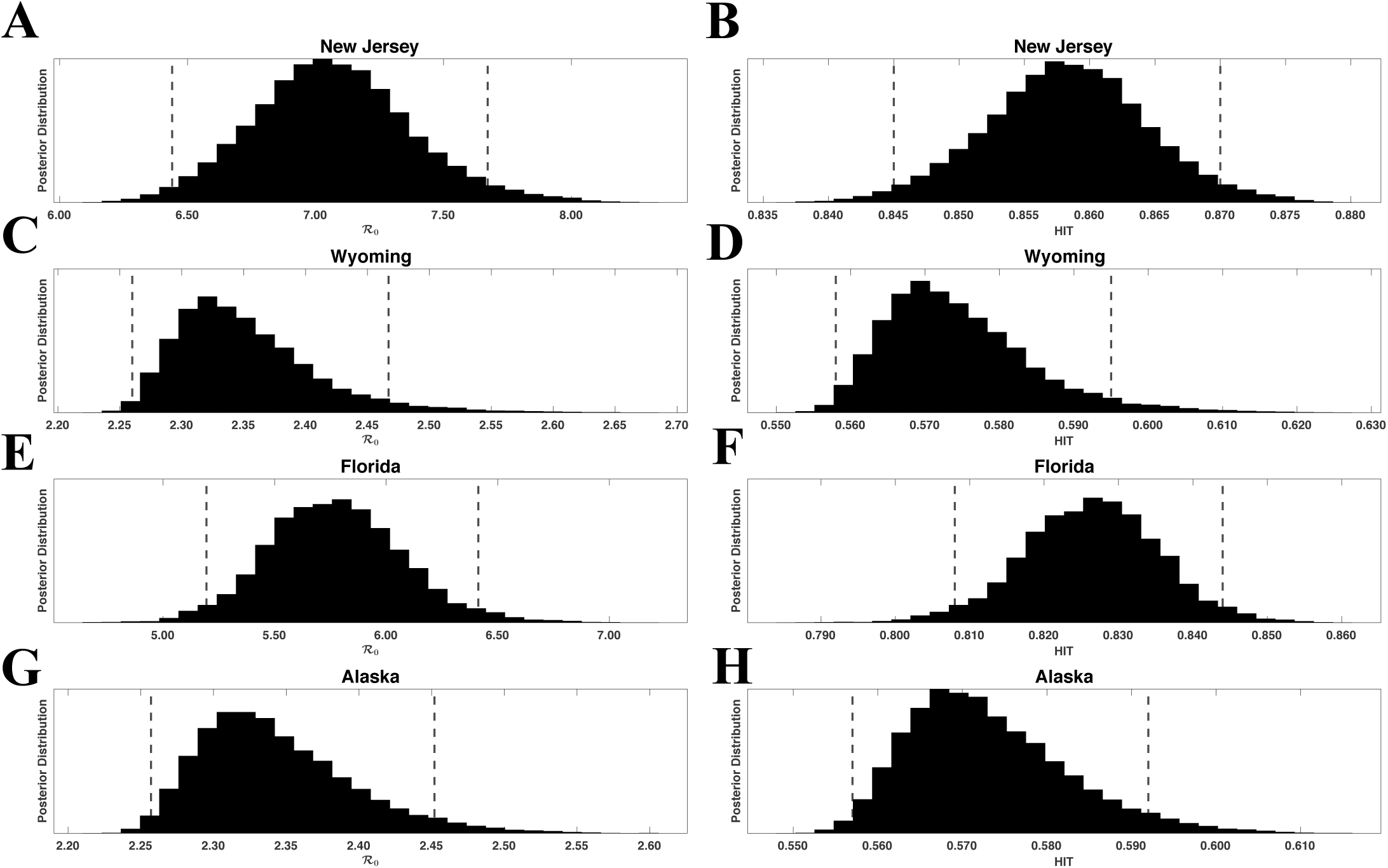
Marginal posterior distributions of ℛ_0_ (left panels) and HIT (right panels) for ancestral strains of SARS-CoV-2 in four US states: (A, B) New Jersey, (C, D) Wyoming, (E, F) Florida, and (G, H) Alaska. Inferences are based on daily reports of new cases from January 21 to June 21, 2020. Each ℛ_0_ posterior was obtained from the corresponding marginal posterior for β and Eq. 1. Each HIT posterior was obtained from the relation HIT *= 1* − *1*/ℛ_0_ and the corresponding marginal posterior for ℛ_0_. The 95% credible intervals for ℛ_0_ are as follows: (6.44, 7.67) for New Jersey, (2.26, 2.47) for Wyoming, (5.20, 6.41) for Florida, and (2.26, 2.45) for Alaska. The 95% credible intervals for the HIT estimates are as follows: (0.84, 0.87) for New Jersey, (0.56, 0.59) for Wyoming, (0.81, 0.84) for Florida, and (0.56, 0.59) for Alaska. For each panel, the endpoints of the corresponding credible interval are indicated with vertical broken lines.

### Region-specific basic reproduction numbers and herd immunity thresholds

To calculate the herd immunity threshold (HIT) for a specific region, we need to know the corresponding region-specific value of the basic reproduction number ℛ_0_, which is given by the following formula (obtained as described in Materials and Methods and SI):

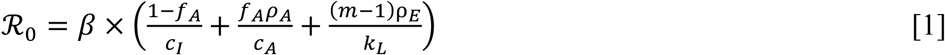

where *β* characterizes the rate of transmission attributable to contacts between persons who are not protected by social distancing, *f*_*A*_ denotes the fraction of infected persons who never develop symptoms (i.e., the fraction of asymptomatic cases), *c*_*A*_ characterizes the rate at which asymptomatic persons recover during the immune clearance phase of infection, *c*_*I*_ characterizes the rate at which symptomatic persons with mild disease recover or progress to severe disease, *ρ*_*E*_ is a constant characterizing the relative infectiousness of presymptomatic persons compared to symptomatic persons (with the same behaviors), *ρ*_*A*_ is a constant characterizing the relative infectiousness of asymptomatic persons compared to symptomatic persons (with the same behaviors), *m* denotes the number of stages in the incubation period, and *k*_*L*_ characterizes disease progression, from one stage of the incubation period to the next and ultimately to an immune clearance phase. The value of ℛ_0_ depends on one inferred region-specific parameter, *β*, and seven fixed parameters, which have values taken to be applicable for all regions (i.e., *f*_*A*_, *c*_*A*_, *c*_*I*_, *ρ*_*E*_, *ρ*_*A*_, *k*_*L*_, and *m*). Estimates of these fixed parameters were taken from Lin et al. (22).

The SARS-CoV-2 variant Delta (lineage B.1.617.2) has been estimated to be 1.64 times more infectious than variant Alpha (lineage B.1.1.7) (28), which has been estimated to be 1.50 times more infectious than ancestral strains (27). Assuming that Delta is the dominant circulating SARS-CoV-2 strain throughout the US (as of September 20, 2021) and that *β* for Delta is 1.64 × 1.50 = 2.46 times greater than *β* for ancestral strains (with other parameters in Eq. 1 remaining the same), the MAP estimate of the Delta-adjusted ℛ_0_ ranges from 5.6 for Wyoming to 18 for New Jersey (from the multiplier given above and SI Table S2). The population-weighted Delta-adjusted ℛ_0_ for the US is 12. These estimates indicate that the herd immunity threshold (HIT) for the Delta variant of SARS-CoV-2 ranges from 82% to 94%.

### Estimates of initial region-specific epidemic growth rates

HIT estimates are directly determined by estimates of the basic reproduction number, which are related to the initial growth rate of the epidemic in a given region. Here, our ℛ_0_ estimates are conditioned on a compartmental model that has been parameterized to reproduce case-reporting data available for each region over a five-month period (January 21 to June 21, 2020). We can use parameter estimates obtained for each region to calculate the initial epidemic growth rate λ, which is directly comparable to early surveillance data (Fig 3 and SI Fig S6). We provide MAP estimates and 95% credible intervals for λ, ℛ_0_, and HIT for selected states in Table 1. MAP estimates and 95% credible intervals for λ, ℛ_0_, and HIT for all states are provided in SI Table S2. These estimates are based on state-specific marginal posteriors for the parameter *β* of our compartmental model. State-specific MAP estimates and 95% credible intervals for *β* (and other adjustable model parameters) are given in SI Table S1. As can be seen (e.g., in Fig 3), our λ estimates are consistent with early case reporting data during the exponential takeoff phase of disease transmission.

**Figure 3.**
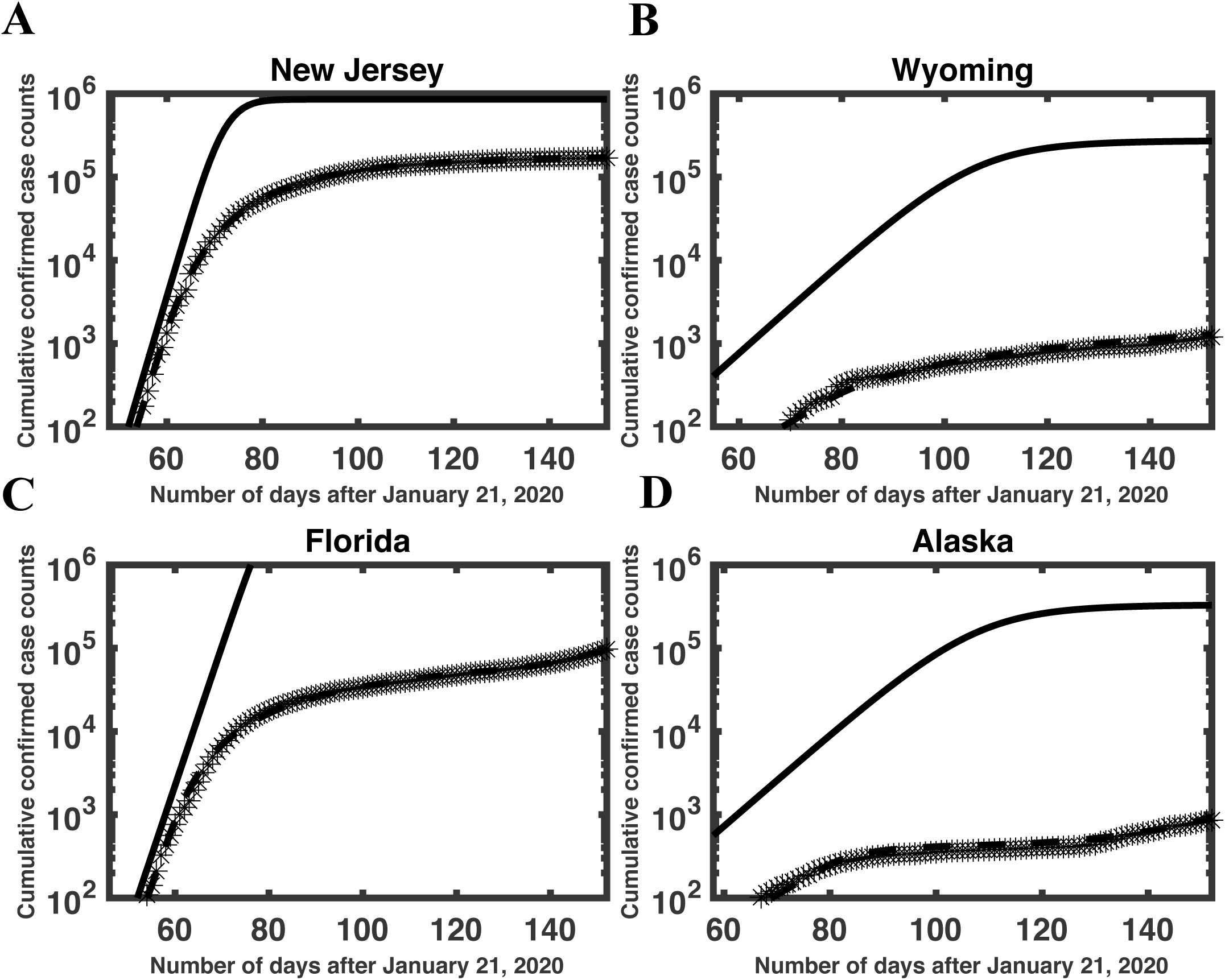
Consistency of model-derived λ estimates with empirical growth rates during initial exponential increase in disease incidence in (A) New Jersey, (B) Wyoming, (C) Florida, and (D) Alaska. In each panel, the initial slope of the solid curve corresponds to λ (calculated as described in Materials and Methods), the crosses indicate empirical cumulative case counts, and the broken line is the model prediction based on MAP estimates for adjustable parameters. The solid curve is derived from the reduced model (Eqs. 1-8 in the SI). This curve shows cumulative case counts had there not been any interventions to limit disease transmission. As can be seen, the initial slopes of the solid and broken curves are comparable. We selected *n =* 0 for New Jersey and Wyoming and *n* = 1 for Florida and Alaska. Among 35 states with *n =* 0, New Jersey has the largest inferred λ value (0.45) and Wyoming has the smallest inferred λ value (0.13). Among 15 states with *n* = 1, Florida has the largest inferred value of λ (0.39) and Alaska has the smallest inferred value of λ (0.13). It should be noted that, in contrast with Fig 1, the y-axis here indicates cumulative (vs. daily) number of cases on a logarithmic (vs. linear) scale.

**Table 1.** Maximum *a posteriori* (MAP) estimates and 95% credible intervals for epidemic parameters (β, λ, ℛ_0_, HIT, and Delta-adjusted HIT) for the states of New Jersey, Wyoming, Florida, and Alaska.

| State | $\beta$ ($d^{-1}$) | $\lambda$ ($d^{-1}$)* | $\mathcal{R}_0$** | HIT*** | Delta-adjusted HIT**** |
| --- | --- | --- | --- | --- | --- |
| New Jersey | 0.65 (0.59-0.71) | 0.45 (0.41-0.48) | 7.1 (6.4-7.7) | 0.86 (0.84-0.87) | 0.94 (0.94-0.95) |
| Wyoming | 0.21 (0.21-0.23) | 0.13 (0.13-0.15) | 2.3 (2.3-2.5) | 0.56 (0.56-0.59) | 0.82 (0.82-0.84) |
| Florida | 0.55 (0.48-0.59) | 0.39 (0.34-0.41) | 6.0 (5.2-6.4) | 0.83 (0.81-0.84) | 0.93 (0.92-0.94) |
| Alaska | 0.21 (0.21-0.23) | 0.13 (0.13-0.14) | 2.3 (2.3-2.5) | 0.56 (0.56-0.59) | 0.82 (0.82-0.84) |

In this analysis, we used surveillance data (daily reports of new cases) available from 21-January-2020 to 21-June-2020 (inclusive dates) to estimate parameter values through Bayesian inference. \*Computed as described in SI. \*\*Calculated using Eq. 1. \*\*\*Obtained through the relation $HIT = 1 - 1/\mathcal{R}_0$ . \*\*\*\*Based on Delta being 2.46 times more infectious than ancestral strains.

### Sensitivity of *β* to the surveillance data used in inference

For each state, we used data from January 21 to June 21, 2020 to infer the MAP estimate of *β* (and the values of the other region-specific adjustable model parameters). Thus, our estimates are derived from a particular subset of the available surveillance data. To check the robustness of MAP estimates for *β* to variations in training data, we performed a sensitivity analysis wherein we inferred *β* using data collected over three distinct periods in 2020: 1) January 21 to May 21, 2) January 21 to June 21, and 3) January 21 to July 21. By visualizing our estimates with a rank order plot (Fig 4) and conducting pairwise two-sample Kolmogorov-Smirnov tests (38), we found that the 4-, 5-, and 6-month training datasets yielded estimates for *β* that were not statistically significantly different from each other. The MAP estimates for *β* obtained using the 4-, 5-, and 6-month datasets are listed in SI Table S3. We assessed sensitivity by computing the relative error between the *β* estimates obtained from the 5-month dataset and the average *β* estimate over all datasets considered. We found that none of the state-level MAP estimates for *β* showed sensitivity (i.e., a relative error exceeding 100% in magnitude) to variations in the training data (SI Table S4). The largest relative error was 12% (for Kansas).

**Figure 4.**
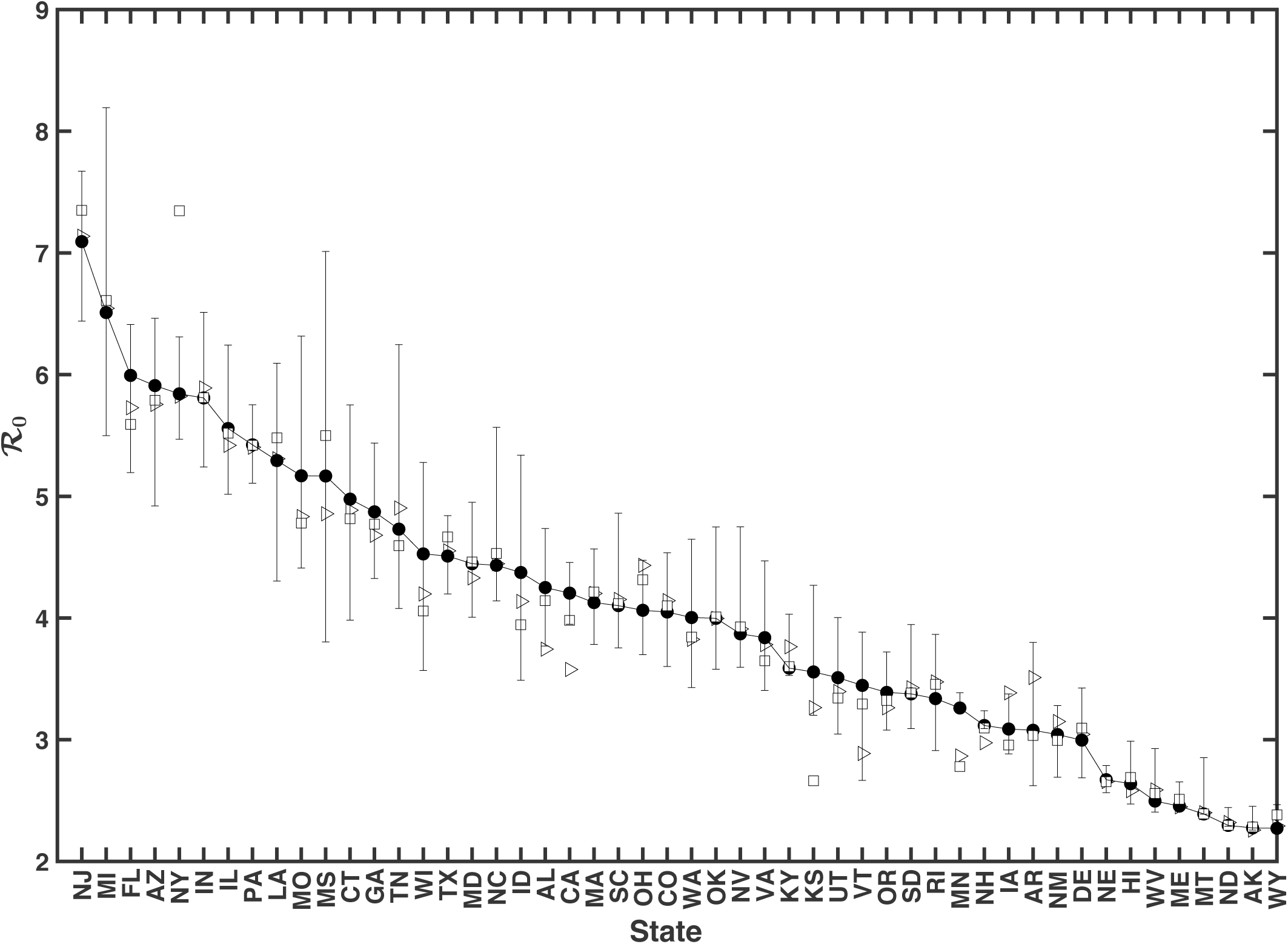
MAP estimates of the basic reproduction number ℛ_0_ for ancestral strains of SARS-CoV-2 in all 50 US states. The different symbols refer to different training datasets used to estimate ℛ_0_. Open triangles correspond to surveillance data collected from January 21 to May 21, 2020, filled circles correspond to surveillance data collected from January 21 to June 21, 2020, and open squares correspond to surveillance data collected from January 21 to July 21, 2020. Estimates of ℛ_0_ are sorted by state from largest to smallest values according to the ℛ_0_ estimates derived from the surveillance data collected for January 21 to June 21, 2020. The whiskers associated with each filled circle indicates the 95% credible interval (inferred from the 5-month dataset). States are indicated using two-letter US postal service state abbreviations (https://about.usps.com/who-we-are/postal-history/state-abbreviations.pdf).

### Global asymptotic stability of the disease-free equilibrium

The model of Lin et al. (22) has a globally asymptotically stable disease-free equilibrium (DFE) if ℛ_0_ < 1, which can be deduced by following the approach of Shuai and van den Driessche (39). As a consequence, the model predicts that the epidemic will be extinguished as the system dynamics are attracted to the DFE.

To confirm that the model behaves as expected around the HIT, we conducted a perturbation analysis for the states of New York (Figs 5A and 5B) and Washington (Figs 5C and 5D). We simulated disease dynamics starting from an arbitrarily chosen initial condition near the HIT number of persons, *S*_*h*_, given by the following formula: *S*_*h*_ = HIT × *S*_0_, where *S*_0_ denotes the population size of the region considered. We defined the size of our perturbation as ε = 0.2 × *S*_*h*_ for Figs 5A and 5C and as *ε* = −0.2 × *S*_*h*_ for Figs 5B and 5D. The initial condition was *S*_0_ − *S*_*h*_ − 1 + ε susceptible persons, 1 infected person, and *S*_*h*_ − *ε* recovered persons. As expected, for *S*_*h*_ < HIT × *S*_0_ (Figs 5A and 5C), the number of infectious persons grows over time, whereas for *S*_*h*_ > HIT × *S*_0_ (Figs 5B and 5D), the number of infectious persons decays over time.

**Figure 5.**
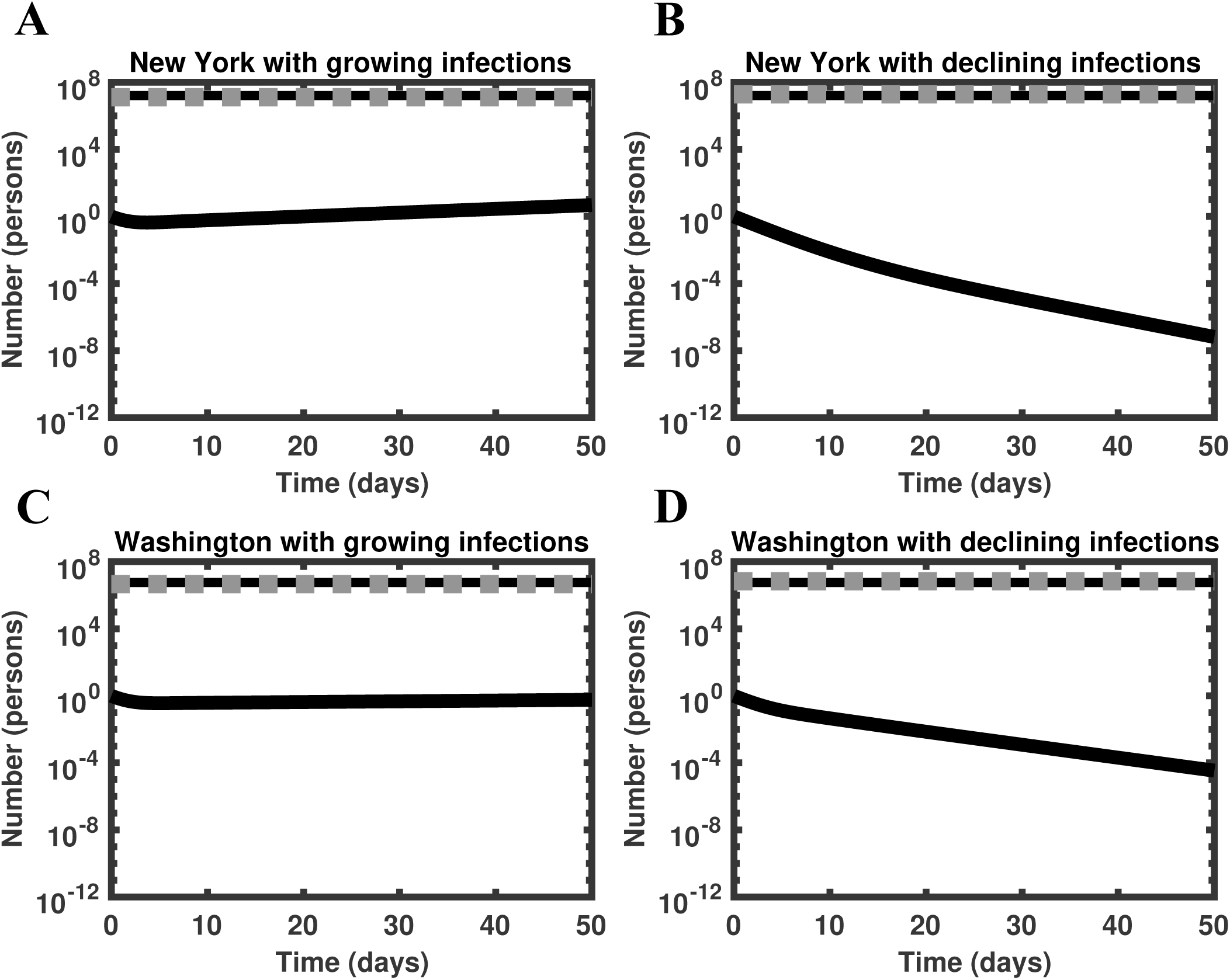
Perturbation analysis using the full model of Lin et al. (22) for the states of New York (panels A and B) and Washington (panels C and D). In each panel, the black solid line represents the number of infectious persons (initially 1), the black broken line represents the threshold number of persons required for herd immunity (i.e., *S*_*h*_), and the gray broken line represents the number of recovered persons (initially *S*_*h*_ − *ε*, obtained as described in Results). Simulations are based on MAP estimates for model parameters obtained using surveillance data collected from January 21 to June 21, 2020.

### Progress toward herd immunity

From our state-specific HIT estimates and other information (discussed below), we were able to calculate percent progress toward herd immunity for each state (Fig 6, SI Table S5). We estimated the percent progress of each state’s population toward herd immunity, 𝒫 ∈ [0%, 100%], using the following equation (the derivation of which is given in the SI):

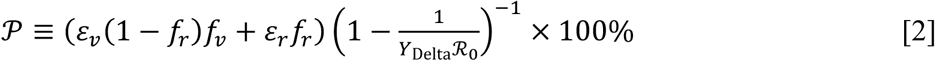

**Figure 6.**
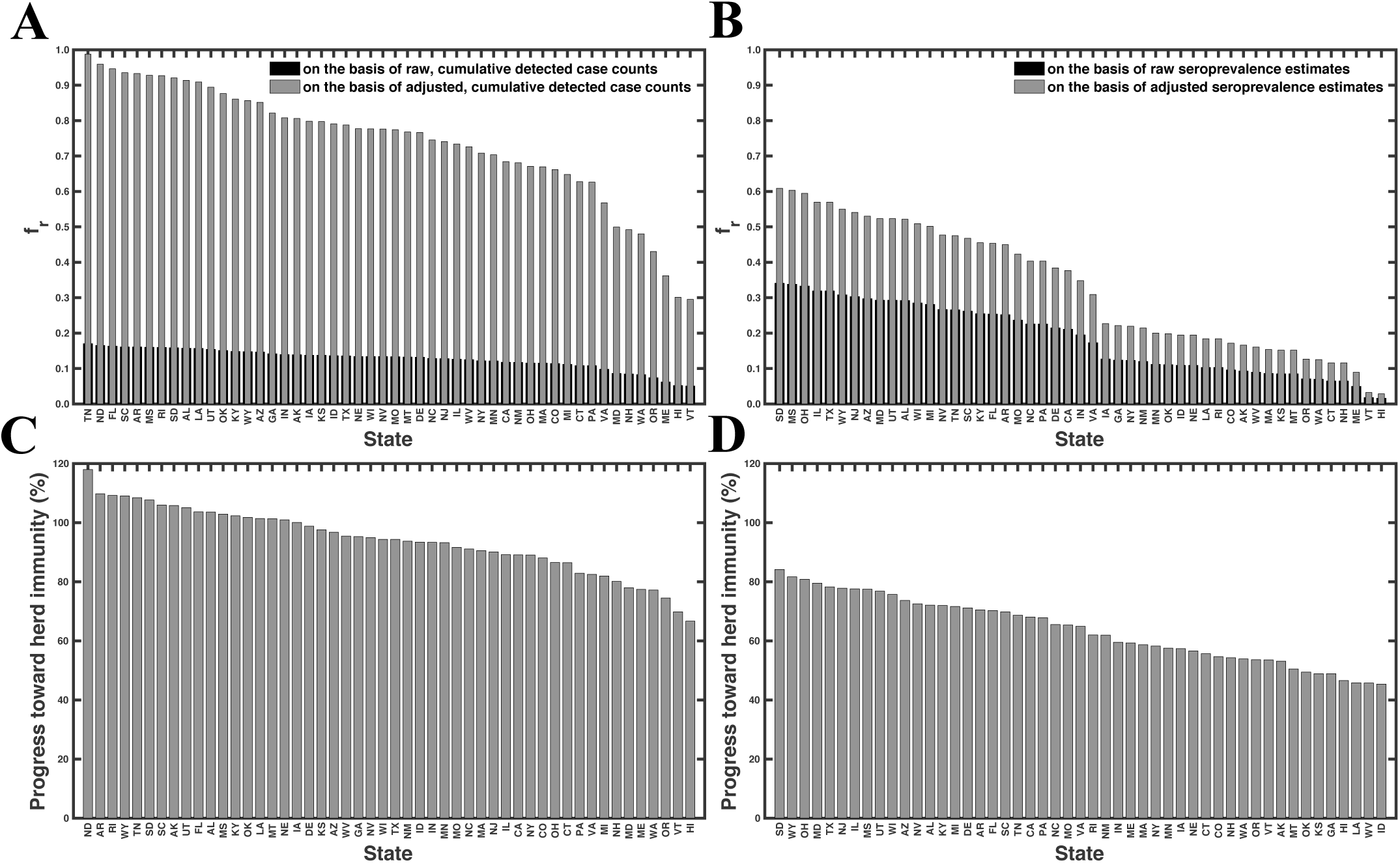
Percent progress toward herd immunity in each of the 50 US states. Percent progress 𝒫indicates the fraction of immune persons required for herd immunity. 𝒫 was calculated using Eq. 2. Black bars (Panel A) correspond to the first scenario (i.e., *f*_*r*_ estimated as the number of detected cases divided by population size), gray bars (Panels A and C) correspond to the second scenario (i.e., *f*_*r*_ estimated as the number of detected cases within a population divided by the population size, adjusted for lack of detection of undiagnosed SARS-CoV-2 infections), black bars (Panel B) correspond to the third scenario (i.e., *f*_*r*_ given by seroprevalence survey results), and gray bars (Panels B and D) correspond to the fourth scenario (i.e., *f*_*r*_ given by seroprevalence survey results adjusted for lack of detection of asymptomatic cases). Estimates for 𝒫 are sorted by state from largest to smallest values according to the second scenario (Panels A and C) and the fourth scenario (Panels B and D). North Dakota was omitted from Panels B and D because a recent estimate of seroprevalence was not available at Ref. (25). States are indicated using two-letter US postal service state abbreviations (https://about.usps.com/who-we-are/postal-history/state-abbreviations.pdf).

where ℛ_0_ is the population-specific basic reproduction number that we estimated for ancestral strains (SI Table S2), *Y*_Delta_ is a multiplier that accounts for the increased transmissibility of SARS-CoV-2 variant Delta, *f*_*r*_ denotes the fraction of the population with immunity acquired through infection, *f*_*v*_ is the fraction of the population that has been vaccinated (24), *ε*_*r*_ is the fraction of infected persons who are protected against productive infection (i.e., an infection that can be transmitted to others), and *ε*_*v*_ is the fraction of vaccinated persons who are protected against productive infection. Recall that we use *Y*_Delta_ = 2.46 (27-28). We estimate that *ε*_*r*_ = 1.0 (40) and *ε*_*v*_ = 0.66 (41). We obtain four different estimates for *f*_*r*_ as follows. In the first case, we obtain *f*_*r*_ as the cumulative number of detected cases within a population divided by the population size. In the second case, we adjust our previous estimate for *f*_*r*_ by a multiplier of 5.8 (42). In other words, we assume that the true disease burden is 5.8 times higher than the detected number of cases. In the third case, we obtain *f*_*r*_ as the fraction of the population that has been infected according to the latest serological survey results reported online at Ref. (25). In the fourth case, we assume *f*_*r*_ = *f*_*r*,0_ /(1 − *f*_*A*_), where *f*_*r*,0_ denotes the estimate of seroprevalence in a given region and *f*_*A*_ denotes the fraction of all cases that are asymptomatic. With this approach, we are assuming that asymptomatic cases are not detected in serological testing (43). We adopt the estimate of Lin et al. (22) that *f*_*A*_ = 0.44.

As can be seen in Fig 6C, which is based on case reporting data, 18 of the 50 states have reached herd immunity. However, in Fig 6D, which is based on serological survey data, none of the states have reached herd immunity. South Dakota is closest to herd immunity, with 84% of the immune persons required for herd immunity. Idaho is furthest from herd immunity, with 45% of the immune persons required for herd immunity. The mean (median) progress toward herd immunity, across all states, is 63% (63%).

## Discussion

One of our most important findings is quantification of how COVID-19 transmissibility, in terms of the basic reproduction number ℛ_0_, varies across the 50 US states. The MAP value of ℛ_0_ for ancestral strains of SARS-CoV-2 ranges from 2.3 for Wyoming to 7.1 for New Jersey. The population-weighted mean for the US is 4.7. These estimates indicate that the herd immunity threshold (HIT) for the Delta variant of SARS-CoV-2 ranges from 82% to 94%, assuming that Delta is 2.46 times more transmissible than ancestral strains. The uncertainty in each ℛ_0_ estimate was quantified: 95% credible intervals are indicated in Figure 4. The 95% credible intervals for ancestral HIT estimates are given in SI Table S2. Because we can estimate the relative effort required to reach herd immunity across the US (in terms of HIT), resources for vaccination campaigns can be targeted to those areas where it is more difficult to achieve herd immunity.

Our ℛ_0_ and HIT estimates differ from estimates given in previous studies. For example, various researchers derived point estimates for ℛ_0_ from data using tools from time-series analysis, without assuming an underlying mechanistic model (13, 15). These tools depend on slope estimation and thus can be expected to depend sensitively on noise and errors in early case-reporting data. Ives and Bozzuto (16) provided state-level estimates for ℛ_0_ (in 36 states), and Fellows et al. (17) used a Bayesian framework to obtain state-level estimates for ℛ_0_ (in all 50 states). For the 30 states that are considered in Ives and Bozzuto (16), Fellows et al. (17), Milicevic et al. (18), and the present study, our estimates for ℛ_0_ were most similar to those of Milicevic et al. (18) (SI Table S6). Milicevic et al. (18) provided state-level ℛ_0_ point estimates (for 45 states) that are statistically consistent with our MAP estimates of ℛ_0_ for ancestral strains of SARS-CoV-2. The main points of difference between these earlier studies and the present study are as follows. Our ℛ_0_ and HIT estimates were obtained from a model consistent with new case-reporting data, as illustrated in Figs 1 and 3. We were able to provide estimates for all 50 states (Fig 4, SI Table S2), and we were able to obtain a Bayesian quantification of the uncertainty in each estimate (Fig 4, SI Table S2).

In the face of Delta, the estimates of Fig6C (based on case reporting data) suggest that a majority of states have yet to achieve herd immunity, and the estimates of Fig 6D (based on serological survey results) suggest that no state in the US has achieved herd immunity as of September 20, 2021. In either case, persons in the US lacking immunity are still at risk (44). The perspective provided by Fig 6D is consistent with the study of Moghadas et al. (45) indicating that only 62% of persons in the US had some form of immunity as of July 15, 2021 (either through infection or vaccination). Given that the percentage of immune persons required for herd immunity according to Fig 6D ranges from 84% for South Dakota to 45% for Idaho (Fig 6D) ∼20 months (counting from January 2020) into the COVID-19 pandemic and ∼9 months after vaccines became widely available, it seems that this situation will persist for months, if not years. How can the US accelerate the approach to herd immunity?

Policies that encourage infection of children and vaccinated persons who have healthy immune systems may be rationalized because such persons seem to be well-protected against severe (but not mild) disease (46) and infected persons seem to have greater protection against productive infection (40). However, this approach has obvious drawbacks, starting with the risks of infection. Another is that non-immune persons may not be able to self-identify as such. Unfortunately, it seems that we cannot rely on currently available vaccines to stop community transmission. Delta-adjusted HITs are mathematically impossible to achieve through vaccination alone because these HITs are close to 1 (SI Table S2) and vaccine protection against productive infection is imperfect (i.e., *ε*_*v*_ is significantly less than 1) (41). Thus, use of Delta-targeted vaccines may be needed to accelerate the approach to herd immunity and to minimize COVID-19 impacts.

One potential benefit of our comprehensive state-level ℛ_0_ estimates is that they quantify how differences in social structure and contact patterns across the US—the factors presumably underlying the spatial heterogeneity in *β* and ℛ_0_ —influence the spread of an aerosol-transmitted virus (47-48). This information, by identifying the regions in the US where transmission is likely to be highest, could be useful for responding to future pandemics caused by viruses similar to SARS-CoV-2.

Our study has several notable limitations. Our HIT estimates are potentially biased downward because of general awareness within the US of the impacts of COVID-19 in other countries (e.g., China and Italy), which could have resulted in a fraction of the US population changing their behaviors to protect themselves from COVID-19 before the start of the local epidemic. In addition, our estimation of percent progress toward herd immunity crucially depends on seroprevalence estimates of the true disease burden. These estimates are associated with some uncertainty (49-51). As illustrated in Fig 6, percent progress toward herd immunity is underestimated if serological tests fail to detect all cases of infection. The reader must also be cautioned that our analysis depends on a number of assumptions. For example, we considered a compartmental model in which populations are taken to be well-mixed and to lack age structure. This is clearly a simplification. More refined estimates could be obtained by making the model more realistic, but this would have the drawback of increasing the complexity of inference, which at some point would make inference impracticable.

## Data Availability

Inferences were obtained using problem-specific code. The functionality of the code has been added to a freely available open-source software package (PyBioNetFit, version 1.1.9). We have confirmed that the results of the problem-specific code are reproduced by PyBioNetFit.

https://github.com/lanl/PyBNF

## Author contributions

A.M., R.G.P, Y.T.L., and W.S.H. designed research; A.M., J.N., E.F.M., Y.C., R.G.P., Y.T.L., and W.S.H. performed research; A.M., J.N., Y.T.L., and W.S.H. analyzed data; and A.M. and W.S.H. wrote the paper.

## Acknowledgements

A.M. was supported by the 2020 Mathematical Sciences Graduate Internship program, which is sponsored by the Division of Mathematical Sciences of the National Science Foundation. E.F.M, J.N., Y.C., R.G.P, and W.S.H. were supported by NIH/NIGMS Grant R01GM111510. Y.T.L. was supported by the Laboratory Directed Research and Development (LDRD) program at Los Alamos National Laboratory. Computational resources for this study consisted of the FARM cluster, a Linux-based supercomputing cluster for the University of California at Davis, and Northern Arizona University’s Monsoon cluster, which is funded by Arizona’s Technology and Research Initiative Fund.

## Supplementary Information

### Reduced Model

We derive ℛ_0_ from a simplified form of the compartmental model of Lin et al. (1). The reduced model is obtained by omitting variables and terms for interventions, including social distancing, quarantine, and self-isolation. Thus, the reduced model describes disease transmission dynamics in the absence of interventions. The equations of the reduced model are as follows:

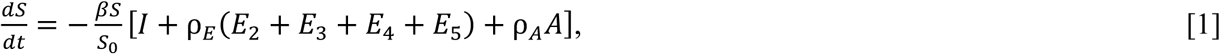

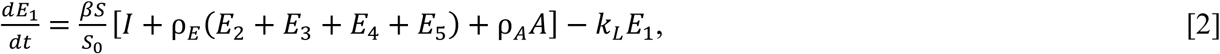

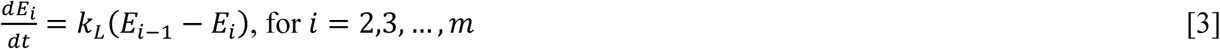

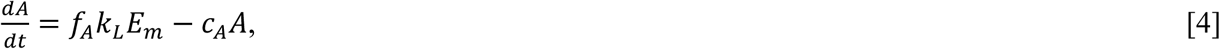

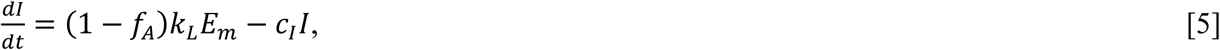

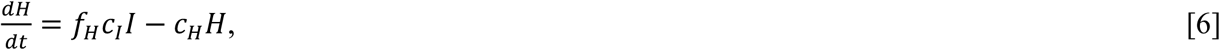

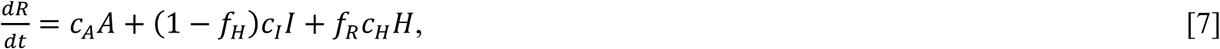

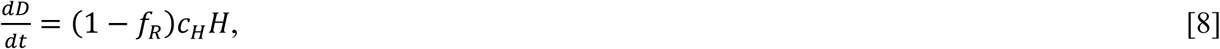

where *t* denotes time, *β, S*_0_, ρ_*E*_, ρ_*A*_, *k*_*L*_, *f*_*A*_, *f*_*H*_, *f*_*R*_, *c*_*A*_, *c*_*I*_, and *c*_*H*_ are positive-valued time-invariant parameters, as defined in Lin et al. (1), and *m* denotes the number of stages taken to comprise the incubation period. Here and in the study of Lin et al. (1), *m* = 5. The values of *β* (a rate constant characterizing disease transmission) and *S*_0_ (the total population) are taken to be region-specific; the other parameters have values that are taken to be universal (i.e., applicable to all regions of interest). The variable *S* denotes the population of susceptible persons. The variables *E*_1_ to *E*_m_ denote populations of exposed persons, e.g., persons incubating virus but not symptomatic. As noted earlier, the incubation period is divided into *m* stages. The variable *A* denotes the population of persons who have progressed through the incubation period but will never develop symptoms (i.e., persons with asymptomatic infections). The variable *I* denotes the population of persons with mild symptomatic disease. The variable *H* denotes the population of persons with severe disease who are hospitalized or isolated at home. The variable *R* denotes the population of recovered persons, and the variable *D* denotes the population of deceased persons.

### Basic Reproduction Number

The basic reproduction number, ℛ_0_, is defined as the number of secondary infections caused by an infected person during the entire period of infectiousness when introduced into a population consisting of susceptible persons only and there are no interventions to limit disease transmission. Here, we use the next-generation matrix method to compute ℛ_0_ (2). The model has a disease-free equilibrium (DFE) *x*_0_ with *S* = *S*_0_, where *S*_0_ is the total population and the remaining populations (*E*_1_, *E*_2_, …, *E*_m_, *A, I, H, R, D*) are equal to 0.

To use the next-generation matrix method, we let *x* = (*E*_1_, *E*_2_, *E*_3_, *E*_4_, *E*_5_, *A, I*) denote the vector of state variables corresponding to compartments containing infected persons. For each infected compartment *i*, we define *f*_*i*_ as the rate of entry of newly infected persons into compartment *i* and *v*_*i*_ as the net transfer of persons out of the *i*^*th*^ compartment. Then, we have *dx*_*i*_/*dt* = *f*_*i*_(*x*) – *v*_*i*_(*x*). Now, we let *F* and *V* denote the Jacobians of *f* and *v* evaluated at the disease-free equilibrium *x*_0_. The (*i, j*) entry of the matrix *F* is the rate at which infected persons in the *j*^*th*^ compartment produce a new infection in the *i*^*th*^ compartment. The (j, *k*) entry of the matrix *V*^−1^ is the expected amount of time that a person introduced to the *k*^*th*^ compartment will spend in a single visit to the *j*^*th*^ compartment. The matrix *F*, which is non-negative, is defined as follows:

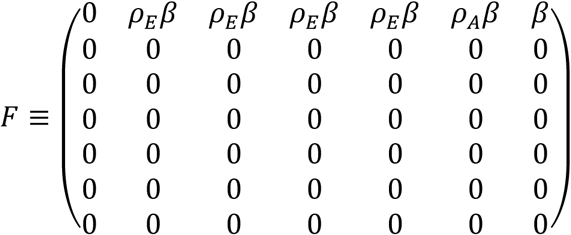

The matrix *V*, which is non-singular (i.e., invertible), is defined as follows:

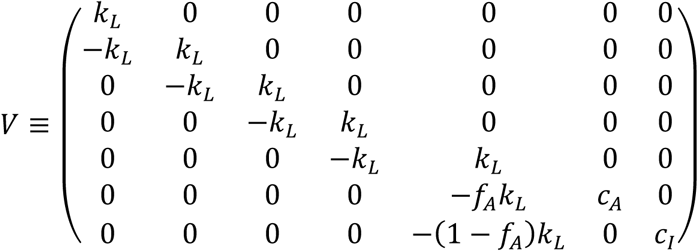

We find ℛ_0_ as the spectral radius (i.e., the dominant eigenvalue) of the matrix *FV*^−1^ (2), which is given by Eq. 1 in the main text.

### Epidemic growth rate

The epidemic growth rate λ is defined as the dominant eigenvalue of the Jacobian of the reduced model linearized at the disease-free equilibrium (DFE). Thus, λ is the largest root of the characteristic polynomial for the 7-dimensional Jacobian matrix *J*, which is equivalent to *F* − *V*. We used the *CharacteristicPolynomial* function in Mathematica (3) to find *J*:

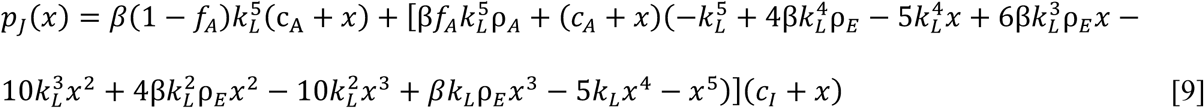

The largest root was found numerically. Solutions were based on state-specific estimates for *β* and the estimates of Lin et al. (1) for other parameters in Eq. 9.

### Progress toward herd immunity

In this section, we explain the assumptions and derive the formula for our metric of progress toward herd immunity (Eq. 2 in the main text). First, we define the variables used in our analysis. For a given region, *S*_0_ denotes the total population size, *N*_*d*_ denotes the cumulative number of cases detected, *N*_*a*_ denotes the cumulative number of asymptomatic cases, *N*_*v*_ denotes the cumulative number of vaccinations completed, *N*_*v,s*_ denotes the number of persons who were susceptible at the time of vaccination, *N*_*v,r*_ denotes the number of persons who had recovered from infection at the time of vaccination, *N*_*c*_ denotes the cumulative number of all cases, ε_*v*_ denotes the fraction of vaccinated individuals protected from productive infection (i.e., an infection that can be transmitted to others), ε_*r*_ denotes the fraction of recovered individuals protected from productive infection, *N*_*r*_ denotes the number of individuals who have recovered from infection, HIT denotes the herd immunity threshold for ancestral strains, *Y*_Delta_ denotes the infectiousness of SARS-CoV-2 variant Delta relative to ancestral strains, *S*_*h*_ ≡ HIT × *S*_0_ denotes the threshold number of persons with immunity needed for herd immunity (in the face of ancestral strains), *S*_*i*_ denotes the estimated number of persons with immunity, *f*_*A*_ ≡ *N*_*a*_/*N*_*c*_ denotes the fraction of all cases that are asymptomatic, *f*_*r*_ ≡ *N*_*r*_/*S*_0_ denotes the fraction of the population with immunity acquired through infection, and *f*_*v*_ ≡ *N*_*v*_/*S*_0_ denotes the fraction of the population that has been vaccinated.

We assume that *S*_0_ is constant. We take *N*_*r*_ = *N*_*c*_ to be a good approximation. We assume that we know *S*_0_, *N*_*d*_, and *N*_*v*_. We assume that susceptible and recovered individuals have the same probability of being vaccinated. From our assumption that susceptible and recovered individuals have the same probability of being vaccinated, it follows that *N*_*v,s*_ = (1 − *f*_*r*_)*N*_*v*_ and *N*_*v,r*_ = *f*_*r*_*N*_*v*_. These relations are consistent with *N*_*v*_ ≡ *N*_*v,s*_ + *N*_*v,r*_. The number of individuals with immunity (protection from productive infection) is given by

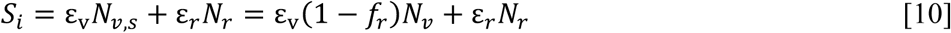

We assume that *Y*_Delta_ gives the value of *β* for SARS-CoV-2 variant Delta relative to *β* for ancestral strains. We assume all other model parameters are the same for Delta. Thus, *Y*_Delta_ℛ_0_ is the basic reproduction number in the face of Delta. We define 𝒫, percent progress toward herd immunity, as

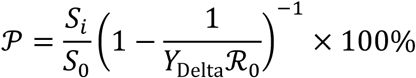

Using the expression given above for *S*_*i*_ (Eq. 10), 1 − 1/(*Y*_Delta_ℛ_0_) as the Delta-adjusted HIT, and *S*_5_ = HIT × *S*_0_, we find Eq. 2 in the main text.

## SI Figure Legends

**Figure S1.** Consistency of results obtained from different codes used to perform Markov chain Monte Carlo (MCMC) sampling. Shown here are 1-dimensional marginal posteriors of parameters for Wyoming (*n* = 0) derived using the Python code of Lin et al. (1) (blue) and PyBioNetFit (4) (red).

**Figure S2.** Markov chain log-likelihood trace plots for each of the 50 US states. Bayesian inference was conditioned on the compartmental model of Lin et al. (1). Bayesian inference was performed as described by Lin et al. (1) except that training data consisted of daily COVID-19 case counts for states (vs. case counts for metropolitan statistical areas). The compartmental model accounts for an initial social distancing period followed by *n* additional periods. We considered *n* = 0, 1 and 2 and selected the best *n* using the model selection procedure described by Lin et al. (1). The number of epochs (or iterations) used for each state was chosen so that convergence was achieved in each case. Inferences are based on daily reports of new cases of COVID-19 from January 21 to June 21, 2020.

**Figure S3.** Parameter trace plots for each of the 50 US states. These parameter trace plots are matched to the likelihood trace plots of Fig S2. It should be noted that the number of parameters varies across the states depending on the selected value of *n*. See the caption of Fig S2 for additional details.

**Figure S4.** Matrix of 1- and 2-dimensional marginalizations of the posterior samples obtained for the adjustable parameters associated with the compartmental model for each of the 50 US states. Inferences are based on daily reports of new cases of COVID-19 from January 21 to June 21, 2020. Plots of marginal posteriors (1-dimensional marginalizations) are shown on the diagonal from top left to bottom right. Other plots are 2-dimensional marginalizations (presented as histograms) indicating the correlations between parameter estimates. Brightness indicates higher probability density. A compact bright area indicates low correlation. An extended, asymmetric bright area indicates high correlation. The pairs plots shown here are matched to the trace plots of Figs S3 and S4. See the caption of Fig S2 for additional details.

**Figure S5.** Posterior predictive checking. The time-dependent predictive posterior distribution for daily number of COVID-19 cases detected is visualized for all states except New Jersey, Wyoming, Florida, and Alaska, which are considered in Fig 1 of the main text. Inferences are based on daily reports of new COVID-19 cases from January 21 to June 21, 2020 (inclusive dates). The compartmental model (1) accounts for an initial social distancing period followed by *n* additional periods. We considered *n* = 0, 1 and 2 and selected the best *n* using the model selection procedure of Lin et al. (1). Crosses indicate observed daily case reports. The shaded region indicates the prediction uncertainty and inferred noise in detection of new cases. The color-coded bands within the shaded region indicate the median and different credible intervals (e.g., dark purple corresponds to the median, the lightest shade of yellow corresponds to the 95% credible interval, and gradations of color between these two extremes correspond to different credible intervals as indicated in the legend). In each panel, the vertical broken line indicates the onset time of the first social-distancing period. For states with *n* = 1, there is an additional (rightmost) broken line, which indicates the onset time of the second social-distancing period. The model was used to make forecasts of new case detection for 14 days after June 21, 2020. The last prediction date was July 5, 2020.

**Figure S6.** Consistency of model-derived λ estimates with empirical growth rates during initial exponential increase in disease incidence in 46 states of the US (i.e., excluding New Jersey, Wyoming, Florida, and Alaska; see Fig 3 in the main text). In each panel, the initial slope of the solid curve corresponds to *λ* (calculated as described in Materials and Methods), the crosses indicate empirical cumulative case counts, and the broken line is the model prediction based on MAP estimates for adjustable parameters. The solid curve is derived from the reduced model (Eqs. 1-8 in the SI). This curve shows cumulative case counts had there not been any interventions to limit disease transmission. As can be seen, the initial slopes of the solid and broken curves are comparable. It should be noted that, in contrast with Fig S5, the y-axis here indicates cumulative (vs. daily) number of cases on a logarithmic (vs. linear) scale.

