## Supplementary Tables S1-S6 and Supplementary Figures S1-S6 for "Bayesian Inference of State-Level COVID-19 Basic Reproduction Numbers across the United States"

Table S1. Estimates of state-specific model parameters. MAP estimates and 95% credible intervals (within parentheses) are given for the inferred parameters.

| State | S_0 | I_0 | m_b | \rho_E | \rho_A | k_L | k_Q | j_Q | f_A | f_H | f_R | c_A | c_I | c_H | t_0 | tau_1-sigma | beta | lambda_0 | p_0 | lambda_1 | p_1 | f_D | r |
|---|---|---|---|---|---|---|---|---|---|---|---|---|---|---|---|---|---|---|---|---|---|---|---|
| Alabama | 4903185 | 1 | 0.1 | 1.1 | 0.9 | 0.94 | 0.0038 | 0.4 | 0.44 | 0.054 | 0.79 | 0.26 | 0.12 | 0.17 | 30.984(22.360-34.336) | N/A | 0.391(0.347-0.436) | 3.499(1.001-9.868) | 0.413(0.369-0.452) | N/A | N/A | 0.967(0.239-1.000) | 7.552(5.428-9.683) |
| Alaska | 731545 | 1 | 0.1 | 1.1 | 0.9 | 0.94 | 0.0038 | 0.4 | 0.44 | 0.054 | 0.79 | 0.26 | 0.12 | 0.17 | 0.389(0.000-8.320) | 24.592(20.416-31.857) | 0.209(0.208-0.226) | 8.181(1.843-9.917) | 0.424(0.388-0.545) | 0.252(0.153-9.449) | 0.042(0.008-0.102) | 0.946(0.538-1.000) | 3.105(1.884-4.686) |
| Arizona | 7278717 | 1 | 0.1 | 1.1 | 0.9 | 0.94 | 0.0038 | 0.4 | 0.44 | 0.054 | 0.79 | 0.26 | 0.12 | 0.17 | 34.393(31.555-40.781) | 61.085(57.441-63.526) | 0.544(0.453-0.595) | 6.487(0.816-6.945) | 0.518(0.469-0.552) | 5.772(0.660-9.581) | 0.422(0.375-0.476) | 0.136(0.039-0.931) | 10.369(7.094-12.786) |
| Arkansas | 3017804 | 1 | 0.1 | 1.1 | 0.9 | 0.94 | 0.0038 | 0.4 | 0.44 | 0.054 | 0.79 | 0.26 | 0.12 | 0.17 | 17.086(0.673-27.769) | N/A | 0.283(0.241-0.350) | 7.148(1.130-9.832) | 0.276(0.210-0.369) | N/A | N/A | 0.977(0.244-0.992) | 2.493(1.777-3.268) |
| California | 39512223 | 1 | 0.1 | 1.1 | 0.9 | 0.94 | 0.0038 | 0.4 | 0.44 | 0.054 | 0.79 | 0.26 | 0.12 | 0.17 | 18.929(12.922-20.468) | N/A | 0.387(0.363-0.410) | 0.091(0.079-0.106) | 0.552(0.529-0.570) | N/A | N/A | 0.955(0.304-1.000) | 13.758(9.699-18.231) |
| Colorado | 5758736 | 1 | 0.1 | 1.1 | 0.9 | 0.94 | 0.0038 | 0.4 | 0.44 | 0.054 | 0.79 | 0.26 | 0.12 | 0.17 | 11.114(4.946-18.122) | N/A | 0.373(0.331-0.417) | 0.164(0.093-0.261) | 0.486(0.435-0.555) | N/A | N/A | 0.047(0.029-0.156) | 12.605(9.102-16.019) |
| Connecticut | 3565287 | 1 | 0.1 | 1.1 | 0.9 | 0.94 | 0.0038 | 0.4 | 0.44 | 0.054 | 0.79 | 0.26 | 0.12 | 0.17 | 24.579(19.192-34.957) | N/A | 0.458(0.366-0.529) | 0.163(0.067-0.256) | 0.537(0.487-0.694) | N/A | N/A | 0.069(0.049-0.910) | 3.867(2.720-4.822) |
| Delaware | 973764 | 1 | 0.1 | 1.1 | 0.9 | 0.94 | 0.0038 | 0.4 | 0.44 | 0.054 | 0.79 | 0.26 | 0.12 | 0.17 | 15.707(1.352-25.519) | N/A | 0.276(0.247-0.315) | 0.063(0.037-0.125) | 0.513(0.377-0.569) | N/A | N/A | 0.539(0.052-0.935) | 3.984(2.789-5.207) |
| Florida | 21477737 | 1 | 0.1 | 1.1 | 0.9 | 0.94 | 0.0038 | 0.4 | 0.44 | 0.054 | 0.79 | 0.26 | 0.12 | 0.17 | 33.729(26.441-34.913) | 63.884(63.829-72.644) | 0.551(0.478-0.590) | 0.169(0.130-0.202) | 0.674(0.637-0.685) | 0.044(0.003-8.701) | 0.548(0.380-0.505) | 0.889(0.158-0.999) | 10.547(7.282-13.066) |
| Georgia | 10617423 | 1 | 0.1 | 1.1 | 0.9 | 0.94 | 0.0038 | 0.4 | 0.44 | 0.054 | 0.79 | 0.26 | 0.12 | 0.17 | 30.334(24.584-33.012) | N/A | 0.448(0.398-0.500) | 0.176(0.134-0.235) | 0.586(0.554-0.613) | N/A | N/A | 0.960(0.303-1.000) | 6.577(4.880-8.300) |
| Hawaii | 1415872 | 1 | 0.1 | 1.1 | 0.9 | 0.94 | 0.0038 | 0.4 | 0.44 | 0.054 | 0.79 | 0.26 | 0.12 | 0.17 | 0.250(0.005-18.577) | 43.824(40.640-50.707) | 0.243(0.227-0.275) | 8.435(1.200-9.819) | 0.462(0.429-0.531) | 0.368(0.208-9.438) | 0.065(0.000-0.133) | 0.269(0.148-0.989) | 7.416(3.538-12.138) |
| Idaho | 1787065 | 1 | 0.1 | 1.1 | 0.9 | 0.94 | 0.0038 | 0.4 | 0.44 | 0.054 | 0.79 | 0.26 | 0.12 | 0.17 | 30.384(22.035-41.502) | 26.118(16.787-44.416) | 0.402(0.321-0.491) | 1.650(0.579-9.408) | 0.556(0.451-0.647) | 0.128(0.002-8.144) | 0.524(0.327-0.561) | 0.206(0.063-0.967) | 3.660(2.494-4.612) |
| Illinois | 12671821 | 1 | 0.1 | 1.1 | 0.9 | 0.94 | 0.0038 | 0.4 | 0.44 | 0.054 | 0.79 | 0.26 | 0.12 | 0.17 | 23.804(20.353-27.454) | N/A | 0.511(0.462-0.574) | 0.271(0.186-0.404) | 0.506(0.476-0.539) | N/A | N/A | 0.047(0.043-0.051) | 16.940(11.817-20.504) |
| Indiana | 6733219 | 1 | 0.1 | 1.1 | 0.9 | 0.94 | 0.0038 | 0.4 | 0.44 | 0.054 | 0.79 | 0.26 | 0.12 | 0.17 | 30.947(28.069-34.415) | N/A | 0.534(0.482-0.599) | 0.273(0.185-0.383) | 0.582(0.553-0.620) | N/A | N/A | 0.061(0.043-0.097) | 23.028(16.373-27.360) |
| Iowa | 3155070 | 1 | 0.1 | 1.1 | 0.9 | 0.94 | 0.0038 | 0.4 | 0.44 | 0.054 | 0.79 | 0.26 | 0.12 | 0.17 | 20.864(4.957-26.906) | N/A | 0.284(0.265-0.311) | 0.051(0.031-0.077) | 0.513(0.400-0.559) | N/A | N/A | 0.914(0.070-0.976) | 6.652(4.727-8.375) |
| Kansas | 2913314 | 1 | 0.1 | 1.1 | 0.9 | 0.94 | 0.0038 | 0.4 | 0.44 | 0.054 | 0.79 | 0.26 | 0.12 | 0.17 | 9.116(0.040-21.513) | N/A | 0.327(0.294-0.393) | 8.963(0.715-9.752) | 0.288(0.236-0.394) | N/A | N/A | 0.020(0.016-0.031) | 2.446(1.811-3.124) |
| Kentucky | 4467673 | 1 | 0.1 | 1.1 | 0.9 | 0.94 | 0.0038 | 0.4 | 0.44 | 0.054 | 0.79 | 0.26 | 0.12 | 0.17 | 14.497(23.657-31.344) | N/A | 0.330(0.336-0.371) | 0.247(1.525-6.814) | 0.426(0.430-0.476) | N/A | N/A | 0.065(0.409-0.809) | 2.832(2.534-2.852) |
| Louisiana | 4648794 | 1 | 0.1 | 1.1 | 0.9 | 0.94 | 0.0038 | 0.4 | 0.44 | 0.054 | 0.79 | 0.26 | 0.12 | 0.17 | 29.321(23.099-34.600) | 23.063(22.072-41.478) | 0.487(0.396-0.561) | 0.068(0.050-0.143) | 0.991(0.724-0.999) | 0.044(0.009-9.149) | 0.621(0.428-0.647) | 0.449(0.166-0.996) | 3.851(2.476-4.454) |
| Maine | 1344212 | 1 | 0.1 | 1.1 | 0.9 | 0.94 | 0.0038 | 0.4 | 0.44 | 0.054 | 0.79 | 0.26 | 0.12 | 0.17 | 0.056(0.001-7.602) | N/A | 0.226(0.225-0.244) | 9.273(2.139-9.983) | 0.232(0.226-0.271) | N/A | N/A | 0.982(0.519-1.000) | 6.302(4.256-8.785) |
| Maryland | 6045680 | 1 | 0.1 | 1.1 | 0.9 | 0.94 | 0.0038 | 0.4 | 0.44 | 0.054 | 0.79 | 0.26 | 0.12 | 0.17 | 20.912(16.191-25.682) | N/A | 0.409(0.369-0.455) | 0.180(0.110-0.286) | 0.464(0.420-0.502) | N/A | N/A | 0.057(0.047-0.071) | 13.997(10.360-18.506) |
| Massachusetts | 6892503 | 1 | 0.1 | 1.1 | 0.9 | 0.94 | 0.0038 | 0.4 | 0.44 | 0.054 | 0.79 | 0.26 | 0.12 | 0.17 | 22.670(11.016-25.519) | N/A | 0.380(0.348-0.420) | 0.055(0.042-0.096) | 0.680(0.481-0.706) | N/A | N/A | 0.946(0.059-0.914) | 4.847(3.404-5.928) |
| Michigan | 9986857 | 1 | 0.1 | 1.1 | 0.9 | 0.94 | 0.0038 | 0.4 | 0.44 | 0.054 | 0.79 | 0.26 | 0.12 | 0.17 | 36.349(29.724-39.519) | N/A | 0.599(0.506-0.754) | 0.219(0.154-0.357) | 0.685(0.641-0.721) | N/A | N/A | 0.952(0.105-0.968) | 2.915(2.098-3.639) |
| Minnesota | 5639632 | 1 | 0.1 | 1.1 | 0.9 | 0.94 | 0.0038 | 0.4 | 0.44 | 0.054 | 0.79 | 0.26 | 0.12 | 0.17 | 0.105(0.000-3.412) | N/A | 0.300(0.298-0.311) | 9.571(2.160-10.000) | 0.212(0.204-0.238) | N/A | N/A | 0.023(0.021-0.026) | 8.521(6.229-10.348) |
| Mississippi | 2976149 | 1 | 0.1 | 1.1 | 0.9 | 0.94 | 0.0038 | 0.4 | 0.44 | 0.054 | 0.79 | 0.26 | 0.12 | 0.17 | 27.049(17.259-40.172) | N/A | 0.475(0.350-0.645) | 9.133(0.804-9.688) | 0.450(0.367-0.583) | N/A | N/A | 0.048(0.033-0.269) | 2.518(1.782-3.260) |
| Missouri | 6137428 | 1 | 0.1 | 1.1 | 0.9 | 0.94 | 0.0038 | 0.4 | 0.44 | 0.054 | 0.79 | 0.26 | 0.12 | 0.17 | 37.235(30.639-41.290) | N/A | 0.475(0.406-0.581) | 0.937(0.298-7.115) | 0.523(0.479-0.566) | N/A | N/A | 0.985(0.228-0.996) | 8.290(5.755-10.350) |
| Montana | 1068778 | 1 | 0.1 | 1.1 | 0.9 | 0.94 | 0.0038 | 0.4 | 0.44 | 0.054 | 0.79 | 0.26 | 0.12 | 0.17 | 0.754(0.002-16.742) | 32.524(27.801-36.190) | 0.220(0.220-0.262) | 9.528(1.637-9.996) | 0.495(0.476-0.612) | 5.366(0.234-9.455) | 0.056(0.033-0.157) | 0.971(0.257-1.000) | 4.025(1.952-5.840) |
| Nebraska | 1934408 | 1 | 0.1 | 1.1 | 0.9 | 0.94 | 0.0038 | 0.4 | 0.44 | 0.054 | 0.79 | 0.26 | 0.12 | 0.17 | 0.036(0.000-14.804) | N/A | 0.246(0.236-0.256) | 0.020(0.011-0.032) | 0.628(0.464-0.938) | N/A | N/A | 0.180(0.088-0.917) | 6.131(4.385-7.965) |
| Nevada | 3080156 | 1 | 0.1 | 1.1 | 0.9 | 0.94 | 0.0038 | 0.4 | 0.44 | 0.054 | 0.79 | 0.26 | 0.12 | 0.17 | 26.225(17.540-32.919) | 65.190(58.061-72.778) | 0.356(0.331-0.437) | 0.508(0.237-9.075) | 0.452(0.408-0.493) | 6.065(0.219-9.464) | 0.320(0.267-0.418) | 0.802(0.093-0.981) | 8.104(5.467-10.151) |
| New Hampshire | 1359711 | 1 | 0.1 | 1.1 | 0.9 | 0.94 | 0.0038 | 0.4 | 0.44 | 0.054 | 0.79 | 0.26 | 0.12 | 0.17 | 0.002(0.000-3.737) | N/A | 0.287(0.284-0.298) | 9.176(1.949-9.968) | 0.267(0.255-0.294) | N/A | N/A | 0.026(0.021-0.033) | 10.142(6.935-12.710) |
| New Jersey | 8882190 | 1 | 0.1 | 1.1 | 0.9 | 0.94 | 0.0038 | 0.4 | 0.44 | 0.054 | 0.79 | 0.26 | 0.12 | 0.17 | 32.076(30.841-36.593) | N/A | 0.652(0.592-0.706) | 0.143(0.113-0.165) | 0.710(0.686-0.768) | N/A | N/A | 0.173(0.102-0.917) | 16.197(11.032-20.226) |
| New Mexico | 2096829 | 1 | 0.1 | 1.1 | 0.9 | 0.94 | 0.0038 | 0.4 | 0.44 | 0.054 | 0.79 | 0.26 | 0.12 | 0.17 | 2.728(0.179-20.746) | N/A | 0.280(0.248-0.302) | 0.173(0.057-0.273) | 0.342(0.325-0.451) | N/A | N/A | 0.052(0.032-0.910) | 10.695(7.485-13.903) |
| New York | 19453561 | 1 | 0.1 | 1.1 | 0.9 | 0.94 | 0.0038 | 0.4 | 0.44 | 0.054 | 0.79 | 0.26 | 0.12 | 0.17 | 26.493(22.339-28.209) | 6.945(5.447-8.628) | 0.537(0.503-0.580) | 3.341(0.002-9.310) | 0.000(0.000-0.047) | 0.173(0.141-0.236) | 0.693(0.649-0.718) | 0.842(0.266-1.000) | 15.497(10.271-17.131) |
| North Carolina | 10488084 | 1 | 0.1 | 1.1 | 0.9 | 0.94 | 0.0038 | 0.4 | 0.44 | 0.054 | 0.79 | 0.26 | 0.12 | 0.17 | 31.206(25.599-36.723) | N/A | 0.408(0.381-0.512) | 0.430(0.239-4.668) | 0.460(0.417-0.498) | N/A | N/A | 0.916(0.178-0.996) | 9.592(6.512-11.786) |
| North Dakota | 762062 | 1 | 0.1 | 1.1 | 0.9 | 0.94 | 0.0038 | 0.4 | 0.44 | 0.054 | 0.79 | 0.26 | 0.12 | 0.17 | 0.121(0.000-6.987) | N/A | 0.211(0.211-0.225) | 0.044(0.025-0.089) | 0.389(0.316-0.484) | N/A | N/A | 0.987(0.504-1.000) | 6.428(4.278-8.309) |
| Ohio | 11689100 | 1 | 0.1 | 1.1 | 0.9 | 0.94 | 0.0038 | 0.4 | 0.44 | 0.054 | 0.79 | 0.26 | 0.12 | 0.17 | 24.363(15.560-29.743) | N/A | 0.374(0.340-0.412) | 0.114(0.091-0.147) | 0.558(0.520-0.586) | N/A | N/A | 0.521(0.110-0.990) | 13.676(9.929-17.656) |
| Oklahoma | 3956971 | 1 | 0.1 | 1.1 | 0.9 | 0.94 | 0.0038 | 0.4 | 0.44 | 0.054 | 0.79 | 0.26 | 0.12 | 0.17 | 31.752(22.607-36.912) | 69.291(66.405-72.574) | 0.368(0.329-0.437) | 0.655(0.398-5.037) | 0.455(0.420-0.680) | 0.675(0.197-9.220) | 0.230(0.016-0.302) | 0.854(0.148-0.992) | 7.919(5.391-10.034) |
| Oregon | 4217737 | 1 | 0.1 | 1.1 | 0.9 | 0.94 | 0.0038 | 0.4 | 0.44 | 0.054 | 0.79 | 0.26 | 0.12 | 0.17 | 16.689(8.407-26.374) | 62.379(58.500-66.910) | 0.312(0.283-0.342) | 0.680(0.342-9.302) | 0.400(0.343-0.416) | 1.426(0.348-9.533) | 0.239(0.179-0.295) | 0.302(0.114-0.997) | 10.100(6.422-13.594) |
| Pennsylvania | 12801989 | 1 | 0.1 | 1.1 | 0.9 | 0.94 | 0.0038 | 0.4 | 0.44 | 0.054 | 0.79 | 0.26 | 0.12 | 0.17 | 33.882(28.338-35.164) | N/A | 0.499(0.470-0.529) | 0.119(0.108-0.135) | 0.682(0.657-0.694) | N/A | N/A | 0.897(0.148-0.980) | 24.128(17.365-30.657) |
| Rhode Island | 1059361 | 1 | 0.1 | 1.1 | 0.9 | 0.94 | 0.0038 | 0.4 | 0.44 | 0.054 | 0.79 | 0.26 | 0.12 | 0.17 | 22.434(5.055-30.168) | N/A | 0.307(0.268-0.356) | 0.059(0.024-0.187) | 0.595(0.300-0.672) | N/A | N/A | 0.616(0.050-0.925) | 1.722(1.190-2.274) |
| South Carolina | 5148714 | 1 | 0.1 | 1.1 | 0.9 | 0.94 | 0.0038 | 0.4 | 0.44 | 0.054 | 0.79 | 0.26 | 0.12 | 0.17 | 27.886(20.463-34.331) | 53.614(50.857-61.892) | 0.377(0.345-0.447) | 0.301(0.182-1.122) | 0.497(0.450-0.531) | 0.327(0.000-9.392) | 0.370(0.256-0.423) | 0.578(0.080-0.985) | 10.621(7.582-13.534) |
| South Dakota | 884659 | 1 | 0.1 | 1.1 | 0.9 | 0.94 | 0.0038 | 0.4 | 0.44 | 0.054 | 0.79 | 0.26 | 0.12 | 0.17 | 33.934(21.169-39.697) | N/A | 0.311(0.284-0.363) | 0.244(0.124-1.721) | 0.444(0.375-0.473) | N/A | N/A | 0.943(0.096-0.963) | 4.223(2.790-5.426) |
| Tennessee | 6829174 | 1 | 0.1 | 1.1 | 0.9 | 0.94 | 0.0038 | 0.4 | 0.44 | 0.054 | 0.79 | 0.26 | 0.12 | 0.17 | 31.625(23.354-37.080) | N/A | 0.435(0.375-0.575) | 0.334(0.151-5.156) | 0.519(0.468-0.560) | N/A | N/A | 0.996(0.118-0.969) | 7.464(5.170-9.169) |
| Texas | 28995881 | 1 | 0.1 | 1.1 | 0.9 | 0.94 | 0.0038 | 0.4 | 0.44 | 0.054 | 0.79 | 0.26 | 0.12 | 0.17 | 24.420(21.333-29.131) | 61.271(60.063-80.583) | 0.415(0.386-0.445) | 0.123(0.101-0.167) | 0.578(0.542-0.596) | 0.083(0.000-9.067) | 0.486(0.262-0.504) | 0.337(0.131-0.963) | 12.400(7.972-15.013) |
| Utah | 3205958 | 1 | 0.1 | 1.1 | 0.9 | 0.94 | 0.0038 | 0.4 | 0.44 | 0.054 | 0.79 | 0.26 | 0.12 | 0.17 | 15.334(3.259-26.542) | 56.229(49.991-62.279) | 0.323(0.280-0.368) | 0.421(0.161-2.055) | 0.396(0.329-0.428) | 0.008(0.000-8.346) | 0.727(0.167-0.670) | 0.210(0.018-0.897) | 13.792(9.650-16.554) |
| Vermont | 623989 | 1 | 0.1 | 1.1 | 0.9 | 0.94 | 0.0038 | 0.4 | 0.44 | 0.054 | 0.79 | 0.26 | 0.12 | 0.17 | 5.435(2.059-32.206) | 43.446(35.682-49.688) | 0.317(0.245-0.357) | 9.312(0.546-9.589) | 0.437(0.408-0.564) | 4.301(0.224-9.482) | 0.172(0.170-0.363) | 0.011(0.012-0.938) | 2.375(1.397-2.952) |
| Virginia | 8535519 | 1 | 0.1 | 1.1 | 0.9 | 0.94 | 0.0038 | 0.4 | 0.44 | 0.054 | 0.79 | 0.26 | 0.12 | 0.17 | 11.702(4.752-20.999) | N/A | 0.353(0.313-0.411) | 0.178(0.060-0.480) | 0.393(0.335-0.479) | N/A | N/A | 0.036(0.027-0.066) | 5.054(3.505-6.291) |
| Washington | 7614893 | 1 | 0.1 | 1.1 | 0.9 | 0.94 | 0.0038 | 0.4 | 0.44 | 0.054 | 0.79 | 0.26 | 0.12 | 0.17 | 15.968(5.728-20.448) | N/A | 0.368(0.315-0.428) | 0.157(0.110-0.242) | 0.527(0.477-0.569) | N/A | N/A | 0.982(0.264-1.000) | 4.574(3.333-5.664) |
| West Virginia | 1792147 | 1 | 0.1 | 1.1 | 0.9 | 0.94 | 0.0038 | 0.4 | 0.44 | 0.054 | 0.79 | 0.26 | 0.12 | 0.17 | 6.096(0.001-19.932) | N/A | 0.230(0.221-0.269) | 8.721(1.382-9.895) | 0.251(0.233-0.324) | N/A | N/A | 0.932(0.231-1.000) | 4.054(2.716-5.348) |
| Wisconsin | 5822534 | 1 | 0.1 | 1.1 | 0.9 | 0.94 | 0.0038 | 0.4 | 0.44 | 0.054 | 0.79 | 0.26 | 0.12 | 0.17 | 19.169(8.024-29.143) | N/A | 0.416(0.328-0.486) | 8.494(0.409-9.195) | 0.411(0.337-0.487) | N/A | N/A | 0.046(0.031-0.164) | 5.939(4.220-7.690) |
| Wyoming | 578759 | 1 | 0.1 | 1.1 | 0.9 | 0.94 | 0.0038 | 0.4 | 0.44 | 0.054 | 0.79 | 0.26 | 0.12 | 0.17 | 0.192(0.000-10.292) | N/A | 0.209(0.208-0.227) | 9.361(1.886-9.972) | 0.213(0.203-0.259) | N/A | N/A | 0.984(0.517-1.000) | 2.564(1.670-3.464) |

**Table S2.** State-specific MAP estimates and 95% credible intervals (within parentheses) of the initial epidemic growth rate $\lambda$ (/d), basic reproduction number $\mathcal{R}_0$, and herd immunity threshold (HIT) for SARS-CoV-2 ancestral strains. Estimates are based on state-specific daily reports of new COVID-19 cases from January 21, 2020 to June 21, 2020 (inclusive dates).

| State | $\lambda$ (/d) | $\mathcal{R}_0$ | HIT |
|---|---|---|---|
| Alabama | 0.278(0.245-0.310) | 4.250(3.770-4.735) | 0.765(0.735-0.789) |
| Alaska | 0.127(0.126-0.142) | 2.275(2.257-2.452) | 0.560(0.557-0.592) |
| Arizona | 0.382(0.322-0.414) | 5.910(4.921-6.463) | 0.831(0.797-0.845) |
| Arkansas | 0.193(0.157-0.247) | 3.078(2.622-3.800) | 0.675(0.619-0.737) |
| California | 0.275(0.257-0.292) | 4.204(3.948-4.456) | 0.762(0.747-0.776) |
| Colorado | 0.264(0.232-0.297) | 4.049(3.601-4.536) | 0.753(0.722-0.780) |
| Connecticut | 0.325(0.260-0.373) | 4.977(3.982-5.752) | 0.799(0.749-0.826) |
| Delaware | 0.187(0.162-0.220) | 2.996(2.687-3.425) | 0.666(0.628-0.708) |
| Florida | 0.387(0.339-0.411) | 5.994(5.195-6.413) | 0.833(0.808-0.844) |
| Georgia | 0.319(0.283-0.354) | 4.872(4.324-5.438) | 0.795(0.769-0.816) |
| Hawaii | 0.158(0.144-0.186) | 2.638(2.471-2.988) | 0.621(0.595-0.665) |
| Idaho | 0.286(0.224-0.348) | 4.374(3.488-5.338) | 0.771(0.713-0.813) |
| Illinois | 0.361(0.328-0.401) | 5.557(5.017-6.242) | 0.820(0.801-0.840) |
| Indiana | 0.376(0.342-0.416) | 5.808(5.241-6.512) | 0.828(0.809-0.846) |
| Iowa | 0.194(0.178-0.216) | 3.088(2.884-3.376) | 0.676(0.653-0.704) |
| Kansas | 0.229(0.203-0.279) | 3.557(3.201-4.269) | 0.719(0.688-0.766) |
| Kentucky | 0.232(0.236-0.263) | 3.588(3.648-4.031) | 0.721(0.726-0.752) |
| Louisiana | 0.345(0.282-0.393) | 5.294(4.303-6.093) | 0.811(0.768-0.836) |
| Maine | 0.143(0.142-0.159) | 2.454(2.450-2.653) | 0.593(0.592-0.623) |
| Maryland | 0.291(0.261-0.324) | 4.446(4.006-4.951) | 0.775(0.750-0.798) |
| Massachusetts | 0.270(0.246-0.299) | 4.127(3.783-4.567) | 0.758(0.736-0.781) |
| Michigan | 0.416(0.358-0.505) | 6.510(5.498-8.194) | 0.846(0.818-0.878) |
| Minnesota | 0.207(0.205-0.217) | 3.261(3.237-3.386) | 0.693(0.691-0.705) |
| Mississippi | 0.337(0.247-0.444) | 5.167(3.804-7.012) | 0.806(0.737-0.857) |
| Missouri | 0.337(0.289-0.405) | 5.169(4.411-6.316) | 0.807(0.773-0.842) |
| Montana | 0.137(0.137-0.175) | 2.390(2.392-2.852) | 0.582(0.582-0.649) |
| Nebraska | 0.161(0.152-0.170) | 2.671(2.565-2.788) | 0.626(0.610-0.641) |
| Nevada | 0.252(0.232-0.311) | 3.870(3.595-4.750) | 0.742(0.722-0.789) |
| New Hampshire | 0.196(0.194-0.206) | 3.117(3.092-3.238) | 0.679(0.677-0.691) |
| New Jersey | 0.448(0.412-0.479) | 7.093(6.440-7.673) | 0.859(0.845-0.870) |
| New Mexico | 0.190(0.162-0.209) | 3.042(2.692-3.280) | 0.671(0.629-0.695) |
| New York | 0.378(0.356-0.405) | 5.842(5.469-6.310) | 0.829(0.817-0.842) |
| North Carolina | 0.290(0.270-0.362) | 4.433(4.141-5.567) | 0.774(0.758-0.820) |
| North Dakota | 0.129(0.128-0.142) | 2.296(2.289-2.443) | 0.564(0.563-0.591) |
| Ohio | 0.265(0.239-0.293) | 4.063(3.698-4.475) | 0.754(0.730-0.777) |
| Oklahoma | 0.261(0.231-0.311) | 3.998(3.579-4.748) | 0.750(0.721-0.789) |

| | | | |
|---|---|---|---|
| Oregon | 0.217(0.193-0.241) | 3.389(3.078-3.721) | 0.705(0.675-0.731) |
| Pennsylvania | 0.353(0.334-0.373) | 5.423(5.108-5.753) | 0.816(0.804-0.826) |
| Rhode Island | 0.213(0.180-0.251) | 3.338(2.911-3.865) | 0.700(0.656-0.741) |
| South Carolina | 0.268(0.244-0.318) | 4.102(3.755-4.862) | 0.756(0.734-0.794) |
| South Dakota | 0.216(0.194-0.257) | 3.377(3.091-3.947) | 0.704(0.677-0.747) |
| Tennessee | 0.310(0.266-0.401) | 4.730(4.079-6.247) | 0.789(0.755-0.840) |
| Texas | 0.295(0.274-0.317) | 4.509(4.197-4.841) | 0.778(0.762-0.793) |
| Utah | 0.226(0.191-0.261) | 3.510(3.046-4.004) | 0.715(0.672-0.750) |
| Vermont | 0.221(0.160-0.253) | 3.446(2.665-3.884) | 0.710(0.625-0.743) |
| Virginia | 0.250(0.218-0.293) | 3.839(3.404-4.469) | 0.739(0.706-0.776) |
| Washington | 0.261(0.220-0.304) | 4.004(3.428-4.648) | 0.750(0.708-0.785) |
| West Virginia | 0.146(0.138-0.181) | 2.496(2.406-2.927) | 0.599(0.584-0.658) |
| Wisconsin | 0.296(0.230-0.344) | 4.527(3.570-5.278) | 0.779(0.720-0.811) |
| Wyoming | 0.127(0.126-0.144) | 2.272(2.260-2.467) | 0.560(0.558-0.595) |

**Table S3.** State-specific MAP estimates of $\beta$ obtained using different datasets. Each 4-month dataset consists of state-specific daily case counts from January 21, 2020 to May 21, 2020. Each 5-month dataset consists of state-specific daily case counts from January 21, 2020 to June 21, 2020. Each 6-month dataset consists of state-specific daily case counts from January 21, 2020 to July 21, 2020.

| | $\beta$ | | |
|---|---|---|---|
| State | 4-month dataset | 5-month dataset | 6-month dataset |
| Alabama | 0.344425688 | 0.391000116 | 0.381114062 |
| Alaska | 0.207653693 | 0.209267205 | 0.210030786 |
| Arizona | 0.529445483 | 0.543664926 | 0.532514897 |
| Arkansas | 0.32298932 | 0.28311954 | 0.279105356 |
| California | 0.329080014 | 0.386768663 | 0.366219921 |
| Colorado | 0.381094706 | 0.372505137 | 0.377135113 |
| Connecticut | 0.449485806 | 0.457835438 | 0.442979285 |
| Delaware | 0.280036868 | 0.27564221 | 0.284739604 |
| Florida | 0.526921359 | 0.551352675 | 0.514304171 |
| Georgia | 0.430531087 | 0.448214949 | 0.438936107 |
| Hawaii | 0.23746958 | 0.242697953 | 0.247395063 |
| Idaho | 0.380490164 | 0.402373556 | 0.36290268 |
| Illinois | 0.498497271 | 0.511225388 | 0.50775665 |
| Indiana | 0.541857154 | 0.534310158 | 0.534529103 |
| Iowa | 0.311378197 | 0.284051758 | 0.271908187 |
| Kansas | 0.300249291 | 0.327199897 | 0.244883859 |
| Kentucky | 0.346224636 | 0.330082083 | 0.331292779 |
| Louisiana | 0.488353117 | 0.487004484 | 0.504124433 |
| Maine | 0.225236222 | 0.225787998 | 0.230958926 |
| Maryland | 0.398259593 | 0.409012941 | 0.410424365 |
| Massachusetts | 0.386526299 | 0.379636348 | 0.387649785 |
| Michigan | 0.602060282 | 0.59887731 | 0.608052362 |
| Minnesota | 0.263631203 | 0.299947476 | 0.255634342 |
| Mississippi | 0.446758703 | 0.475310237 | 0.505863769 |
| Missouri | 0.444622556 | 0.475476797 | 0.439740679 |
| Montana | 0.220783058 | 0.219834578 | 0.219902109 |
| Nebraska | 0.244391787 | 0.2457406 | 0.244258417 |
| Nevada | 0.359673276 | 0.35601223 | 0.361269375 |
| New Hampshire | 0.273648163 | 0.286714521 | 0.284787414 |
| New Jersey | 0.656582201 | 0.652491027 | 0.676182424 |
| New Mexico | 0.289726267 | 0.279879781 | 0.275387246 |
| New York | 0.535623994 | 0.537427785 | 0.675754416 |
| North Carolina | 0.409013369 | 0.407806714 | 0.416748691 |
| North Dakota | 0.213348899 | 0.211192131 | 0.211282563 |

| | | | |
|---|---|---|---|
| Ohio | 0.407742398 | 0.373799884 | 0.396832606 |
| Oklahoma | 0.367641453 | 0.367742577 | 0.368838695 |
| Oregon | 0.30011376 | 0.311724057 | 0.305668351 |
| Pennsylvania | 0.497075654 | 0.49886789 | 0.498607161 |
| Rhode Island | 0.319663632 | 0.307048589 | 0.317887767 |
| South Carolina | 0.382005702 | 0.377319773 | 0.378889298 |
| South Dakota | 0.315190829 | 0.310612853 | 0.311311794 |
| Tennessee | 0.451102391 | 0.435139557 | 0.422615302 |
| Texas | 0.418736232 | 0.414805718 | 0.429223299 |
| Utah | 0.31236819 | 0.32286498 | 0.307458597 |
| Vermont | 0.265564455 | 0.317029863 | 0.303010374 |
| Virginia | 0.347873875 | 0.353117278 | 0.335591603 |
| Washington | 0.351809052 | 0.368329925 | 0.353636894 |
| West Virginia | 0.238034407 | 0.229590329 | 0.235258658 |
| Wisconsin | 0.386164126 | 0.416429043 | 0.373319249 |
| Wyoming | 0.210764739 | 0.209001836 | 0.219153861 |

**Table S4.** Sensitivity of MAP estimate of $\beta$ to training dataset. Sensitivity was assessed by computing the relative error between the $\beta$ estimates obtained from the 5-month dataset and the average $\beta$ estimate over all datasets considered (i.e., 4-, 5-, and 6-month periods). Each 4-month dataset consists of state-specific daily case counts from January 21, 2020 to May 21, 2020. Each 5-month dataset consists of state-specific daily case counts from January 21, 2020 to June 21, 2020. Each 6-month dataset consists of state-specific daily case counts from January 21, 2020 to July 21, 2020.

| State | Relative error (%) |
|---|---|
| Alabama | 5.056736841 |
| Alaska | 0.135565694 |
| Arizona | 1.580036956 |
| Arkansas | 4.050499367 |
| California | 7.230353936 |
| Colorado | 1.169110891 |
| Connecticut | 1.71856443 |
| Delaware | 1.605396619 |
| Florida | 3.860395675 |
| Georgia | 2.046222168 |
| Hawaii | 0.07301952 |
| Idaho | 5.354867104 |
| Illinois | 1.067352626 |
| Indiana | 0.482148062 |
| Iowa | 1.750513215 |
| Kansas | 12.52579428 |
| Kentucky | 1.722236898 |
| Louisiana | 1.248313993 |
| Maine | 0.677311718 |
| Maryland | 0.767179733 |
| Massachusetts | 1.291664711 |
| Michigan | 0.683144998 |
| Minnesota | 9.842300289 |
| Mississippi | 0.140202483 |
| Missouri | 4.896925896 |
| Montana | 0.153819893 |
| Nebraska | 0.385488999 |
| Nevada | 0.828093273 |
| New Hampshire | 1.774059408 |
| New Jersey | 1.399445444 |
| New Mexico | 0.633608589 |
| New York | 7.806630712 |

| | |
|---|---|
| North Carolina | 0.822704993 |
| North Dakota | 0.353431338 |
| Ohio | 4.835068796 |
| Oklahoma | 0.090108154 |
| Oregon | 1.9254369 |
| Pennsylvania | 0.137363347 |
| Rhode Island | 2.482979345 |
| South Carolina | 0.549584631 |
| South Dakota | 0.563102005 |
| Tennessee | 0.262716142 |
| Texas | 1.453009268 |
| Utah | 2.747788206 |
| Vermont | 7.394371088 |
| Virginia | 2.196551742 |
| Washington | 2.906929126 |
| West Virginia | 2.007787943 |
| Wisconsin | 6.239810825 |
| Wyoming | 1.86485316 |

**Table S5.** Percent progress $\mathcal{P}$ toward herd immunity for each of the 50 states. This table provides the inputs to Eq. 2 that were used to generate the results plotted in Fig. 6. In Case 1, $f_r$ is given in column 6. In Case 2, $f_r$ is given in column 8. Note that $f_A = 0.44$. In both cases, $\varepsilon_v = 0.66$ and $\varepsilon_r = 1.0$.

| State | $S_0$ | $Y_{\text{Delta}}$ | $1 - \frac{1}{Y_{\text{Delta}}\mathcal{R}_0}$ | $N_d/S_0$ | $\left(\frac{N_d}{S_0}\right) \times 5.8$ | $f_s$ | $f_s/(1-f_A)$ | $f_v$ | $\mathcal{P}$ (Case 1) | $\mathcal{P}$ (Case 2) |
|---|---|---|---|---|---|---|---|---|---|---|
| Alabama | 4903185 | 2.46 | 0.9044 | 0.1575 | 0.9136 | 0.292 | 0.5214 | 0.4127 | 103.6217 | 72.0726 |
| Alaska | 731545 | 2.46 | 0.8213 | 0.1390 | 0.8063 | 0.093 | 0.1661 | 0.4909 | 105.8143 | 53.1165 |
| Arizona | 7278717 | 2.46 | 0.9312 | 0.1468 | 0.8517 | 0.297 | 0.5304 | 0.5029 | 96.7460 | 73.6934 |
| Arkansas | 3017804 | 2.46 | 0.8679 | 0.1609 | 0.9330 | 0.252 | 0.4500 | 0.4460 | 109.7714 | 70.5032 |
| California | 39512223 | 2.46 | 0.9033 | 0.1180 | 0.6844 | 0.211 | 0.3768 | 0.5788 | 89.1154 | 68.0675 |
| Colorado | 5758736 | 2.46 | 0.8996 | 0.1141 | 0.6616 | 0.096 | 0.1714 | 0.5857 | 88.0859 | 54.6621 |
| Connecticut | 3565287 | 2.46 | 0.9183 | 0.1082 | 0.6276 | 0.065 | 0.1161 | 0.6774 | 86.4714 | 55.6717 |
| Delaware | 973764 | 2.46 | 0.8643 | 0.1322 | 0.7665 | 0.215 | 0.3839 | 0.5679 | 98.8090 | 71.1368 |
| Florida | 21477737 | 2.46 | 0.9322 | 0.1631 | 0.9462 | 0.254 | 0.4536 | 0.5584 | 103.6342 | 70.2618 |
| Georgia | 10617423 | 2.46 | 0.9166 | 0.1416 | 0.8212 | 0.124 | 0.2214 | 0.4407 | 95.2713 | 48.8630 |
| Hawaii | 1415872 | 2.46 | 0.8459 | 0.0520 | 0.3014 | 0.016 | 0.0286 | 0.5701 | 66.7006 | 46.5866 |
| Idaho | 1787065 | 2.46 | 0.9071 | 0.1363 | 0.7908 | 0.109 | 0.1946 | 0.4078 | 93.3873 | 45.3530 |
| Illinois | 12671821 | 2.46 | 0.9269 | 0.1265 | 0.7339 | 0.319 | 0.5696 | 0.5270 | 89.1686 | 77.6101 |
| Indiana | 6733219 | 2.46 | 0.9300 | 0.1393 | 0.8080 | 0.195 | 0.3482 | 0.4768 | 93.3745 | 59.4952 |
| Iowa | 3155070 | 2.46 | 0.8684 | 0.1376 | 0.7983 | 0.127 | 0.2268 | 0.5318 | 100.0827 | 57.3692 |
| Kansas | 2913314 | 2.46 | 0.8857 | 0.1375 | 0.7972 | 0.085 | 0.1518 | 0.5020 | 97.5952 | 48.8669 |
| Kentucky | 4467673 | 2.46 | 0.8867 | 0.1484 | 0.8607 | 0.255 | 0.4554 | 0.5083 | 102.3341 | 71.9576 |
| Louisiana | 4648794 | 2.46 | 0.9232 | 0.1568 | 0.9093 | 0.103 | 0.1839 | 0.4436 | 101.3706 | 45.8047 |
| Maine | 1344212 | 2.46 | 0.8344 | 0.0624 | 0.3621 | 0.05 | 0.0893 | 0.6748 | 77.4412 | 59.3086 |
| Maryland | 6045680 | 2.46 | 0.9086 | 0.0861 | 0.4993 | 0.293 | 0.5232 | 0.6328 | 77.9720 | 79.5039 |
| Massachusetts | 6892503 | 2.46 | 0.9015 | 0.1154 | 0.6694 | 0.086 | 0.1536 | 0.6725 | 90.5334 | 58.7074 |
| Michigan | 9986857 | 2.46 | 0.9376 | 0.1117 | 0.6480 | 0.281 | 0.5018 | 0.5172 | 81.9332 | 71.6587 |
| Minnesota | 5639632 | 2.46 | 0.8753 | 0.1213 | 0.7036 | 0.112 | 0.2000 | 0.5752 | 93.2384 | 57.5475 |
| Mississippi | 2976149 | 2.46 | 0.9213 | 0.1600 | 0.9278 | 0.338 | 0.6036 | 0.4213 | 102.8846 | 77.4756 |
| Missouri | 6137428 | 2.46 | 0.9214 | 0.1335 | 0.7744 | 0.237 | 0.4232 | 0.4700 | 91.6428 | 65.3515 |
| Montana | 1068778 | 2.46 | 0.8299 | 0.1324 | 0.7678 | 0.085 | 0.1518 | 0.4774 | 101.3321 | 50.4968 |

| | | | | | | | | | | |
|---|---|---|---|---|---|---|---|---|---|---|
| Nebraska | 1934408 | 2.46 | 0.8478 | 0.1340 | 0.7772 | 0.109 | 0.1946 | 0.5367 | 100.9768 | 56.6056 |
| Nevada | 3080156 | 2.46 | 0.8950 | 0.1339 | 0.7764 | 0.267 | 0.4768 | 0.4982 | 94.9681 | 72.4986 |
| New Hampshire | 1359711 | 2.46 | 0.8696 | 0.0849 | 0.4923 | 0.065 | 0.1161 | 0.6099 | 80.1139 | 54.2680 |
| New Jersey | 8882190 | 2.46 | 0.9427 | 0.1278 | 0.7410 | 0.303 | 0.5411 | 0.6347 | 90.1171 | 77.7900 |
| New Mexico | 2096829 | 2.46 | 0.8664 | 0.1174 | 0.6811 | 0.12 | 0.2143 | 0.6215 | 93.7109 | 61.9322 |
| New York | 19453561 | 2.46 | 0.9304 | 0.1221 | 0.7080 | 0.123 | 0.2196 | 0.6256 | 89.0551 | 58.2348 |
| North Carolina | 10488084 | 2.46 | 0.9083 | 0.1286 | 0.7456 | 0.226 | 0.4036 | 0.4868 | 91.0880 | 65.5284 |
| North Dakota | 762062 | 2.46 | 0.8229 | 0.1654 | 0.9594 | - | - | 0.4333 | 117.9901 | 34.7532 |
| Ohio | 11689100 | 2.46 | 0.9000 | 0.1157 | 0.6709 | 0.333 | 0.5946 | 0.4961 | 86.5202 | 80.8211 |
| Oklahoma | 3956971 | 2.46 | 0.8983 | 0.1511 | 0.8766 | 0.111 | 0.1982 | 0.4647 | 101.7986 | 49.4386 |
| Oregon | 4217737 | 2.46 | 0.8800 | 0.0742 | 0.4306 | 0.071 | 0.1268 | 0.5987 | 74.4978 | 53.6129 |
| Pennsylvania | 12801989 | 2.46 | 0.9250 | 0.1080 | 0.6265 | 0.226 | 0.4036 | 0.5691 | 82.8942 | 67.8431 |
| Rhode Island | 1059361 | 2.46 | 0.8782 | 0.1599 | 0.9272 | 0.103 | 0.1839 | 0.6696 | 109.2411 | 62.0112 |
| South Carolina | 5148714 | 2.46 | 0.9009 | 0.1613 | 0.9353 | 0.262 | 0.4679 | 0.4595 | 105.9958 | 69.8451 |
| South Dakota | 884659 | 2.46 | 0.8796 | 0.1587 | 0.9205 | 0.341 | 0.6089 | 0.5076 | 107.6770 | 84.1227 |
| Tennessee | 6829174 | 2.46 | 0.9141 | 0.1703 | 0.9880 | 0.266 | 0.4750 | 0.4411 | 108.4681 | 68.6860 |
| Texas | 28995881 | 2.46 | 0.9098 | 0.1358 | 0.7879 | 0.319 | 0.5696 | 0.5019 | 94.3166 | 78.2778 |
| Utah | 3205958 | 2.46 | 0.8842 | 0.1542 | 0.8946 | 0.293 | 0.5232 | 0.4954 | 105.0729 | 76.8066 |
| Vermont | 623989 | 2.46 | 0.8820 | 0.0509 | 0.2952 | 0.018 | 0.0321 | 0.6893 | 69.8241 | 53.5661 |
| Virginia | 8535519 | 2.46 | 0.8941 | 0.0980 | 0.5682 | 0.173 | 0.3089 | 0.5953 | 82.5214 | 64.9181 |
| Washington | 7614893 | 2.46 | 0.8985 | 0.0828 | 0.4802 | 0.07 | 0.1250 | 0.6223 | 77.2079 | 53.9088 |
| West Virginia | 1792147 | 2.46 | 0.8371 | 0.1252 | 0.7263 | 0.09 | 0.1607 | 0.4015 | 95.4285 | 45.7627 |
| Wisconsin | 5822534 | 2.46 | 0.9102 | 0.1339 | 0.7769 | 0.285 | 0.5089 | 0.5563 | 94.3549 | 75.7240 |
| Wyoming | 578759 | 2.46 | 0.8211 | 0.1477 | 0.8568 | 0.308 | 0.5500 | 0.4063 | 109.0295 | 81.6820 |

**Table S6.** Comparison of estimates of the basic reproduction number $\mathcal{R}_0$ for ancestral strains of SARS-CoV-2 from four different studies for each of the 30 states considered in all of the studies.

| | $\mathcal{R}_0$ estimate | | | |
|---|---|---|---|---|
| State | This study | Ref. (16) | Ref. (17) | Ref. (18) |
| Kansas | 4.25 | 1.418 | 2.363 | 4.212 |
| Michigan | 5.91 | 1.299 | 2.234 | 4.166 |
| Montana | 4.204 | 2.447 | 2.196 | 3.294 |
| Maine | 4.049 | 2.077 | 2.286 | 3.463 |
| Colorado | 4.977 | 3.18 | 2.675 | 5.143 |
| Tennessee | 2.996 | 1.387 | 1.886 | 3.109 |
| Louisiana | 5.994 | 1.803 | 2.285 | 5.284 |
| Kentucky | 4.872 | 2.914 | 2.311 | 5.629 |
| Iowa | 5.294 | 4.558 | 2.376 | 3.703 |
| Connecticut | 4.446 | 3.145 | 2.584 | 4.133 |
| Arizona | 4.127 | 3.011 | 2.805 | 3.309 |
| Ohio | 6.51 | 5.083 | 2.002 | 7.533 |
| Hawaii | 3.261 | 1.599 | 2.467 | 3.013 |
| Massachusetts | 5.167 | 1.314 | 2.247 | 7.707 |
| Mississippi | 5.169 | 1.258 | 2.234 | 5.277 |
| Nebraska | 3.87 | 1.217 | 2.131 | 3.759 |
| Alaska | 7.093 | 1.357 | 2.961 | 6.814 |
| South Dakota | 3.042 | 4.558 | 1.899 | 2.9 |
| Alabama | 5.842 | 6.463 | 3.275 | 4.246 |
| Florida | 4.433 | 2.774 | 2.558 | 4.895 |
| Delaware | 4.063 | 1.473 | 2.596 | 4.938 |
| Nevada | 3.998 | 2.368 | 2.118 | 4.903 |
| California | 5.423 | 1.028 | 2.714 | 4.839 |
| Maryland | 3.338 | 2.82 | 2.247 | 2.817 |
| Oregon | 4.102 | 1.321 | 1.963 | 4.647 |
| Idaho | 4.73 | 1.191 | 2.428 | 5.07 |
| Illinois | 4.509 | 0.947 | 2.402 | 4.396 |
| New Jersey | 3.839 | 1.403 | 2.079 | 3.453 |
| Arizona | 4.004 | 1.652 | 1.274 | 3.46 |
| Georgia | 4.527 | 2.267 | 2.493 | 5.406 |

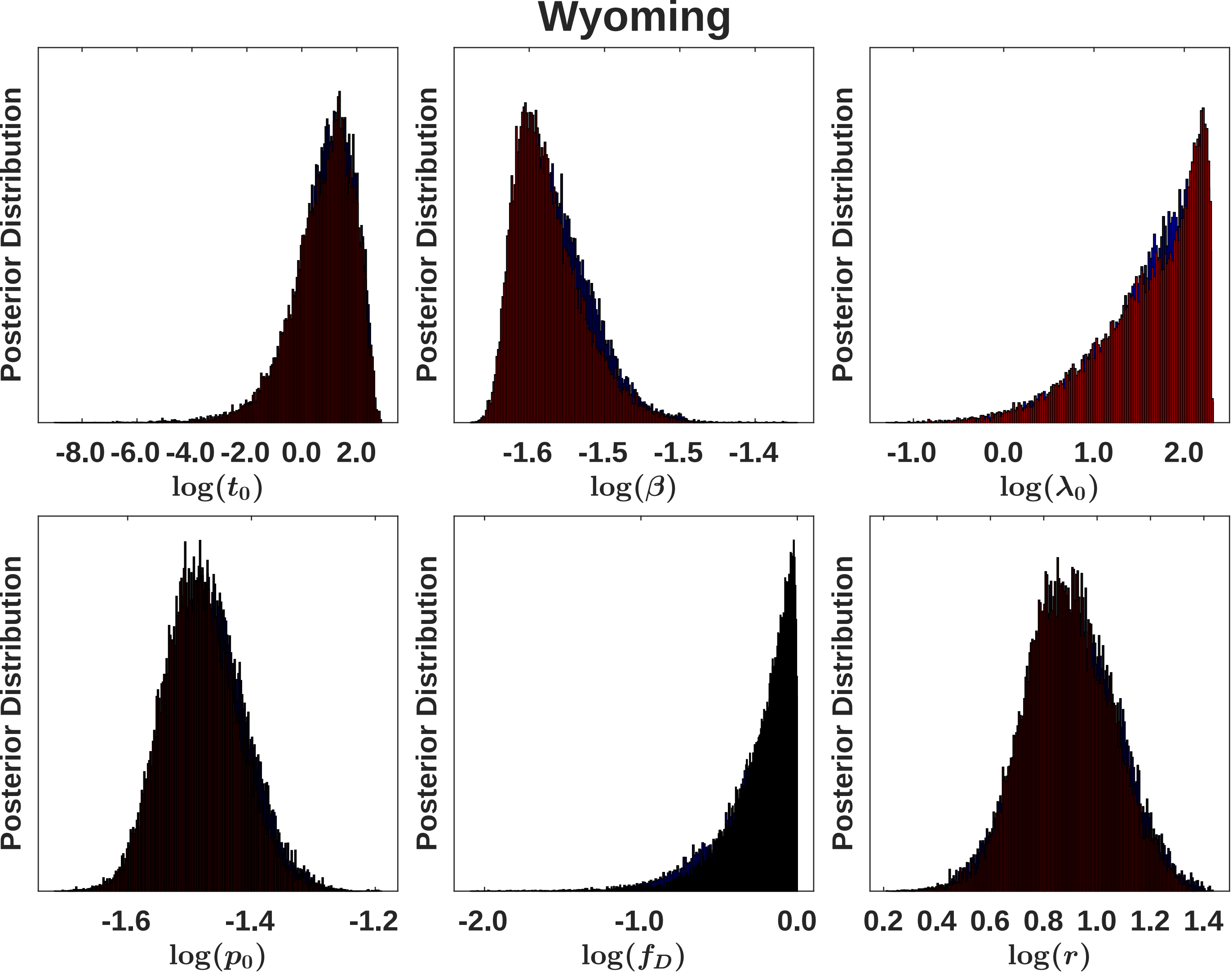

Wyoming
Posterior Distribution
-8.0 -6.0 -4.0 -2.0 0.0 2.0
$\log(t_0)$
-1.6 -1.5 -1.5 -1.4
$\log(\beta)$
-1.0 0.0 1.0 2.0
$\log(\lambda_0)$
-1.6 -1.4 -1.2
$\log(p_0)$
-2.0 -1.0 0.0
$\log(f_D)$
0.2 0.4 0.6 0.8 1.0 1.2 1.4
$\log(r)$

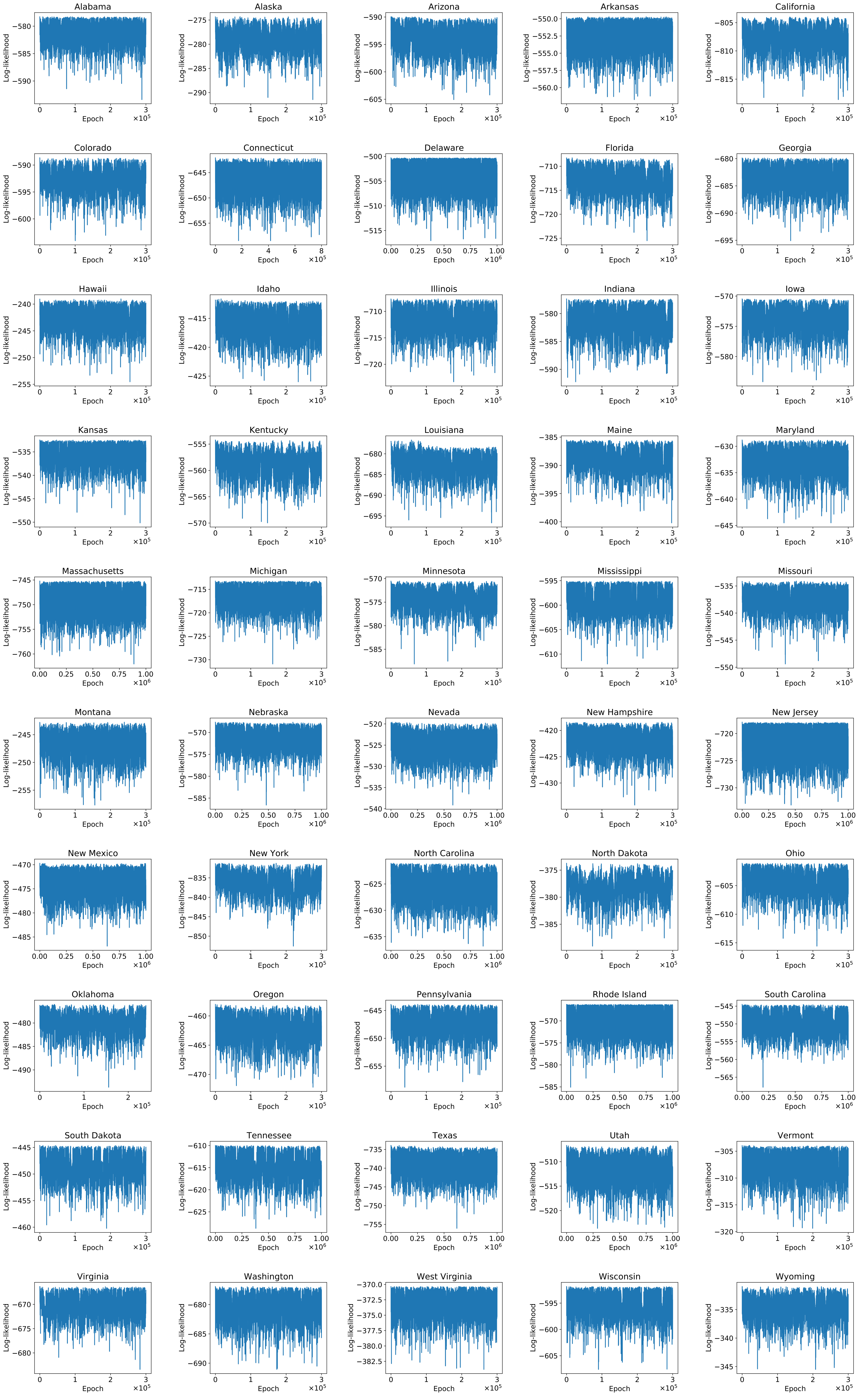
Alabama
Alaska
Arizona
Arkansas
California
Colorado
Connecticut
Delaware
Florida
Georgia
Hawaii
Idaho
Illinois
Indiana
Iowa
Kansas
Kentucky
Louisiana
Maine
Maryland
Massachusetts
Michigan
Minnesota
Mississippi
Missouri
Montana
Nebraska
Nevada
New Hampshire
New Jersey
New Mexico
New York
North Carolina
North Dakota
Ohio
Oklahoma
Oregon
Pennsylvania
Rhode Island
South Carolina
South Dakota
Tennessee
Texas
Utah
Vermont
Virginia
Washington
West Virginia
Wisconsin
Wyoming
Log-likelihood
Epoch

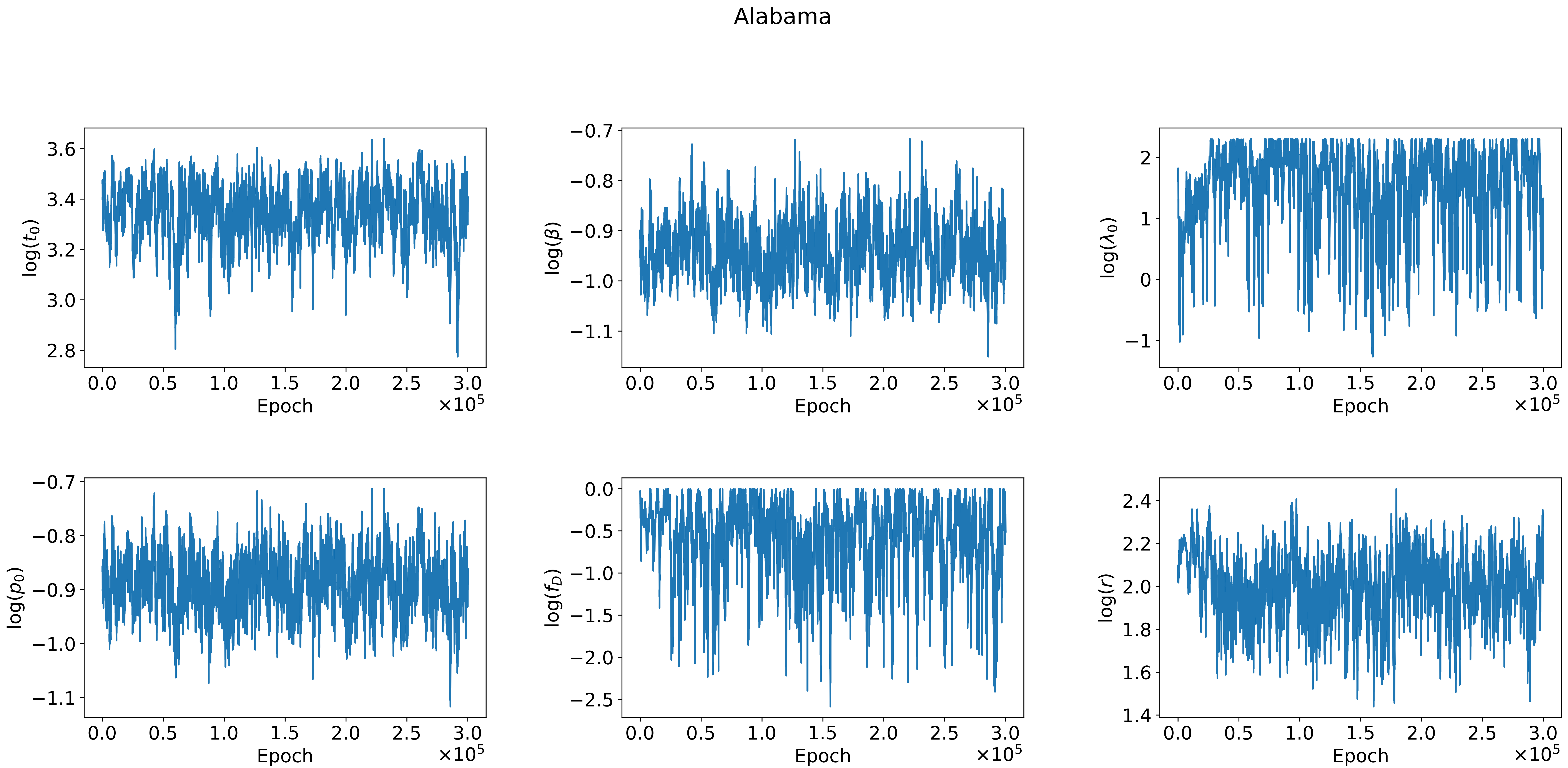

Alabama
log($t_0$)
log($\beta$)
log($\lambda_0$)
log($p_0$)
log($f_D$)
log($r$)
Epoch
$\times 10^5$

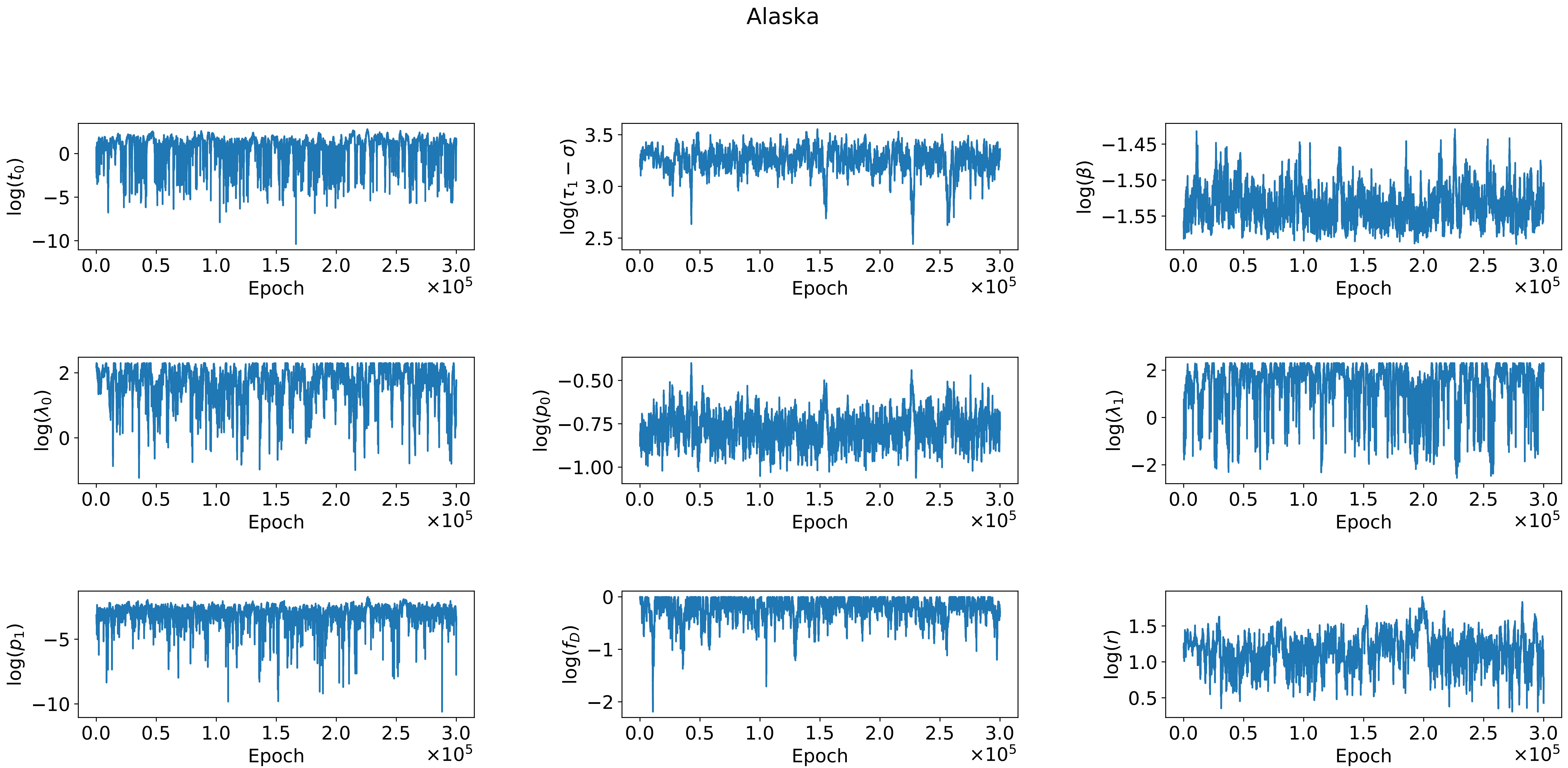

Alaska
log($t_0$)
log($\tau_1 - \sigma$)
log($\beta$)
log($\lambda_0$)
log($p_0$)
log($\lambda_1$)
log($p_1$)
log($f_D$)
log($r$)
Epoch
×10^5

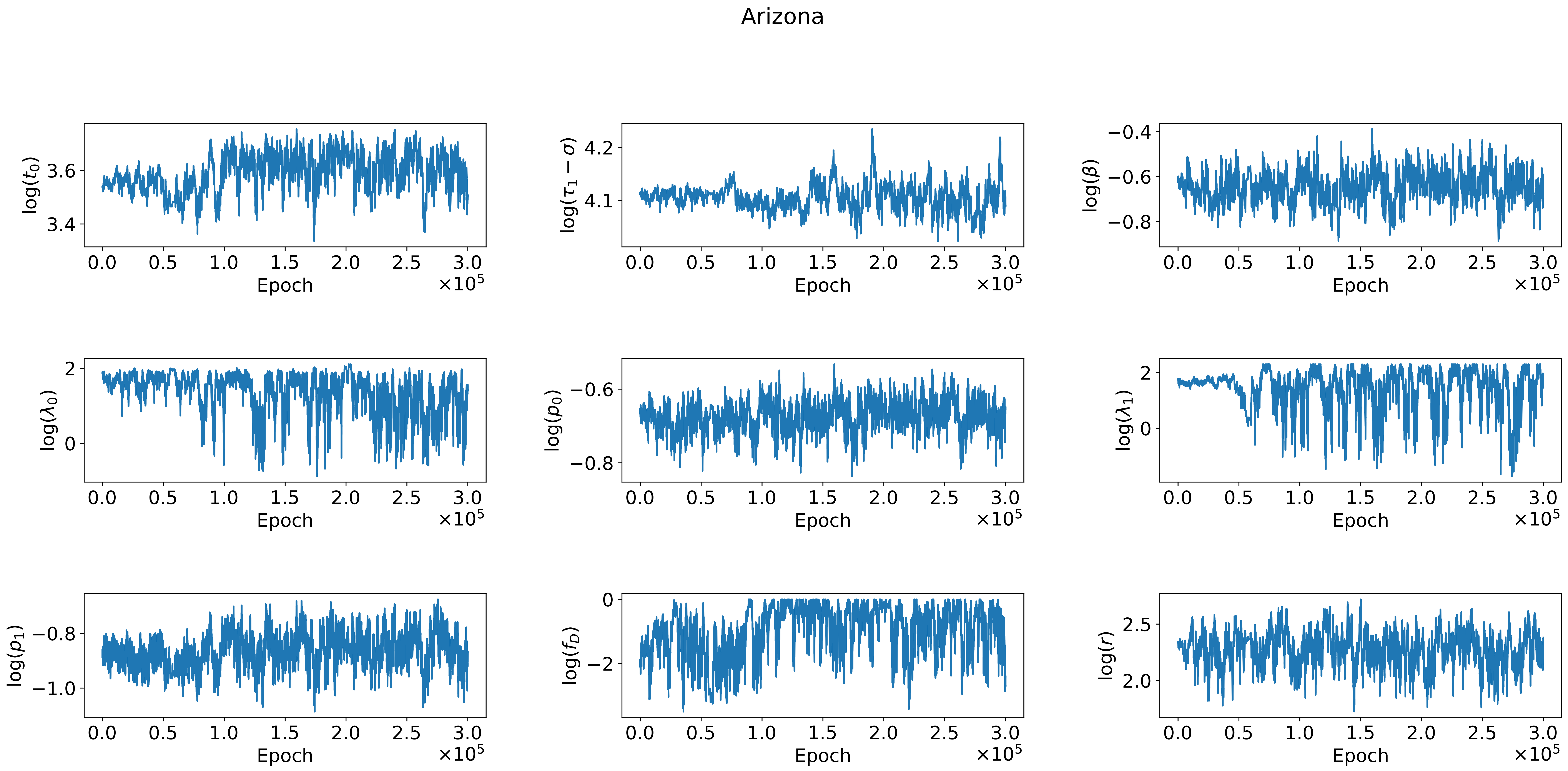
Arizona
log($t_0$)
3.4
3.6
log($\tau_1 - \sigma$)
4.1
4.2
log($\beta$)
−0.8
−0.6
−0.4
log($\lambda_0$)
0
2
log($p_0$)
−0.8
−0.6
log($\lambda_1$)
0
2
log($p_1$)
−1.0
−0.8
log($f_D$)
−2
0
log($r$)
2.0
2.5
0.0
0.5
1.0
1.5
2.0
2.5
3.0
Epoch
×10$^5$

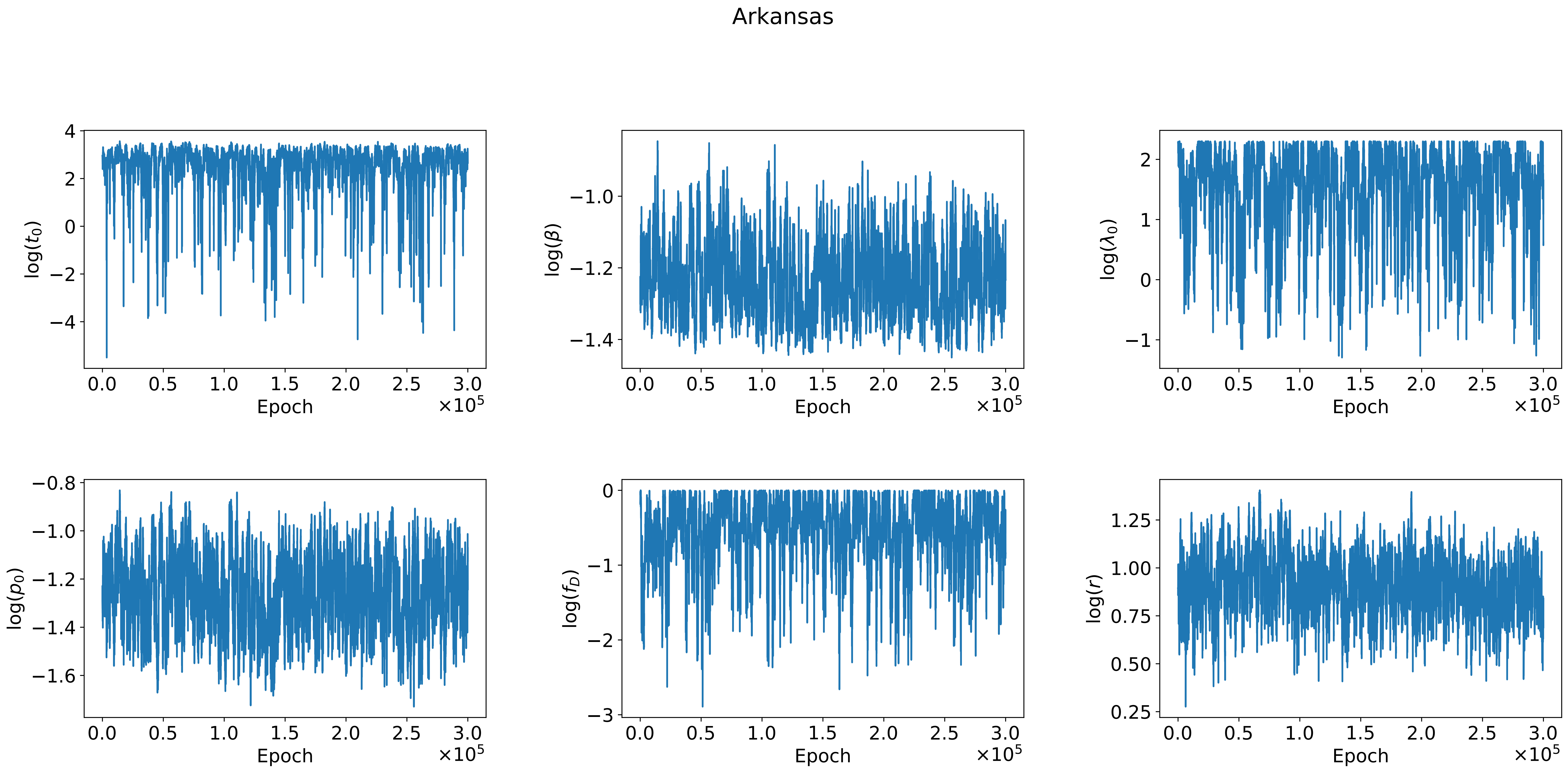
Arkansas
log($t_0$)
log($\beta$)
log($\lambda_0$)
log($p_0$)
log($f_D$)
log($r$)
Epoch
$\times 10^5$

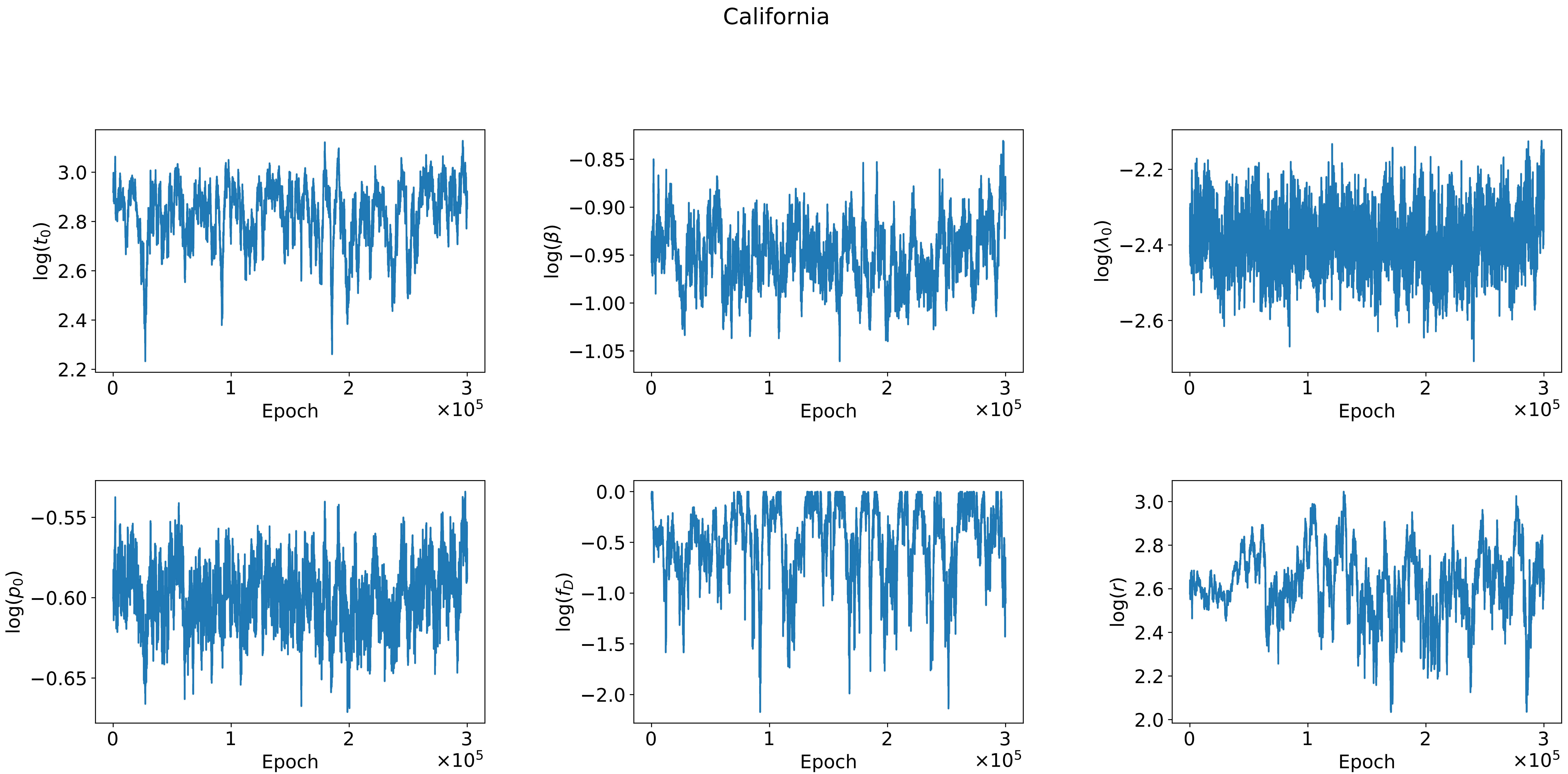

California
3.0
2.8
2.6
2.4
2.2
log($t_0$)
0
1
2
3
Epoch
×10^5
−0.85
−0.90
−0.95
−1.00
−1.05
log($\beta$)
−2.2
−2.4
−2.6
log($\lambda_0$)
−0.55
−0.60
−0.65
log($p_0$)
0.0
−0.5
−1.0
−1.5
−2.0
log($f_D$)
3.0
2.8
2.6
2.4
2.2
2.0
log($r$)

Colorado

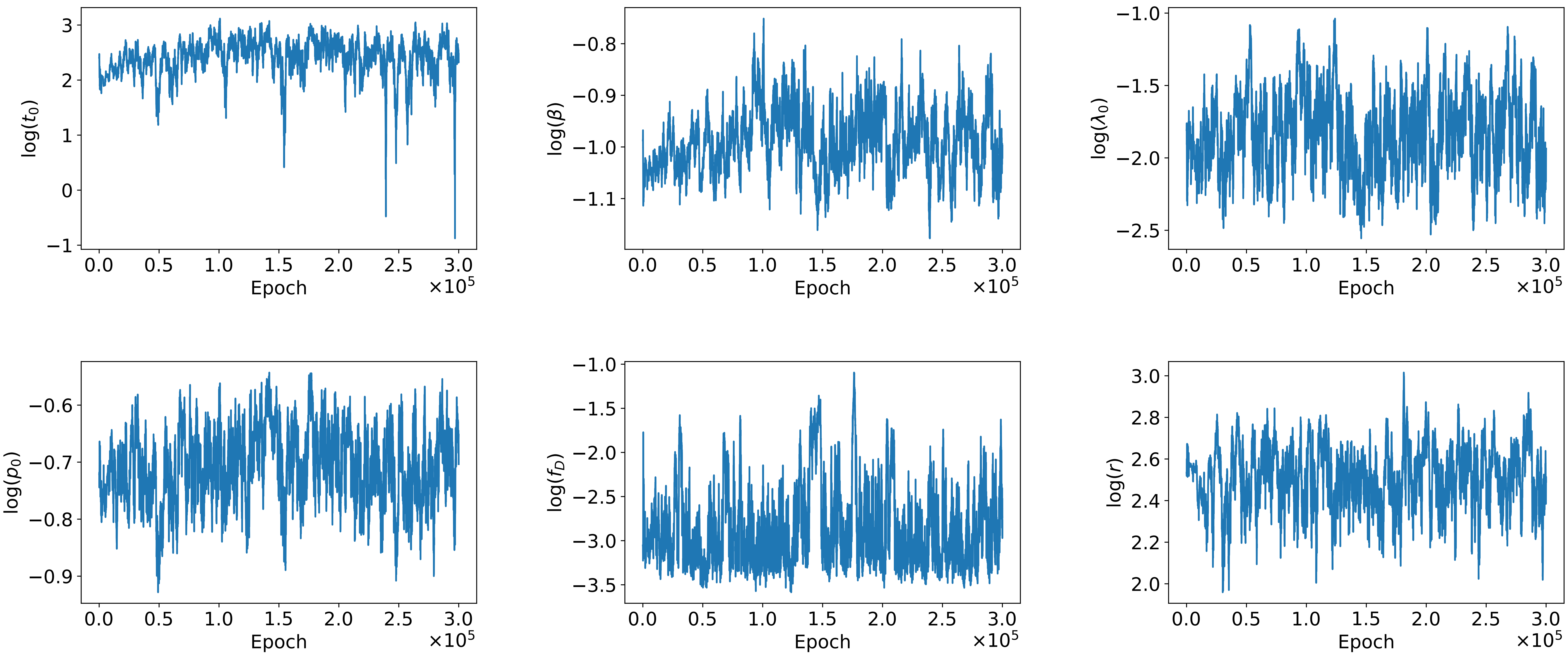

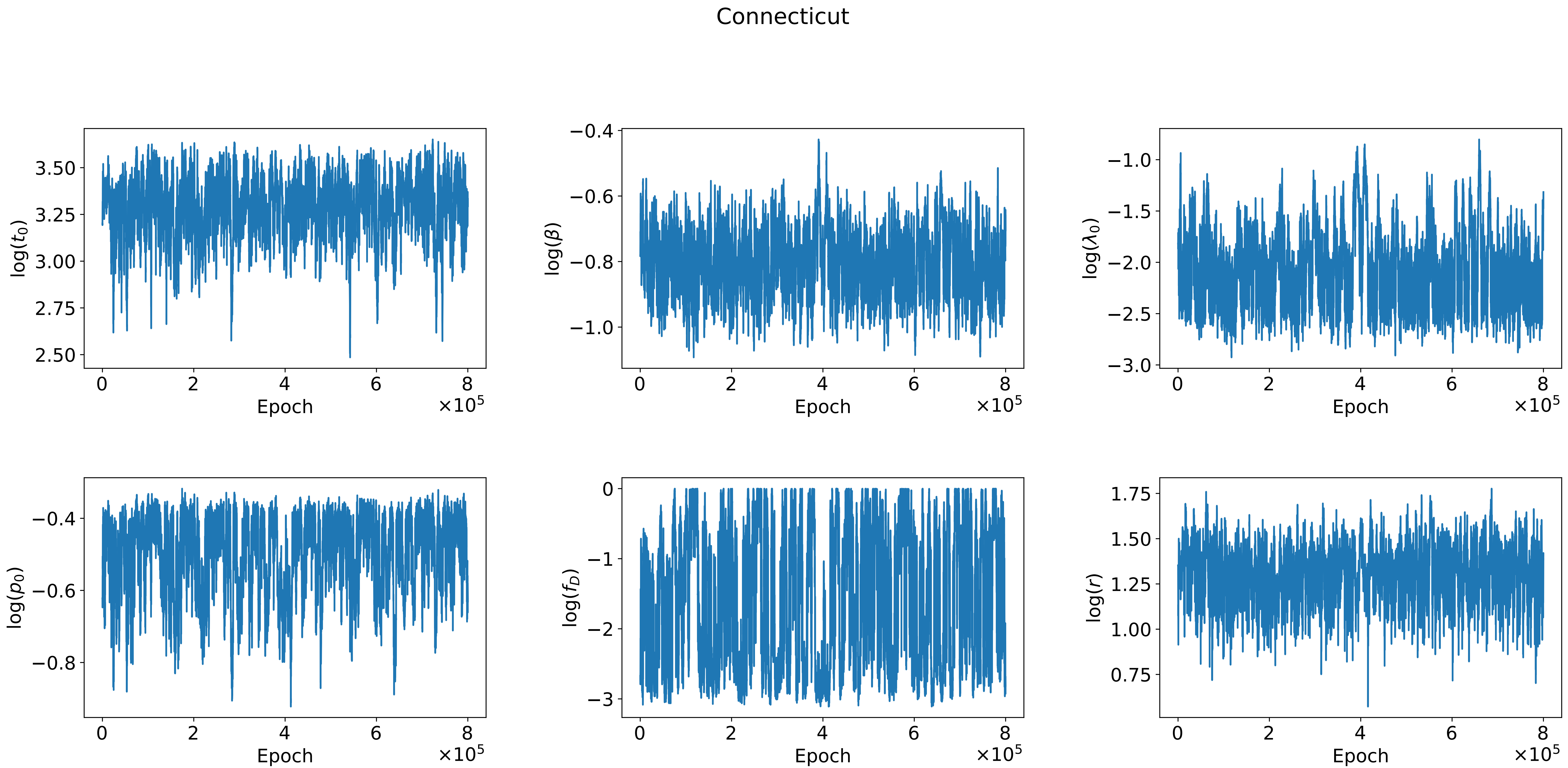

Connecticut
log($t_0$)
log($\beta$)
log($\lambda_0$)
log($p_0$)
log($f_D$)
log($r$)
Epoch
$\times 10^5$

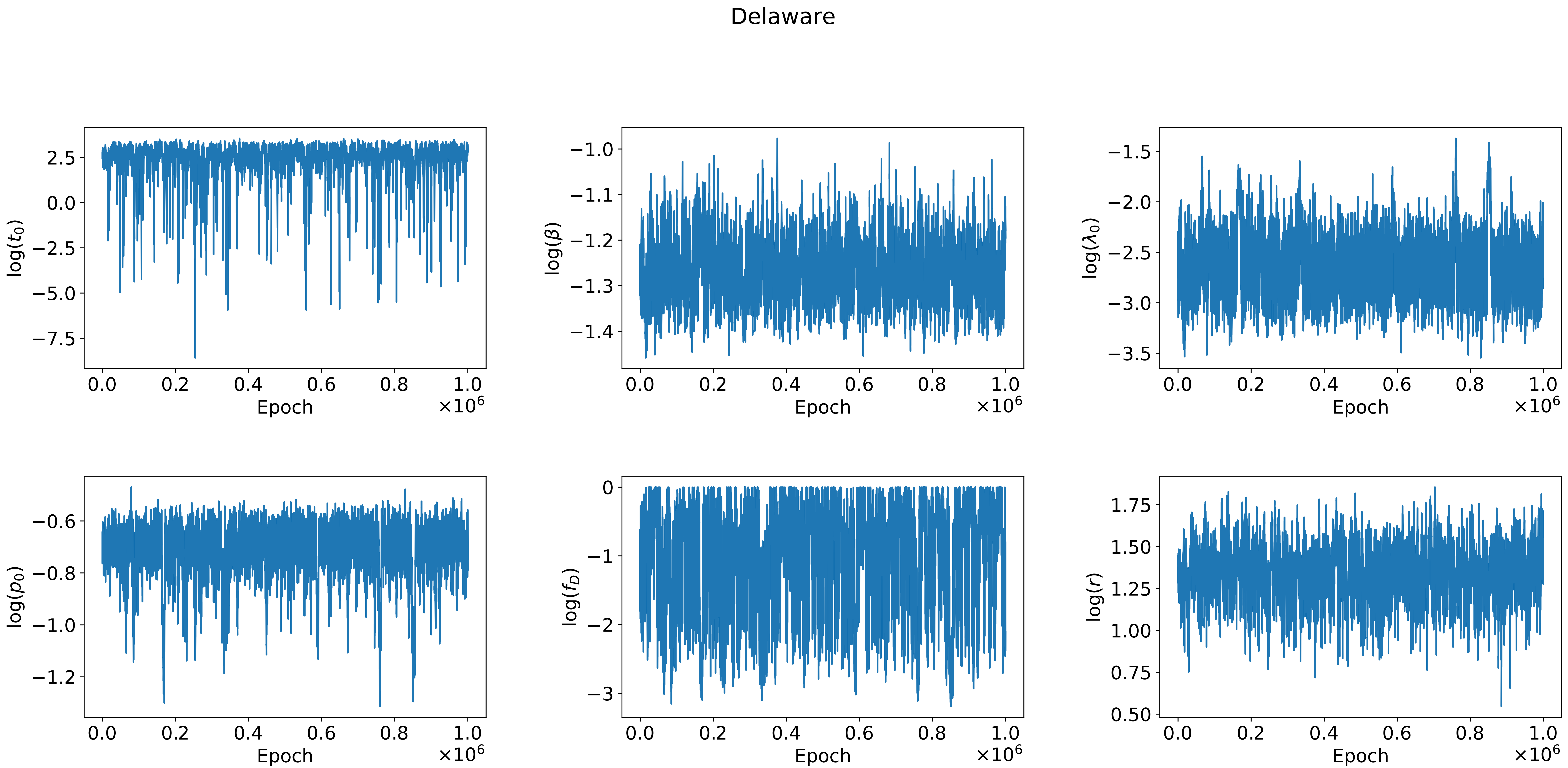

Delaware
log($t_0$)
log($\beta$)
log($\lambda_0$)
log($p_0$)
log($f_D$)
log($r$)
Epoch
$\times 10^6$
2.5
0.0
−2.5
−5.0
−7.5
−1.0
−1.1
−1.2
−1.3
−1.4
−1.5
−2.0
−2.5
−3.0
−3.5
−0.6
−0.8
−1.0
−1.2
0
−1
−2
−3
1.75
1.50
1.25
1.00
0.75
0.50
0.0
0.2
0.4
0.6
0.8
1.0

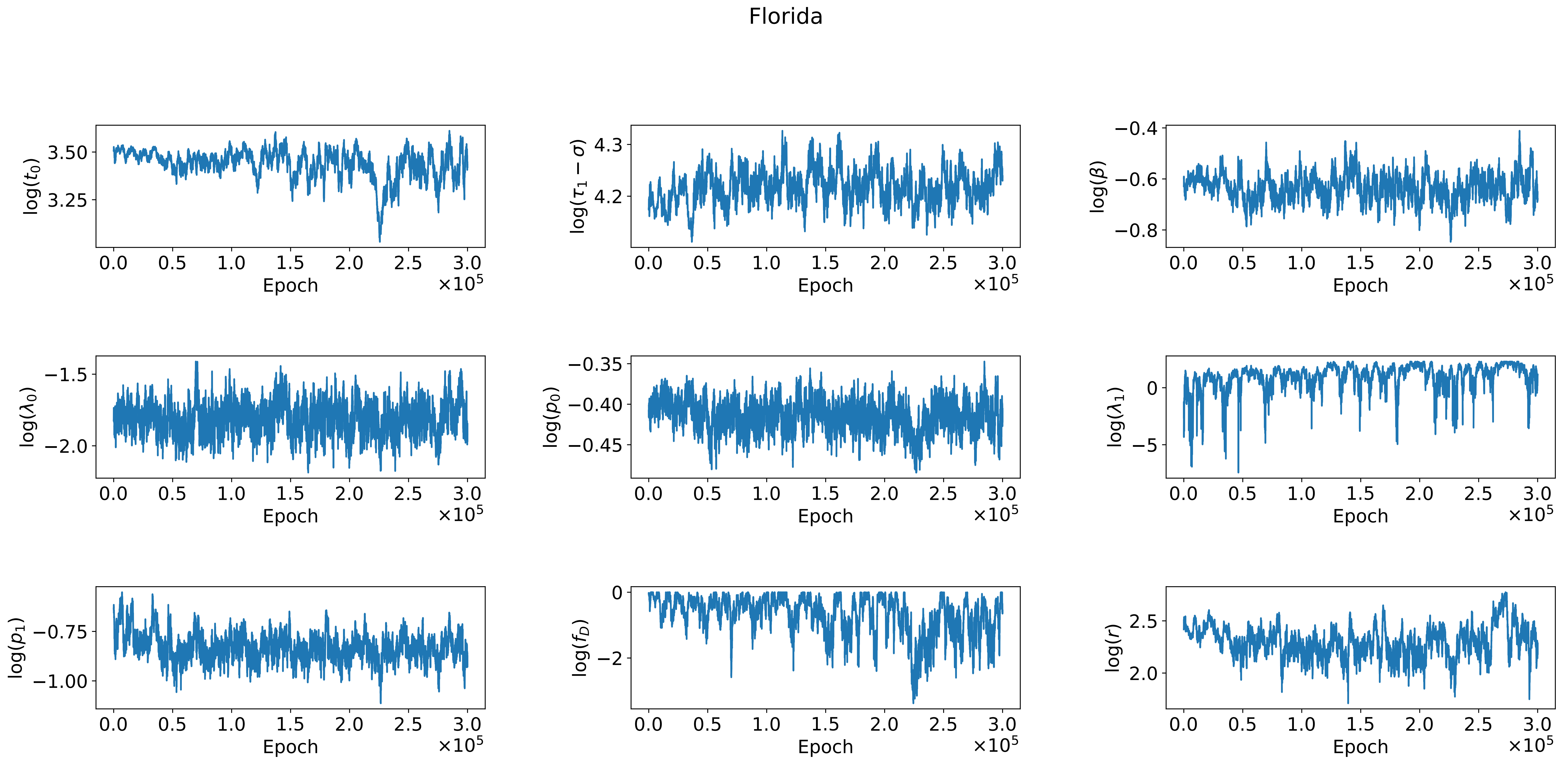
Florida
log($t_0$)
log($\tau_1 - \sigma$)
log($\beta$)
log($\lambda_0$)
log($p_0$)
log($\lambda_1$)
log($p_1$)
log($f_D$)
log($r$)
Epoch
$\times 10^5$

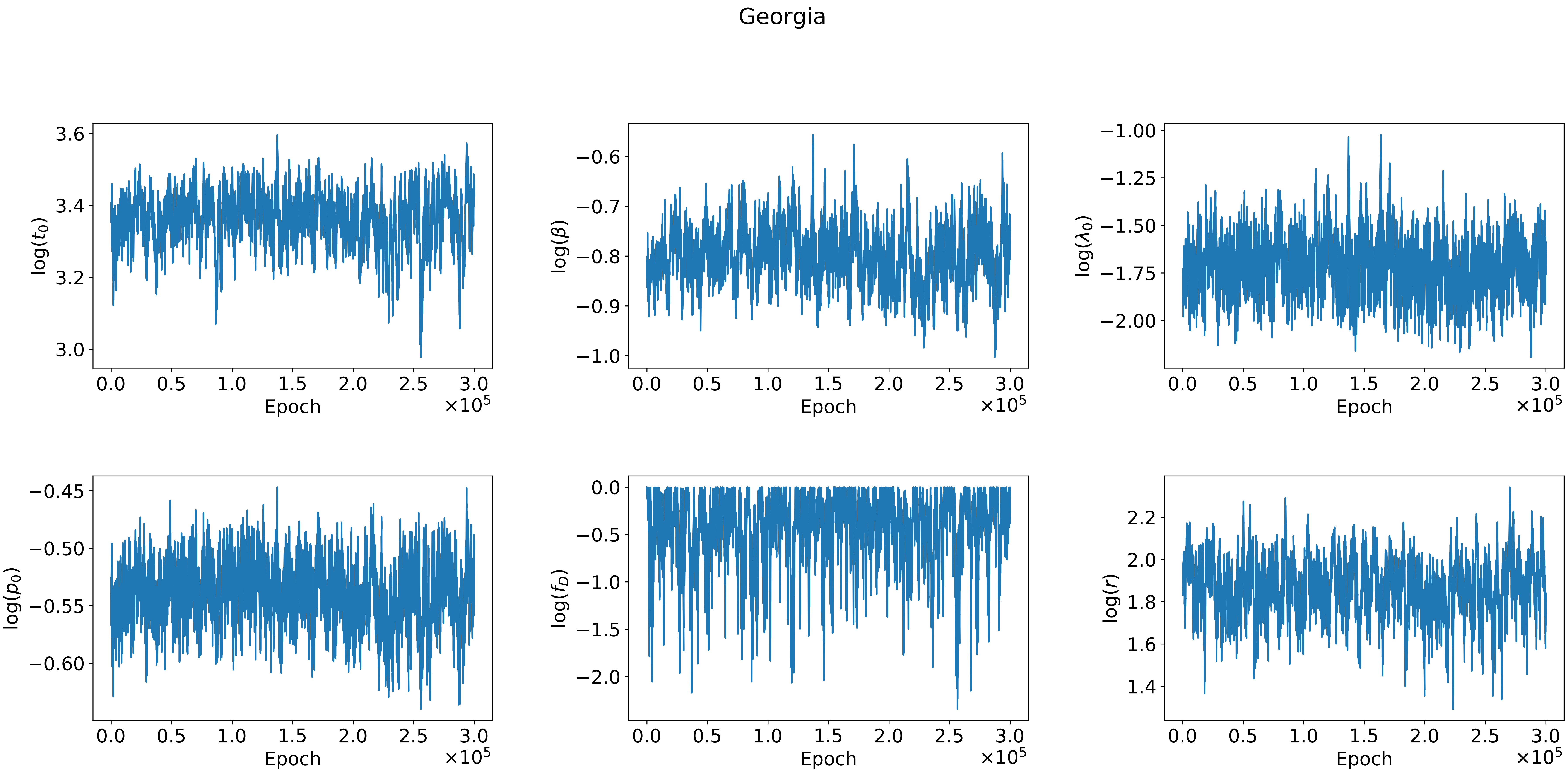
Georgia
log($t_0$)
log($\beta$)
log($\lambda_0$)
log($p_0$)
log($f_D$)
log($r$)
Epoch
$\times 10^5$

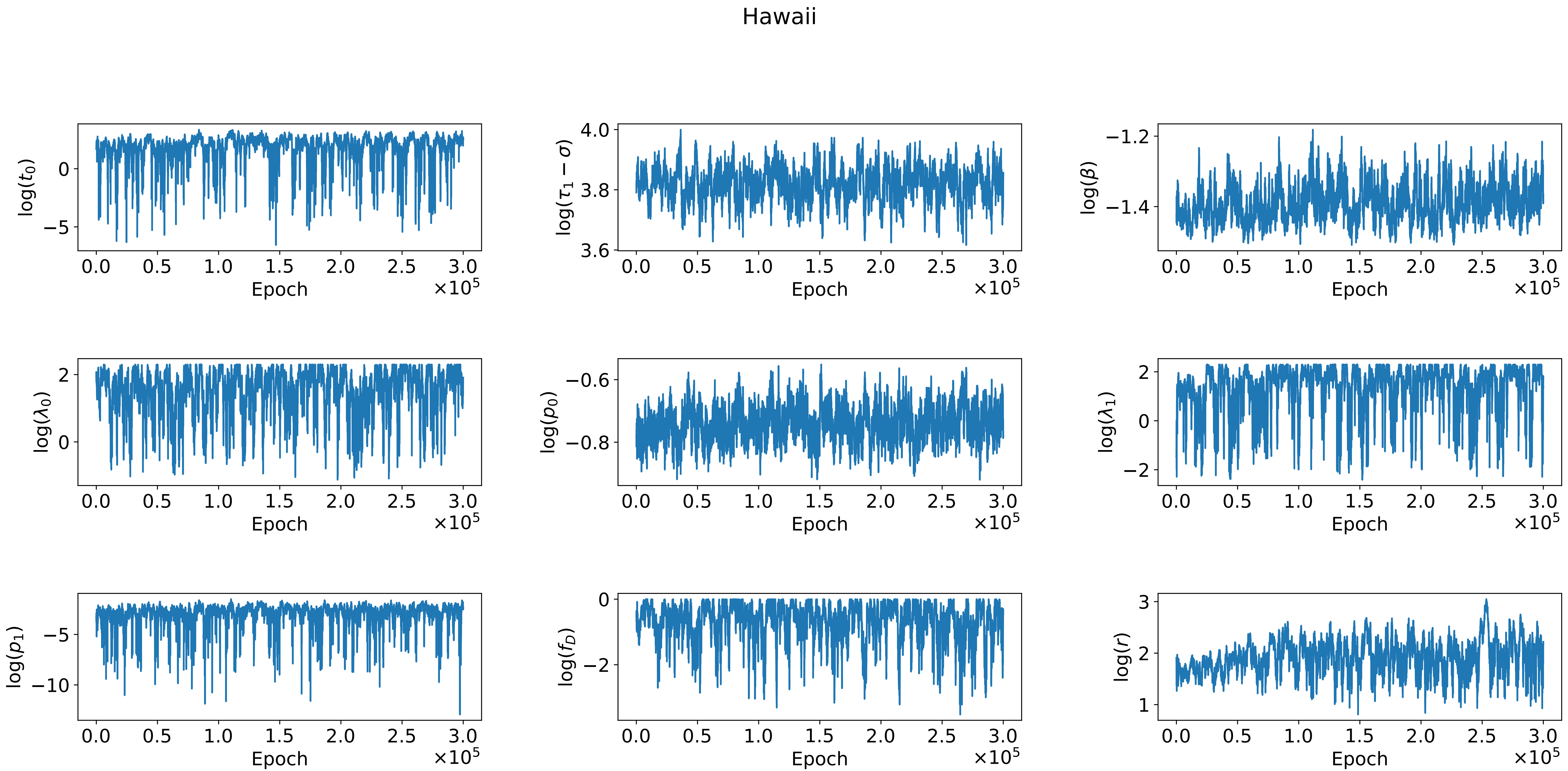

Hawaii
log($t_0$)
log($\tau_1 - \sigma$)
log($\beta$)
log($\lambda_0$)
log($p_0$)
log($\lambda_1$)
log($p_1$)
log($f_D$)
log($r$)
Epoch
$\times 10^5$

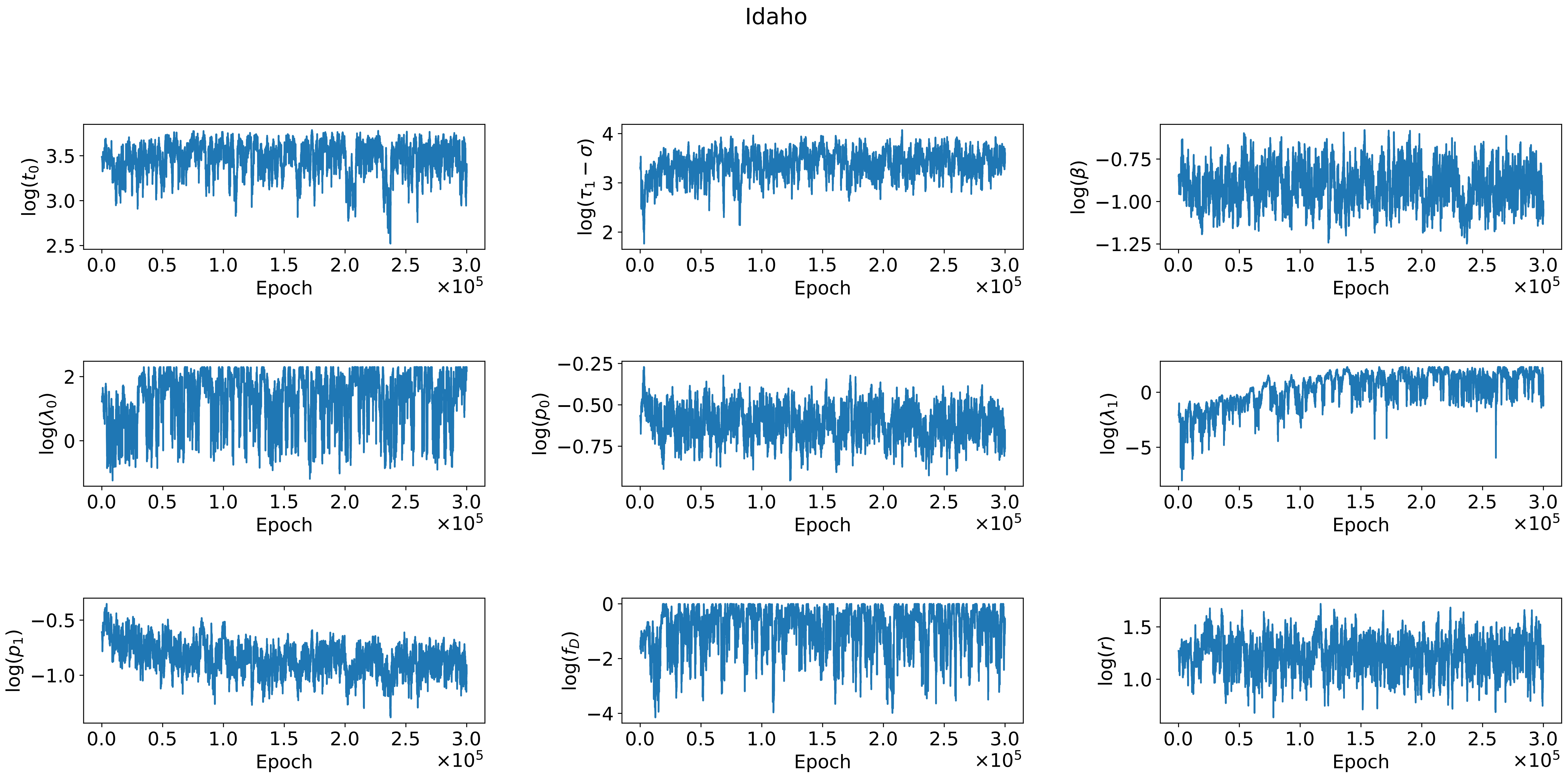

Idaho
log($t_0$)
log($\tau_1 - \sigma$)
log($\beta$)
log($\lambda_0$)
log($p_0$)
log($\lambda_1$)
log($p_1$)
log($f_D$)
log($r$)
Epoch
$\times 10^5$

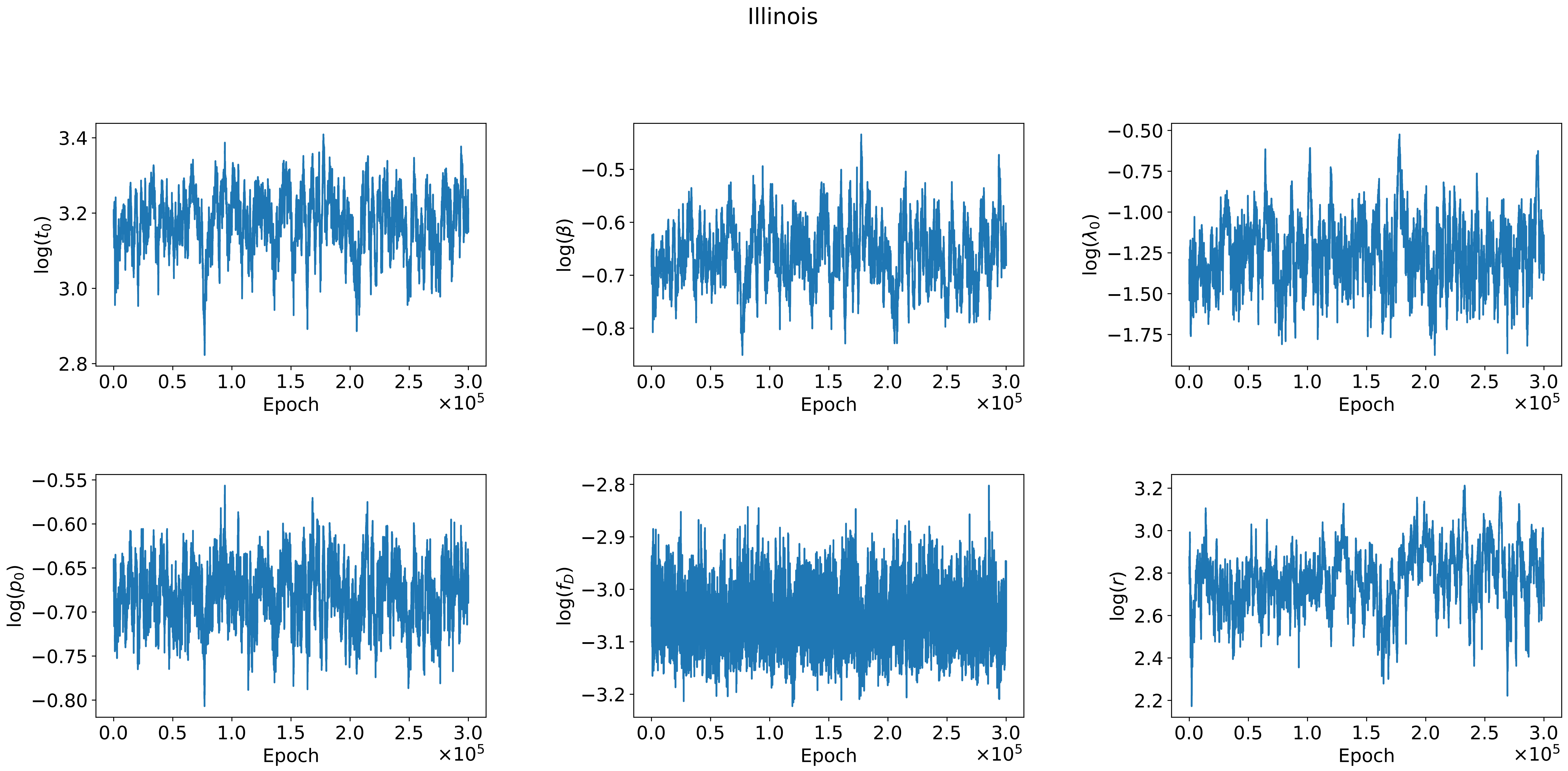
Illinois
log($t_0$)
log($\beta$)
log($\lambda_0$)
log($p_0$)
log($f_D$)
log($r$)
Epoch
$\times 10^5$

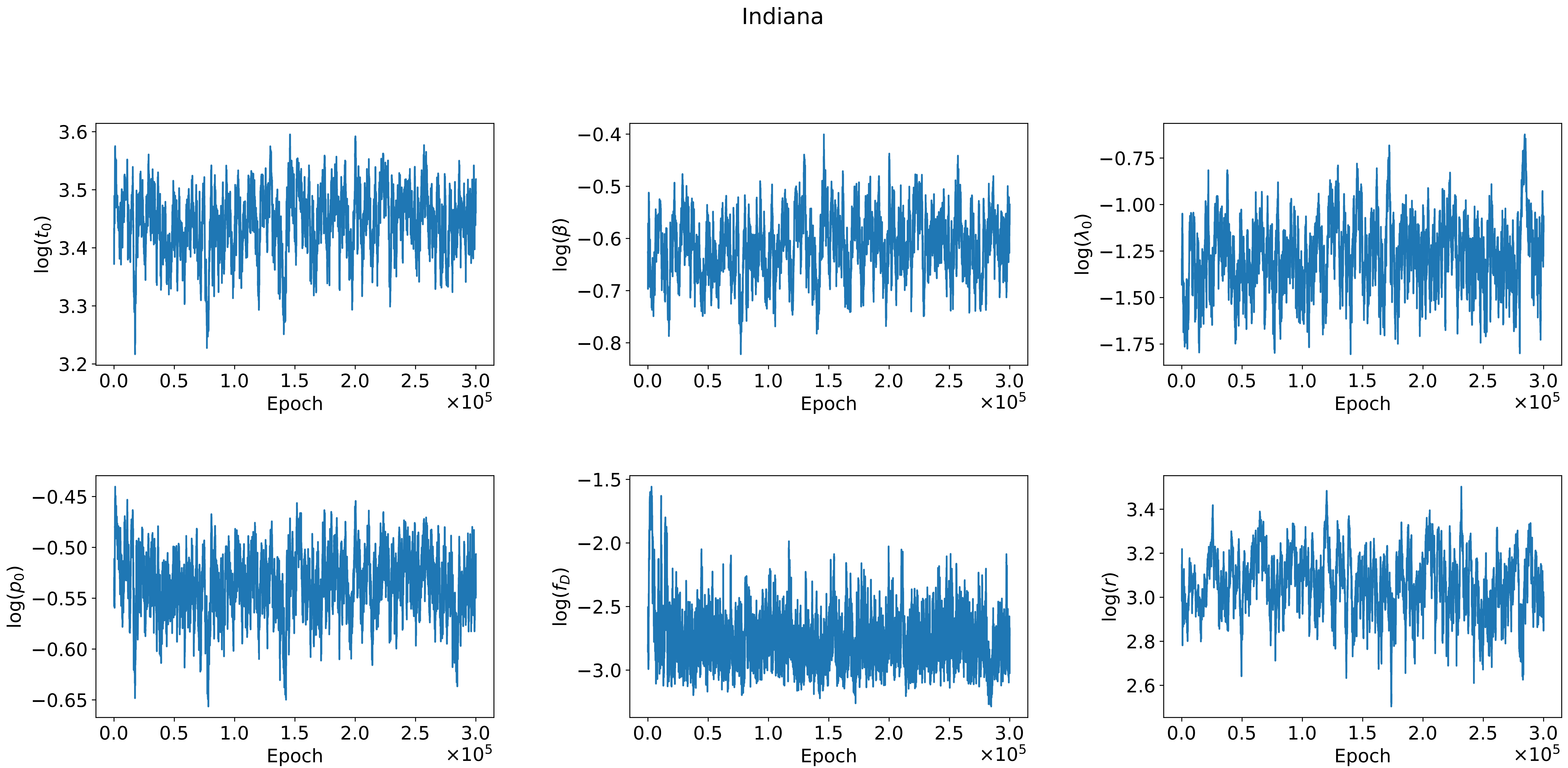
Indiana
3.6
3.5
3.4
3.3
3.2
$\log(t_0)$
0.0
0.5
1.0
1.5
2.0
2.5
3.0
Epoch
$\times10^5$
−0.4
−0.5
−0.6
−0.7
−0.8
$\log(\beta)$
−0.75
−1.00
−1.25
−1.50
−1.75
$\log(\lambda_0)$
−0.45
−0.50
−0.55
−0.60
−0.65
$\log(p_0)$
−1.5
−2.0
−2.5
−3.0
$\log(f_D)$
3.4
3.2
3.0
2.8
2.6
$\log(r)$

Iowa

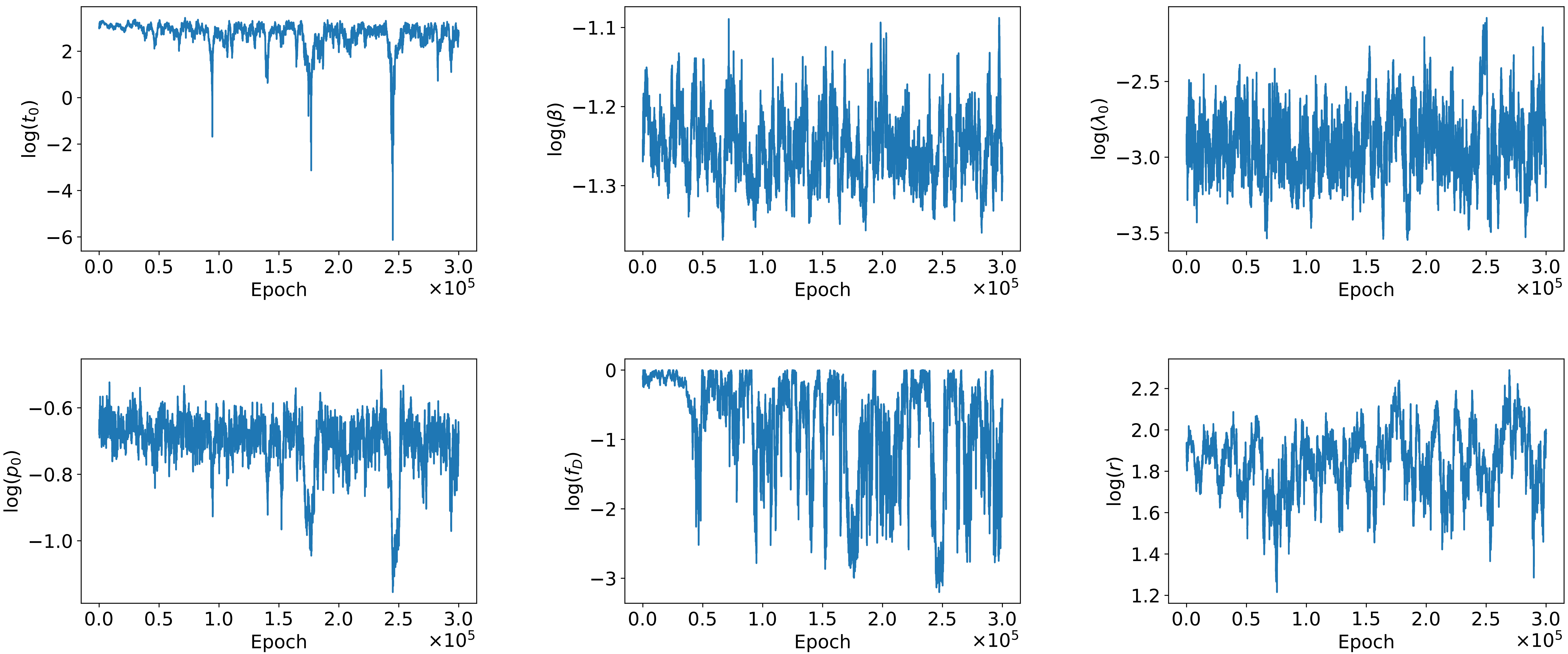

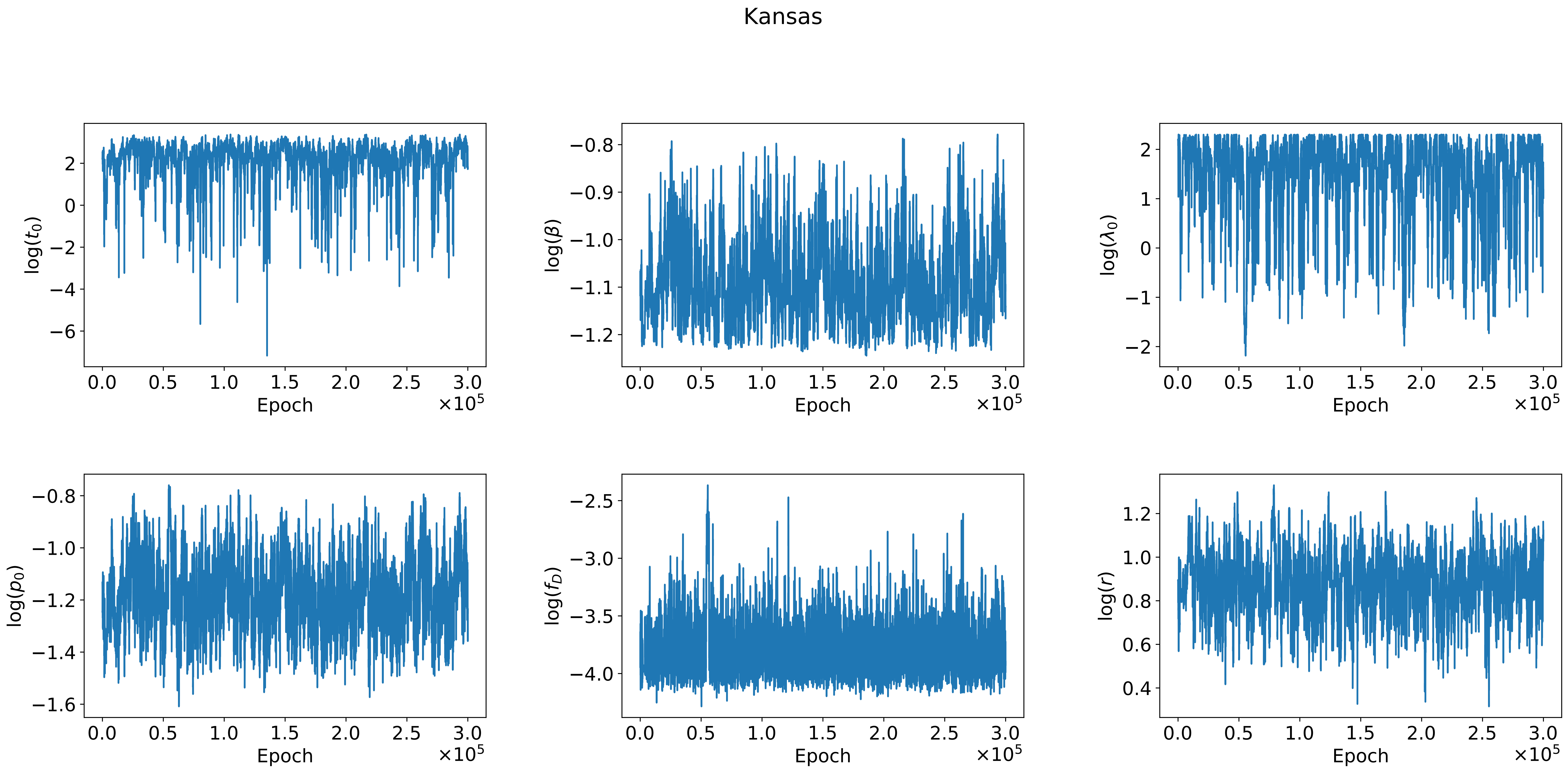

Kansas
log($t_0$)
log($\beta$)
log($\lambda_0$)
log($p_0$)
log($f_D$)
log($r$)
Epoch
$\times 10^5$

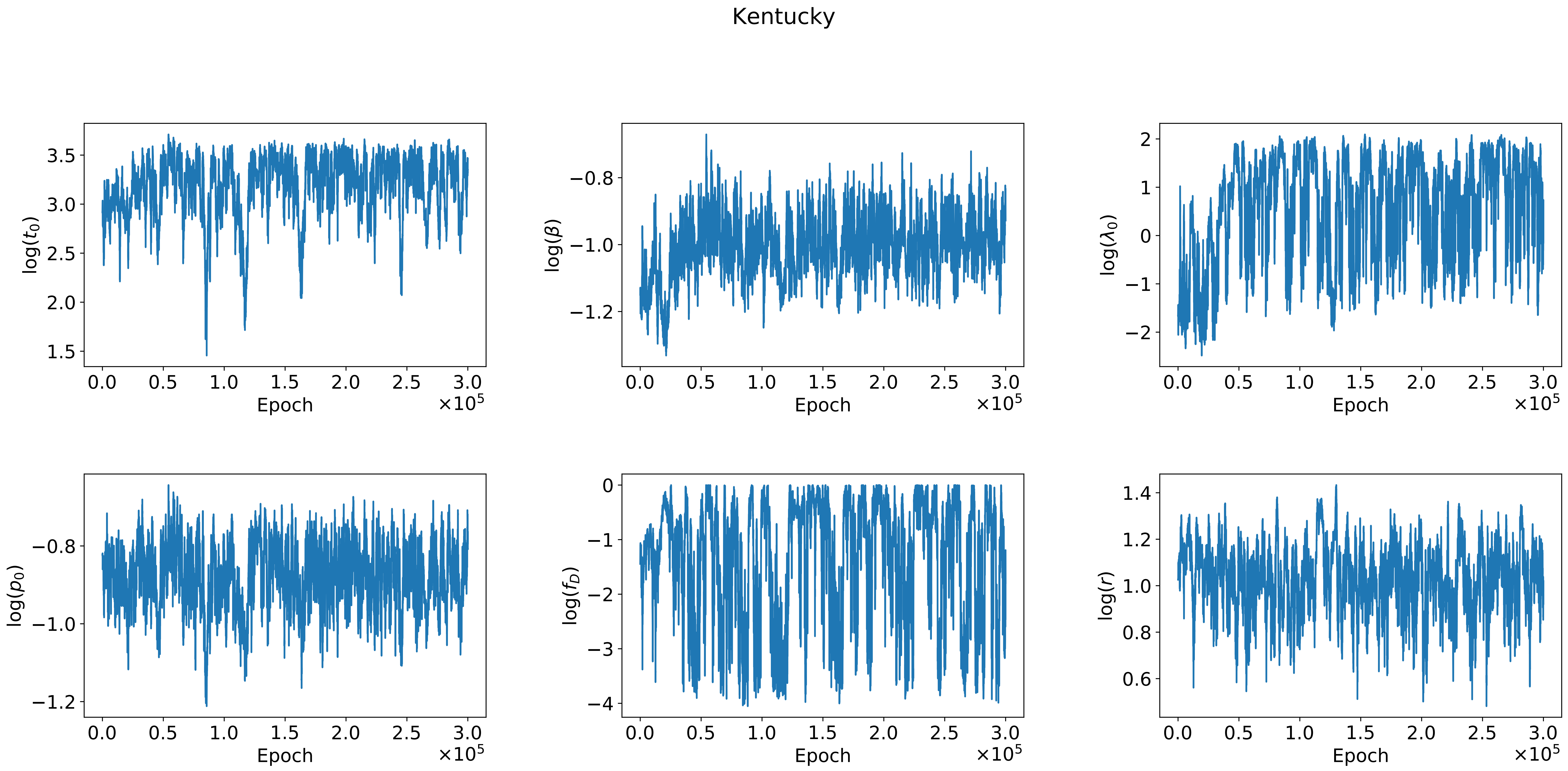
Kentucky
log($t_0$)
log($\beta$)
log($\lambda_0$)
log($p_0$)
log($f_D$)
log($r$)
Epoch
$\times 10^5$

Louisiana

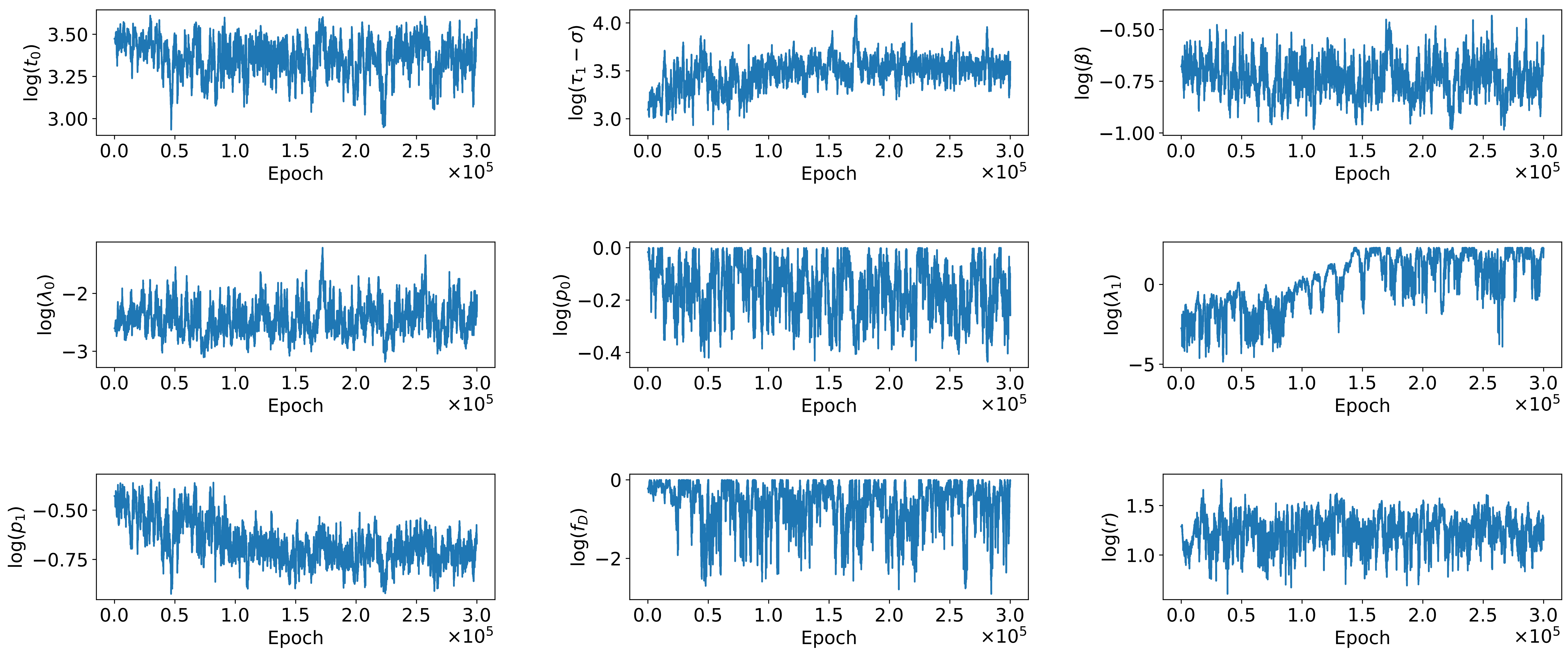

log($t_0$)
3.50
3.25
3.00
log($\tau_1 - \sigma$)
4.0
3.5
3.0
log($\beta$)
−0.50
−0.75
−1.00
log($\lambda_0$)
−2
−3
log($p_0$)
0.0
−0.2
−0.4
log($\lambda_1$)
0
−5
log($p_1$)
−0.50
−0.75
log($f_D$)
0
−2
log($r$)
1.5
1.0
0.0 0.5 1.0 1.5 2.0 2.5 3.0
Epoch
$\times 10^5$

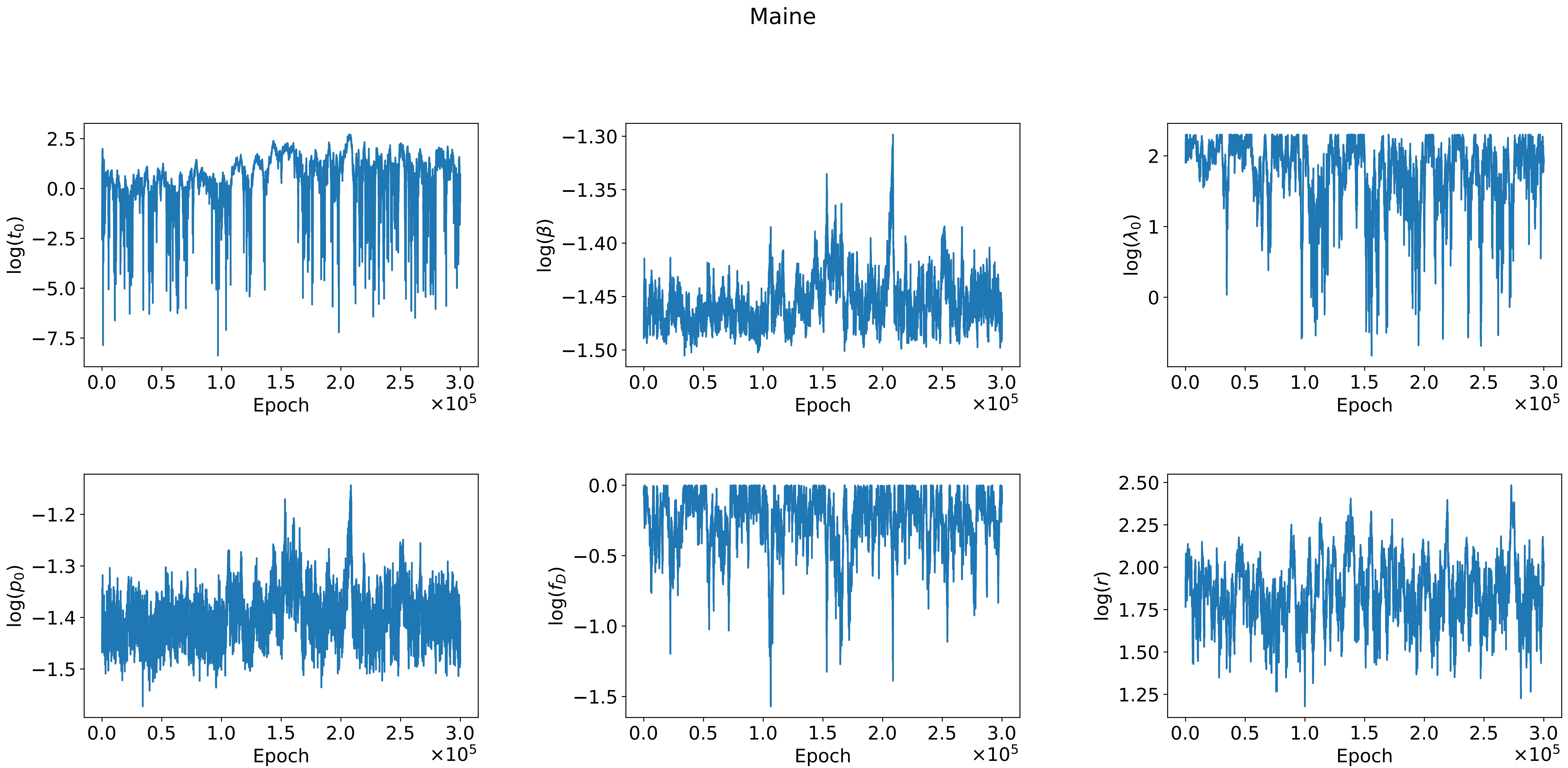

Maine
log($t_0$)
log($\beta$)
log($\lambda_0$)
log($p_0$)
log($f_D$)
log($r$)
Epoch
$\times 10^5$

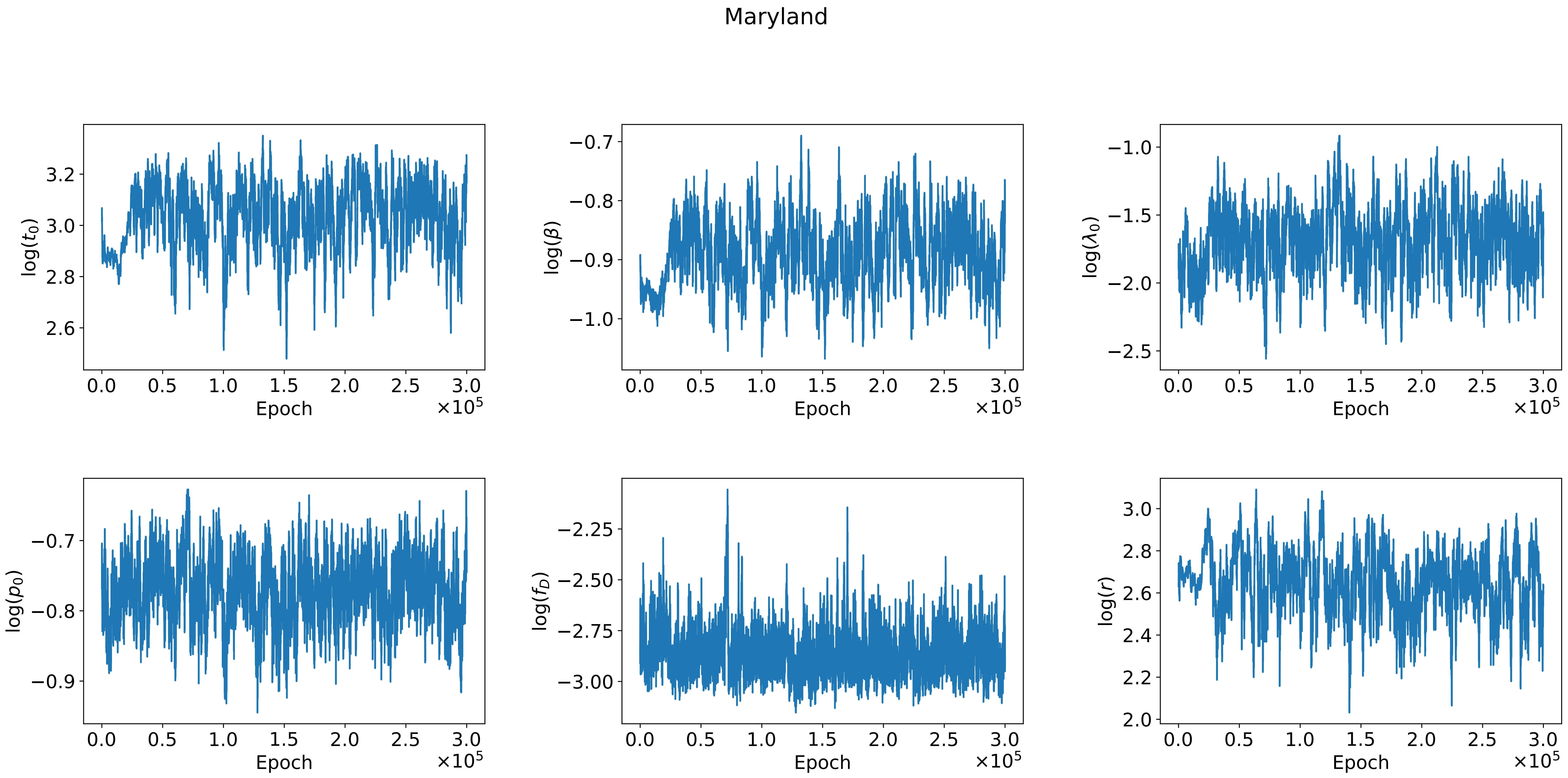

Maryland
log($t_0$)
log($\beta$)
log($\lambda_0$)
log($p_0$)
log($f_D$)
log($r$)
Epoch
$\times 10^5$

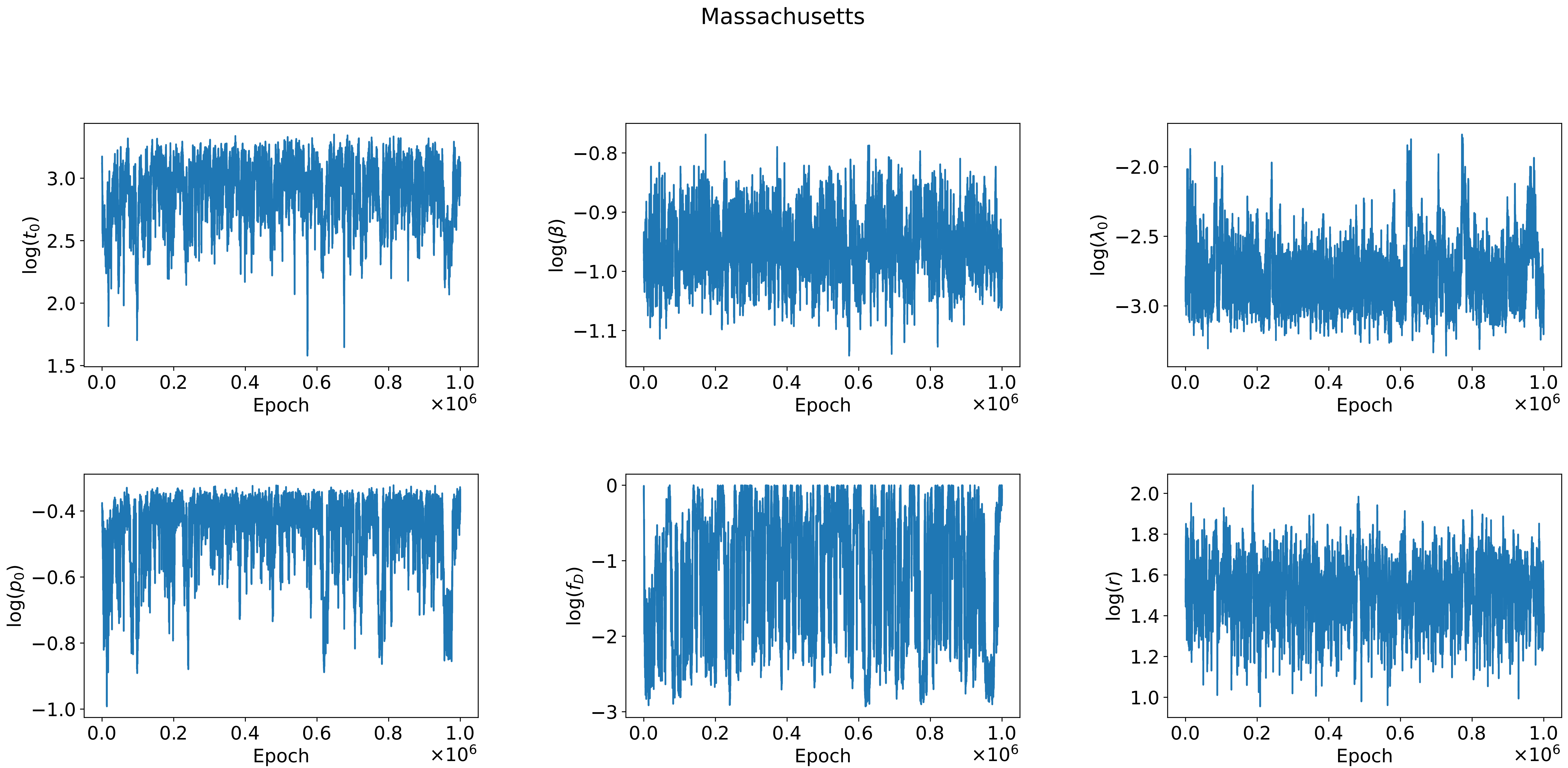
Massachusetts
log($t_0$)
log($\beta$)
log($\lambda_0$)
log($p_0$)
log($f_D$)
log($r$)
Epoch
$\times10^6$

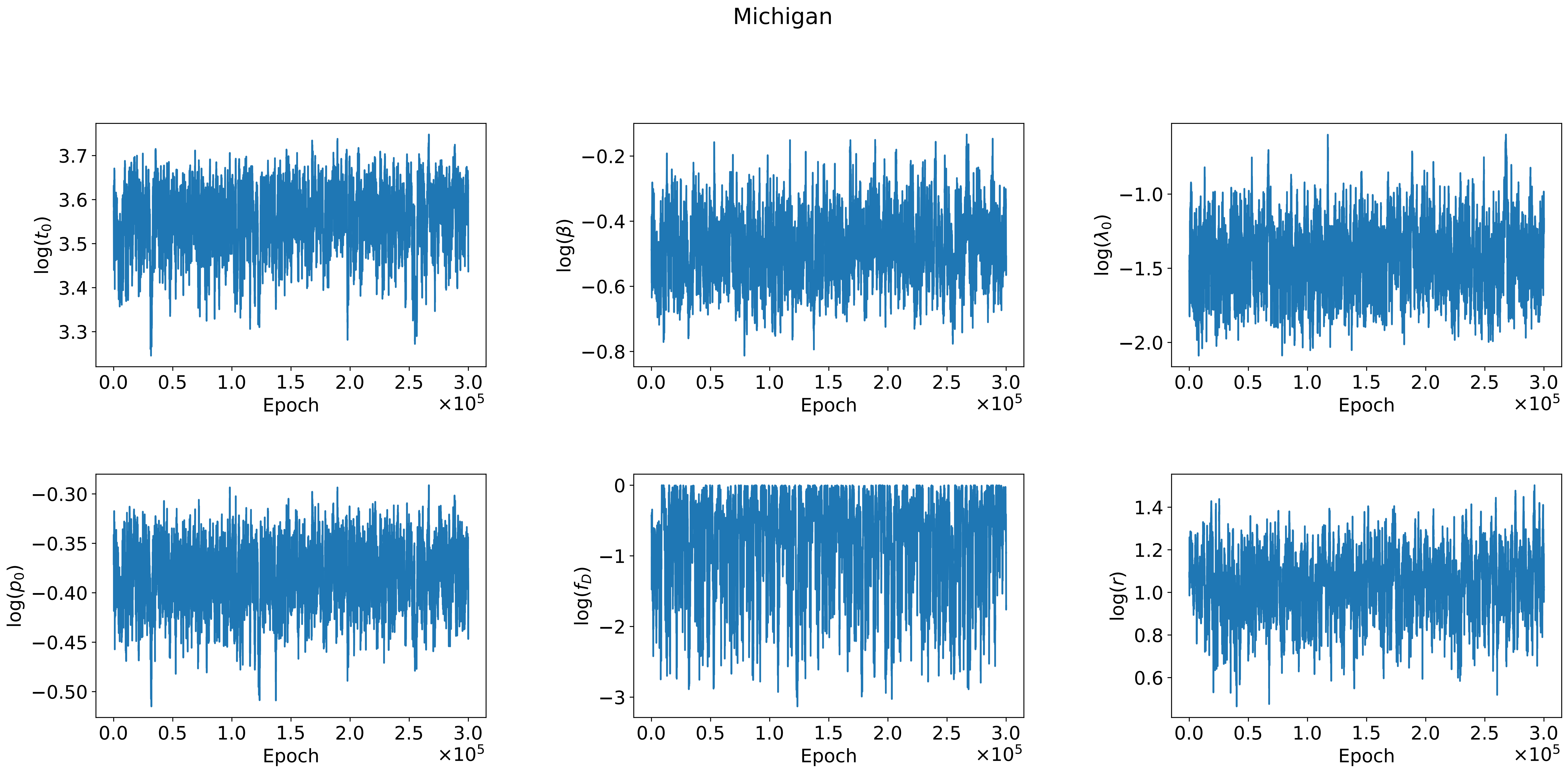
Michigan
log($t_0$)
log($\beta$)
log($\lambda_0$)
log($p_0$)
log($f_D$)
log($r$)
Epoch
$\times 10^5$

Minnesota

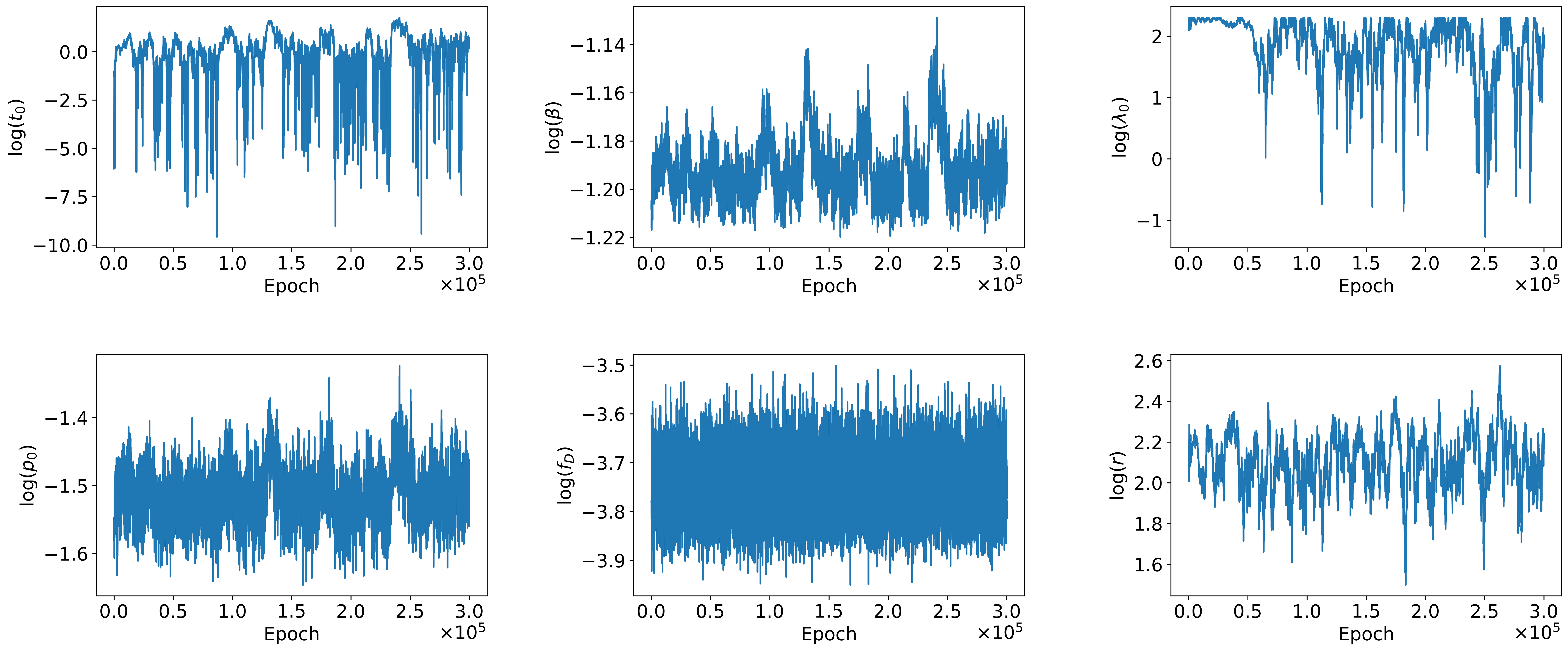

log($t_0$)
0.0
−2.5
−5.0
−7.5
−10.0
log($\beta$)
−1.14
−1.16
−1.18
−1.20
−1.22
log($\lambda_0$)
2
1
0
−1
log($p_0$)
−1.4
−1.5
−1.6
log($f_D$)
−3.5
−3.6
−3.7
−3.8
−3.9
log($r$)
2.6
2.4
2.2
2.0
1.8
1.6
0.0 0.5 1.0 1.5 2.0 2.5 3.0
Epoch
$\times 10^5$

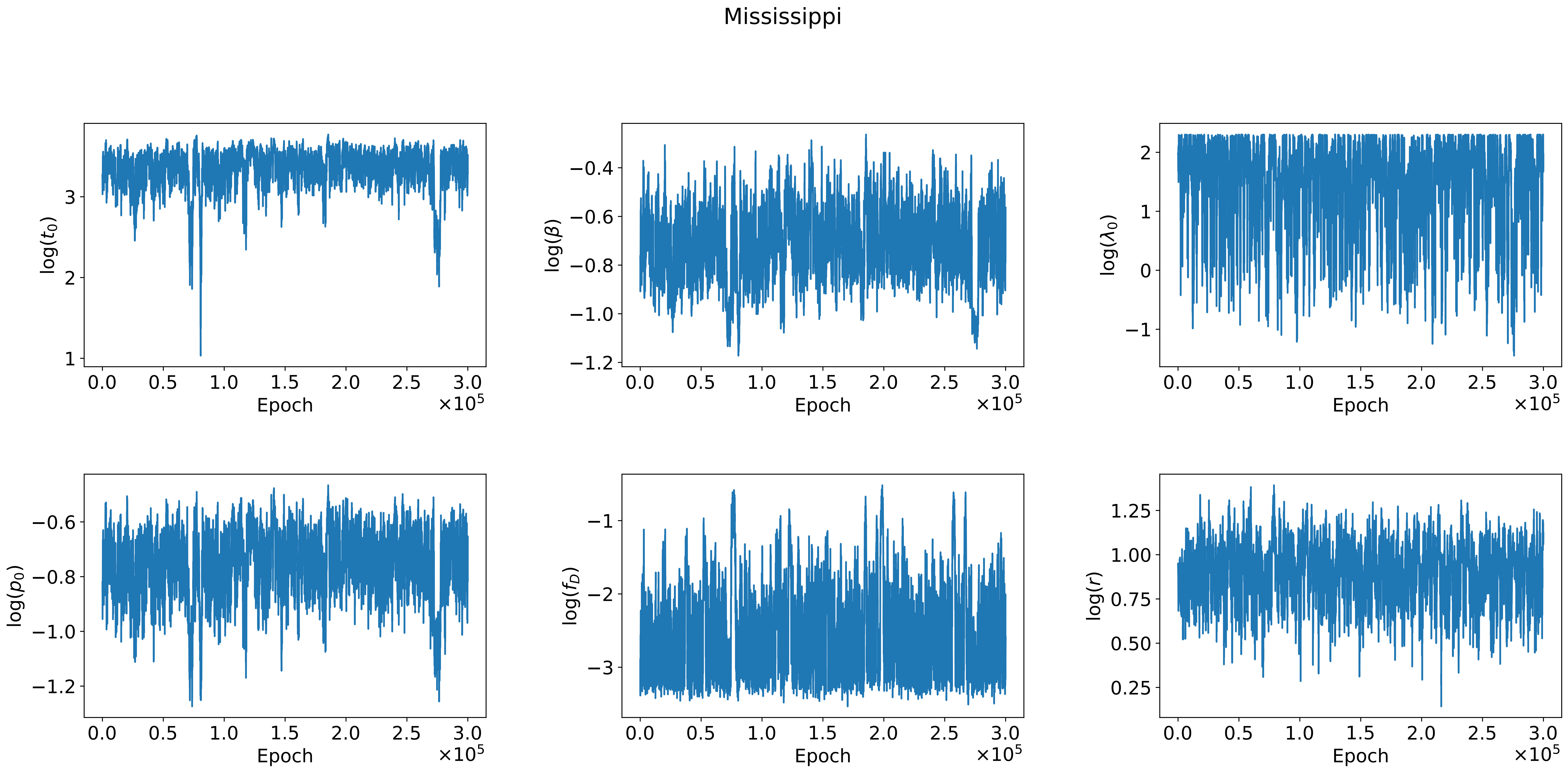
Mississippi
log($t_0$)
log($\beta$)
log($\lambda_0$)
log($p_0$)
log($f_D$)
log($r$)
Epoch
$\times10^5$

Missouri

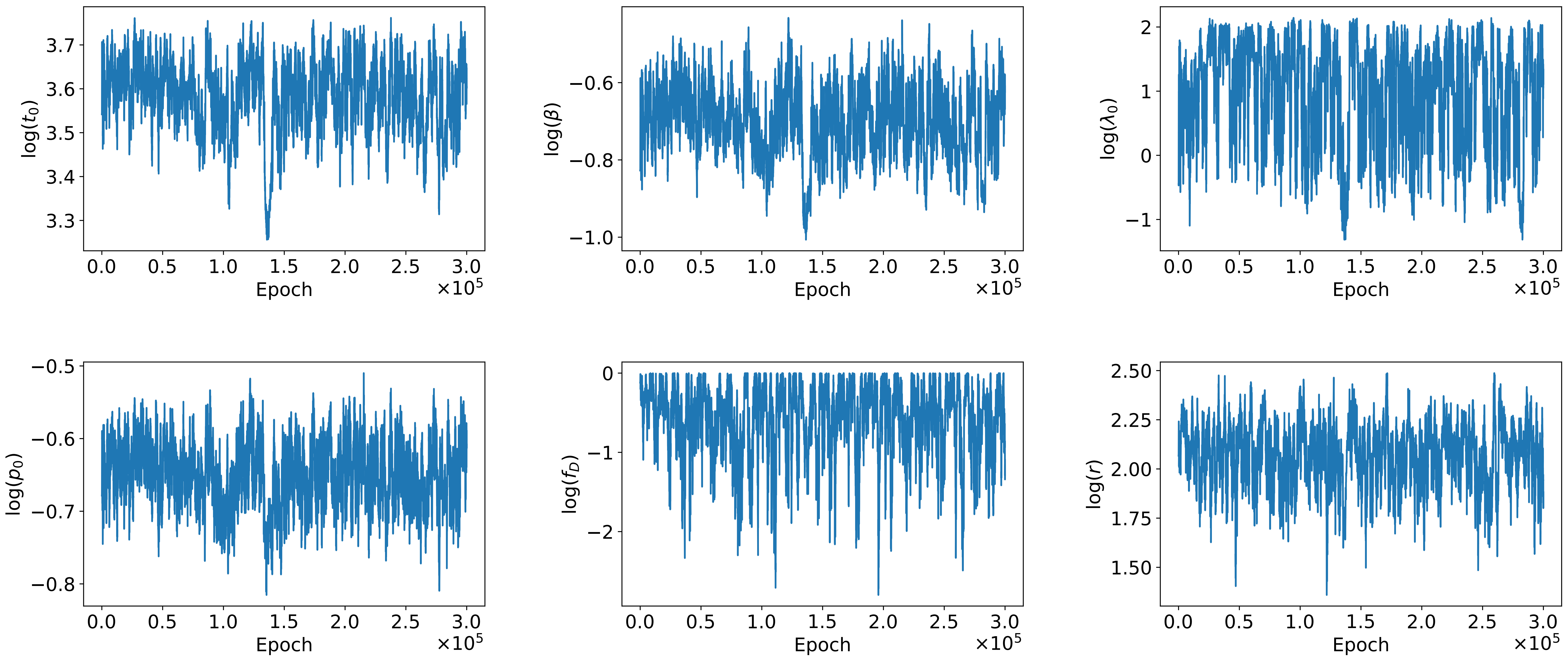

log($t_0$)
3.7
3.6
3.5
3.4
3.3
log($\beta$)
−0.6
−0.8
−1.0
log($\lambda_0$)
2
1
0
−1
log($p_0$)
−0.5
−0.6
−0.7
−0.8
log($f_D$)
0
−1
−2
log($r$)
2.50
2.25
2.00
1.75
1.50
0.0
0.5
1.0
1.5
2.0
2.5
3.0
Epoch
$\times 10^5$

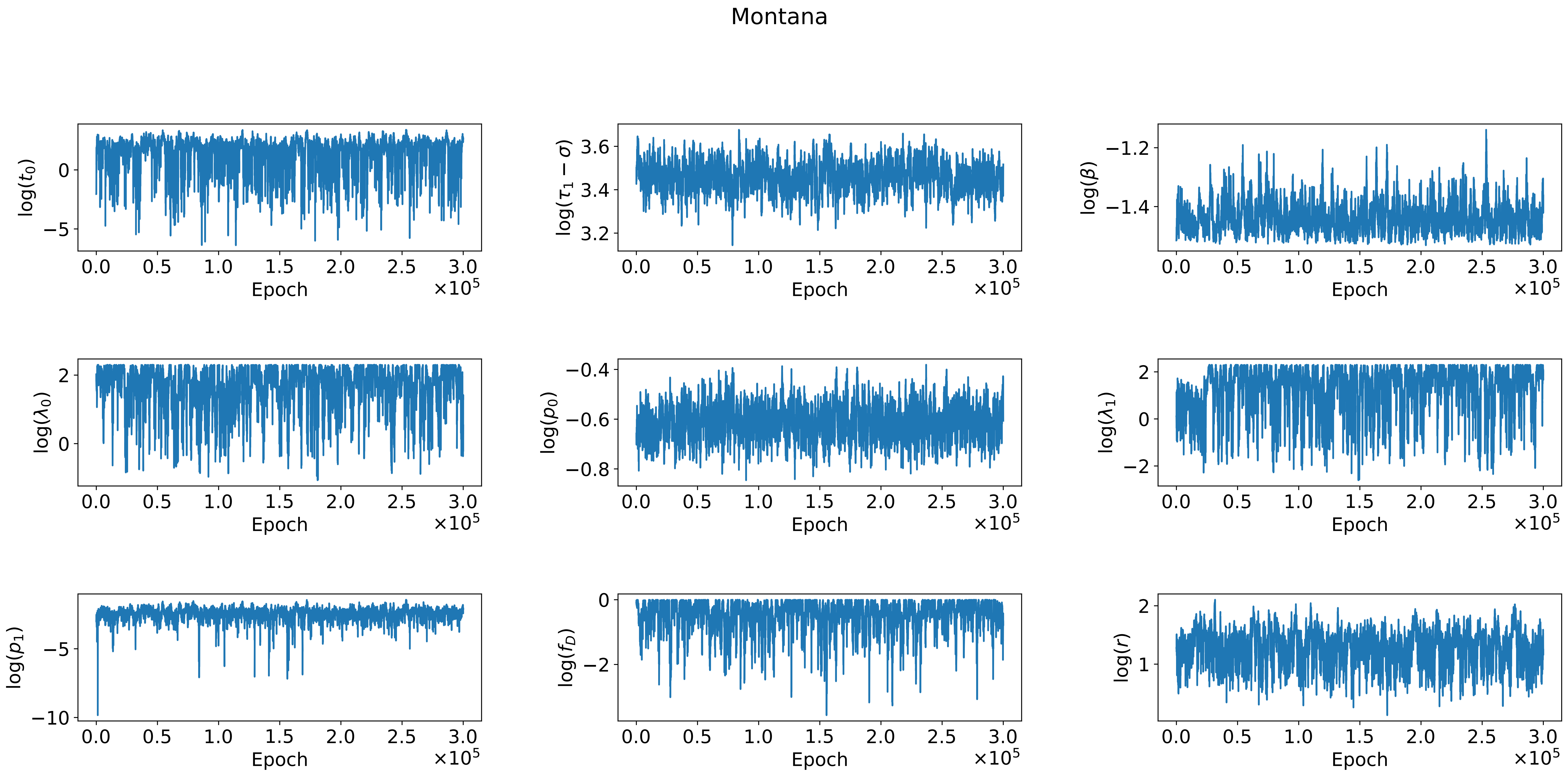

Montana
log($t_0$)
log($\tau_1 - \sigma$)
log($\beta$)
log($\lambda_0$)
log($p_0$)
log($\lambda_1$)
log($p_1$)
log($f_D$)
log($r$)
Epoch
$\times 10^5$

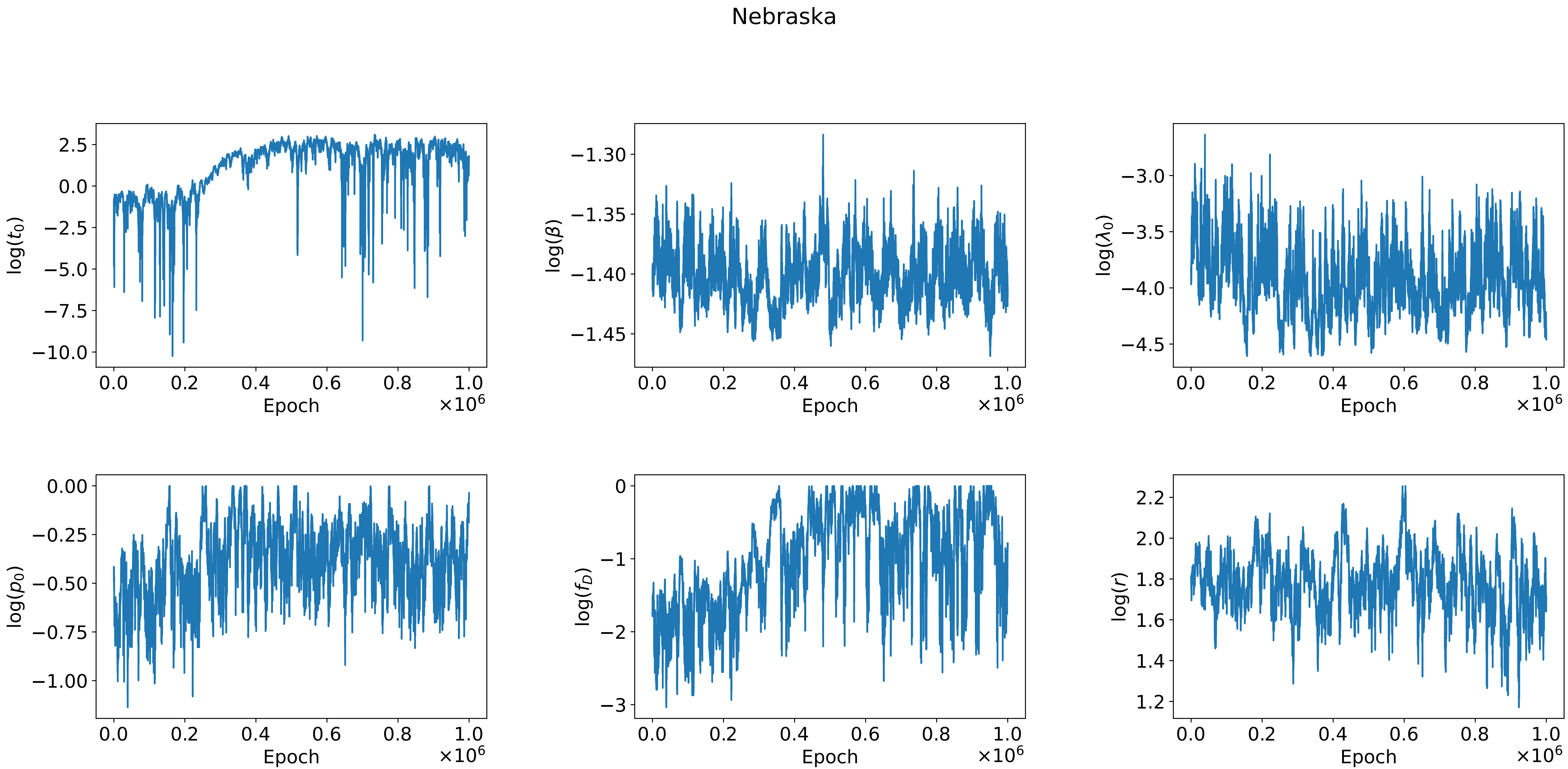

Nebraska
log($t_0$)
log($\beta$)
log($\lambda_0$)
log($p_0$)
log($f_D$)
log($r$)
Epoch
$\times 10^6$

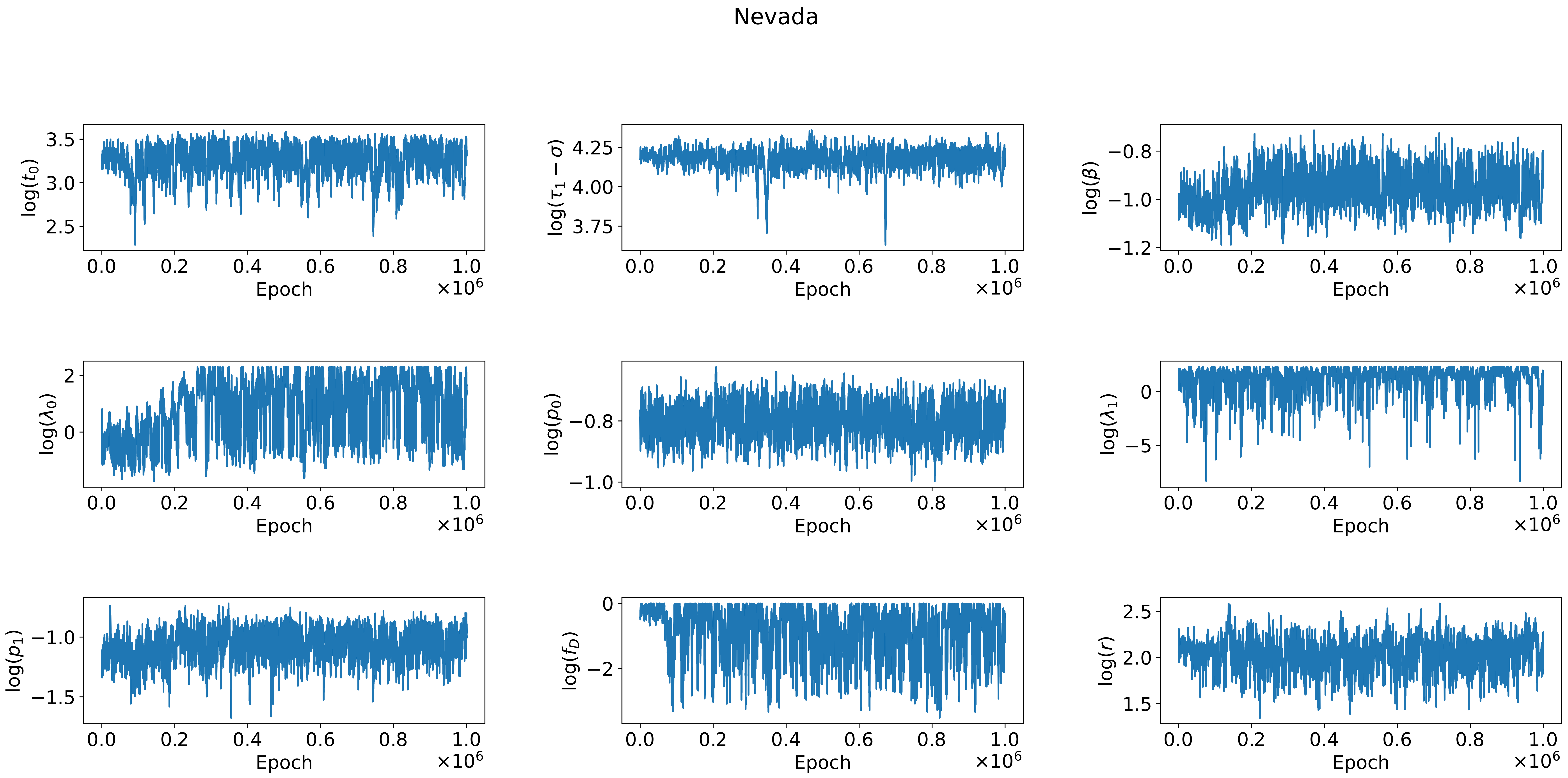
Nevada
log($t_0$)
log($\tau_1 - \sigma$)
log($\beta$)
log($\lambda_0$)
log($p_0$)
log($\lambda_1$)
log($p_1$)
log($f_D$)
log($r$)
Epoch
$\times 10^6$

New Hampshire

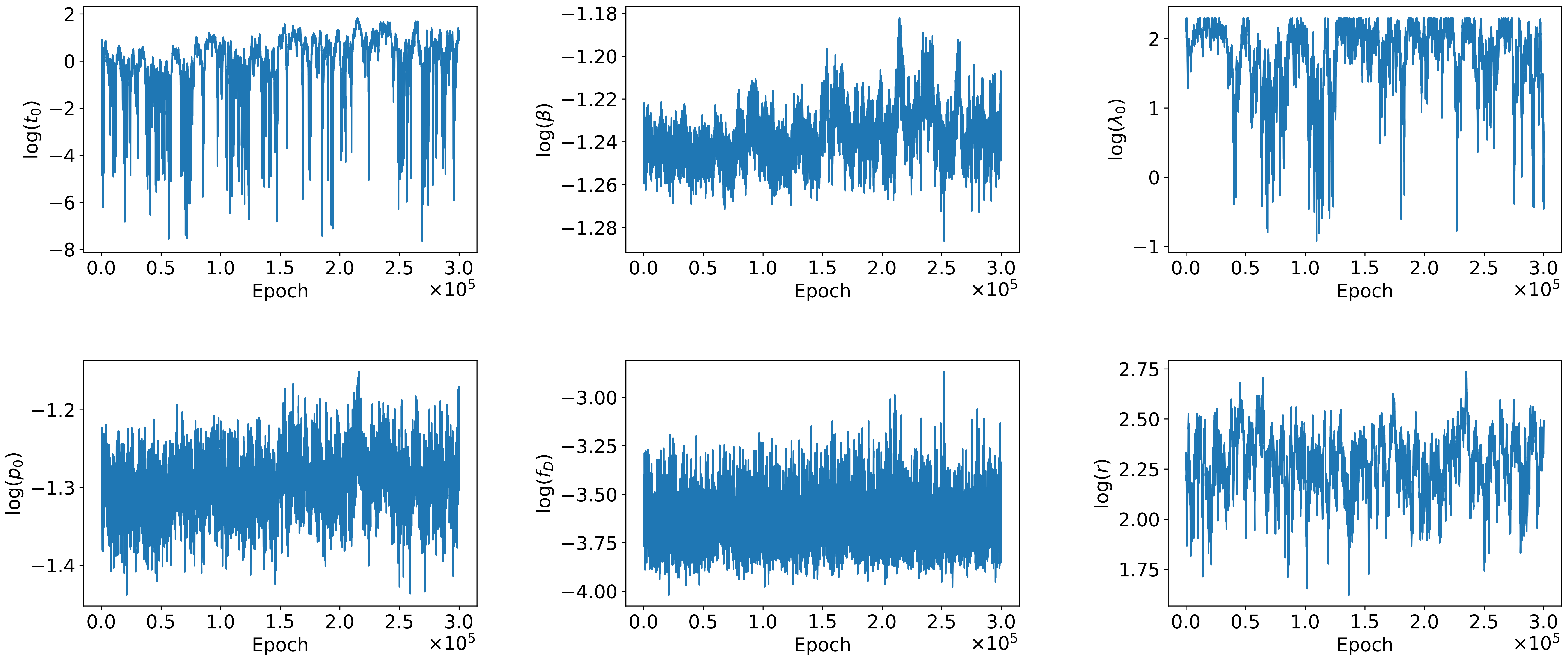

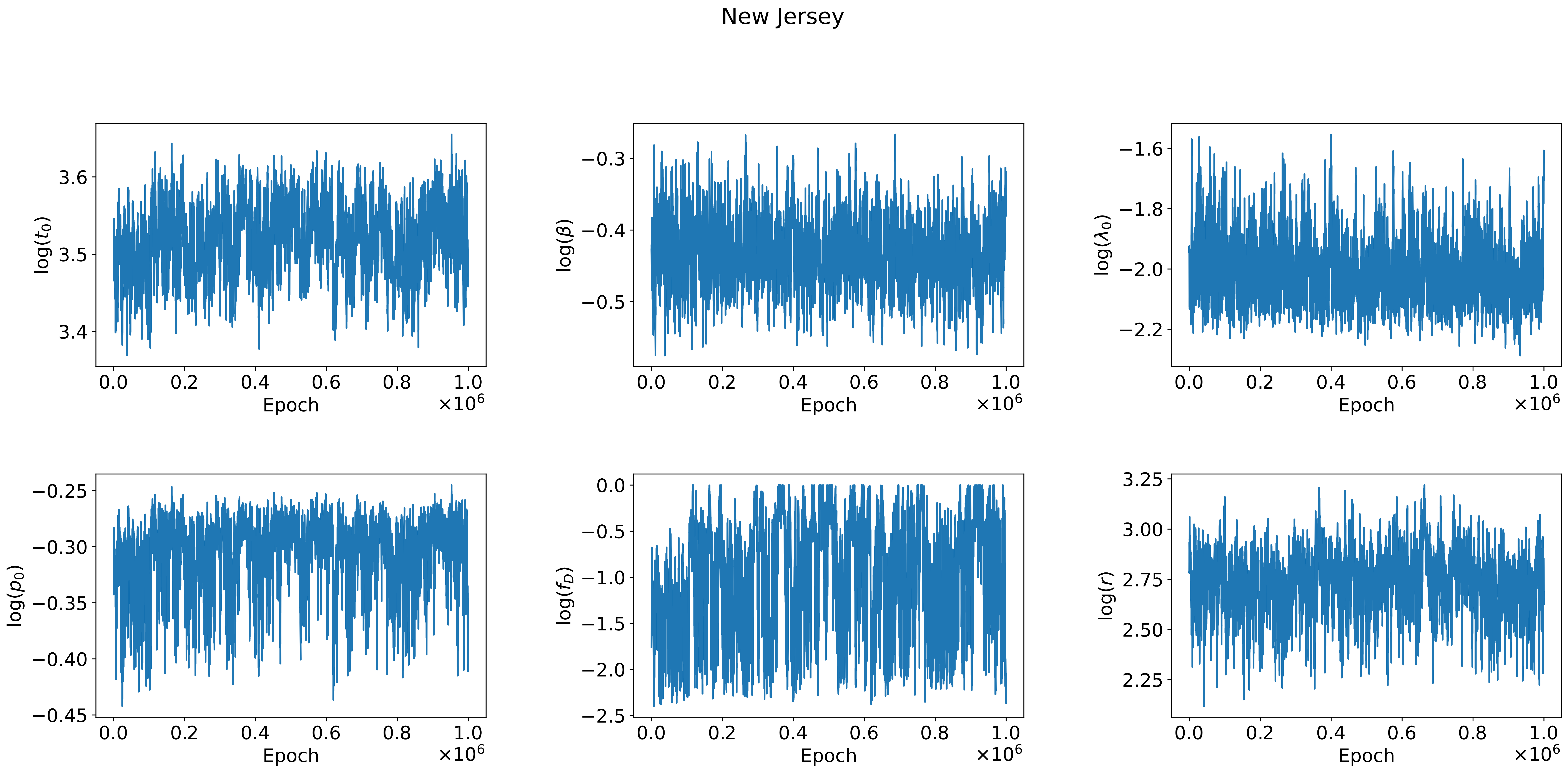
New Jersey
log($t_0$)
log($\beta$)
log($\lambda_0$)
log($p_0$)
log($f_D$)
log($r$)
Epoch
$\times 10^6$
3.6
3.5
3.4
−0.3
−0.4
−0.5
−1.6
−1.8
−2.0
−2.2
−0.25
−0.30
−0.35
−0.40
−0.45
0.0
−0.5
−1.0
−1.5
−2.0
−2.5
3.25
3.00
2.75
2.50
2.25
0.0
0.2
0.4
0.6
0.8
1.0

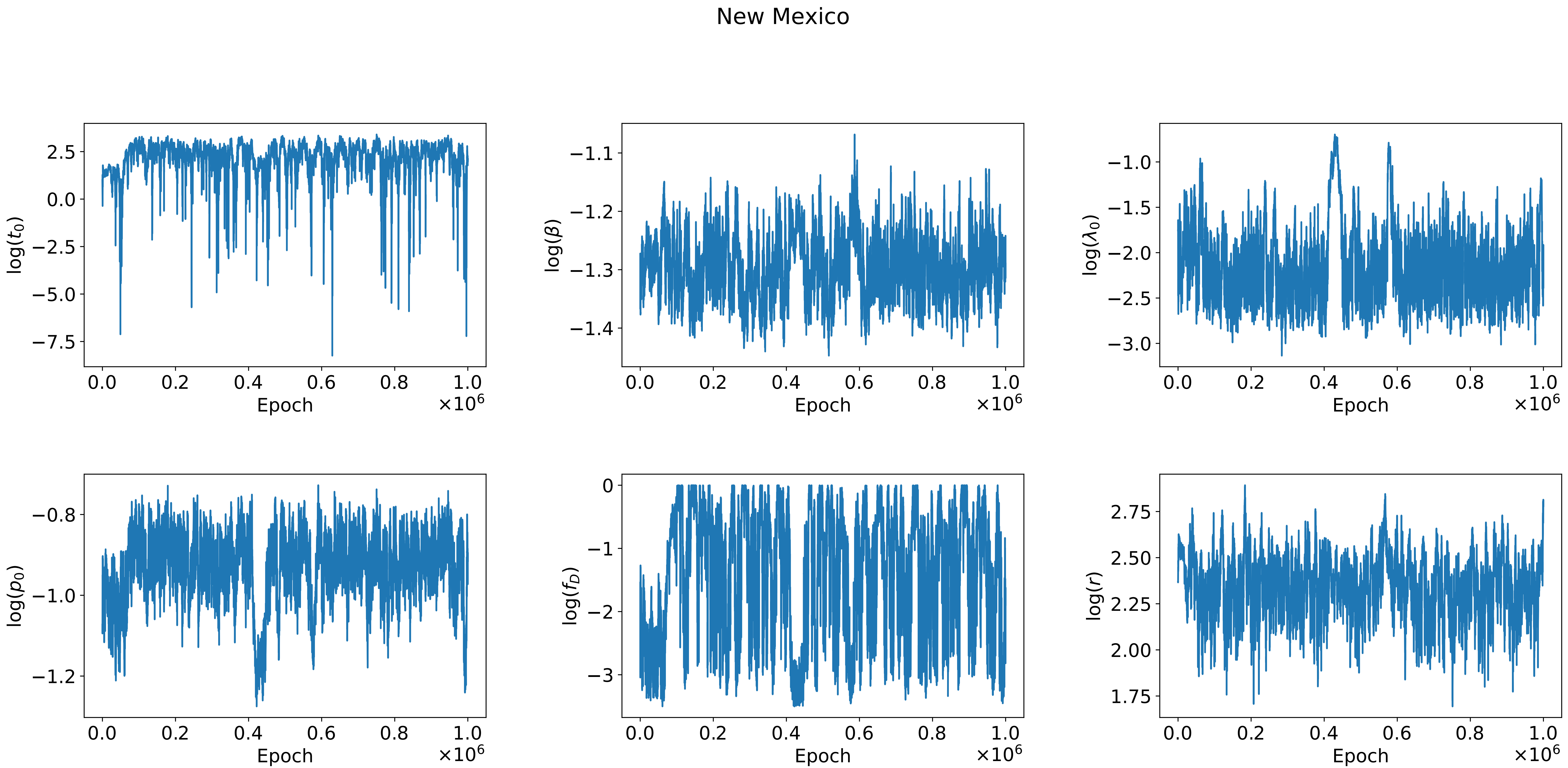
New Mexico
log($t_0$)
log($\beta$)
log($\lambda_0$)
log($p_0$)
log($f_D$)
log($r$)
Epoch
$\times 10^6$

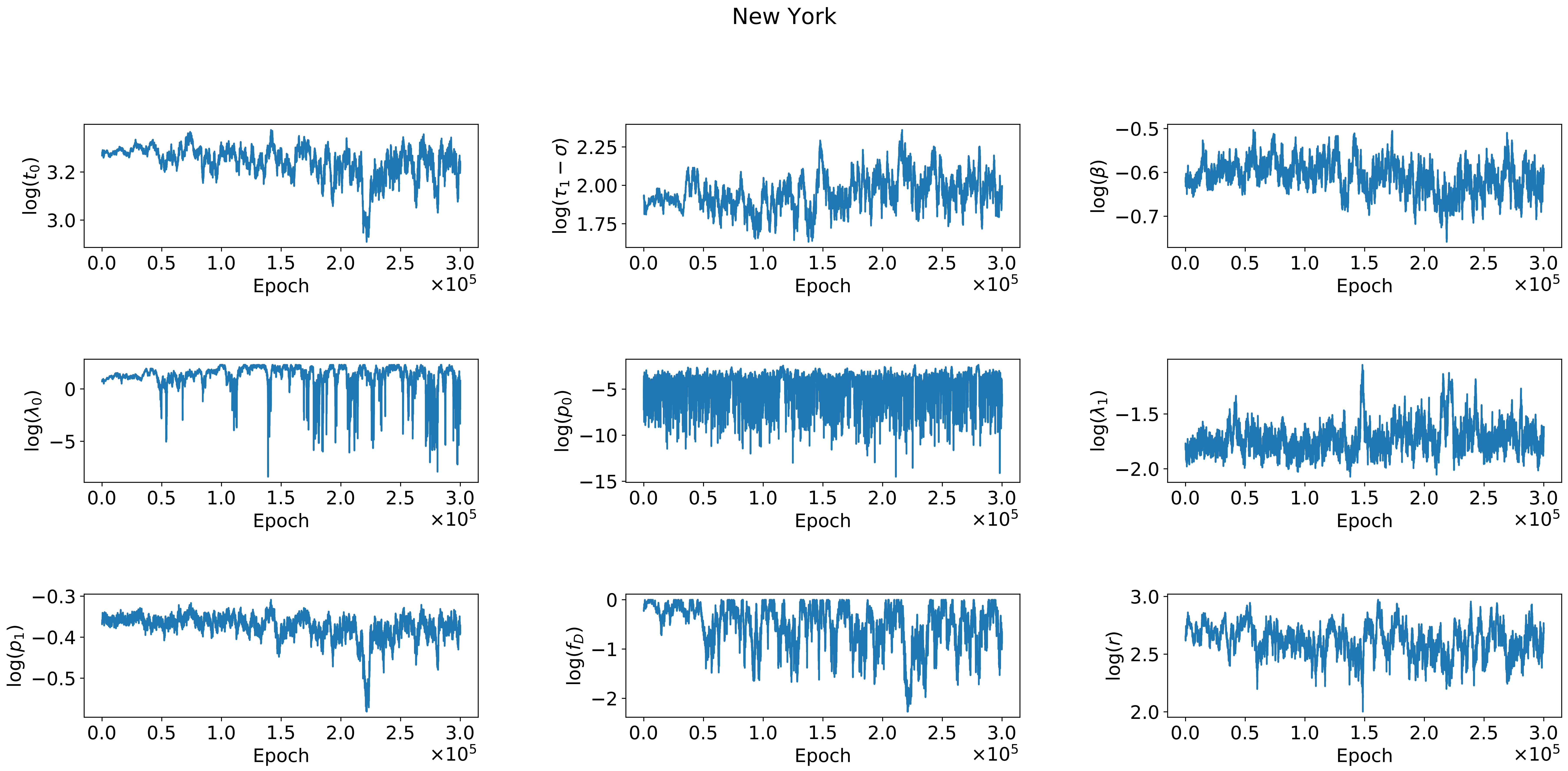
New York
log($t_0$)
log($\tau_1 - \sigma$)
log($\beta$)
log($\lambda_0$)
log($p_0$)
log($\lambda_1$)
log($p_1$)
log($f_D$)
log($r$)
Epoch
$\times 10^5$

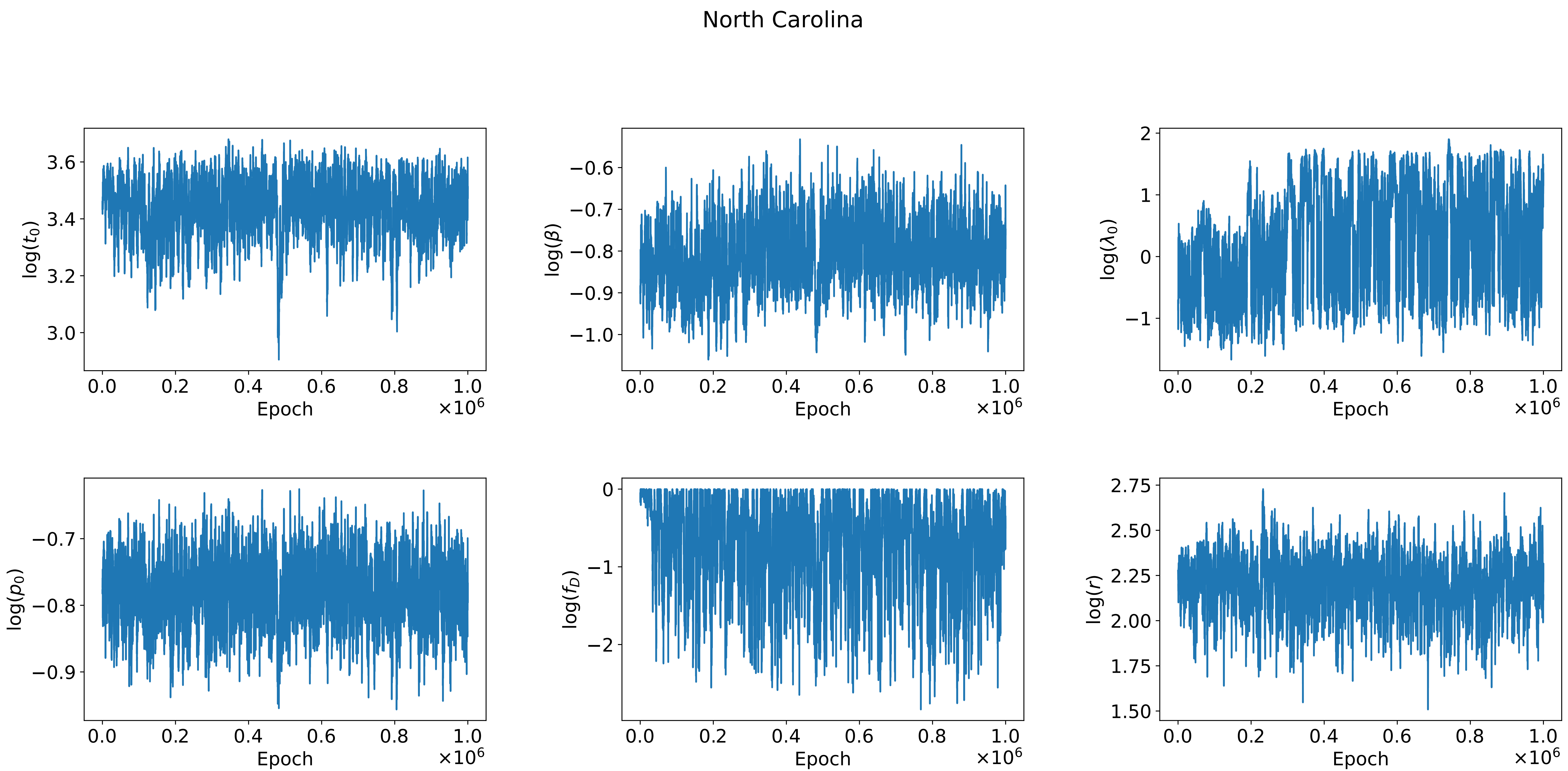
North Carolina
log($t_0$)
3.6
3.4
3.2
3.0
0.0
0.2
0.4
0.6
0.8
1.0
Epoch
$\times 10^6$
log($\beta$)
−0.6
−0.7
−0.8
−0.9
−1.0
0.0
0.2
0.4
0.6
0.8
1.0
Epoch
$\times 10^6$
log($\lambda_0$)
2
1
0
−1
0.0
0.2
0.4
0.6
0.8
1.0
Epoch
$\times 10^6$
log($p_0$)
−0.7
−0.8
−0.9
0.0
0.2
0.4
0.6
0.8
1.0
Epoch
$\times 10^6$
log($f_D$)
0
−1
−2
0.0
0.2
0.4
0.6
0.8
1.0
Epoch
$\times 10^6$
log($r$)
2.75
2.50
2.25
2.00
1.75
1.50
0.0
0.2
0.4
0.6
0.8
1.0
Epoch
$\times 10^6$

North Dakota

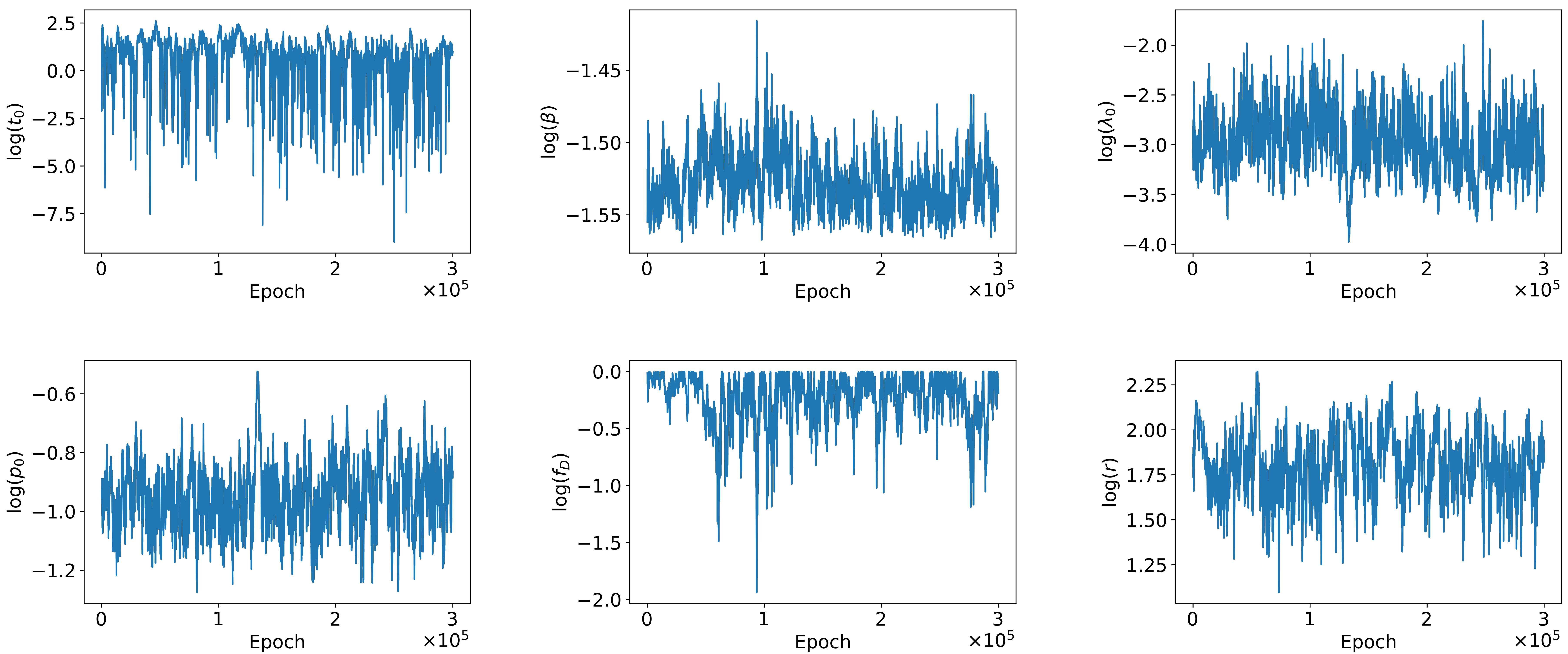

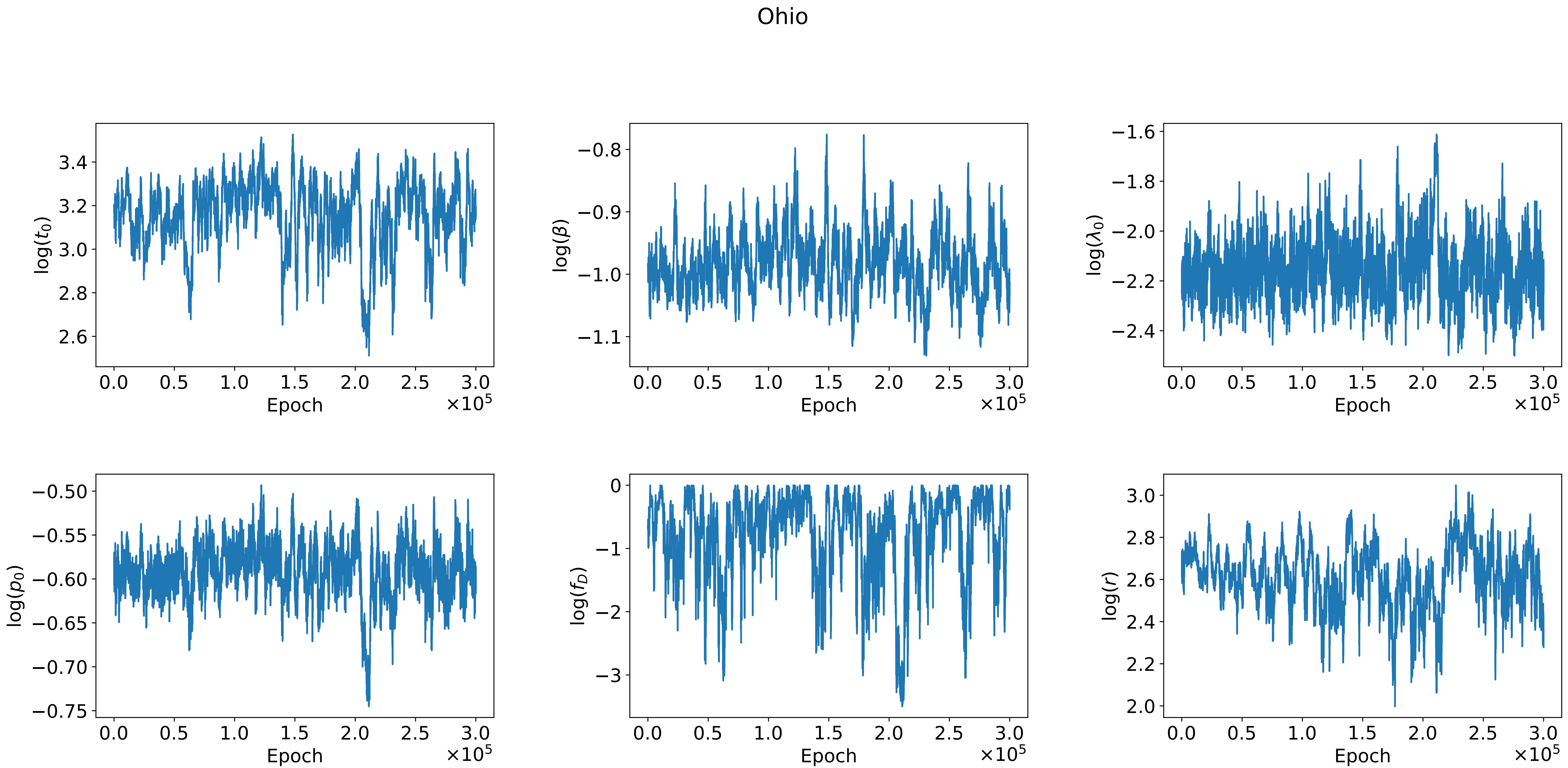
Ohio
log($t_0$)
log($\beta$)
log($\lambda_0$)
log($p_0$)
log($f_D$)
log($r$)
Epoch
$\times 10^5$

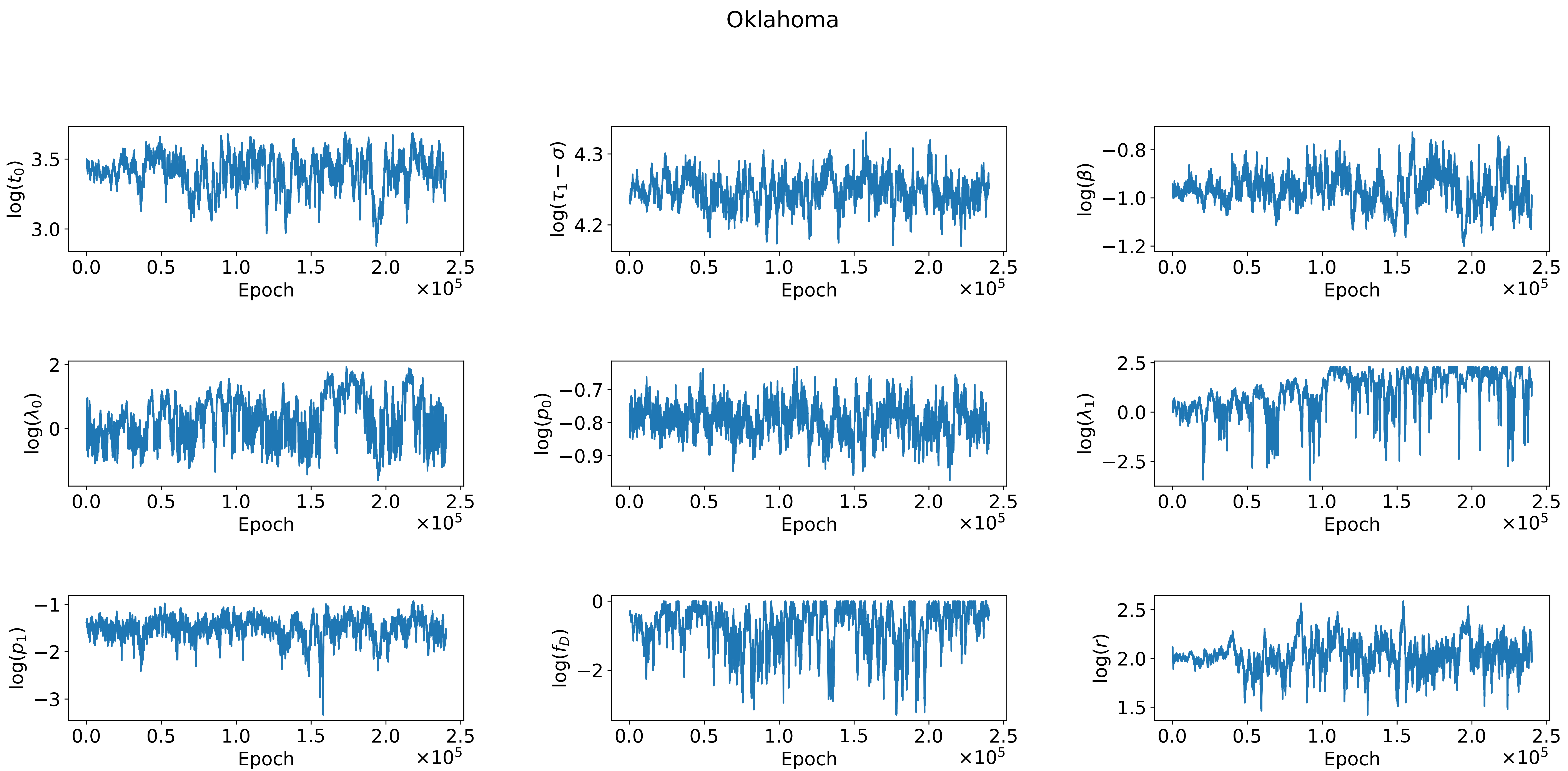
Oklahoma
log($t_0$)
log($\tau_1 - \sigma$)
log($\beta$)
log($\lambda_0$)
log($p_0$)
log($\lambda_1$)
log($p_1$)
log($f_D$)
log($r$)
Epoch
$\times 10^5$

Oregon

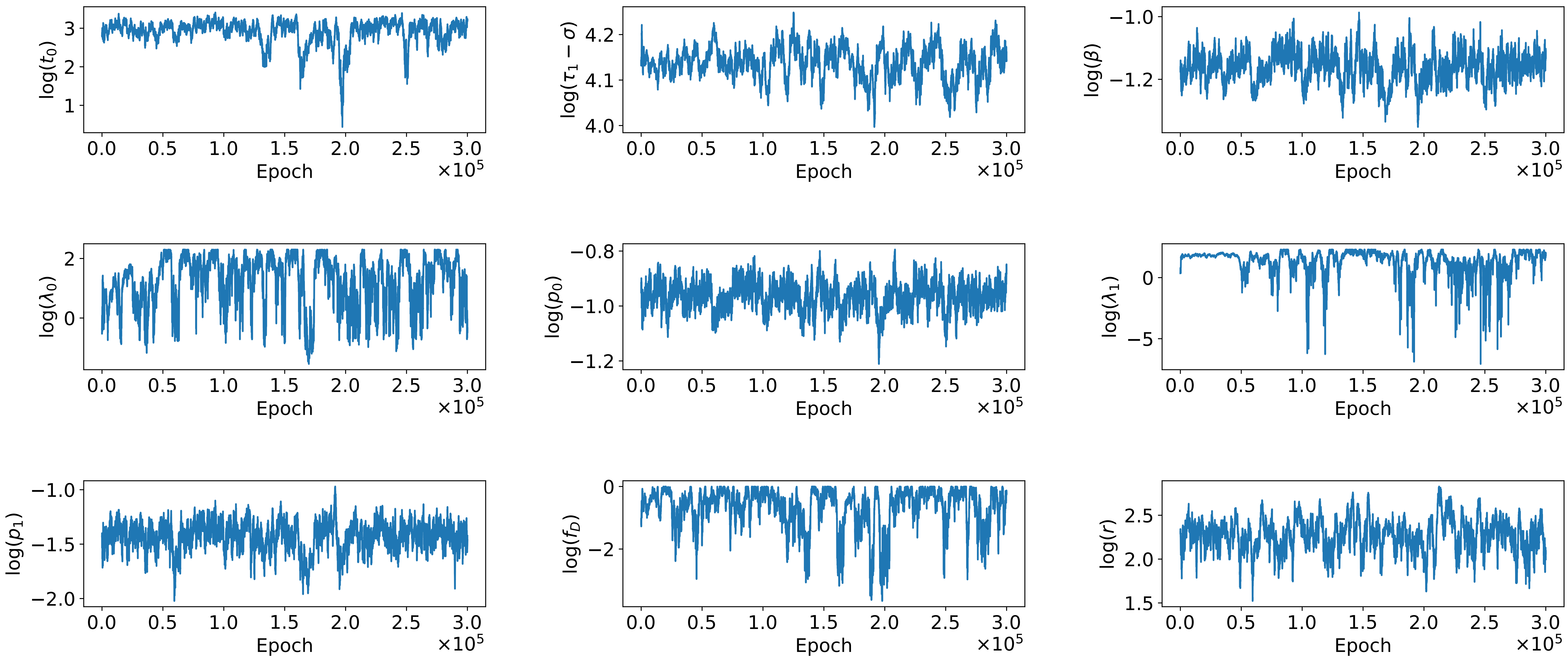

log($t_0$)
log($\tau_1 - \sigma$)
log($\beta$)
log($\lambda_0$)
log($p_0$)
log($\lambda_1$)
log($p_1$)
log($f_D$)
log($r$)
Epoch
×10^5

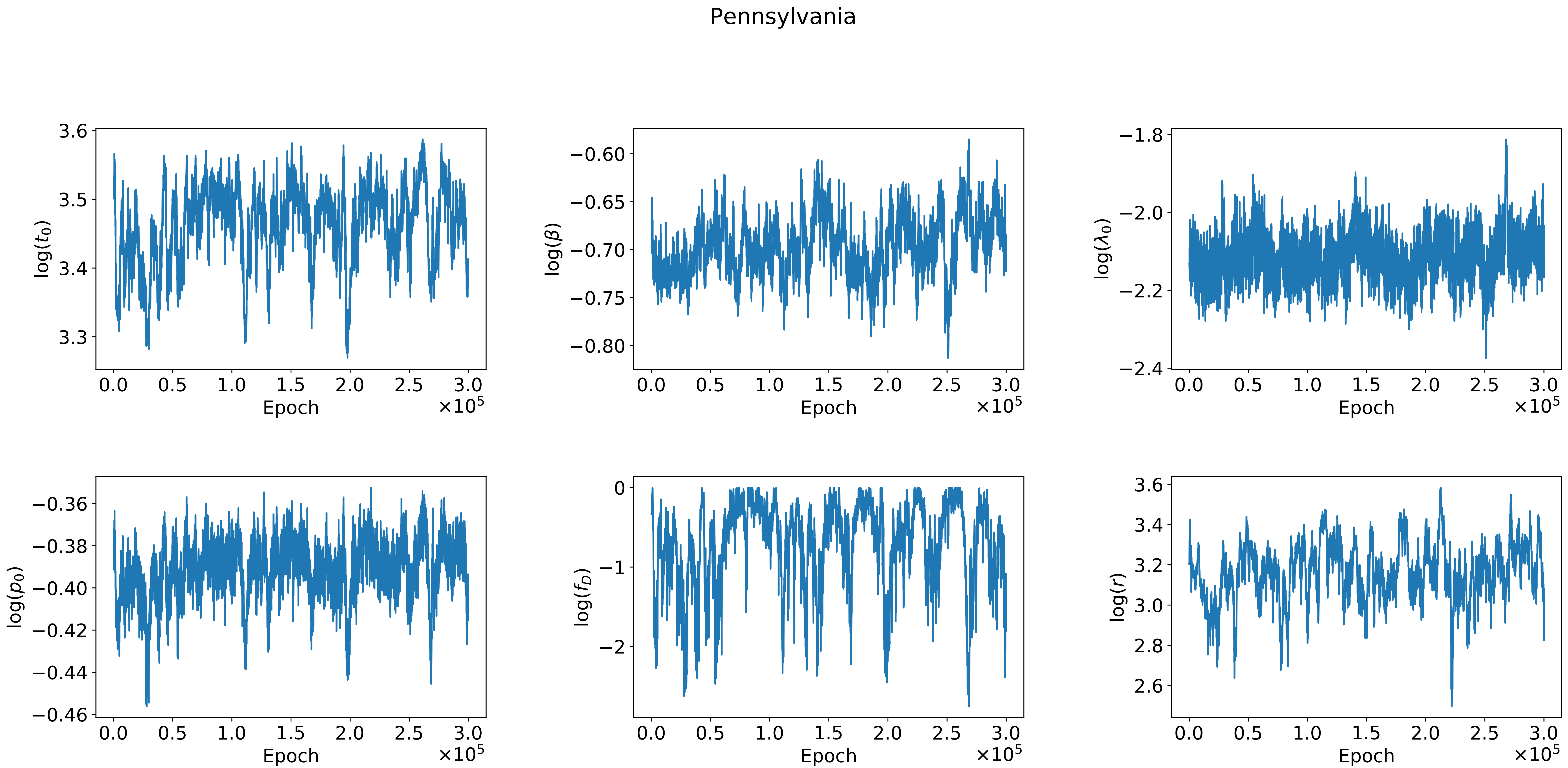

Pennsylvania
log($t_0$)
log($\beta$)
log($\lambda_0$)
log($p_0$)
log($f_D$)
log($r$)
Epoch
$\times 10^5$

Rhode Island

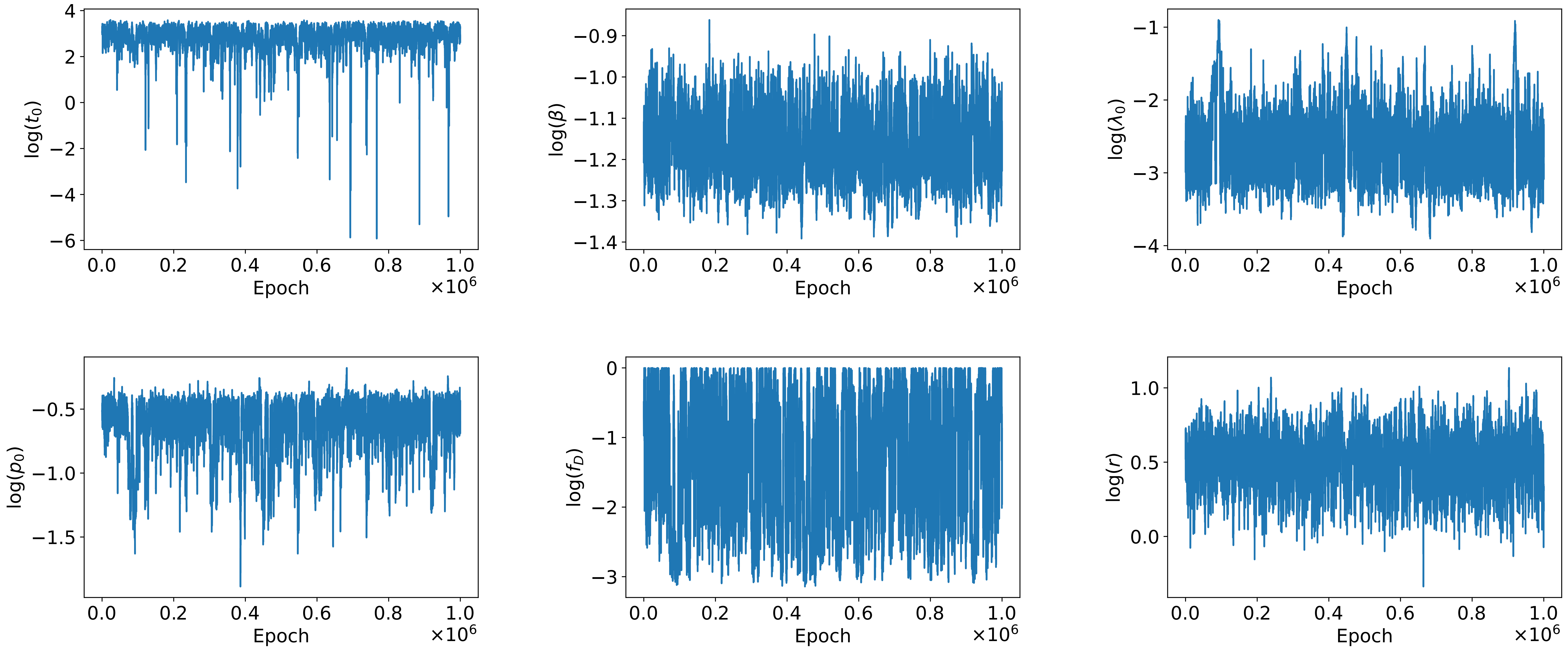

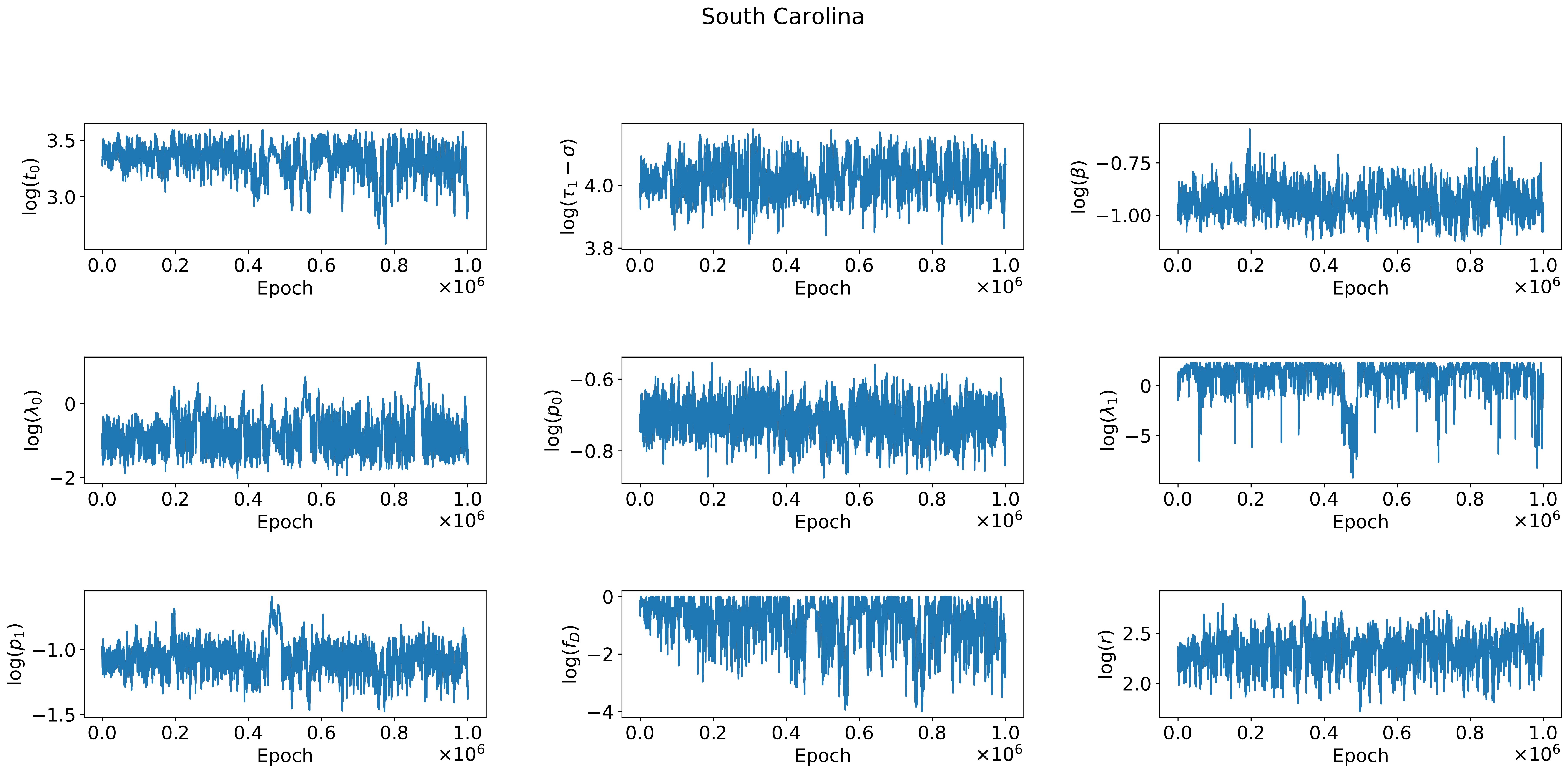

South Carolina
$\log(t_0)$
$\log(\tau_1 - \sigma)$
$\log(\beta)$
$\log(\lambda_0)$
$\log(p_0)$
$\log(\lambda_1)$
$\log(p_1)$
$\log(f_D)$
$\log(r)$
Epoch
$\times 10^6$

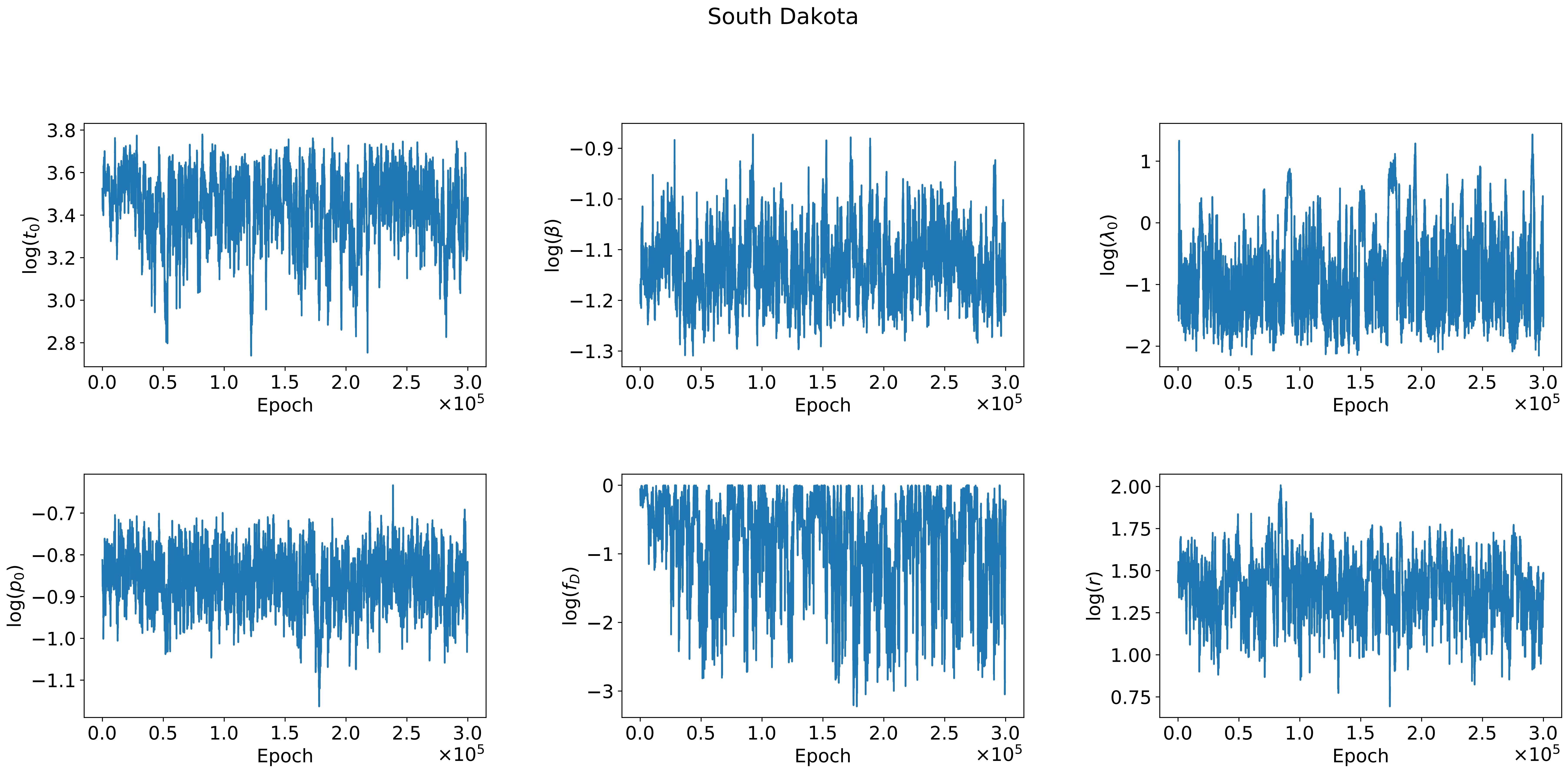

South Dakota
log($t_0$)
log($\beta$)
log($\lambda_0$)
log($p_0$)
log($f_D$)
log($r$)
Epoch
$\times 10^5$

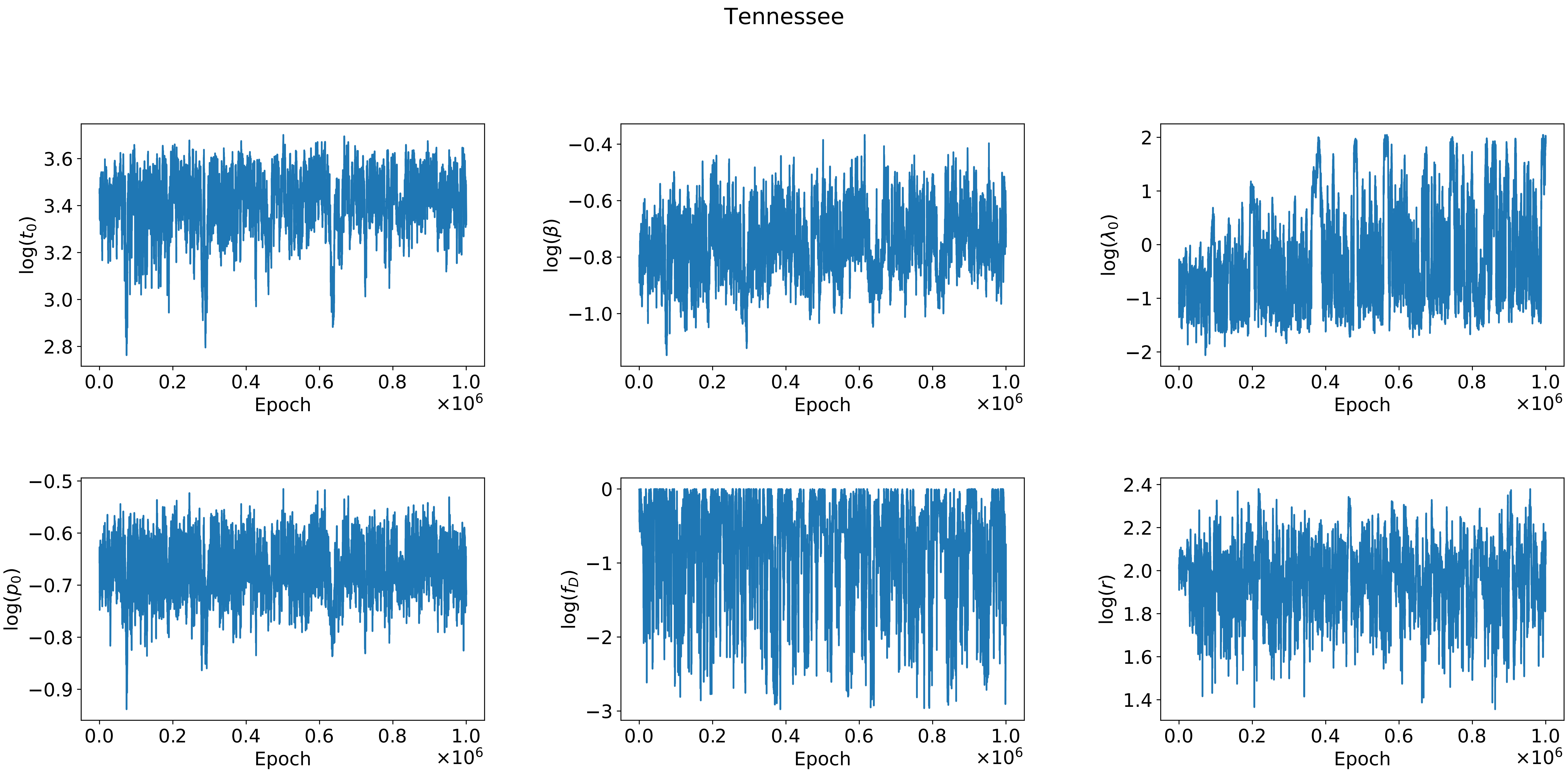

Tennessee
log($t_0$)
log($\beta$)
log($\lambda_0$)
log($p_0$)
log($f_D$)
log($r$)
Epoch
$\times 10^6$

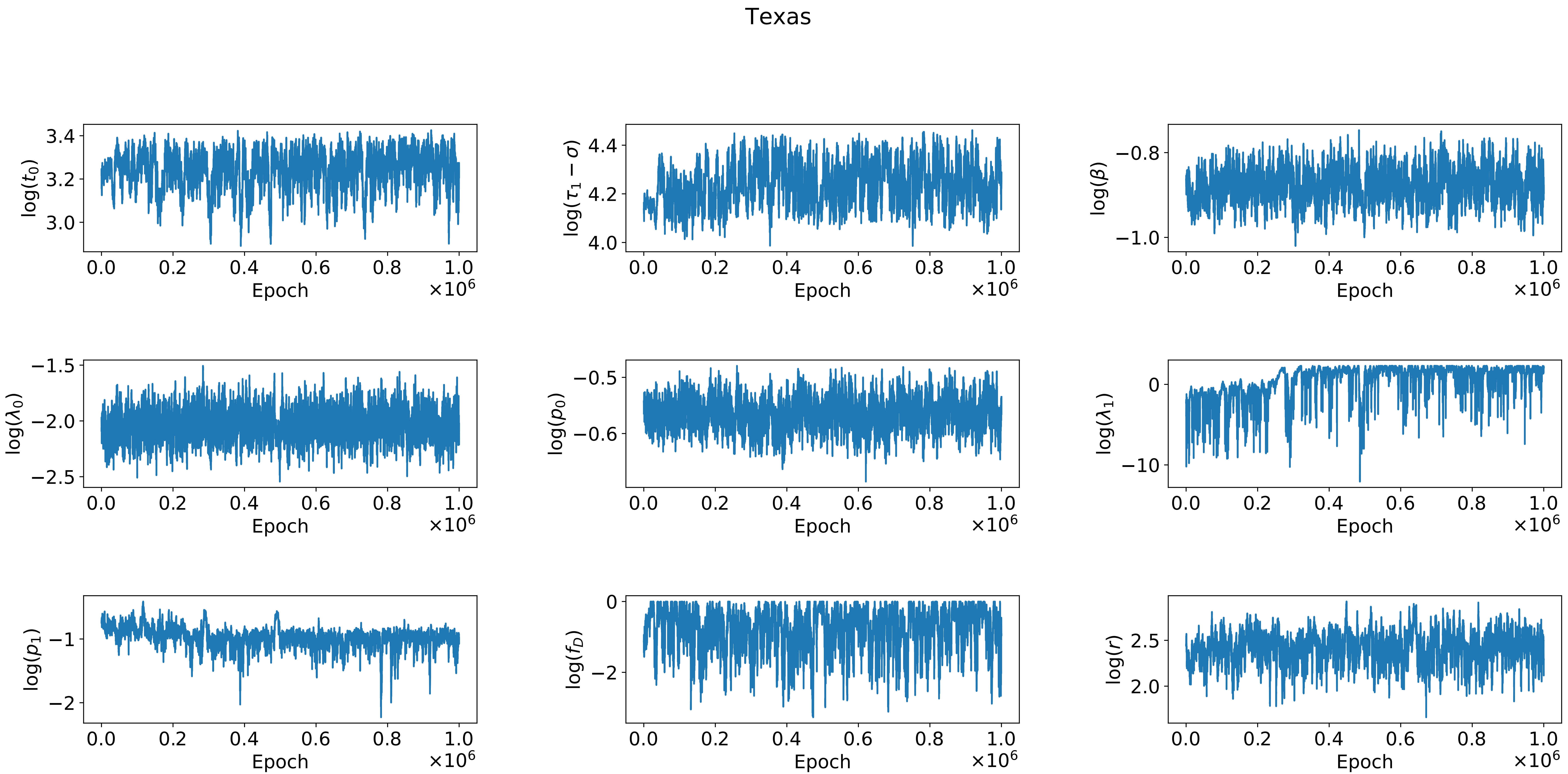

Texas
$\log(t_0)$
$\log(\tau_1 - \sigma)$
$\log(\beta)$
$\log(\lambda_0)$
$\log(p_0)$
$\log(\lambda_1)$
$\log(p_1)$
$\log(f_D)$
$\log(r)$
Epoch
$\times 10^6$

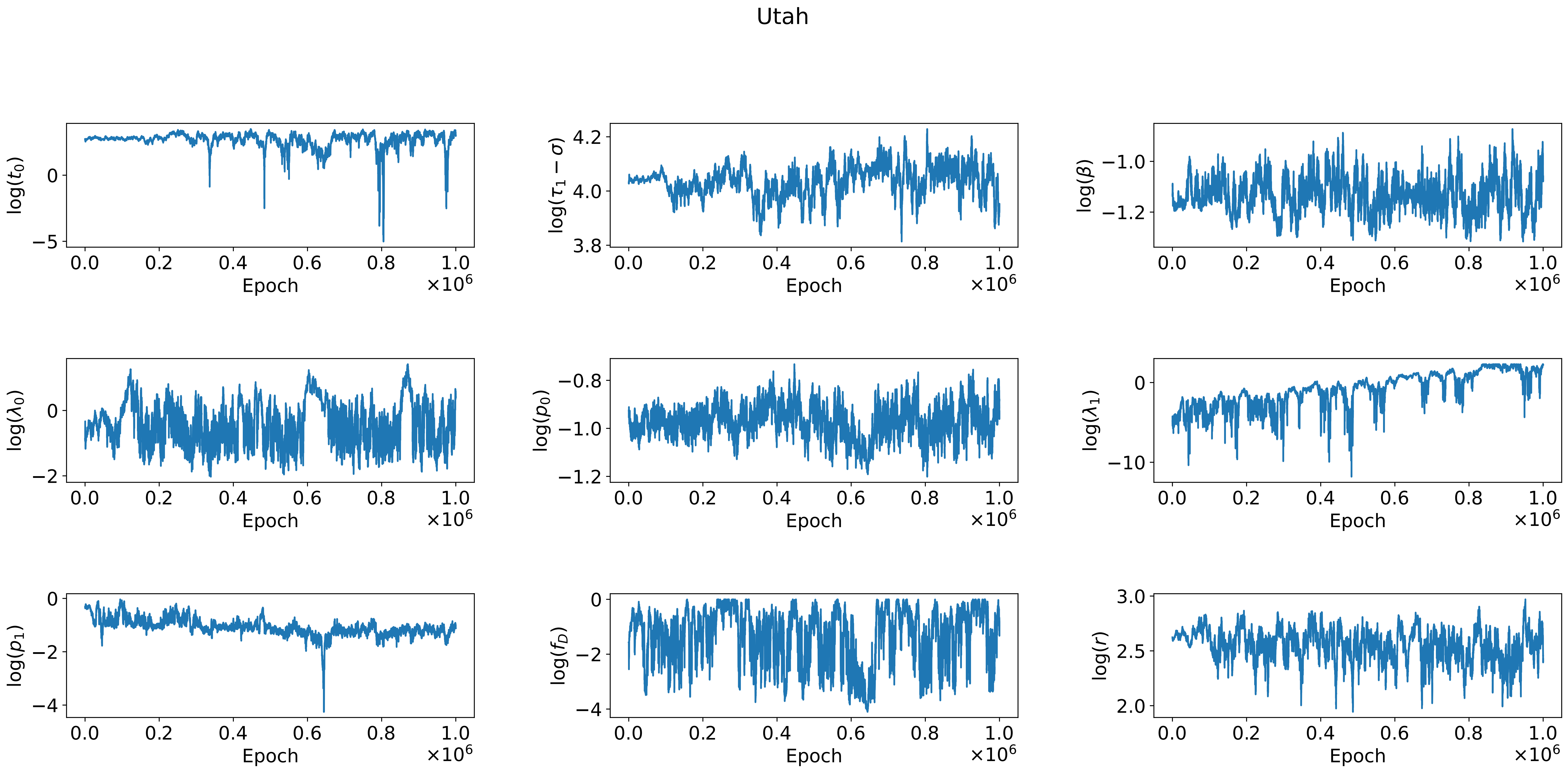

Utah
log(t0)
log(τ1 − σ)
log(β)
log(λ0)
log(p0)
log(λ1)
log(p1)
log(fD)
log(r)
Epoch
×10^6

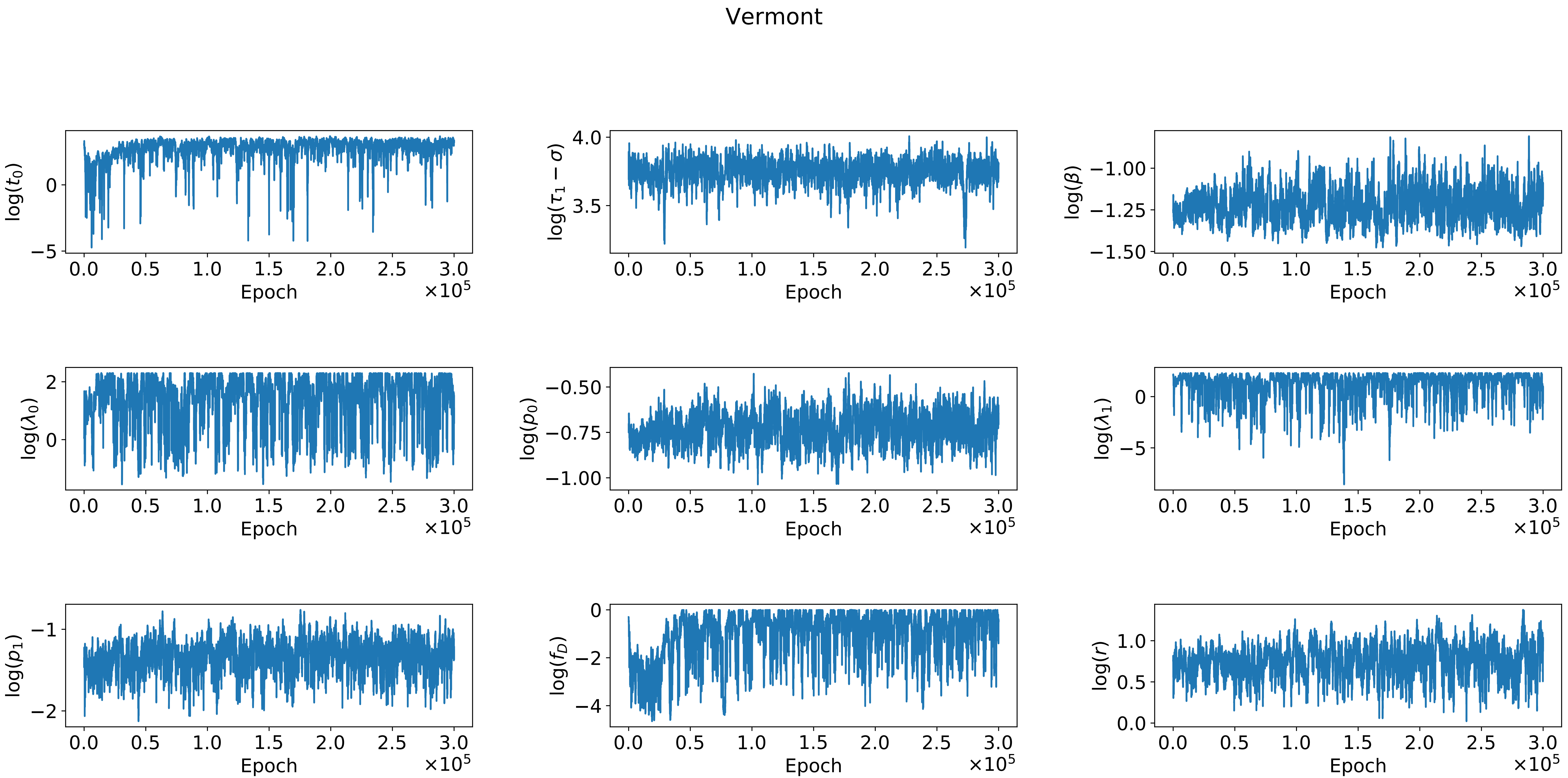
Vermont
log($t_0$)
log($\tau_1 - \sigma$)
log($\beta$)
log($\lambda_0$)
log($p_0$)
log($\lambda_1$)
log($p_1$)
log($f_D$)
log($r$)
Epoch
$\times 10^5$

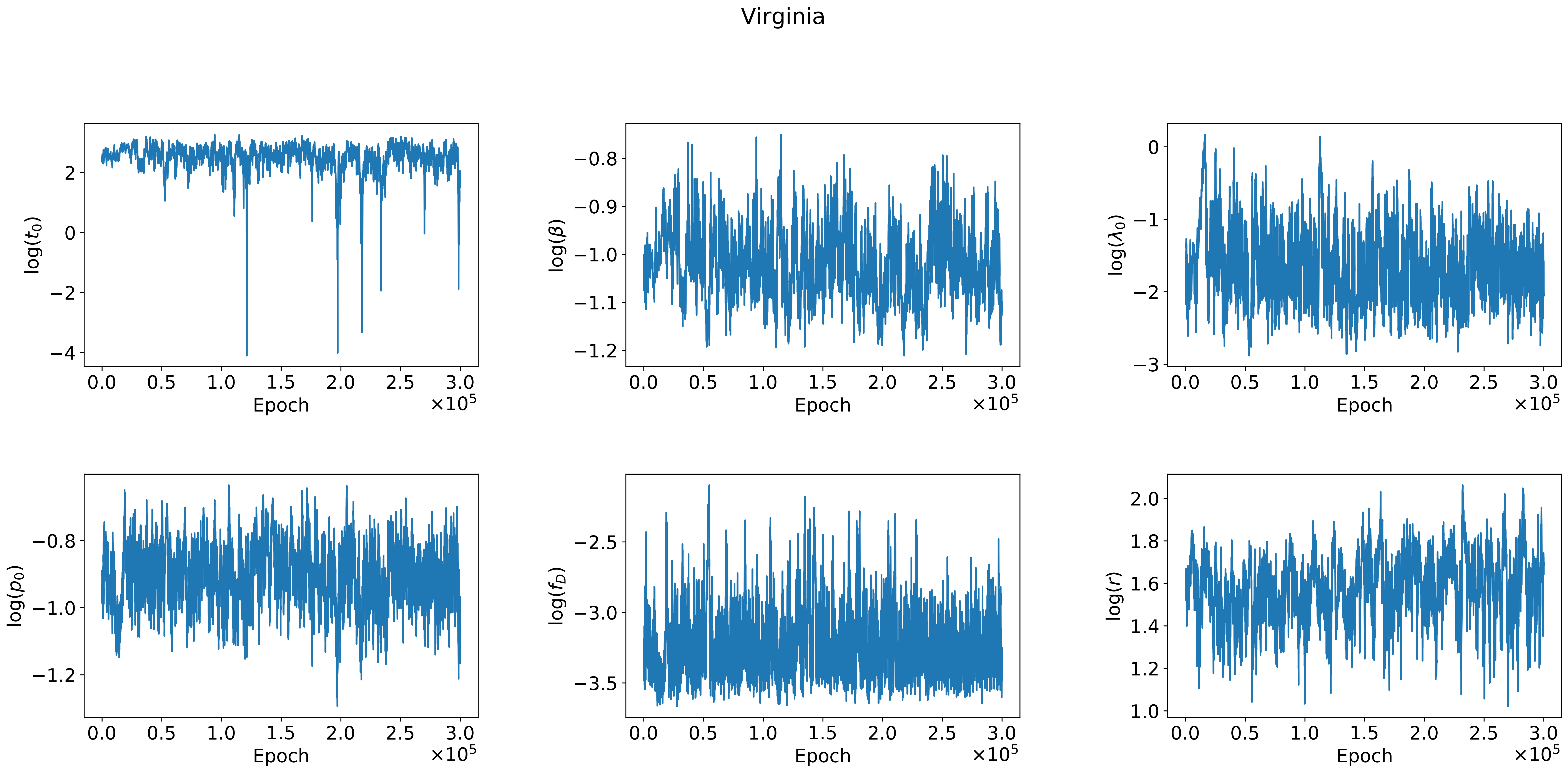
Virginia
log($t_0$)
log($\beta$)
log($\lambda_0$)
log($p_0$)
log($f_D$)
log($r$)
Epoch
$\times 10^5$

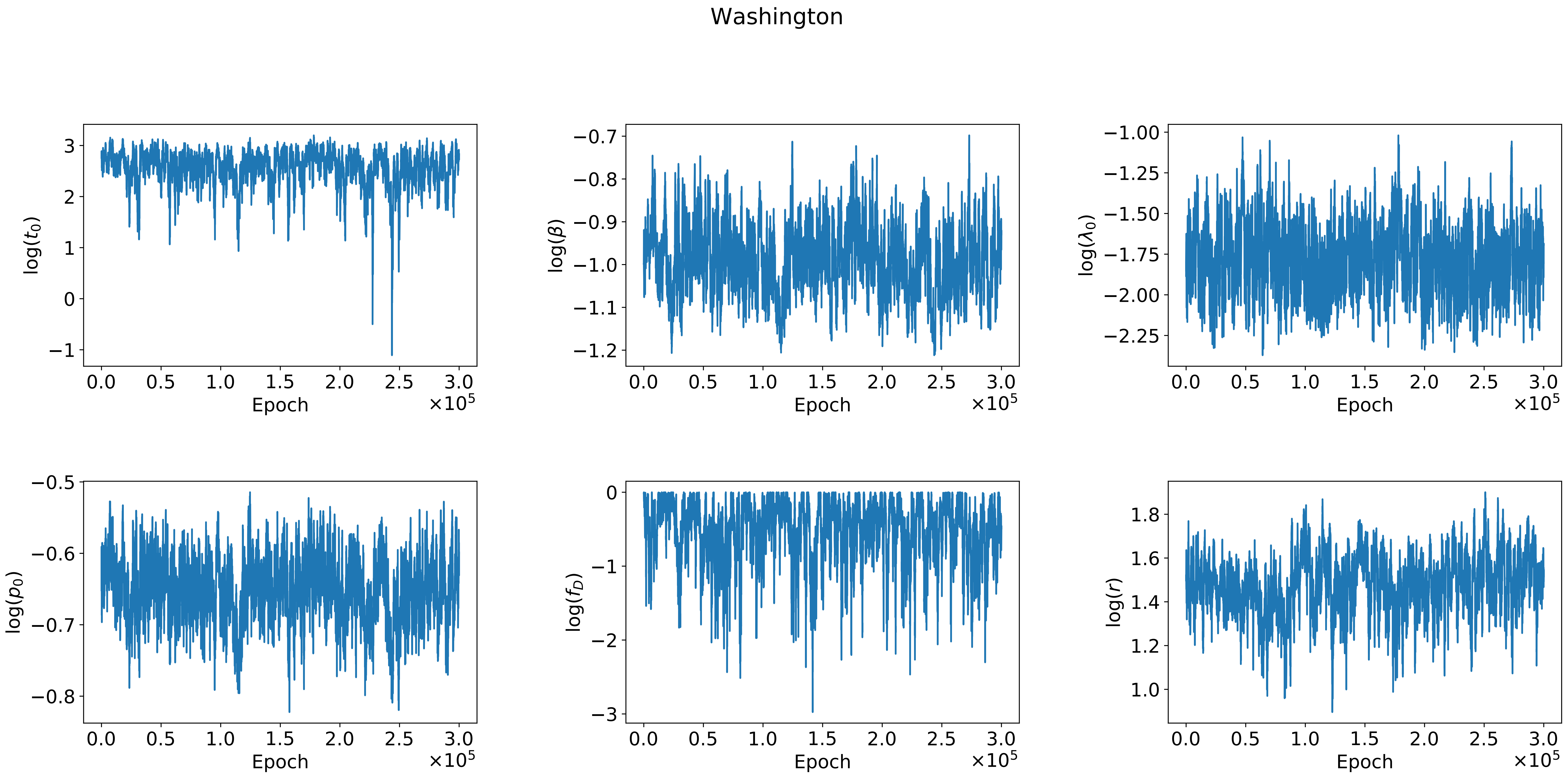
Washington
log($t_0$)
log($\beta$)
log($\lambda_0$)
log($p_0$)
log($f_D$)
log($r$)
Epoch
×10^5

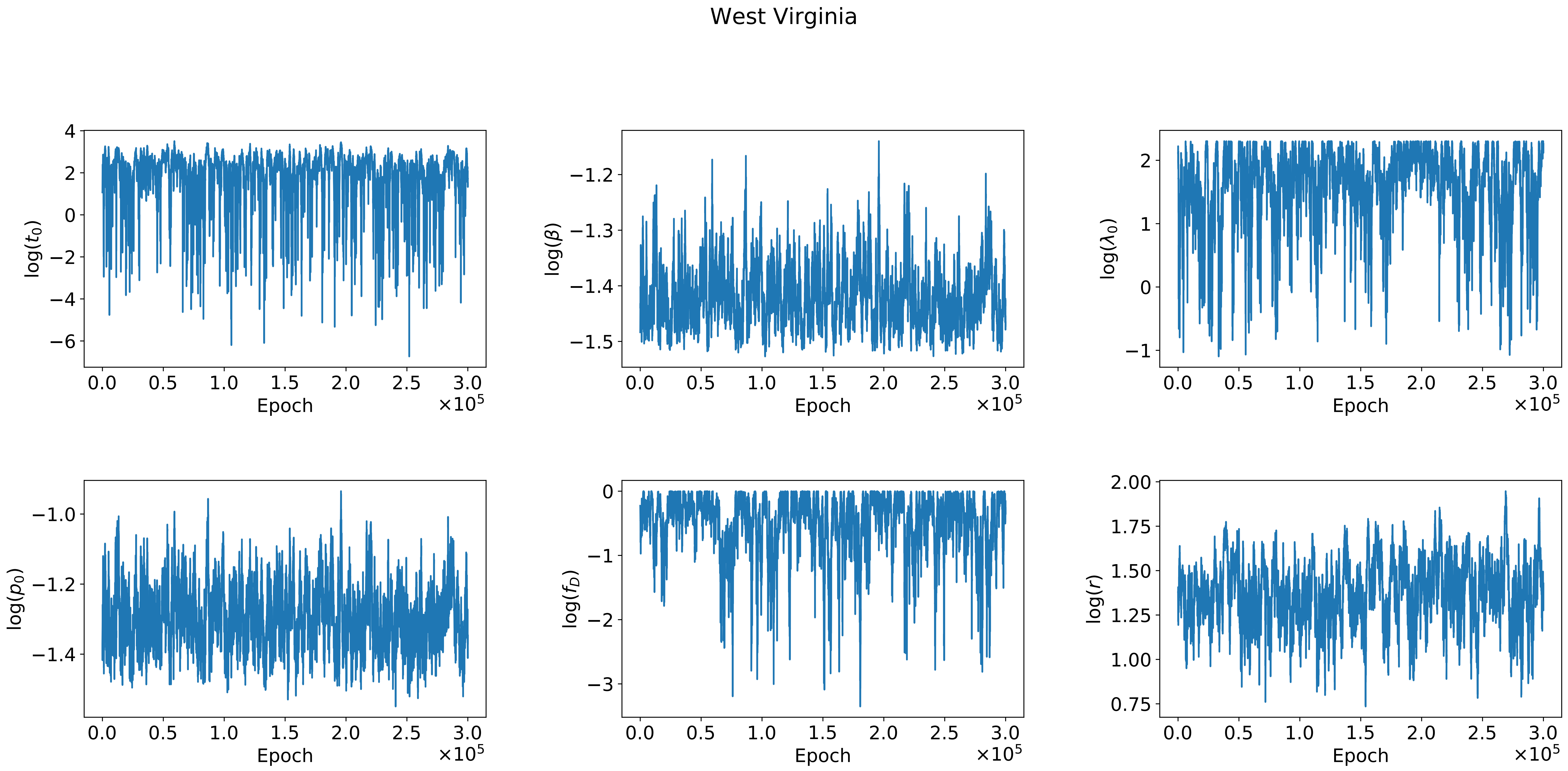

West Virginia
log($t_0$)
log($\beta$)
log($\lambda_0$)
log($p_0$)
log($f_D$)
log($r$)
Epoch
$\times 10^5$

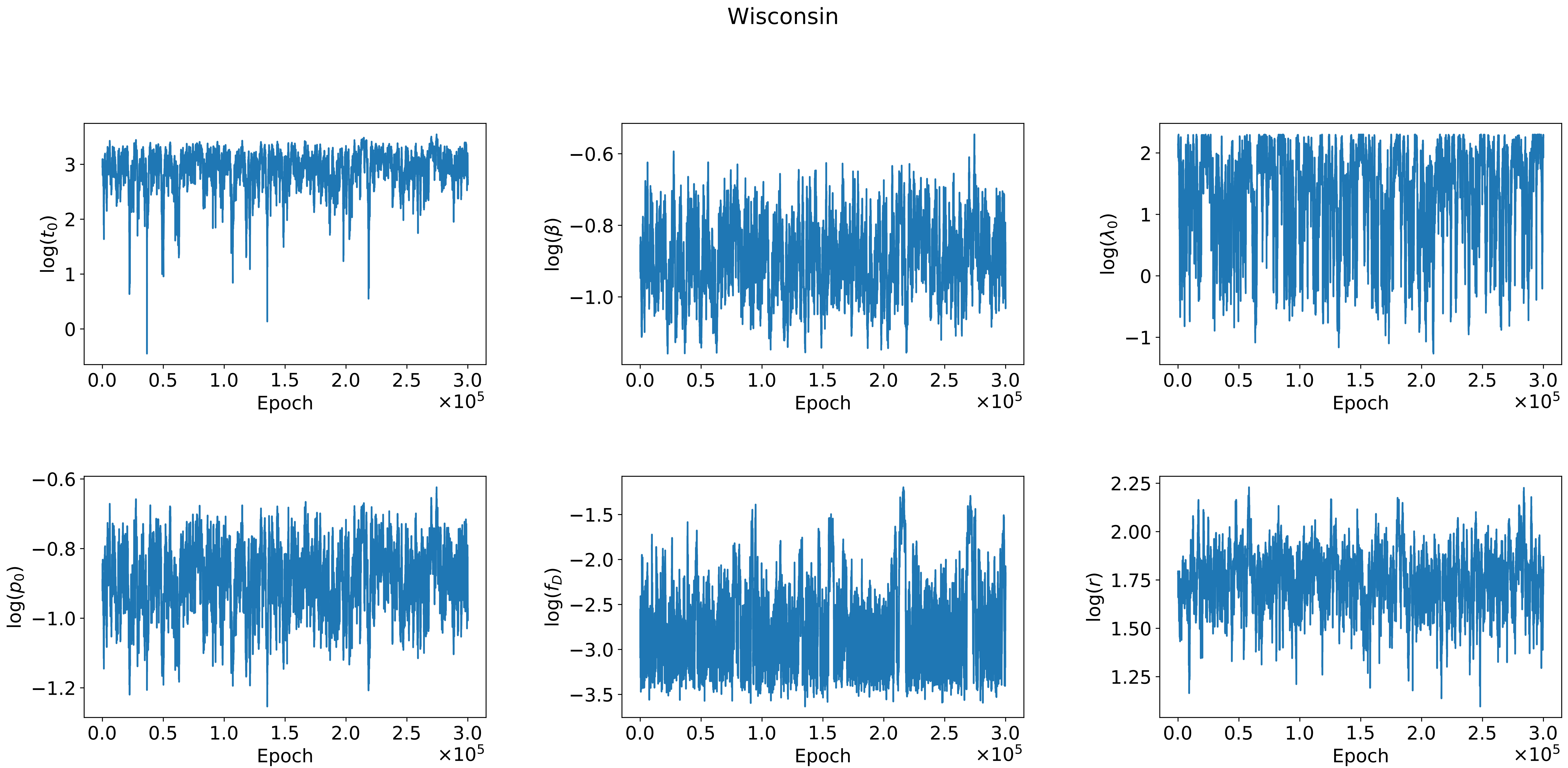

Wisconsin
log($t_0$)
log($\beta$)
log($\lambda_0$)
log($p_0$)
log($f_D$)
log($r$)
Epoch
$\times 10^5$

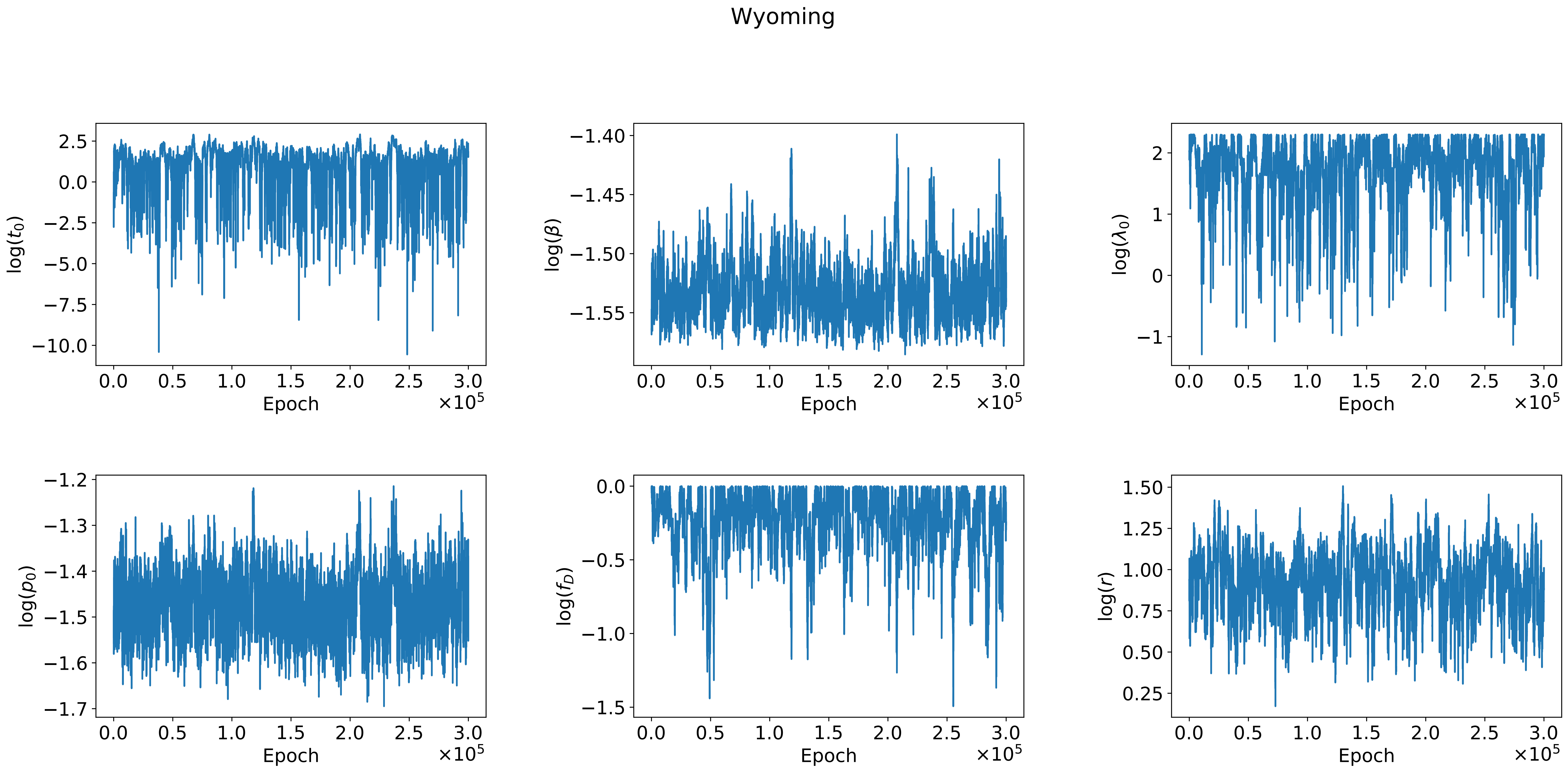

Wyoming
log($t_0$)
2.5
0.0
−2.5
−5.0
−7.5
−10.0
log($\beta$)
−1.40
−1.45
−1.50
−1.55
log($\lambda_0$)
2
1
0
−1
log($p_0$)
−1.2
−1.3
−1.4
−1.5
−1.6
−1.7
log($f_D$)
0.0
−0.5
−1.0
−1.5
log($r$)
1.50
1.25
1.00
0.75
0.50
0.25
0.0
0.5
1.0
1.5
2.0
2.5
3.0
Epoch
$\times 10^5$

Alabama

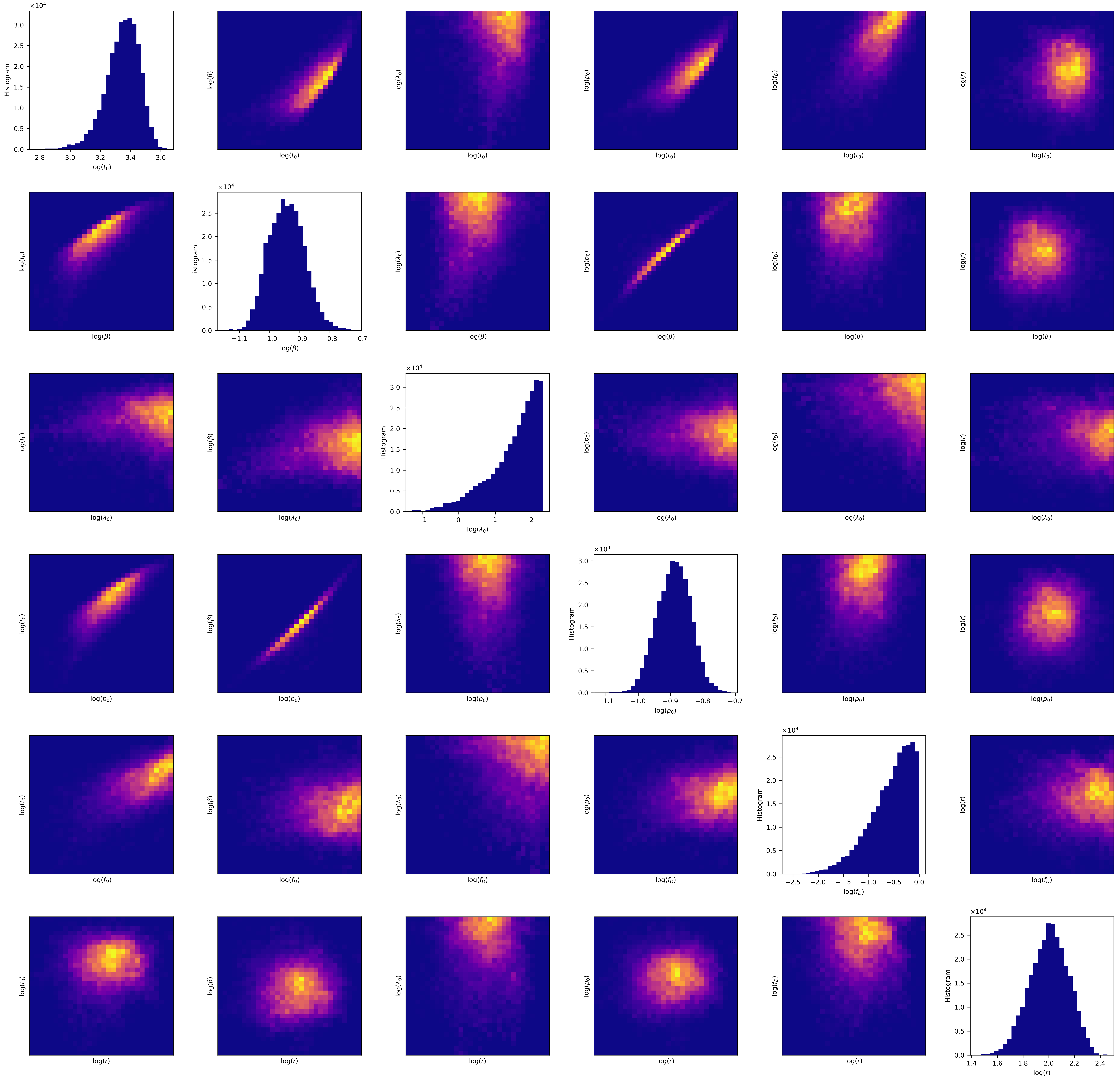

### Alaska

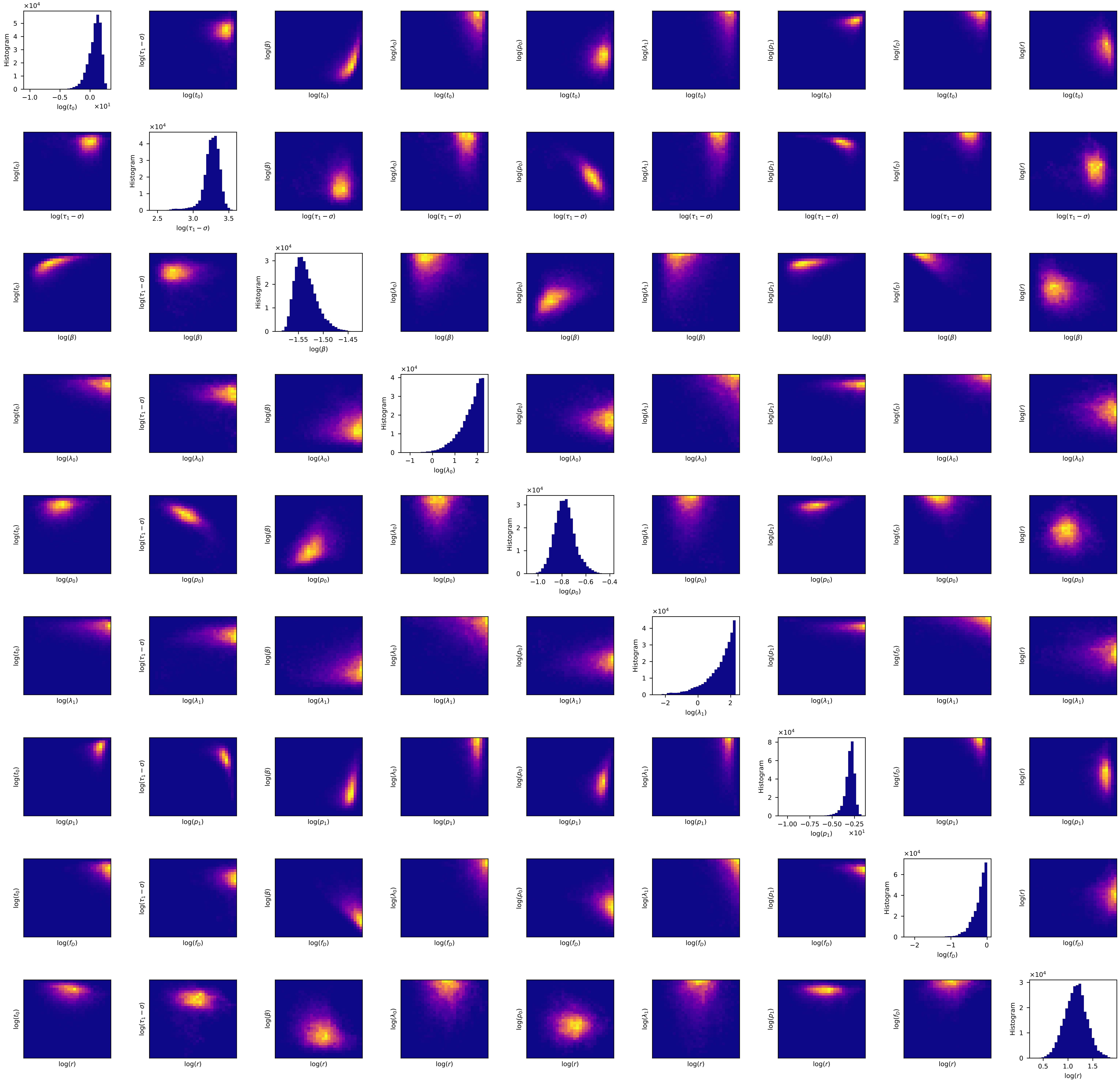
Histogram
log(t_0)
log(τ_1 − σ)
log(β)
log(λ_0)
log(p_0)
log(λ_1)
log(p_1)
log(f_D)
log(r)
×10^4
×10^1

Arizona

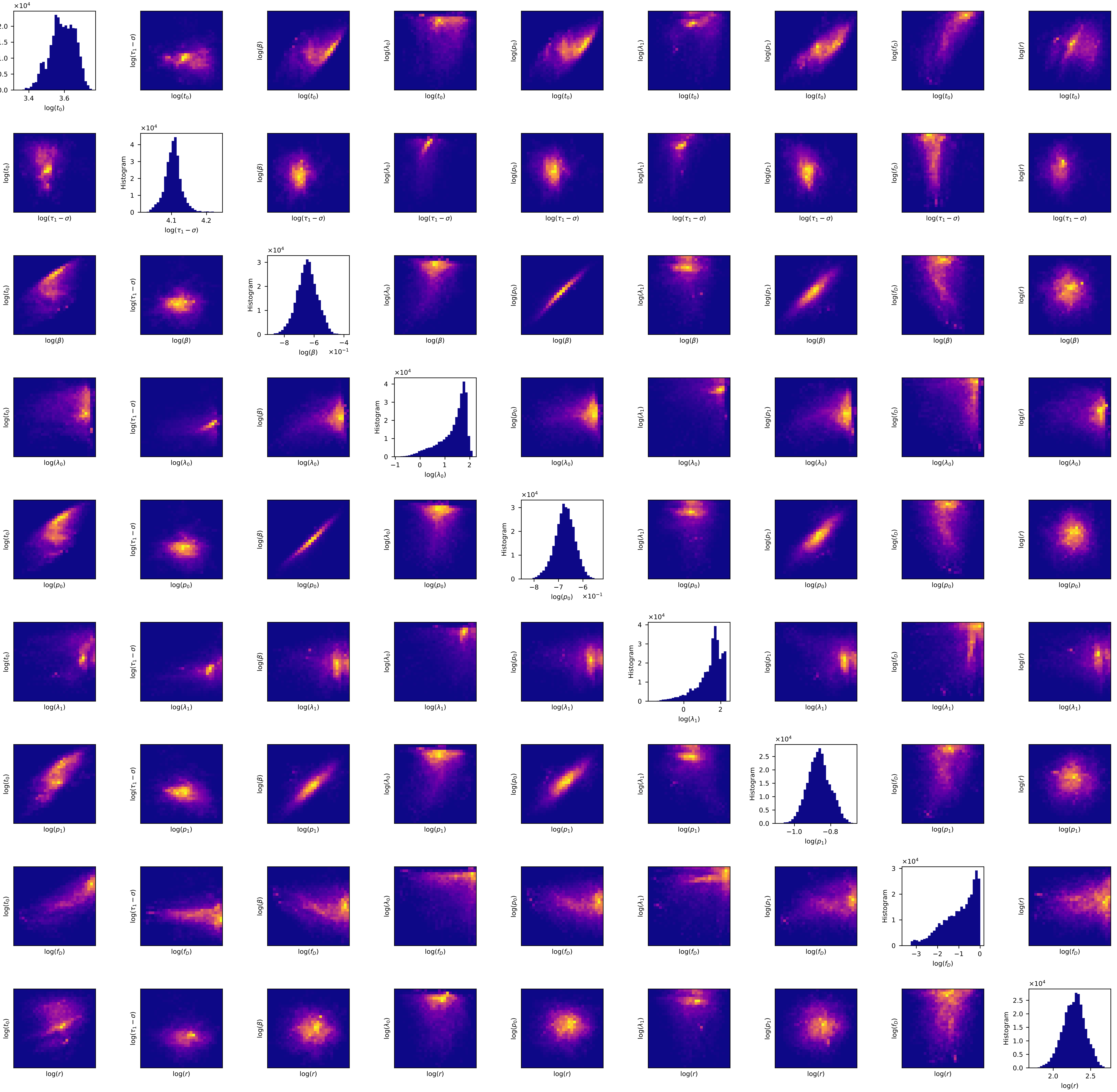

Histogram
log($t_0$)
log($\tau_1 - \sigma$)
log($\beta$)
log($\lambda_0$)
log($p_0$)
log($\lambda_1$)
log($p_1$)
log($f_D$)
log($r$)
×10^4
×10^-1

### Arkansas

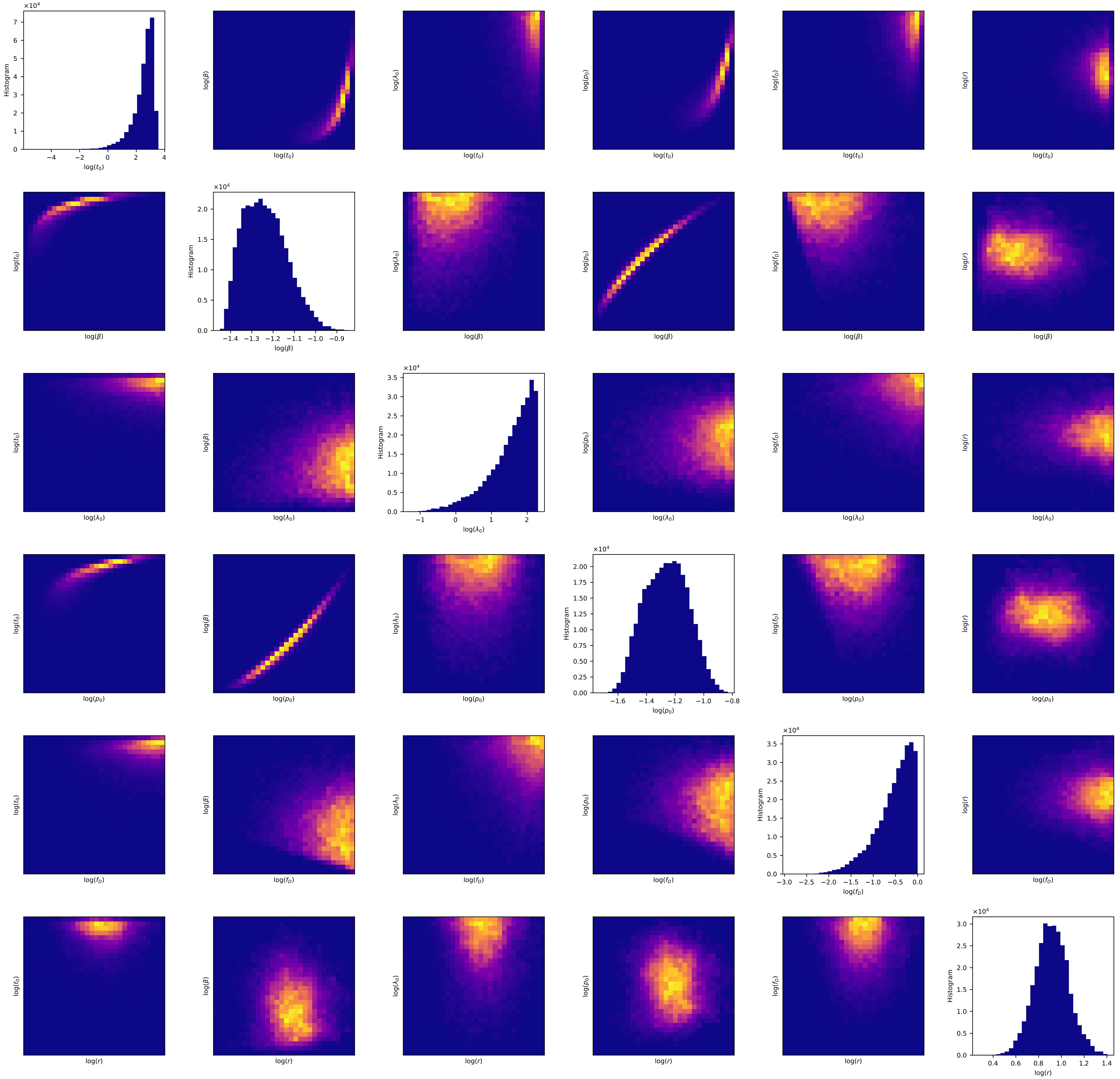

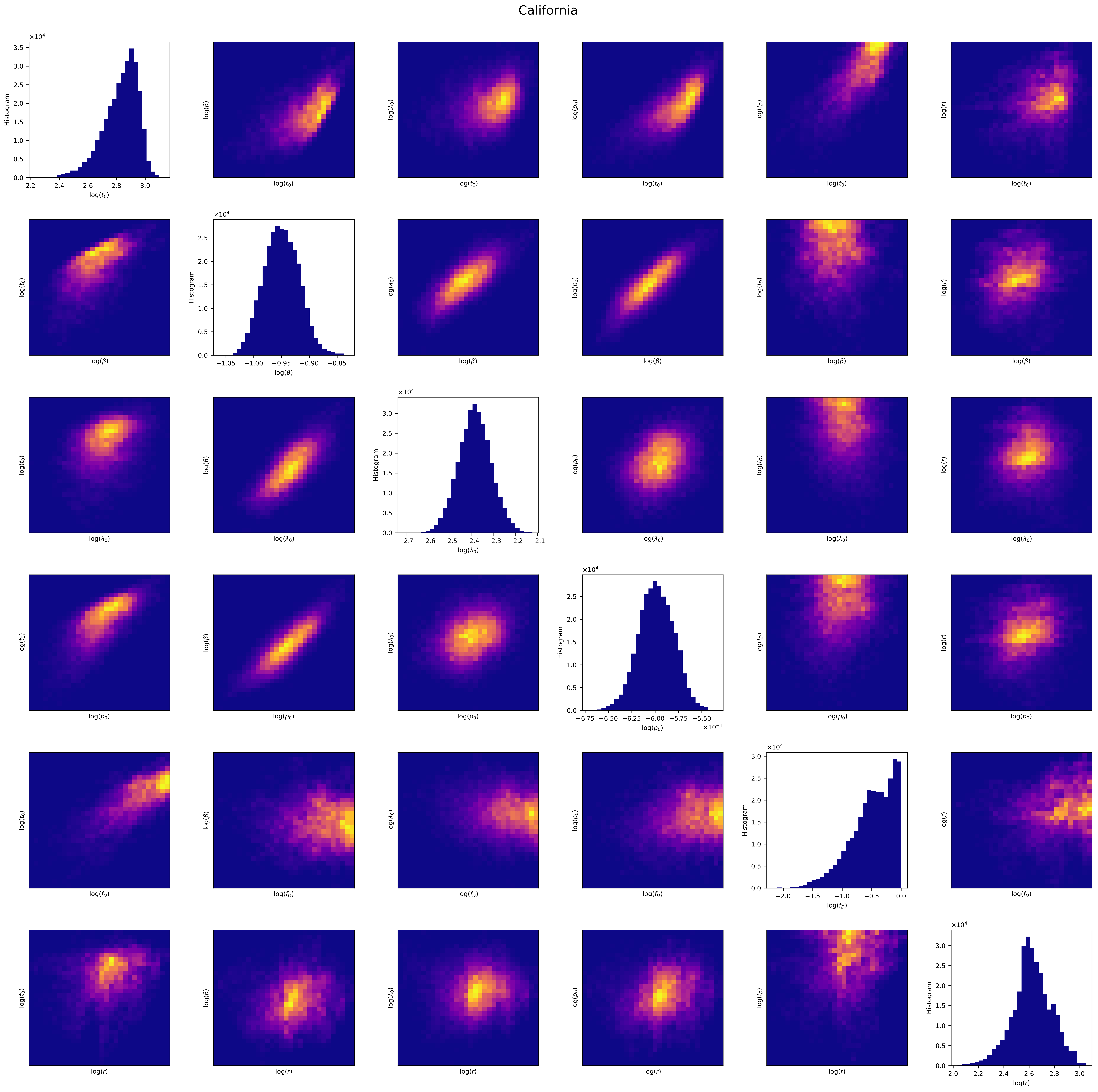
California
Histogram
log($t_0$)
log($\beta$)
log($\lambda_0$)
log($p_0$)
log($f_D$)
log($r$)
×10^4
×10^-1

### Colorado

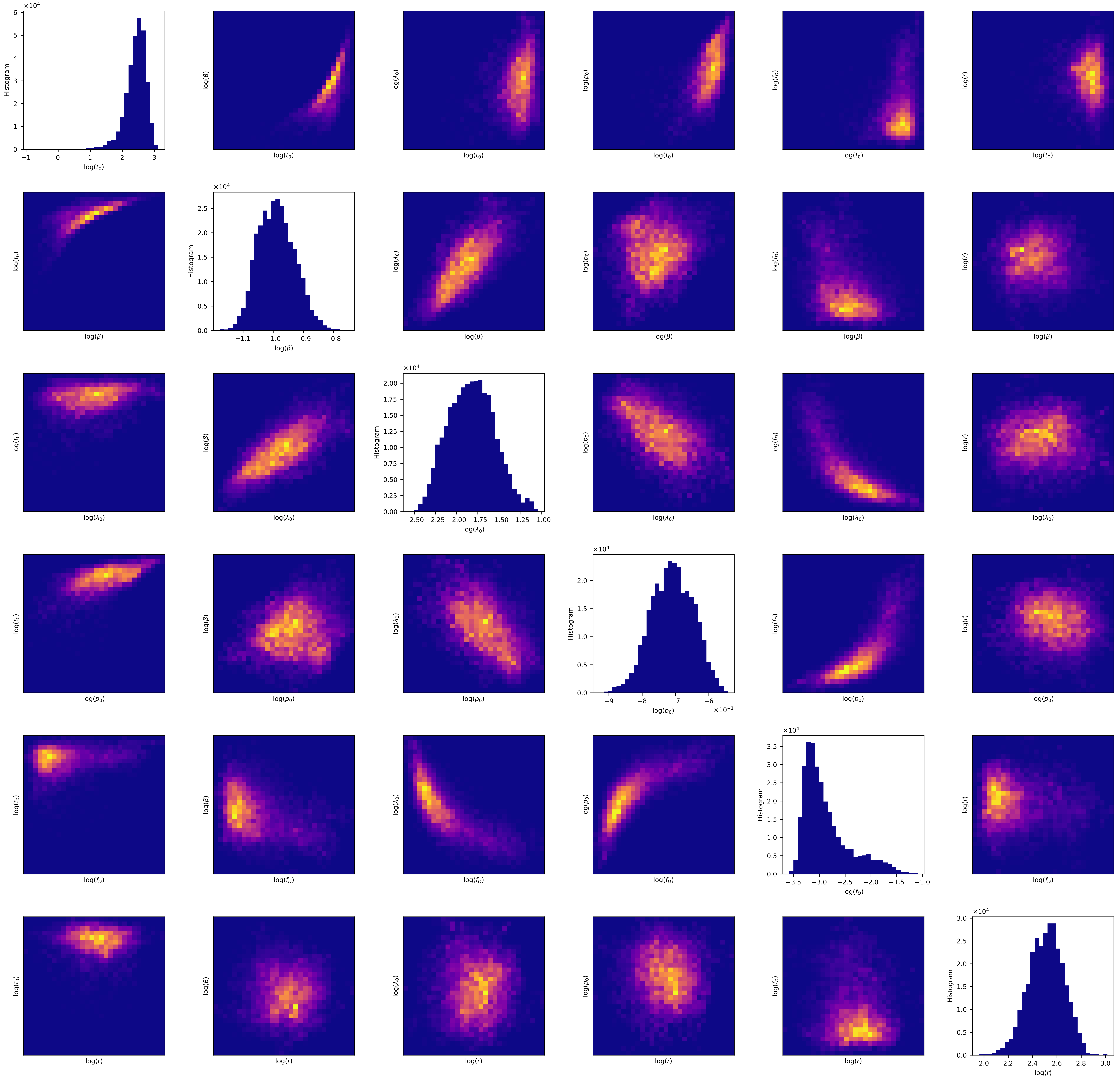

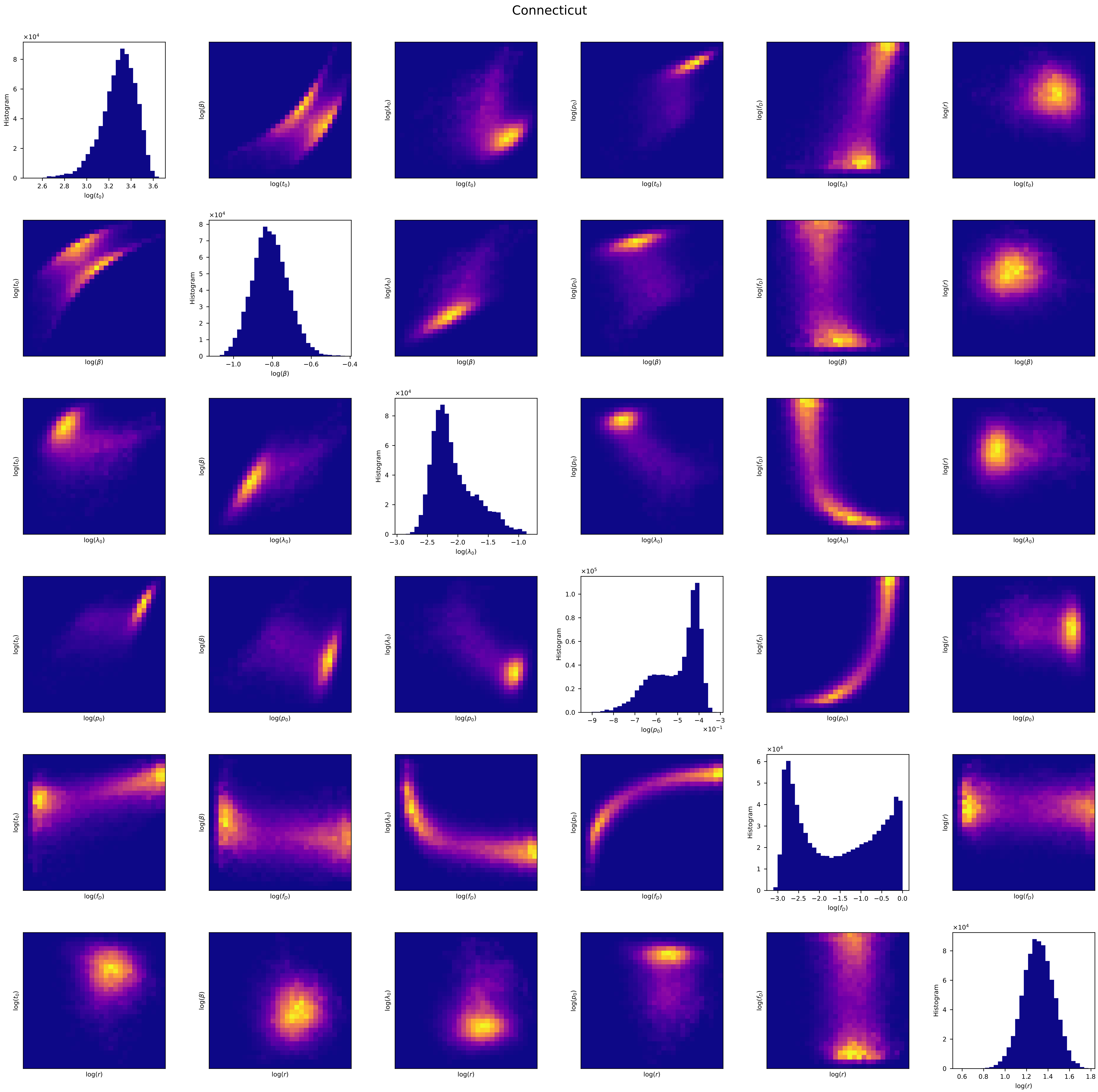
Connecticut
log($t_0$)
log($\beta$)
log($\lambda_0$)
log($p_0$)
log($f_D$)
log($r$)
Histogram

Delaware

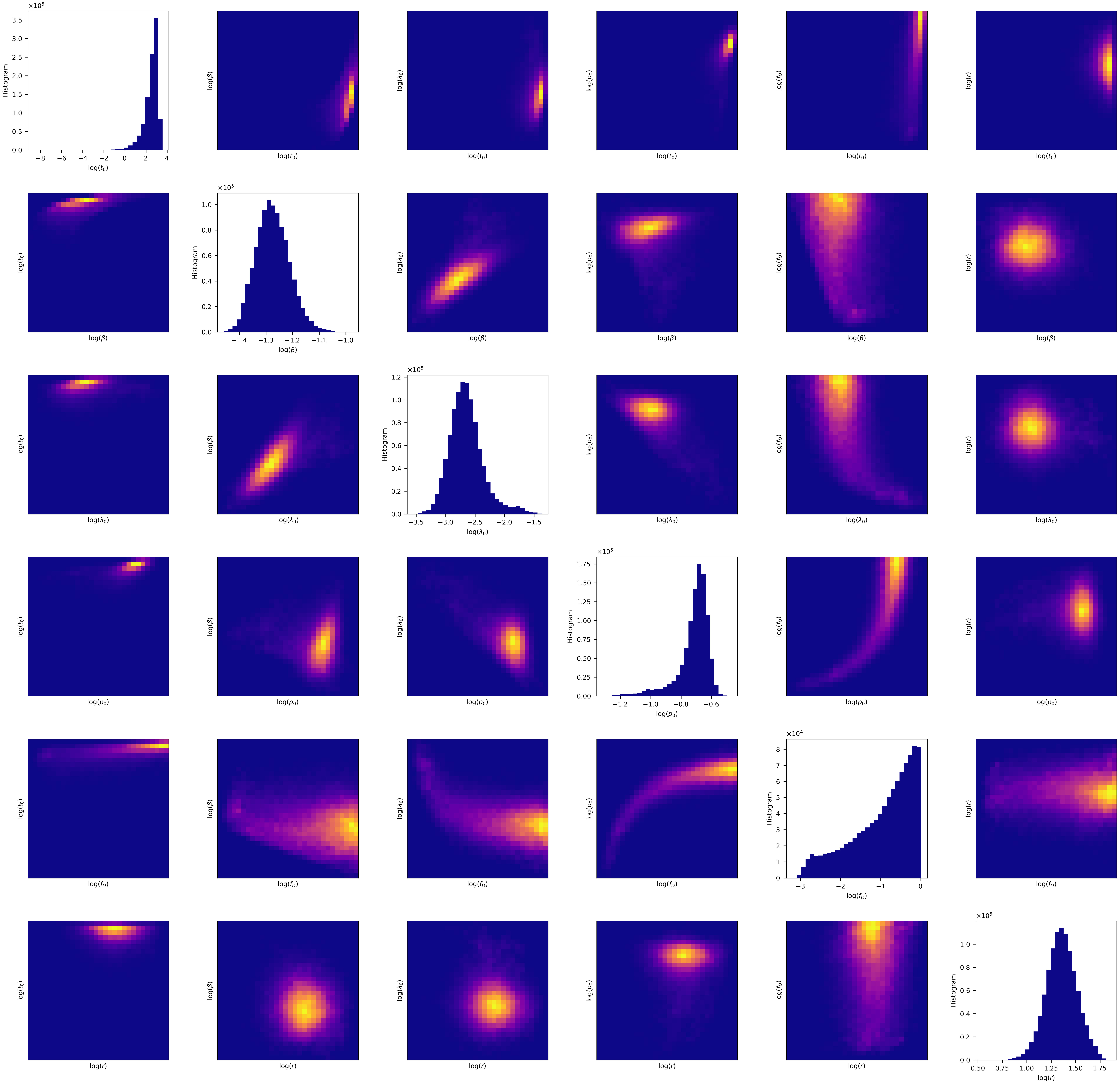

×10^5
Histogram
log($t_0$)
−8 −6 −4 −2 0 2 4
0.0 0.5 1.0 1.5 2.0 2.5 3.0 3.5
log($\beta$)
−1.4 −1.3 −1.2 −1.1 −1.0
0.0 0.2 0.4 0.6 0.8 1.0
log($\lambda_0$)
−3.5 −3.0 −2.5 −2.0 −1.5
0.0 0.2 0.4 0.6 0.8 1.0 1.2
log($p_0$)
−1.2 −1.0 −0.8 −0.6
0.00 0.25 0.50 0.75 1.00 1.25 1.50 1.75
log($f_D$)
×10^4
−3 −2 −1 0
0 1 2 3 4 5 6 7 8
log($r$)
0.50 0.75 1.00 1.25 1.50 1.75
0.0 0.2 0.4 0.6 0.8 1.0

### Florida

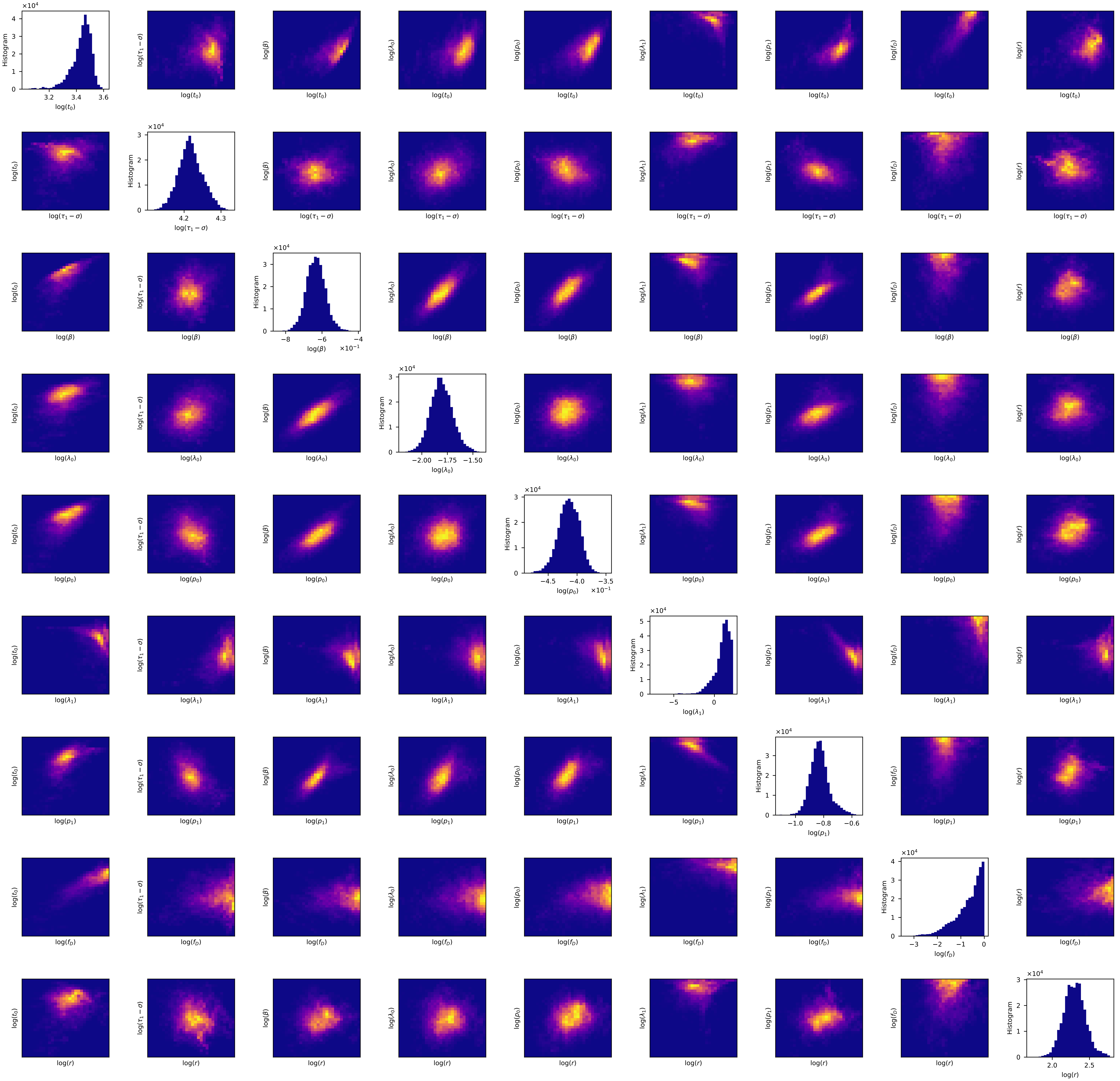
Histogram
log($t_0$)
log($\tau_1-\sigma$)
log($\beta$)
log($\lambda_0$)
log($p_0$)
log($\lambda_1$)
log($p_1$)
log($f_D$)
log($r$)

### Georgia

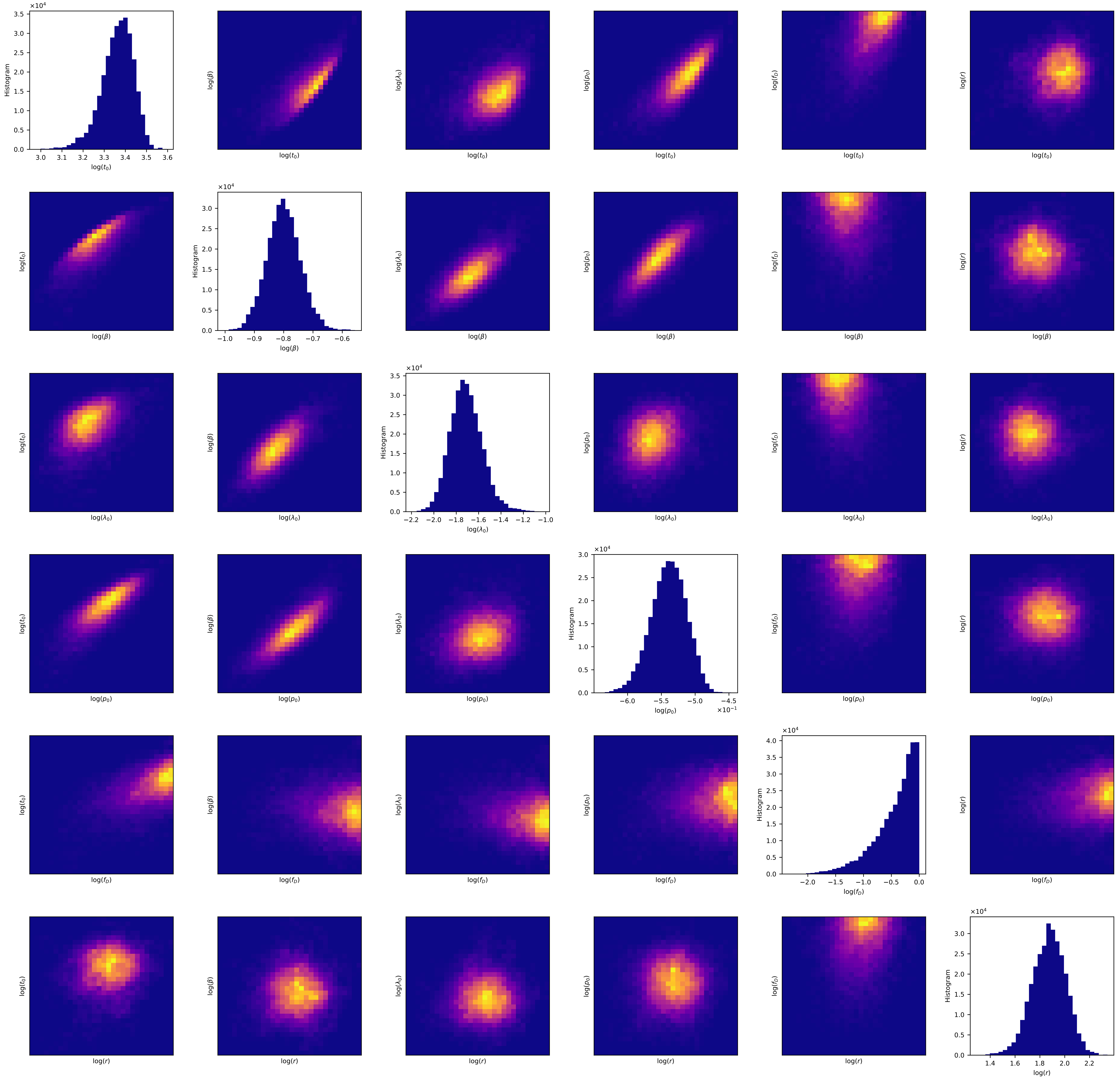

log($t_0$)
log($\beta$)
log($\lambda_0$)
log($p_0$)
log($f_D$)
log($r$)
Histogram
×10^4
×10^-1

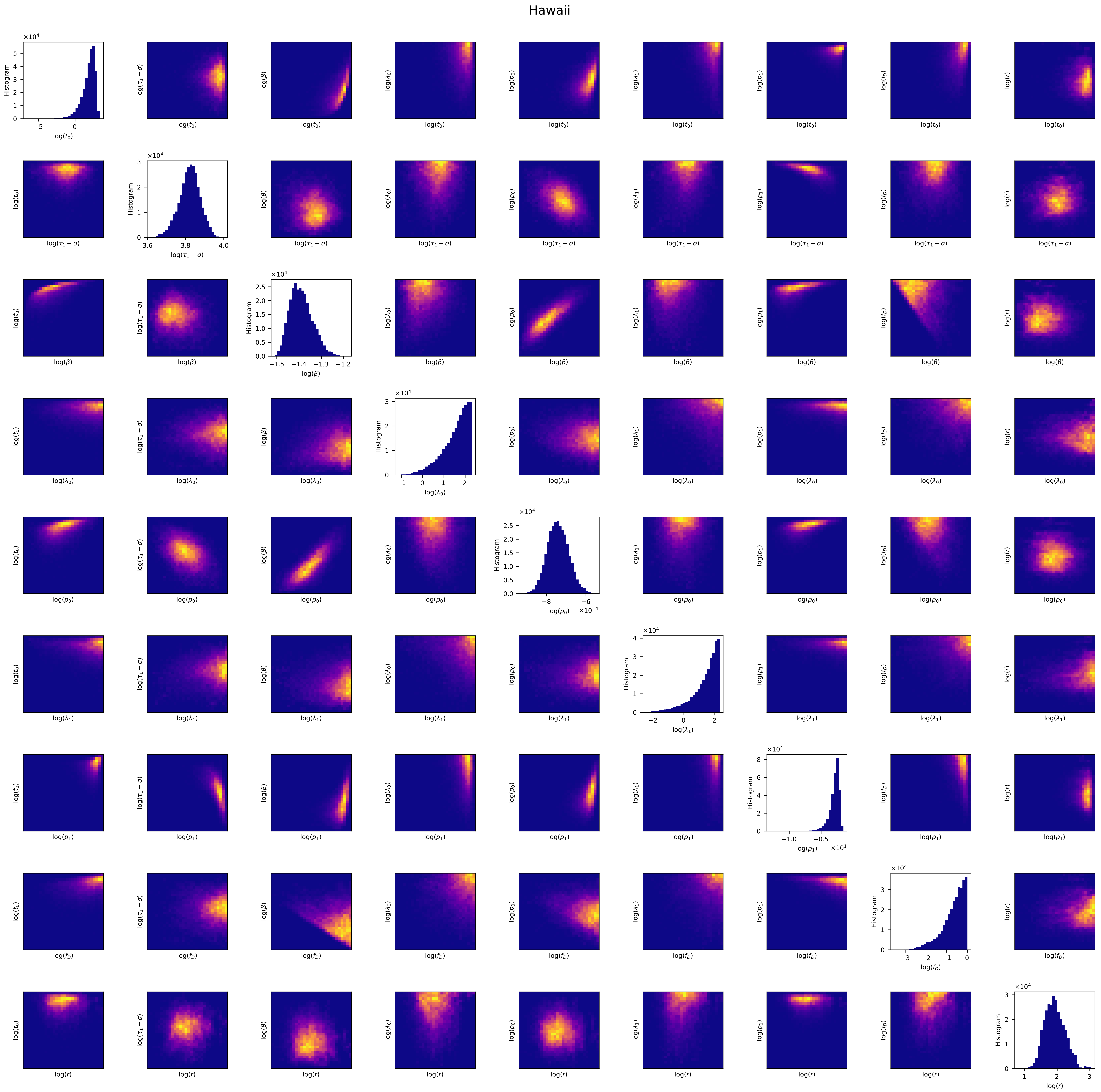
Hawaii
Histogram
log($t_0$)
log($\tau_1 - \sigma$)
log($\beta$)
log($\lambda_0$)
log($p_0$)
log($\lambda_1$)
log($p_1$)
log($f_D$)
log($r$)

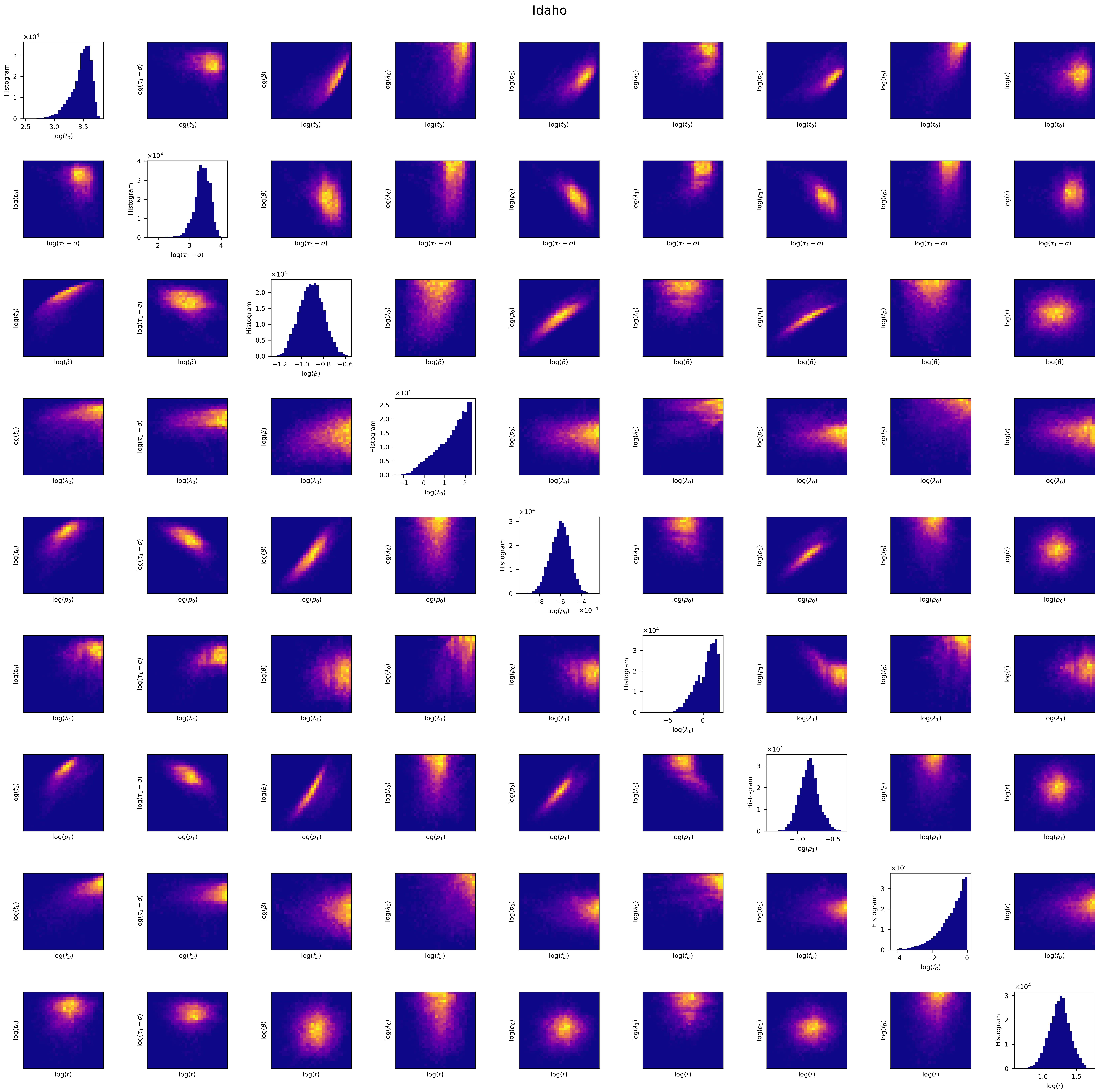
Idaho
log($t_0$)
log($\tau_1 - \sigma$)
log($\beta$)
log($\lambda_0$)
log($p_0$)
log($\lambda_1$)
log($p_1$)
log($f_D$)
log($r$)
Histogram

Illinois

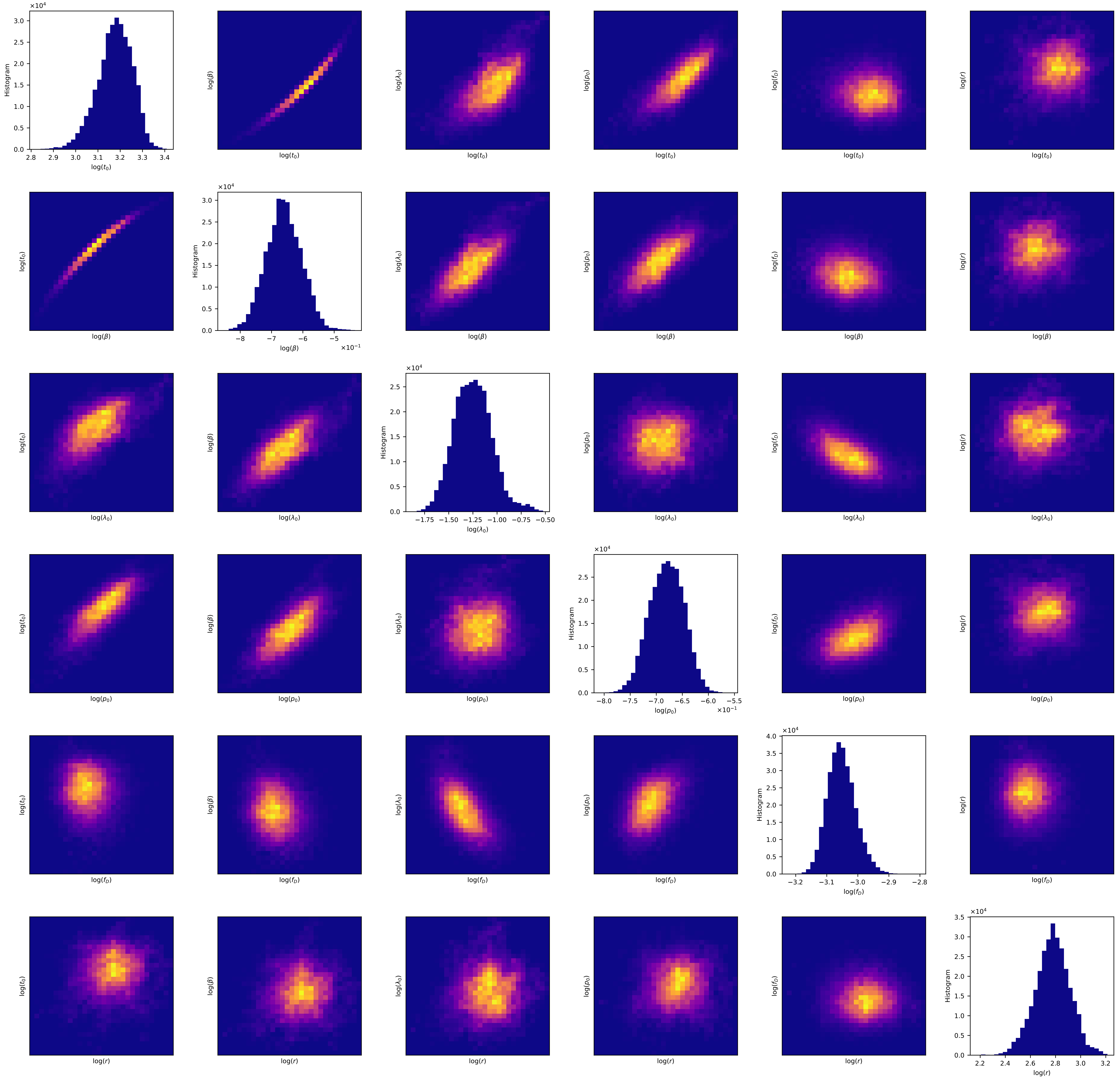

Histogram
log($t_0$)
log($\beta$)
log($\lambda_0$)
log($p_0$)
log($f_D$)
log($r$)
×10^4
×10^-1

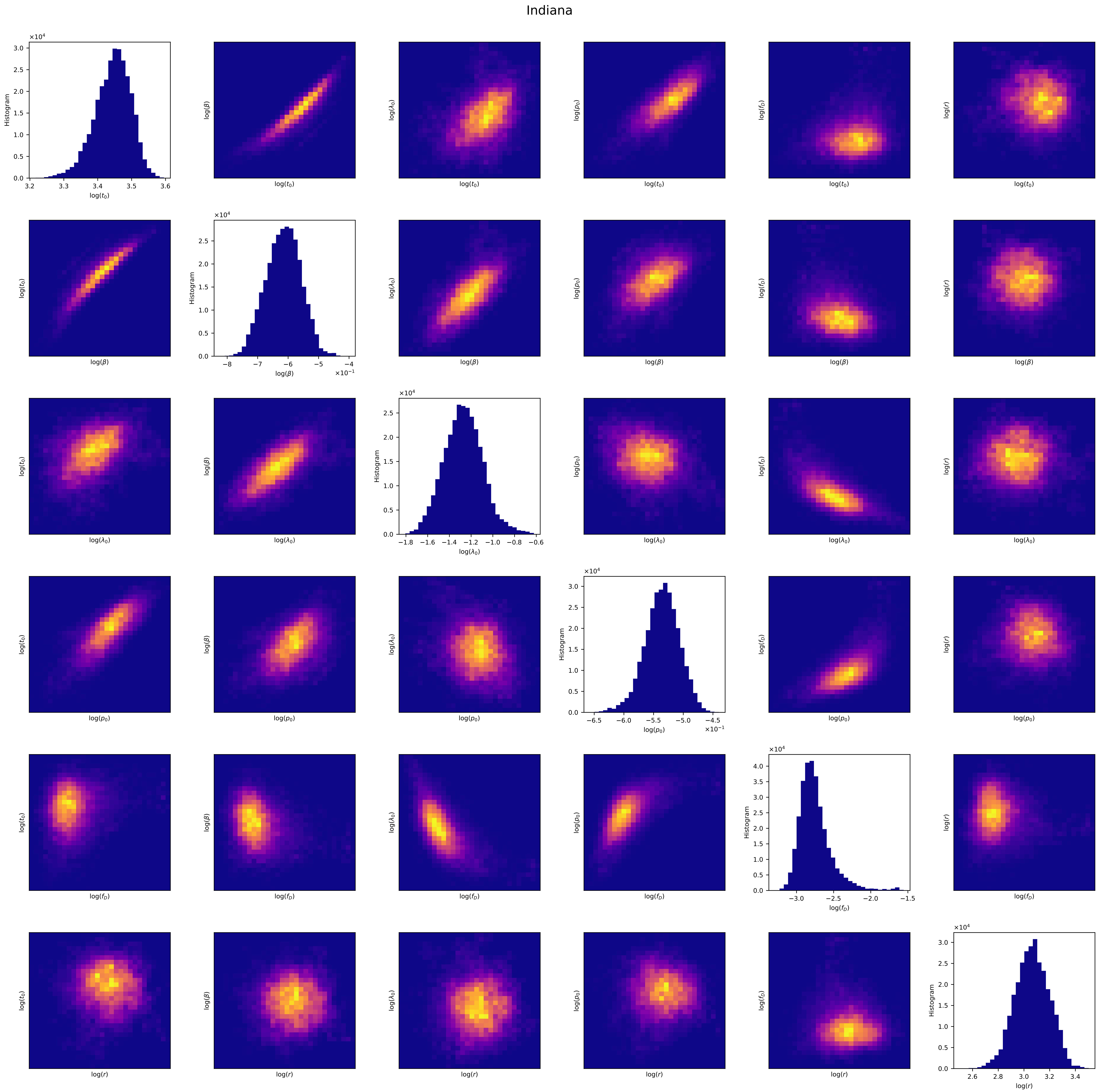

Indiana
Histogram
log($t_0$)
log($\beta$)
log($\lambda_0$)
log($p_0$)
log($f_D$)
log($r$)
×10^4
×10^-1

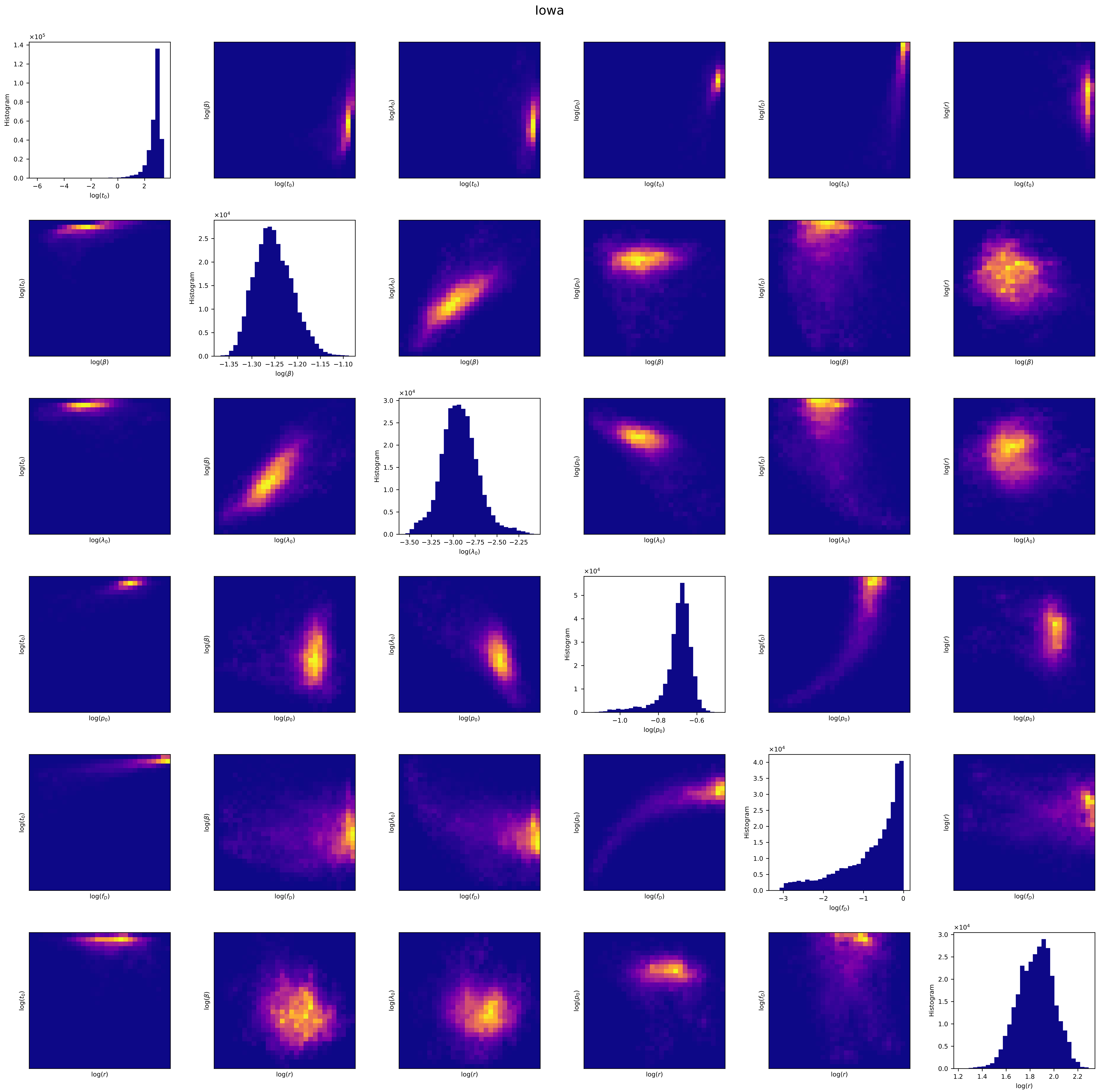
Iowa
Histogram
log($t_0$)
log($\beta$)
log($\lambda_0$)
log($p_0$)
log($f_D$)
log($r$)

Kansas

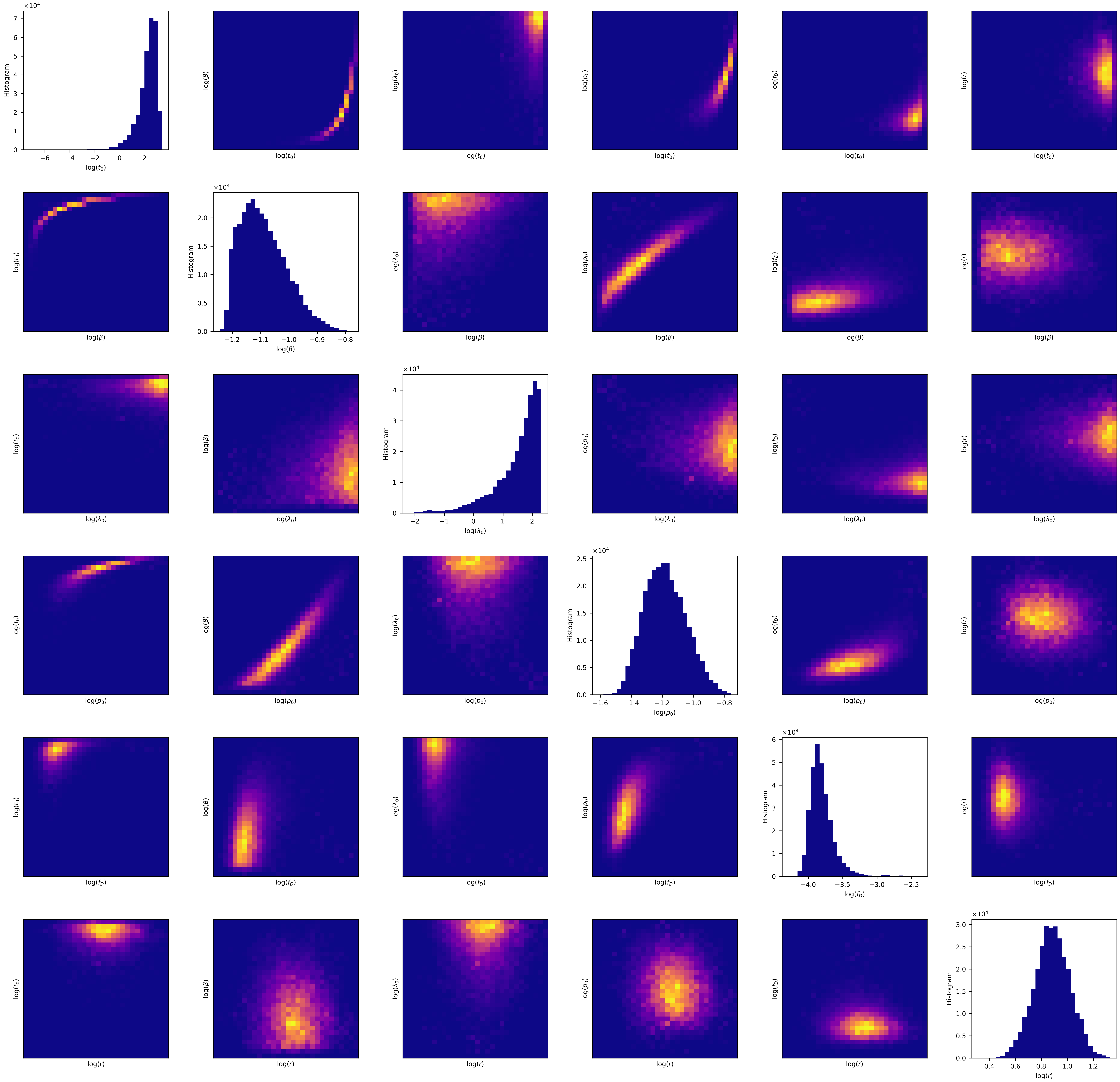

Kentucky

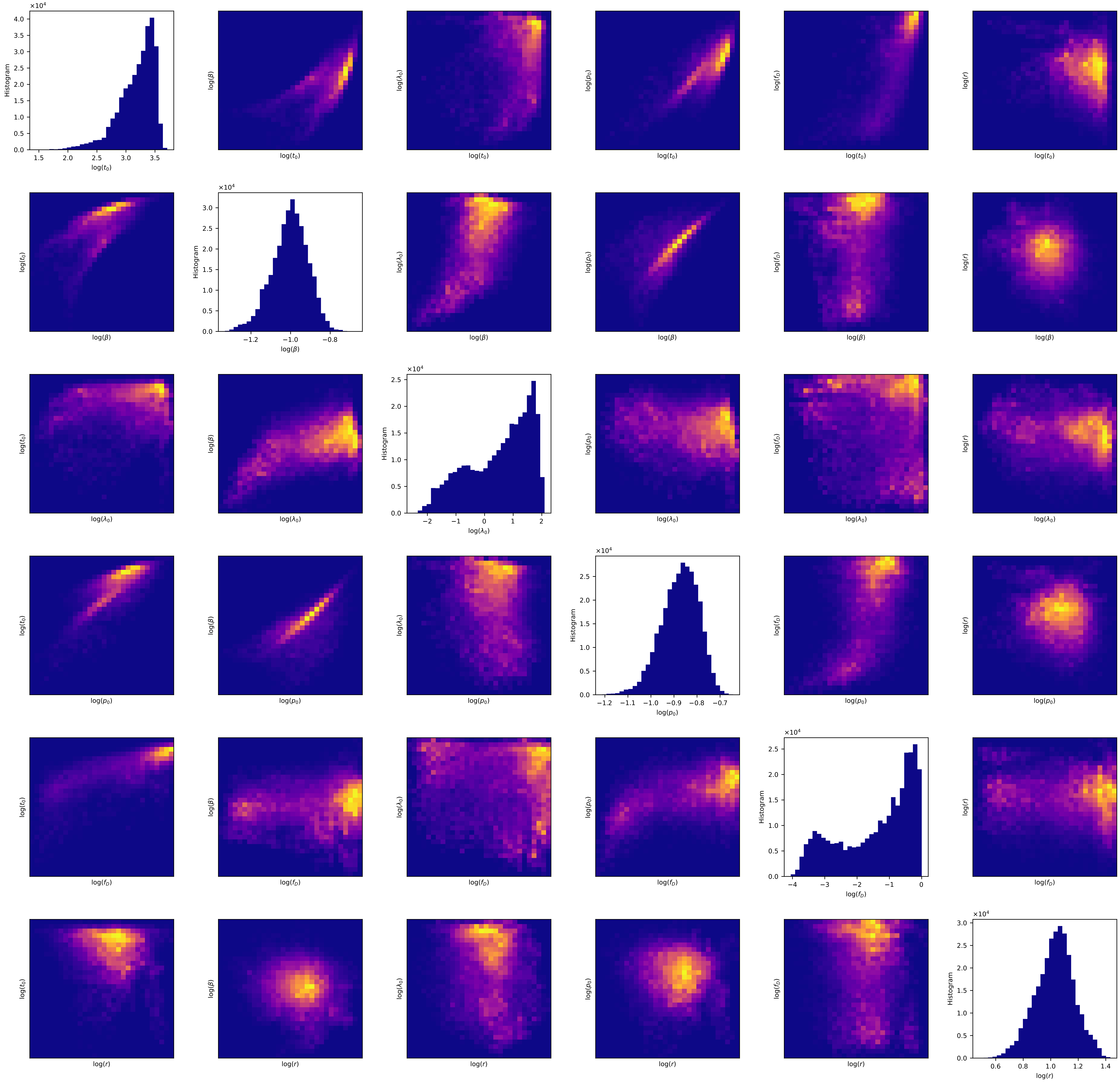
Histogram
log($t_0$)
log($\beta$)
log($\lambda_0$)
log($p_0$)
log($f_D$)
log($r$)
$\times 10^4$

Louisiana

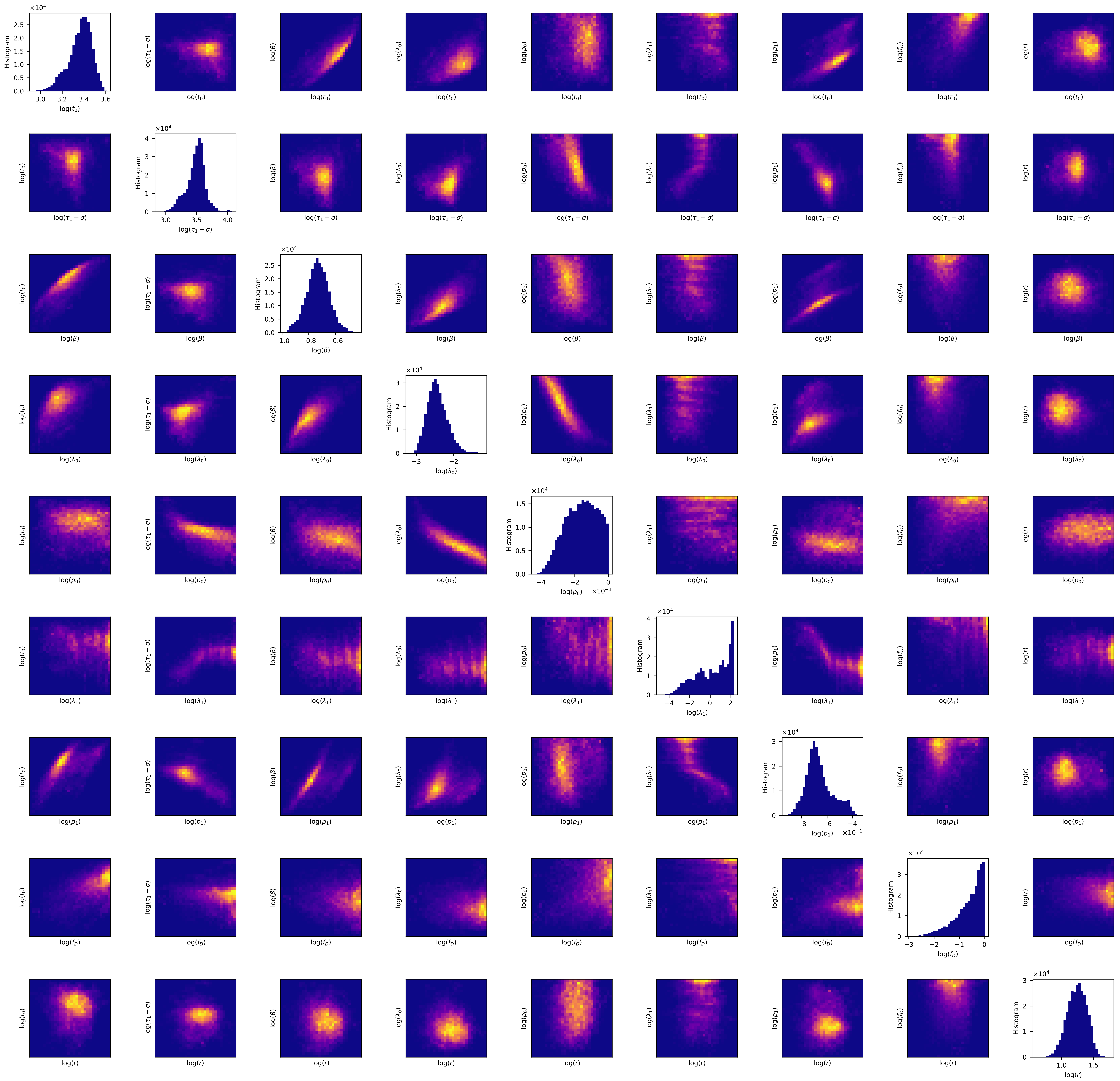

×10⁴
Histogram
log($t_0$)
log($\tau_1 - \sigma$)
log($\beta$)
log($\lambda_0$)
log($p_0$)
log($\lambda_1$)
log($p_1$)
log($f_D$)
log($r$)
0.0 0.5 1.0 1.5 2.0 2.5
3.0 3.2 3.4 3.6
0 1 2 3 4
3.0 3.5 4.0
−1.0 −0.8 −0.6
−3 −2
−4 −2 0
×10⁻¹
−4 −2 0 2
−8 −6 −4
−3 −2 −1 0
1.0 1.5

Maine

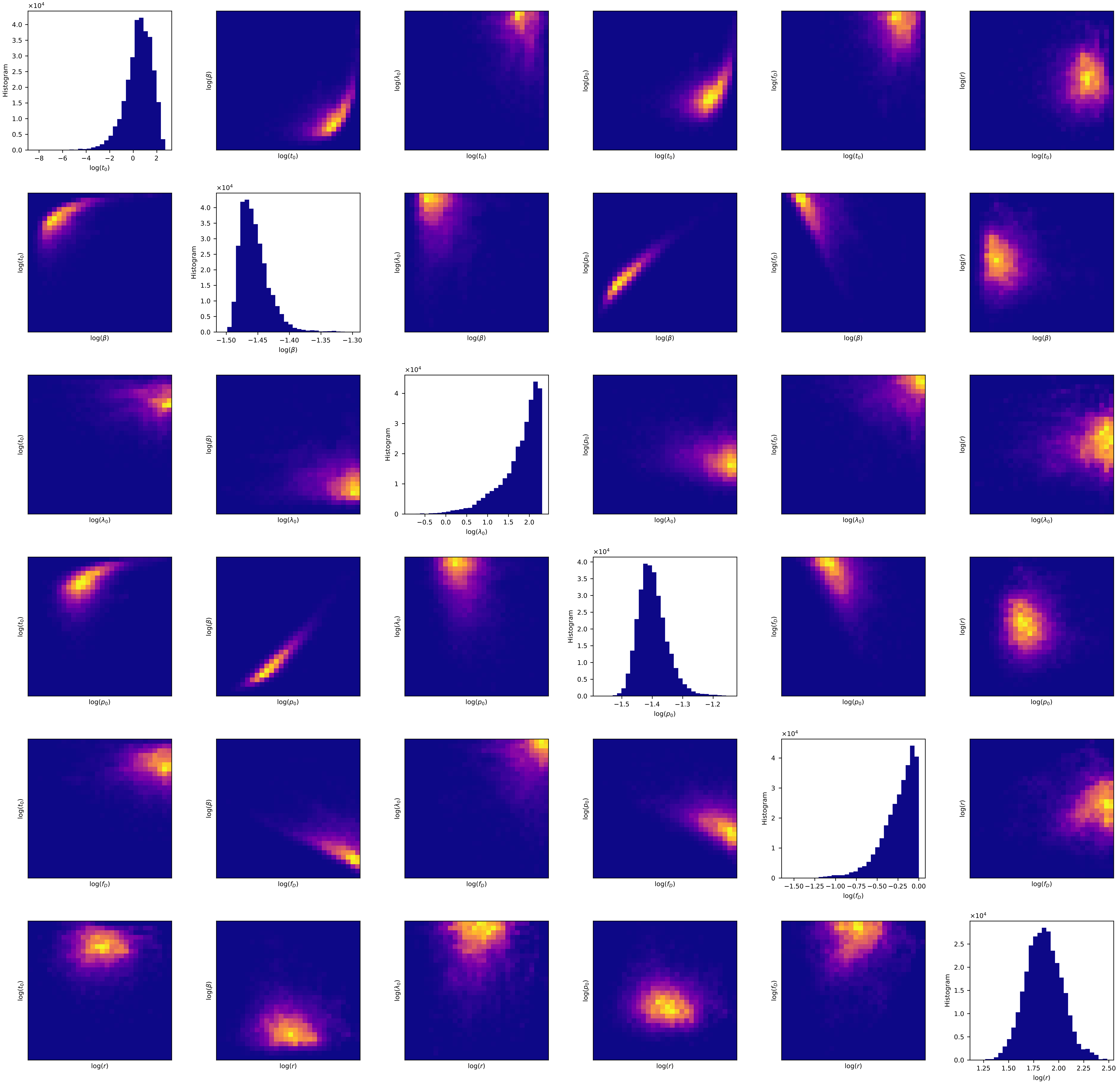

×10^4
Histogram
log(t_0)
log(β)
log(λ_0)
log(p_0)
log(f_D)
log(r)
−8 −6 −4 −2 0 2
0.0 0.5 1.0 1.5 2.0 2.5 3.0 3.5 4.0
−1.50 −1.45 −1.40 −1.35 −1.30
−0.5 0.0 0.5 1.0 1.5 2.0
0 1 2 3 4
−1.5 −1.4 −1.3 −1.2
−1.50 −1.25 −1.00 −0.75 −0.50 −0.25 0.00
1.25 1.50 1.75 2.00 2.25 2.50

#### Maryland

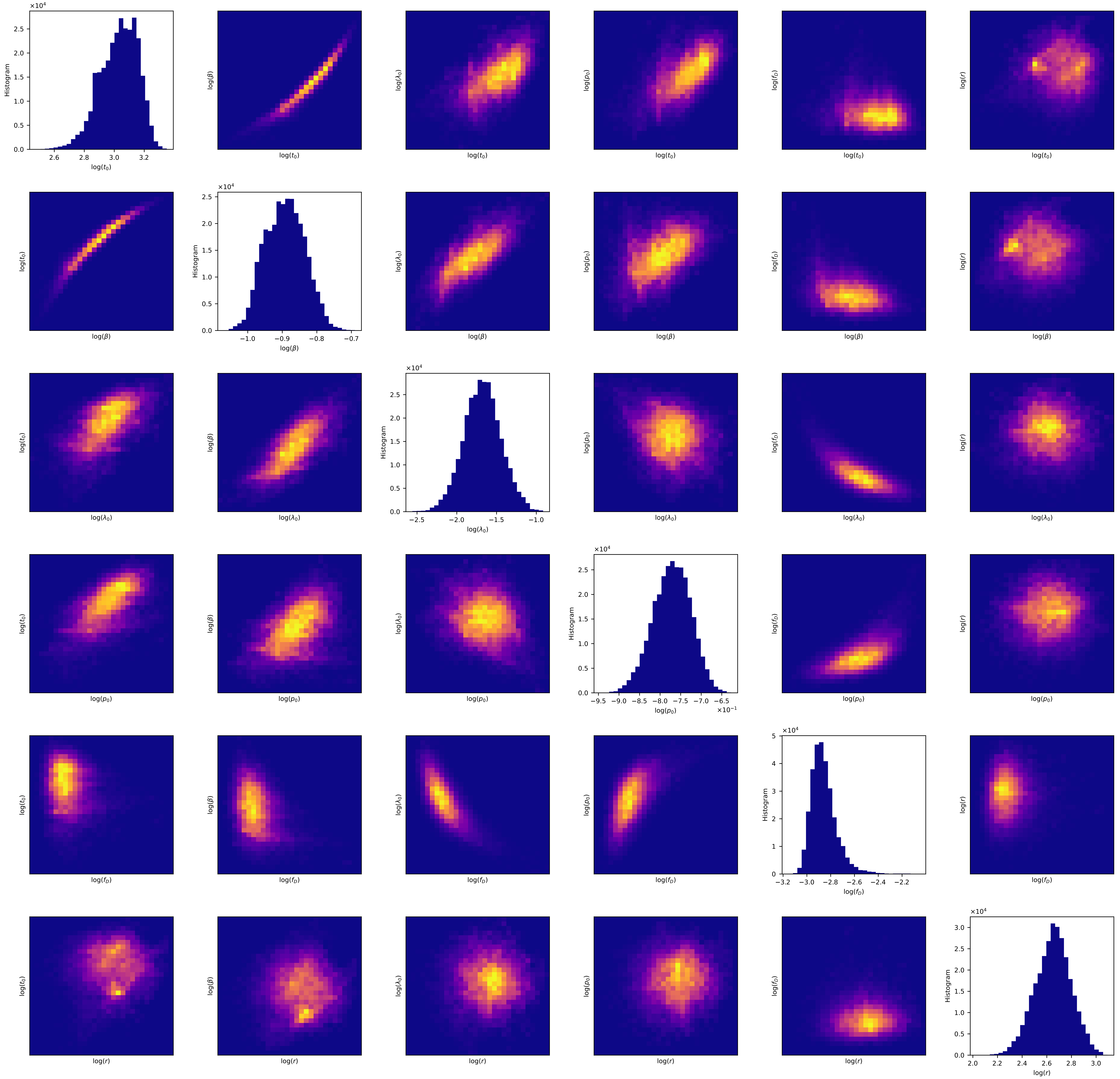

#### Massachusetts

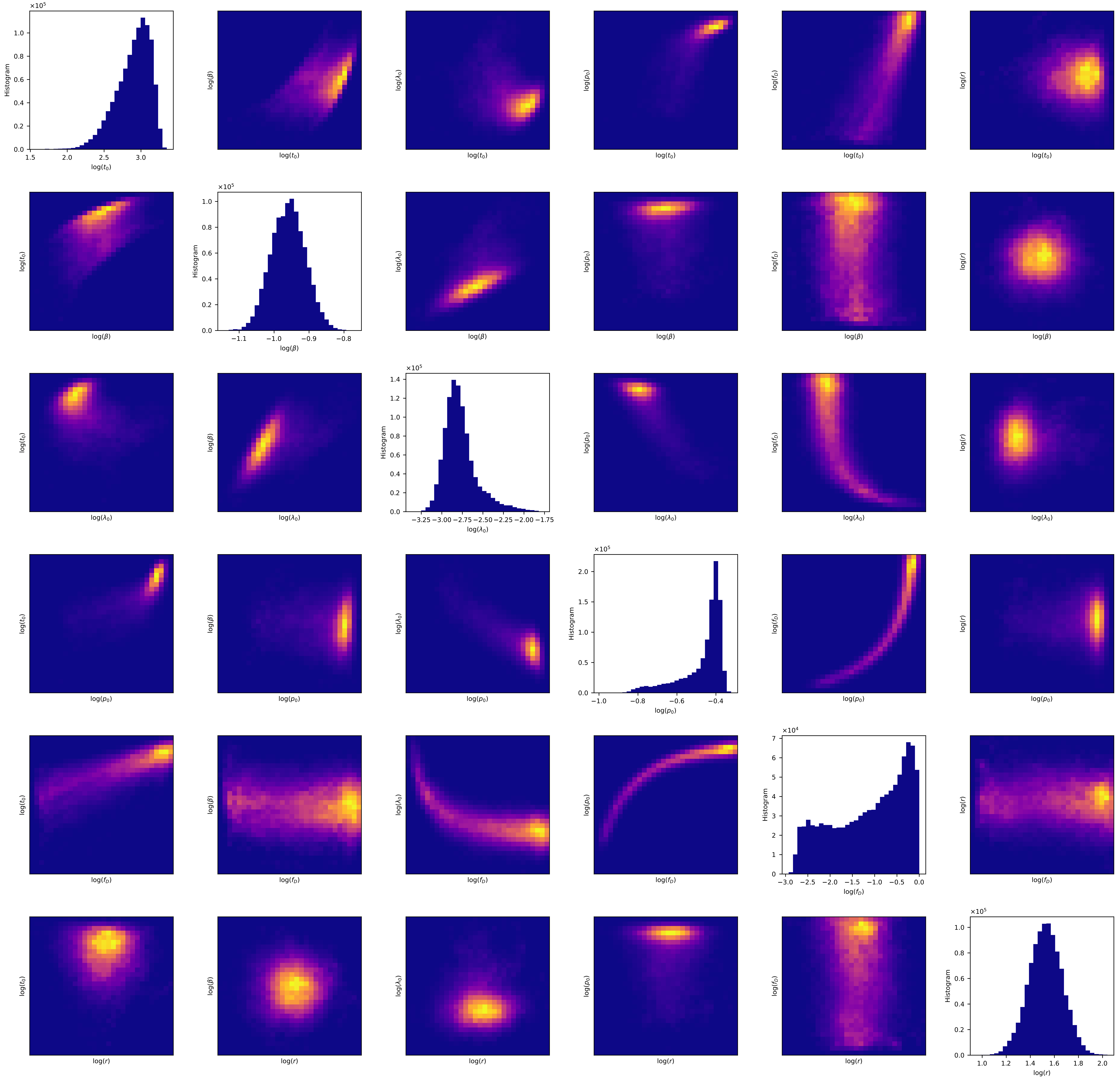

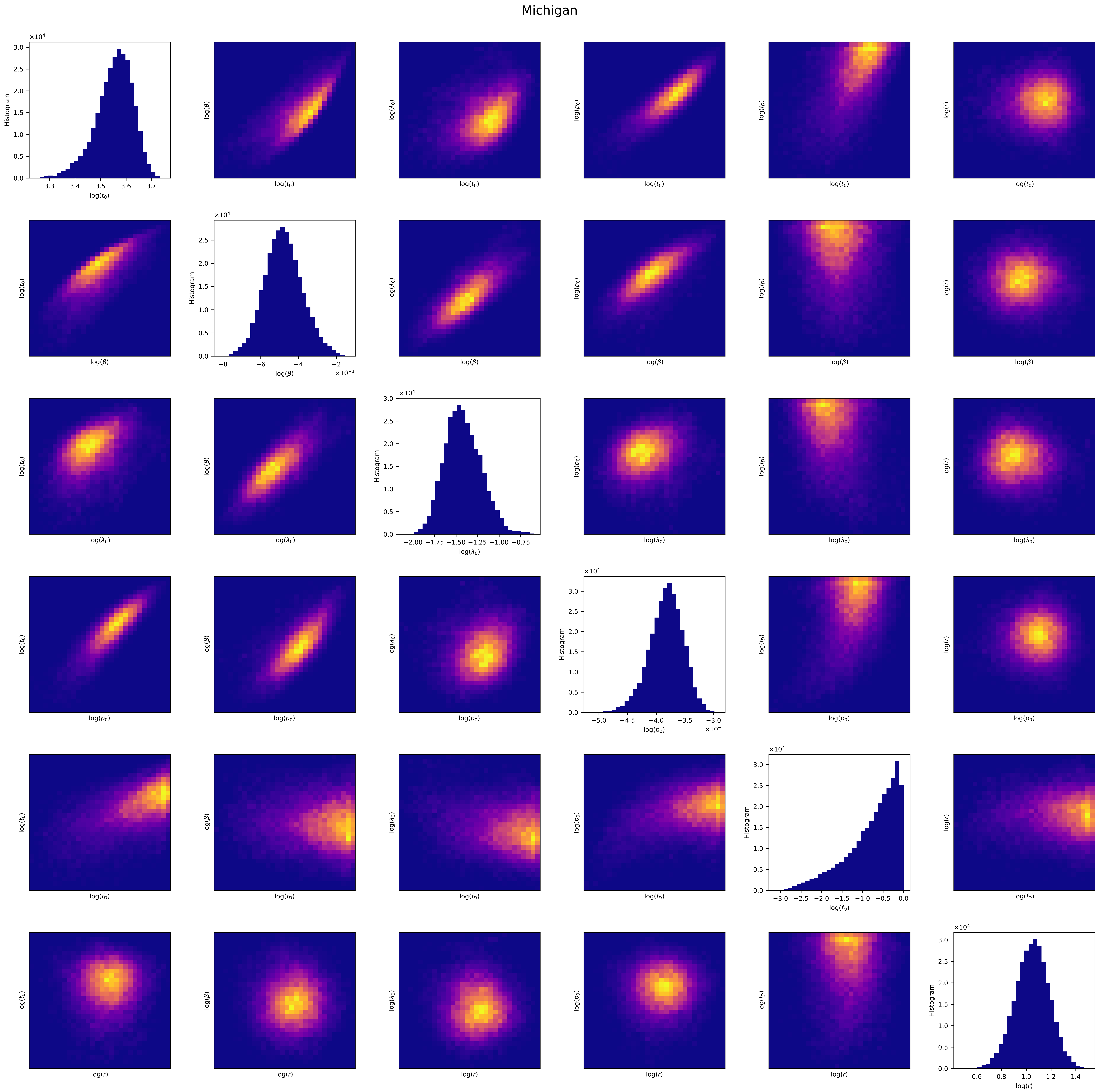
Michigan
Histogram
$\log(t_0)$
$\log(\beta)$
$\log(\lambda_0)$
$\log(p_0)$
$\log(f_D)$
$\log(r)$

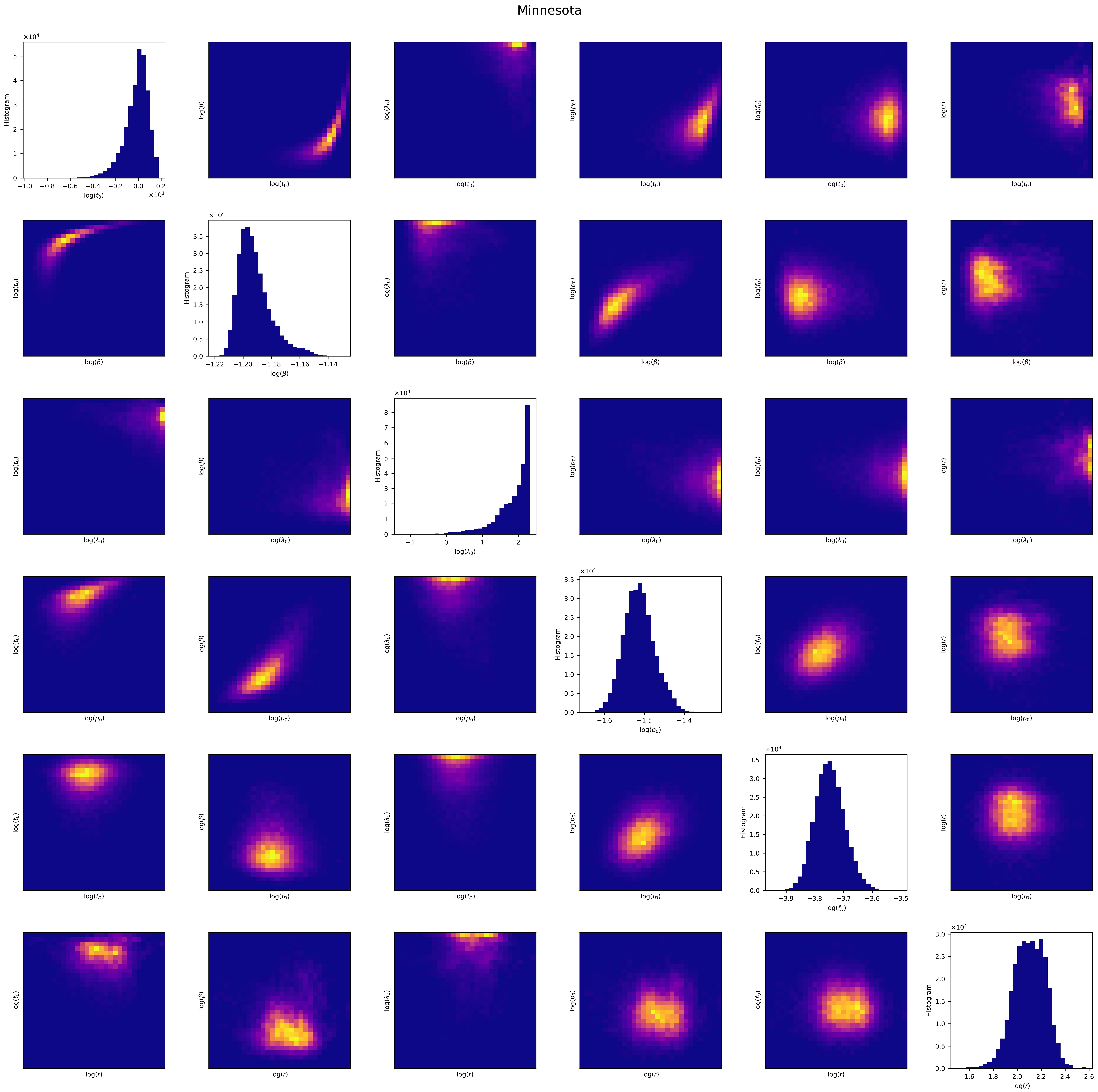
Minnesota
Histogram
log($t_0$)
log($\beta$)
log($\lambda_0$)
log($p_0$)
log($f_D$)
log($r$)
×10^4
×10^1

Mississippi

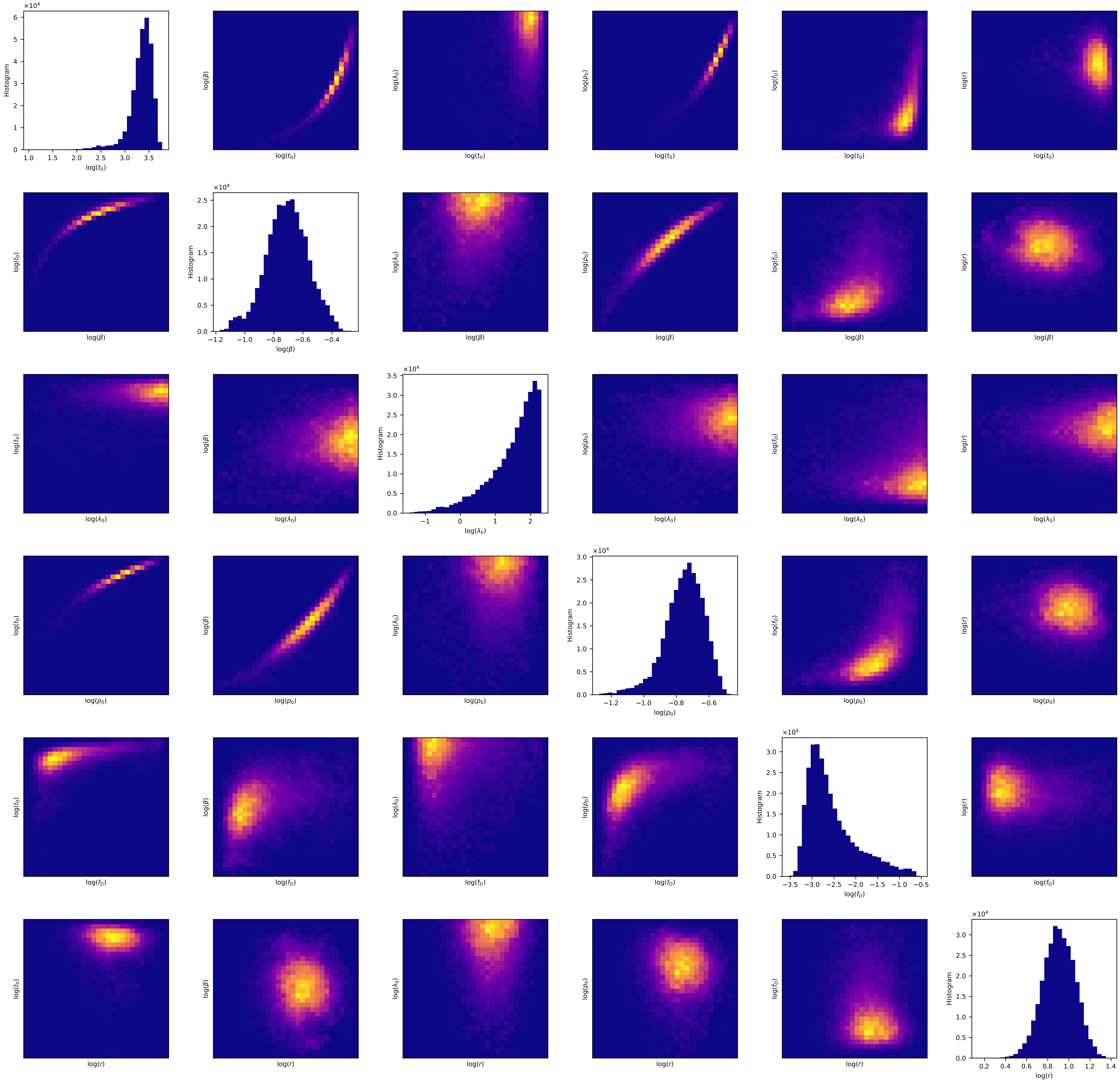

### Missouri

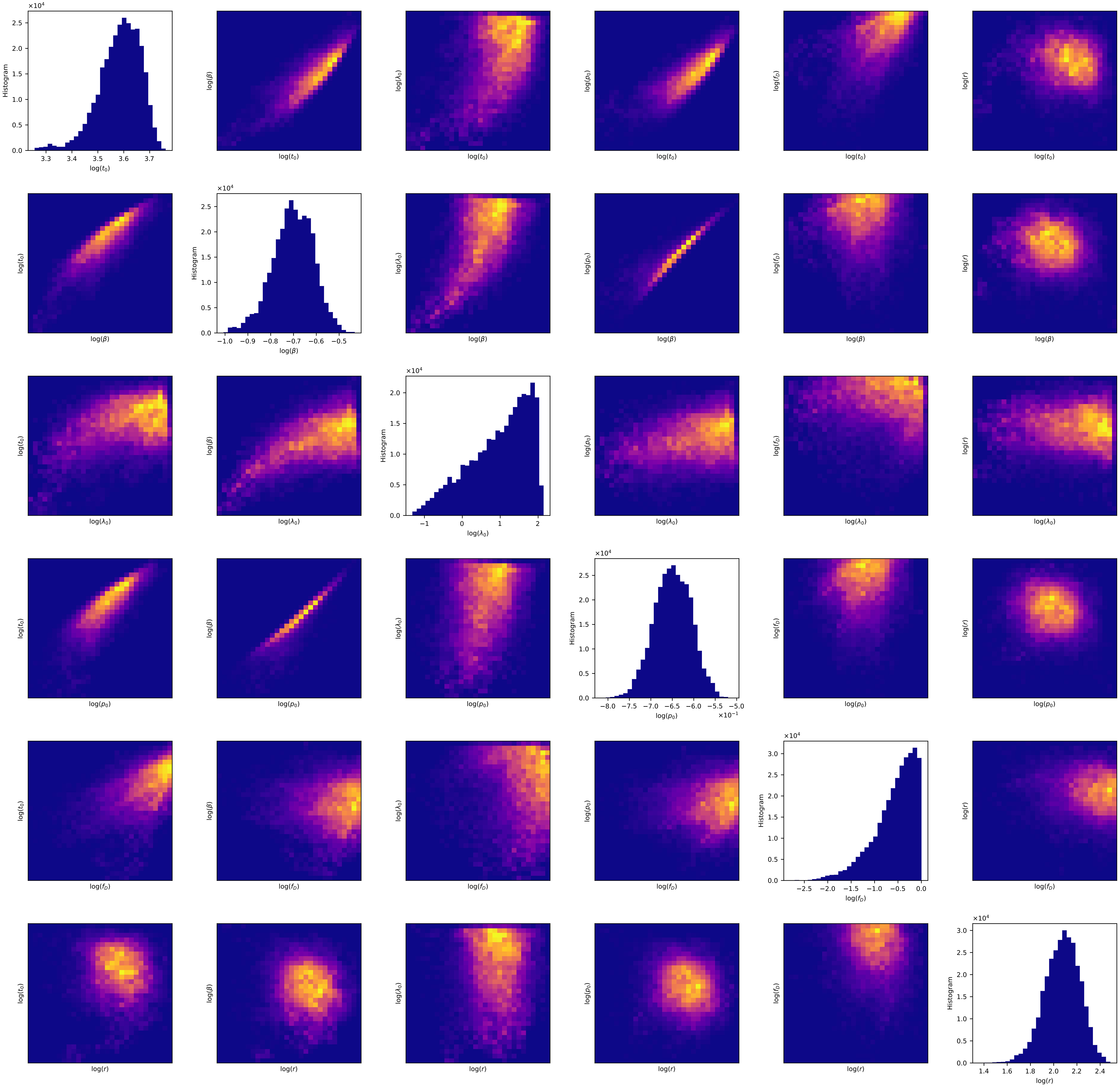

Histogram
log($t_0$)
log($\beta$)
log($\lambda_0$)
log($p_0$)
log($f_D$)
log($r$)
×10^4
×10^-1

### Montana

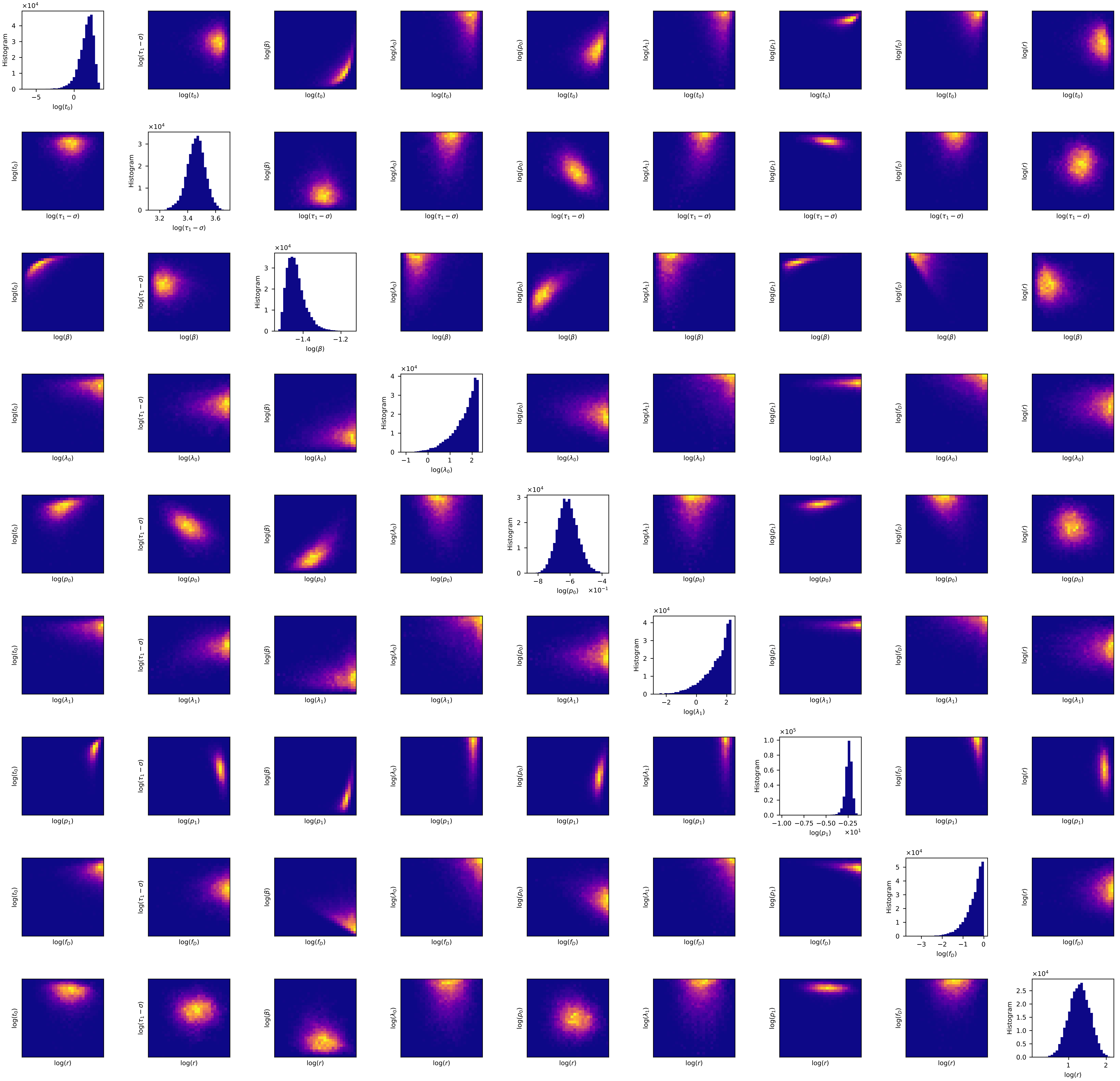
Histogram
log($t_0$)
log($\tau_1-\sigma$)
log($\beta$)
log($\lambda_0$)
log($p_0$)
log($\lambda_1$)
log($p_1$)
log($f_D$)
log($r$)

Nebraska

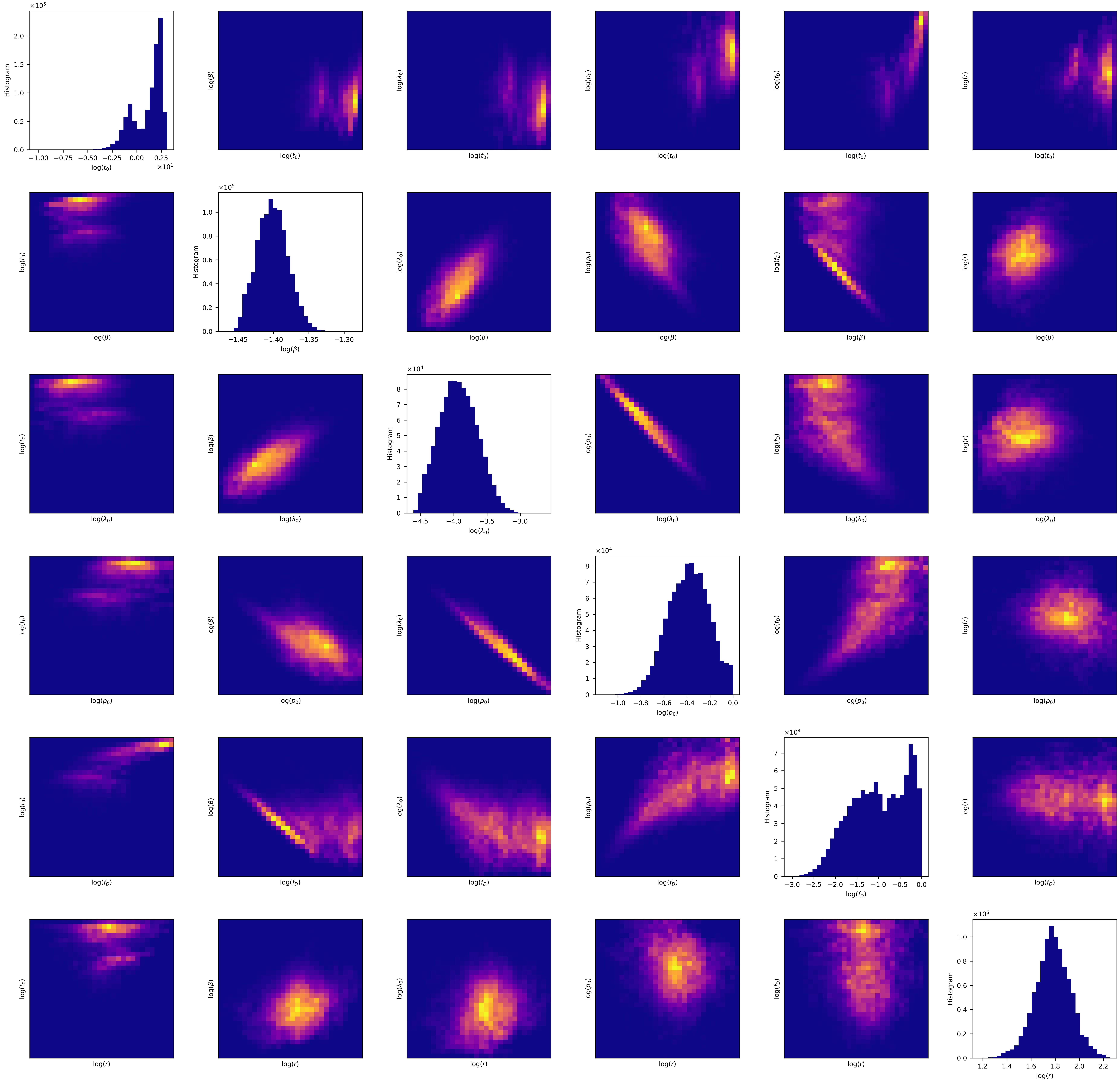

×10^5
Histogram
2.0
1.5
1.0
0.5
0.0
−1.00 −0.75 −0.50 −0.25 0.00 0.25
log(t_0)
×10^1
log(β)
log(λ_0)
log(p_0)
log(f_D)
log(r)
log(t_0)
×10^5
1.0
0.8
0.6
0.4
0.2
0.0
−1.45 −1.40 −1.35 −1.30
×10^4
8
7
6
5
4
3
2
1
0
−4.5 −4.0 −3.5 −3.0
−1.0 −0.8 −0.6 −0.4 −0.2 0.0
−3.0 −2.5 −2.0 −1.5 −1.0 −0.5 0.0
1.2 1.4 1.6 1.8 2.0 2.2

### Nevada

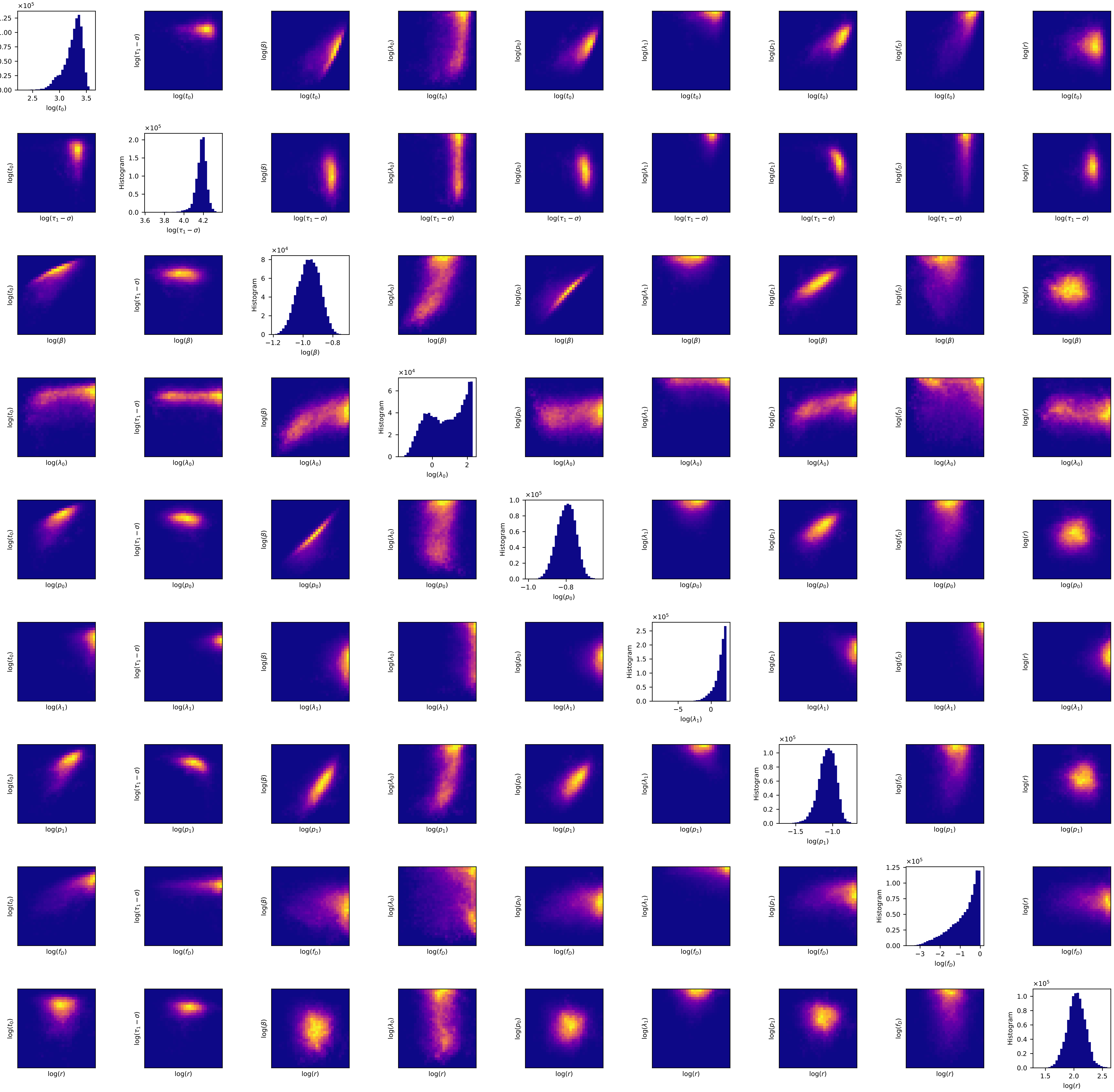
Histogram
log($t_0$)
log($\tau_1 - \sigma$)
log($\beta$)
log($\lambda_0$)
log($p_0$)
log($\lambda_1$)
log($p_1$)
log($f_D$)
log($r$)

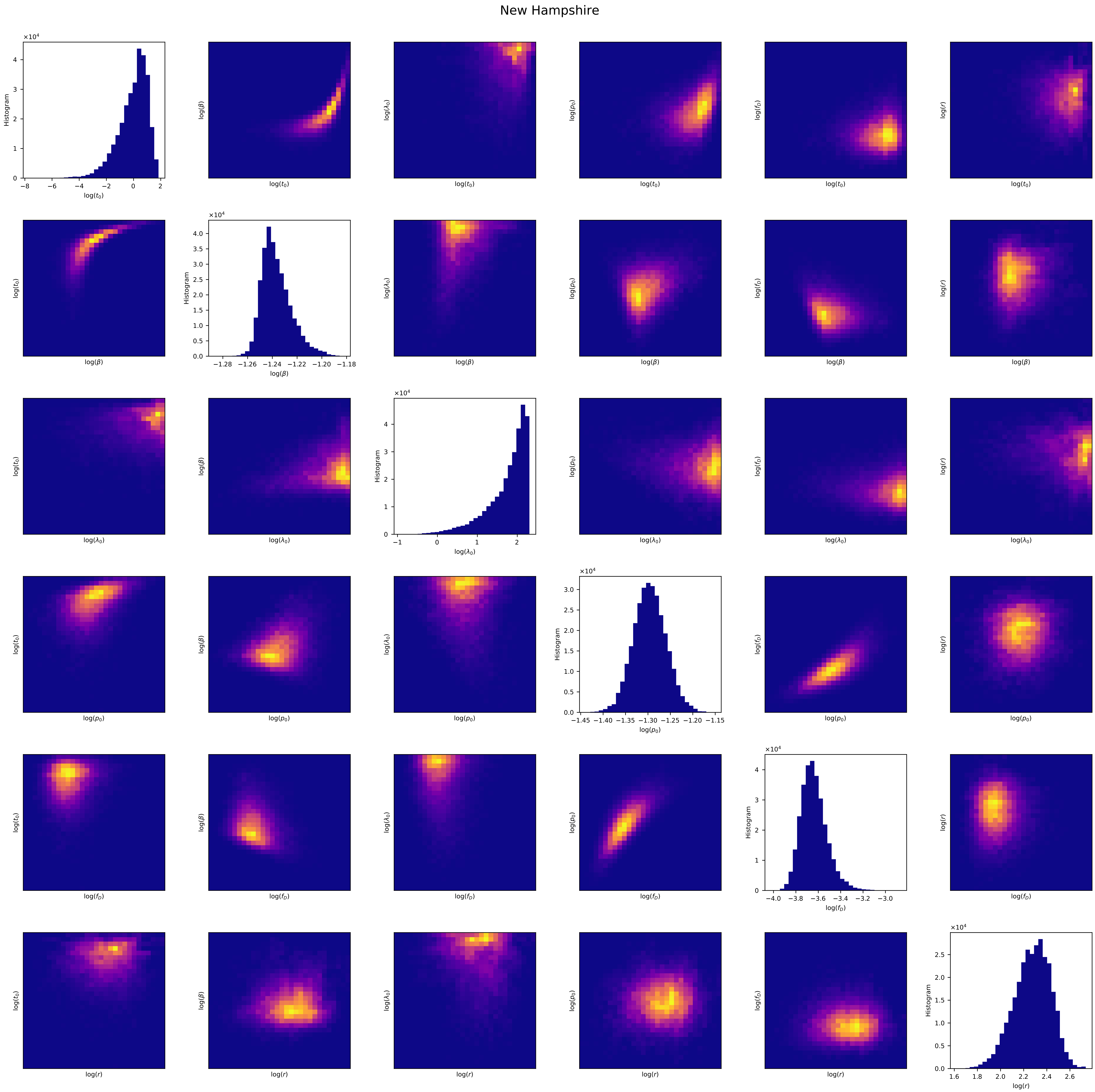
New Hampshire
log($t_0$)
log($\beta$)
log($\lambda_0$)
log($p_0$)
log($f_D$)
log($r$)
Histogram
×10^4

### New Jersey

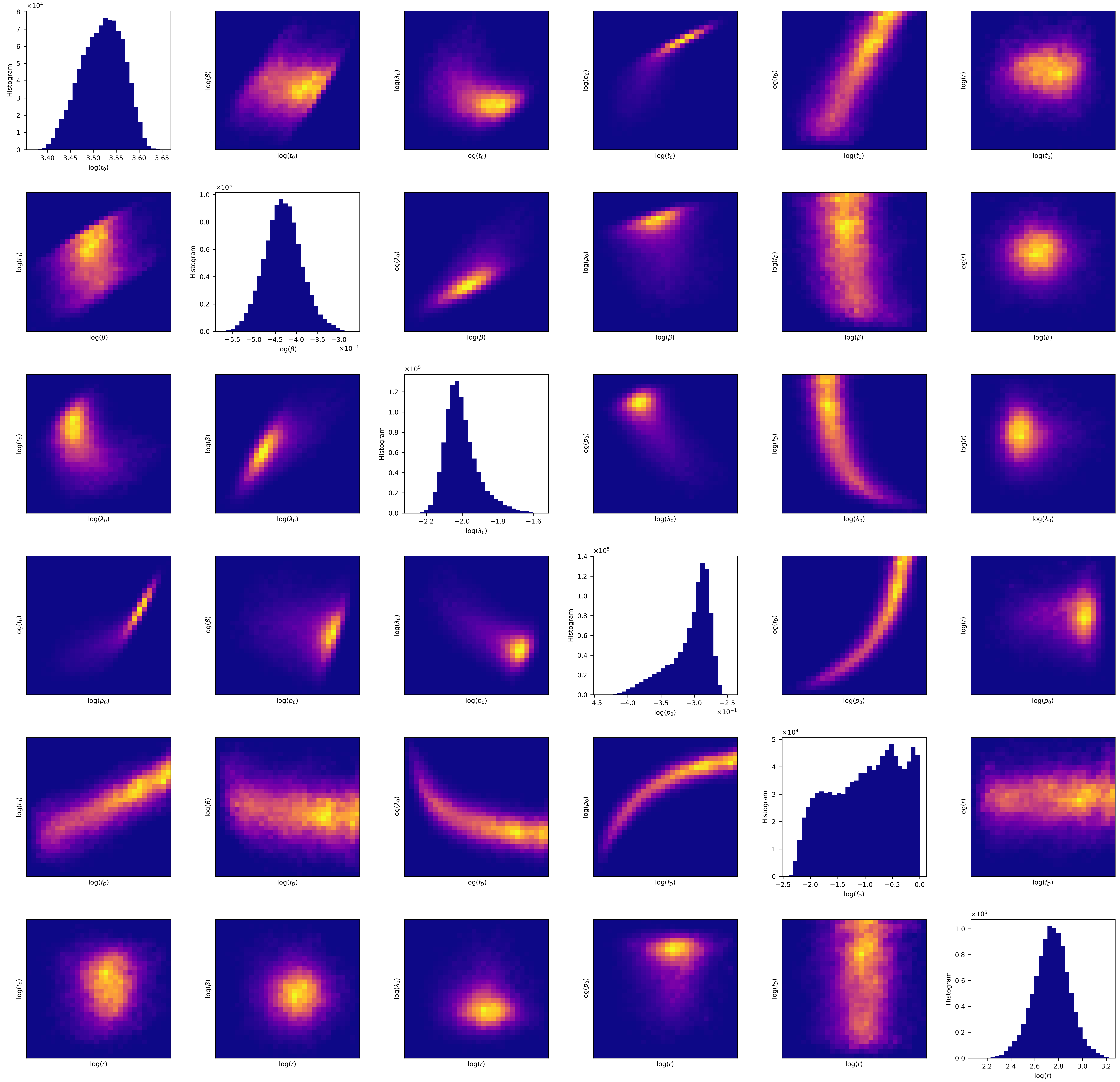

×10^4
Histogram
log($t_0$)
log($\beta$)
log($\lambda_0$)
log($p_0$)
log($f_D$)
log($r$)
×10^5
×10^-1

New Mexico

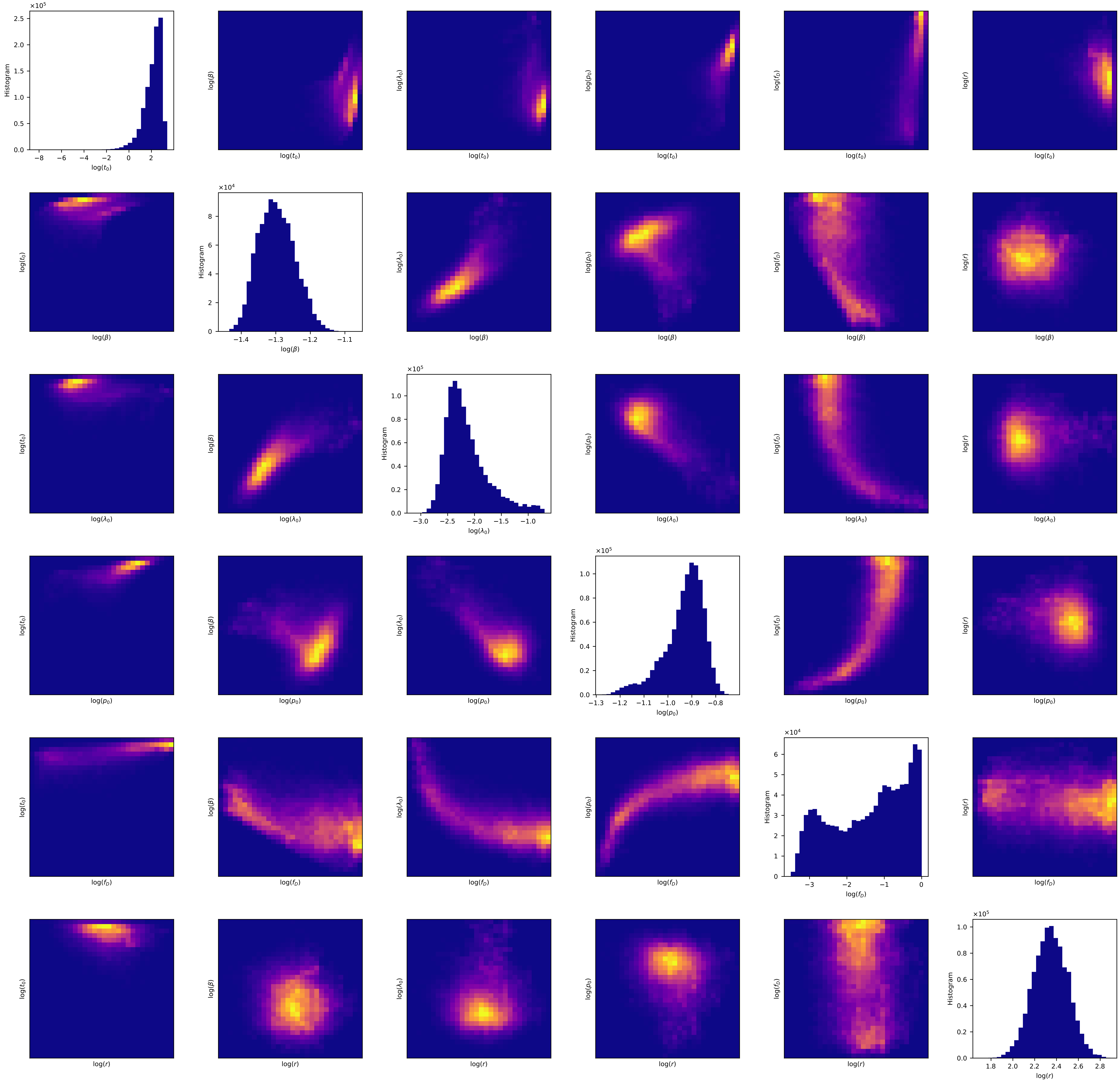

Histogram
log($t_0$)
log($\beta$)
log($\lambda_0$)
log($p_0$)
log($f_D$)
log($r$)
$\times 10^5$
$\times 10^4$

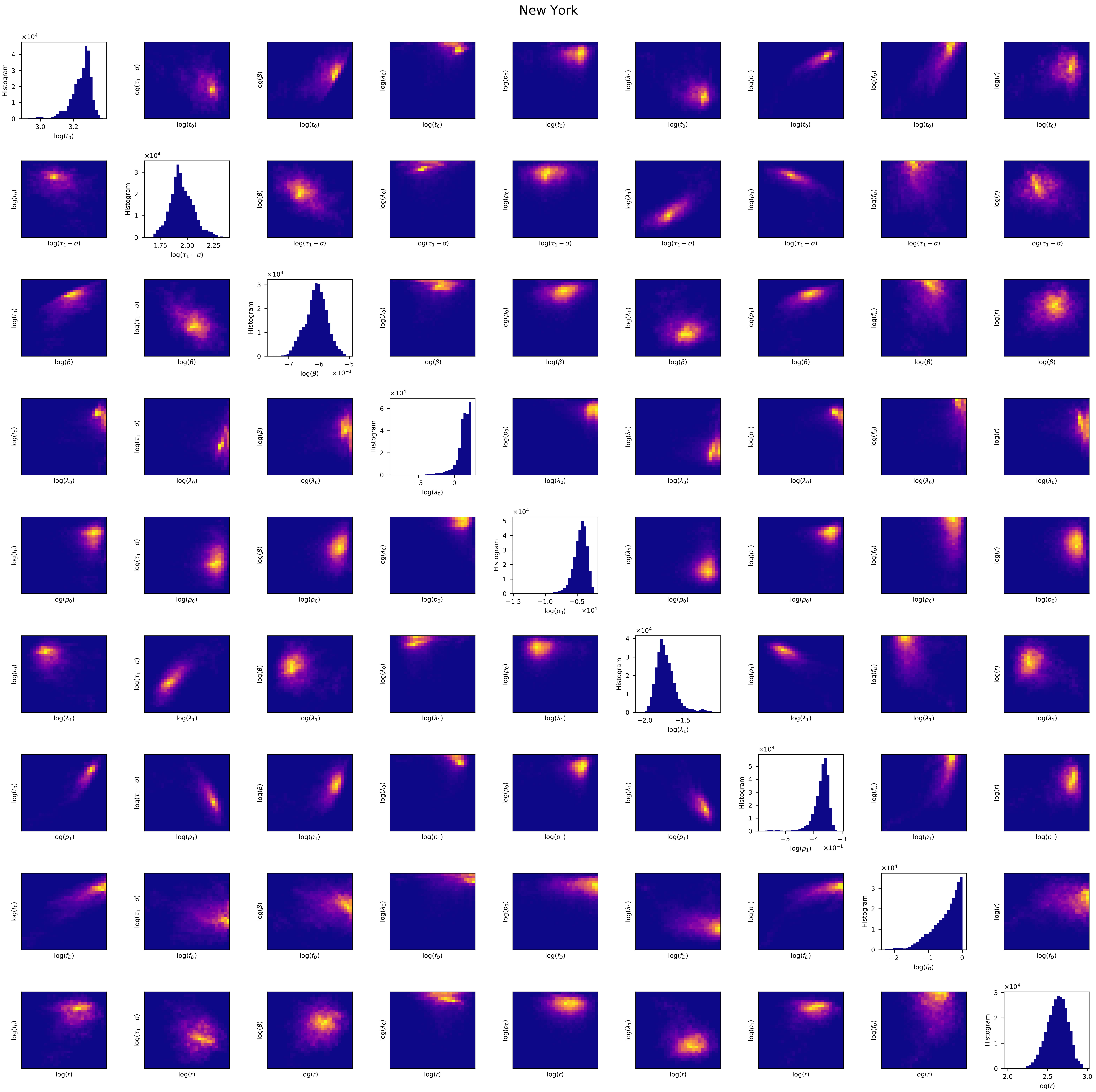
New York
Histogram
$\log(t_0)$
$\log(\tau_1 - \sigma)$
$\log(\beta)$
$\log(\lambda_0)$
$\log(p_0)$
$\log(\lambda_1)$
$\log(p_1)$
$\log(f_D)$
$\log(r)$

North Carolina

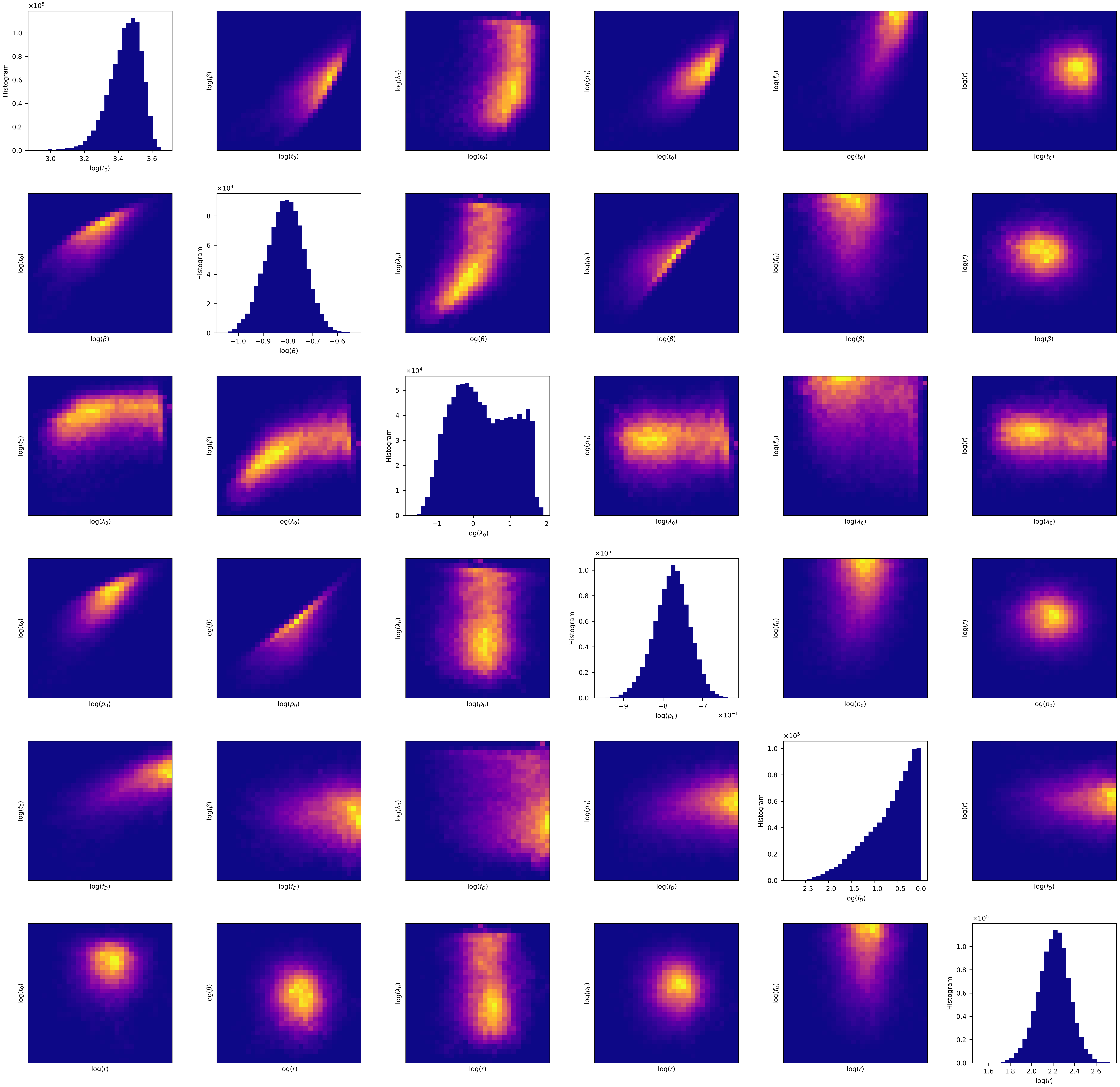

### North Dakota

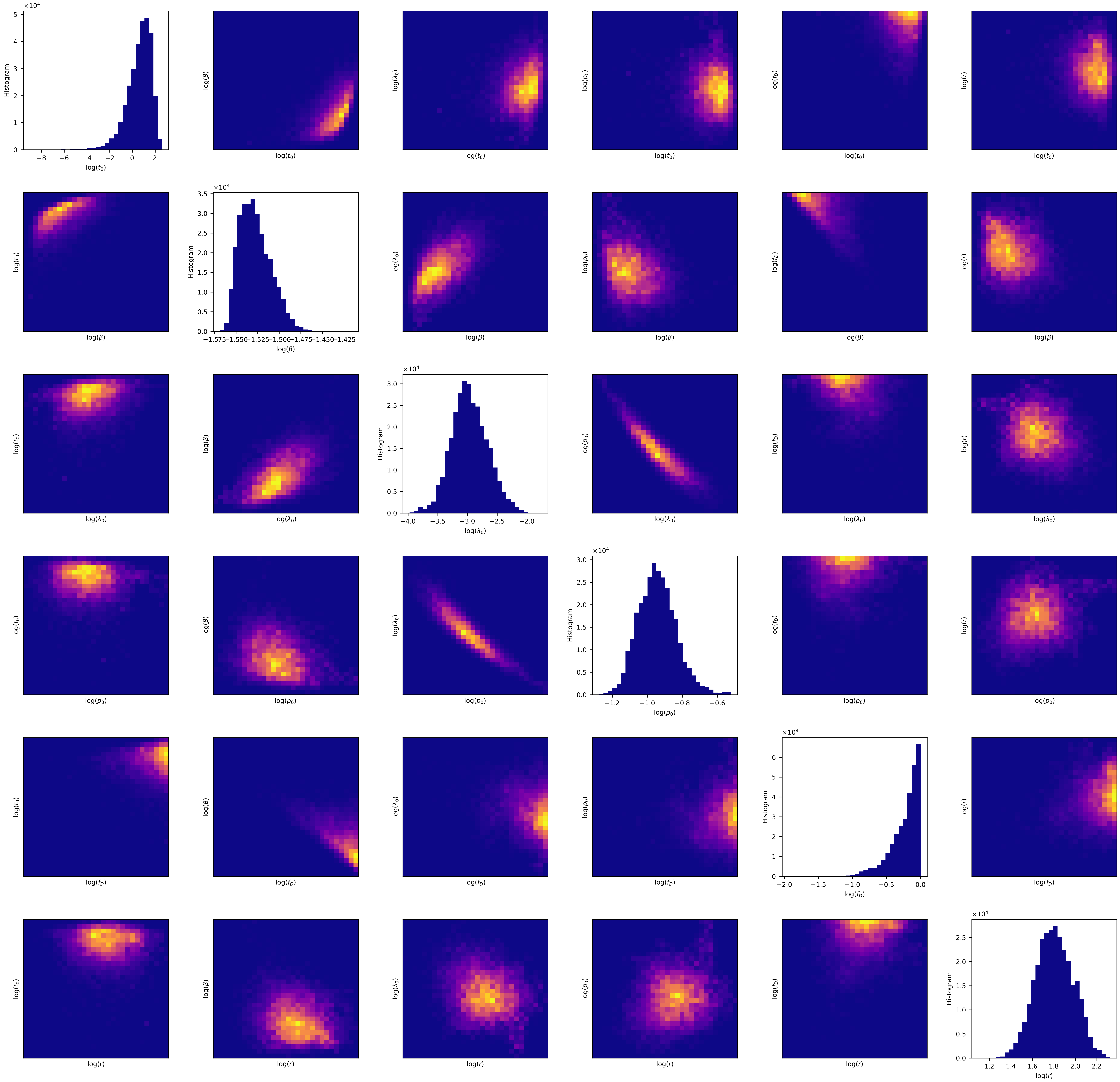

### Ohio

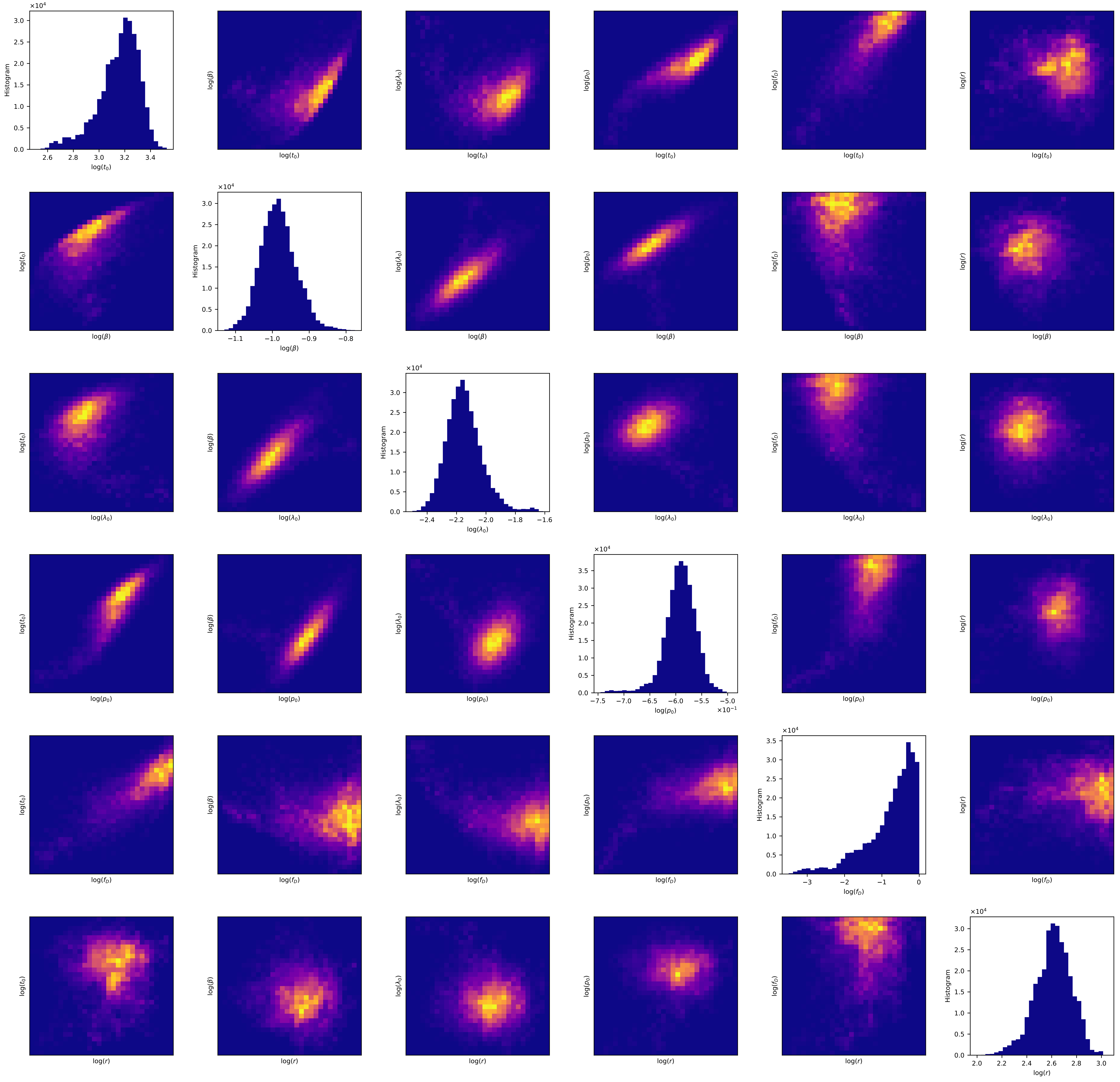

Histogram
×10^4
log($t_0$)
log($\beta$)
log($\lambda_0$)
log($p_0$)
log($f_D$)
log($r$)
×10^−1

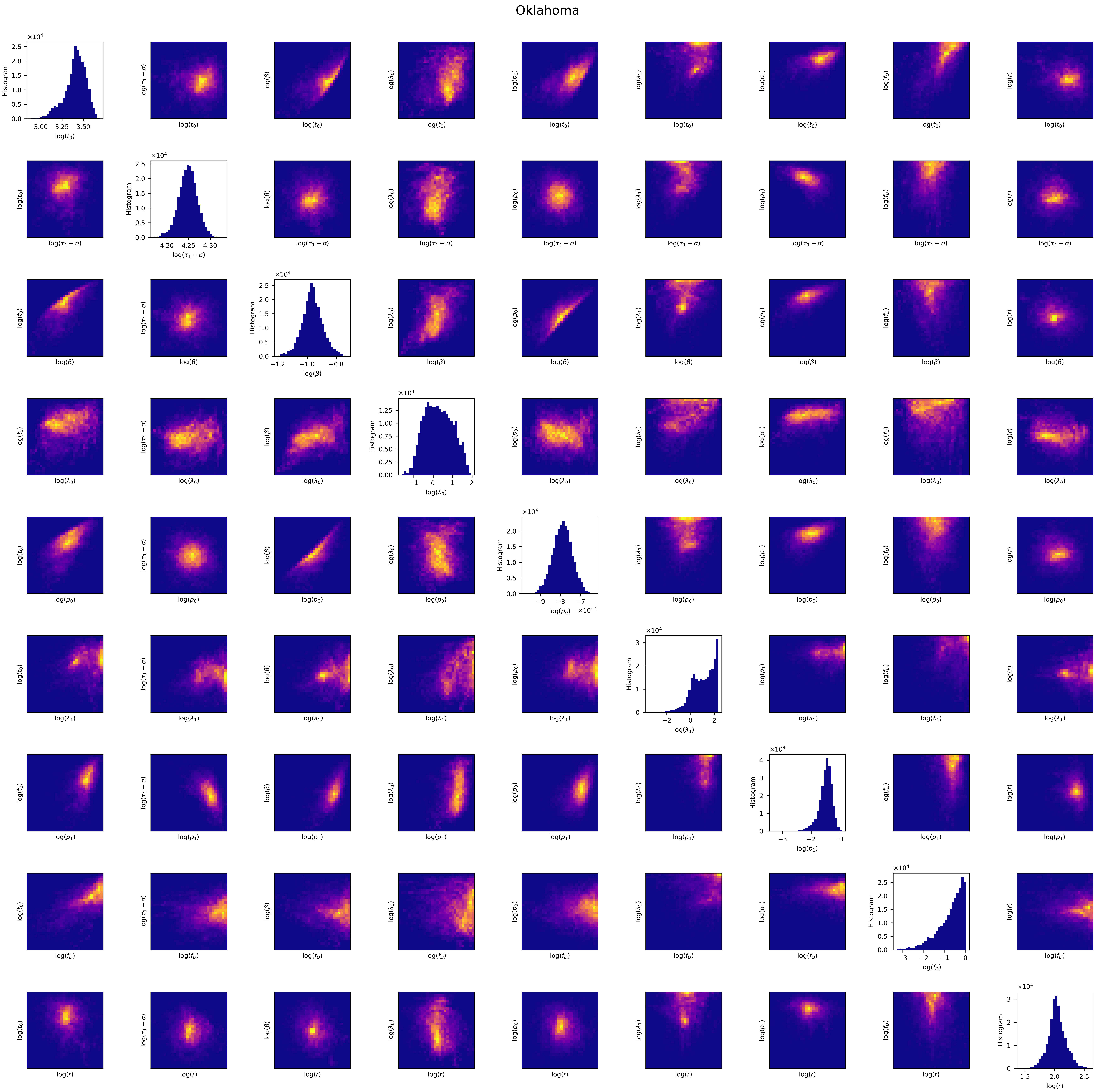
Oklahoma
Histogram
log($t_0$)
log($\tau_1-\sigma$)
log($\beta$)
log($\lambda_0$)
log($p_0$)
log($\lambda_1$)
log($p_1$)
log($f_D$)
log($r$)

Oregon

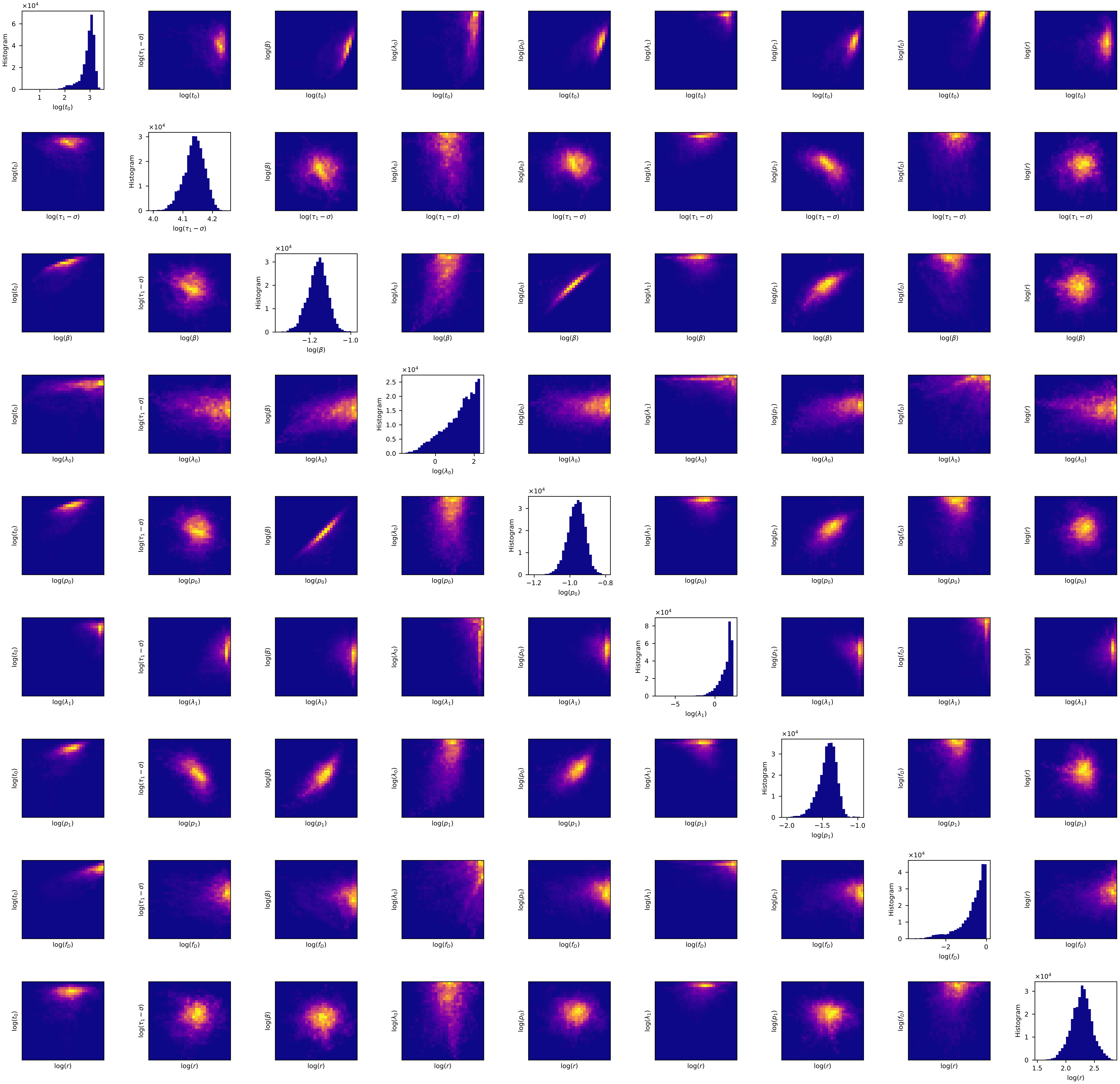

Histogram
×10^4
log($t_0$)
log($\tau_1-\sigma$)
log($\beta$)
log($\lambda_0$)
log($p_0$)
log($\lambda_1$)
log($p_1$)
log($f_D$)
log($r$)

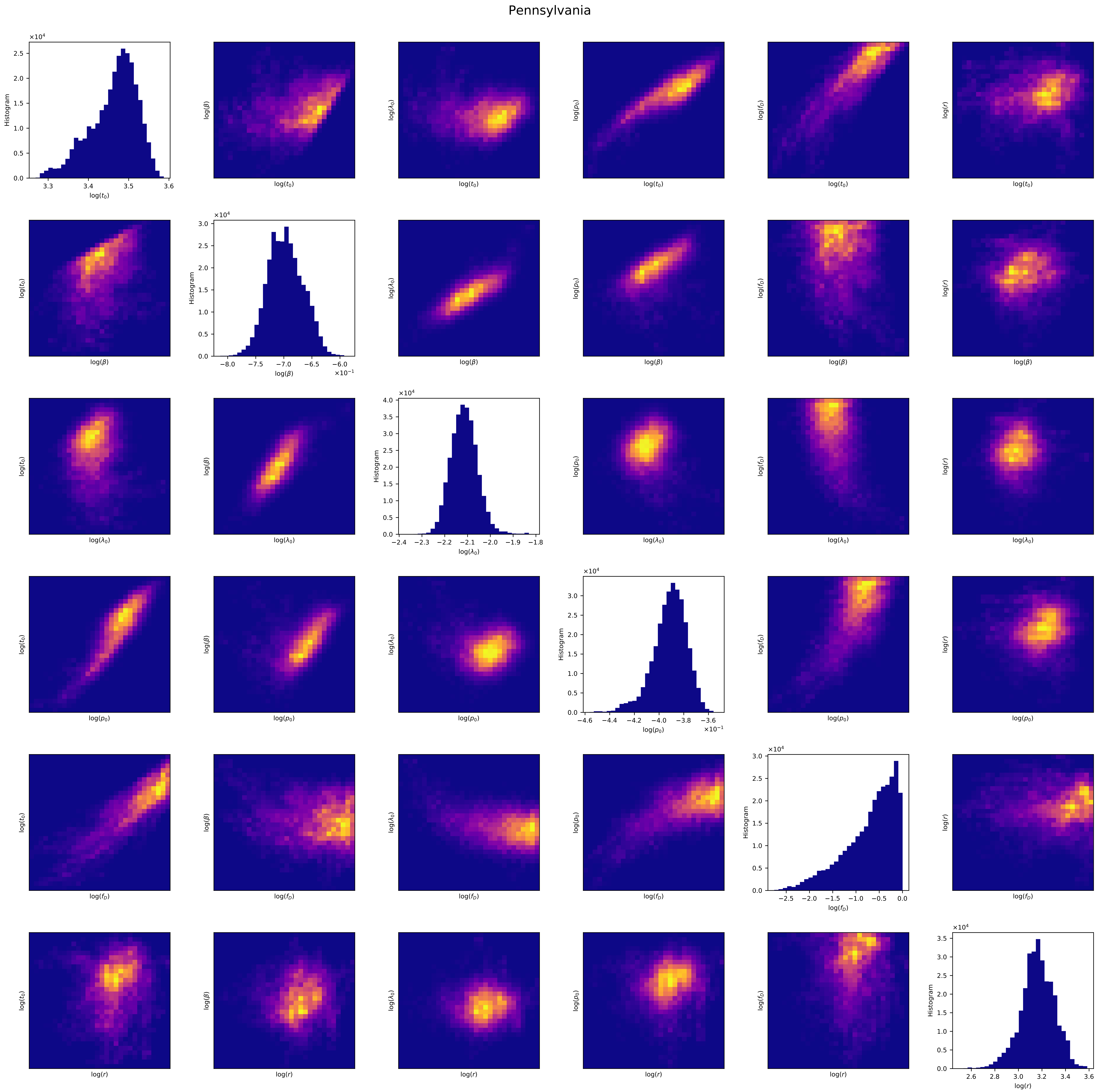
Pennsylvania
Histogram
$\log(t_0)$
$\log(\beta)$
$\log(\lambda_0)$
$\log(p_0)$
$\log(f_D)$
$\log(r)$
$\times 10^4$
$\times 10^{-1}$

### Rhode Island

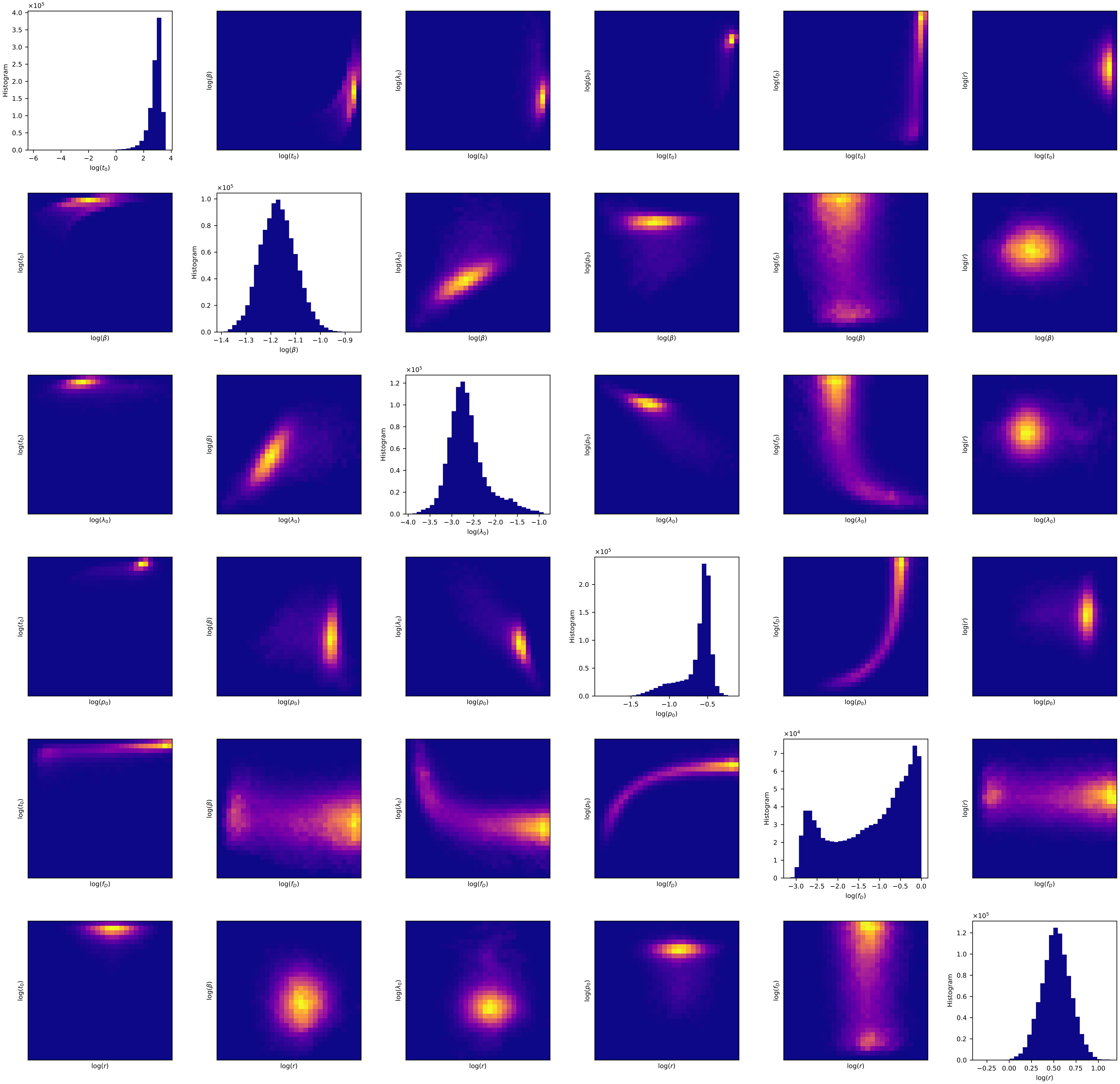

South Carolina

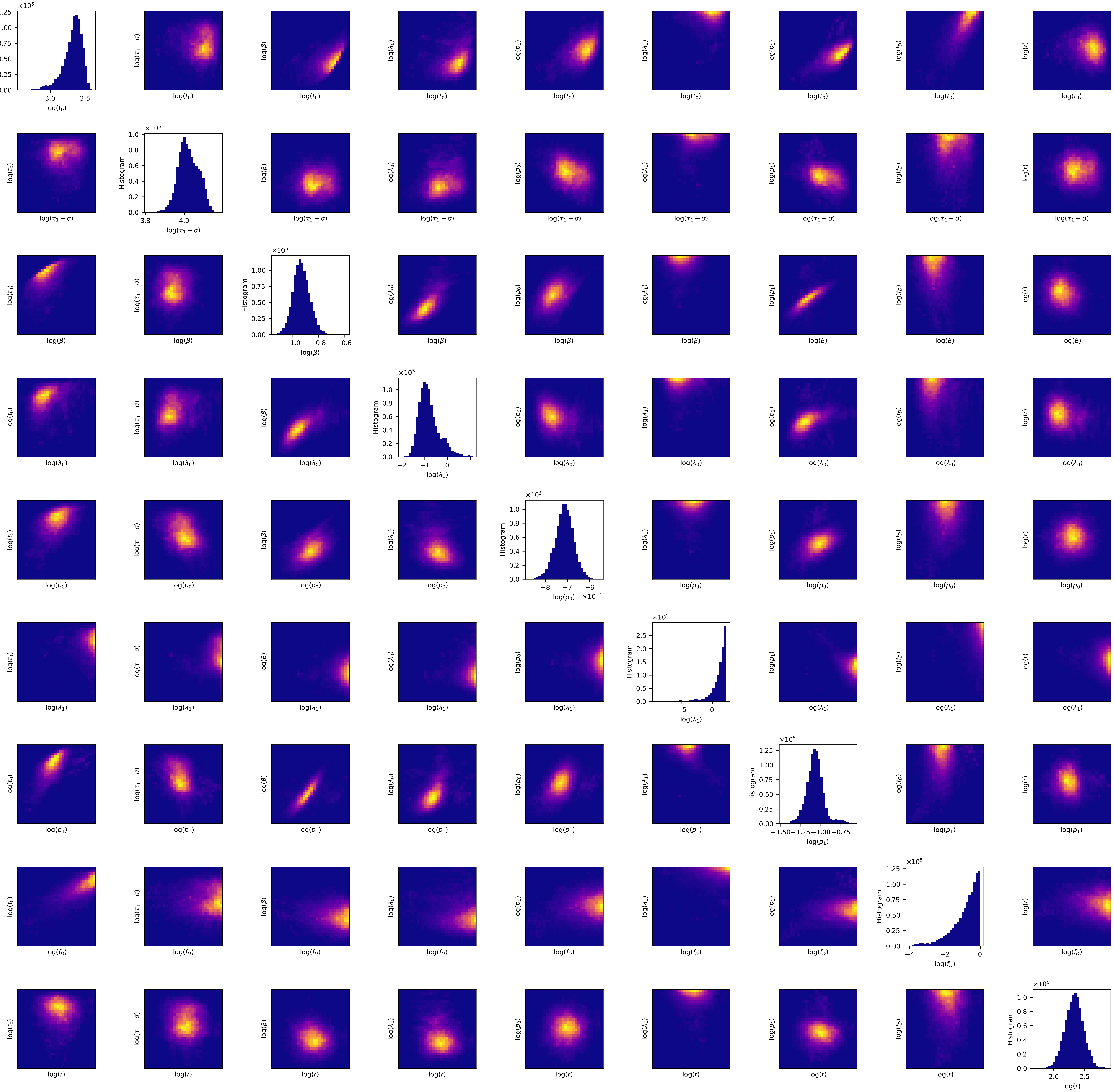

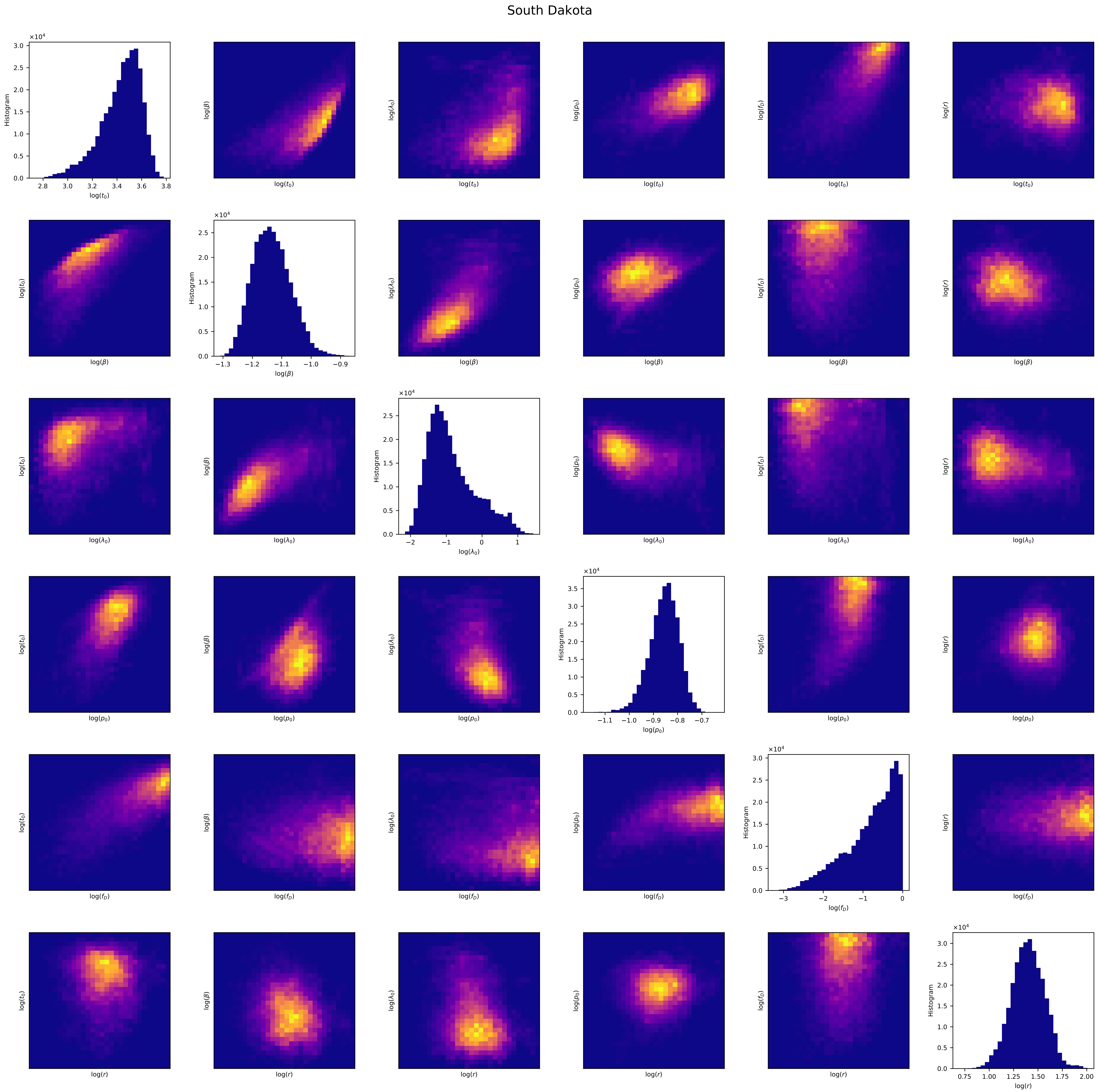
South Dakota
Histogram
log(t0)
log(β)
log(λ0)
log(p0)
log(fD)
log(r)

Tennessee

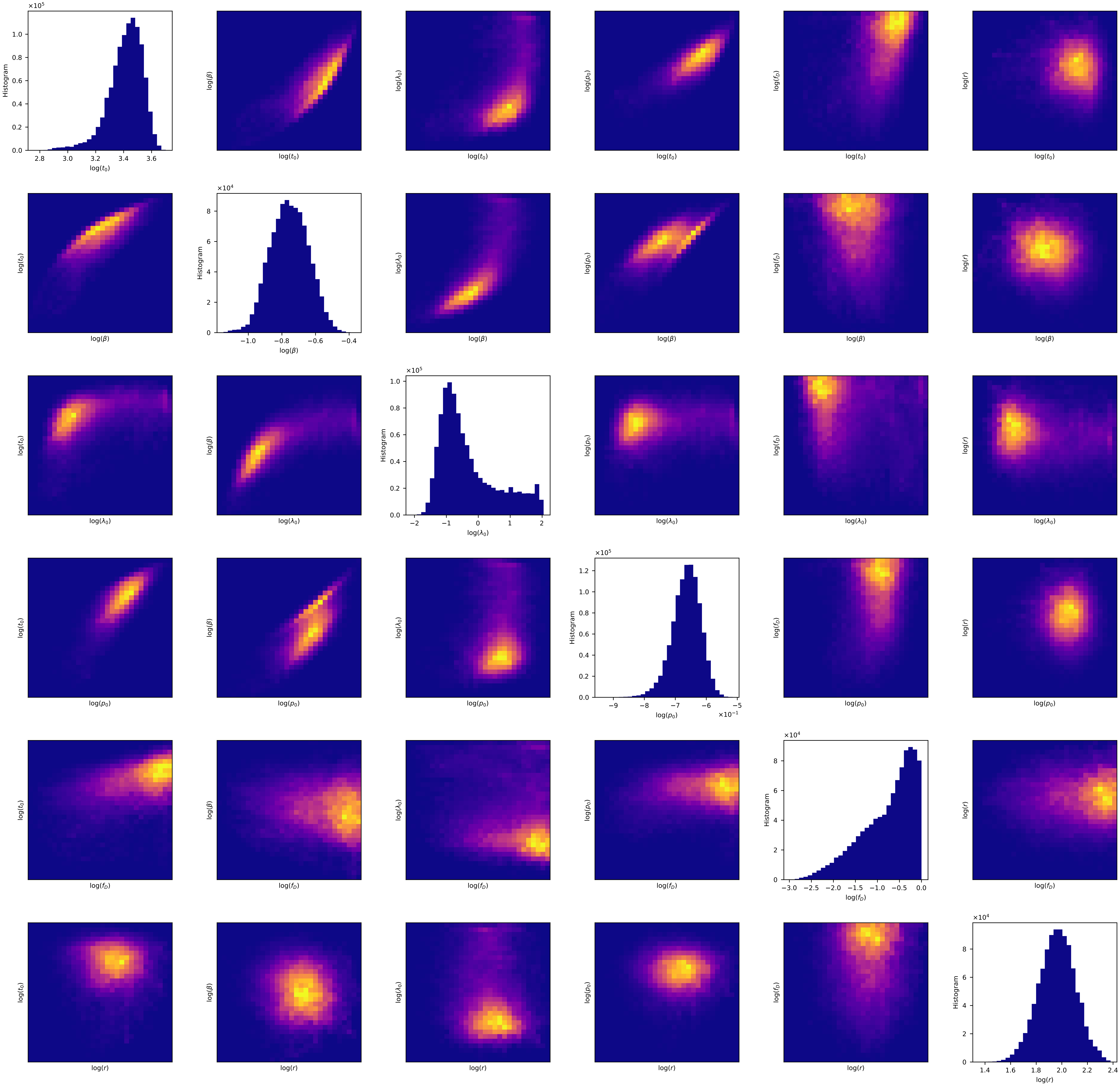
log($t_0$)
log($\beta$)
log($\lambda_0$)
log($p_0$)
log($f_D$)
log($r$)
Histogram
×10^5
×10^4
×10^-1

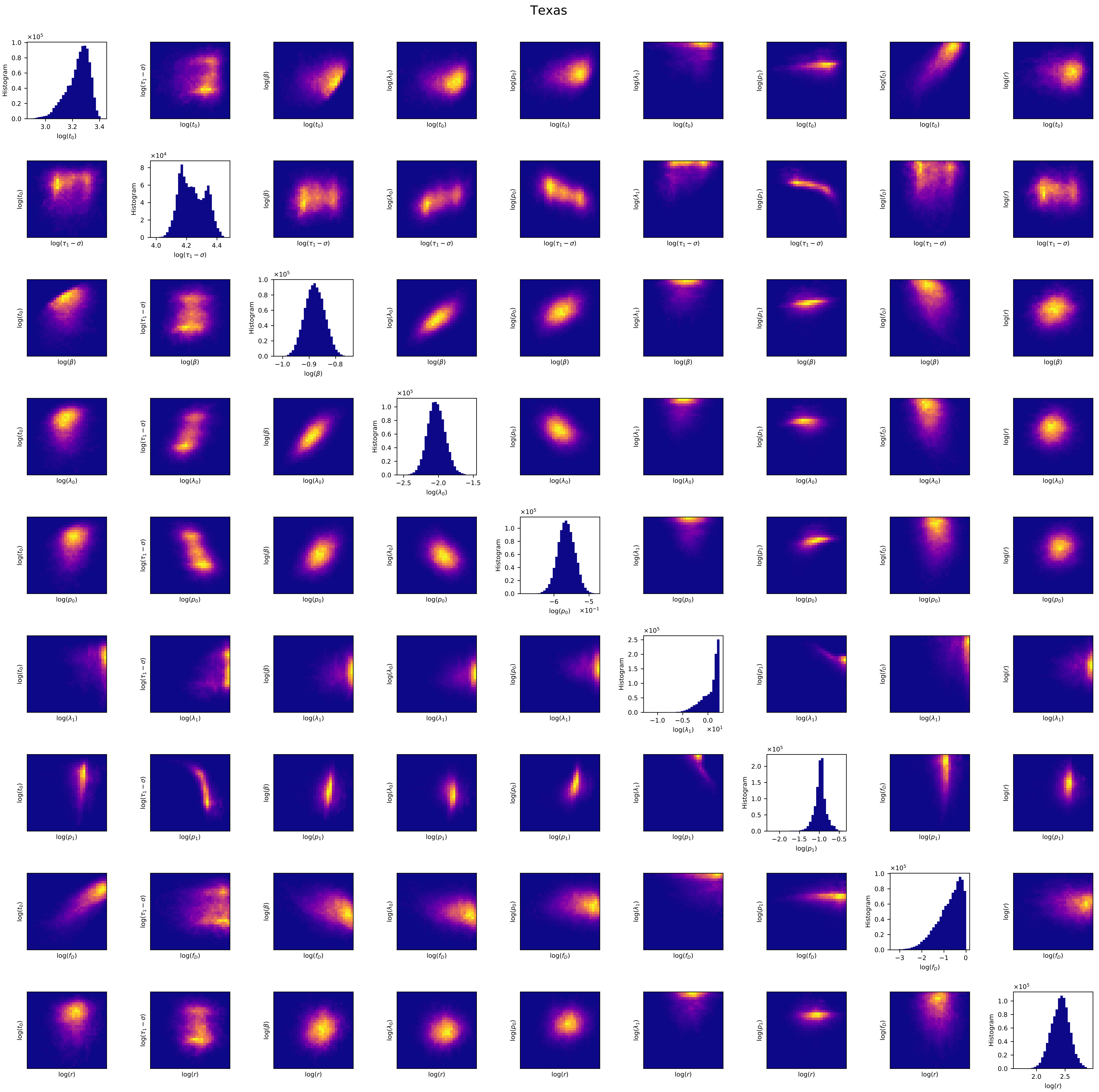
Texas
Histogram
log($t_0$)
log($\tau_1-\sigma$)
log($\beta$)
log($\lambda_0$)
log($p_0$)
log($\lambda_1$)
log($p_1$)
log($f_D$)
log($r$)

### Utah

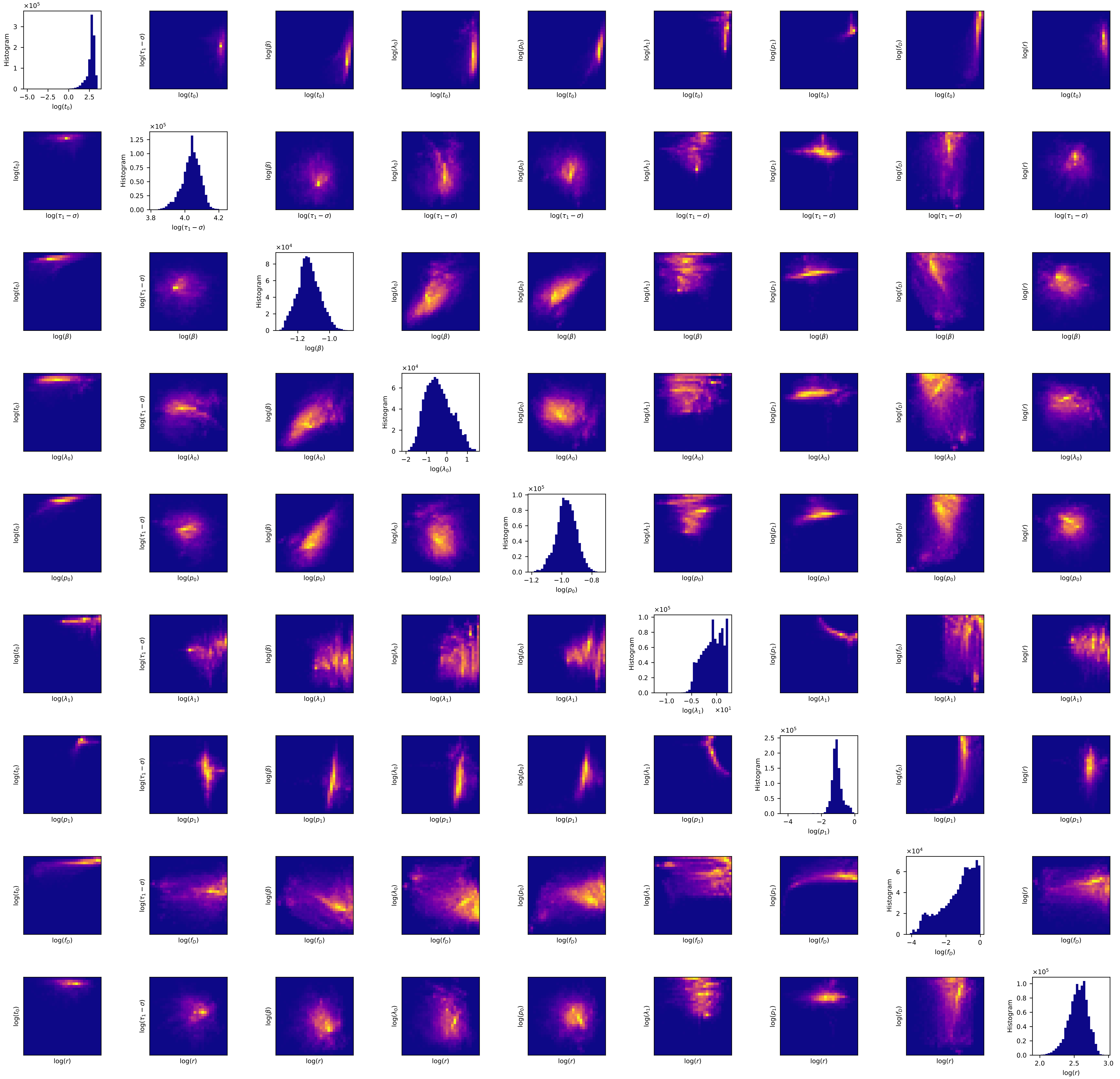
Histogram
log($t_0$)
log($\tau_1-\sigma$)
log($\beta$)
log($\lambda_0$)
log($p_0$)
log($\lambda_1$)
log($p_1$)
log($f_D$)
log($r$)
×10^5
×10^4
×10^1

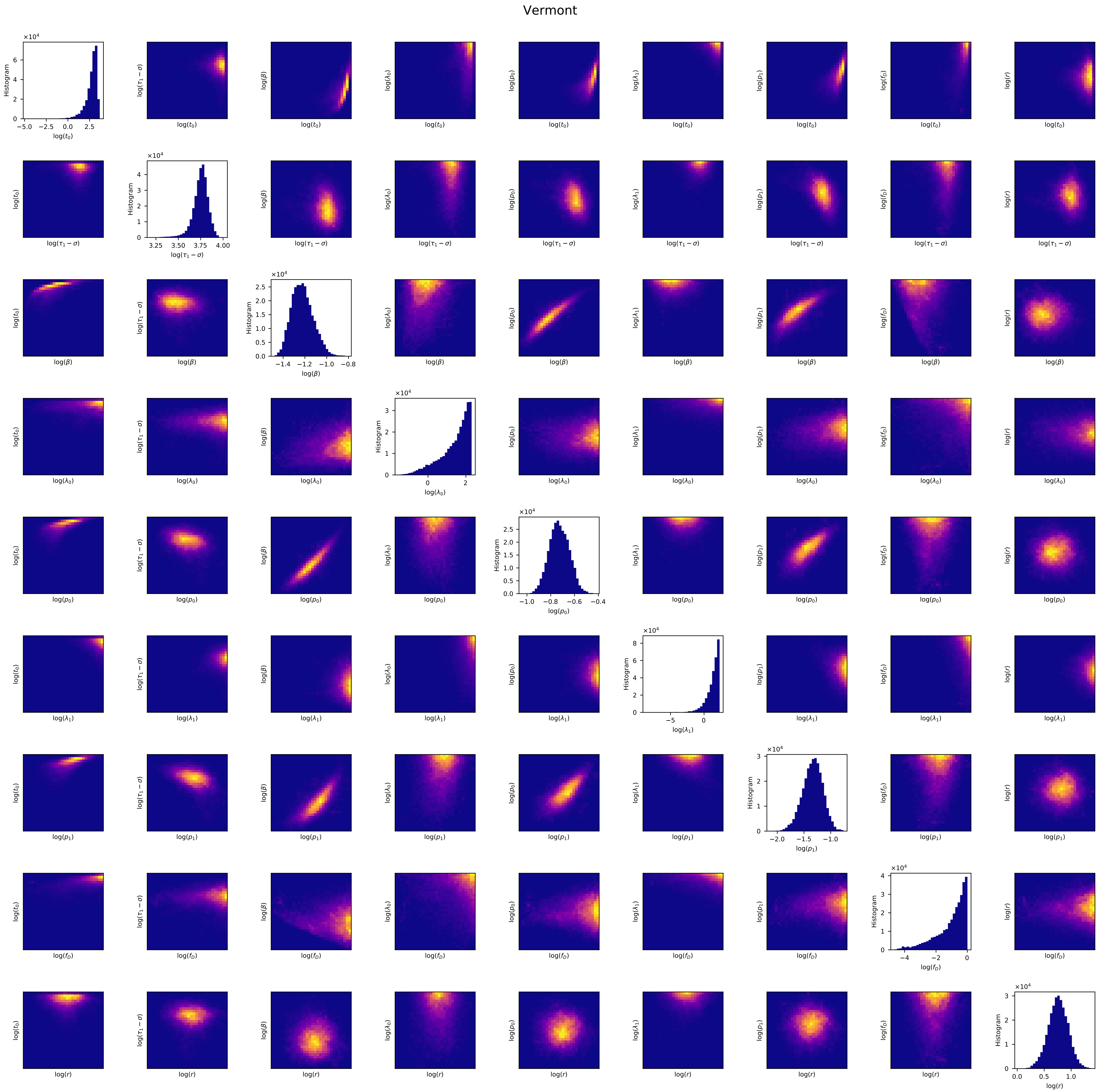
Vermont
Histogram
log($t_0$)
log($\tau_1 - \sigma$)
log($\beta$)
log($\lambda_0$)
log($p_0$)
log($\lambda_1$)
log($p_1$)
log($f_D$)
log($r$)

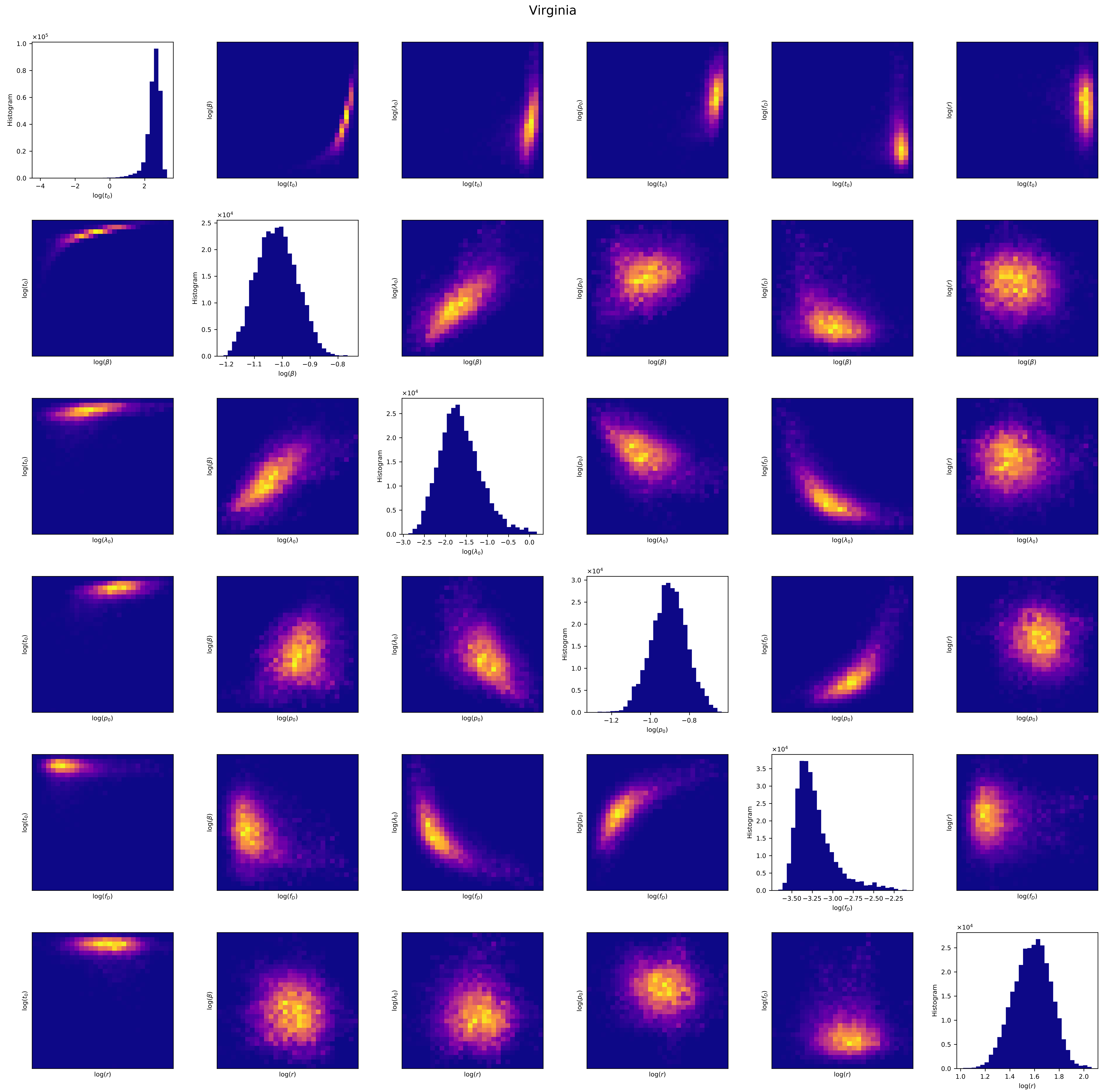

Virginia
Histogram
log($t_0$)
log($\beta$)
log($\lambda_0$)
log($p_0$)
log($f_D$)
log($r$)

#### Washington

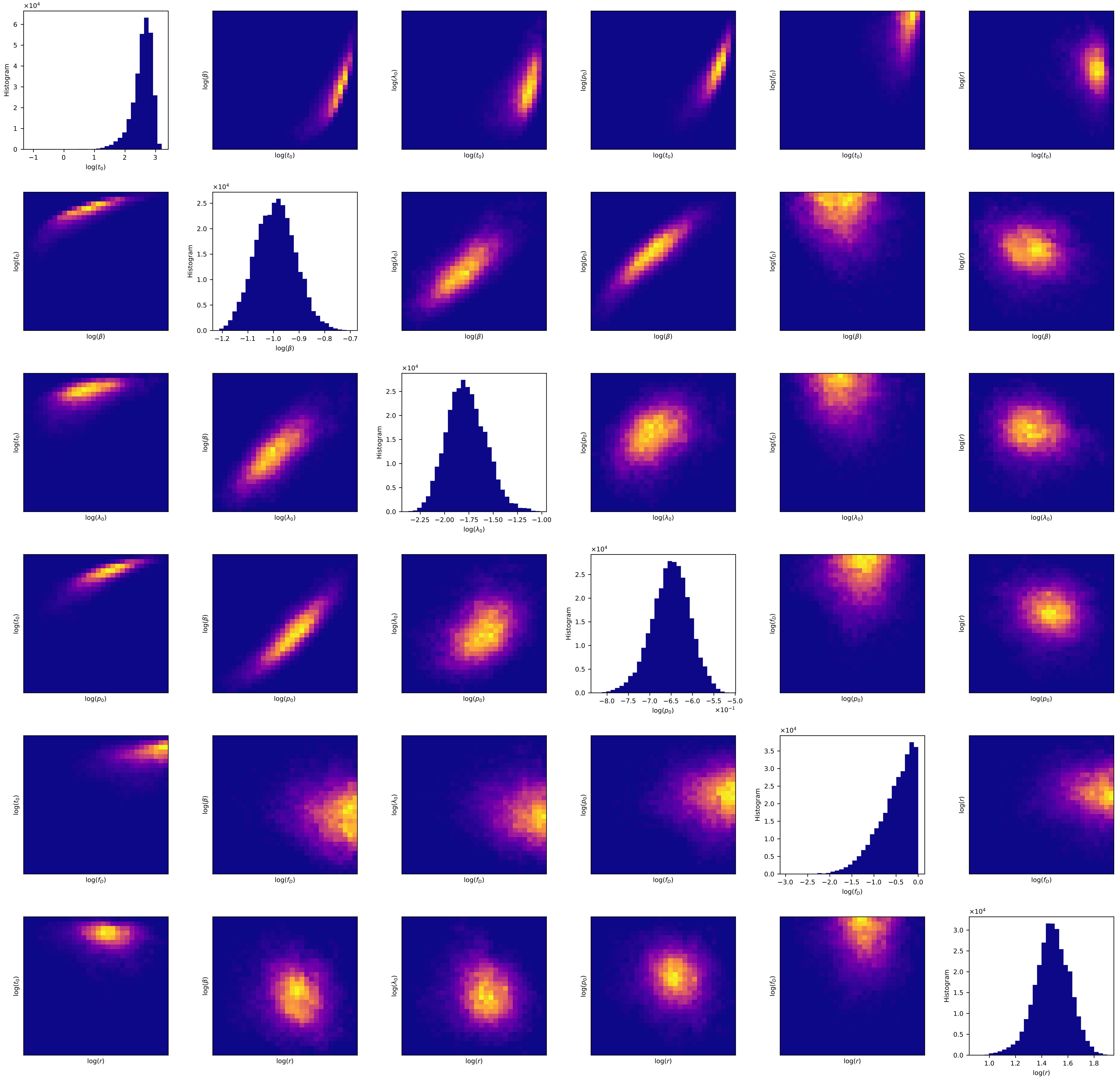

West Virginia

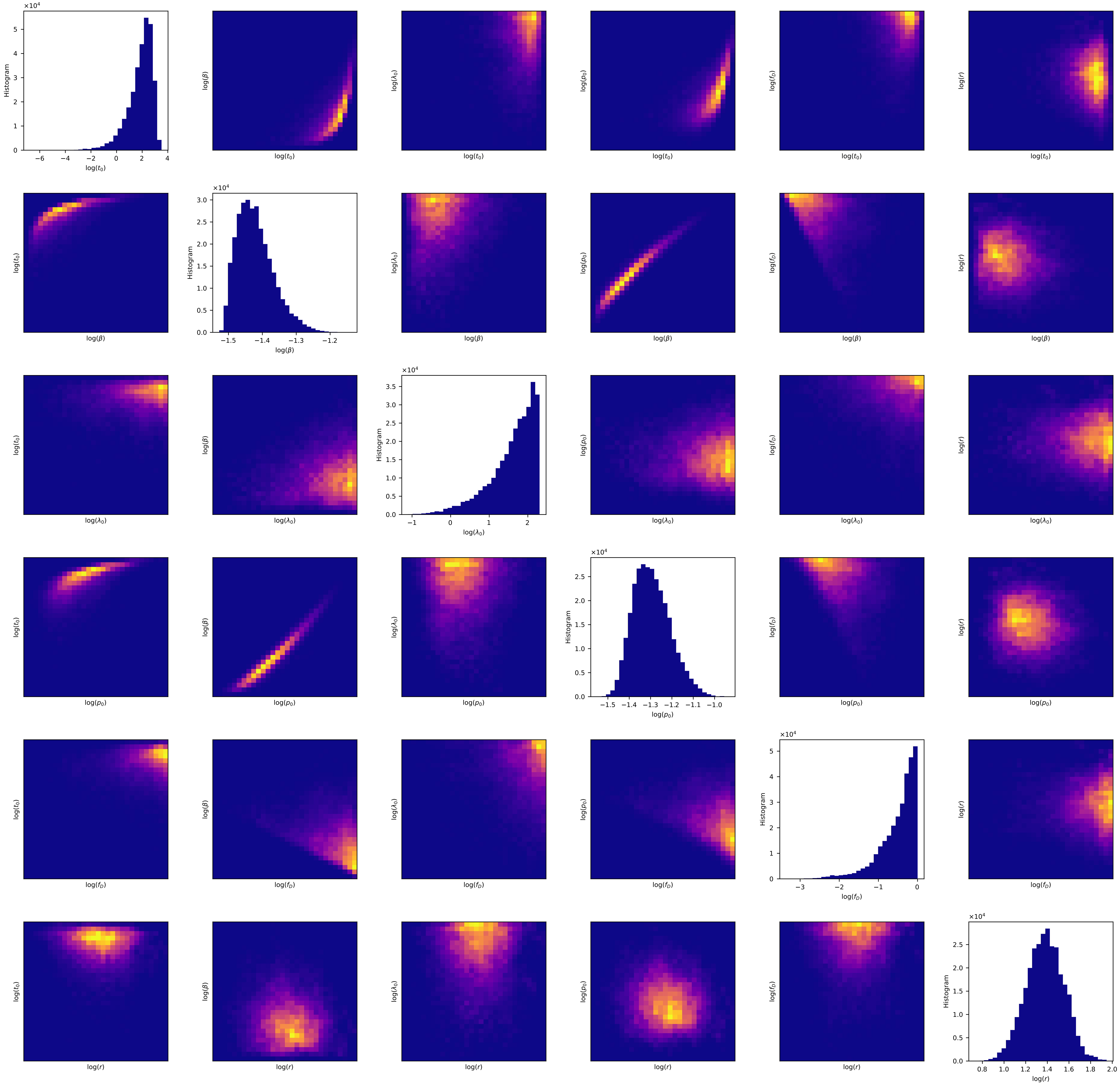

Wisconsin

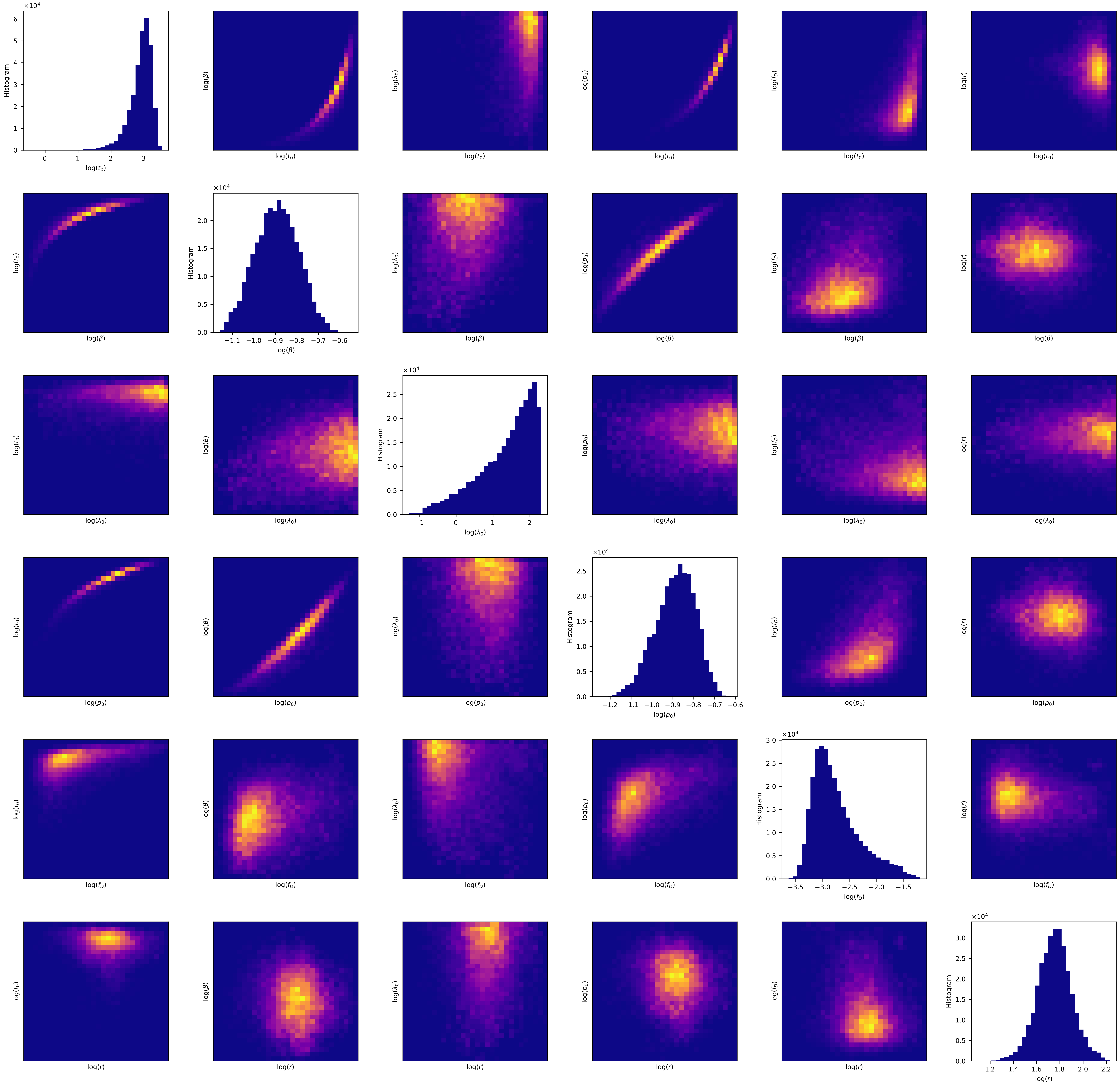

Wyoming

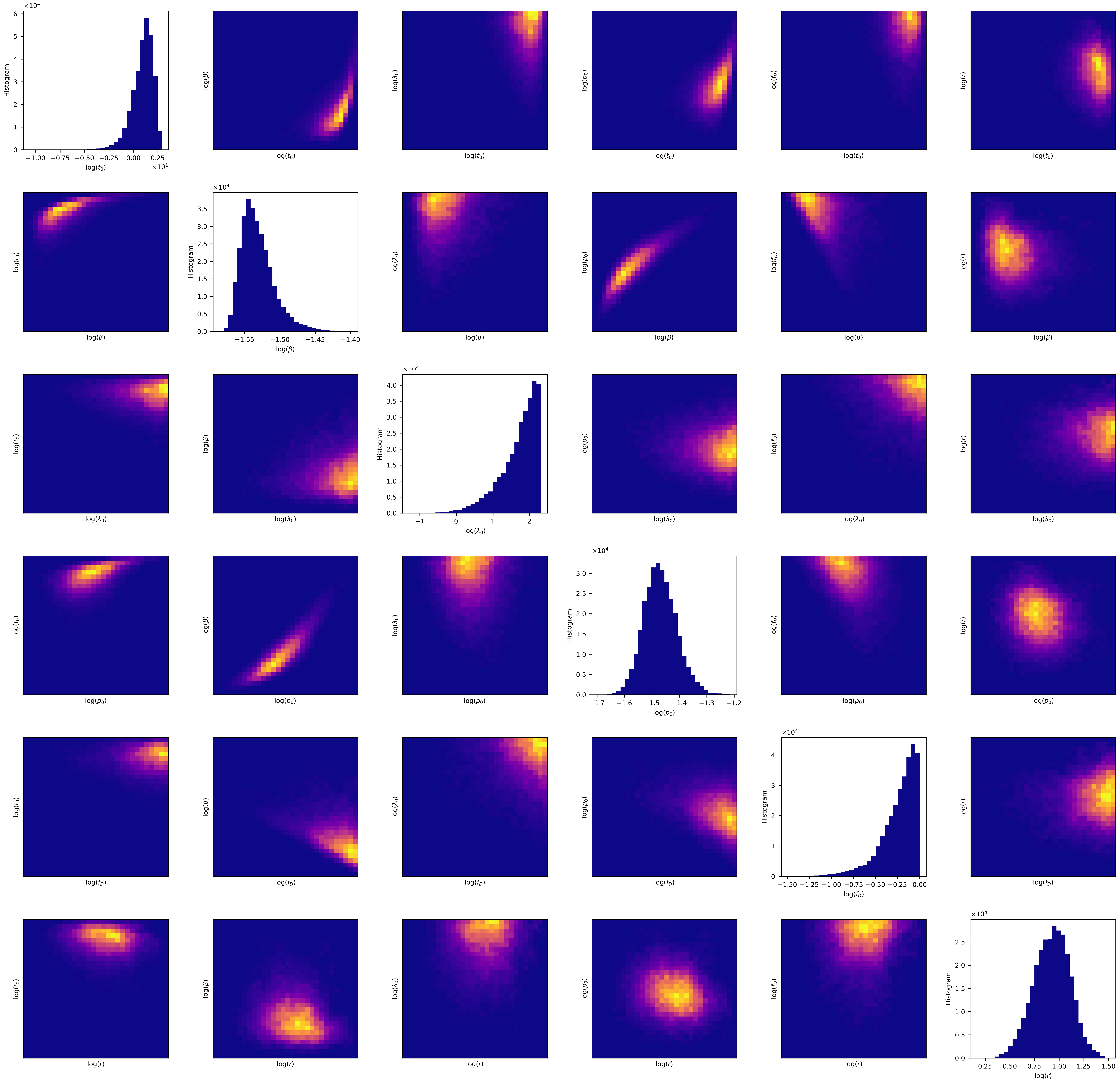
Histogram
log($t_0$)
log($\beta$)
log($\lambda_0$)
log($p_0$)
log($f_D$)
log($r$)
×10^4
×10^1

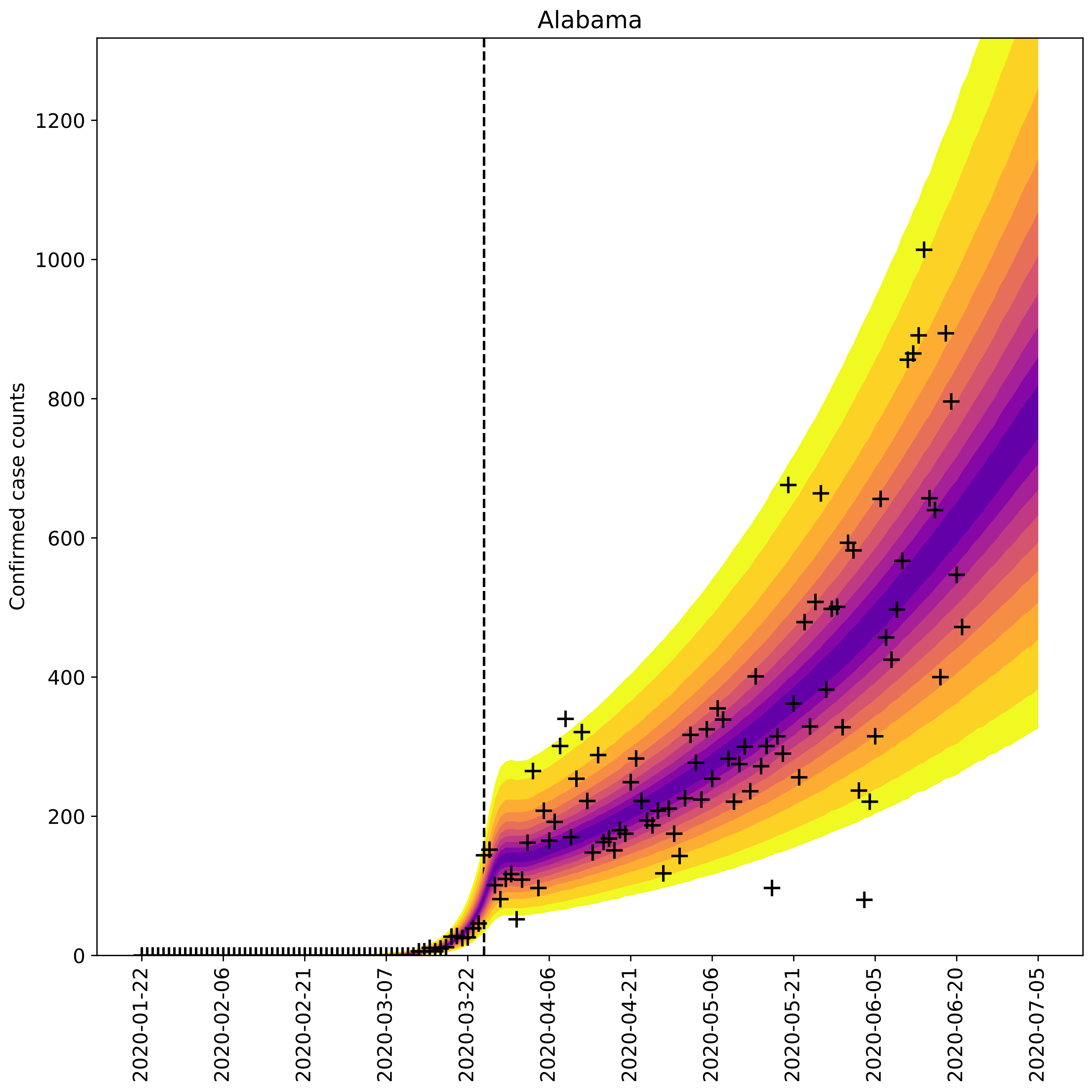

Alabama
Confirmed case counts
0
200
400
600
800
1000
1200
2020-01-22
2020-02-06
2020-02-21
2020-03-07
2020-03-22
2020-04-06
2020-04-21
2020-05-06
2020-05-21
2020-06-05
2020-06-20
2020-07-05

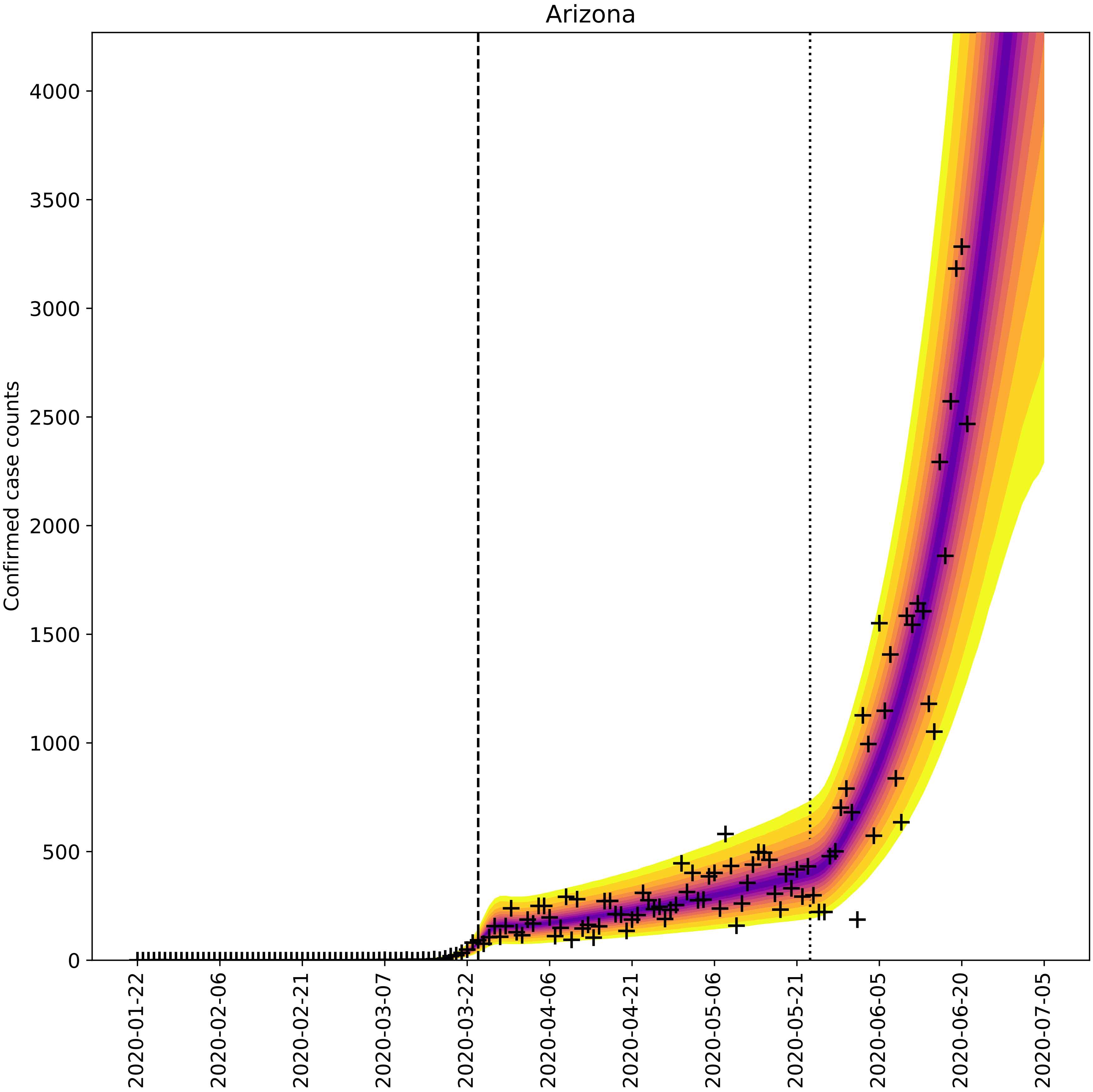
Arizona
Confirmed case counts
0
500
1000
1500
2000
2500
3000
3500
4000
2020-01-22
2020-02-06
2020-02-21
2020-03-07
2020-03-22
2020-04-06
2020-04-21
2020-05-06
2020-05-21
2020-06-05
2020-06-20
2020-07-05

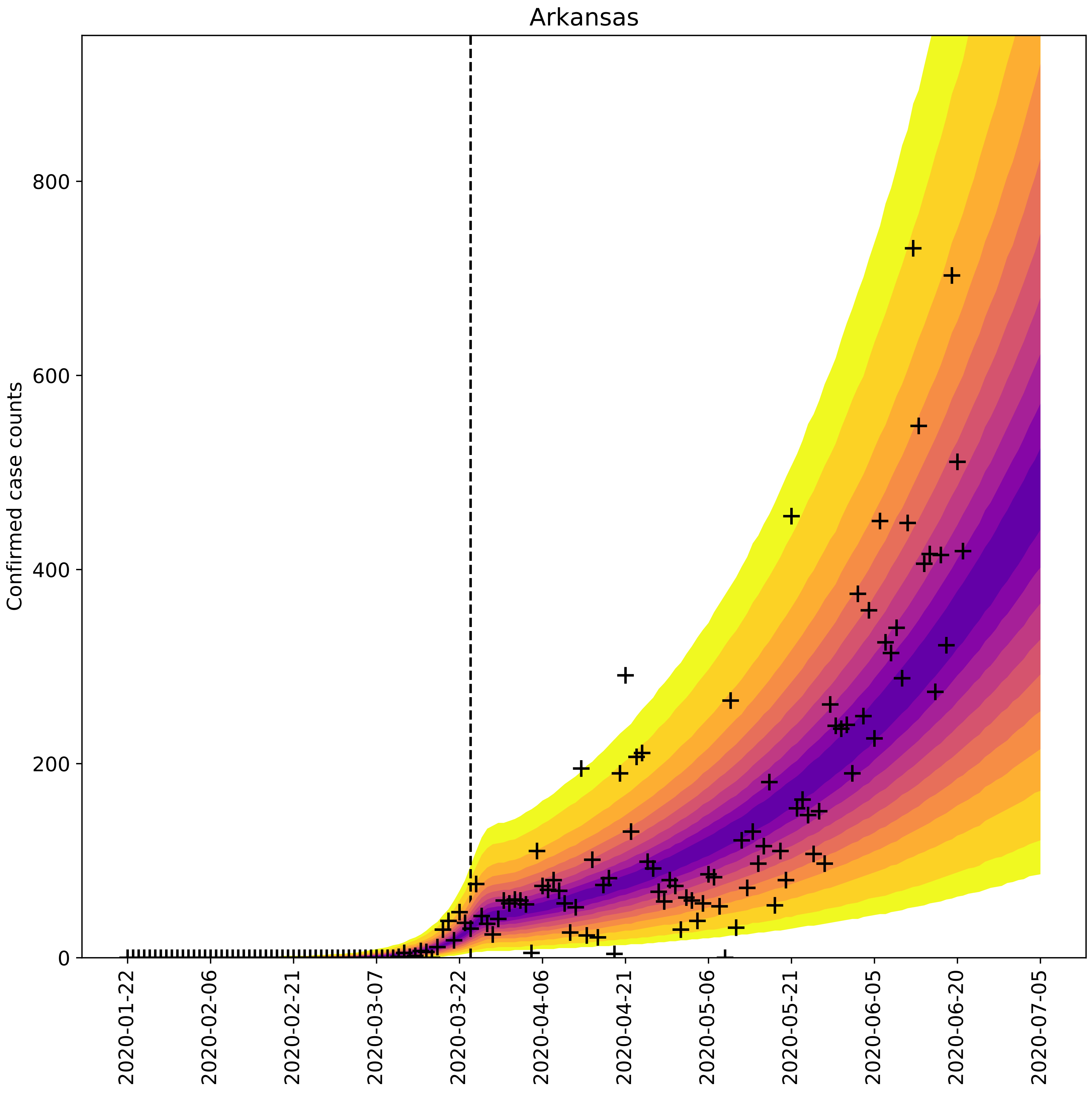

Arkansas
Confirmed case counts
0
200
400
600
800
2020-01-22
2020-02-06
2020-02-21
2020-03-07
2020-03-22
2020-04-06
2020-04-21
2020-05-06
2020-05-21
2020-06-05
2020-06-20
2020-07-05

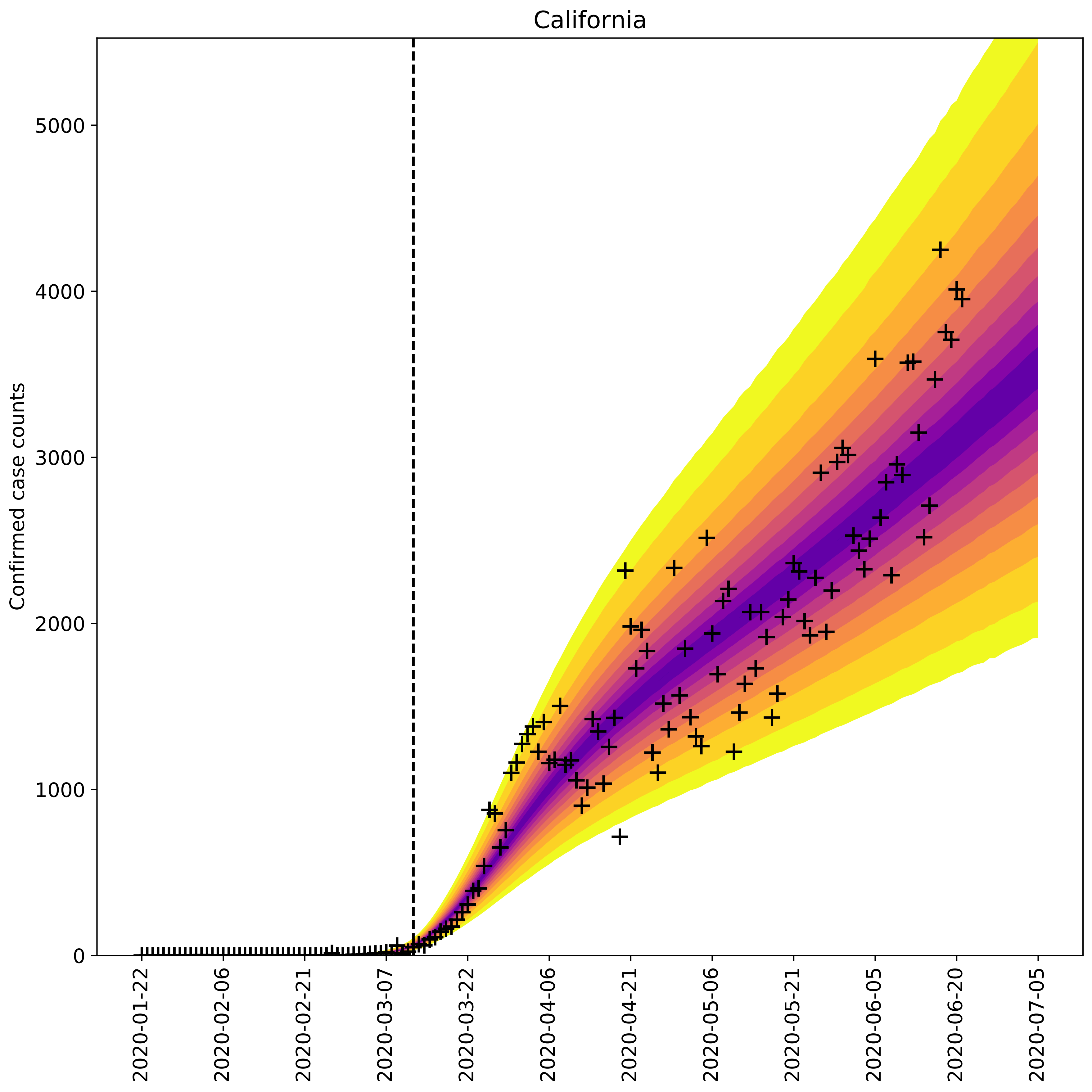

California
Confirmed case counts
0
1000
2000
3000
4000
5000
2020-01-22
2020-02-06
2020-02-21
2020-03-07
2020-03-22
2020-04-06
2020-04-21
2020-05-06
2020-05-21
2020-06-05
2020-06-20
2020-07-05

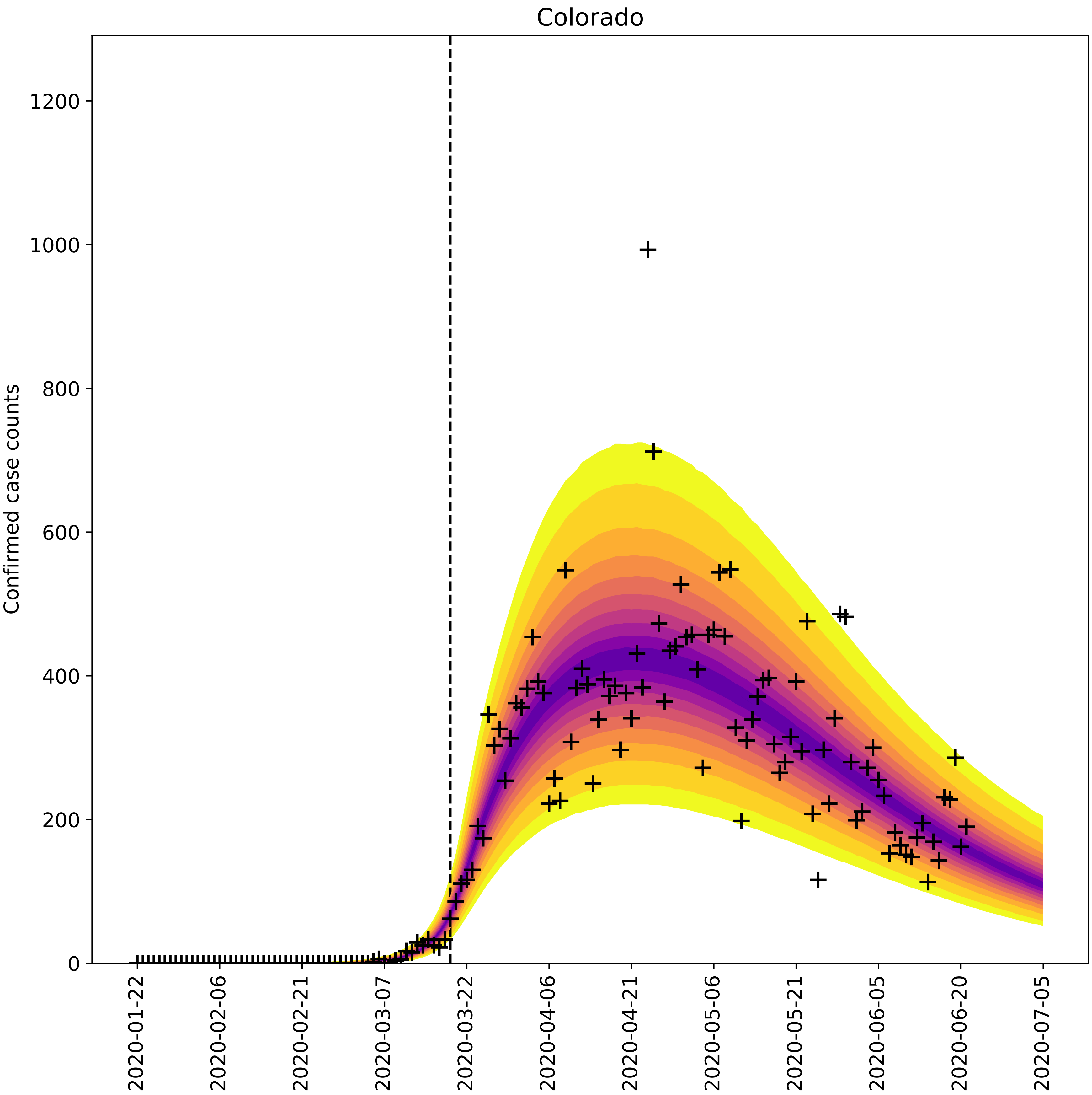

Colorado
Confirmed case counts
0
200
400
600
800
1000
1200
2020-01-22
2020-02-06
2020-02-21
2020-03-07
2020-03-22
2020-04-06
2020-04-21
2020-05-06
2020-05-21
2020-06-05
2020-06-20
2020-07-05

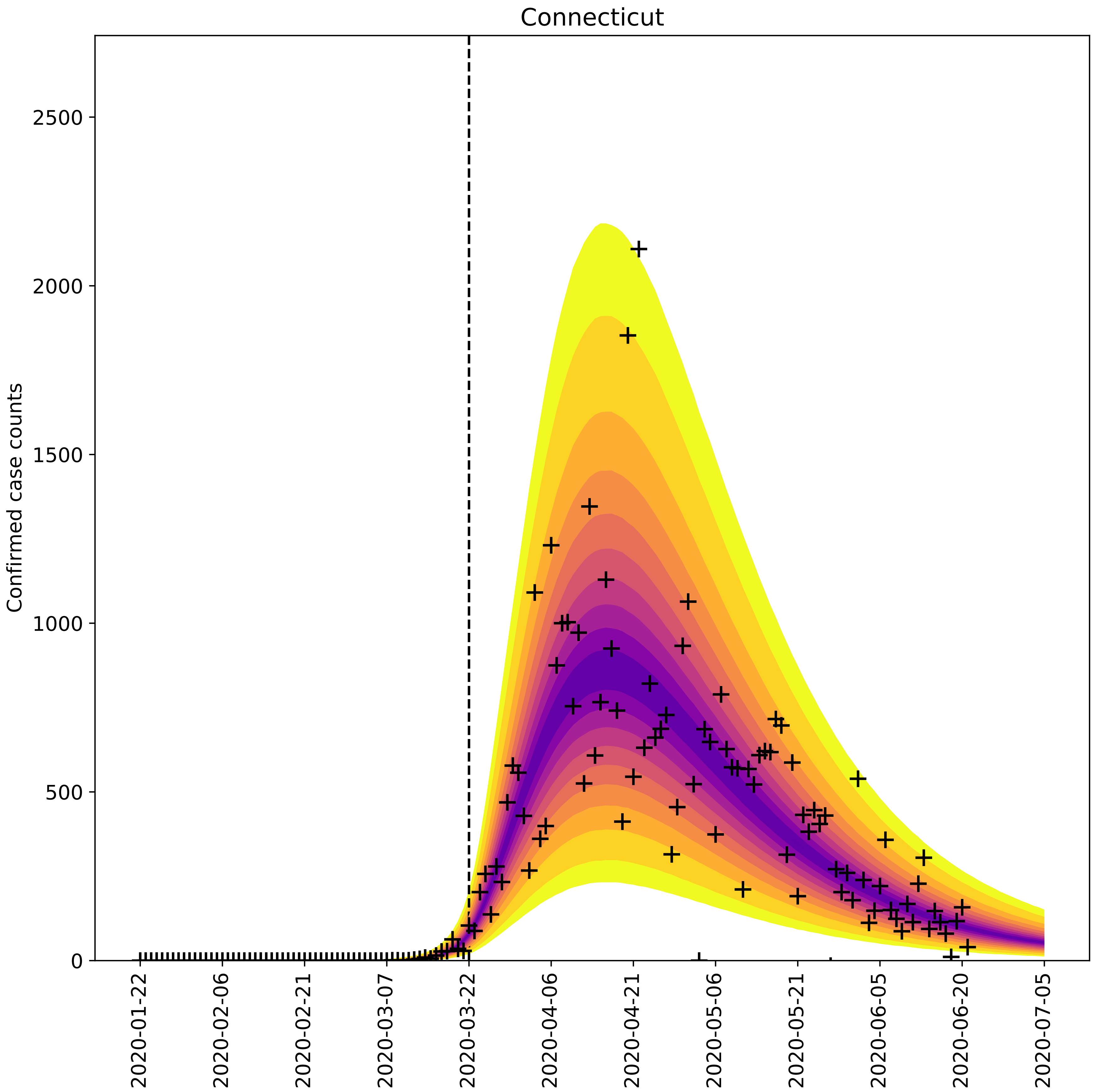

Connecticut
Confirmed case counts
0
500
1000
1500
2000
2500
2020-01-22
2020-02-06
2020-02-21
2020-03-07
2020-03-22
2020-04-06
2020-04-21
2020-05-06
2020-05-21
2020-06-05
2020-06-20
2020-07-05

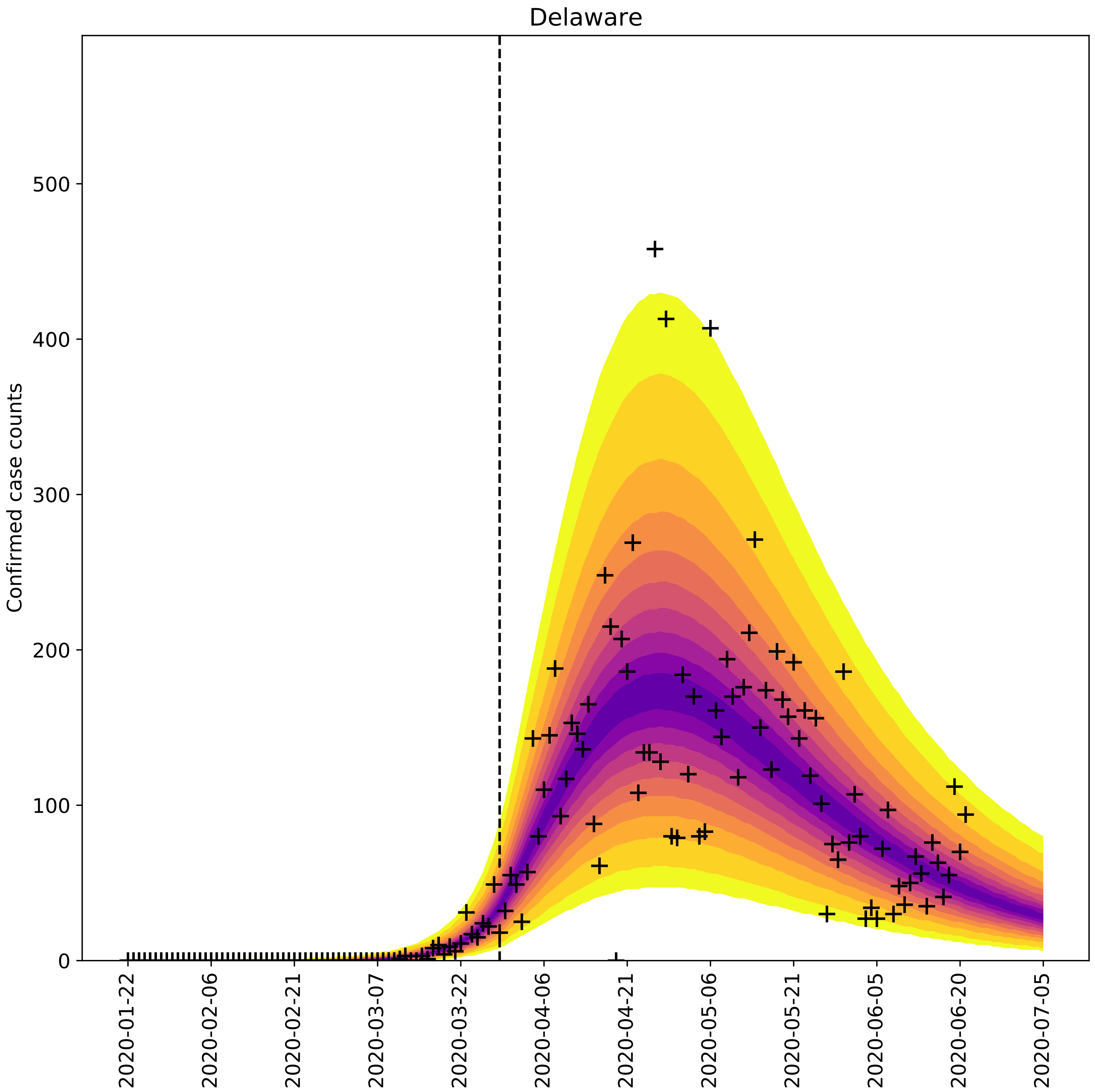

Delaware
Confirmed case counts
0
100
200
300
400
500
2020-01-22
2020-02-06
2020-02-21
2020-03-07
2020-03-22
2020-04-06
2020-04-21
2020-05-06
2020-05-21
2020-06-05
2020-06-20
2020-07-05

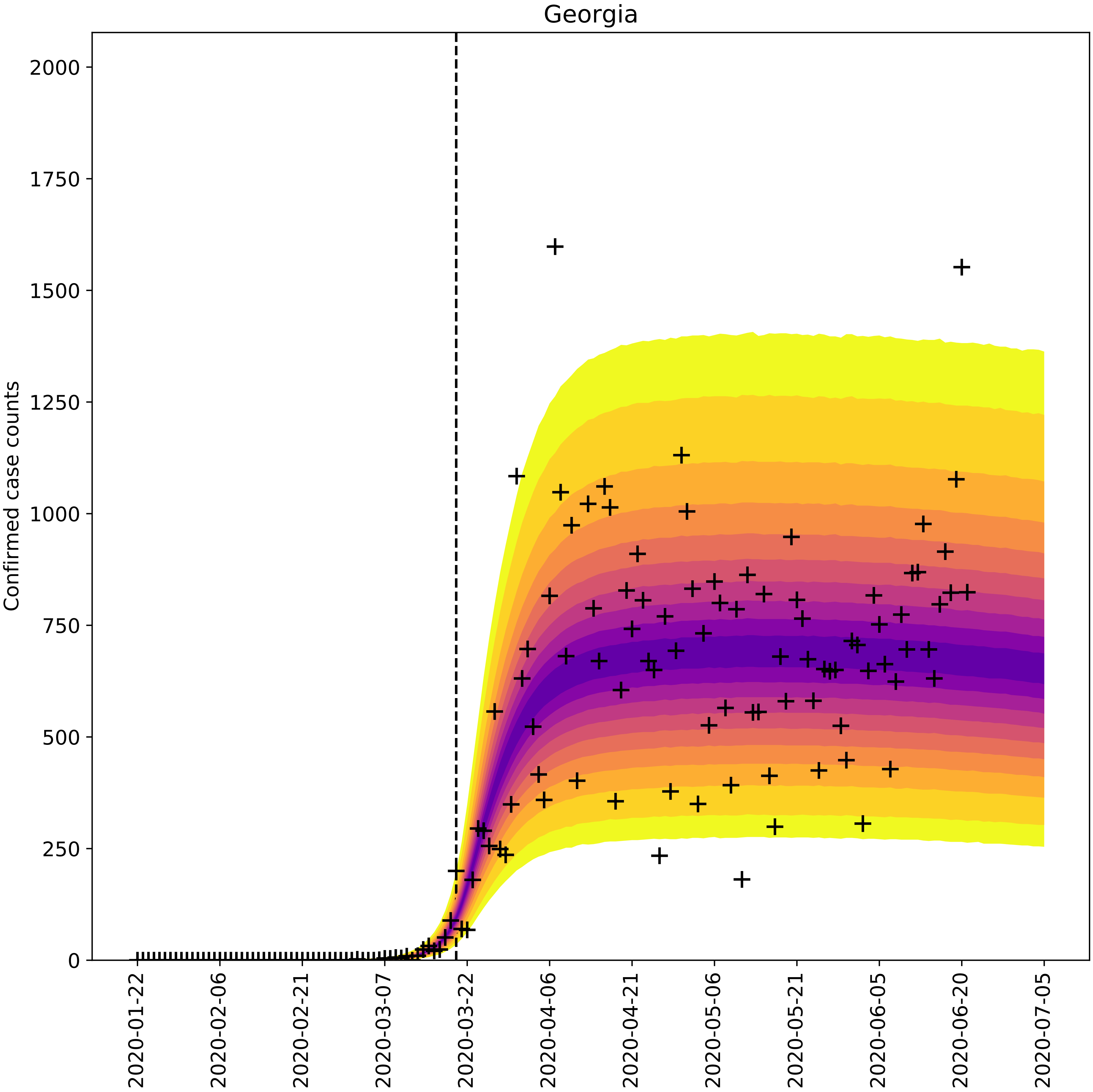
Georgia
Confirmed case counts
0
250
500
750
1000
1250
1500
1750
2000
2020-01-22
2020-02-06
2020-02-21
2020-03-07
2020-03-22
2020-04-06
2020-04-21
2020-05-06
2020-05-21
2020-06-05
2020-06-20
2020-07-05

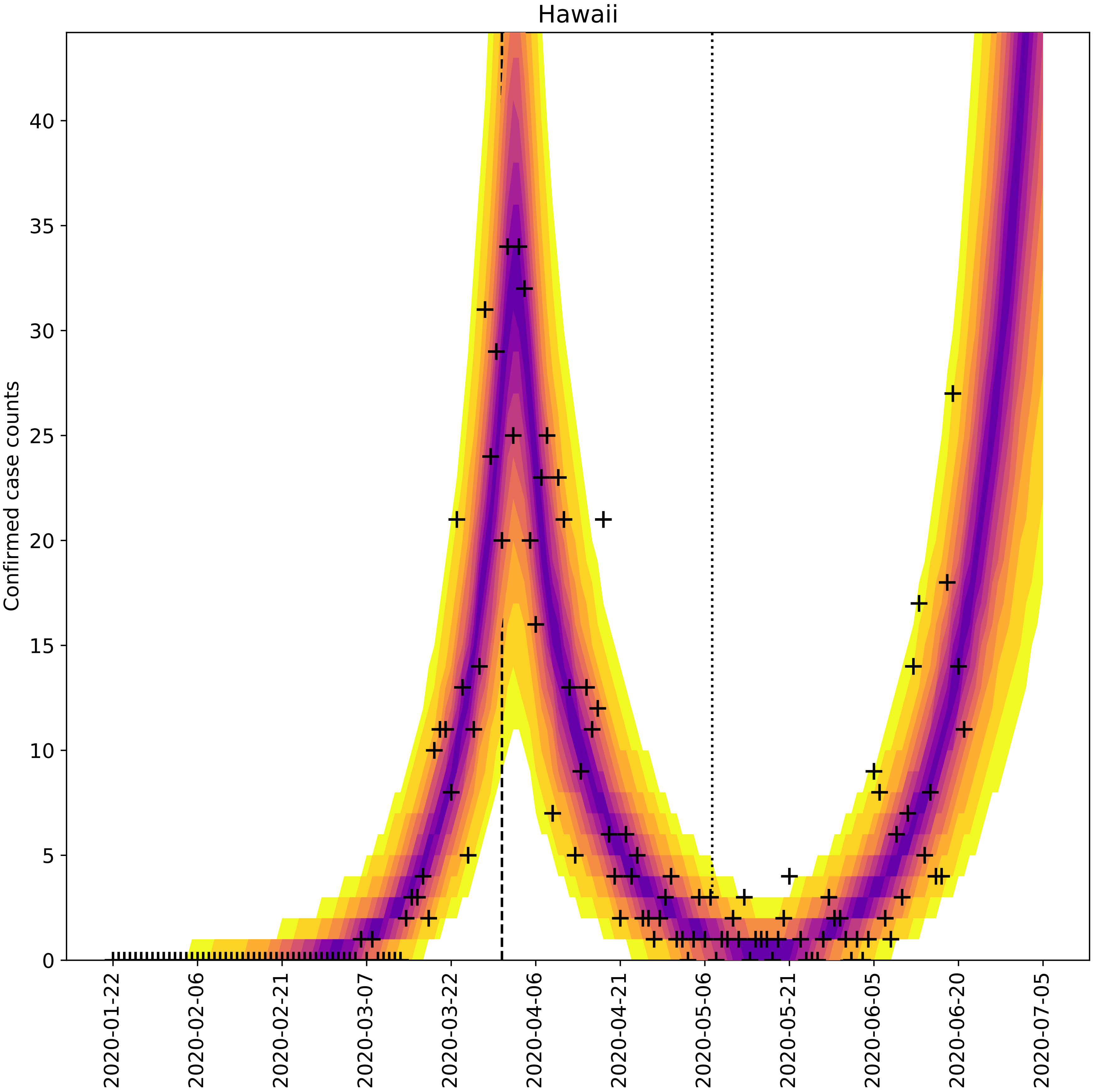

Hawaii
Confirmed case counts
0
5
10
15
20
25
30
35
40
2020-01-22
2020-02-06
2020-02-21
2020-03-07
2020-03-22
2020-04-06
2020-04-21
2020-05-06
2020-05-21
2020-06-05
2020-06-20
2020-07-05

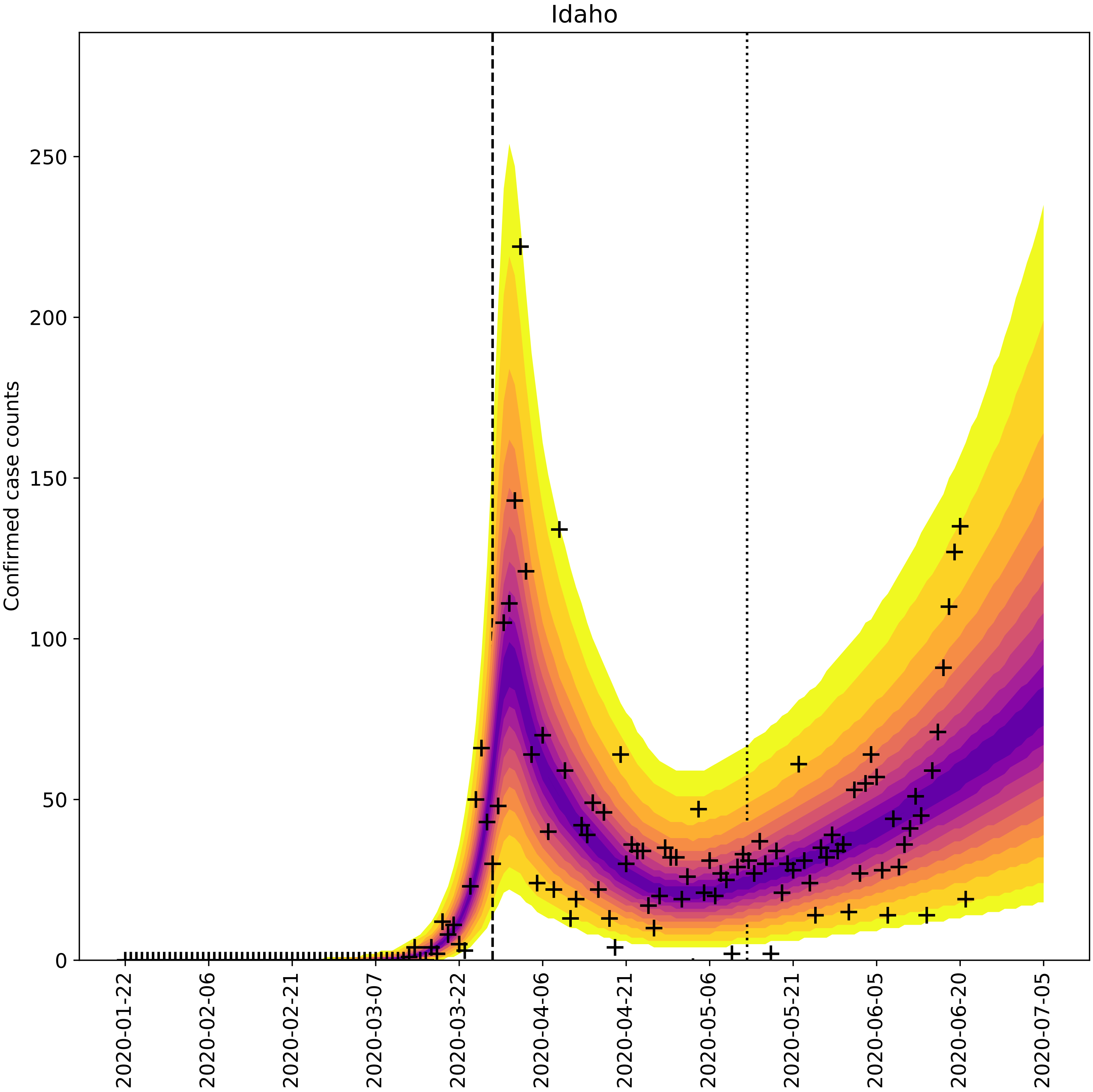

Idaho
Confirmed case counts
0
50
100
150
200
250
2020-01-22
2020-02-06
2020-02-21
2020-03-07
2020-03-22
2020-04-06
2020-04-21
2020-05-06
2020-05-21
2020-06-05
2020-06-20
2020-07-05

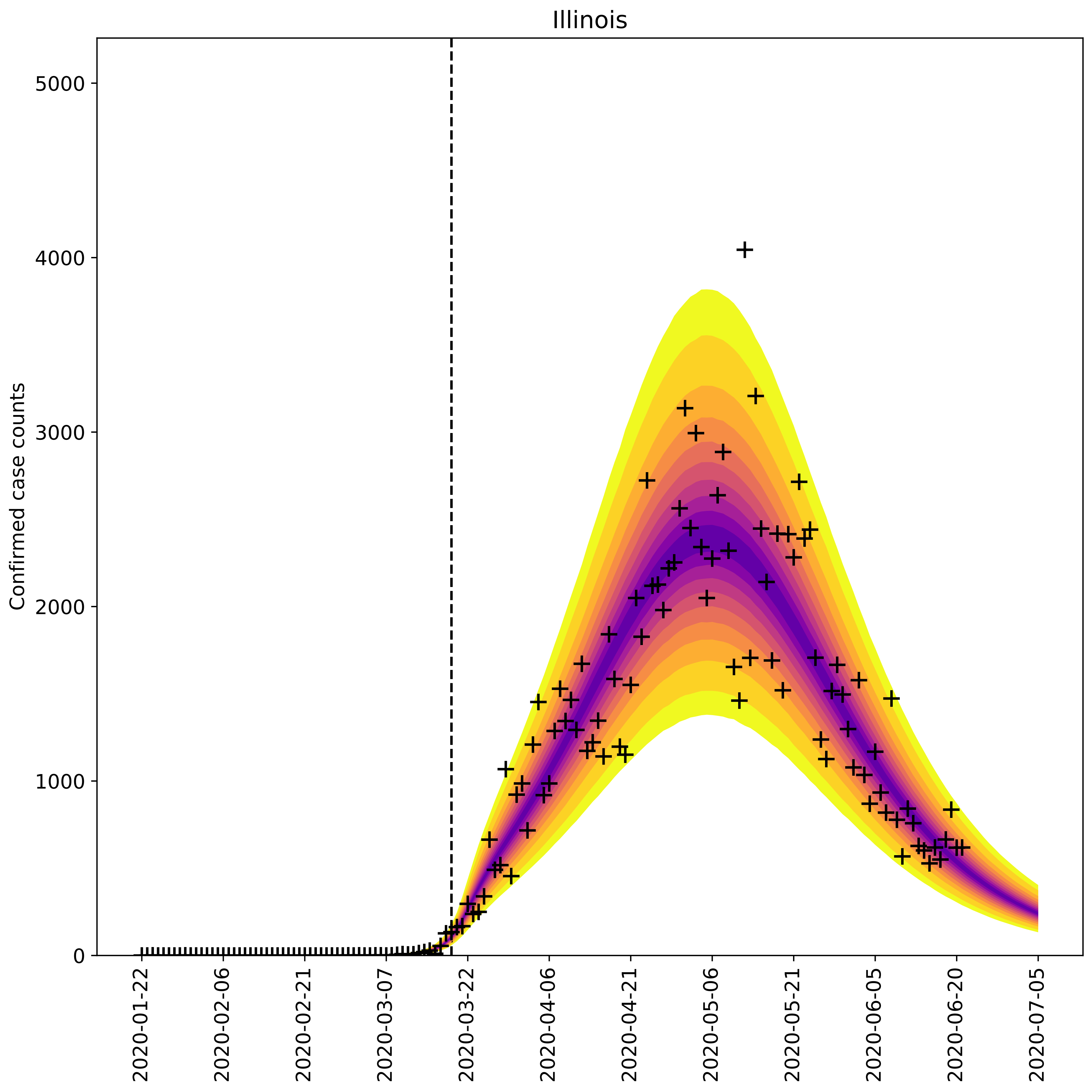
Illinois
Confirmed case counts
0
1000
2000
3000
4000
5000
2020-01-22
2020-02-06
2020-02-21
2020-03-07
2020-03-22
2020-04-06
2020-04-21
2020-05-06
2020-05-21
2020-06-05
2020-06-20
2020-07-05

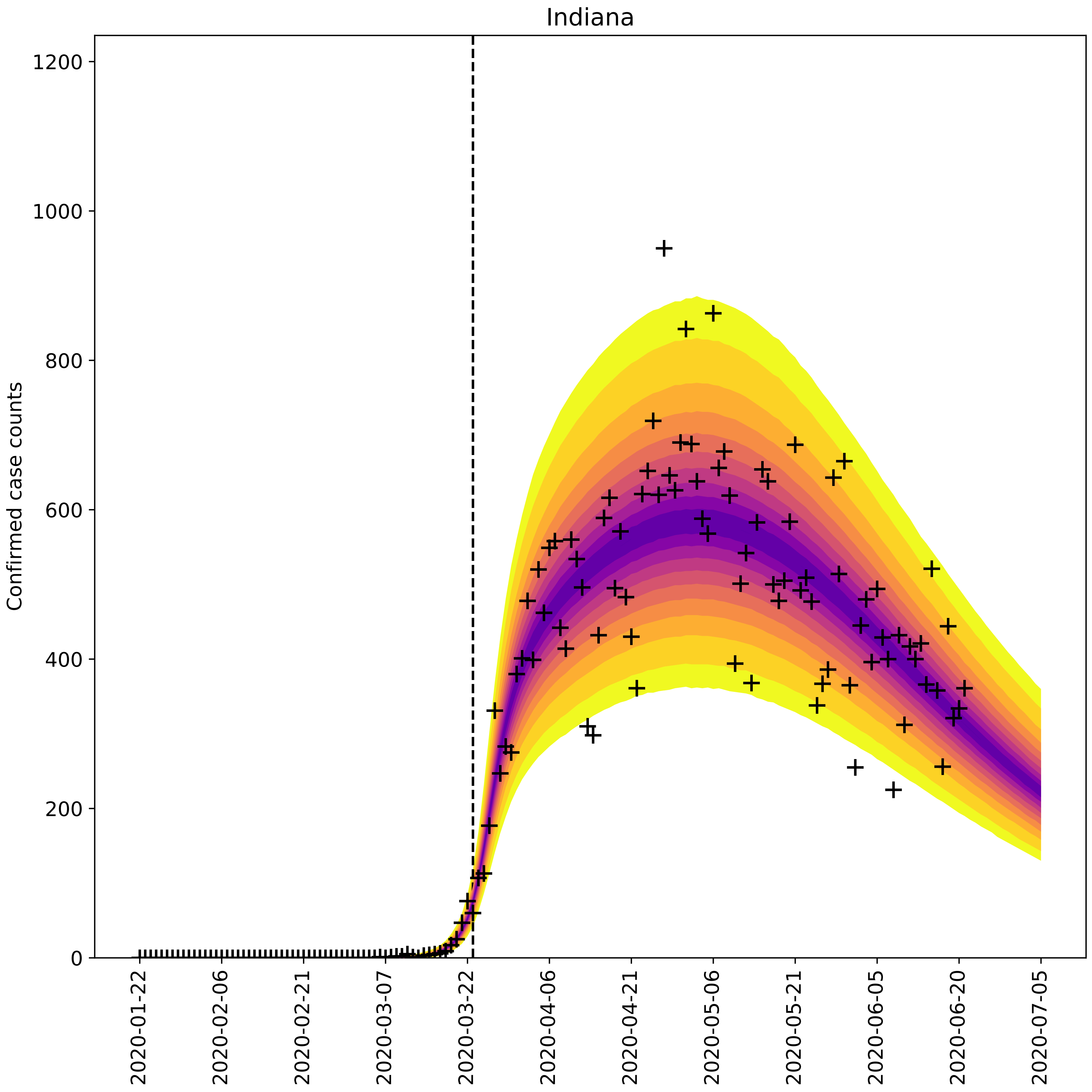

Indiana
Confirmed case counts
0
200
400
600
800
1000
1200
2020-01-22
2020-02-06
2020-02-21
2020-03-07
2020-03-22
2020-04-06
2020-04-21
2020-05-06
2020-05-21
2020-06-05
2020-06-20
2020-07-05

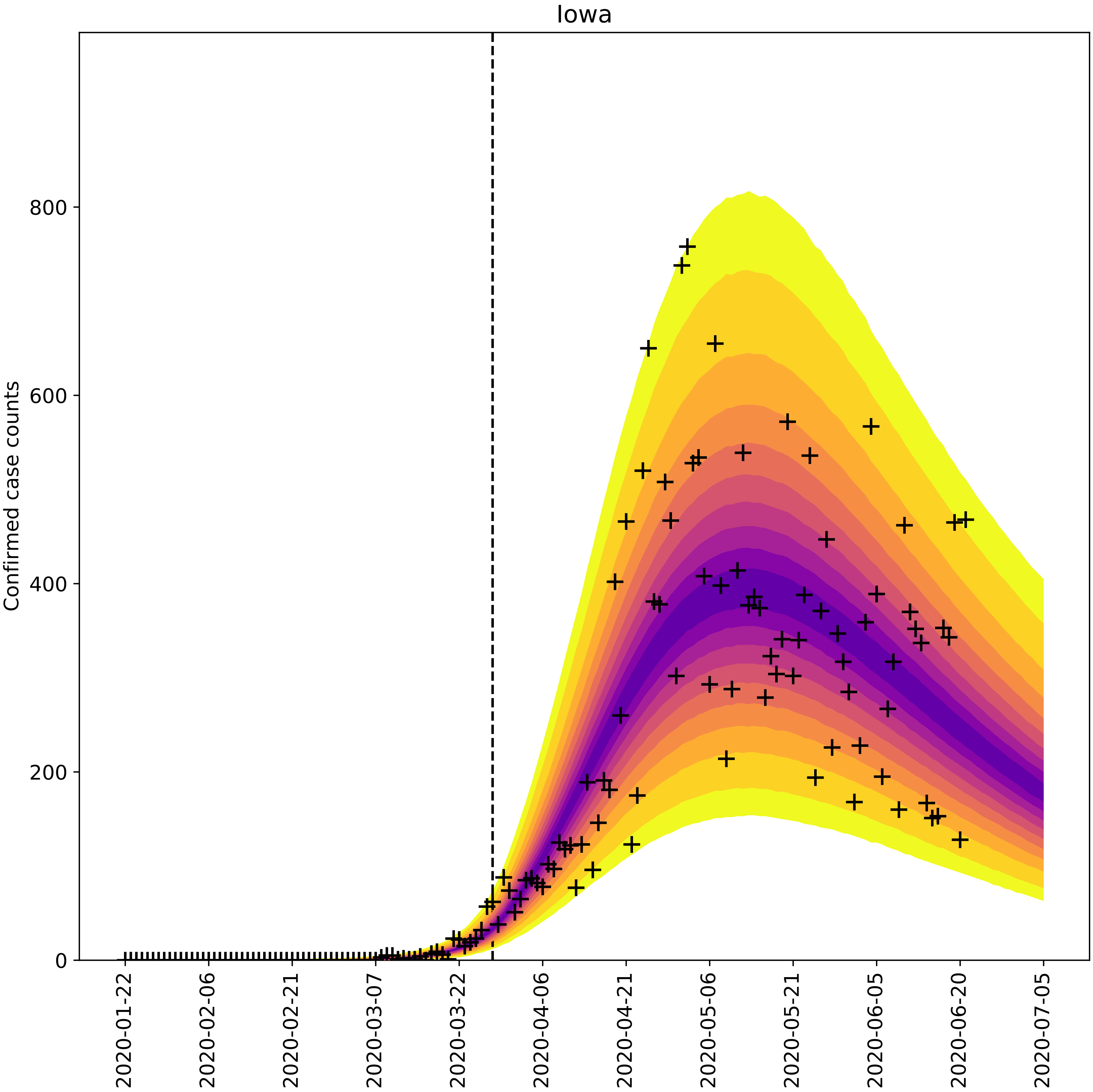

Iowa
Confirmed case counts
0
200
400
600
800
2020-01-22
2020-02-06
2020-02-21
2020-03-07
2020-03-22
2020-04-06
2020-04-21
2020-05-06
2020-05-21
2020-06-05
2020-06-20
2020-07-05

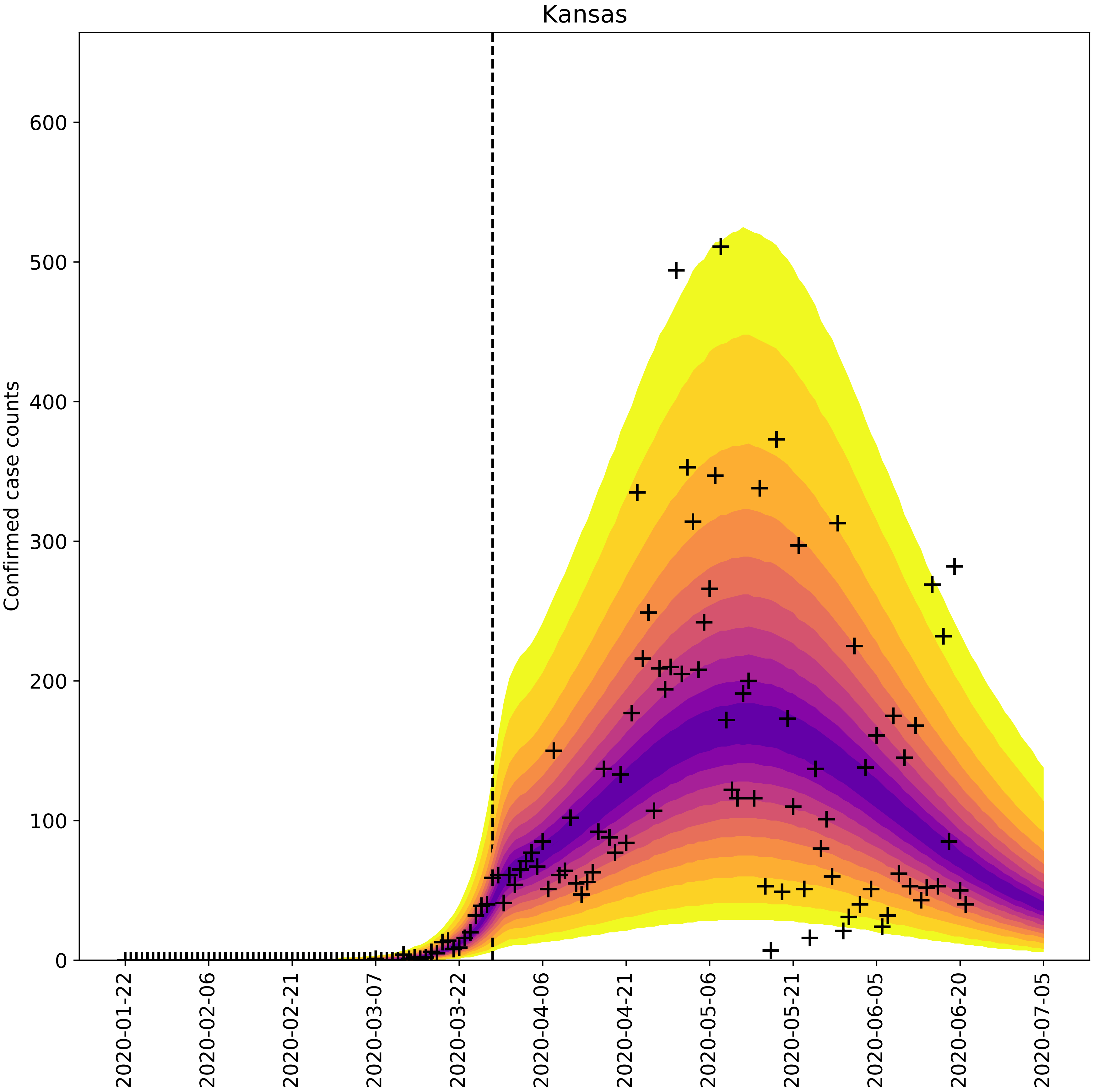
Kansas
Confirmed case counts
0
100
200
300
400
500
600
2020-01-22
2020-02-06
2020-02-21
2020-03-07
2020-03-22
2020-04-06
2020-04-21
2020-05-06
2020-05-21
2020-06-05
2020-06-20
2020-07-05

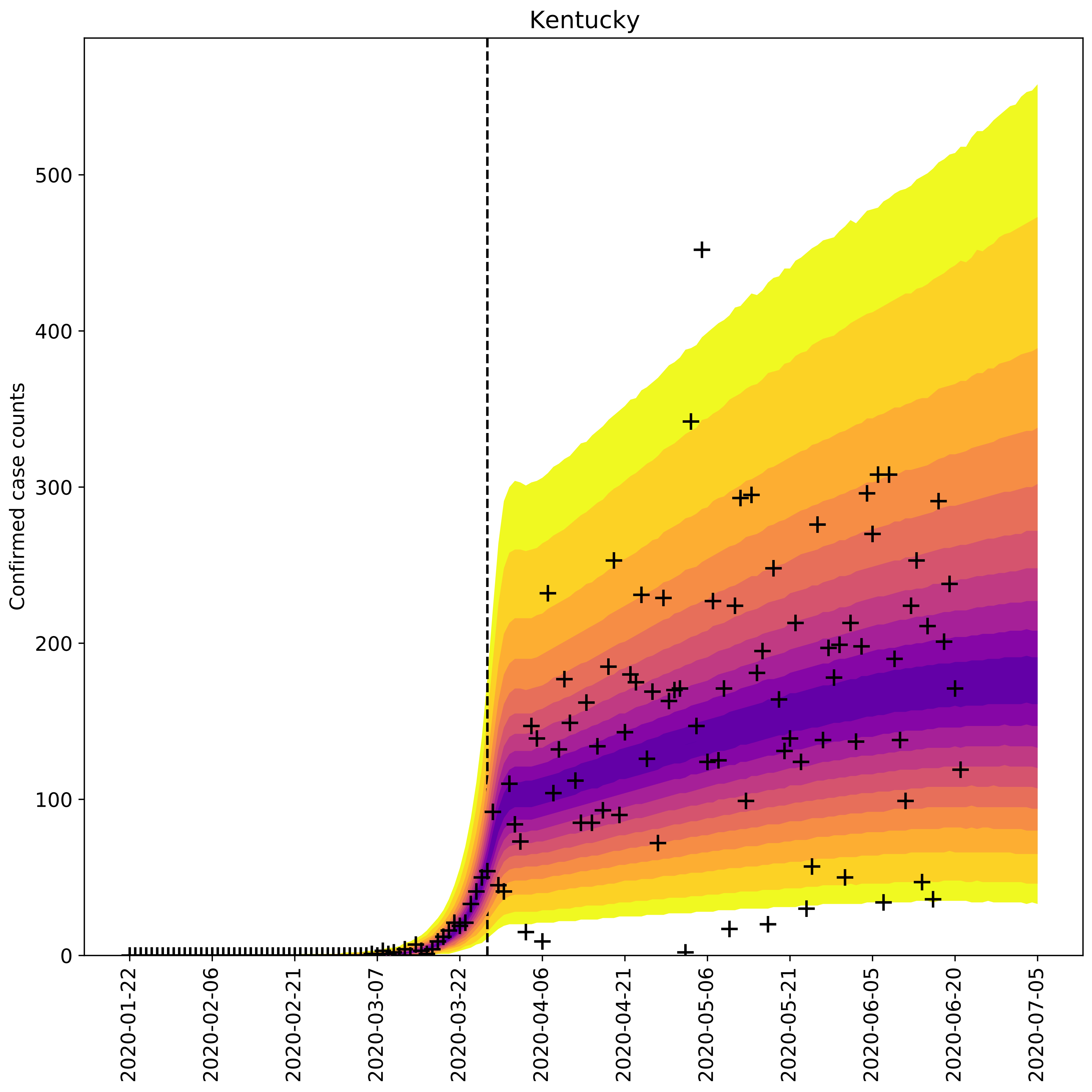

Kentucky
Confirmed case counts
0
100
200
300
400
500
2020-01-22
2020-02-06
2020-02-21
2020-03-07
2020-03-22
2020-04-06
2020-04-21
2020-05-06
2020-05-21
2020-06-05
2020-06-20
2020-07-05

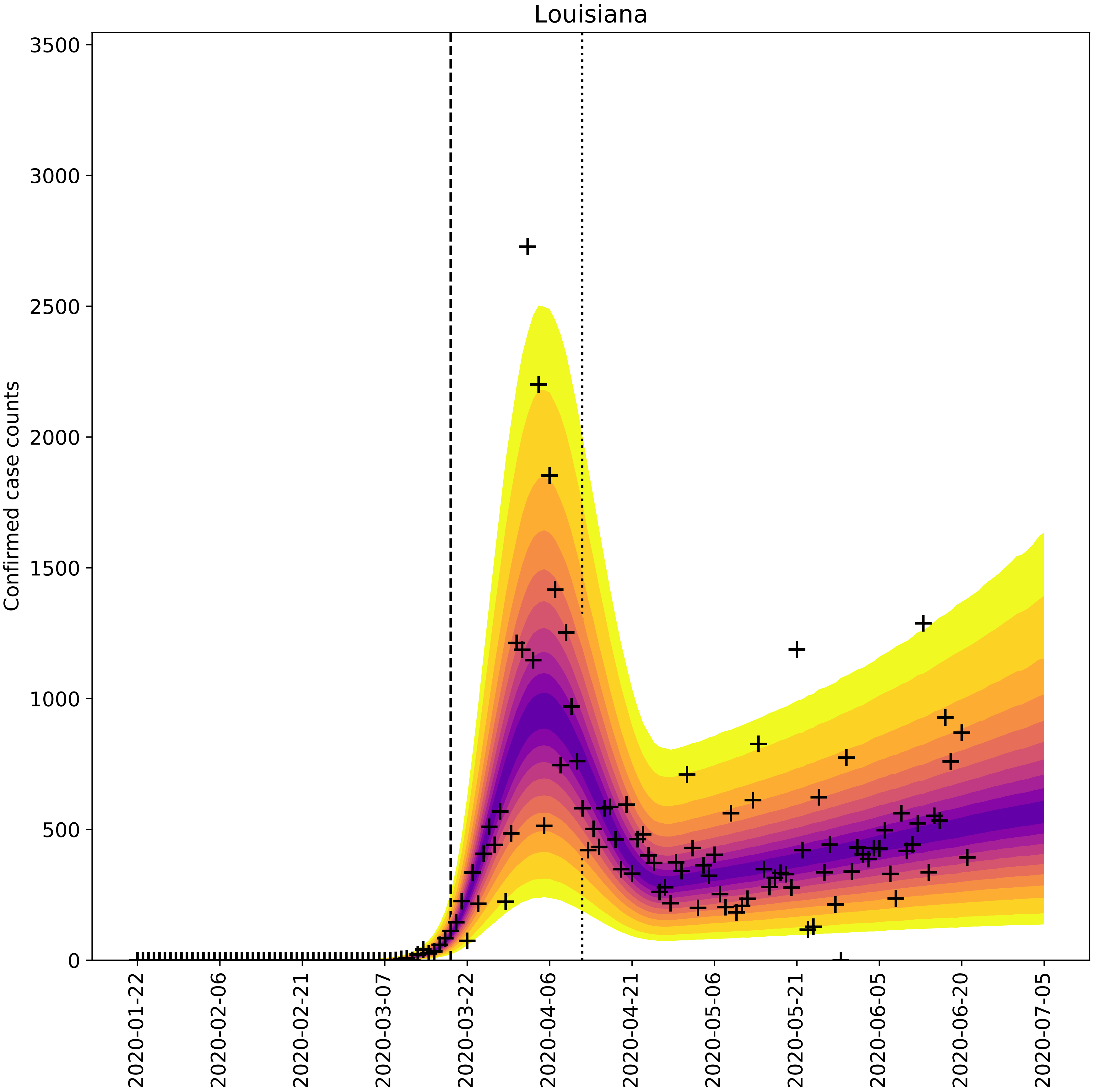

Louisiana
Confirmed case counts
0
500
1000
1500
2000
2500
3000
3500
2020-01-22
2020-02-06
2020-02-21
2020-03-07
2020-03-22
2020-04-06
2020-04-21
2020-05-06
2020-05-21
2020-06-05
2020-06-20
2020-07-05

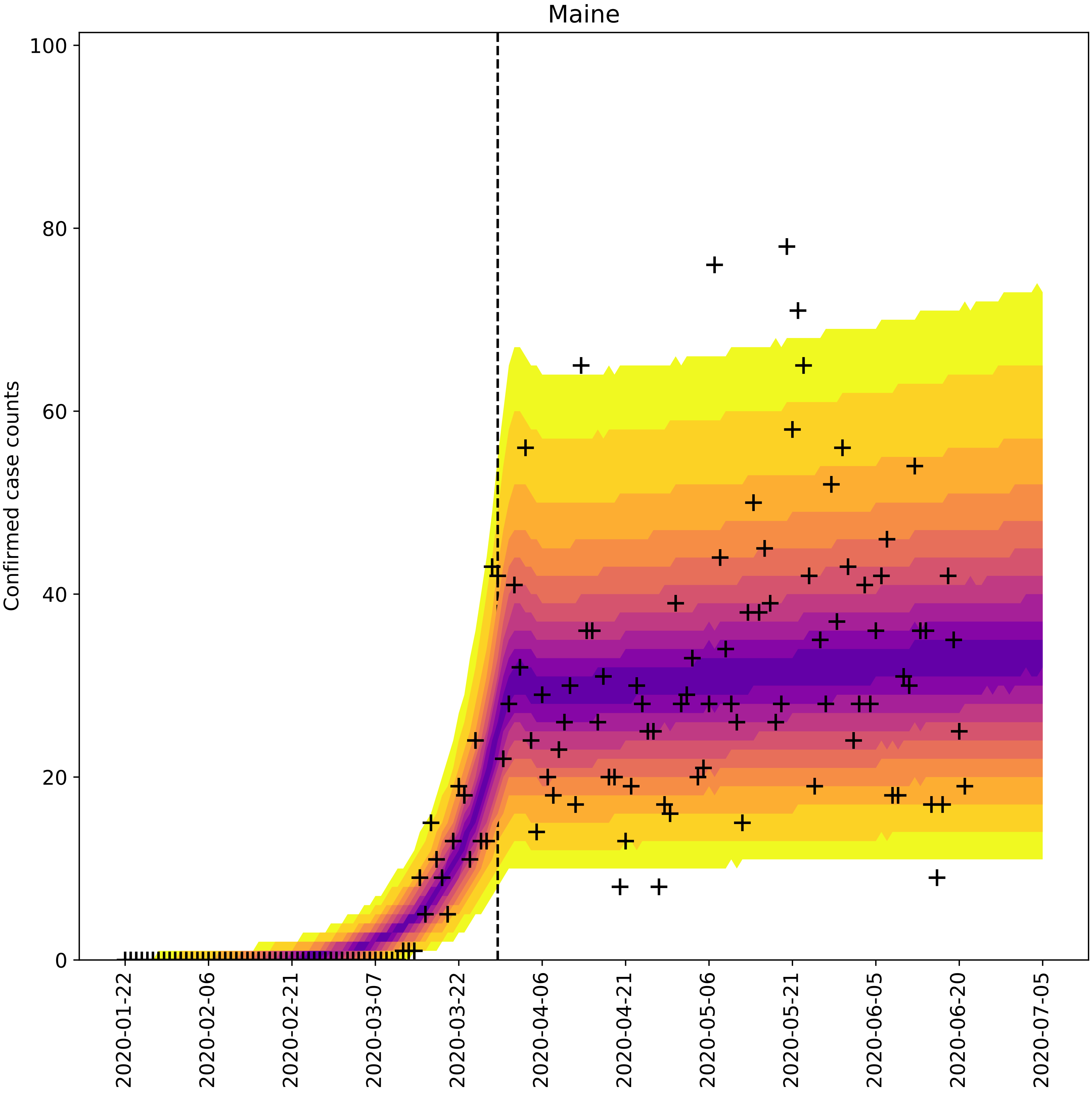

Maine
Confirmed case counts
0
20
40
60
80
100
2020-01-22
2020-02-06
2020-02-21
2020-03-07
2020-03-22
2020-04-06
2020-04-21
2020-05-06
2020-05-21
2020-06-05
2020-06-20
2020-07-05

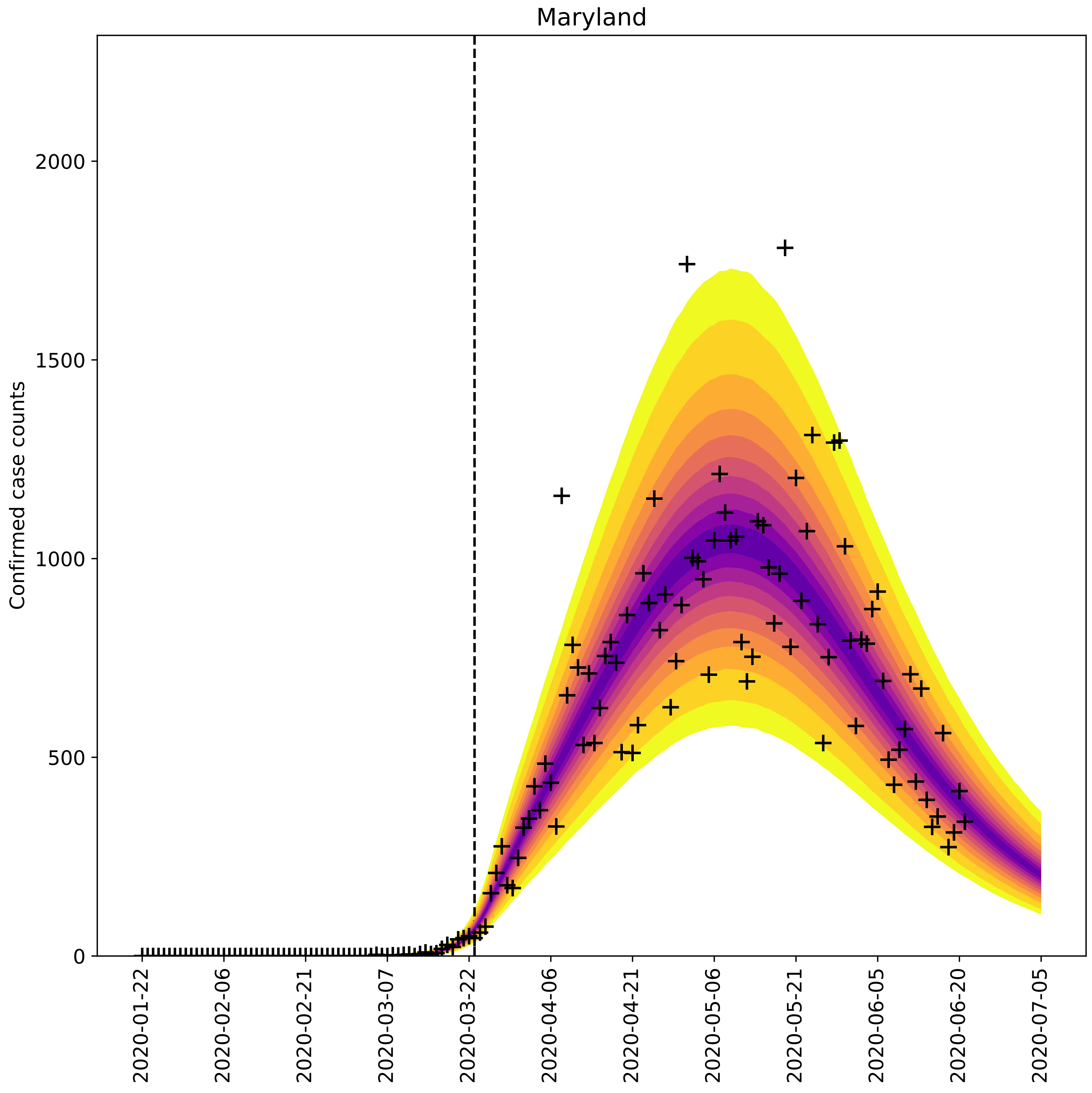

Maryland
Confirmed case counts
0
500
1000
1500
2000
2020-01-22
2020-02-06
2020-02-21
2020-03-07
2020-03-22
2020-04-06
2020-04-21
2020-05-06
2020-05-21
2020-06-05
2020-06-20
2020-07-05

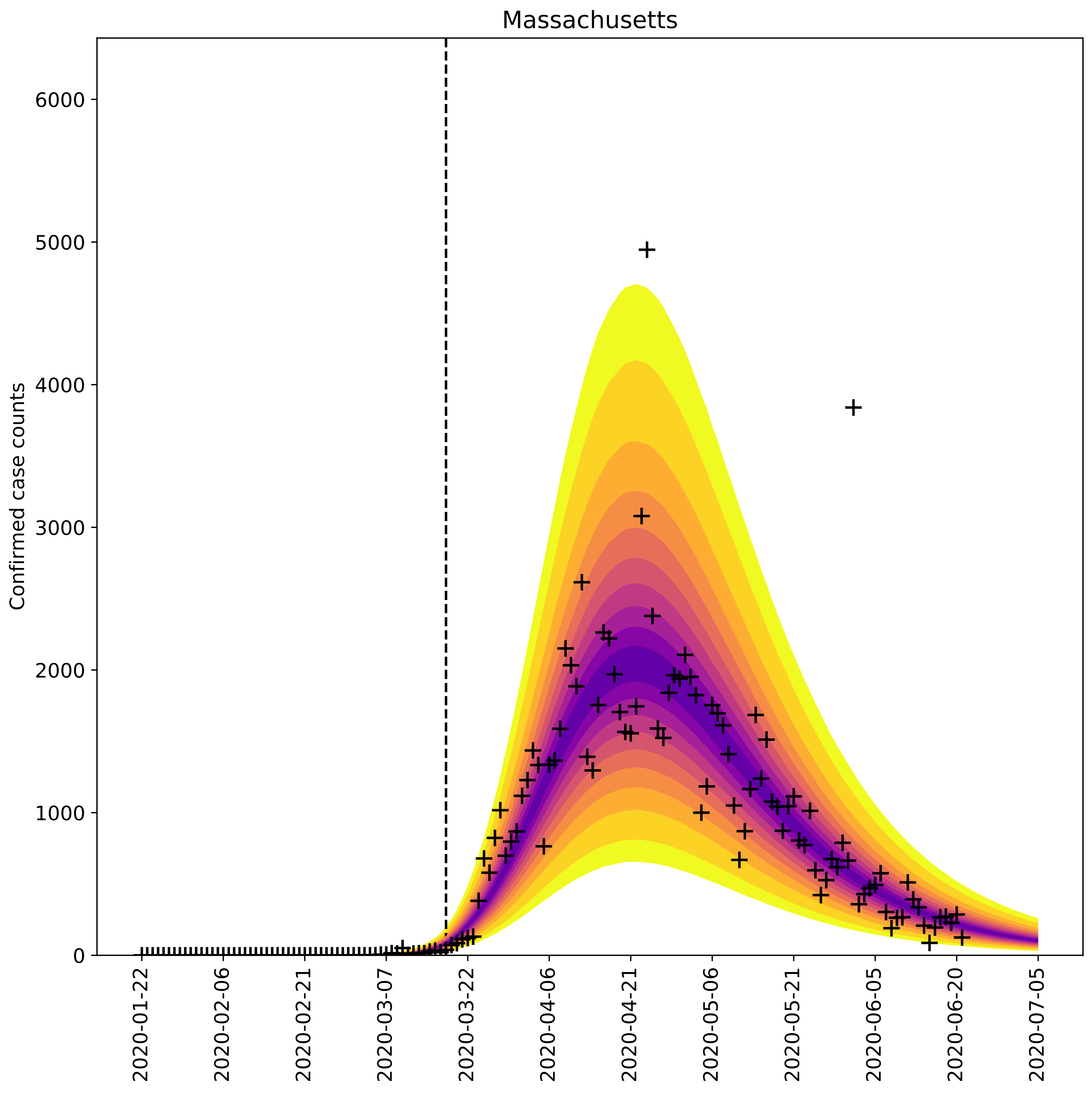

Massachusetts
Confirmed case counts
6000
5000
4000
3000
2000
1000
0
2020-01-22
2020-02-06
2020-02-21
2020-03-07
2020-03-22
2020-04-06
2020-04-21
2020-05-06
2020-05-21
2020-06-05
2020-06-20
2020-07-05

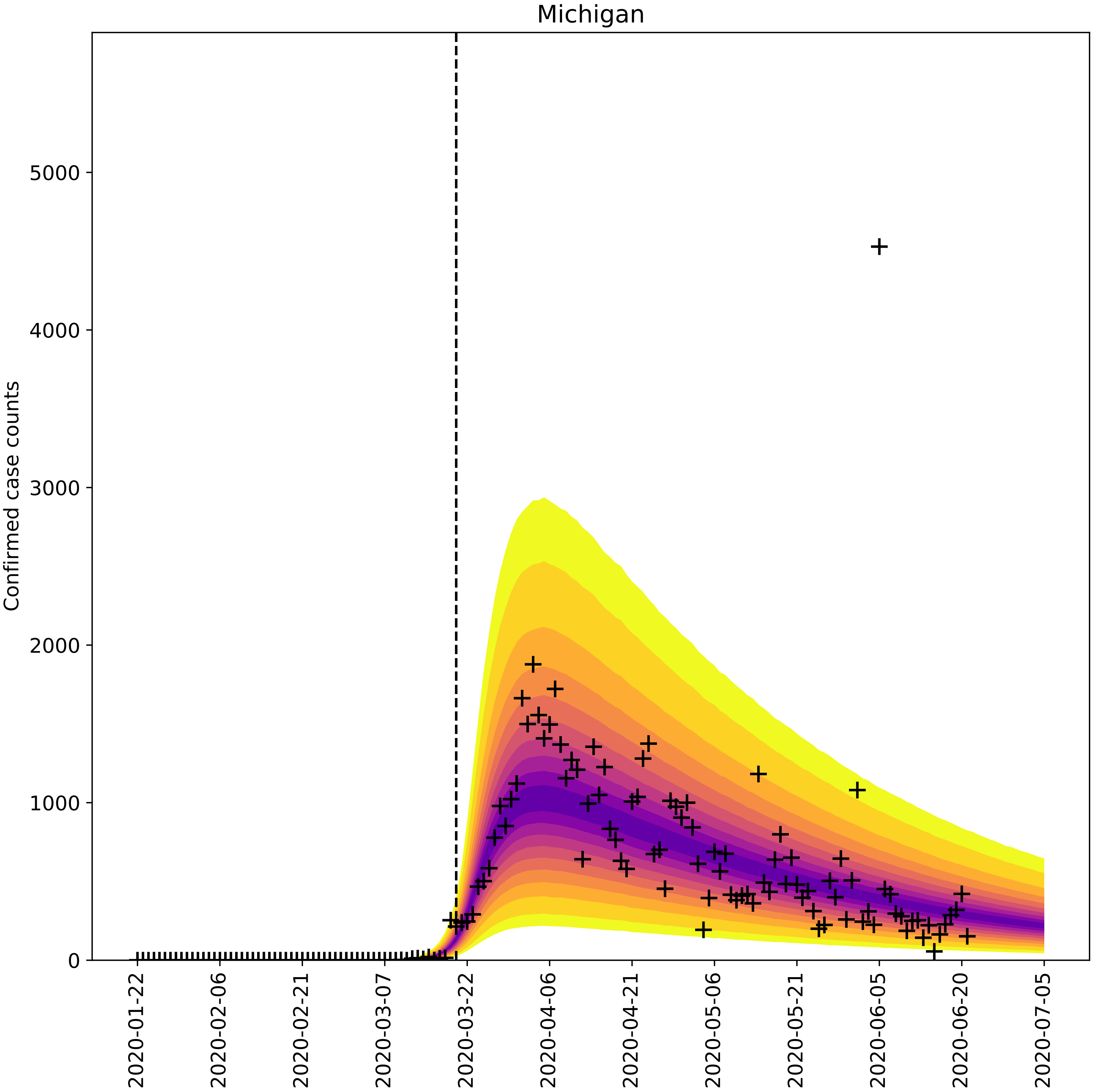

Michigan
Confirmed case counts
0
1000
2000
3000
4000
5000
2020-01-22
2020-02-06
2020-02-21
2020-03-07
2020-03-22
2020-04-06
2020-04-21
2020-05-06
2020-05-21
2020-06-05
2020-06-20
2020-07-05

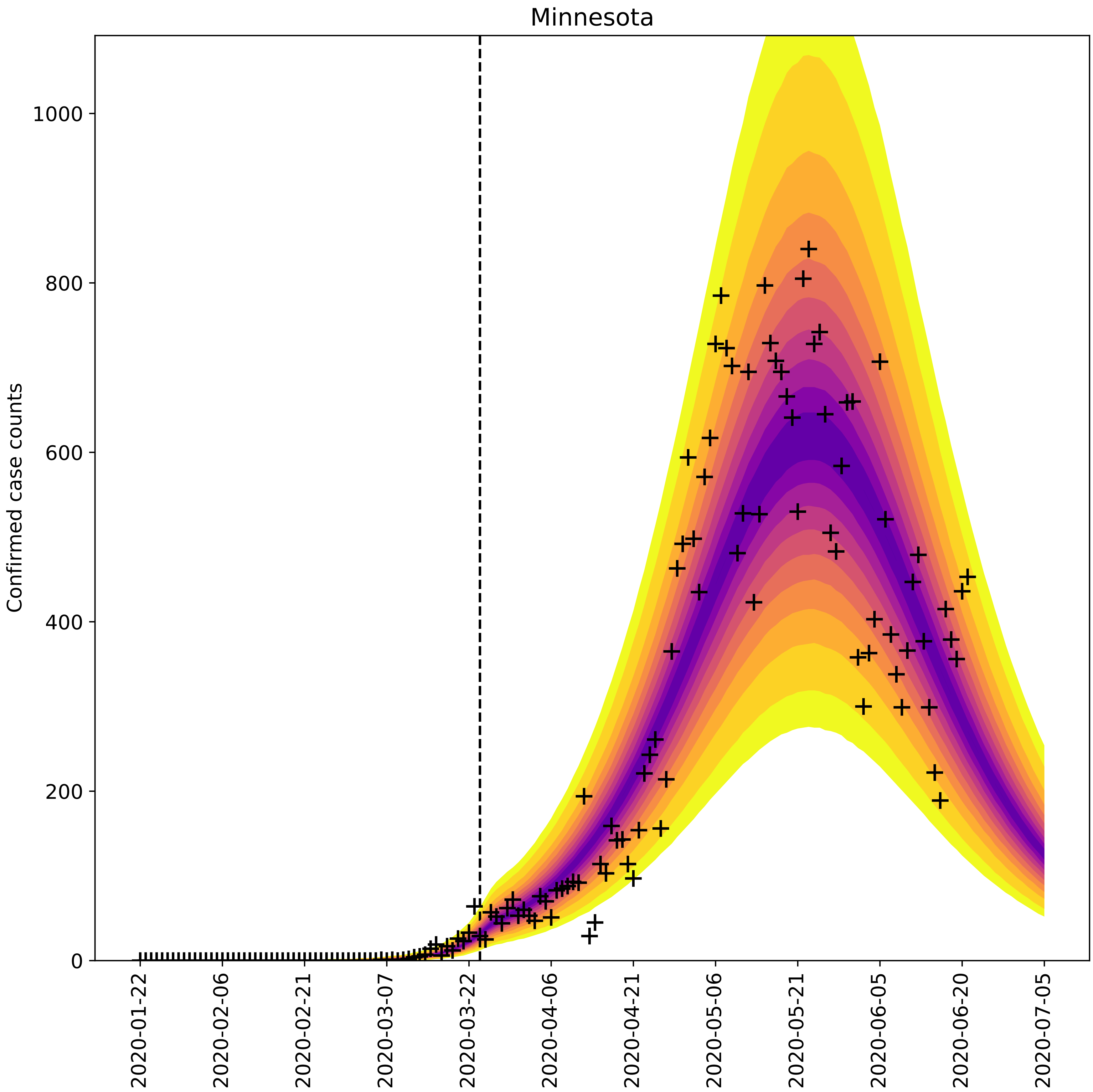

Minnesota
Confirmed case counts
0
200
400
600
800
1000
2020-01-22
2020-02-06
2020-02-21
2020-03-07
2020-03-22
2020-04-06
2020-04-21
2020-05-06
2020-05-21
2020-06-05
2020-06-20
2020-07-05

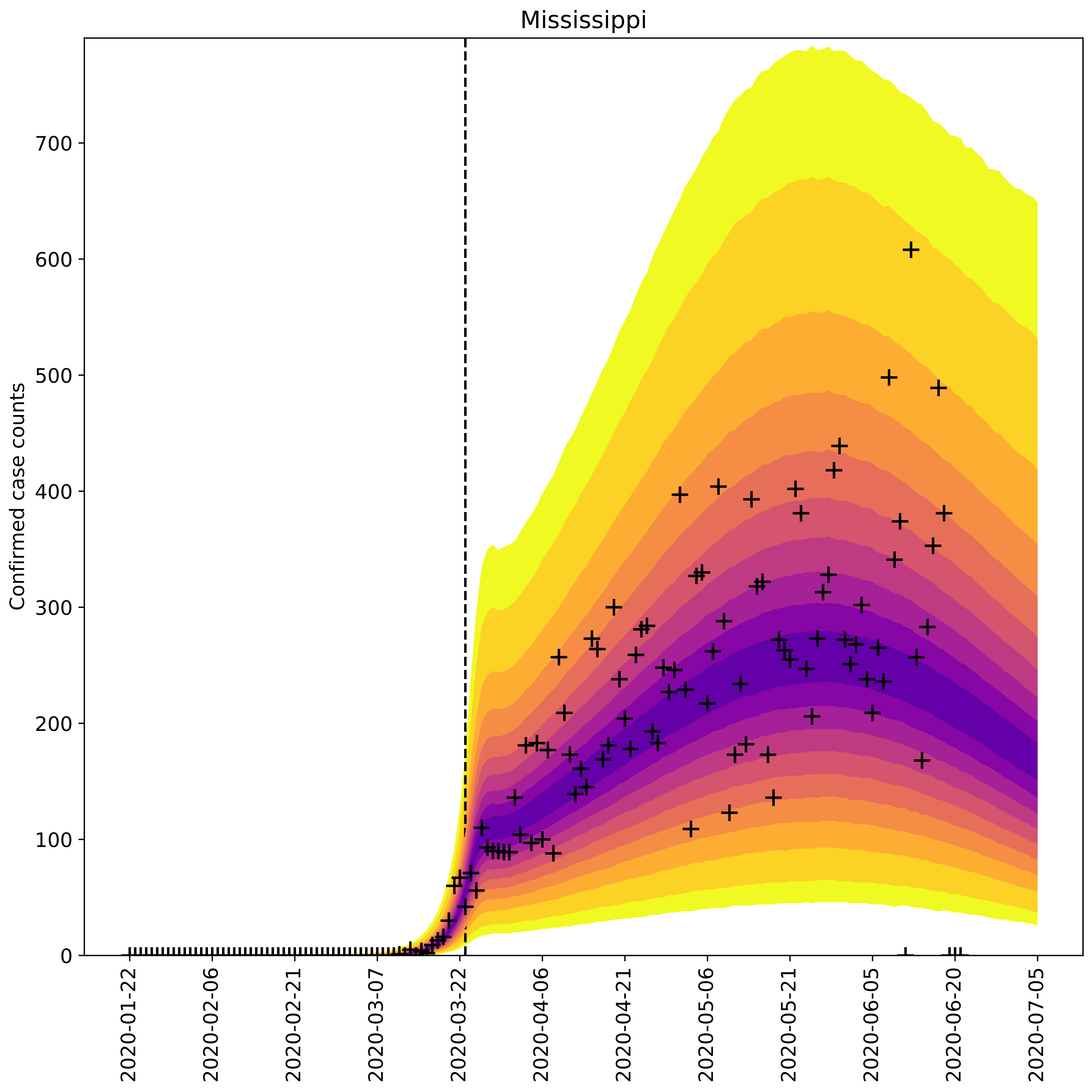
Mississippi
Confirmed case counts
0
100
200
300
400
500
600
700
2020-01-22
2020-02-06
2020-02-21
2020-03-07
2020-03-22
2020-04-06
2020-04-21
2020-05-06
2020-05-21
2020-06-05
2020-06-20
2020-07-05

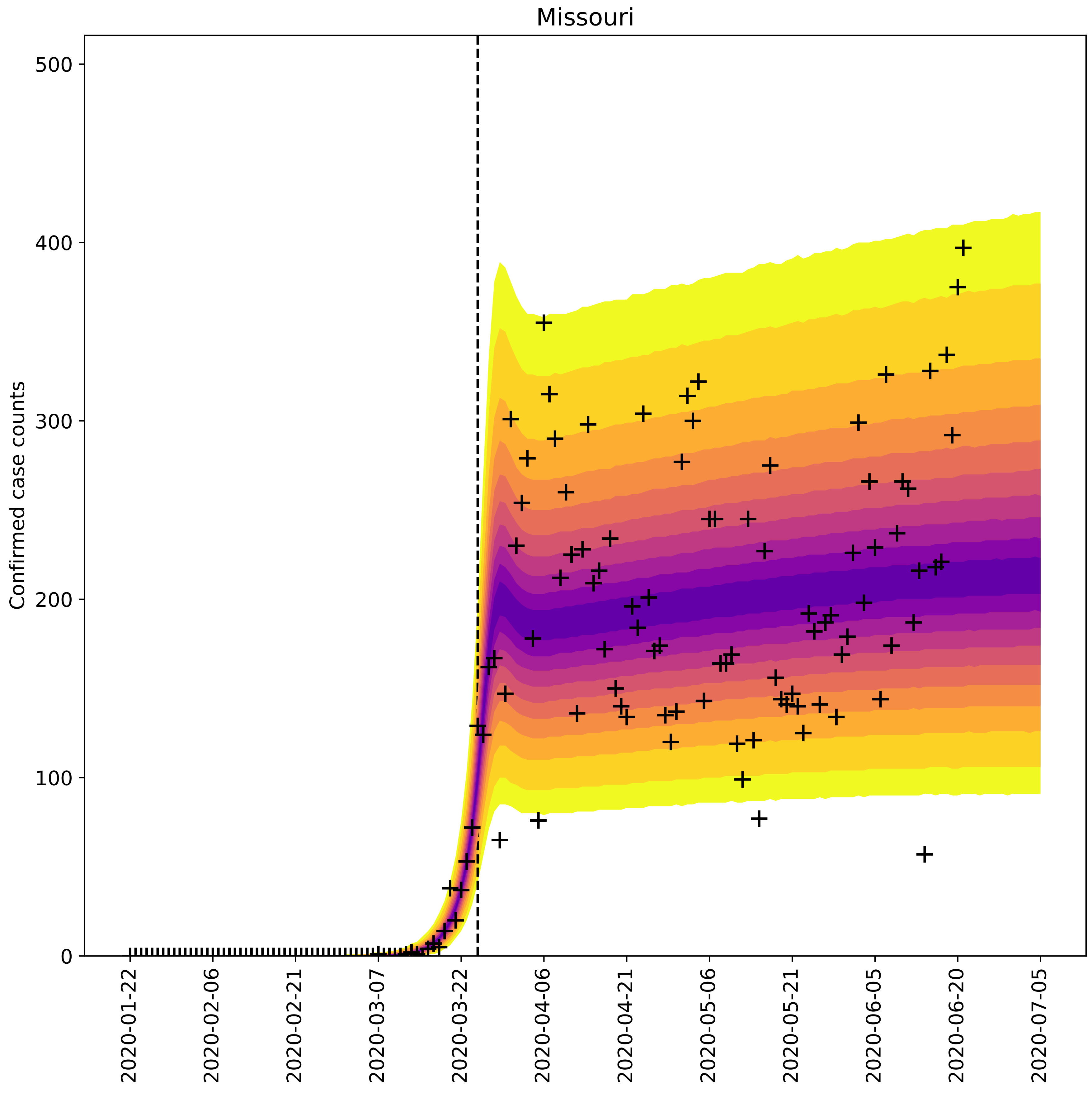

Missouri
Confirmed case counts
0
100
200
300
400
500
2020-01-22
2020-02-06
2020-02-21
2020-03-07
2020-03-22
2020-04-06
2020-04-21
2020-05-06
2020-05-21
2020-06-05
2020-06-20
2020-07-05

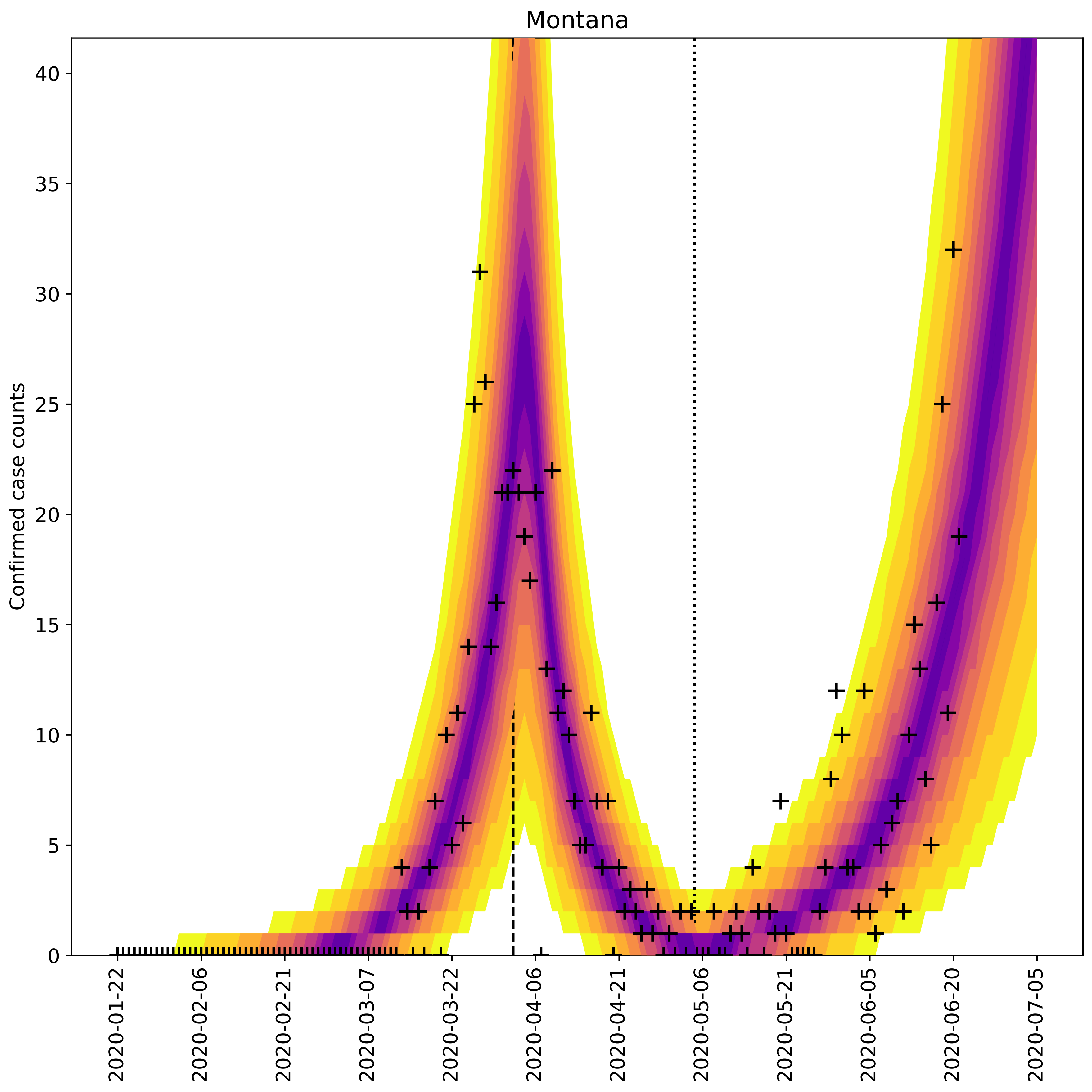

Montana
Confirmed case counts
0
5
10
15
20
25
30
35
40
2020-01-22
2020-02-06
2020-02-21
2020-03-07
2020-03-22
2020-04-06
2020-04-21
2020-05-06
2020-05-21
2020-06-05
2020-06-20
2020-07-05

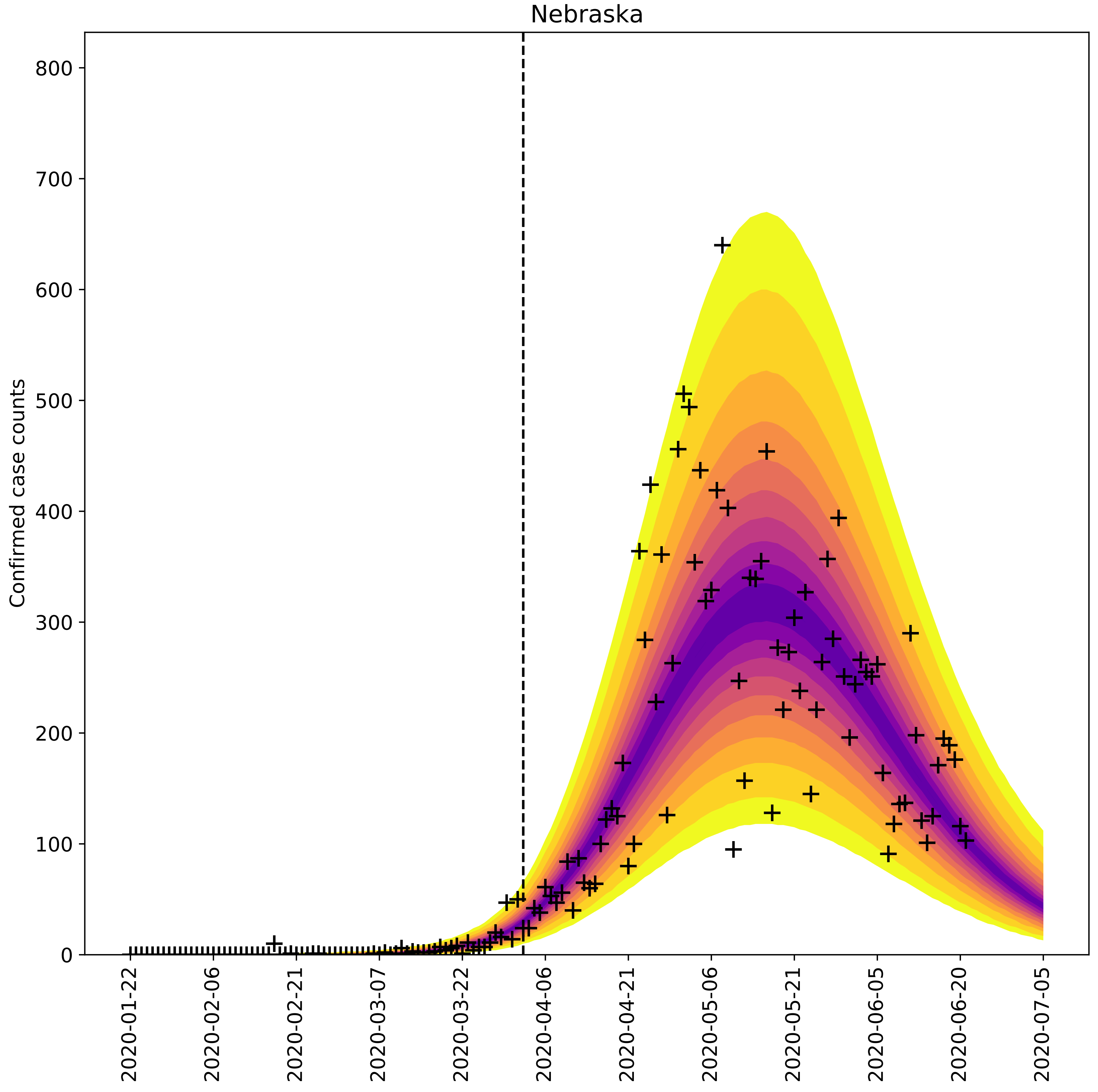

Nebraska
Confirmed case counts
0
100
200
300
400
500
600
700
800
2020-01-22
2020-02-06
2020-02-21
2020-03-07
2020-03-22
2020-04-06
2020-04-21
2020-05-06
2020-05-21
2020-06-05
2020-06-20
2020-07-05

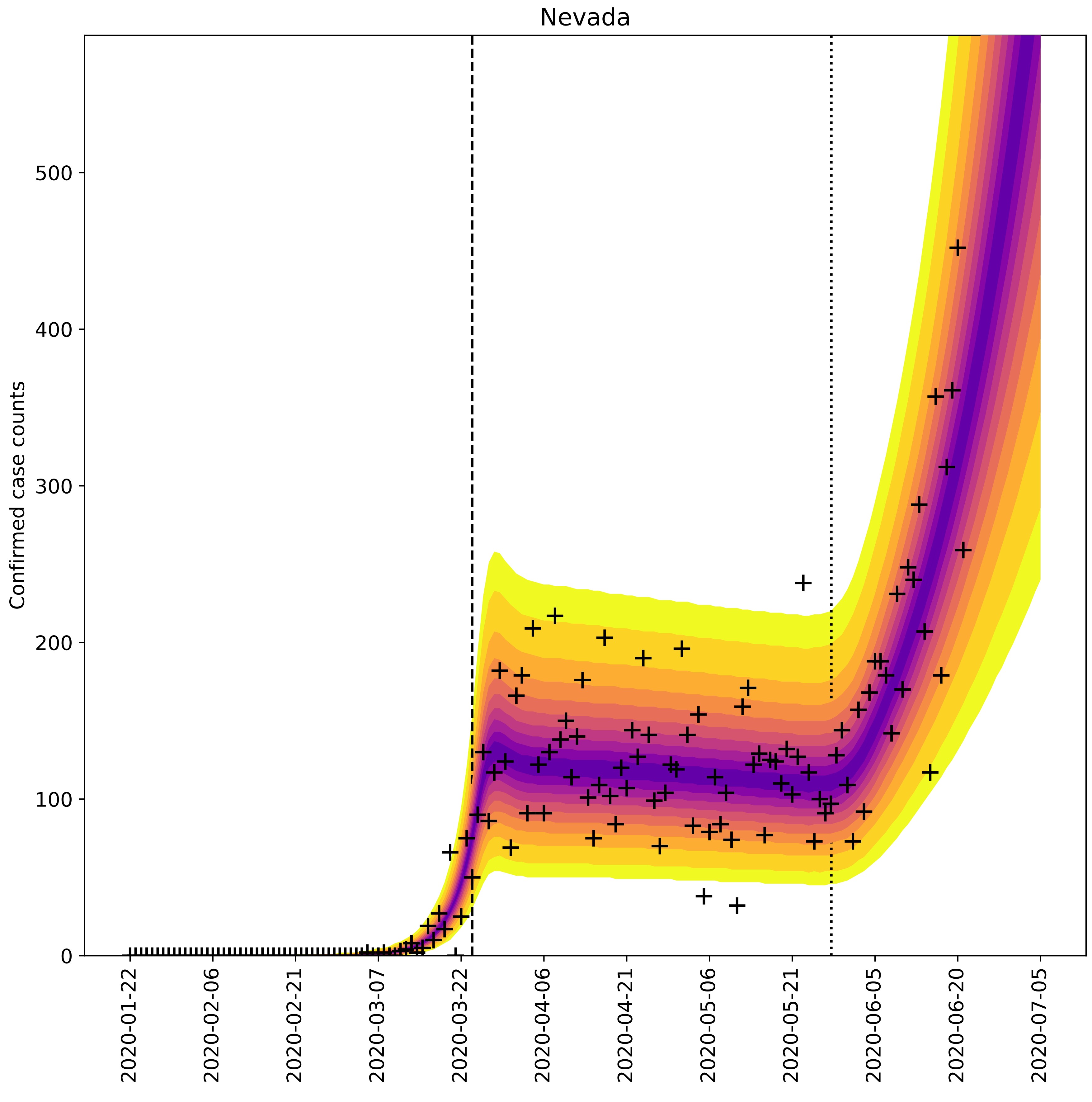

Nevada
Confirmed case counts
0
100
200
300
400
500
2020-01-22
2020-02-06
2020-02-21
2020-03-07
2020-03-22
2020-04-06
2020-04-21
2020-05-06
2020-05-21
2020-06-05
2020-06-20
2020-07-05

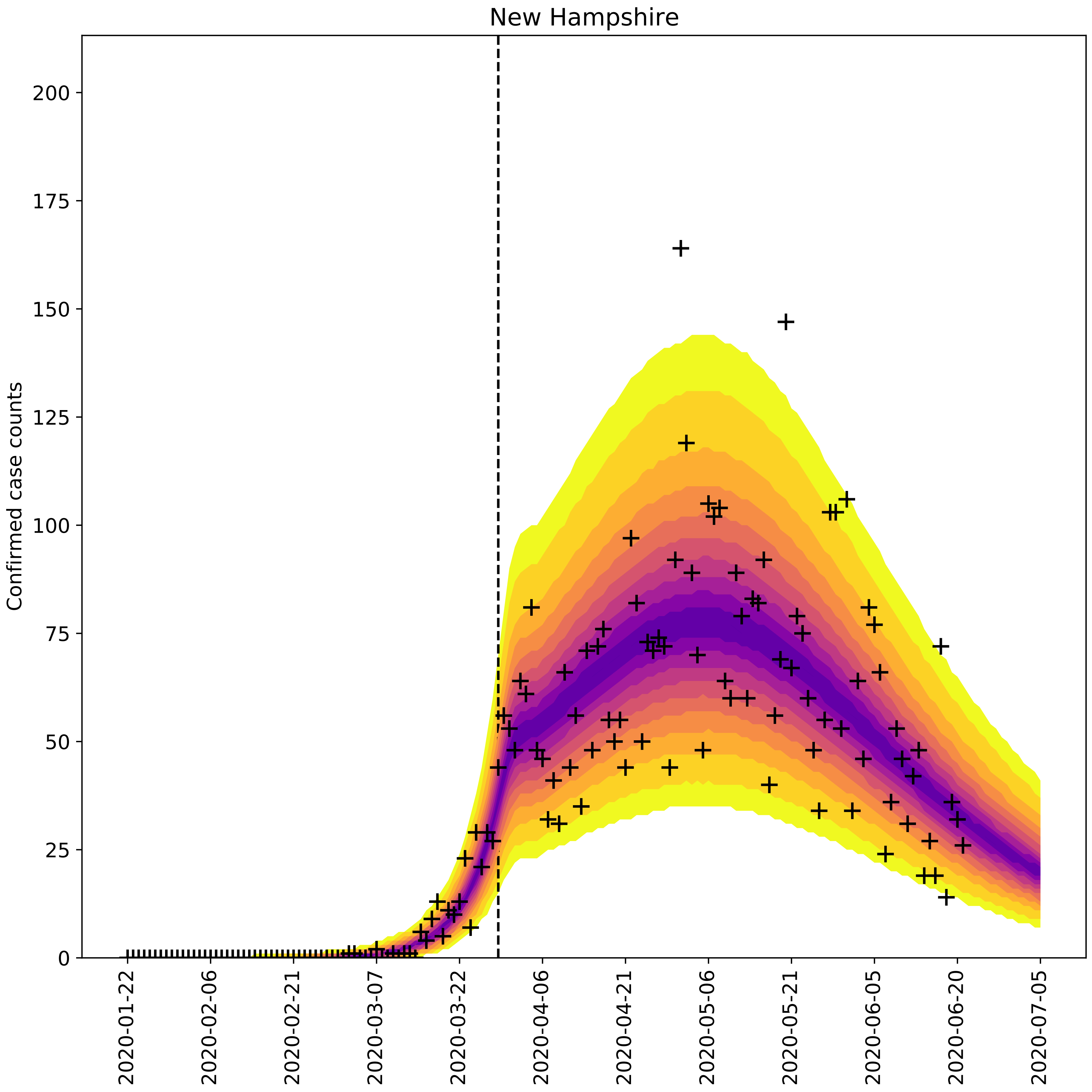
New Hampshire
Confirmed case counts
0
25
50
75
100
125
150
175
200
2020-01-22
2020-02-06
2020-02-21
2020-03-07
2020-03-22
2020-04-06
2020-04-21
2020-05-06
2020-05-21
2020-06-05
2020-06-20
2020-07-05

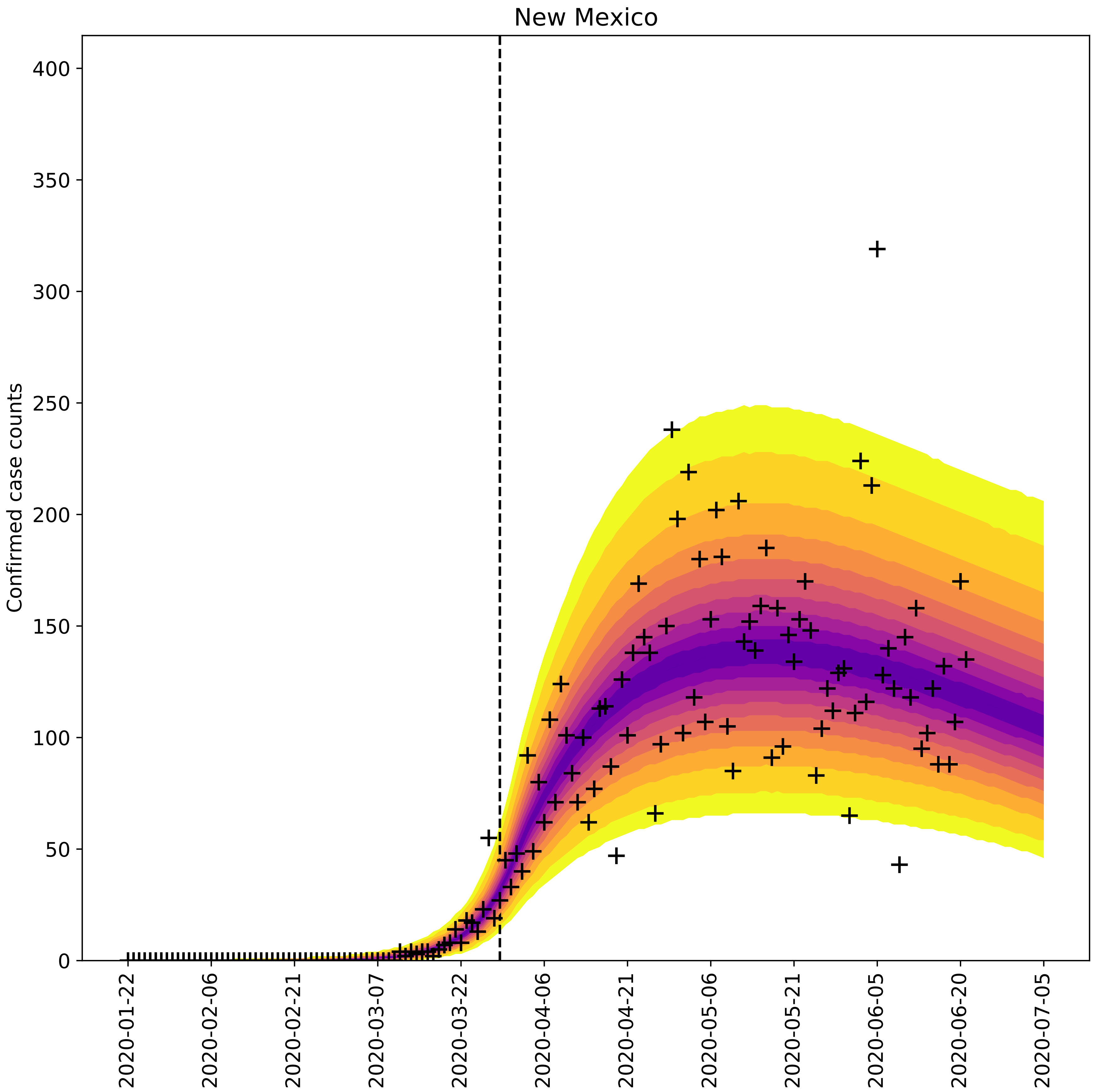

New Mexico
Confirmed case counts
0
50
100
150
200
250
300
350
400
2020-01-22
2020-02-06
2020-02-21
2020-03-07
2020-03-22
2020-04-06
2020-04-21
2020-05-06
2020-05-21
2020-06-05
2020-06-20
2020-07-05

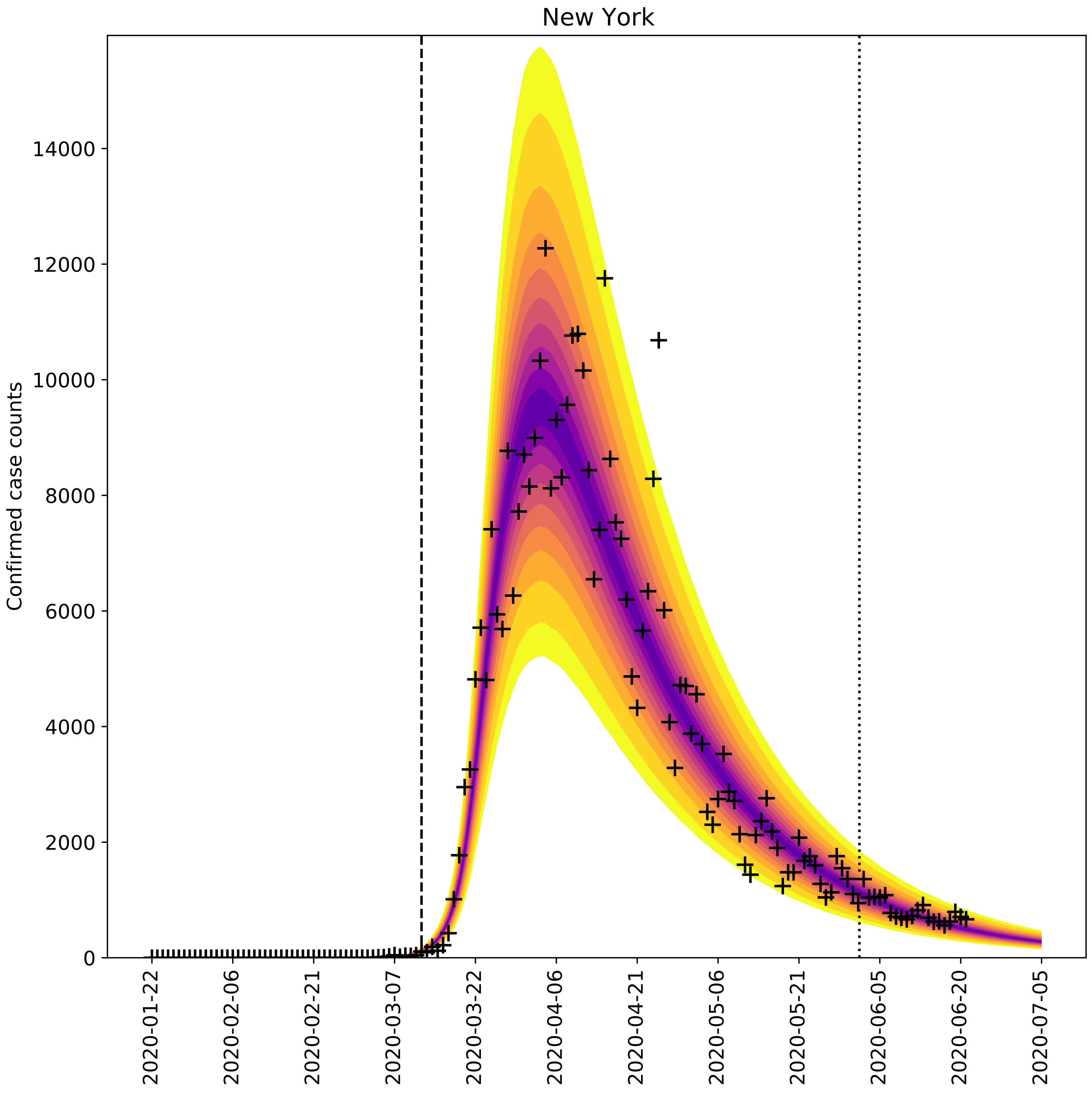
New York
Confirmed case counts
0
2000
4000
6000
8000
10000
12000
14000
2020-01-22
2020-02-06
2020-02-21
2020-03-07
2020-03-22
2020-04-06
2020-04-21
2020-05-06
2020-05-21
2020-06-05
2020-06-20
2020-07-05

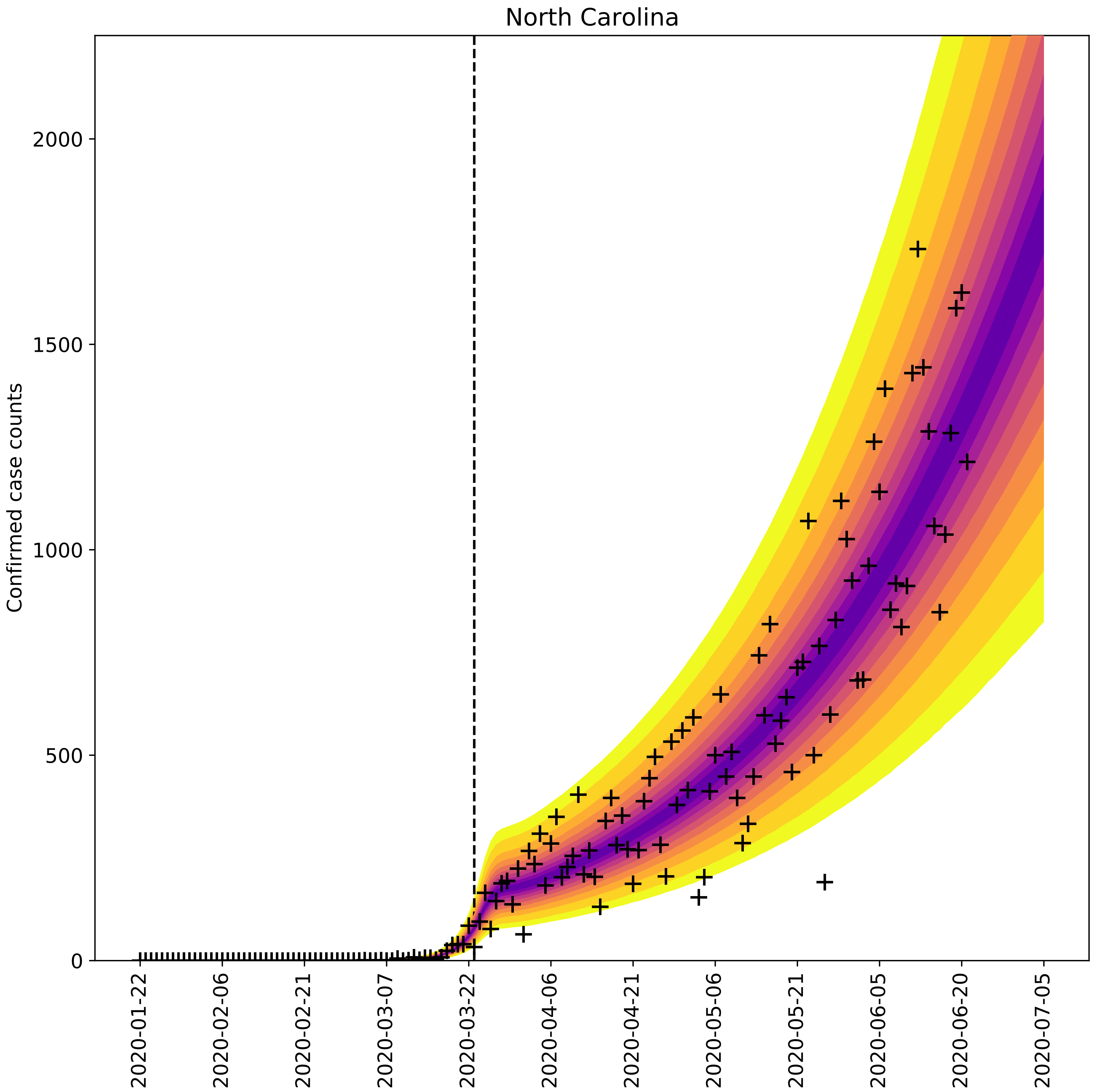
North Carolina
Confirmed case counts
0
500
1000
1500
2000
2020-01-22
2020-02-06
2020-02-21
2020-03-07
2020-03-22
2020-04-06
2020-04-21
2020-05-06
2020-05-21
2020-06-05
2020-06-20
2020-07-05

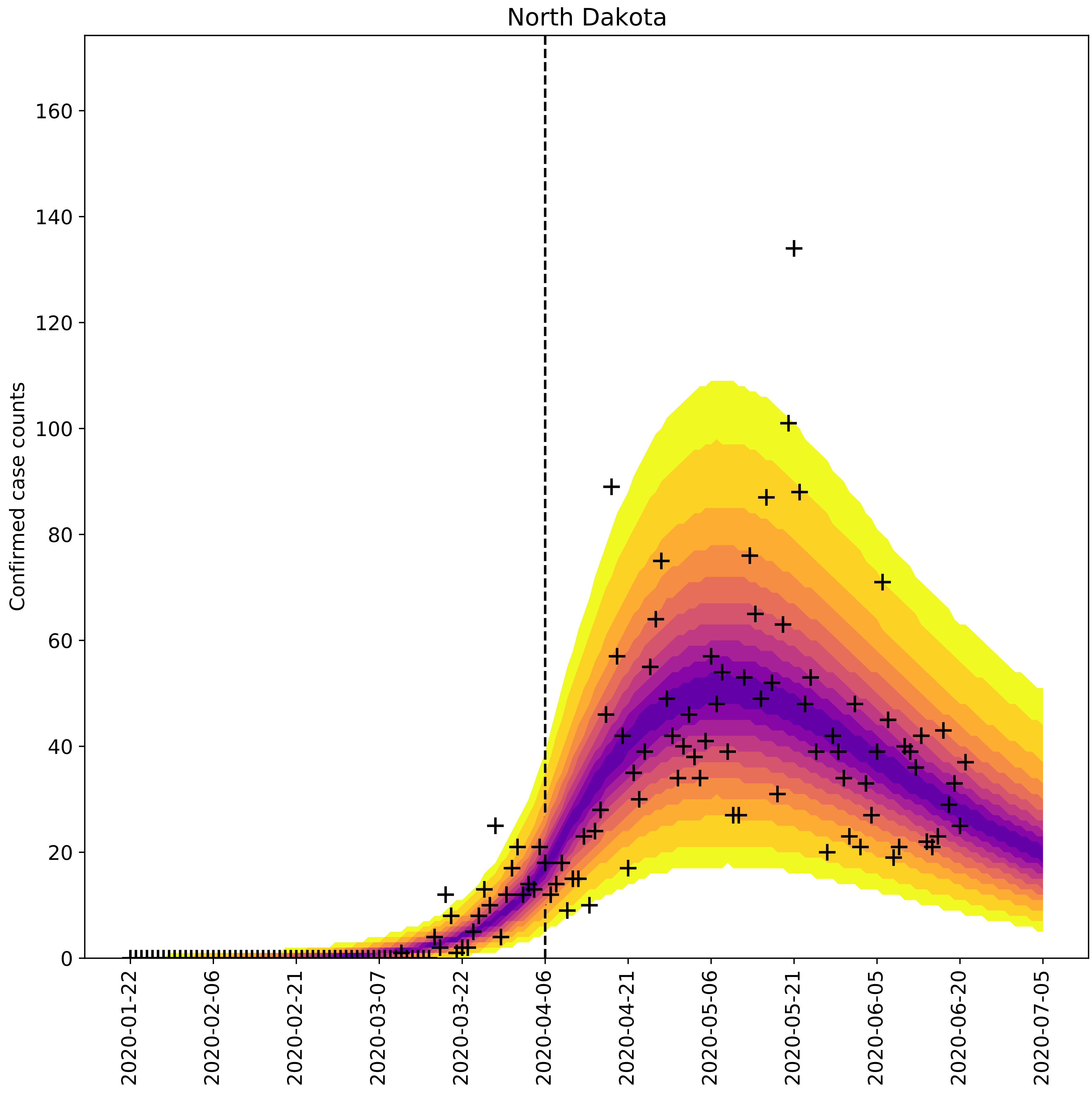
North Dakota
Confirmed case counts
0
20
40
60
80
100
120
140
160
2020-01-22
2020-02-06
2020-02-21
2020-03-07
2020-03-22
2020-04-06
2020-04-21
2020-05-06
2020-05-21
2020-06-05
2020-06-20
2020-07-05

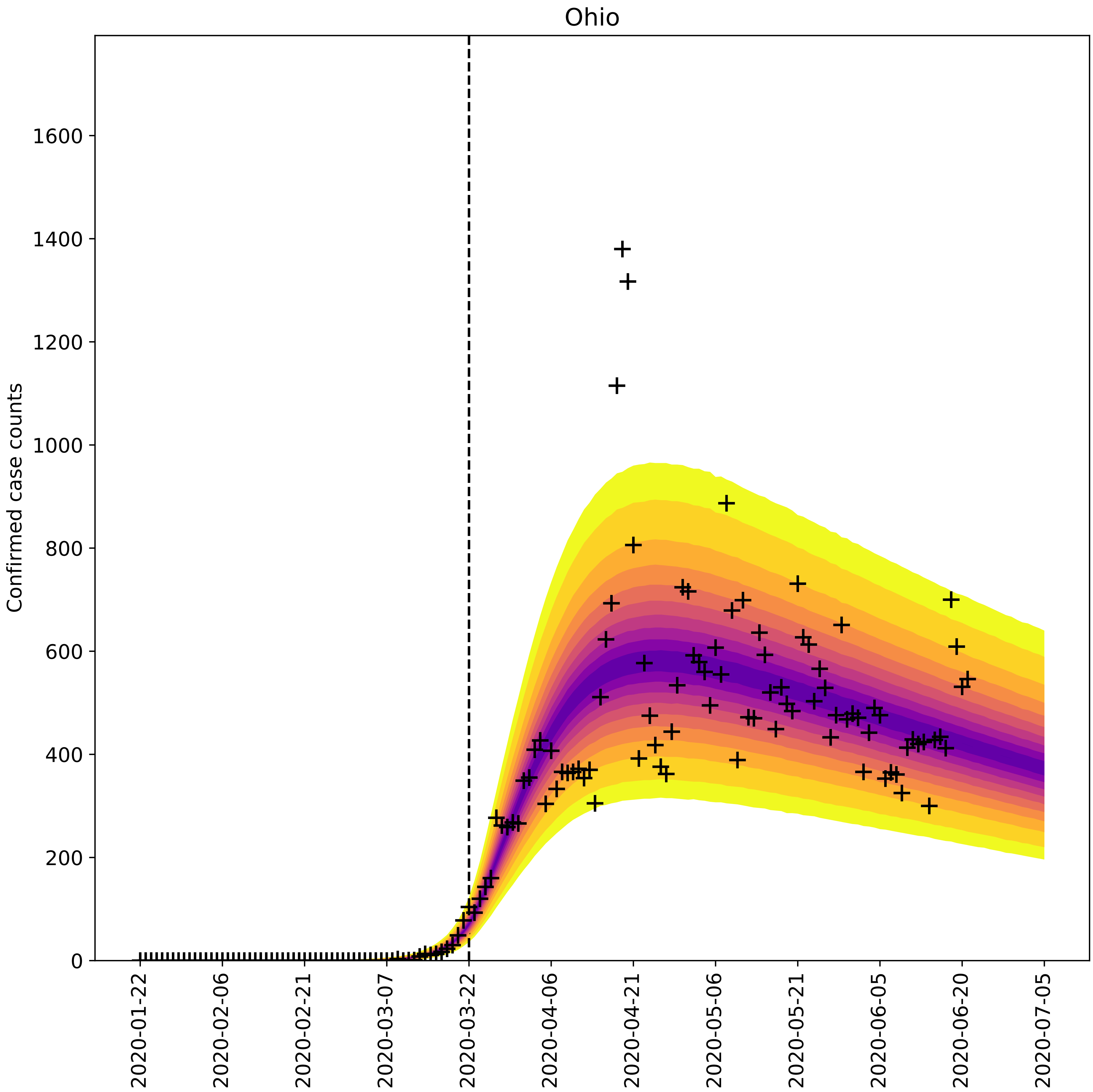

Ohio
Confirmed case counts
0
200
400
600
800
1000
1200
1400
1600
2020-01-22
2020-02-06
2020-02-21
2020-03-07
2020-03-22
2020-04-06
2020-04-21
2020-05-06
2020-05-21
2020-06-05
2020-06-20
2020-07-05

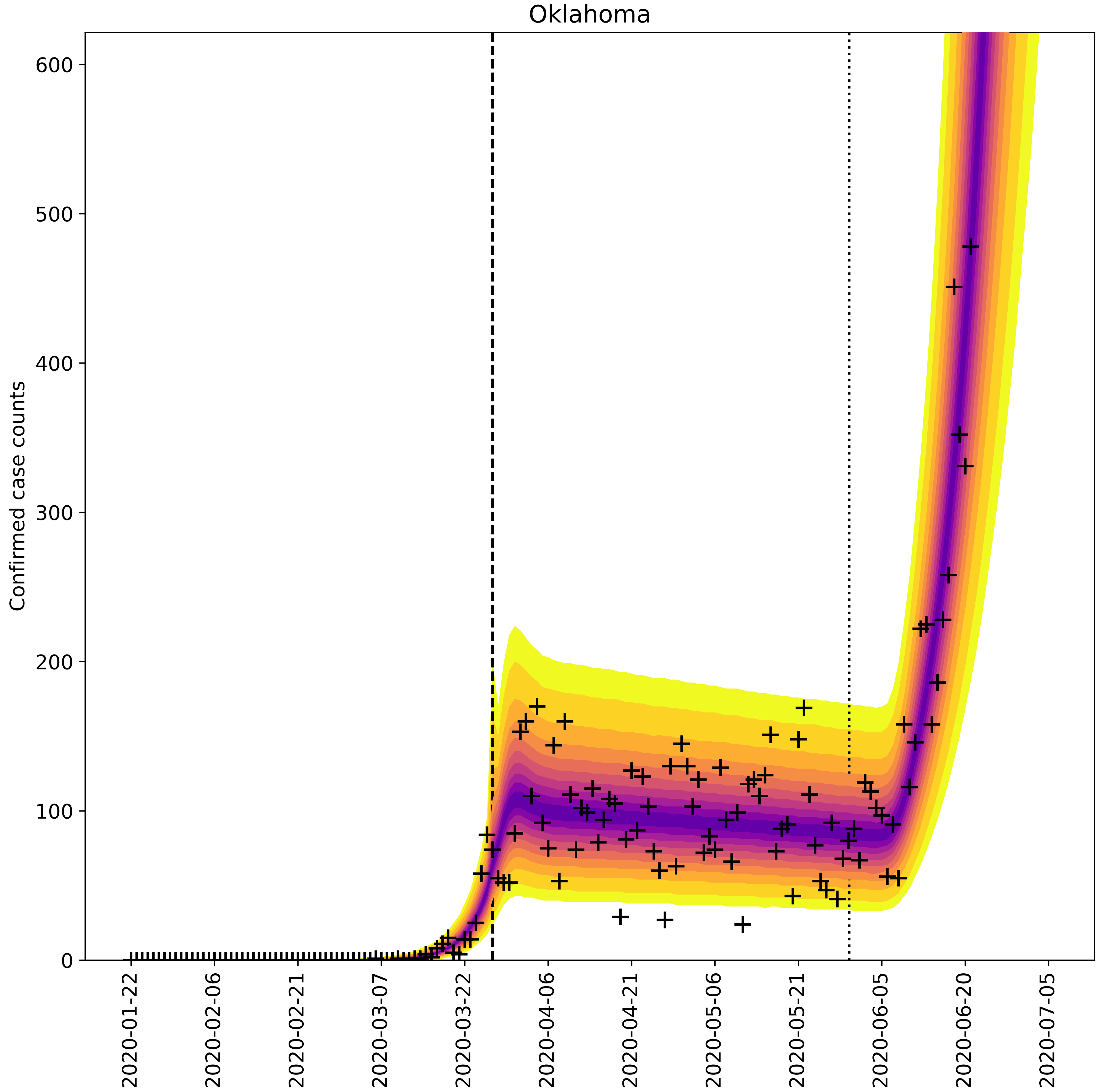
Oklahoma
Confirmed case counts
0
100
200
300
400
500
600
2020-01-22
2020-02-06
2020-02-21
2020-03-07
2020-03-22
2020-04-06
2020-04-21
2020-05-06
2020-05-21
2020-06-05
2020-06-20
2020-07-05

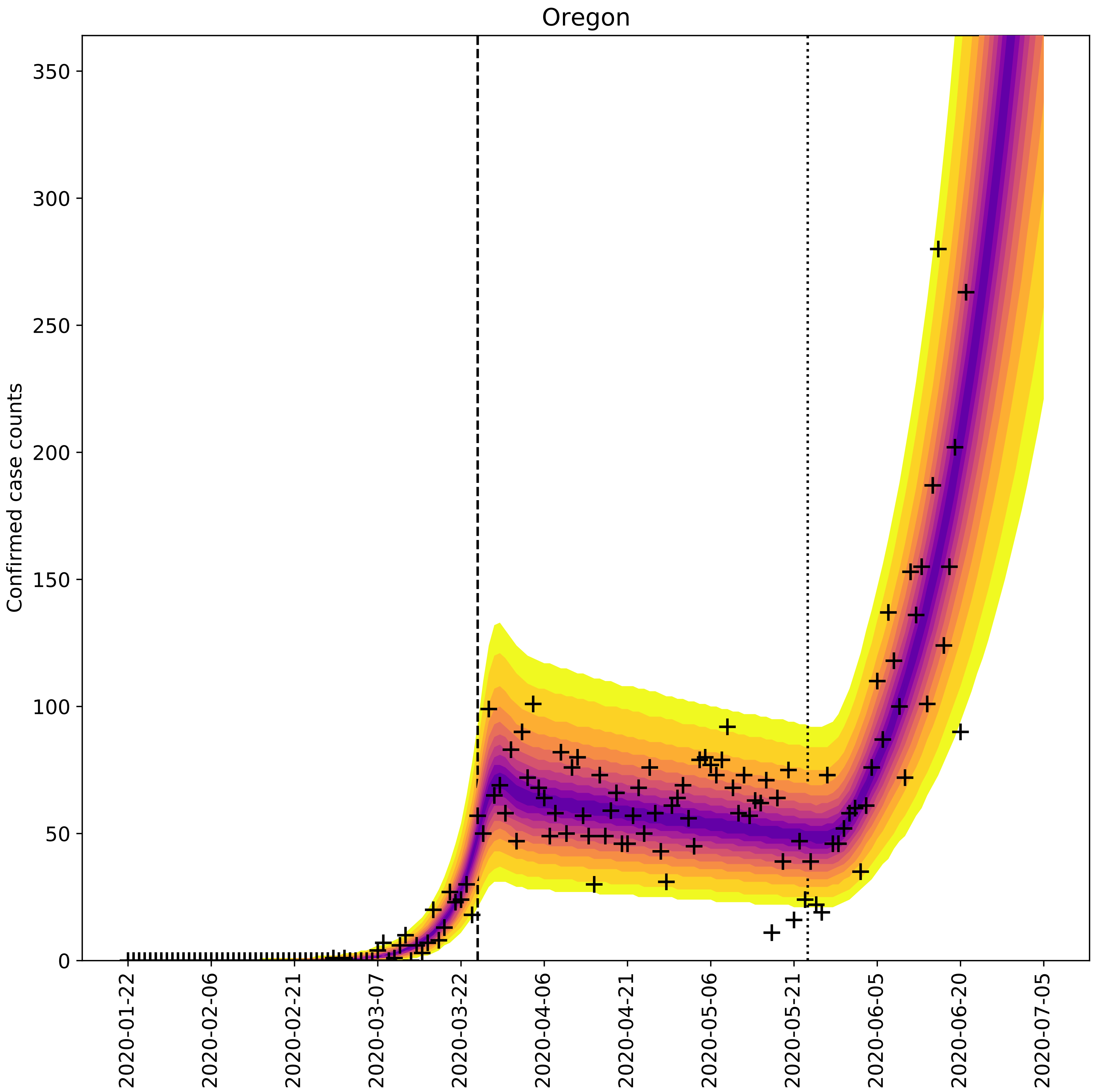

Oregon
Confirmed case counts
0
50
100
150
200
250
300
350
2020-01-22
2020-02-06
2020-02-21
2020-03-07
2020-03-22
2020-04-06
2020-04-21
2020-05-06
2020-05-21
2020-06-05
2020-06-20
2020-07-05

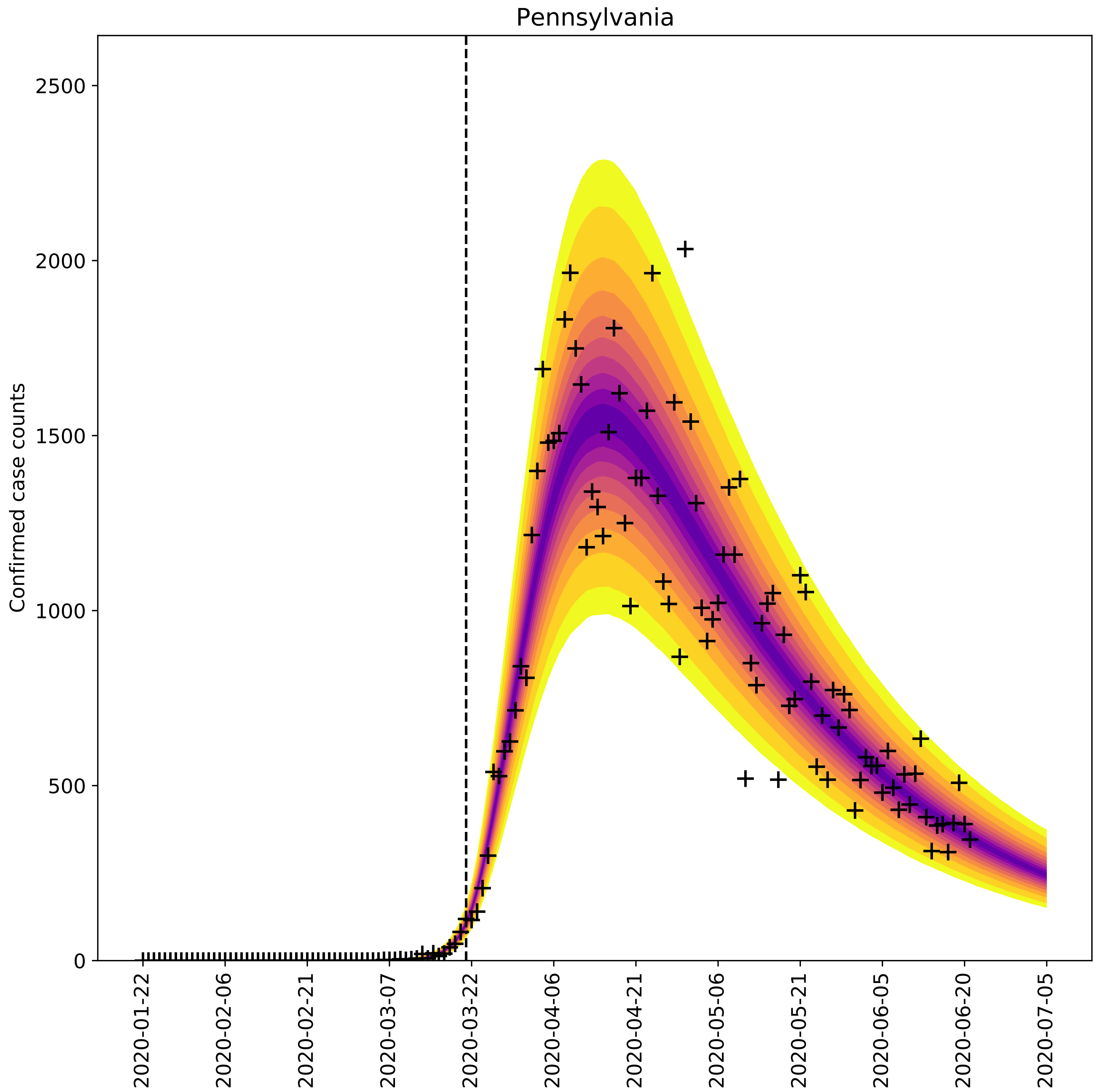

Pennsylvania
Confirmed case counts
0
500
1000
1500
2000
2500
2020-01-22
2020-02-06
2020-02-21
2020-03-07
2020-03-22
2020-04-06
2020-04-21
2020-05-06
2020-05-21
2020-06-05
2020-06-20
2020-07-05

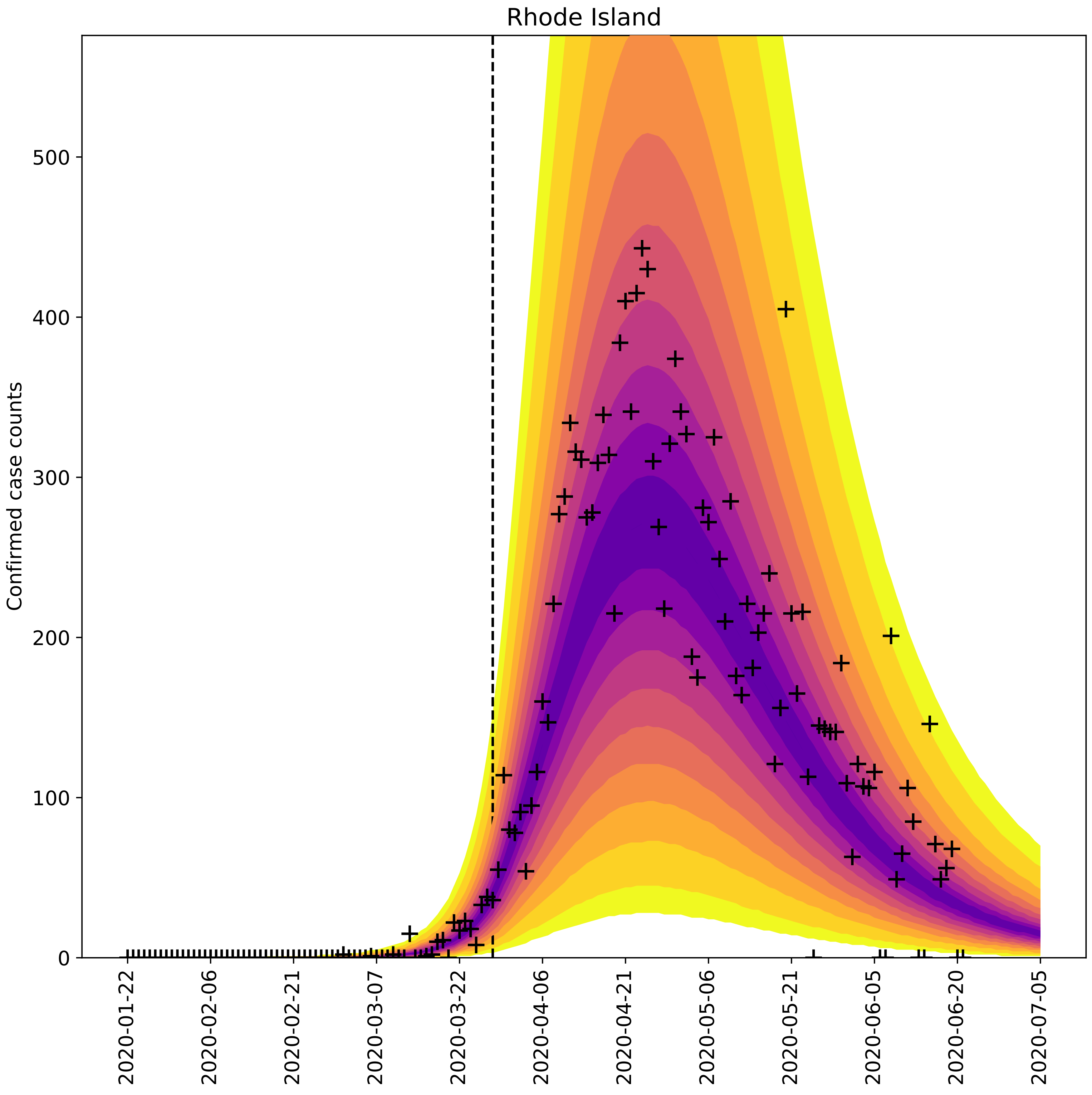

Rhode Island
Confirmed case counts
0
100
200
300
400
500
2020-01-22
2020-02-06
2020-02-21
2020-03-07
2020-03-22
2020-04-06
2020-04-21
2020-05-06
2020-05-21
2020-06-05
2020-06-20
2020-07-05

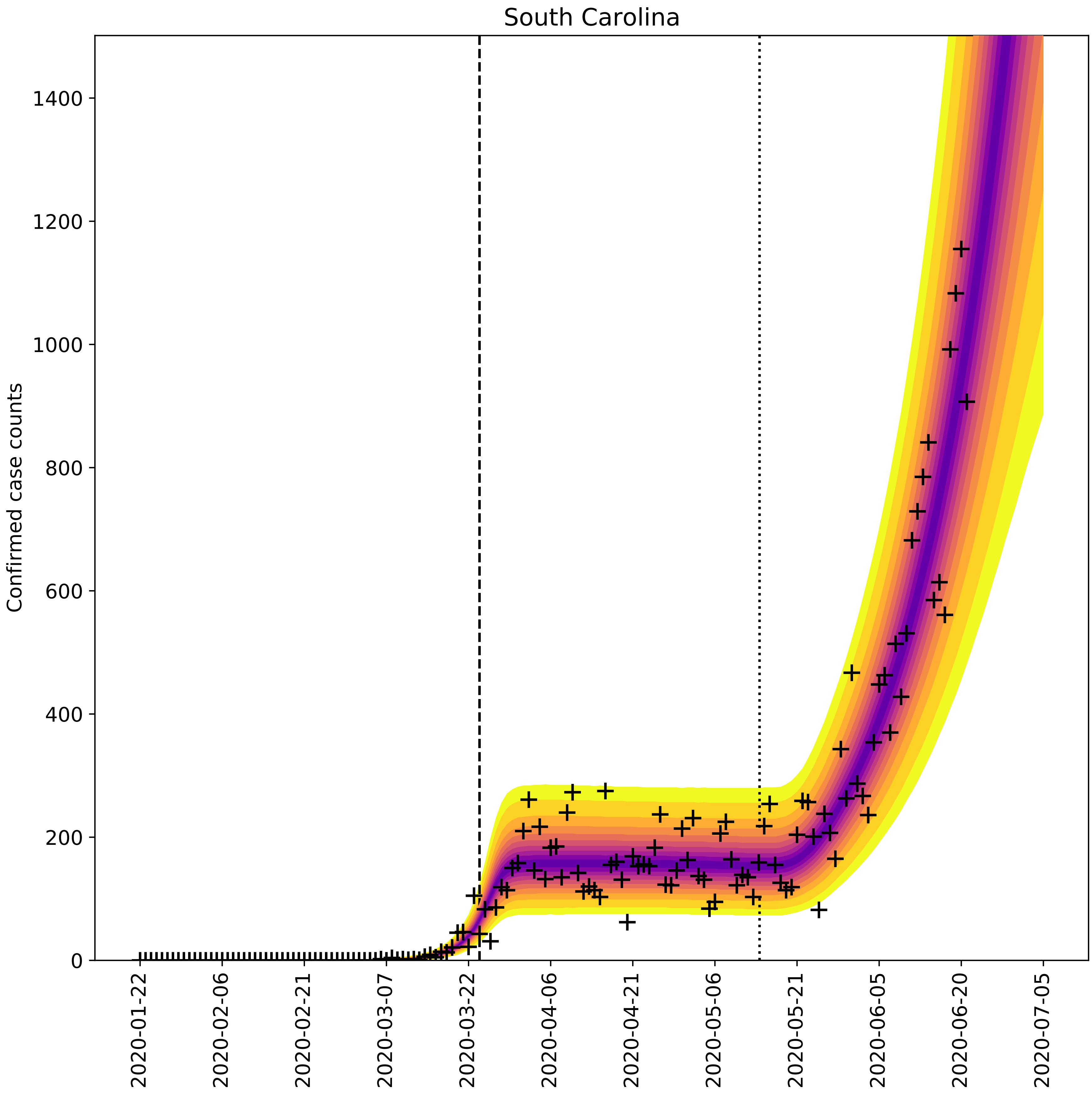

South Carolina
Confirmed case counts
0
200
400
600
800
1000
1200
1400
2020-01-22
2020-02-06
2020-02-21
2020-03-07
2020-03-22
2020-04-06
2020-04-21
2020-05-06
2020-05-21
2020-06-05
2020-06-20
2020-07-05

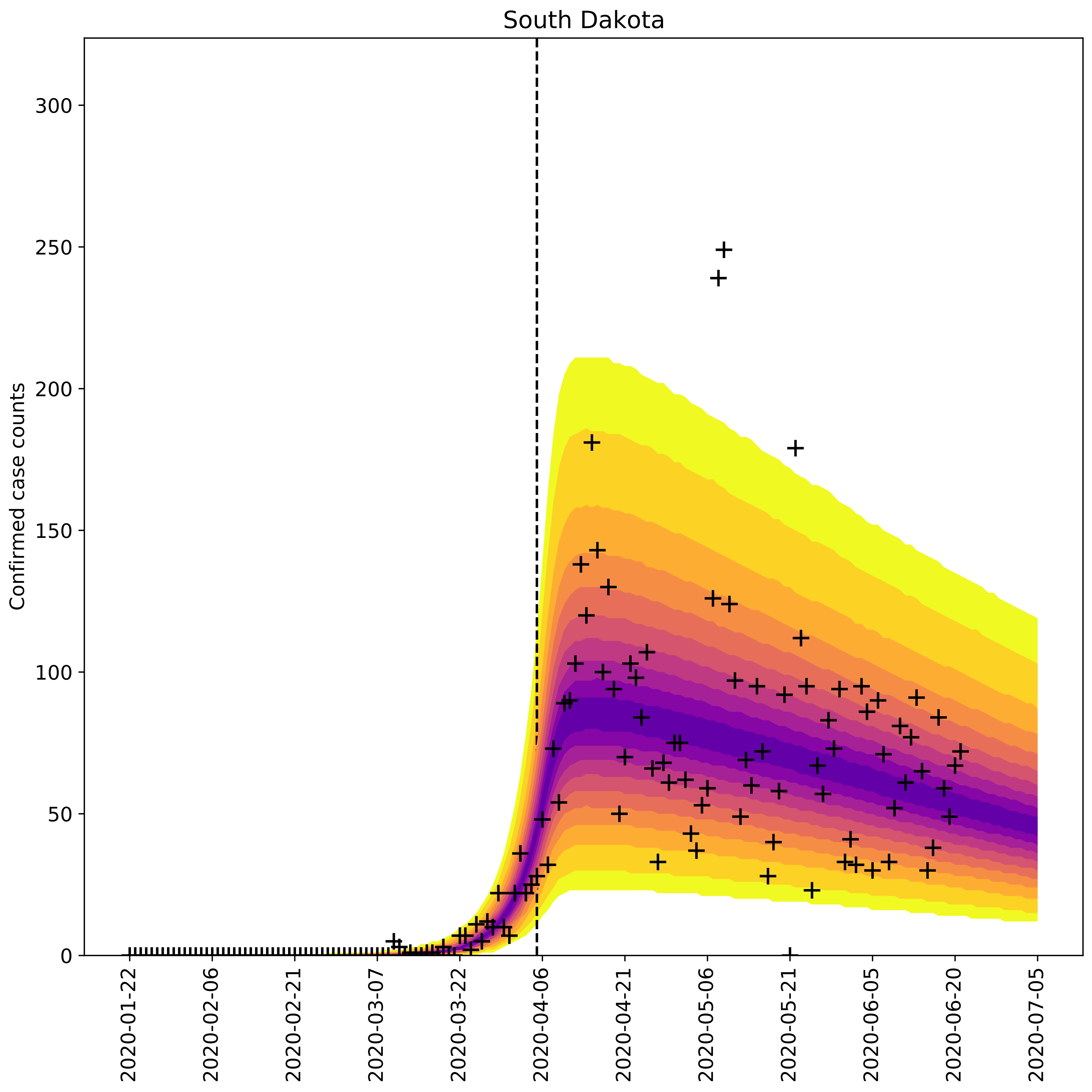

South Dakota
Confirmed case counts
0
50
100
150
200
250
300
2020-01-22
2020-02-06
2020-02-21
2020-03-07
2020-03-22
2020-04-06
2020-04-21
2020-05-06
2020-05-21
2020-06-05
2020-06-20
2020-07-05

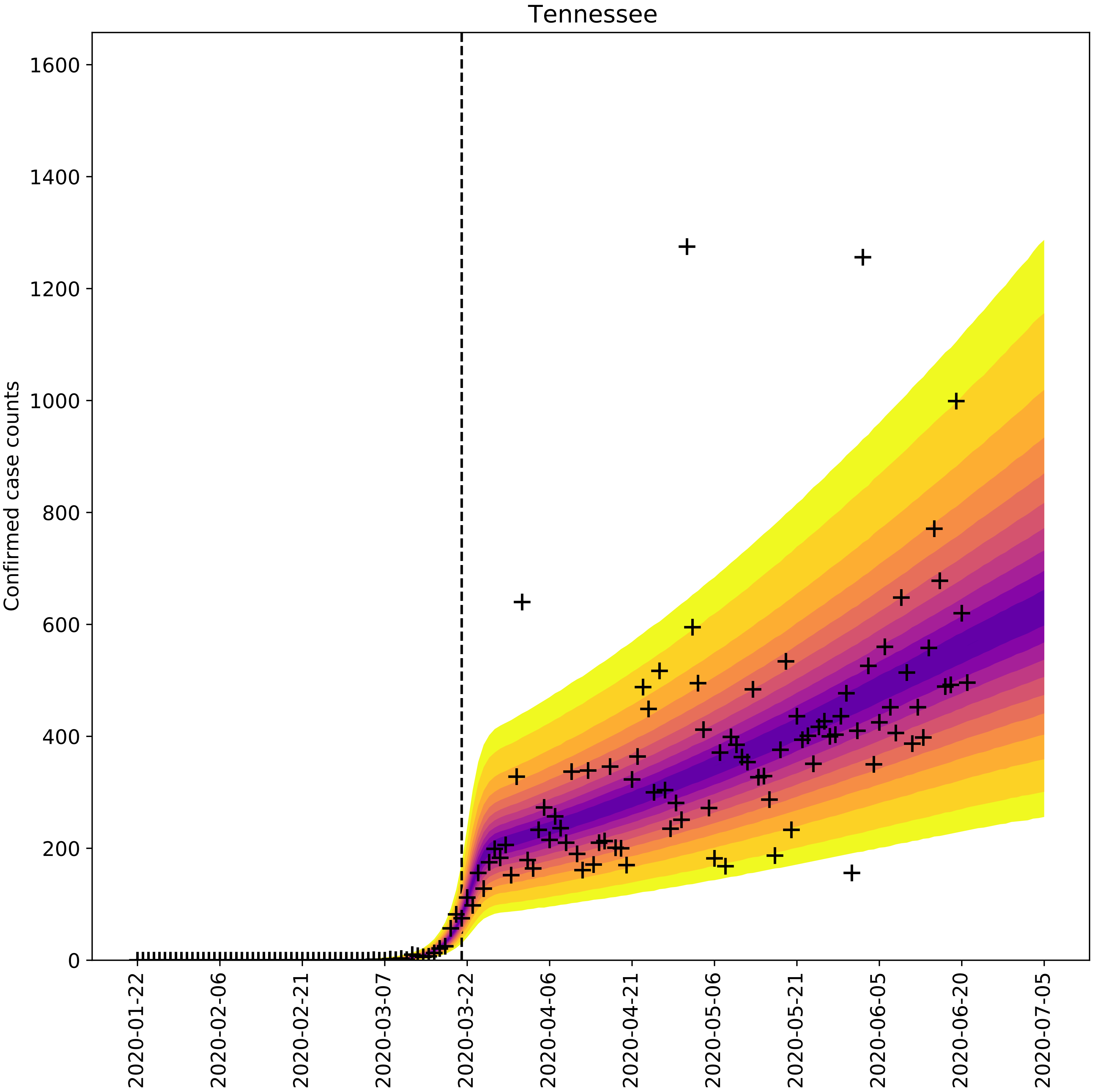

Tennessee
Confirmed case counts
0
200
400
600
800
1000
1200
1400
1600
2020-01-22
2020-02-06
2020-02-21
2020-03-07
2020-03-22
2020-04-06
2020-04-21
2020-05-06
2020-05-21
2020-06-05
2020-06-20
2020-07-05

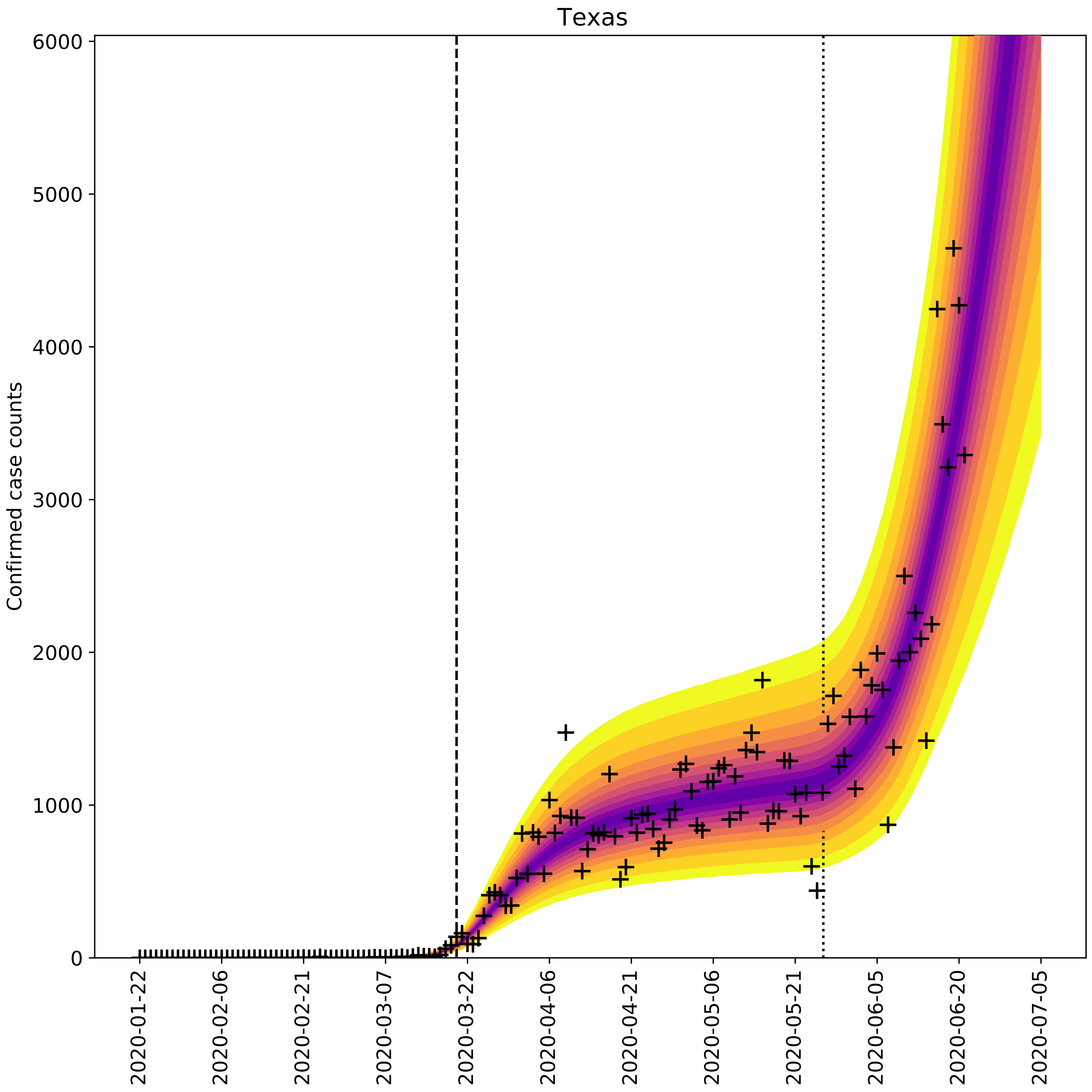

Texas
Confirmed case counts
0
1000
2000
3000
4000
5000
6000
2020-01-22
2020-02-06
2020-02-21
2020-03-07
2020-03-22
2020-04-06
2020-04-21
2020-05-06
2020-05-21
2020-06-05
2020-06-20
2020-07-05

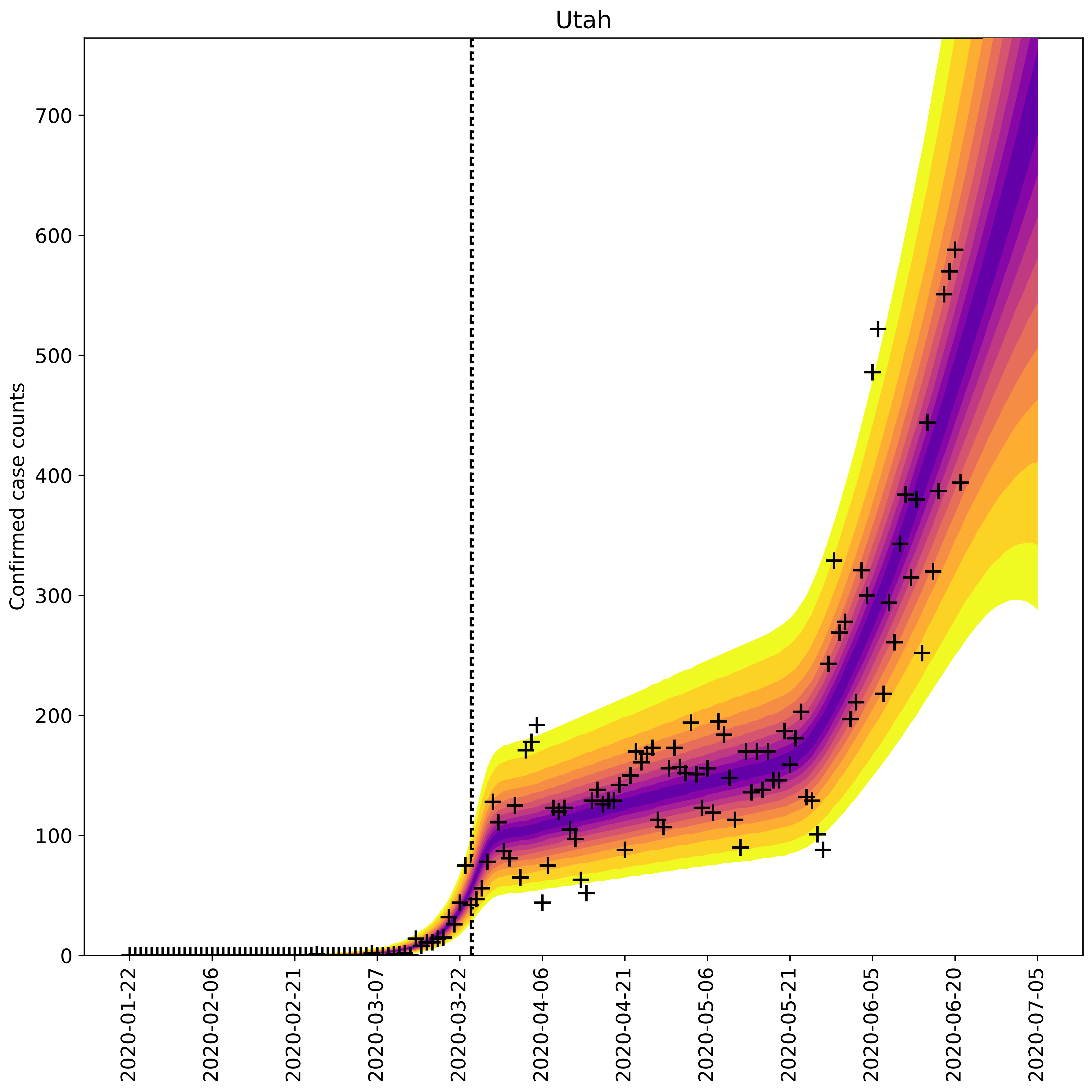

Utah
Confirmed case counts
0
100
200
300
400
500
600
700
2020-01-22
2020-02-06
2020-02-21
2020-03-07
2020-03-22
2020-04-06
2020-04-21
2020-05-06
2020-05-21
2020-06-05
2020-06-20
2020-07-05

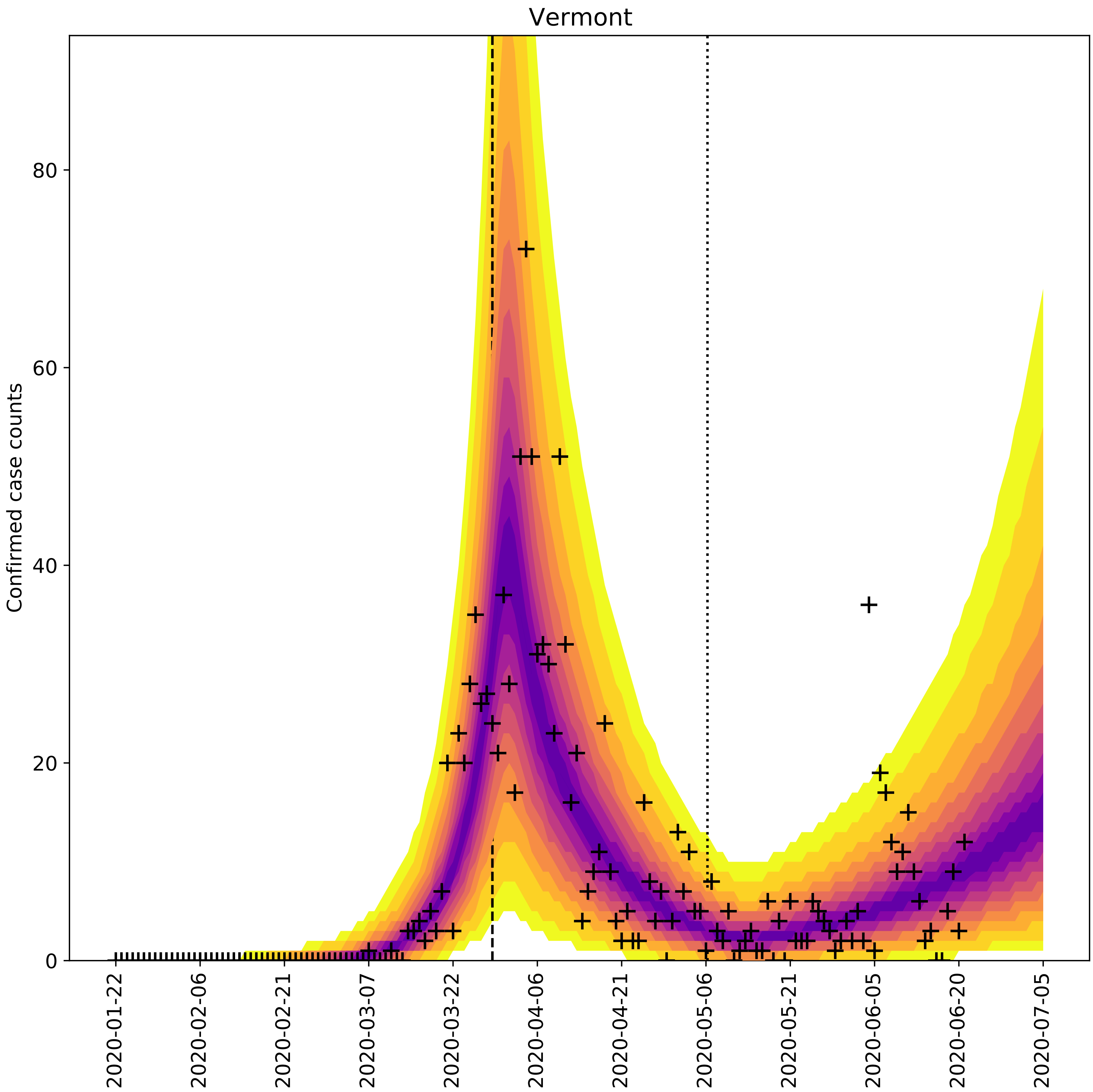

Vermont
Confirmed case counts
0
20
40
60
80
2020-01-22
2020-02-06
2020-02-21
2020-03-07
2020-03-22
2020-04-06
2020-04-21
2020-05-06
2020-05-21
2020-06-05
2020-06-20
2020-07-05

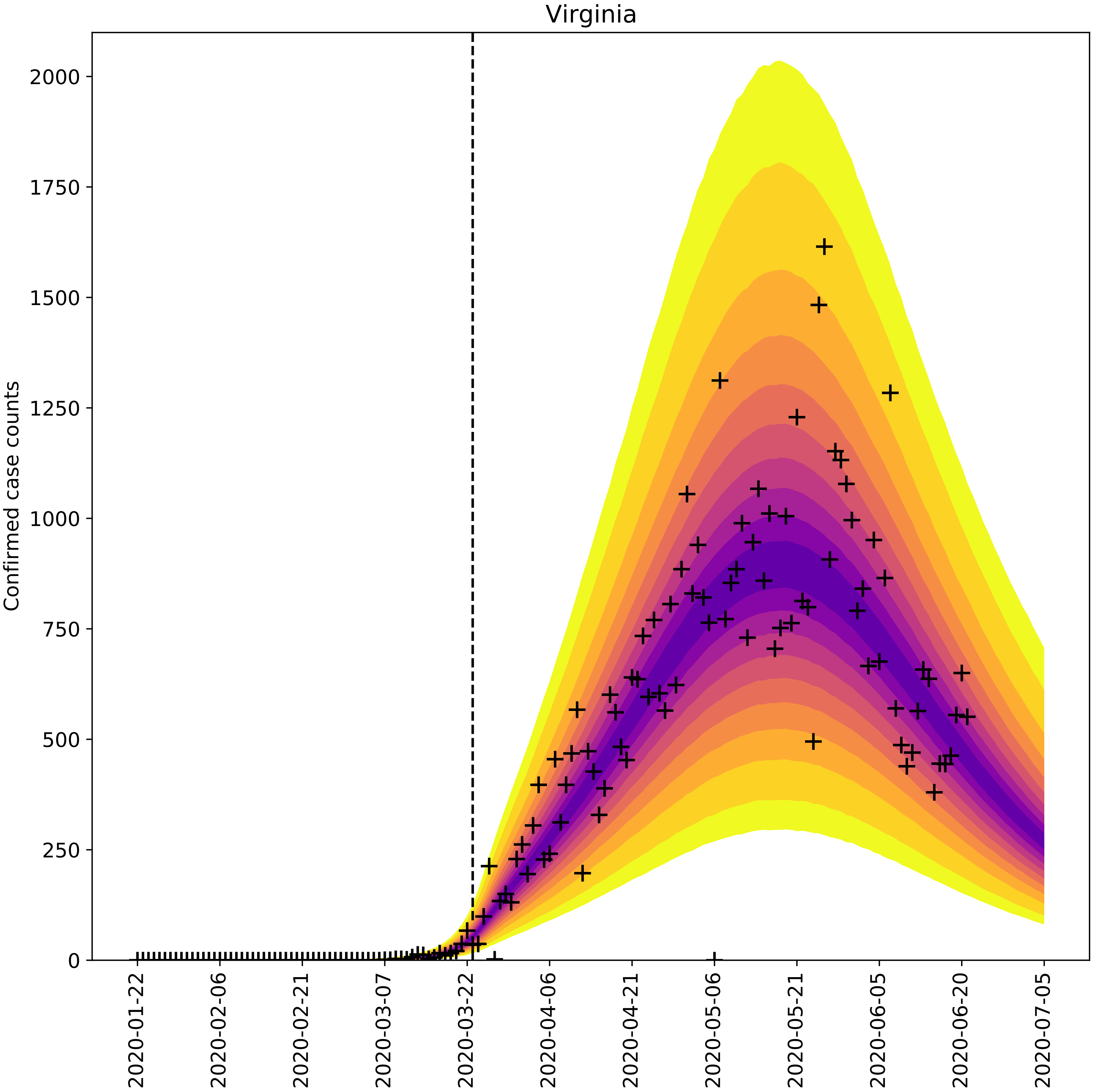
Virginia
Confirmed case counts
0
250
500
750
1000
1250
1500
1750
2000
2020-01-22
2020-02-06
2020-02-21
2020-03-07
2020-03-22
2020-04-06
2020-04-21
2020-05-06
2020-05-21
2020-06-05
2020-06-20
2020-07-05

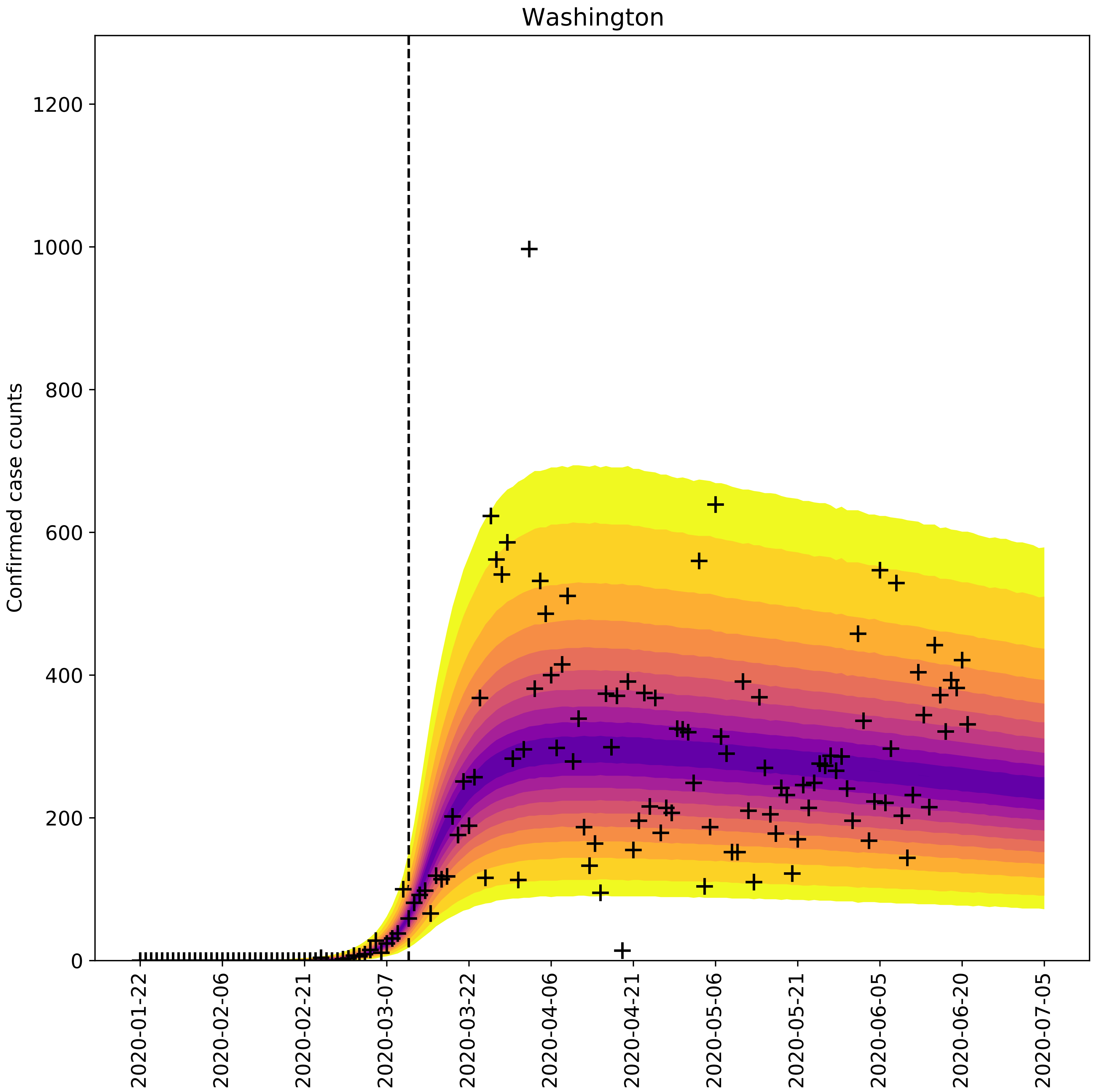

Washington
Confirmed case counts
0
200
400
600
800
1000
1200
2020-01-22
2020-02-06
2020-02-21
2020-03-07
2020-03-22
2020-04-06
2020-04-21
2020-05-06
2020-05-21
2020-06-05
2020-06-20
2020-07-05

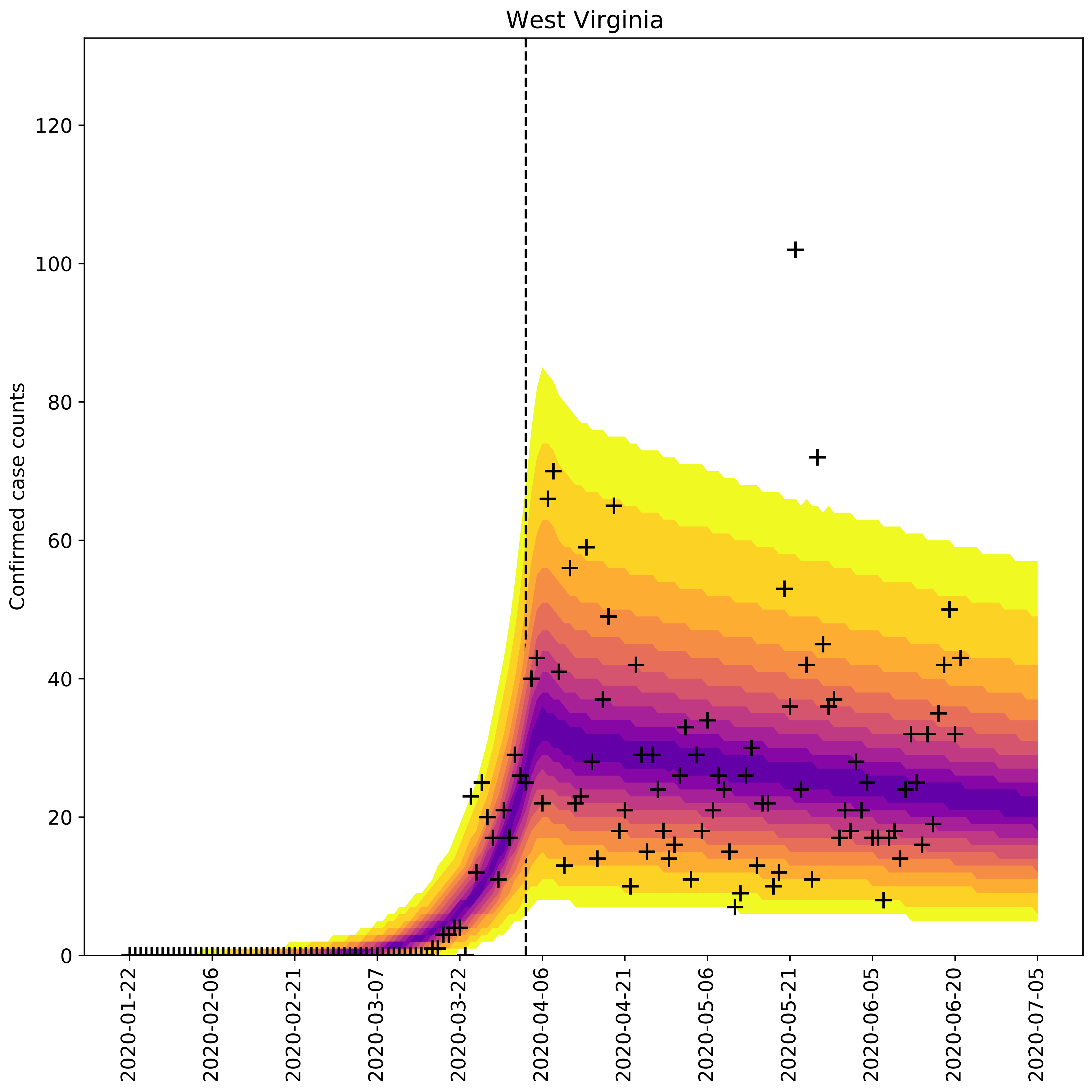

West Virginia
Confirmed case counts
0
20
40
60
80
100
120
2020-01-22
2020-02-06
2020-02-21
2020-03-07
2020-03-22
2020-04-06
2020-04-21
2020-05-06
2020-05-21
2020-06-05
2020-06-20
2020-07-05

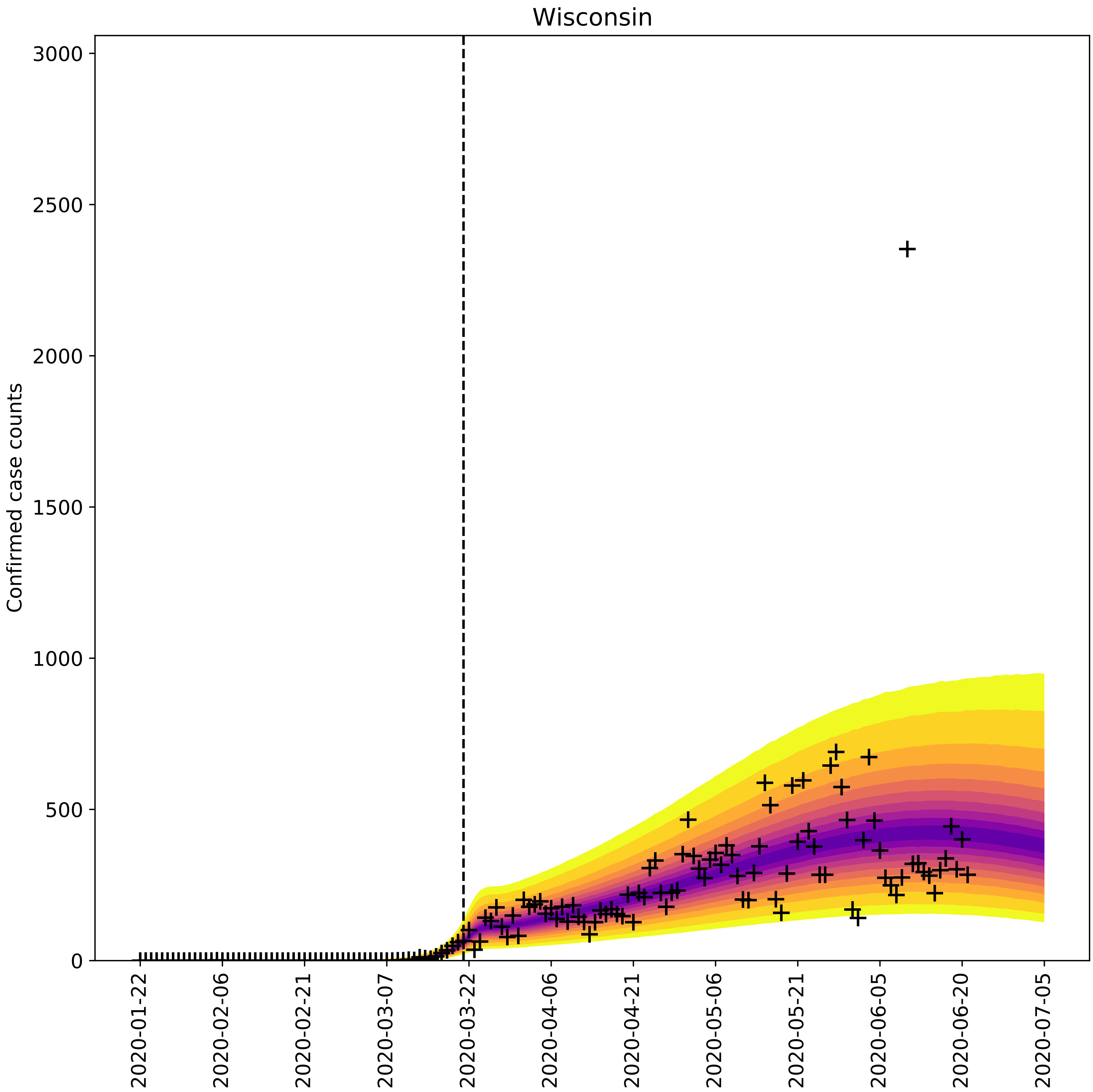

Wisconsin
Confirmed case counts
0
500
1000
1500
2000
2500
3000
2020-01-22
2020-02-06
2020-02-21
2020-03-07
2020-03-22
2020-04-06
2020-04-21
2020-05-06
2020-05-21
2020-06-05
2020-06-20
2020-07-05

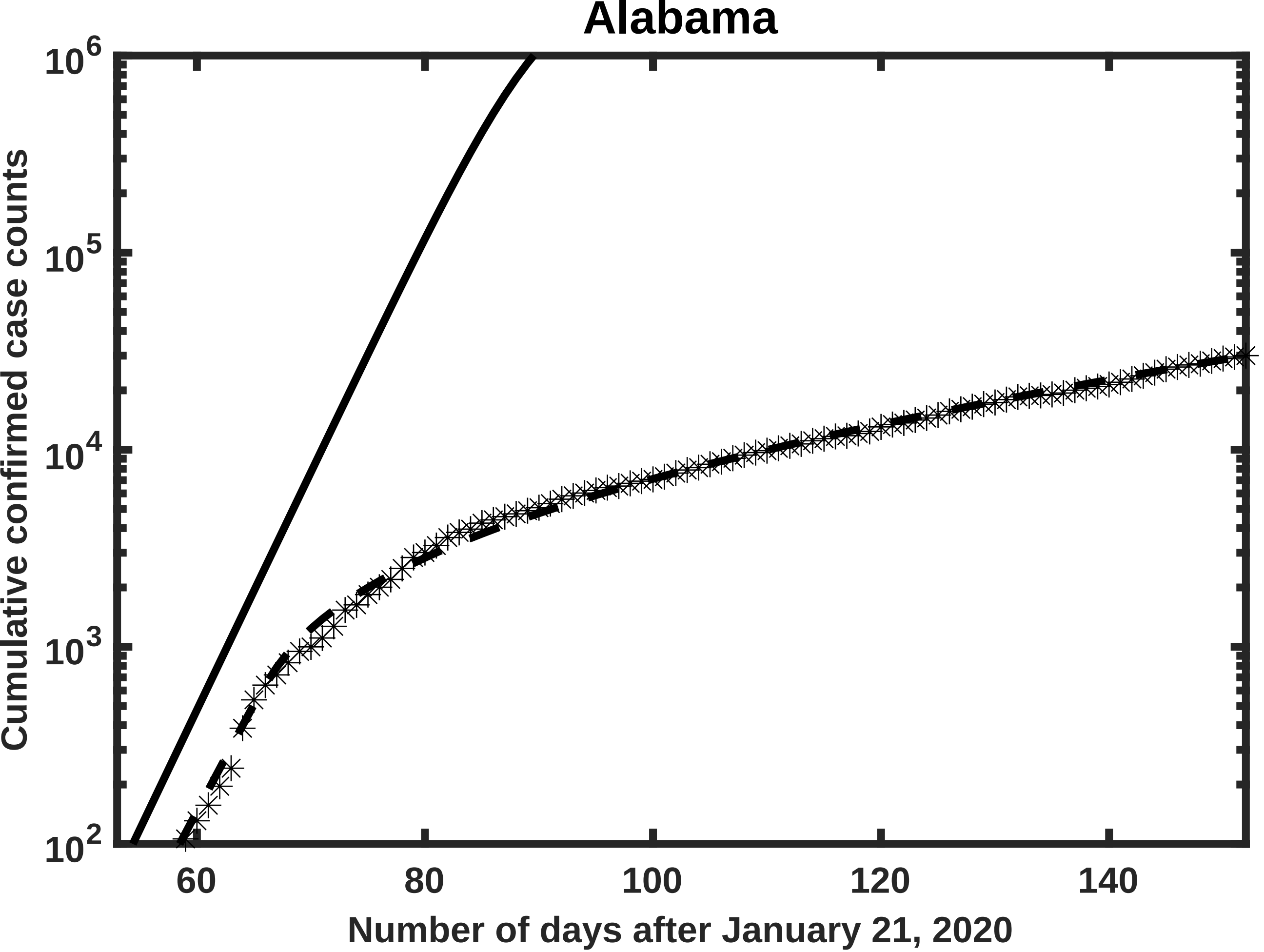

Alabama
Cumulative confirmed case counts
$10^2$
$10^3$
$10^4$
$10^5$
$10^6$
60
80
100
120
140
Number of days after January 21, 2020

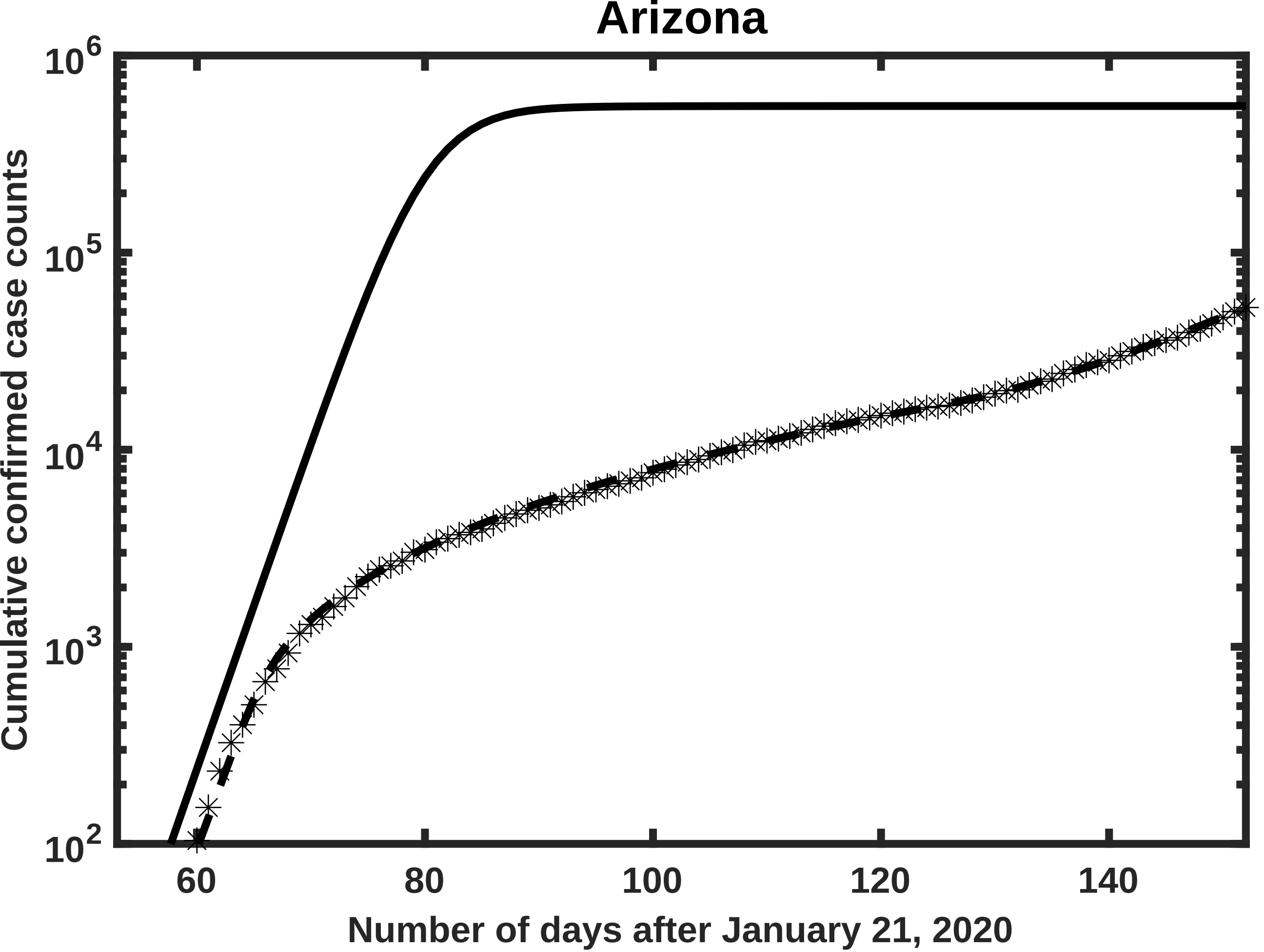

Arizona
Cumulative confirmed case counts
Number of days after January 21, 2020
$10^2$
$10^3$
$10^4$
$10^5$
$10^6$
60
80
100
120
140

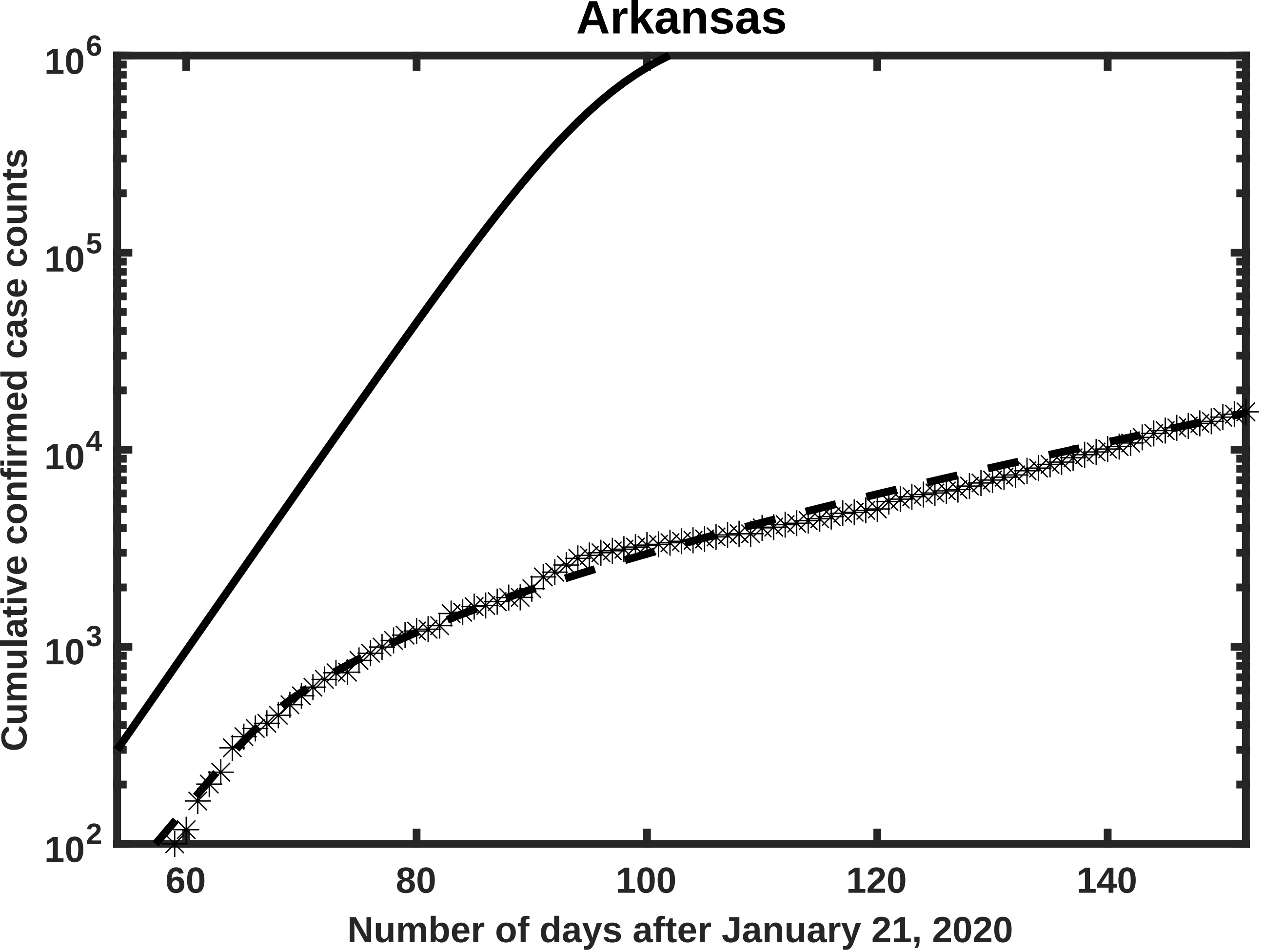

Arkansas
$10^6$
$10^5$
$10^4$
$10^3$
$10^2$
60
80
100
120
140
Number of days after January 21, 2020
Cumulative confirmed case counts

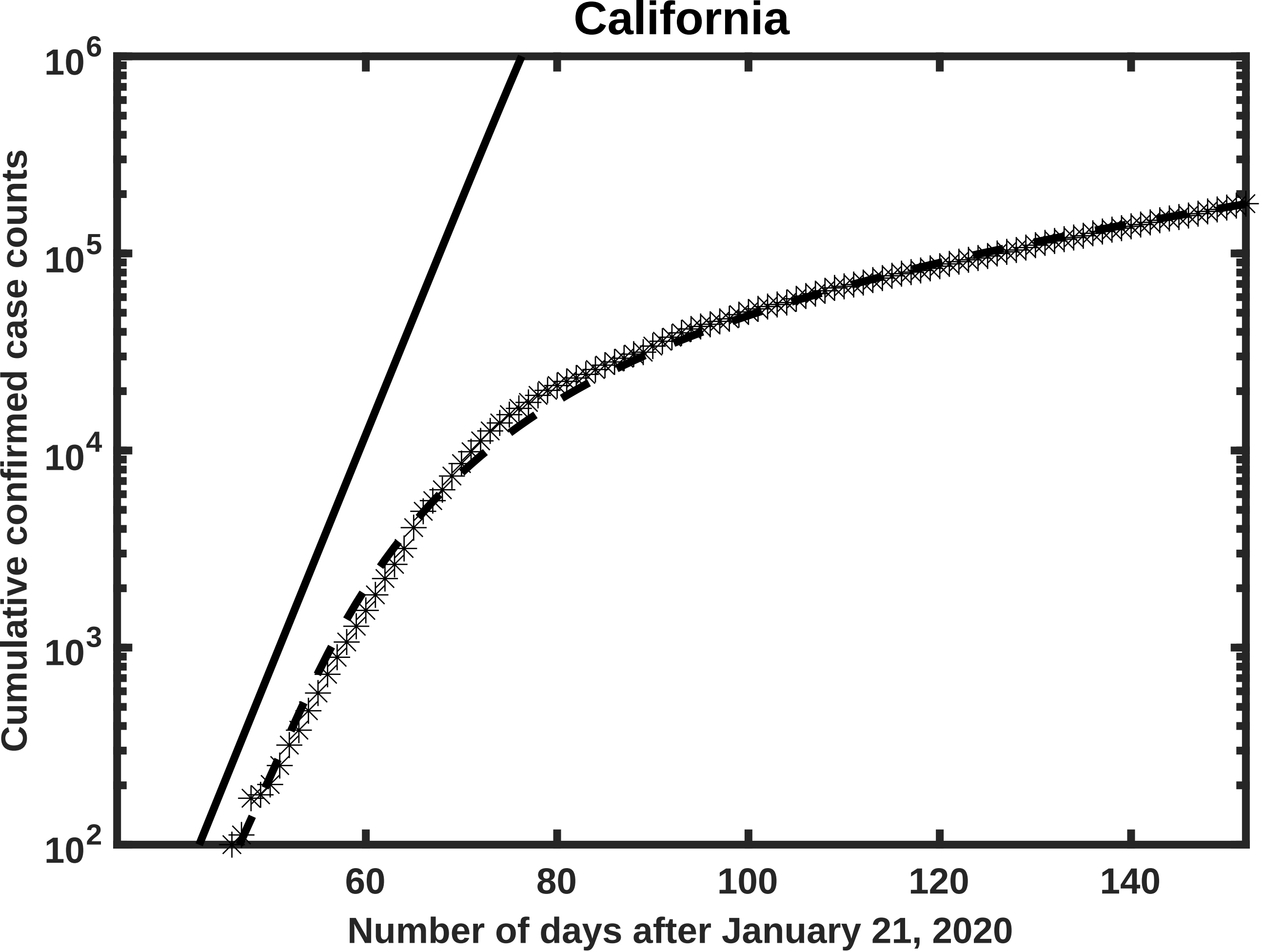
California
Cumulative confirmed case counts
$10^6$
$10^5$
$10^4$
$10^3$
$10^2$
60
80
100
120
140
Number of days after January 21, 2020

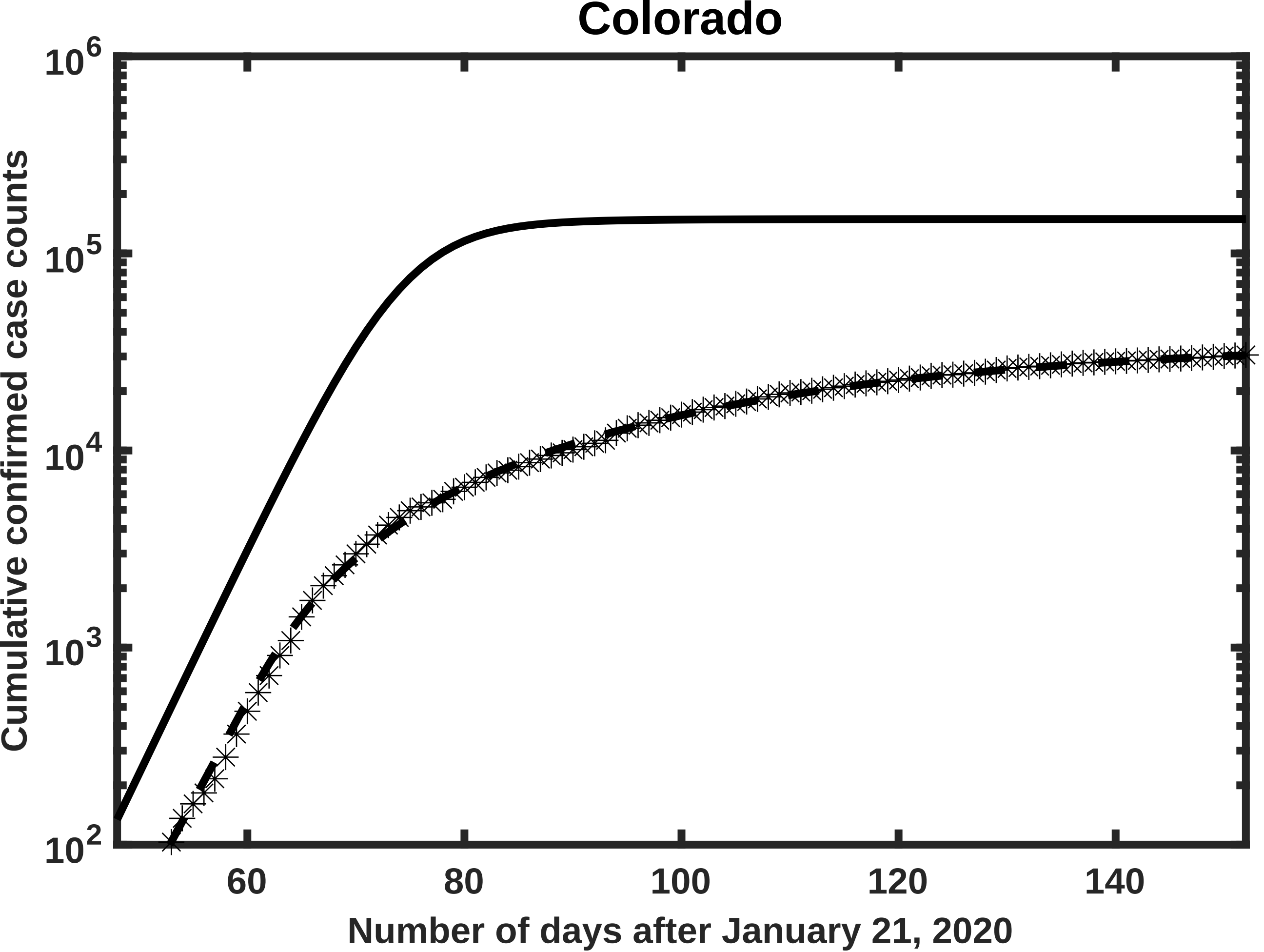

Colorado
$10^6$
$10^5$
$10^4$
$10^3$
$10^2$
60
80
100
120
140
Cumulative confirmed case counts
Number of days after January 21, 2020

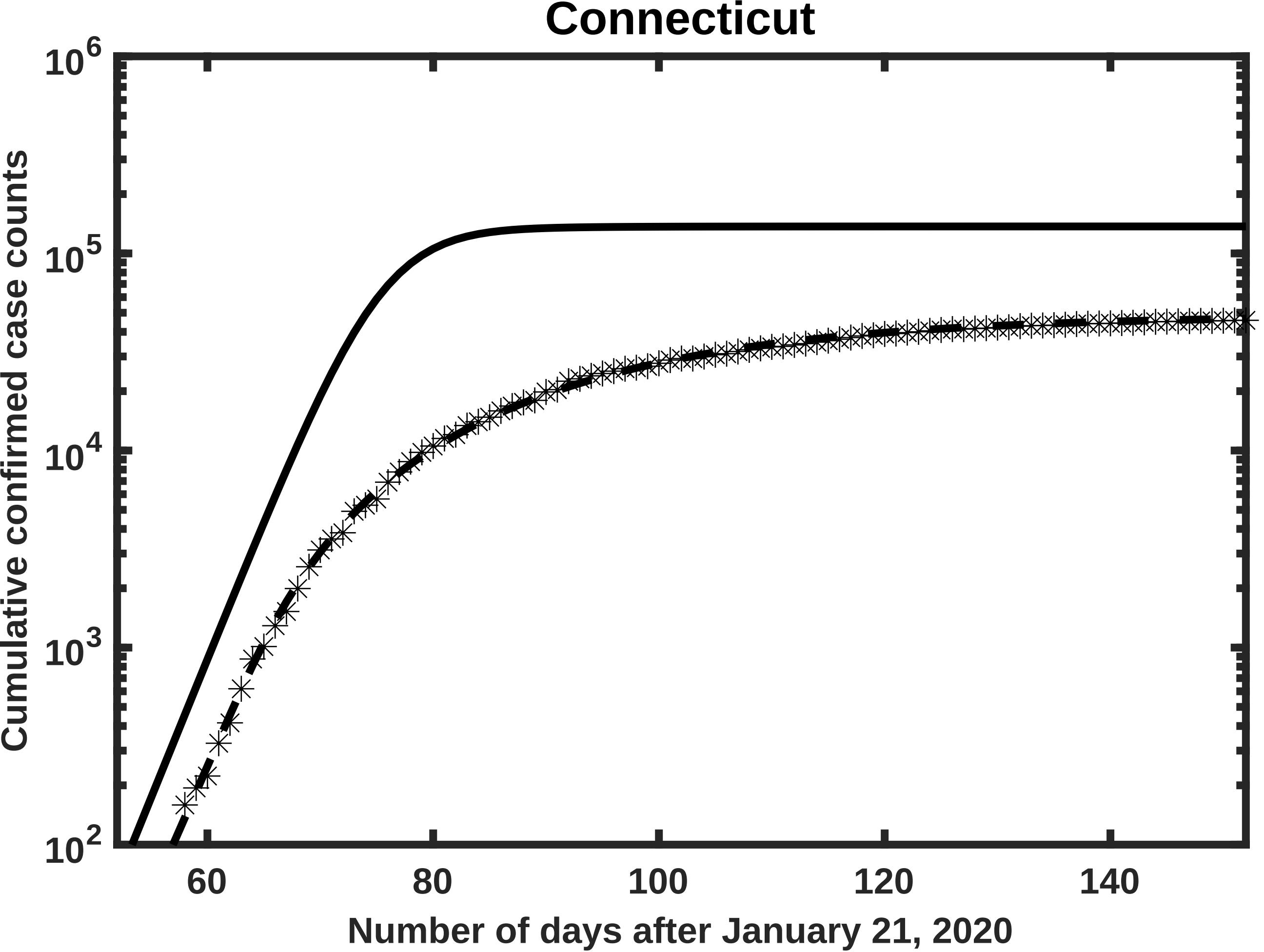

Connecticut
Cumulative confirmed case counts
10^6
10^5
10^4
10^3
10^2
60
80
100
120
140
Number of days after January 21, 2020

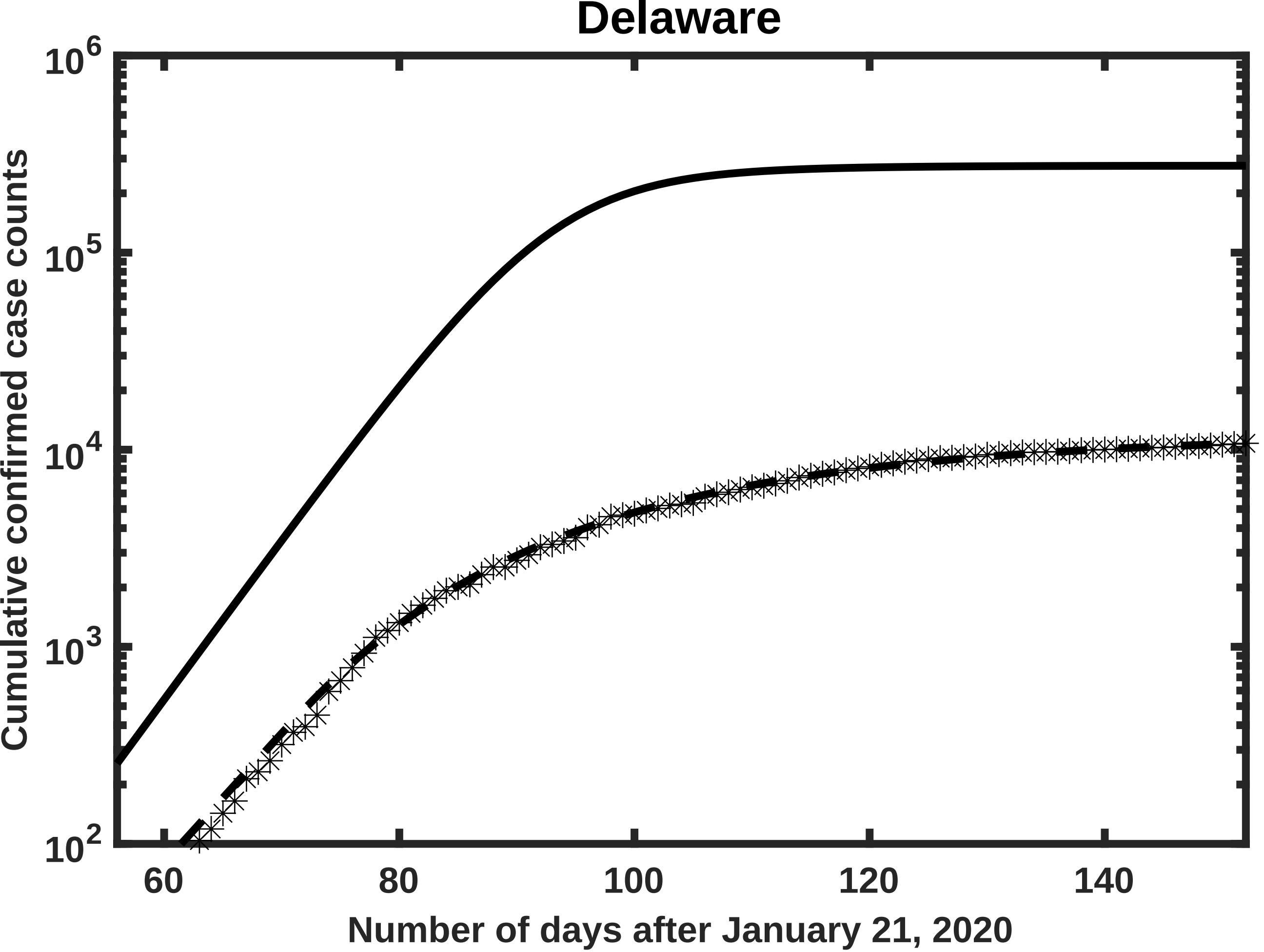
Delaware
10^6
10^5
10^4
10^3
10^2
Cumulative confirmed case counts
60
80
100
120
140
Number of days after January 21, 2020

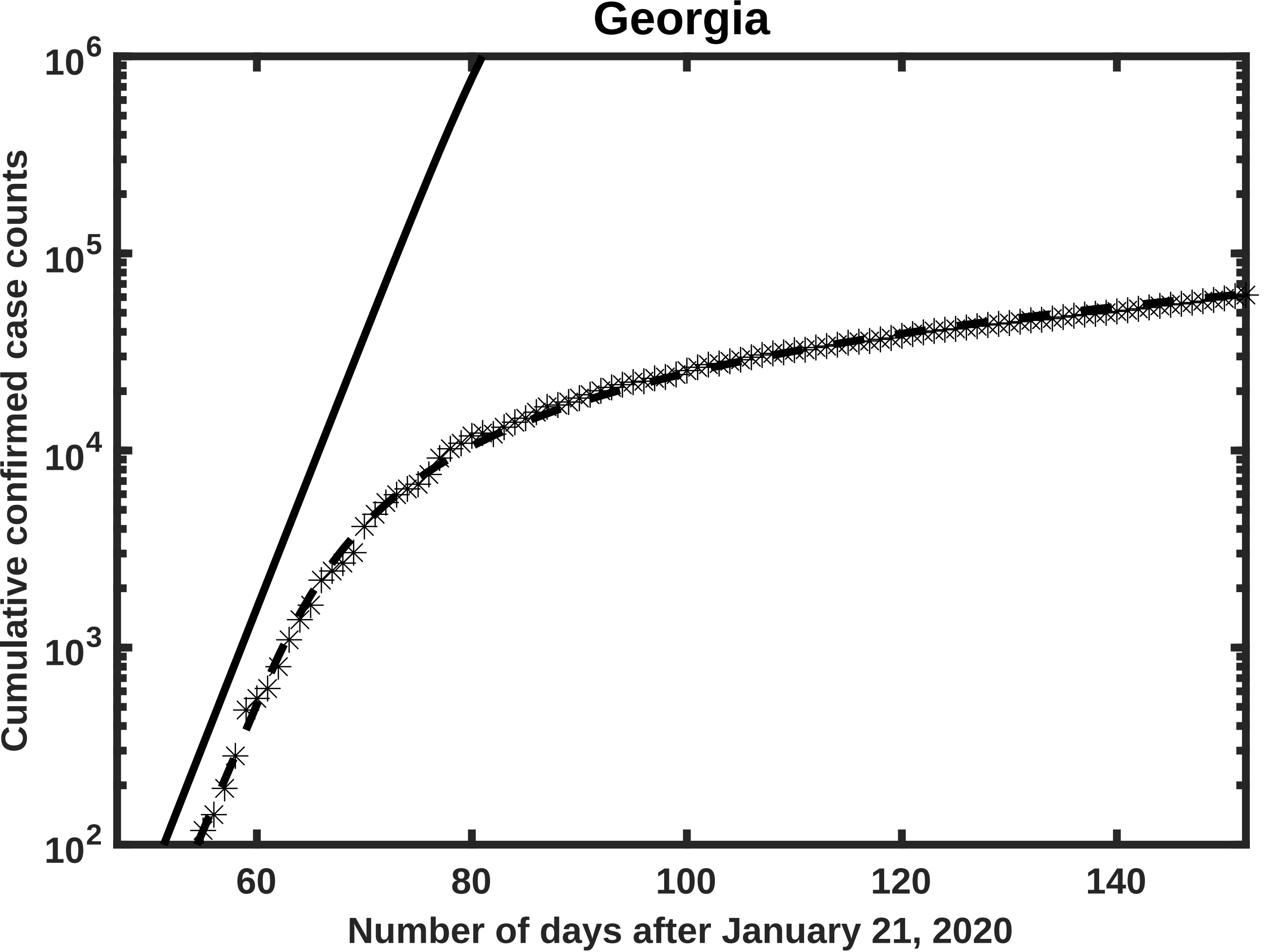

Georgia
$10^6$
$10^5$
$10^4$
$10^3$
$10^2$
Cumulative confirmed case counts
60
80
100
120
140
Number of days after January 21, 2020

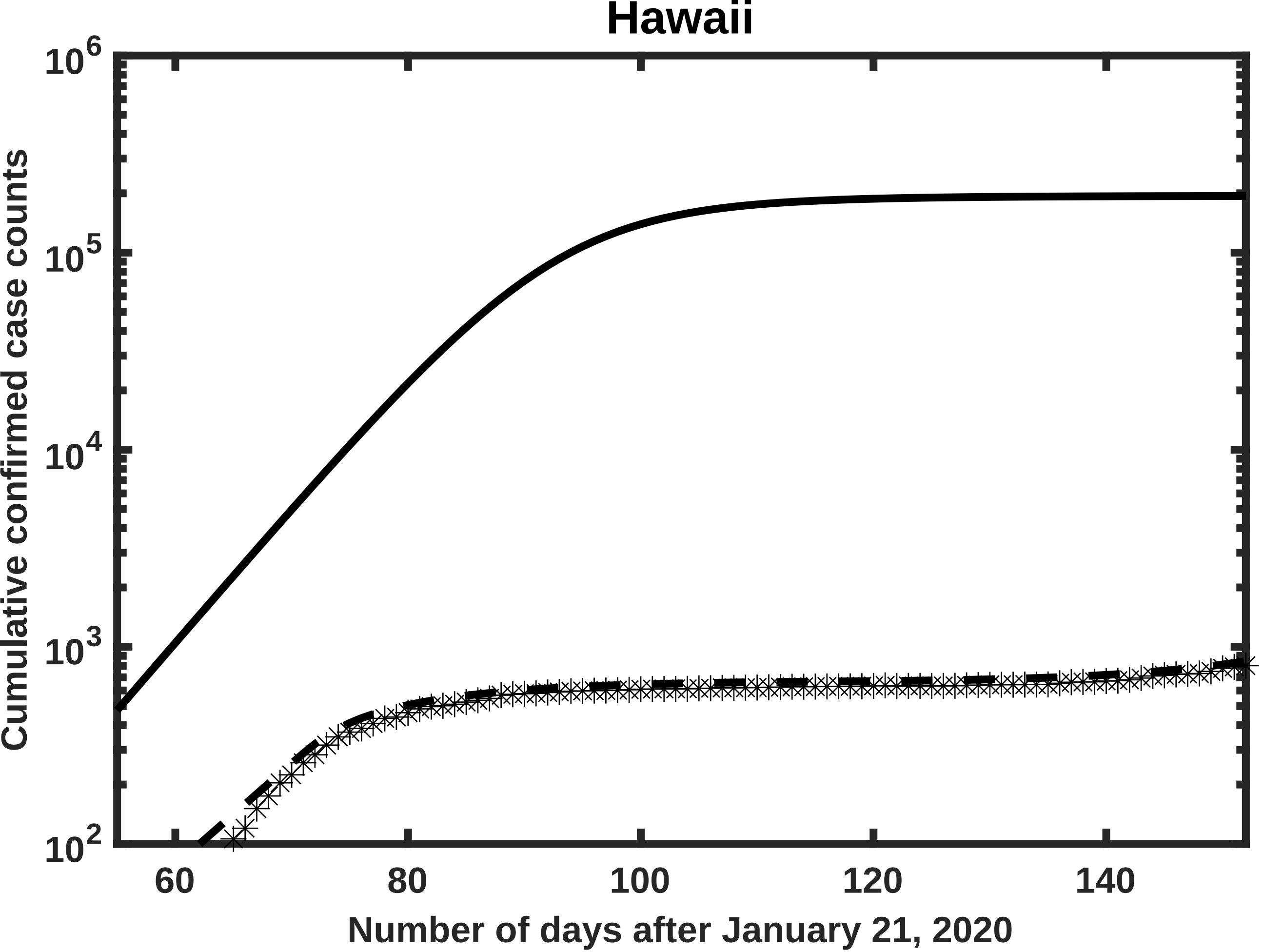
Hawaii
Cumulative confirmed case counts
$10^6$
$10^5$
$10^4$
$10^3$
$10^2$
60
80
100
120
140
Number of days after January 21, 2020

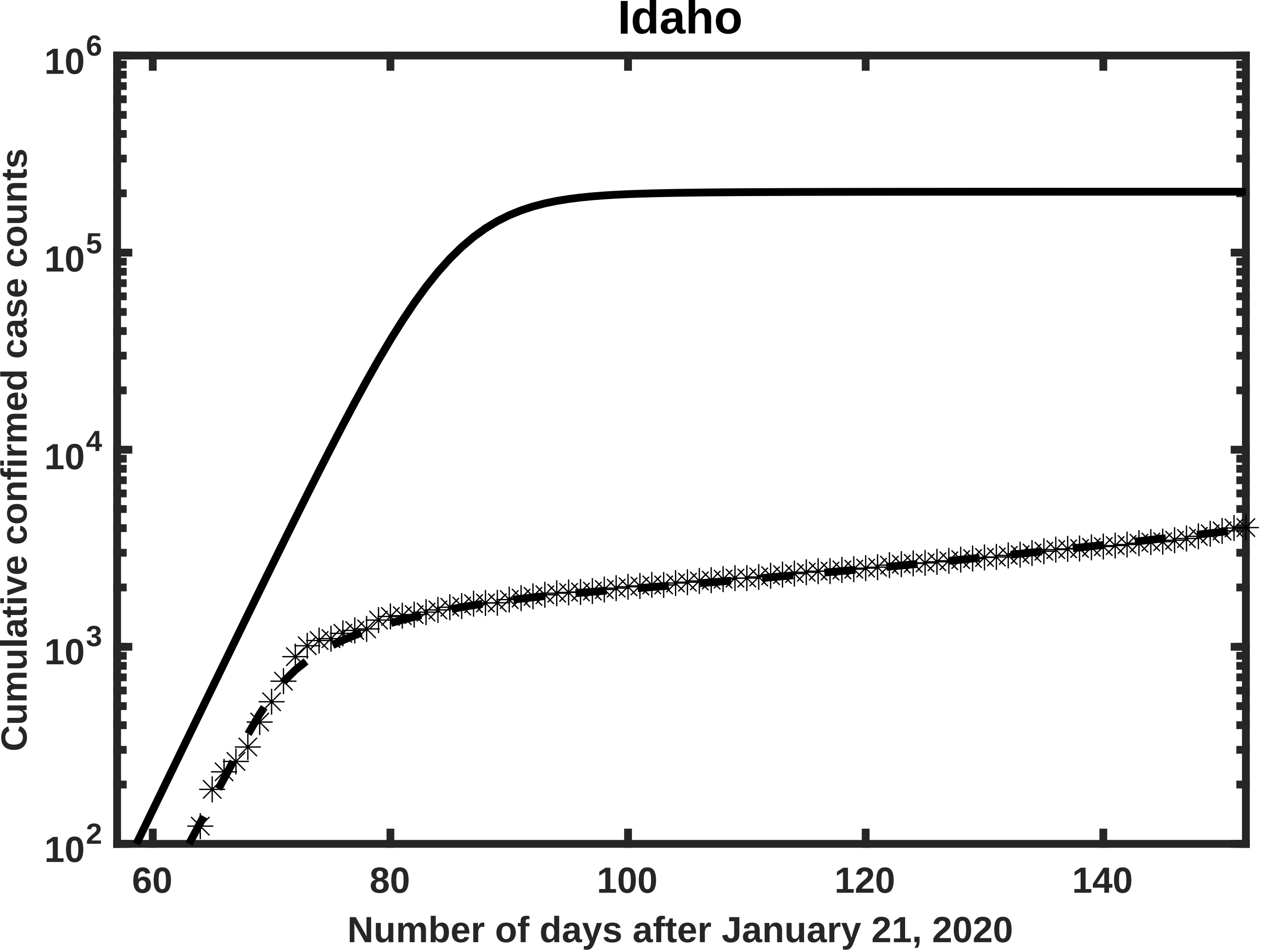

Idaho
Cumulative confirmed case counts
Number of days after January 21, 2020
$10^6$
$10^5$
$10^4$
$10^3$
$10^2$
60
80
100
120
140

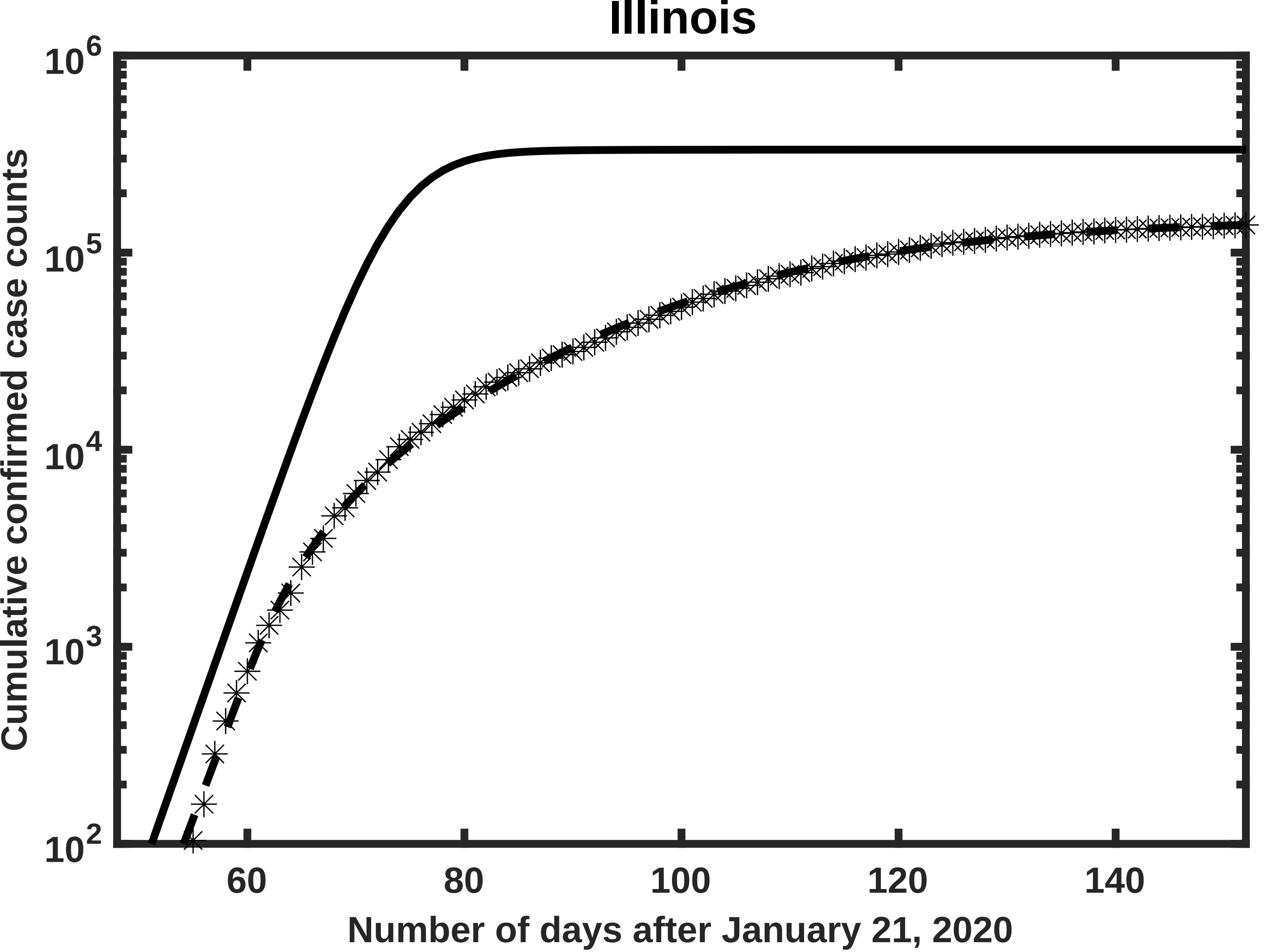

Illinois
Cumulative confirmed case counts
Number of days after January 21, 2020
$10^6$
$10^5$
$10^4$
$10^3$
$10^2$
60
80
100
120
140

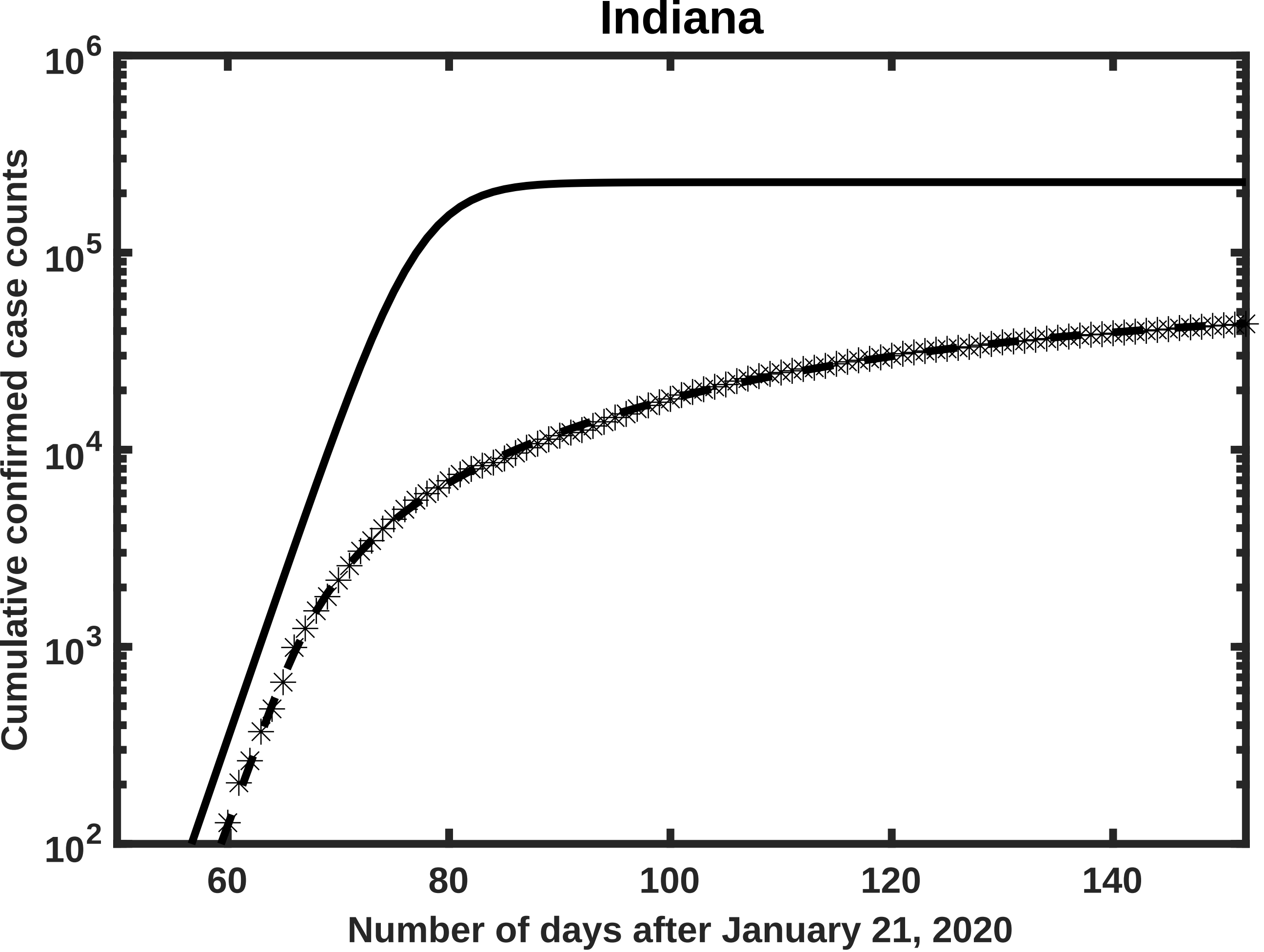
Indiana
Cumulative confirmed case counts
Number of days after January 21, 2020
$10^6$
$10^5$
$10^4$
$10^3$
$10^2$
60
80
100
120
140

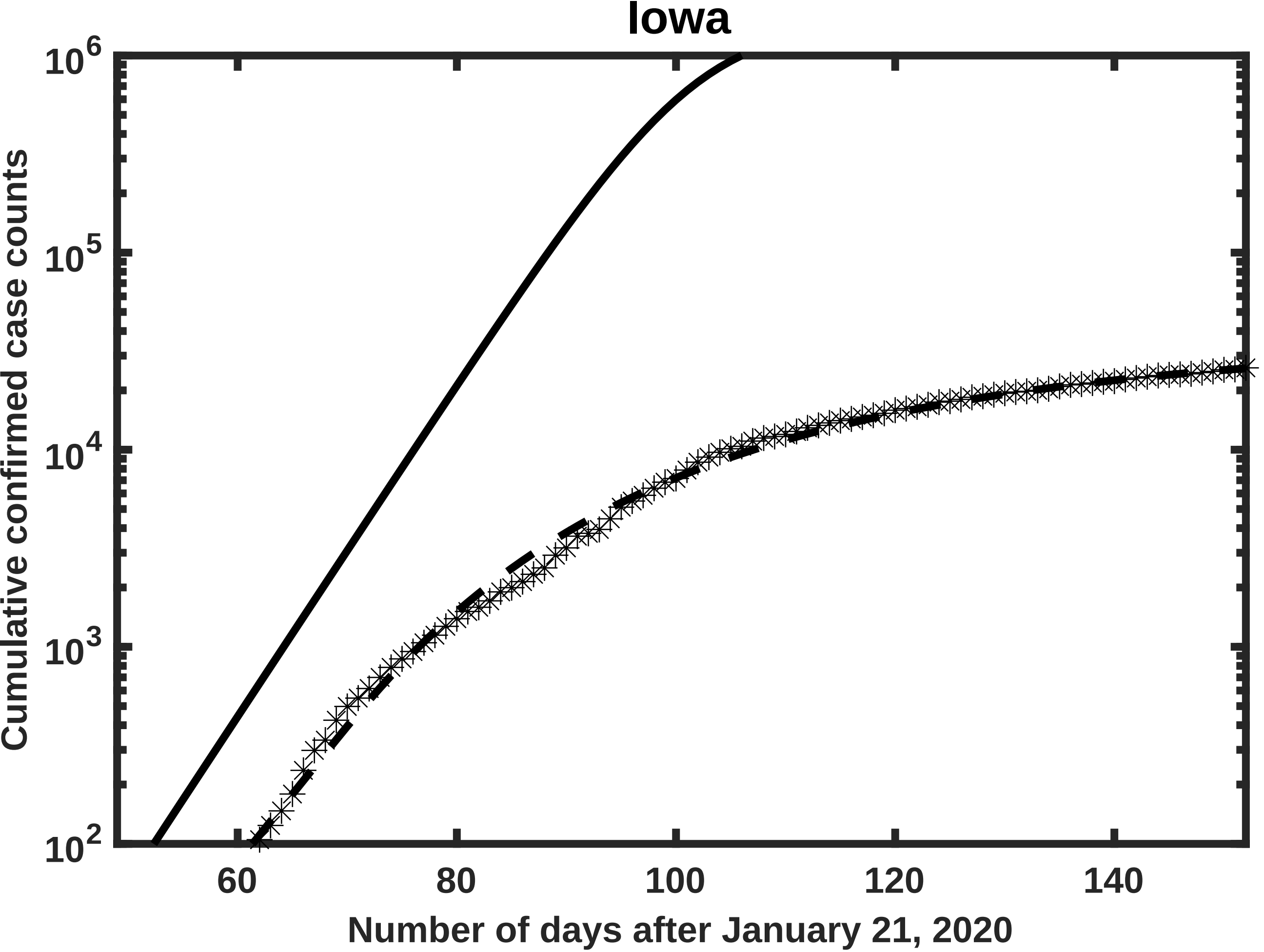

Iowa
Cumulative confirmed case counts
$10^6$
$10^5$
$10^4$
$10^3$
$10^2$
60
80
100
120
140
Number of days after January 21, 2020

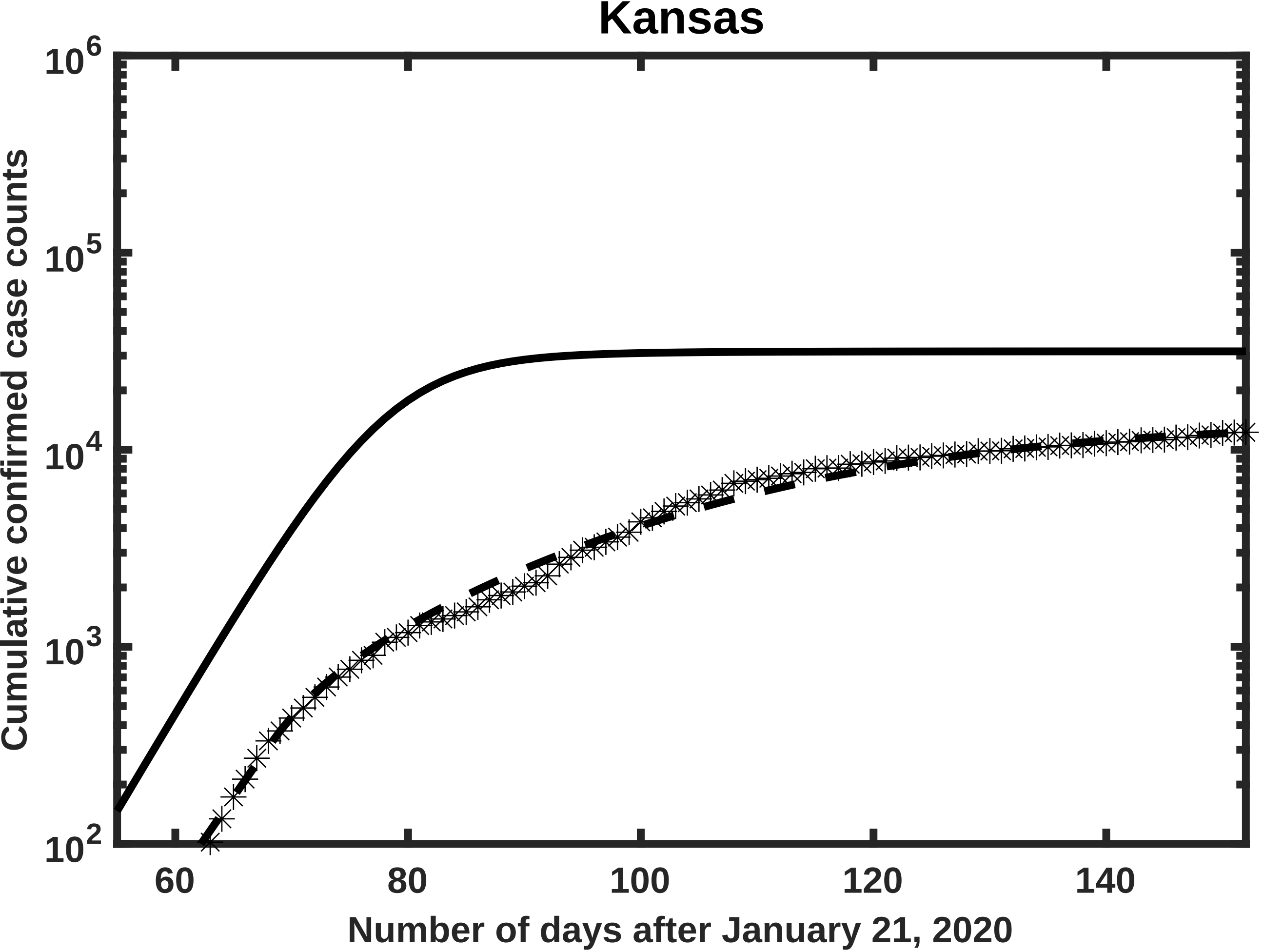

Kansas
10^6
10^5
10^4
10^3
10^2
60
80
100
120
140
Cumulative confirmed case counts
Number of days after January 21, 2020

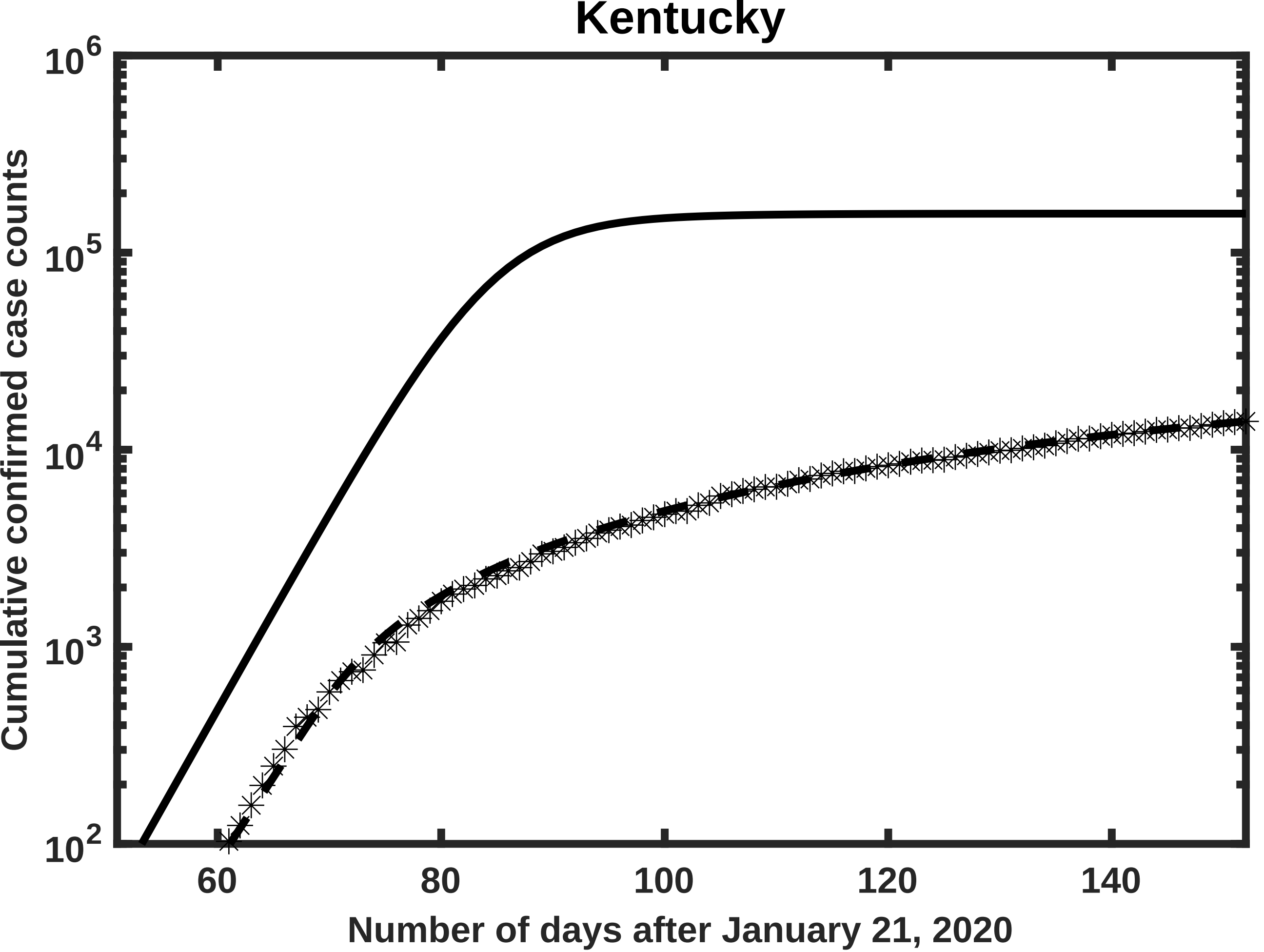
Kentucky
Cumulative confirmed case counts
$10^6$
$10^5$
$10^4$
$10^3$
$10^2$
60
80
100
120
140
Number of days after January 21, 2020

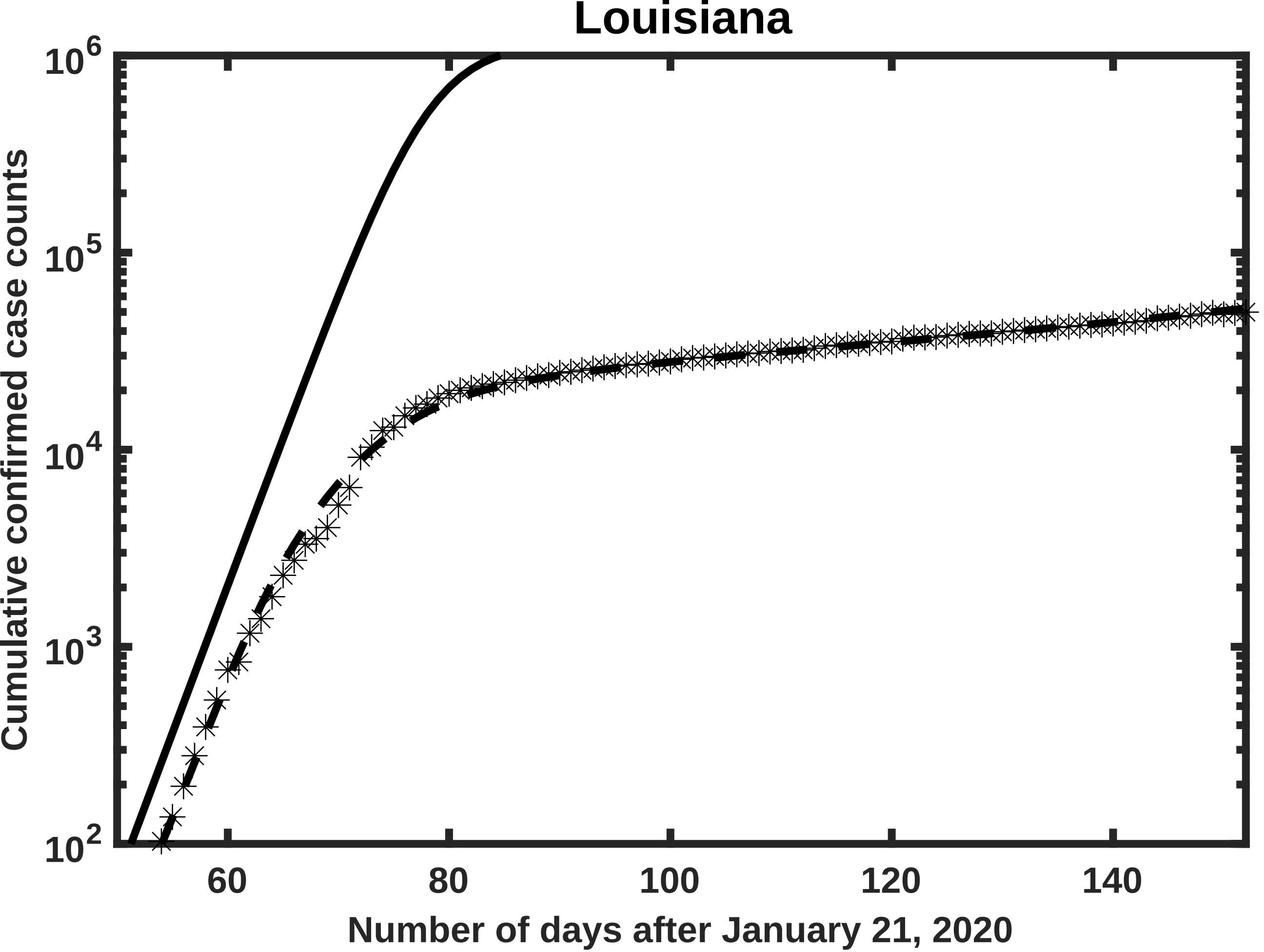

Louisiana
10^6
10^5
10^4
10^3
10^2
Cumulative confirmed case counts
60
80
100
120
140
Number of days after January 21, 2020

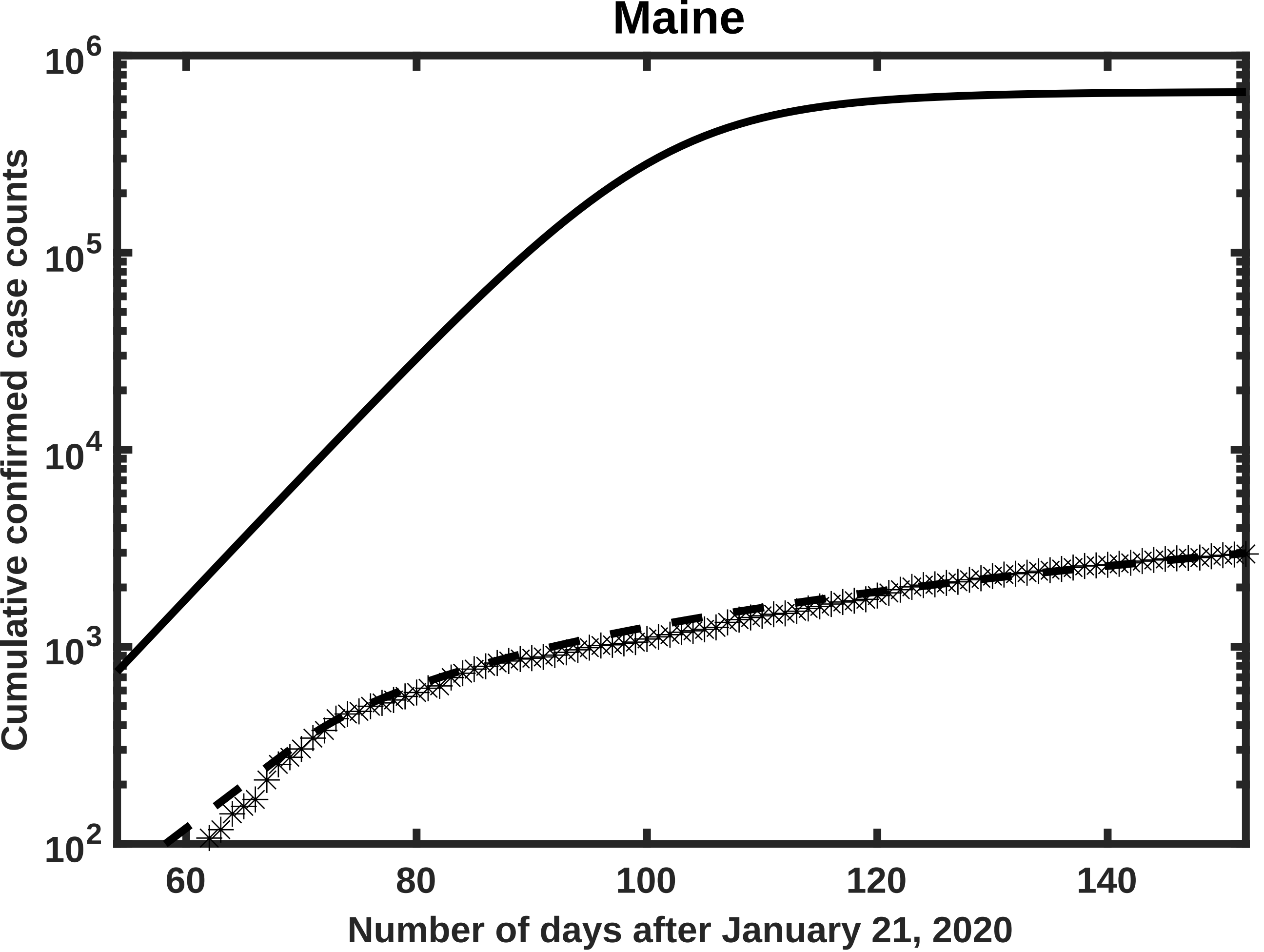
Maine
$10^6$
$10^5$
$10^4$
$10^3$
$10^2$
Cumulative confirmed case counts
60
80
100
120
140
Number of days after January 21, 2020

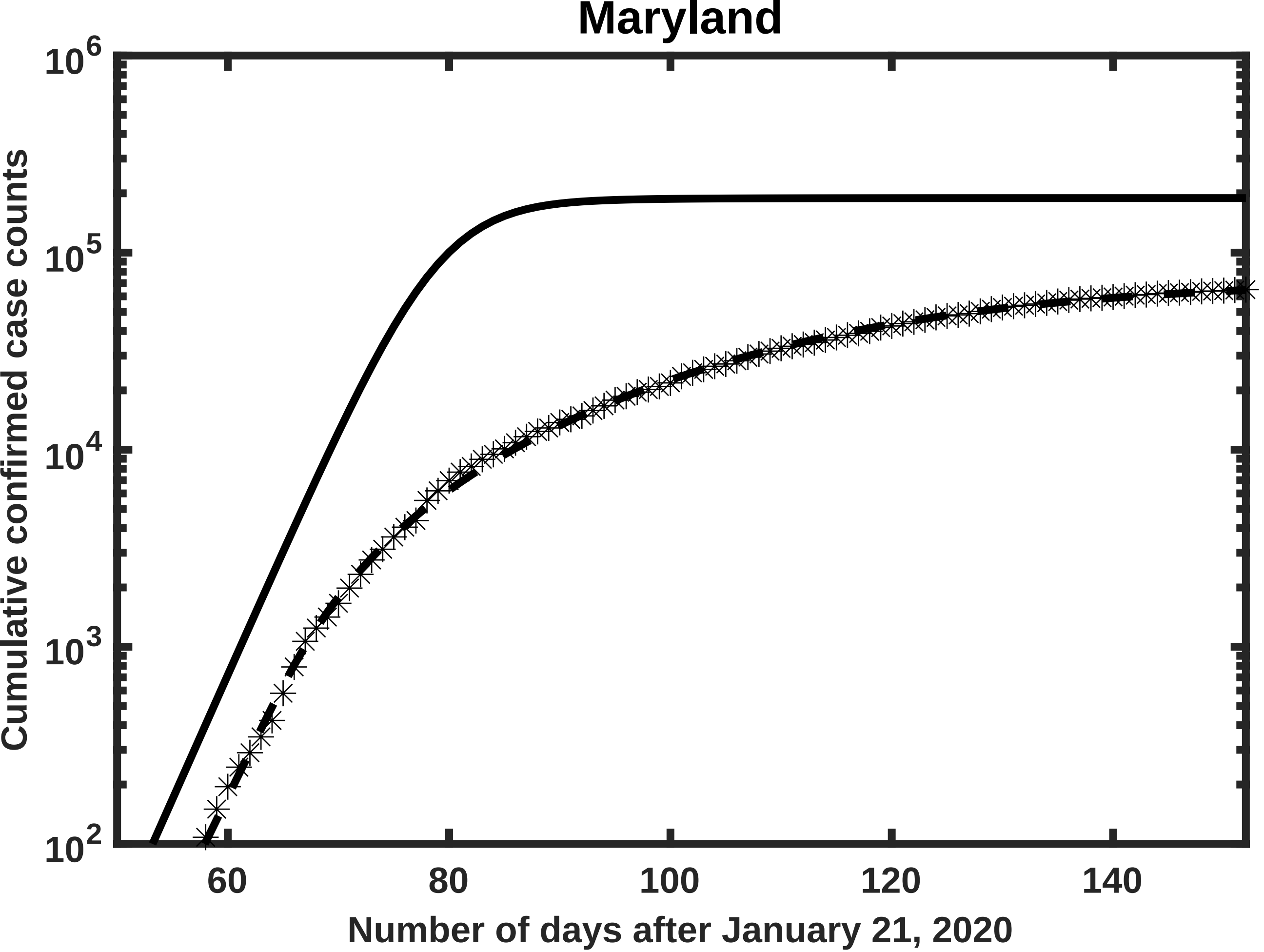

Maryland
$10^6$
$10^5$
$10^4$
$10^3$
$10^2$
Cumulative confirmed case counts
60
80
100
120
140
Number of days after January 21, 2020

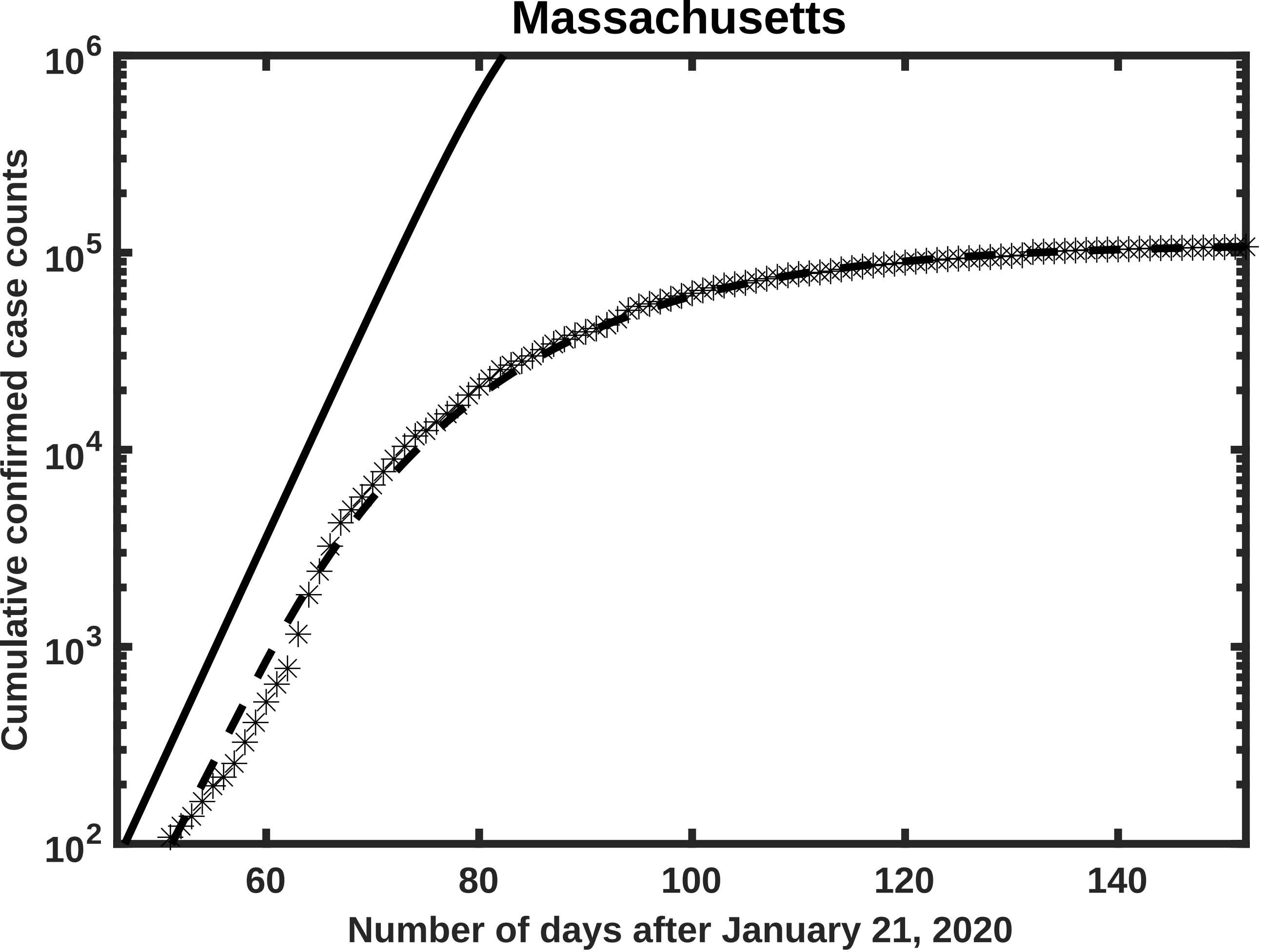
Massachusetts
Cumulative confirmed case counts
$10^2$
$10^3$
$10^4$
$10^5$
$10^6$
60
80
100
120
140
Number of days after January 21, 2020

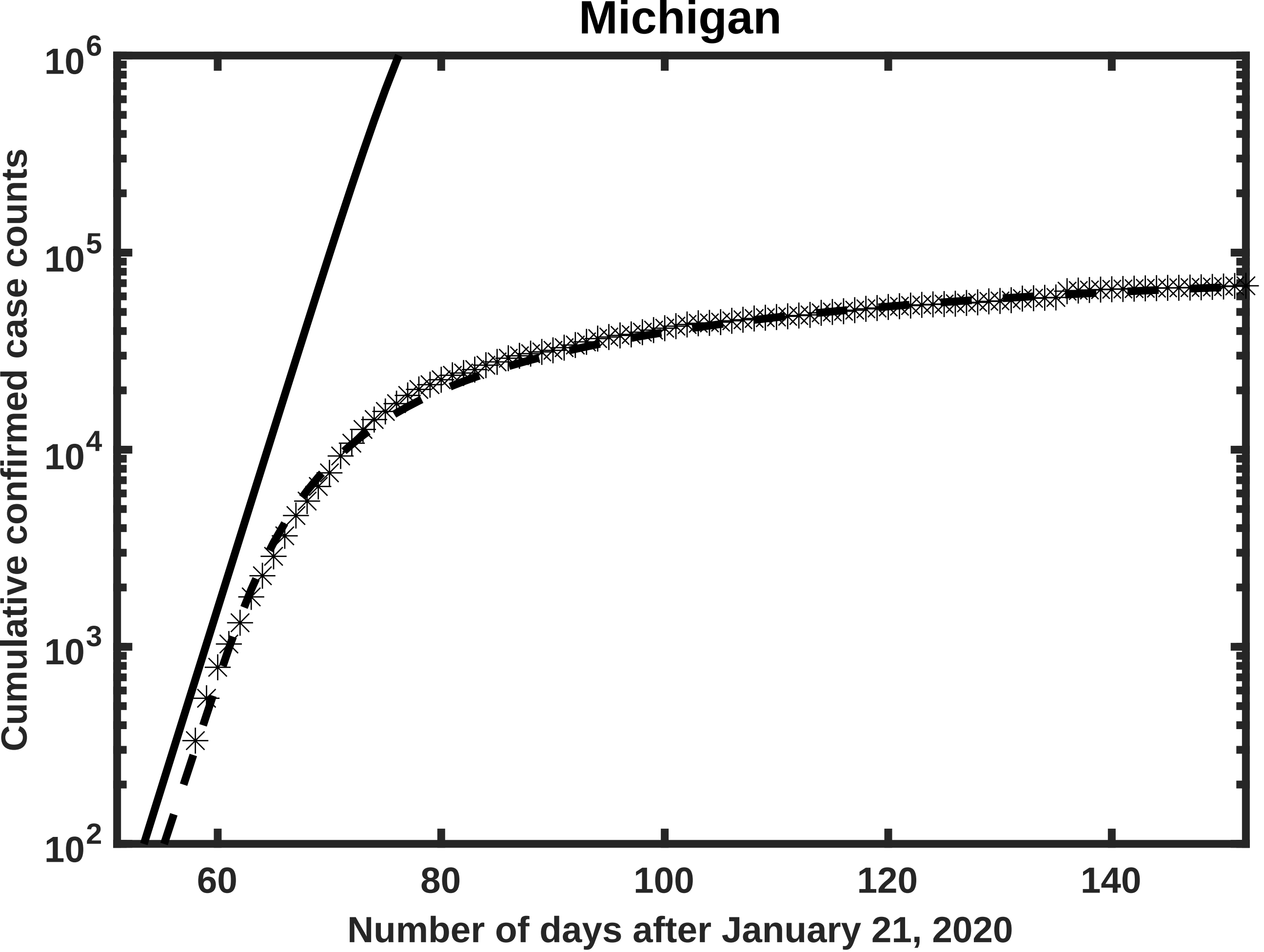

Michigan
Cumulative confirmed case counts
Number of days after January 21, 2020
$10^6$
$10^5$
$10^4$
$10^3$
$10^2$
60
80
100
120
140

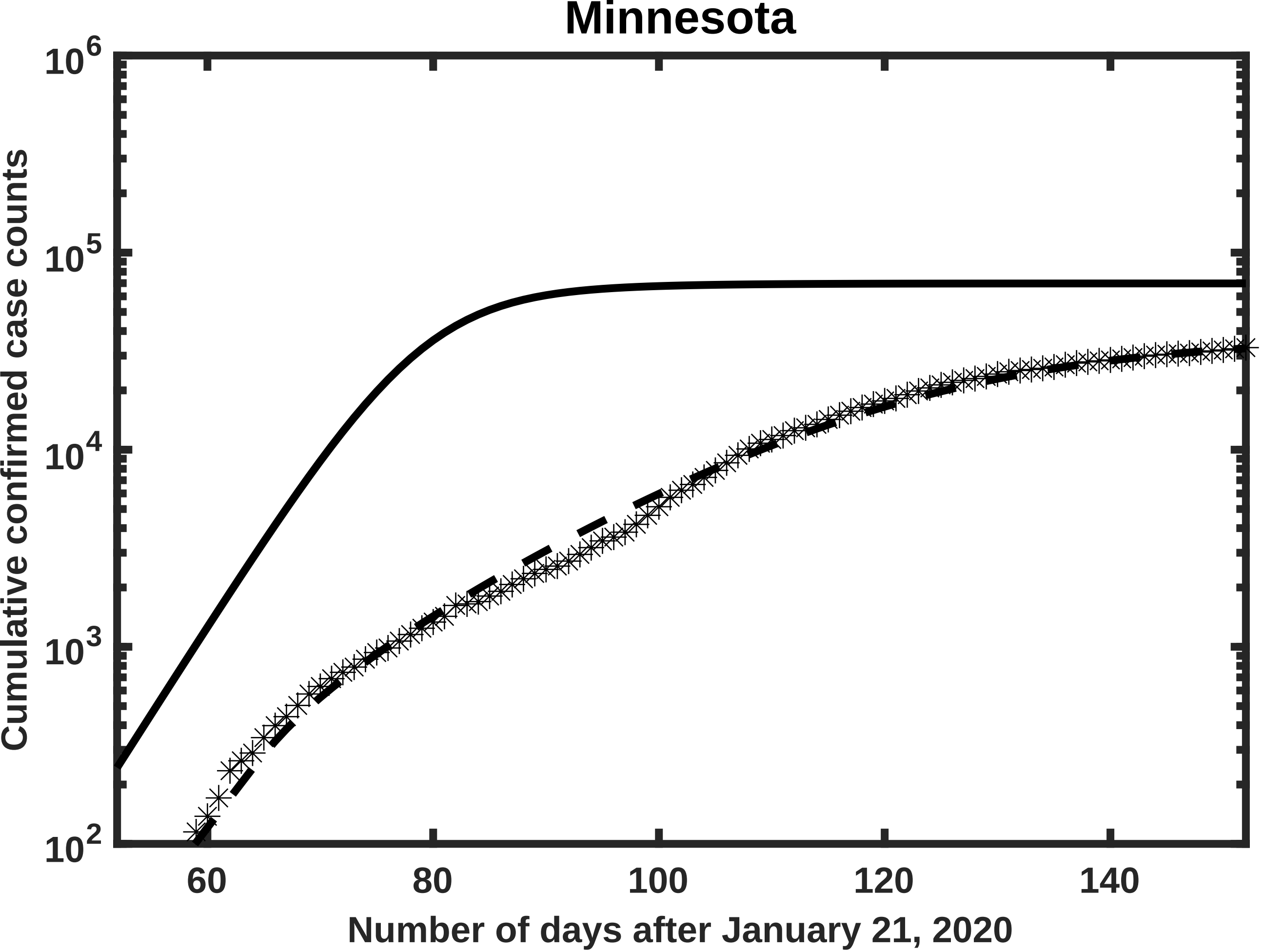
Minnesota
$10^6$
$10^5$
$10^4$
$10^3$
$10^2$
Cumulative confirmed case counts
60
80
100
120
140
Number of days after January 21, 2020

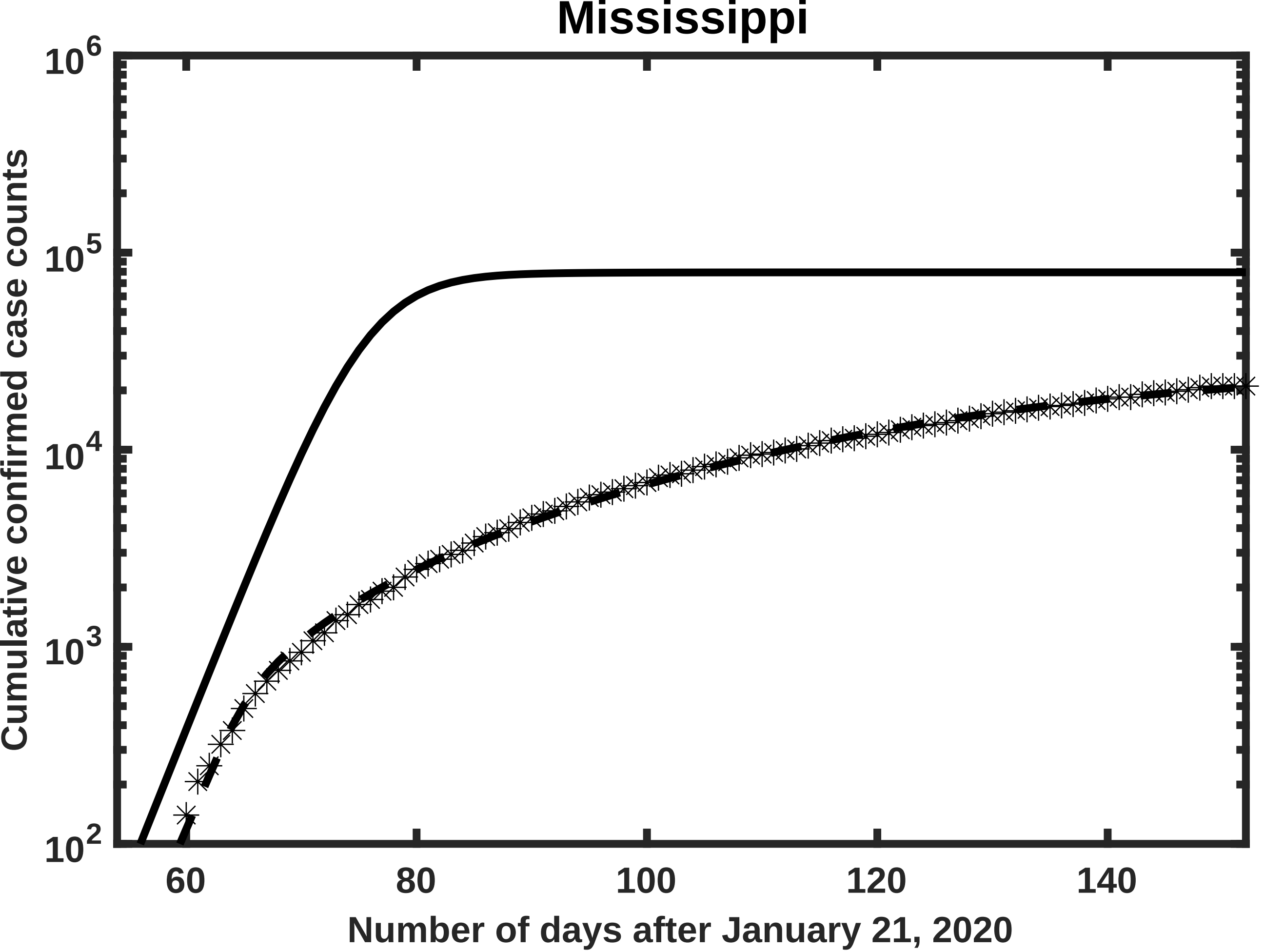
Mississippi
Cumulative confirmed case counts
$10^6$
$10^5$
$10^4$
$10^3$
$10^2$
60
80
100
120
140
Number of days after January 21, 2020

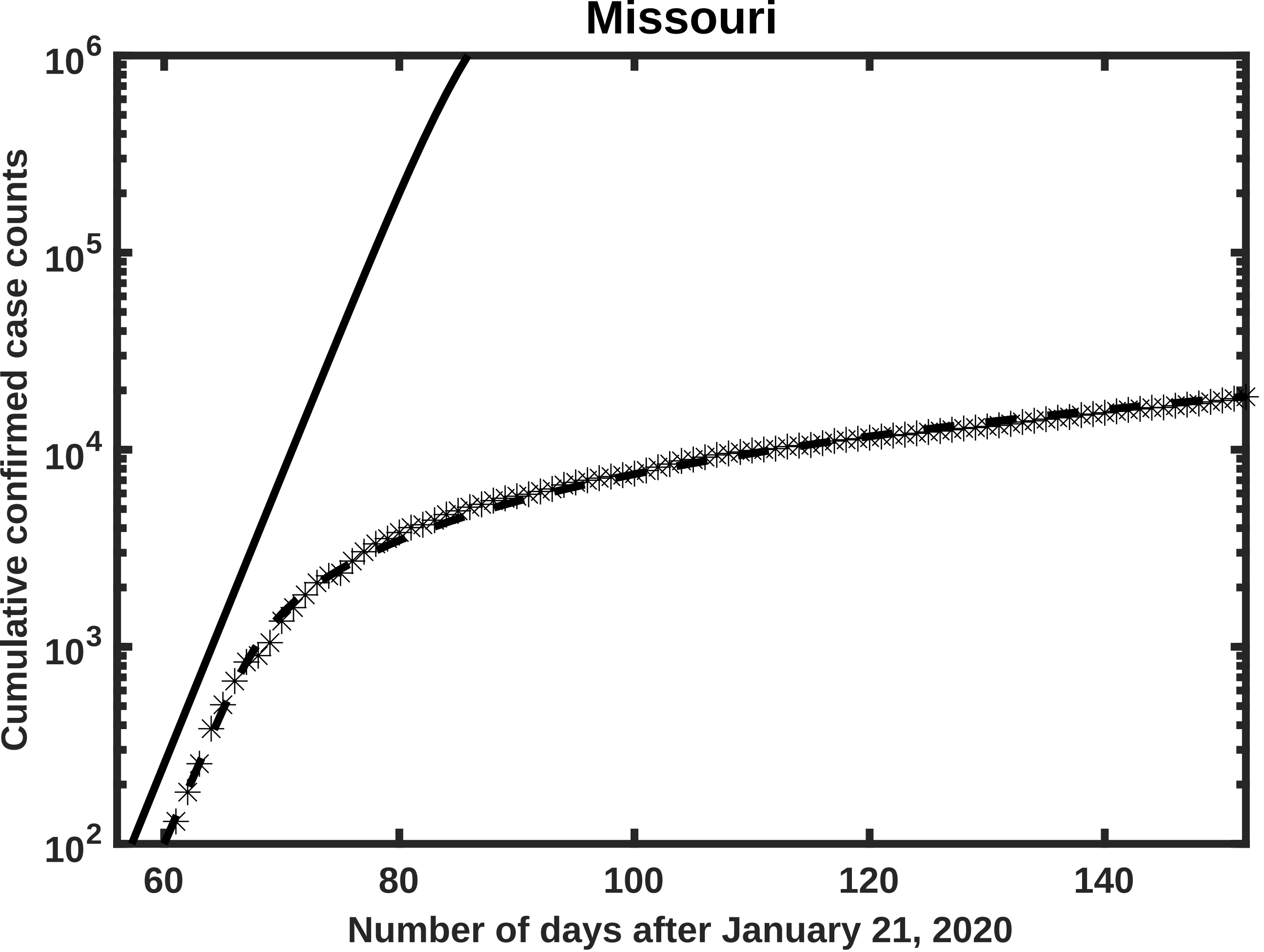
Missouri
Cumulative confirmed case counts
$10^6$
$10^5$
$10^4$
$10^3$
$10^2$
60
80
100
120
140
Number of days after January 21, 2020

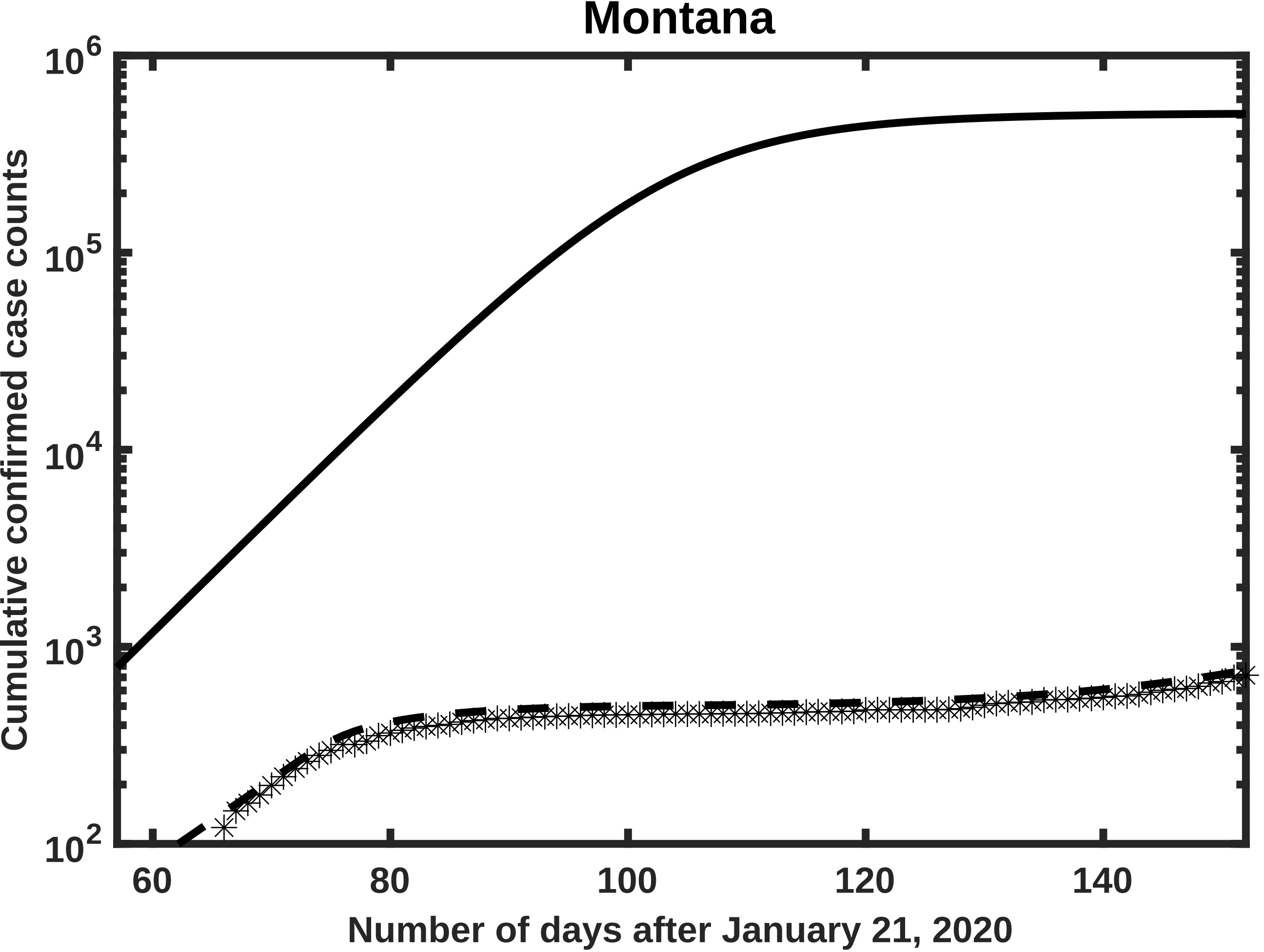
Montana
Cumulative confirmed case counts
$10^6$
$10^5$
$10^4$
$10^3$
$10^2$
60
80
100
120
140
Number of days after January 21, 2020

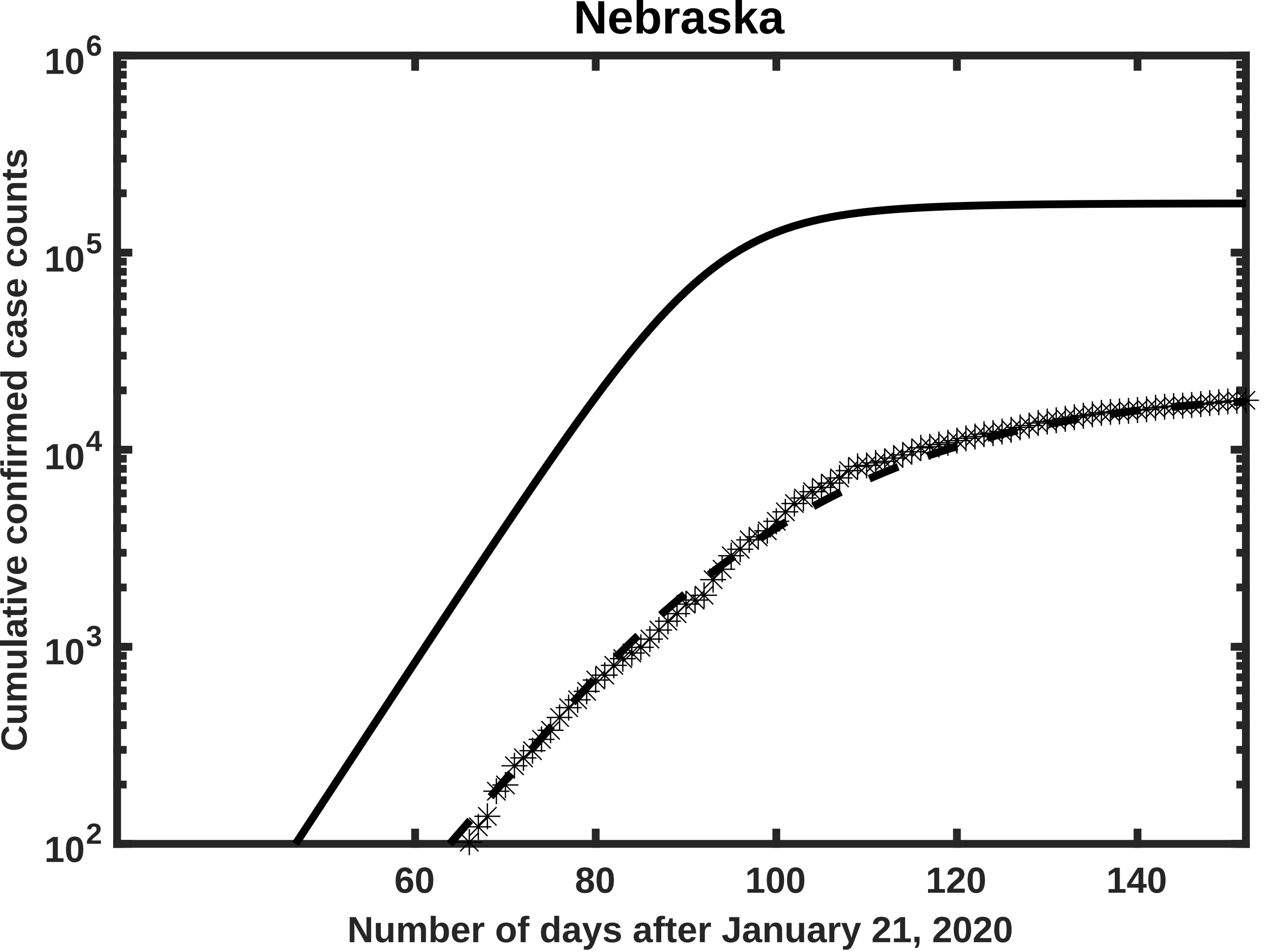

Nebraska
10^6
10^5
10^4
10^3
10^2
60
80
100
120
140
Number of days after January 21, 2020
Cumulative confirmed case counts

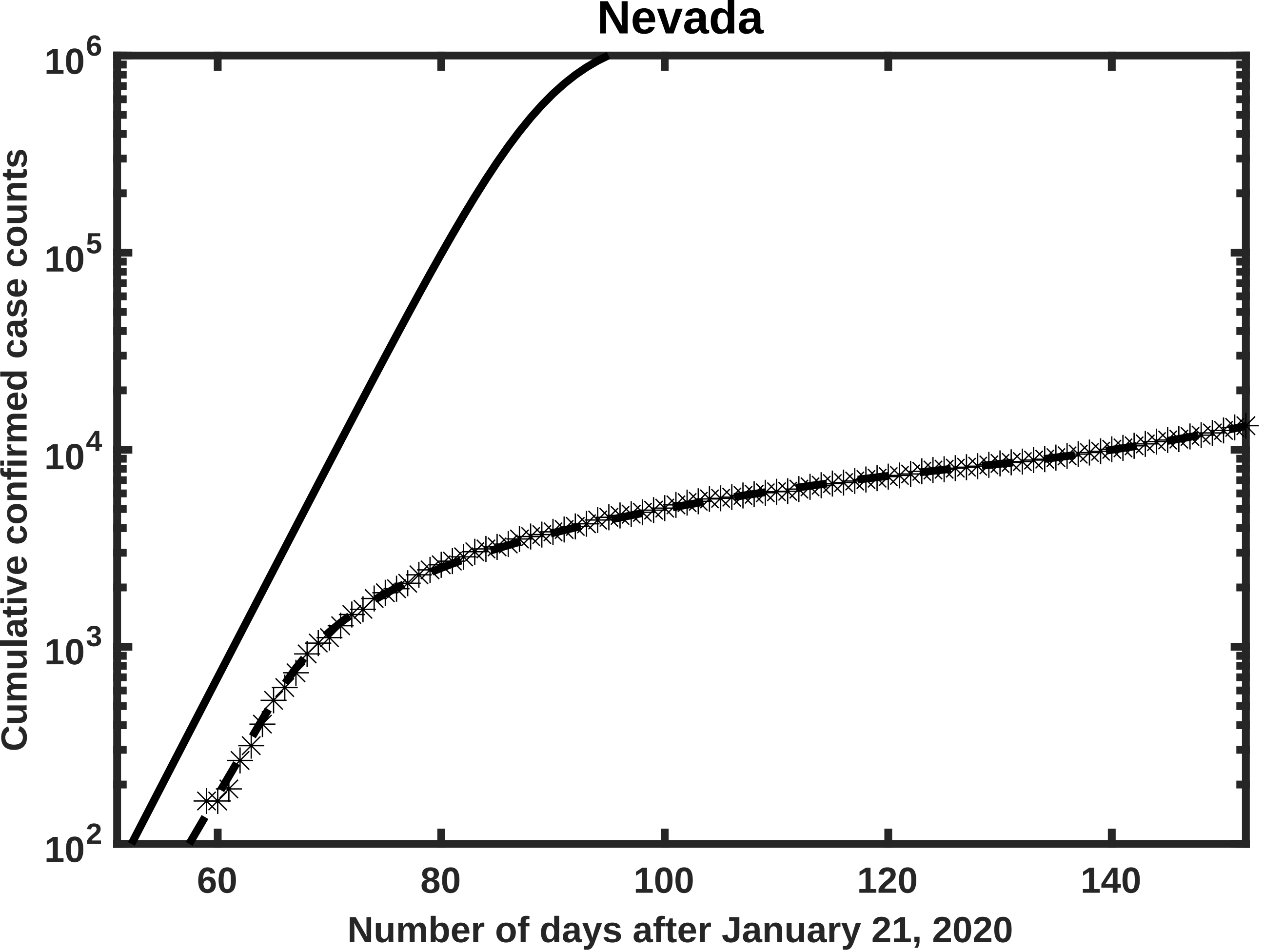

Nevada
10^6
10^5
10^4
10^3
10^2
60
80
100
120
140
Number of days after January 21, 2020
Cumulative confirmed case counts

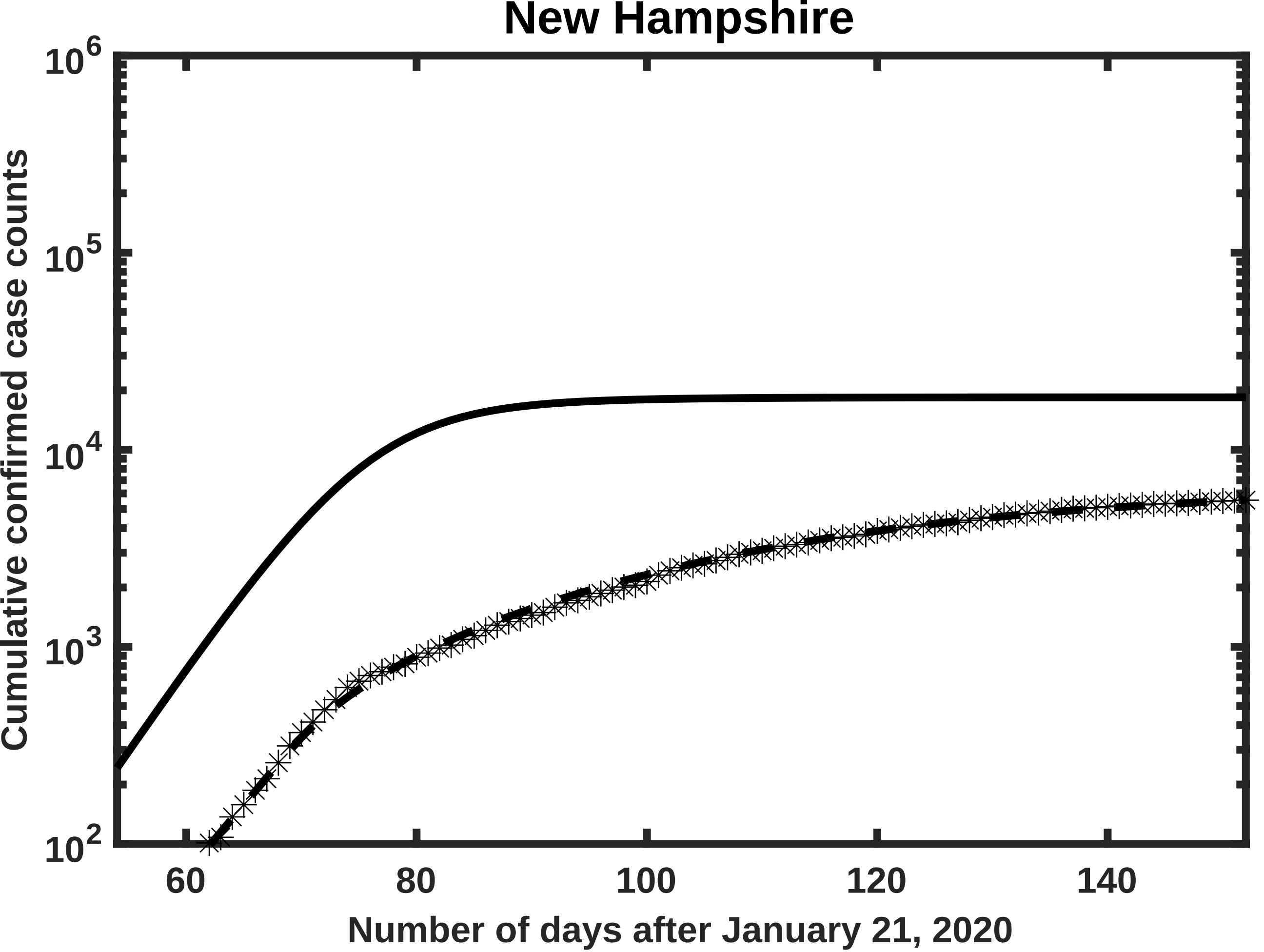

New Hampshire
Cumulative confirmed case counts
$10^6$
$10^5$
$10^4$
$10^3$
$10^2$
60
80
100
120
140
Number of days after January 21, 2020

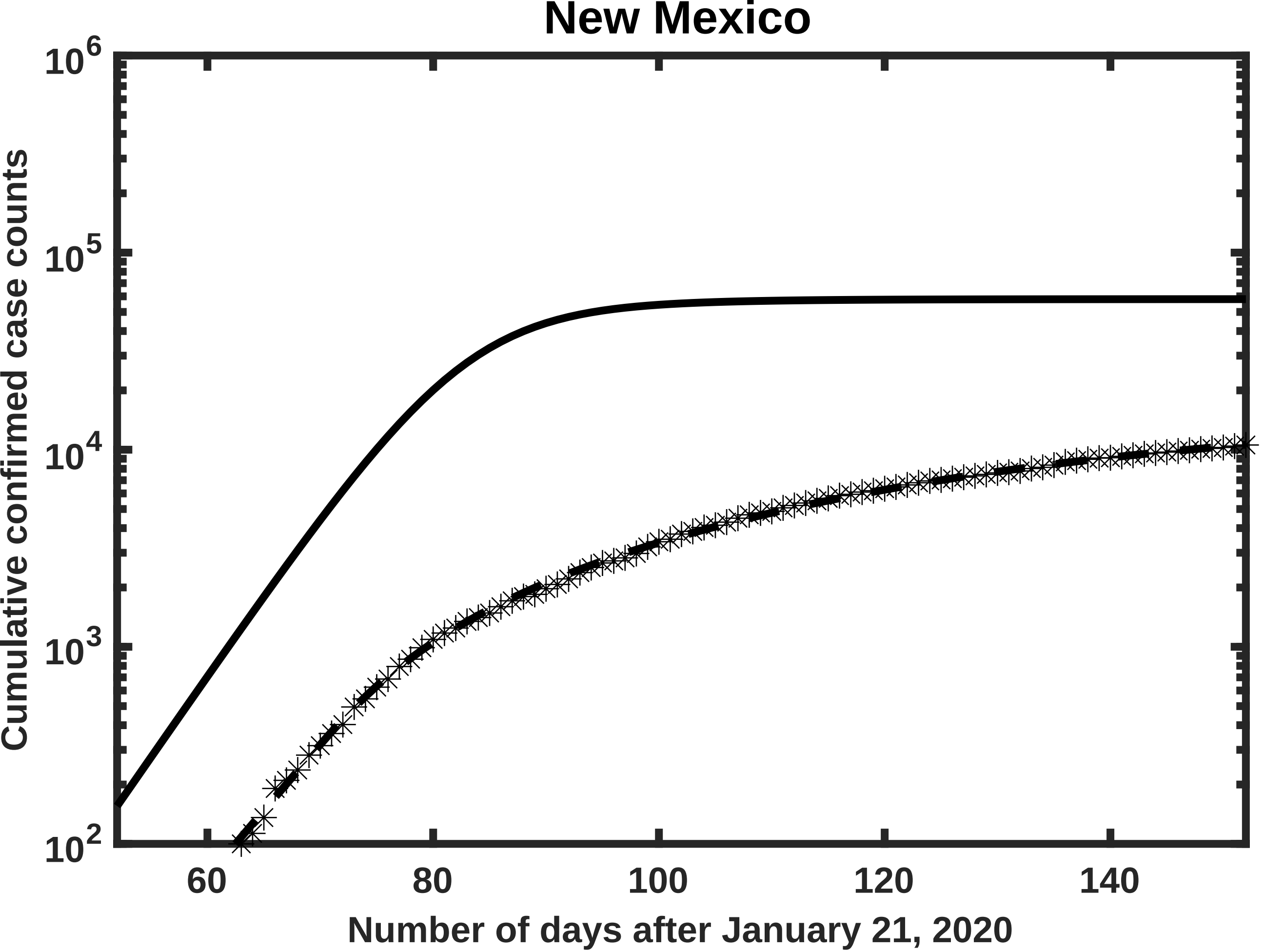
New Mexico
Cumulative confirmed case counts
$10^2$
$10^3$
$10^4$
$10^5$
$10^6$
60
80
100
120
140
Number of days after January 21, 2020

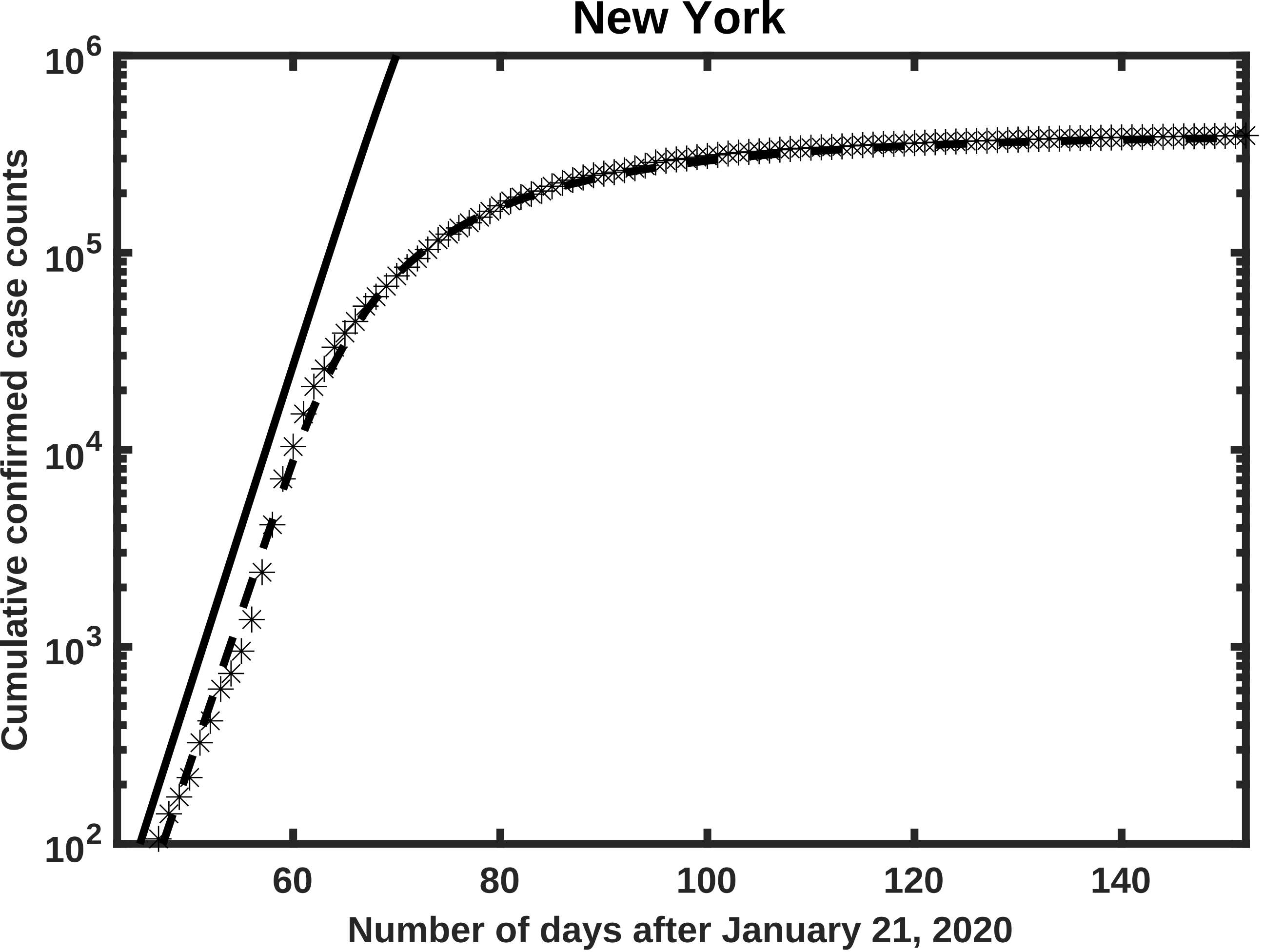
New York
$10^6$
$10^5$
$10^4$
$10^3$
$10^2$
60
80
100
120
140
Cumulative confirmed case counts
Number of days after January 21, 2020

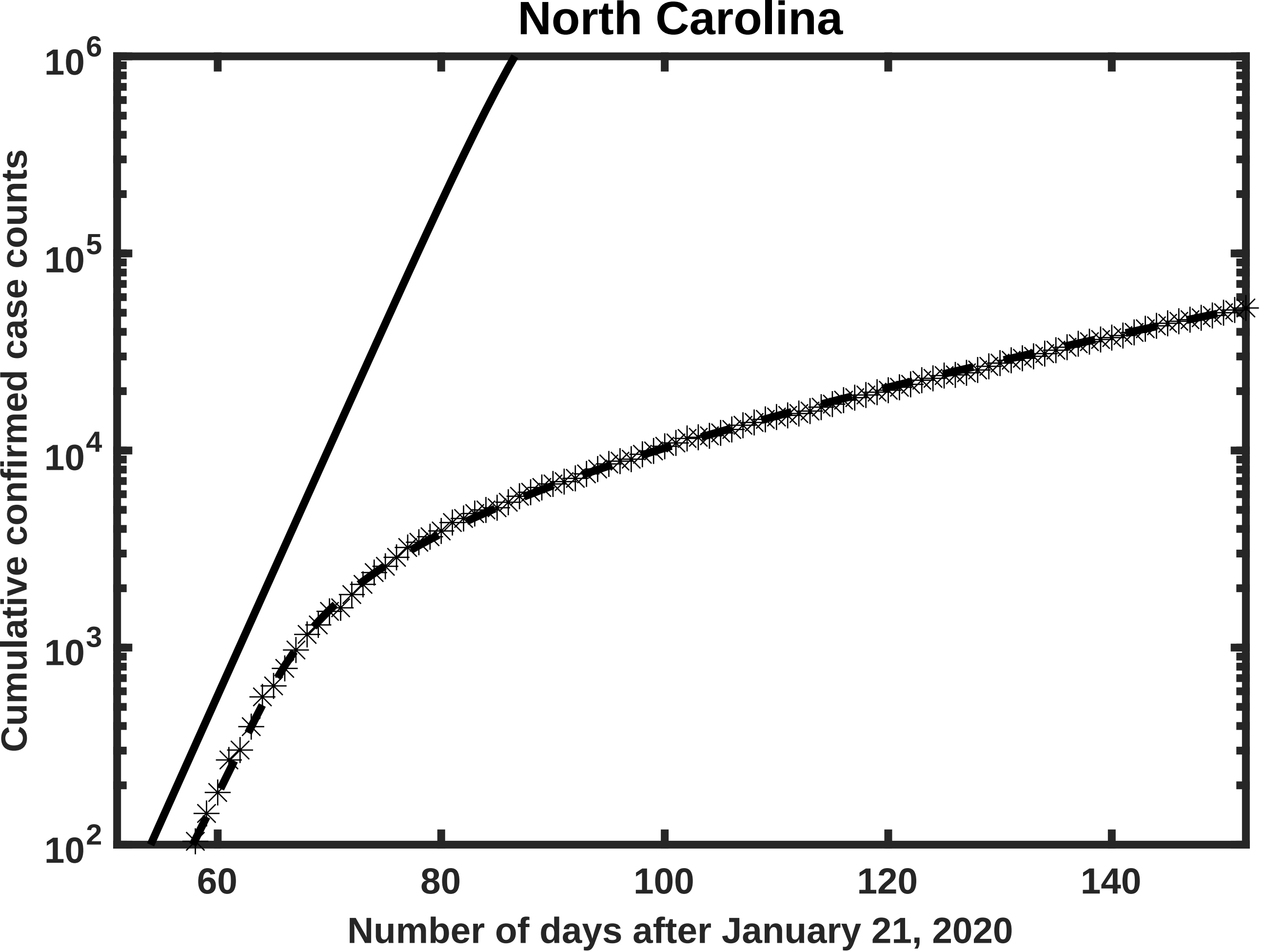
North Carolina
Cumulative confirmed case counts
Number of days after January 21, 2020
$10^6$
$10^5$
$10^4$
$10^3$
$10^2$
60
80
100
120
140

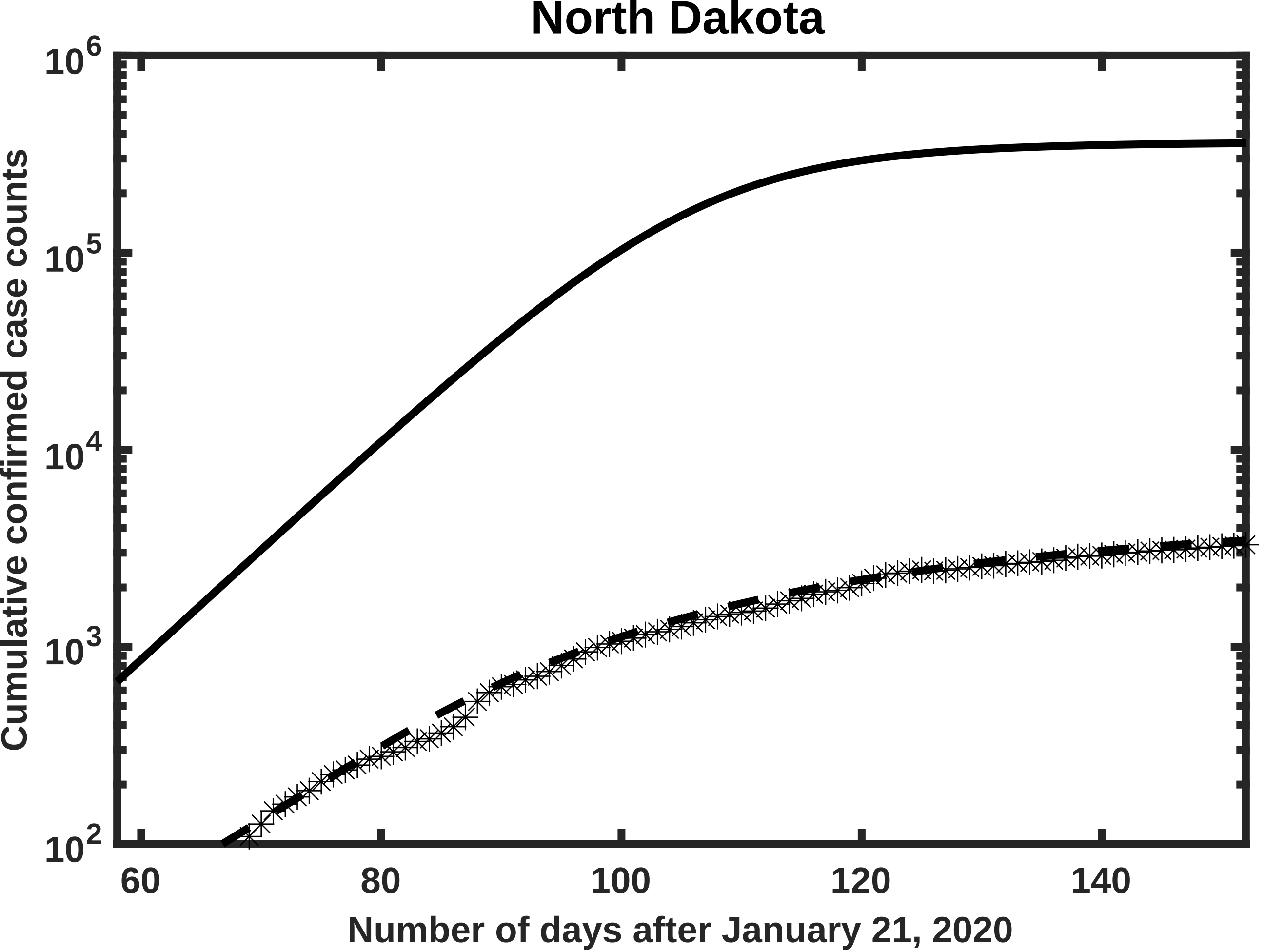
North Dakota
Cumulative confirmed case counts
$10^6$
$10^5$
$10^4$
$10^3$
$10^2$
60
80
100
120
140
Number of days after January 21, 2020

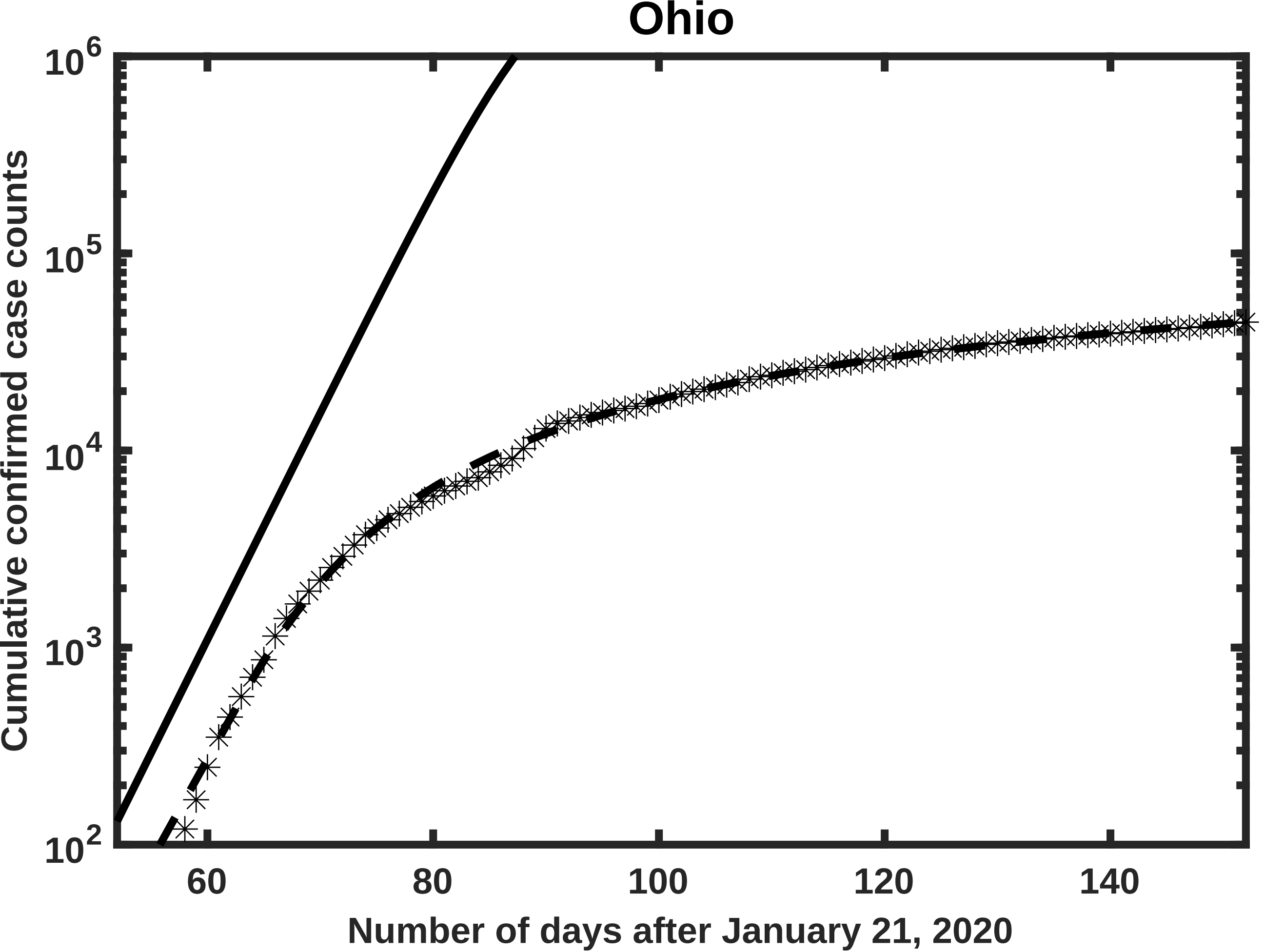

Ohio
Cumulative confirmed case counts
$10^6$
$10^5$
$10^4$
$10^3$
$10^2$
60
80
100
120
140
Number of days after January 21, 2020

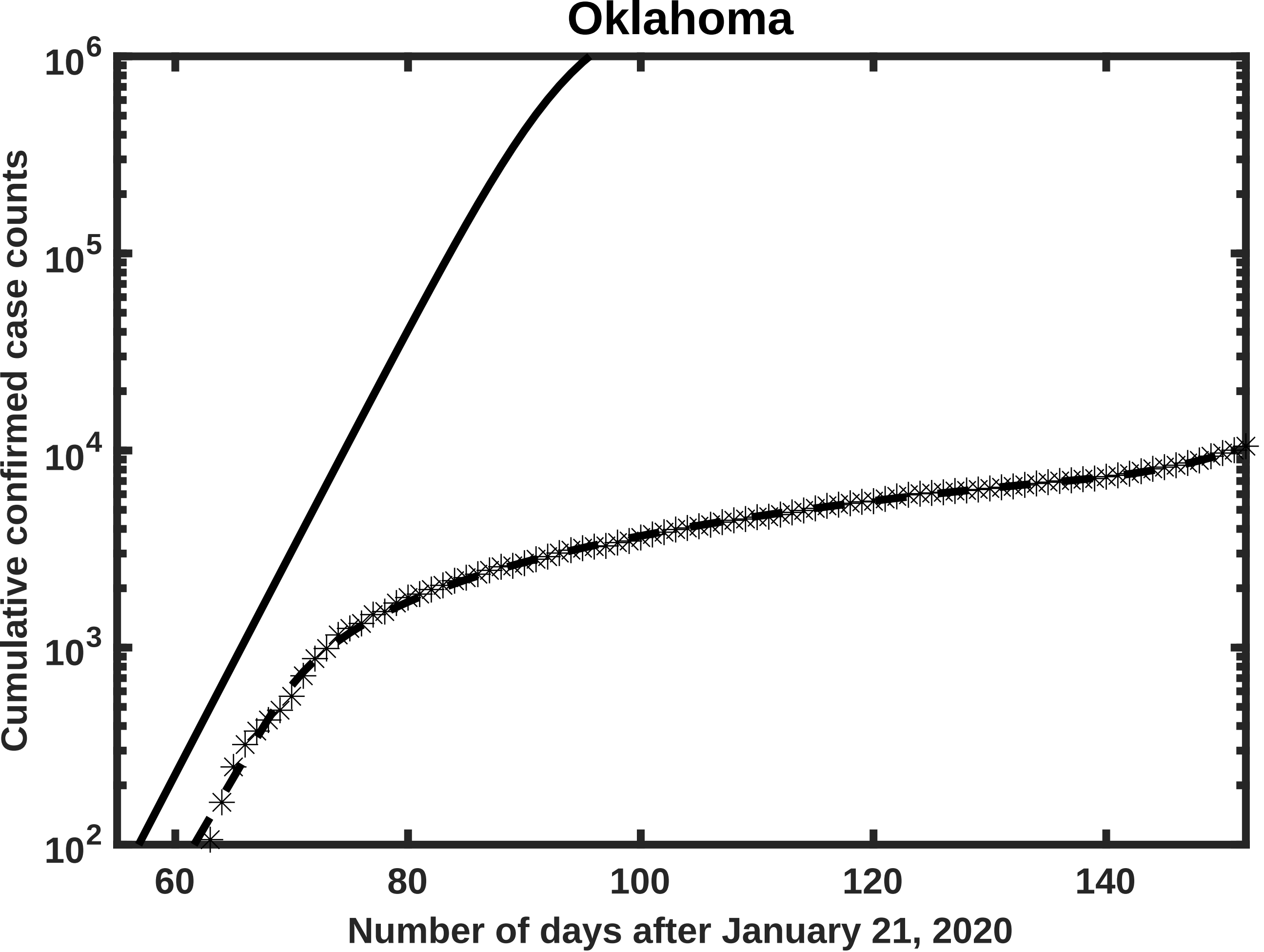
Oklahoma
Cumulative confirmed case counts
Number of days after January 21, 2020
$10^6$
$10^5$
$10^4$
$10^3$
$10^2$
60
80
100
120
140

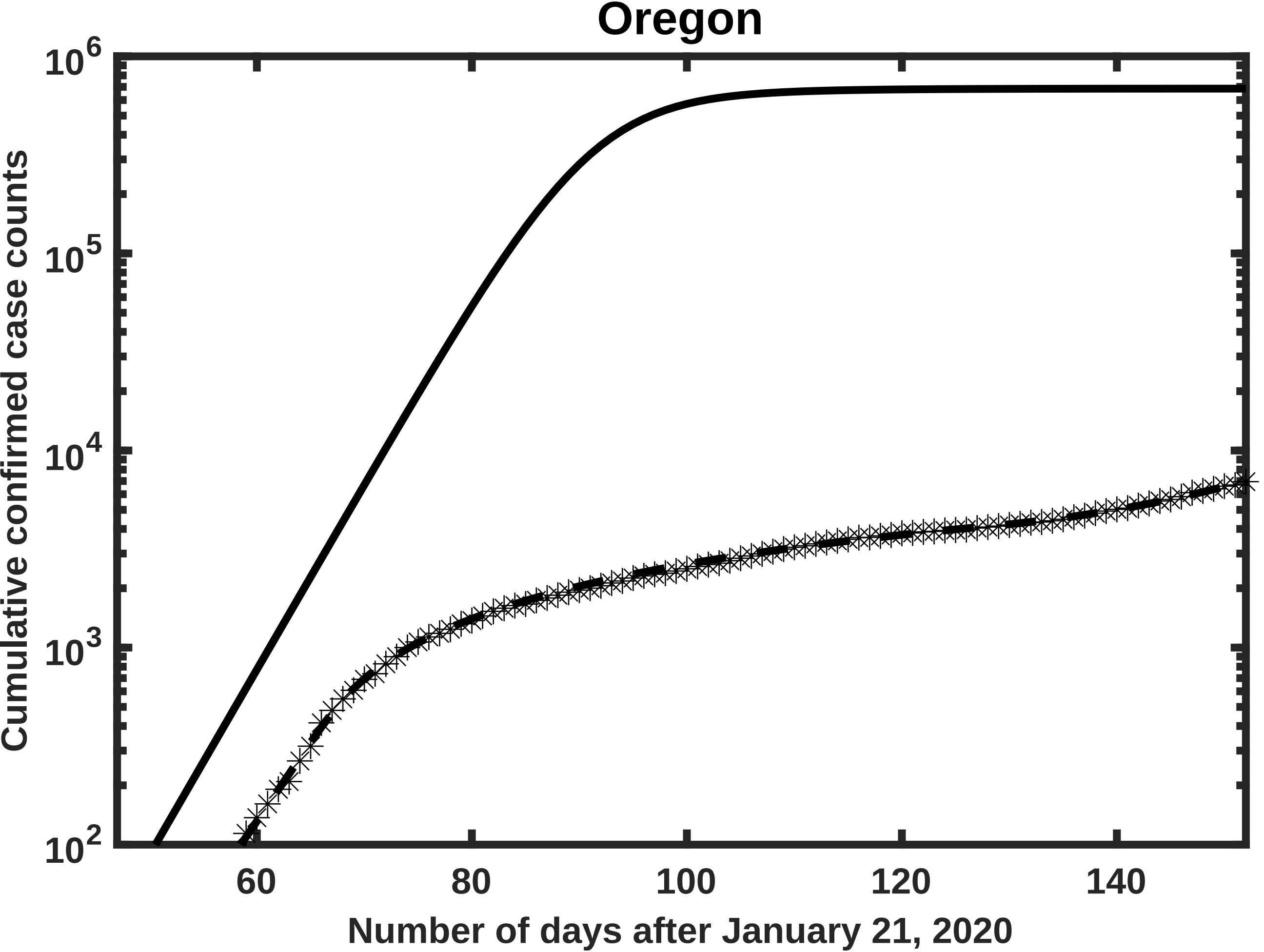
Oregon
Cumulative confirmed case counts
$10^2$
$10^3$
$10^4$
$10^5$
$10^6$
60
80
100
120
140
Number of days after January 21, 2020

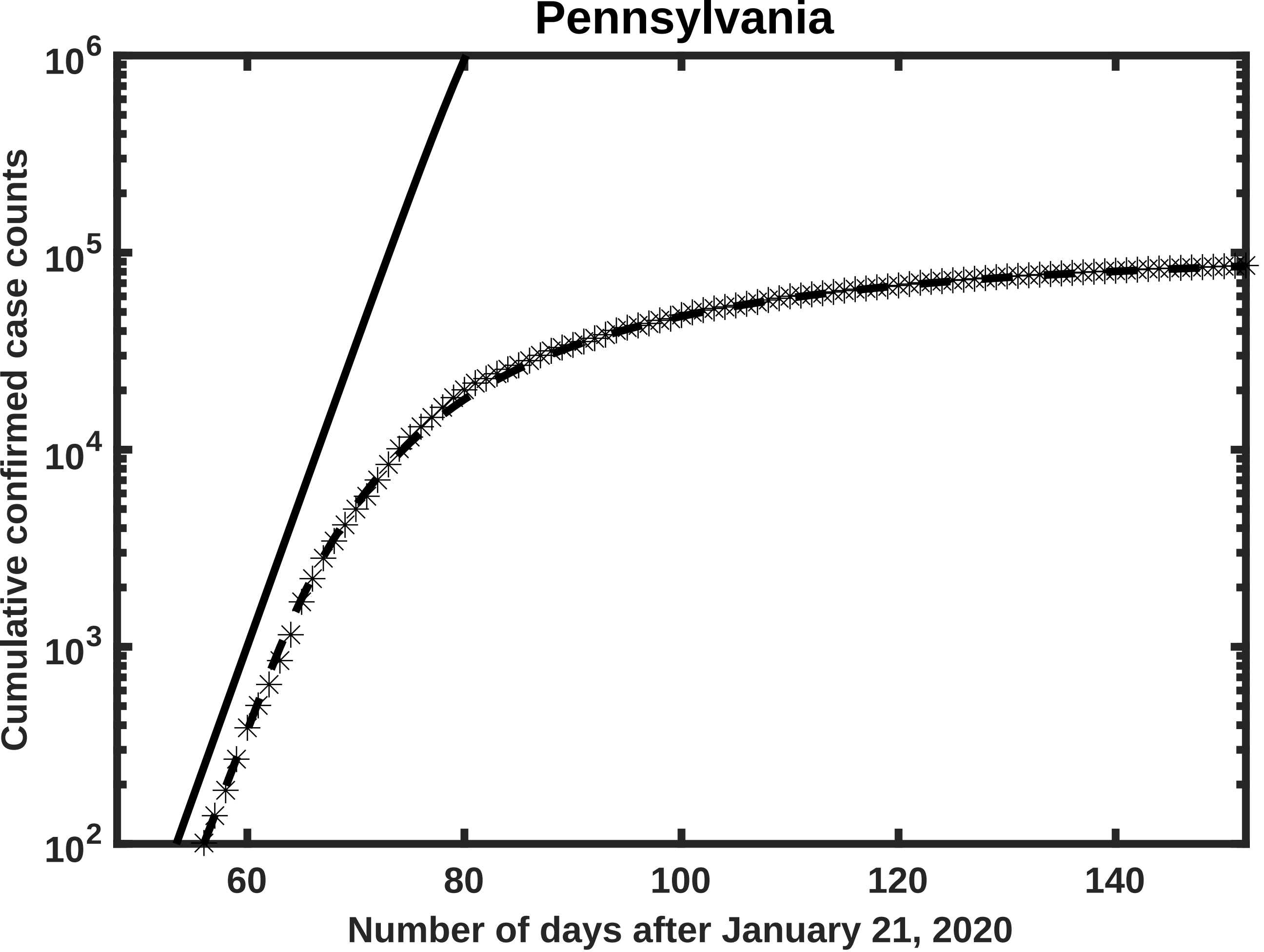

Pennsylvania
$10^6$
$10^5$
$10^4$
$10^3$
$10^2$
Cumulative confirmed case counts
60
80
100
120
140
Number of days after January 21, 2020

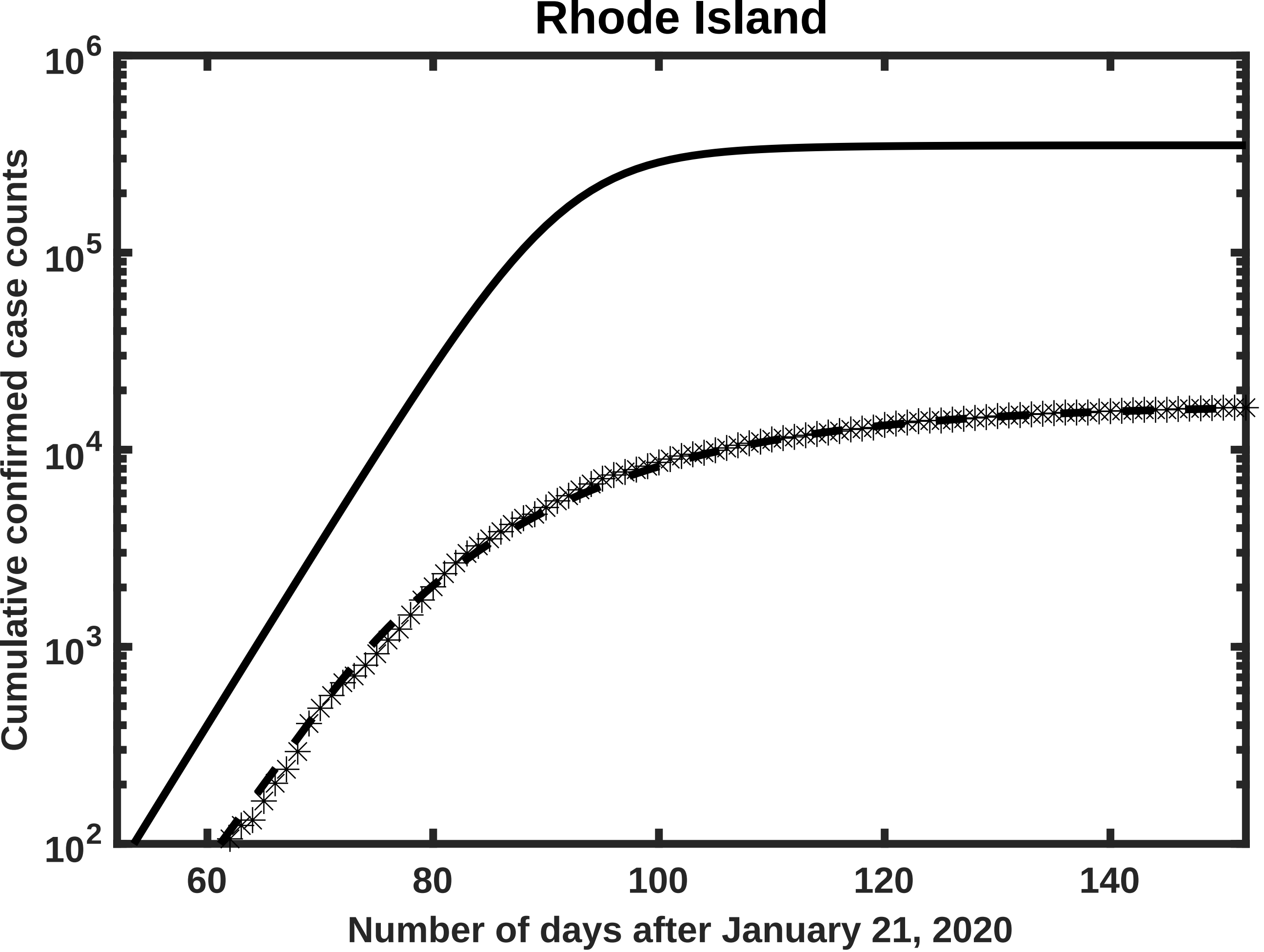
Rhode Island
Cumulative confirmed case counts
$10^2$
$10^3$
$10^4$
$10^5$
$10^6$
60
80
100
120
140
Number of days after January 21, 2020

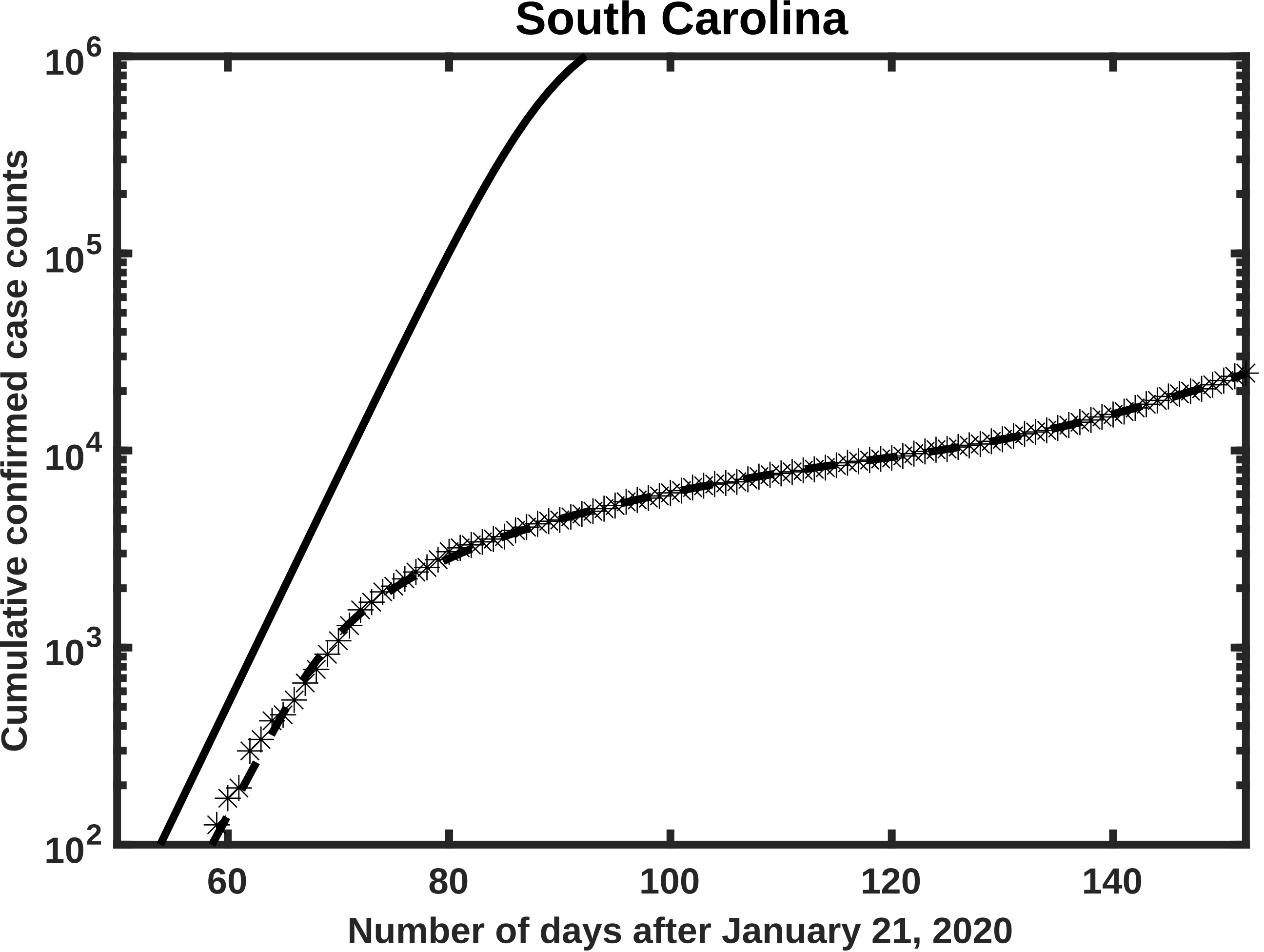
South Carolina
Cumulative confirmed case counts
$10^6$
$10^5$
$10^4$
$10^3$
$10^2$
60
80
100
120
140
Number of days after January 21, 2020

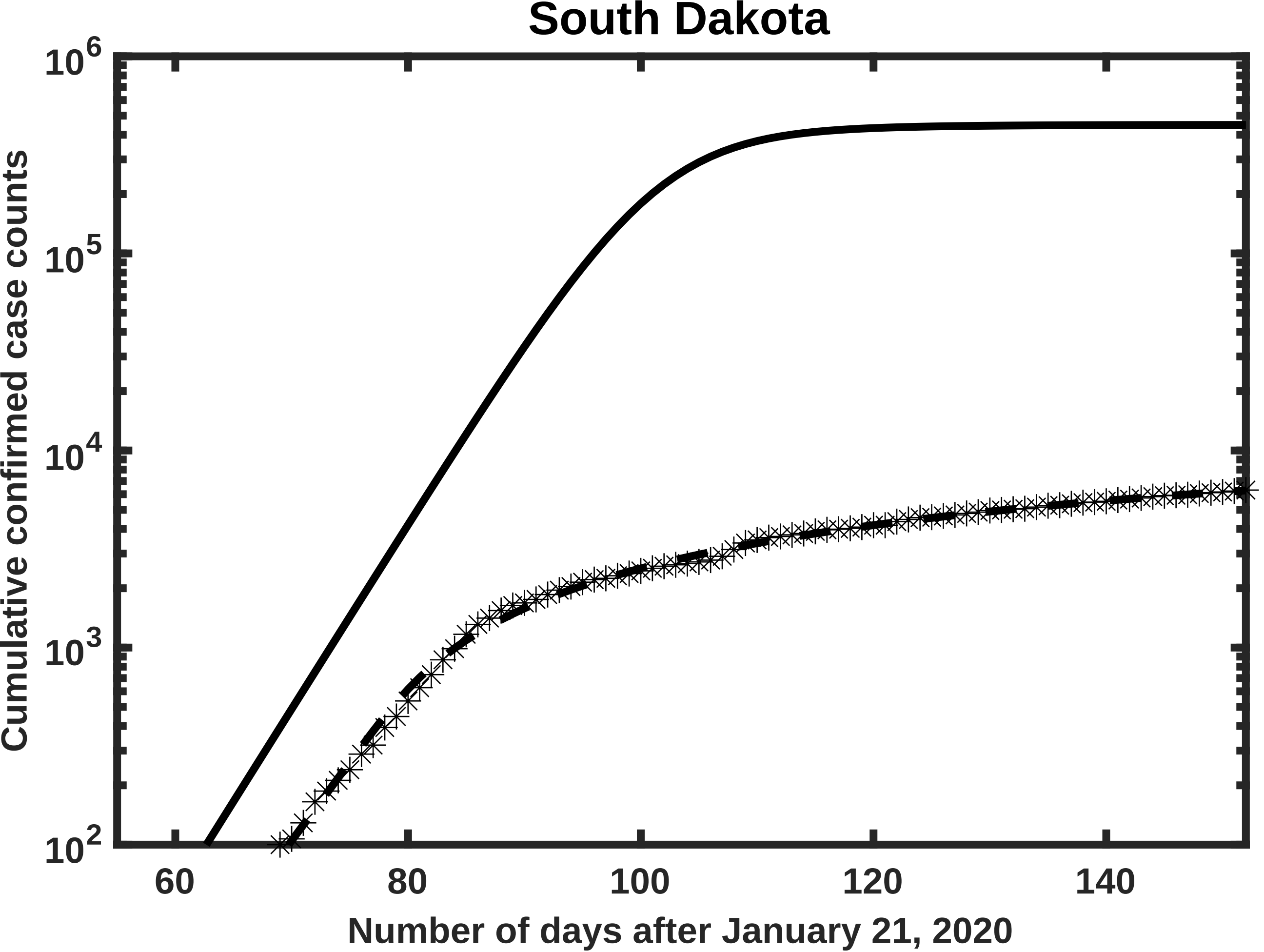
South Dakota
Cumulative confirmed case counts
$10^6$
$10^5$
$10^4$
$10^3$
$10^2$
60
80
100
120
140
Number of days after January 21, 2020

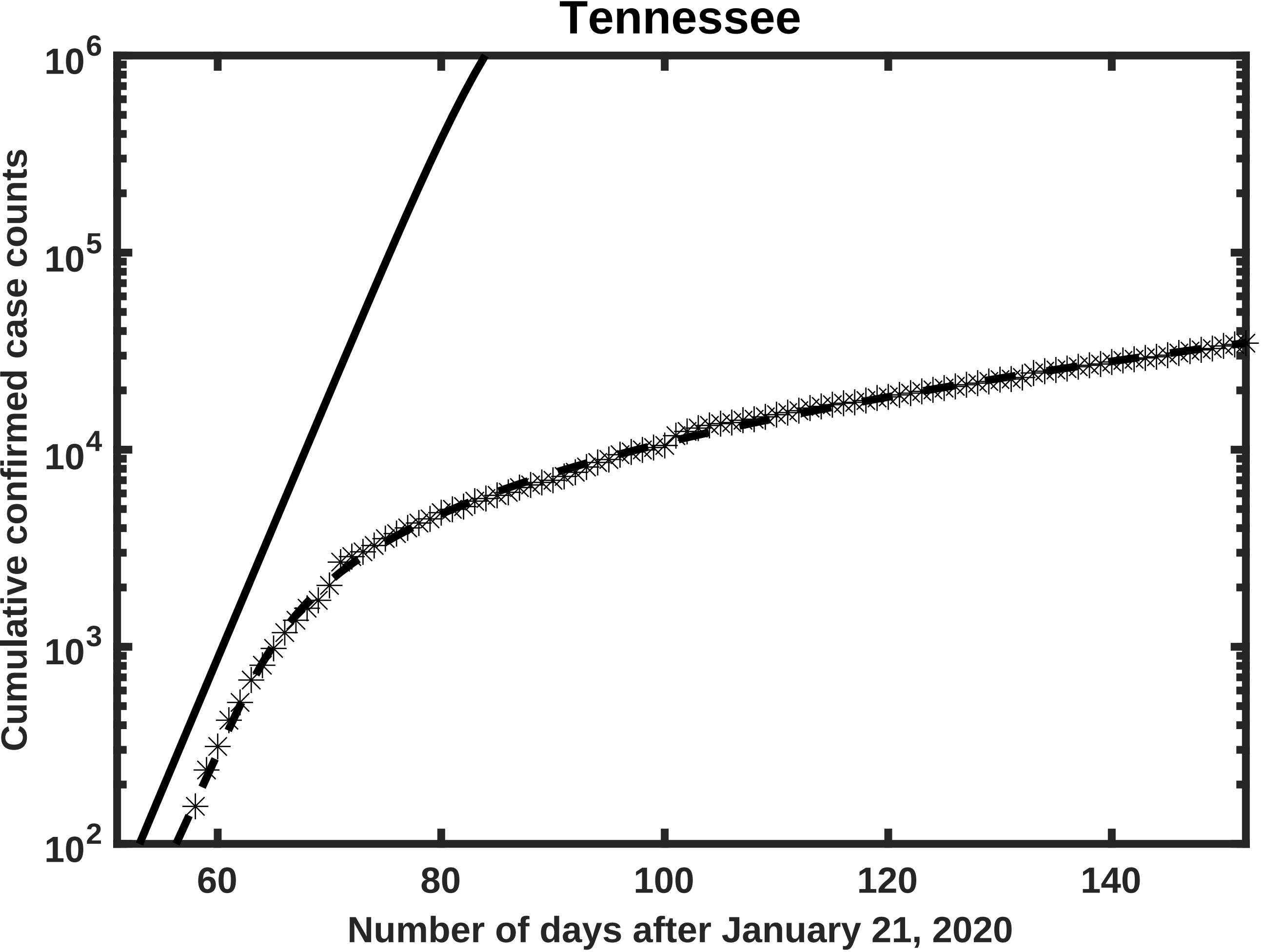
Tennessee
$10^6$
$10^5$
$10^4$
$10^3$
$10^2$
Cumulative confirmed case counts
60
80
100
120
140
Number of days after January 21, 2020

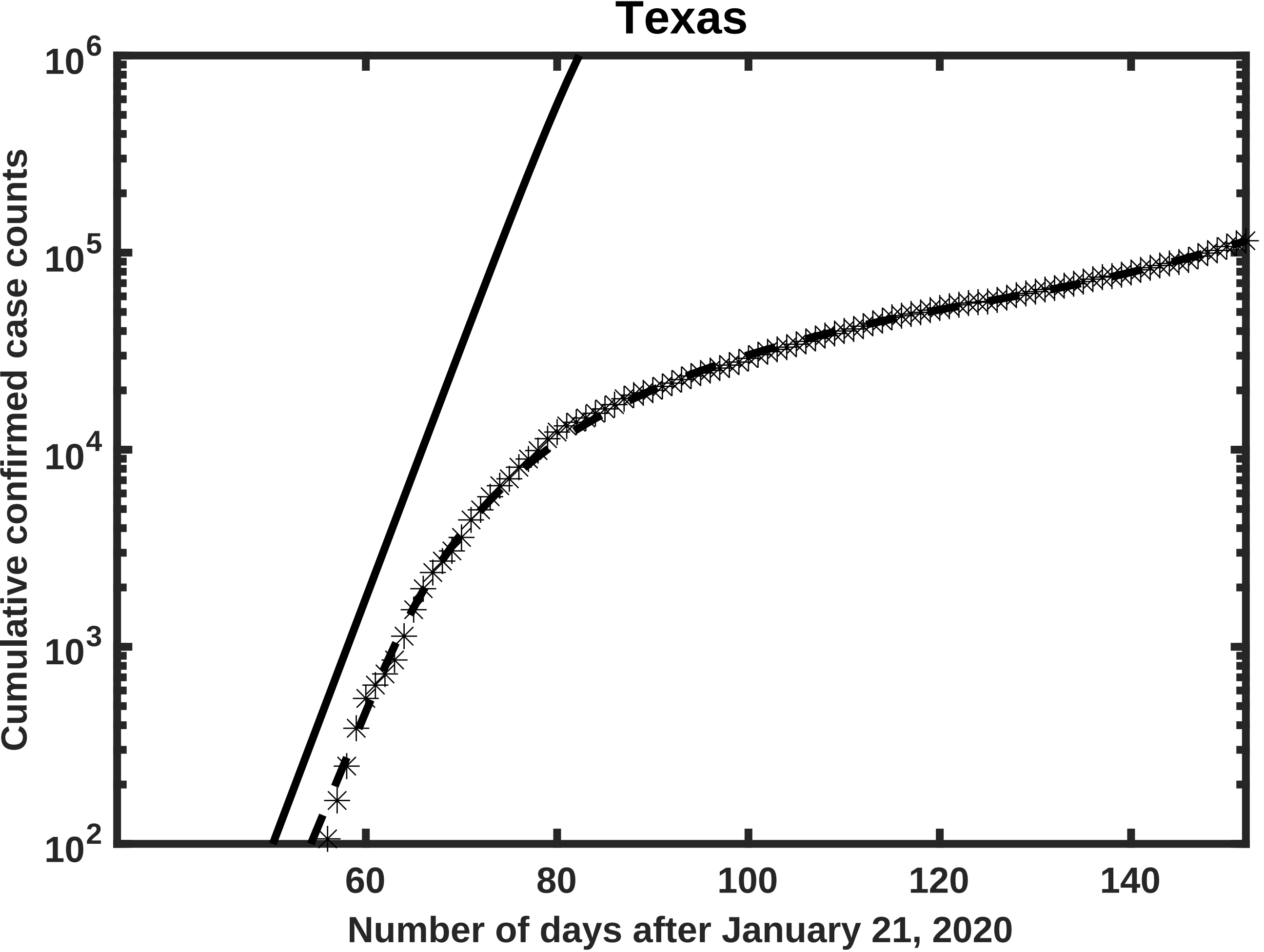

Texas
Cumulative confirmed case counts
Number of days after January 21, 2020
$10^6$
$10^5$
$10^4$
$10^3$
$10^2$
60
80
100
120
140

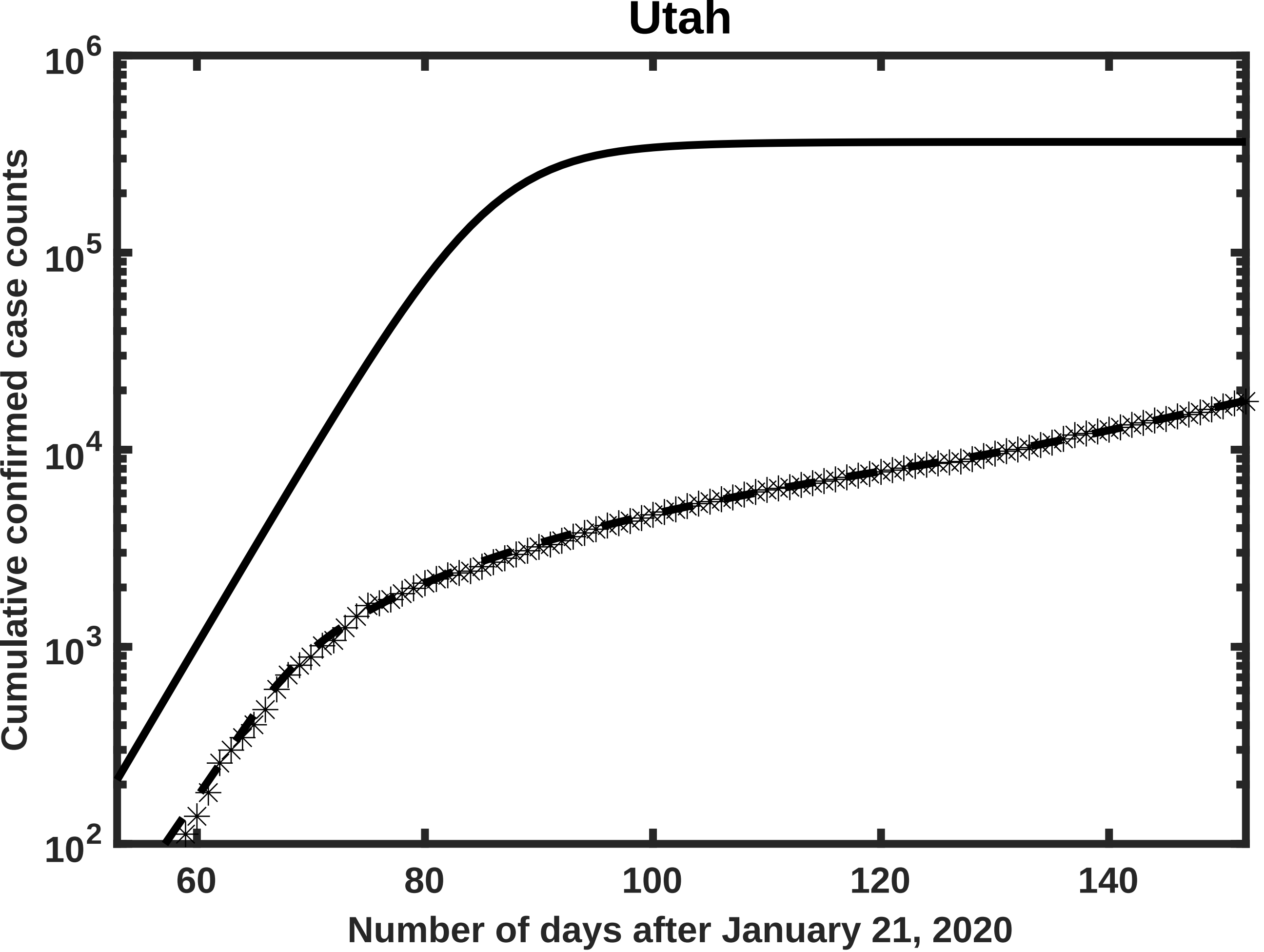

Utah
$10^6$
$10^5$
$10^4$
$10^3$
$10^2$
Cumulative confirmed case counts
60
80
100
120
140
Number of days after January 21, 2020

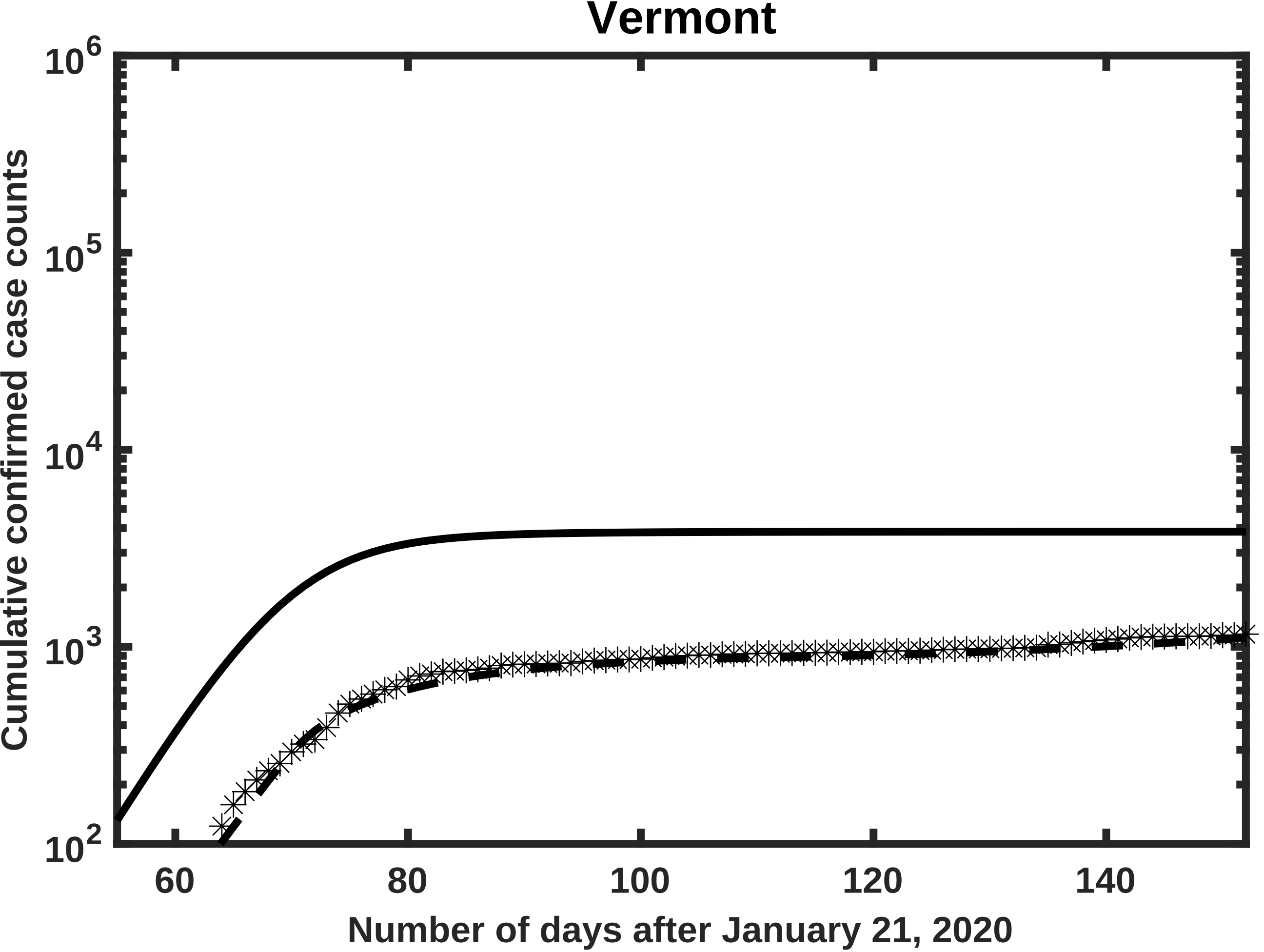

Vermont
Cumulative confirmed case counts
$10^6$
$10^5$
$10^4$
$10^3$
$10^2$
60
80
100
120
140
Number of days after January 21, 2020

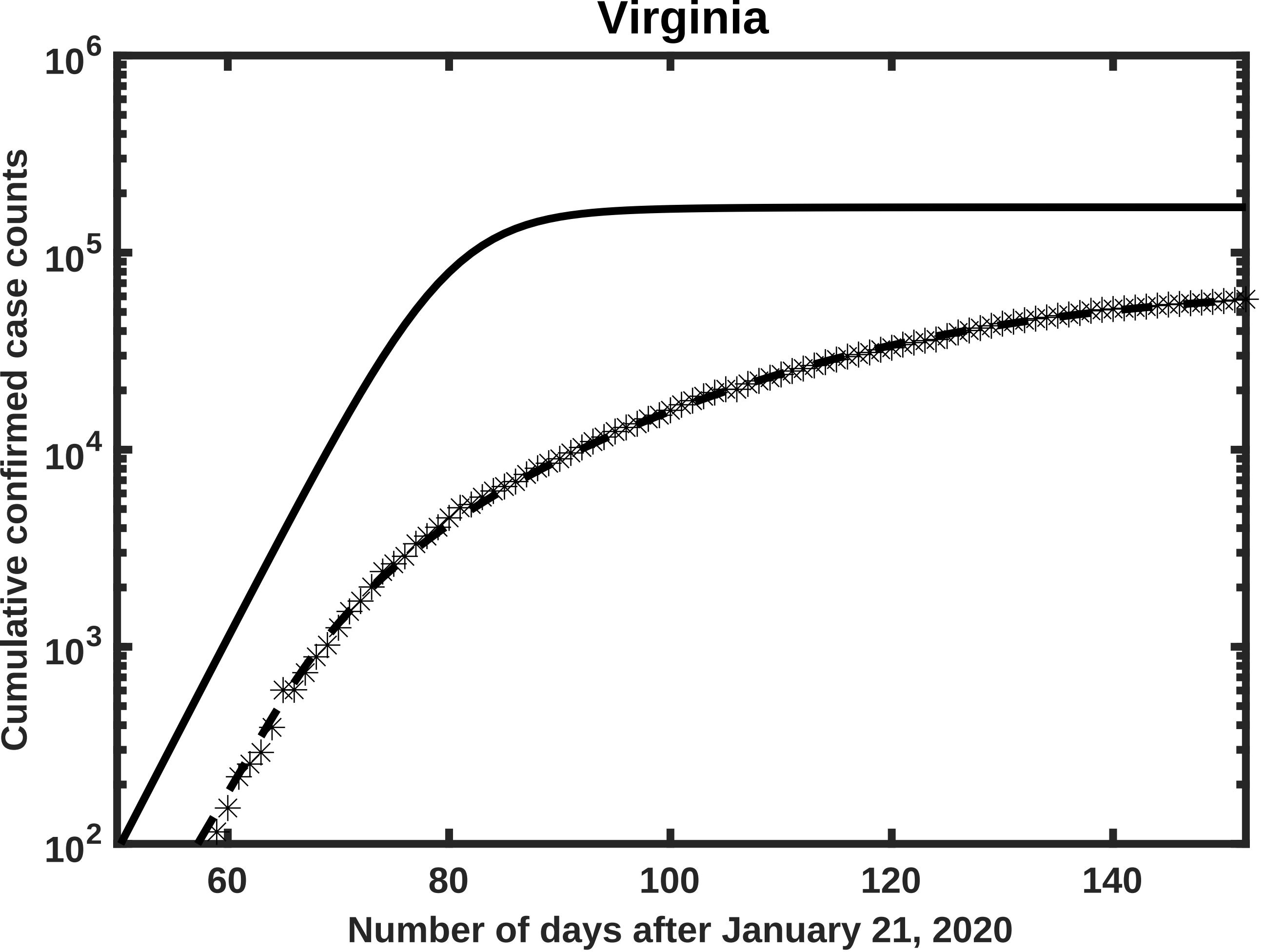
Virginia
$10^6$
$10^5$
$10^4$
$10^3$
$10^2$
60
80
100
120
140
Cumulative confirmed case counts
Number of days after January 21, 2020

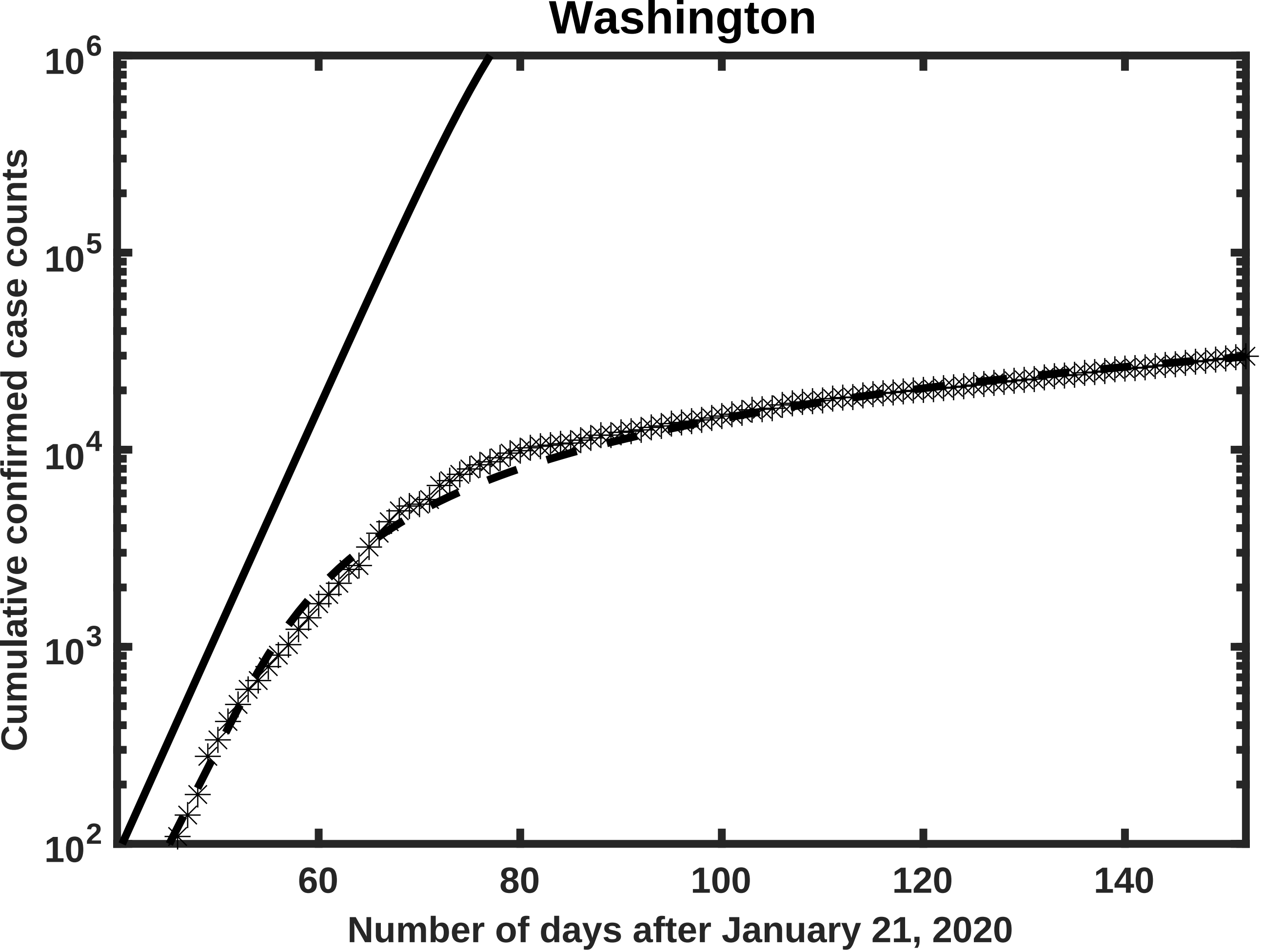
Washington
Cumulative confirmed case counts
10^6
10^5
10^4
10^3
10^2
60
80
100
120
140
Number of days after January 21, 2020

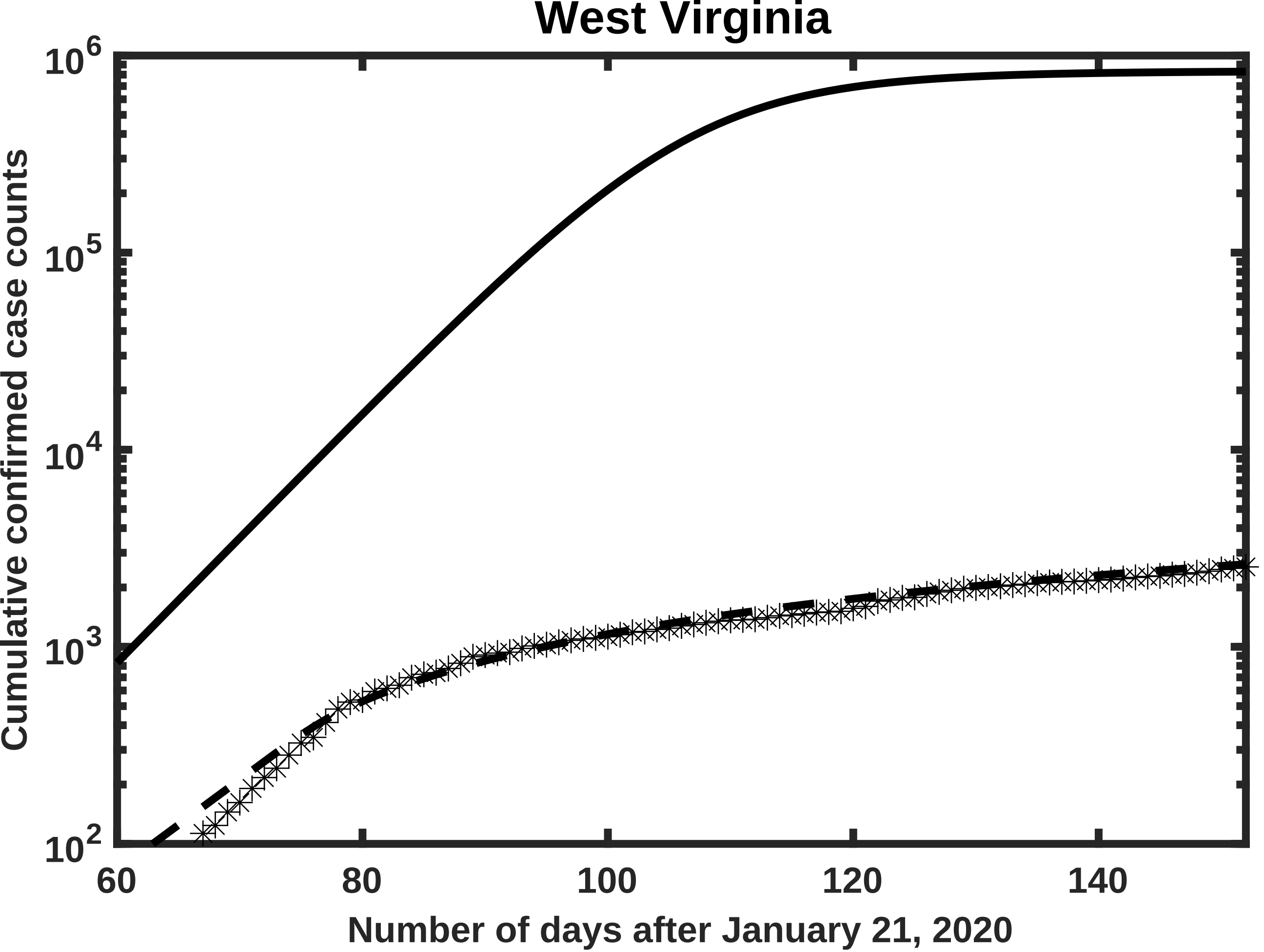

West Virginia
Cumulative confirmed case counts
10^6
10^5
10^4
10^3
10^2
60
80
100
120
140
Number of days after January 21, 2020

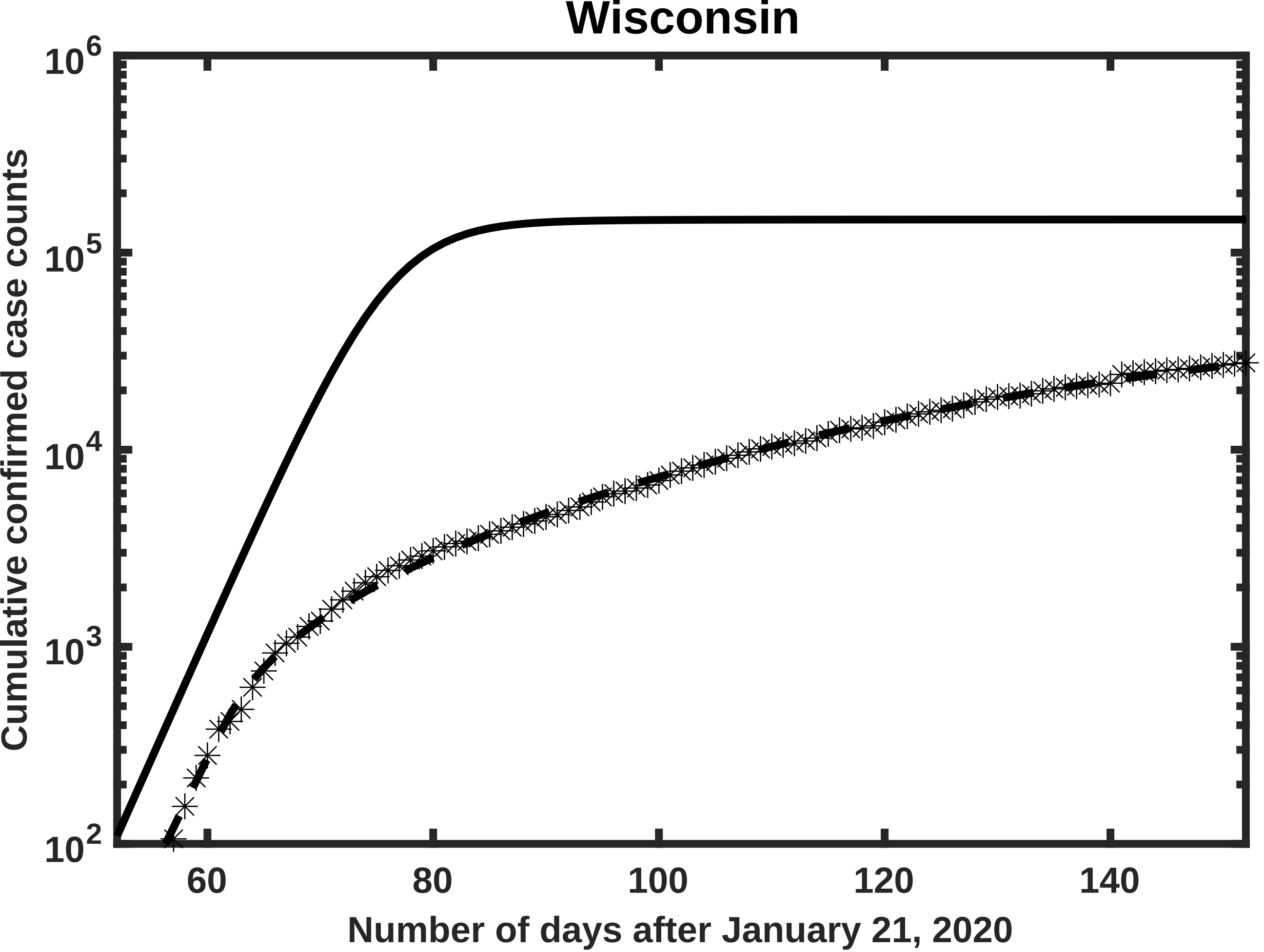
Wisconsin
Cumulative confirmed case counts
$10^6$
$10^5$
$10^4$
$10^3$
$10^2$
60
80
100
120
140
Number of days after January 21, 2020
